# Sex and gender differences in OCD: A scoping review and a two subtypes hypothesis

**DOI:** 10.1101/2025.02.08.25321933

**Authors:** Meghan Van Zandt, Emily Olfson, Brittany Stahnke, Samantha Taylor, Aleigh Wise, Cheyenne Williams, Michael H. Bloch, Christopher Pittenger, Helen Pushkarskaya

## Abstract

This scoping review explores sex and gender differences in obsessive-compulsive disorder (OCD) as reported in the literature between 2009 and 2026. A comprehensive PubMed search was conducted without restrictions, using terms related to sex, gender, and OCD. 8,394 articles were screened by two reviewers, with 1,773 assessed at the full-text level. Of these, 1,038 were included: 72 meta-analyses, 19 systematic reviews, 47 narrative reviews, 3 conceptual and integrative reviews, and 897 original reports. Original reports were categorized by focus — symptomology and burden (120), comorbidities (231), epidemiology (255), human genetics (101), altered neurocognitive and neurobiological functioning (74), treatment (86), and animal models (63) — and assessed for quality using CASP checklists. There was abundant research on sex and gender effects in OCD, but limited synthesis. Identified methodological concerns included inconsistent assessments of sex and gender, variability in OCD symptom measurement, failure to account for menstrual cycle effects, and a lack of genome-wide and full-brain imaging studies. Biases in results may arise from not accounting for gender differences in insight, help-seeking behaviors, and comorbidity. We identify critical knowledge gaps and develop hypotheses for future research, incorporating recommendations for precision medicine approaches. Investigating sex and gender differences in OCD can advance equitable mental health care by addressing the specific needs of diverse populations. Appreciating these effects offers a pathway to better understanding the complexity of OCD, fostering targeted interventions and a more nuanced approach to treatment and prevention.

## 1. Introduction

Obsessive-compulsive disorder (OCD) affects 2–3% of the global population and contributes significantly to worldwide morbidity (Kessler et al., 2012; Murray et al., 1996), severely impacting both the lives of those directly affected and their families and creating substantial burdens on individuals and society at large (Markarian et al., 2010). The effectiveness of evidence-based treatments remains limited (Pittenger, 2023; Reid et al., 2021). Consequently, advancing our understanding of OCD—its diagnosis, underlying mechanisms, and treatment, including the development of precision medicine approaches— is a public health imperative.

The clinical manifestations of OCD are markedly variable. Patients differ in their specific symptoms (Lochner & Stein, 2003), the experienced motivation behind these symptoms [incompleteness/harm avoidance (Ecker & Gönner, 2008)], comorbidity (Diniz et al., 2004), and natural history (Selles et al., 2014). There is a growing appreciation that an individual’s sex and gender may contribute to this heterogeneity (Cherian et al., 2014; Labad et al., 2008; Mathes et al., 2019). Incorporating a ’gendered’ perspective may enhance diagnostic accuracy and treatment selection, for both men and women (Franceschini & Fattore, 2021). Notably, the World Health Organization’s Mental Health Action Plan 2013– 2020 highlighted the critical need for age- and sex-specific data in child mental health (Saxena et al., 2015), supporting such an approach.

Views on sex and gender differences in OCD have evolved over the decades. Early literature argued against the existence of significant effects of sex and gender in OCD (Castle et al., 1995), but more recent reviews have acknowledged mounting evidence supporting their existence and importance (Lochner et al., 2004; Lochner & Stein, 2001; Mathis et al., 2011). An initial focus was on sex and gender effects in the epidemiology and clinical presentation of OCD. More recent literature has increasingly explored differences between men and women across a wider range of domains, drawing on data from self-reports, clinical assessments, behavioral studies, neuroimaging, and biological samples.

In this scoping review, we summarize both original research and literature reviews focused on sex and gender influences on all aspects of OCD over a decade (from July 1^st^, 2009 to May 31^st^, 2026). We have several key goals: (1) to provide a comprehensive and systematic survey of evidence concerning sex and gender effects in OCD; (2) to assess methodological considerations in this body of research; (3) to identify knowledge gaps; and (4) to propose a roadmap for future hypothesis-driven investigations into sex and gender effects in OCD, incorporating recommendations for precision medicine approaches.

## 2. Methods

This study was conducted using the Preferred Reporting Items for Systematic Reviews and Meta-Analyses (PRISMA) guidelines and checklist (Page et al., 2021), as depicted by Figure 1. Its methodology was guided by Arksey and O’Malley (2005). As a scoping review, the study was not preregistered in PROSPERO.

**Figure 1.**
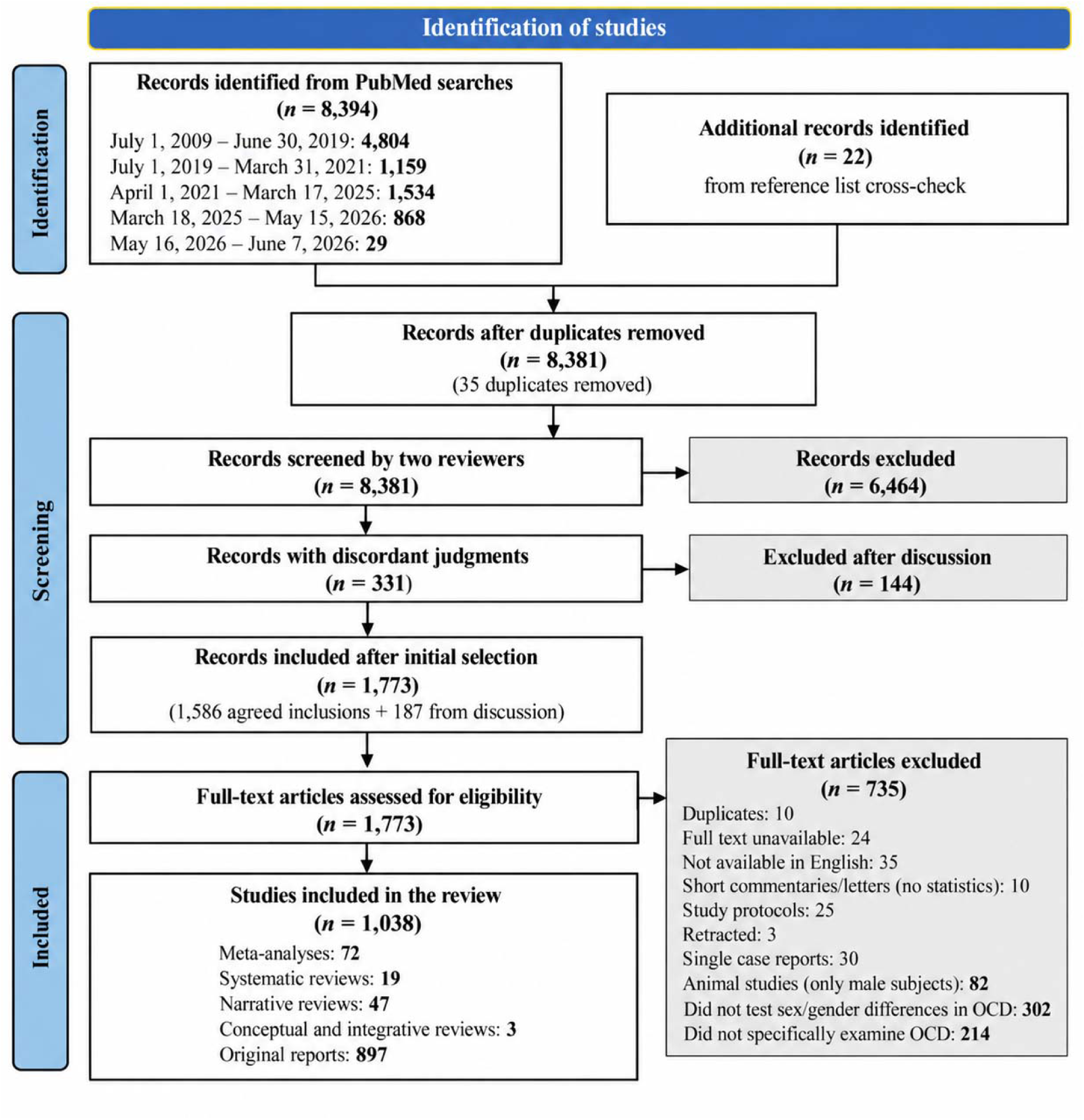
Flowchart of the process of identification of studies for the review.

PubMed was searched with the following search words: (<u>gender</u> OR <u>sex</u> OR <u>male</u> OR <u>female</u>) AND <u>obsessive-compulsive disorder</u>. The search was conducted in five waves. On July 2, 2019, records published from July 1, 2009, to June 30, 2019, were searched; on April 26, 2021, records published from July 1, 2019, to March 31, 2021; and on March 18, 2025, records published from April 1, 2021, to March 17, 2025. Two additional supplementary searches were conducted on May 16, 2026 (covering March 18, 2025, to May 15, 2026) and on August 1, 2026 (covering May 16, 2026, to June 7, 2026).

Included studies:

1. Were published in an English language journal,
2. Met one of the following criteria:

a. Compared OCD to other clinical or non-clinical groups, or
b. Included only individuals with an OCD diagnosis, or
c. Compared OCD-related condition (e.g., tics, anorexia, etc.) to other clinical and non-clinical groups, or
d. Compared a different form psychopathology with comorbid OCD to the same psychopathology without Comorbid OCD; and
3. Met one of the following criteria:

e) Directly compared men with OCD and women with OCD, or
f) Included gender/sex as a moderating factor of the difference between OCD (or other groups of interest described above) and other clinical and non-clinical groups (not just as a control variable), or
g) Included only one sex, men or women (case studies were excluded).

Our search strategy was designed to identify relevant studies in humans, but it also captured a number of animal studies, which we chose to include in this review. Special selection considerations were applied to these studies. We excluded animal studies that only examined male animals; prior to 2015, when use of both male and female animals was made mandatory for NIH-funded research (NIH, 2015), this was common. Studies that examined only female animals were included, if they examined the effects of estrogen, progesterone, or the estrous cycle on OCD-relevant behaviors. Criterion 2 (above) cannot be applied to animal studies and was modified as follows: animal model studies were included if they a) directly compared the effects of sex or the effect of specific sex hormones (e.g. estrogen) on behavior and b) examined brain circuits, signaling pathways or pathological alterations thought to be relevant to OCD. We note that animal studies captured by our search are unlikely to represent a comprehensive summary of all relevant work; it is likely that many animal studies that examine processes of potential relevance do not include the keywords “<u>obsessive-compulsive disorder”</u> and thus may have been missed. This limitation is discussed in more detail in Section 3.8.

During initial selection, two authors (MVZ and HP) independently reviewed each of the papers identified by this PubMed literature search to check whether they met these inclusion criteria. The reference lists in the retrieved articles were checked against the database search, and any other articles that might fulfill the inclusion criteria were retrieved. Any disagreements between MVZ and HP were resolved by group discussion.

Papers selected during initial selection were divided into 7 broad categories (by MVZ), based on the focus of the paper: Symptomology and Burden, Comorbidities, Epidemiology, Genetics, Altered Neurocognitive and Neurobiological functioning, Treatment, and Animal Models. Each category was assigned to one of the authors (MVZ, EO, BS, or HP) for in-depth review (see Figure 1). Each of the researchers prepared a review of the assigned section; if any paper was identified as not meeting inclusion/exclusion criteria or not fitting the assigned category, it was discussed by the group and either excluded or recategorized. HP conducted an additional final review of excluded papers, and any disagreements with prior decisions were discussed with the group. CP and MB conducted the overall editing of the review.

All included papers were assessed by four researchers (BS, ST, AW, and CW) for quality of evidence using the CASP Checklists (Buccheri & Sharifi, 2017; Long et al., 2020); available upon request). Overall patterns, methodological concerns, knowledge gaps, and new hypotheses for future research were discussed and summarized during meetings of all coauthors. These summaries of key considerations are presented at the end of each section.

## 3. Results

The initial PubMed searches identified 8,394 papers: 4,804 papers in the search from July 1, 2009, to June 30, 2019; 1,159 papers in the search from July 1, 2019, to March 31, 2021; 1,534 papers in the search from April 1, 2021, to March 17, 2025; 868 papers in the supplementary search from March 18, 2025, to May 15, 2026; and 29 papers in the supplementary search from May 16, 2026, to June 7, 2026. 35 duplicate records were removed. The reference list cross-check revealed an additional 22 papers. MVZ and HP independently agreed to include 1586 papers and not to include 6464; of the remaining 331 papers on which these two reviewers had discordant judgments, 187 were included following group discussion. Overall, 1773 papers were included during this initial selection step.

During the final selection step, 735 papers were excluded: 10 were duplicates, for 24 full text was unavailable, 35 were not available in English, 10 were short commentaries or letters to the editor with no statistics reported, 25 were study protocols, 3 were retracted, 30 were single case reports, 82 were animal studies with only male subjects, 302 did not test sex/gender differences in OCD (i.e. did not meet the inclusion criteria 3), and 214 did not specifically examine OCD (e.g. focused on obsessive-compulsive personality disorder or pooled OCD with other anxiety disorders).

Overall, 1,038 papers were confirmed to be included: 72 meta-analyses, 19 systematic reviews, 47 narrative reviews, 3 conceptual and integrative reviews, and 897 original reports (Figure 1). A summary of meta-analyses and previous systematic and narrative reviews is reported in Appendix A1. Original reports were reviewed and are reported in seven sections:^1^ the Symptomology and Burden section includes 120 papers (Section 3.2, by BS and HP); the Epidemiology section includes 255 papers (Section 3.3, by MVZ and HP); the Comorbidities section includes 231 papers (Section 3.4, by MVZ and HP); the Human Genetics section includes 101 papers (Section 3.5, by EO); the Altered Neurocognitive and Neurobiological functioning section includes 74 papers (Section 3.6, by HP); the Treatment section included 86 papers (Section 3.7, by BS and HP), and the Animal Models section includes 63 papers (Section 3.8, by MVZ). Thirty three papers are included in more than one section. Figure 2 illustrates that the number of papers on most topics increased over time during the review period, with the most dramatic increase in epidemiological studies (including comorbidities).

**Figure 2.**
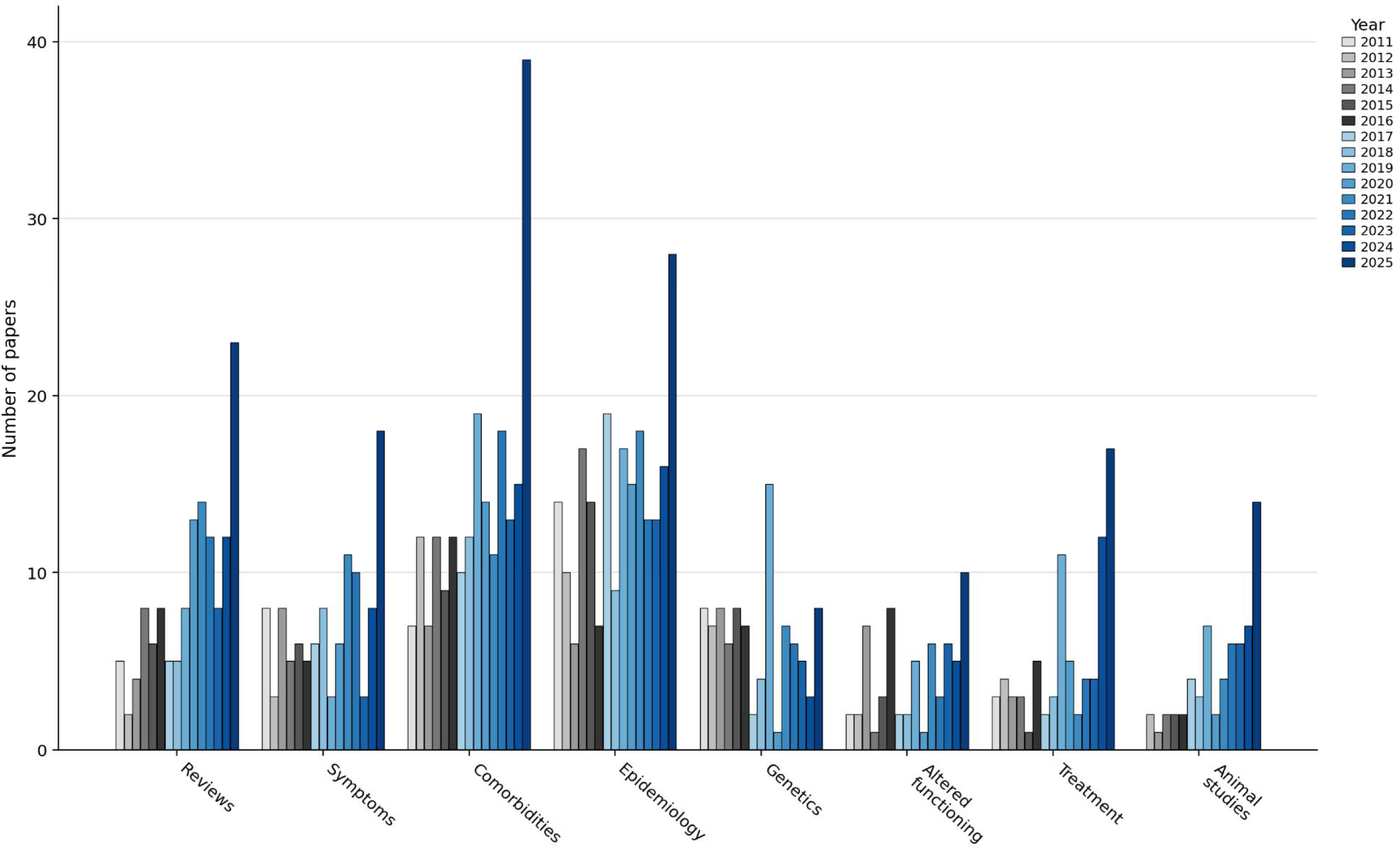
The number of papers on each reviewed topic is presented by year. Publications from 2010 and 2025 are excluded as the review period did not cover all months of these years. In 2015, the NIH mandated the use of both male and female animals in funded research. A two-year lag was included to account for the time required to publish results. Data from years before the policy’s impact are color-coded in grey, while data following the policy’s impact are shown in blue.

Throughout the review, we cited each included paper by the first author’s last name and publication year. Due to the very large number of included papers (N = 912), these papers are listed in the appendices rather than the references. Also, due to the very large number of papers included in the review, only selected key methodological concerns, knowledge gaps, and new hypotheses for future research are presented at the end of each section. Our hope is that by characterizing the overall patterns that emerged in the literature, we will facilitate generation of new hypotheses, more systematic investigations, and a broader discussion on how sex and gender may contribute to the heterogeneity of OCD.

### 3.1. A note on sex and gender

Sex and gender are distinct concepts. Both are complex. ’Sex’ refers to an individual’s biological classification as male or female, determined by sex chromosomes, anatomy, and sex steroid hormones; these usually coincide and allow for ready classification as ‘male’ and ‘female’, but ambiguity arises in edge cases. ’Gender’ relates to individuals’ social identification as men, women, or nonbinary/other and is influenced by sex and by family, society, and culture (Holdcroft, 2007; Lindqvist et al., 2021; Mendrek, 2015). Sex assigned at birth and current gender identification align in most cases; discrepancies can occur in intersex individuals or those identifying as transgender or nonbinary. The interplay between sex and gender influences behavior and brain function over the lifetime. Amongst the hundreds of reviewed studies, only one reported asking participants to report both sex, as assigned at birth, and gender identity. In all other studies, participants were simply asked to report whether they were men (or male) or women (or female); in most studies the details of how this question was posed or a determination was made are not available. For the purpose of this review, the assumption was made that, unless otherwise explicitly reported, human genetic studies and animal model work assessed the effects of sex and all other studies assessed participants’ self-reported gender.

Following APA 7 guidelines (Transue, 2019) terms *male* and *female* are used as nouns when describing a group with a wide age range or as adjectives to describe a specific person. Terms *men* and *women* or *boys* and *girls* are used to describe individuals from a specific age group.

### 3.2 Symptoms and Associated Burden

#### Highlights from Section 3.2 Symptoms and Associated Burden

<u>Symptom presentation</u>: Overall severity is often, but not uniformly, higher in females. Girls may show broader burden in selected domains, while adult gender differences appear more in symptom organization.

<u>Associated burden:</u> Burden is domain-specific. Men show greater loneliness and labor-market marginalization; women show greater marital-role difficulties, poorer quality of life in some domains, and greater family accommodation in some studies.

<u>Key concerns:</u> Findings vary by assessment method, insight may affect reporting, and developmental changes in symptom organization remain understudied. Gender minorities remain substantially understudied in phenomenological studies.

<u>Topics for future study:</u> Gender differences in symptom subtypes and insight; gender differences in subthreshold symptoms and diagnostic thresholds; assessments tailored to developmental and life stages (e.g., puberty, geriatric); relationships between maternal OCD and offspring physical health; and gender differences in help-seeking among youth with OCD.

#### 3.2.1 Symptom Presentation

The clinical presentation of OCD is marked by high heterogeneity, some of which may be linked to sex or gender (Franceschini & Fattore, 2021; Mathis et al., 2011). Initial efforts to classify the heterogeneity of OCD symptoms were based on symptom content and converged on 3–5 dimensions: contamination/cleaning, fear of harm/checking, taboo thoughts, and symmetry/ordering (Bloch et al., 2008). (Hoarding emerged as a separate dimension in early studies but is now considered a separate disorder.) Seventy-nine studies from the review period addressed gender-relevant aspects of OCD symptom presentation and assessment (Table 1, Table A2.1). Of these, 53 studies examined symptom presence or severity (3 across all ages, 15 in children/adolescents, and 35 in adults), 3 examined motivations underlying symptoms (1 across all ages, 1 in children/adolescents, and 1 in adults), 8 used latent-class or other data-driven approaches to examine symptom cooccurrence (2 in children/adolescents and 6 in adults), and 15 focused on assessment-related questions (1 across all ages, 4 in children/adolescents, and 10 in adults).

**Table 1.**
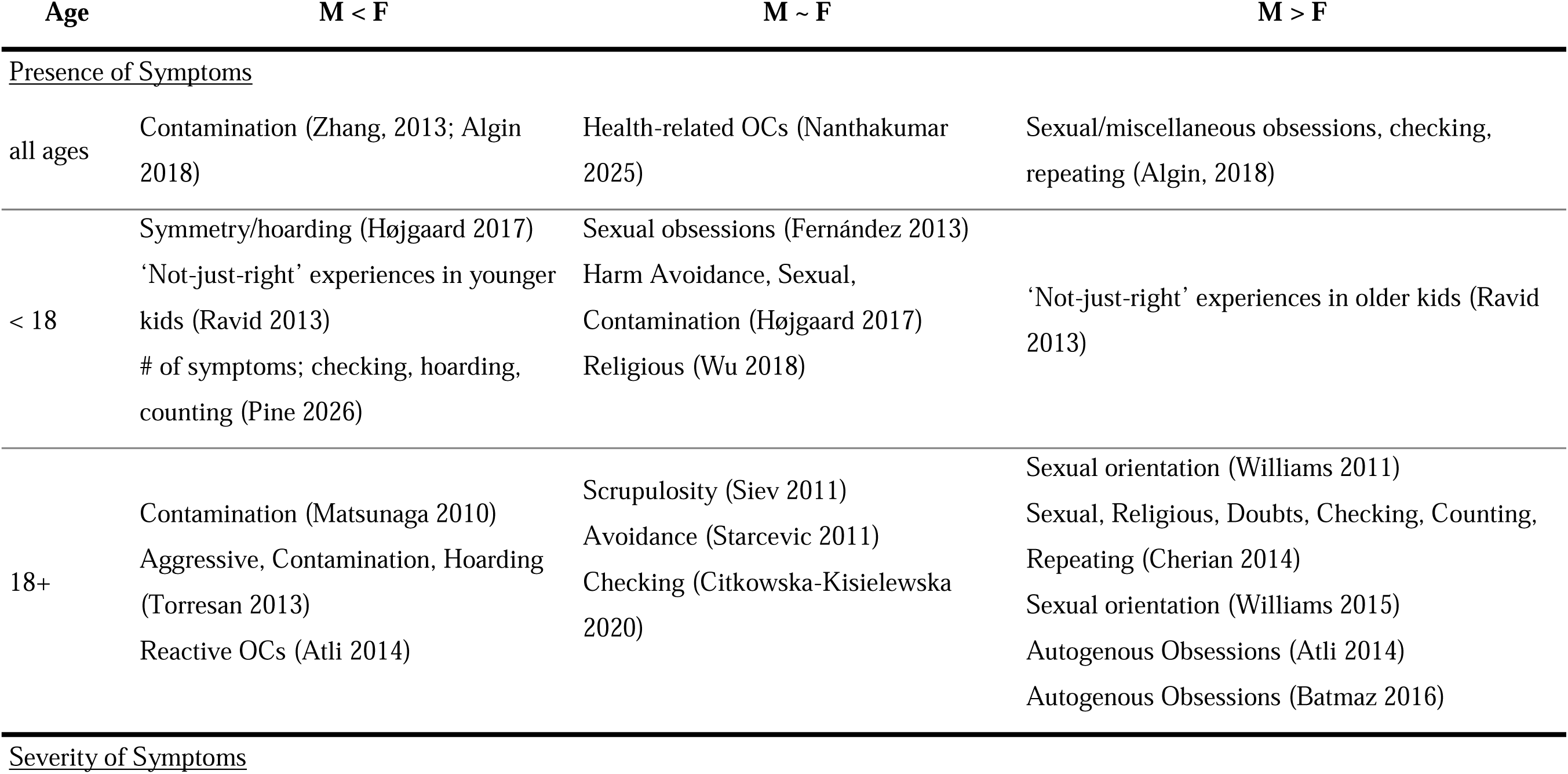
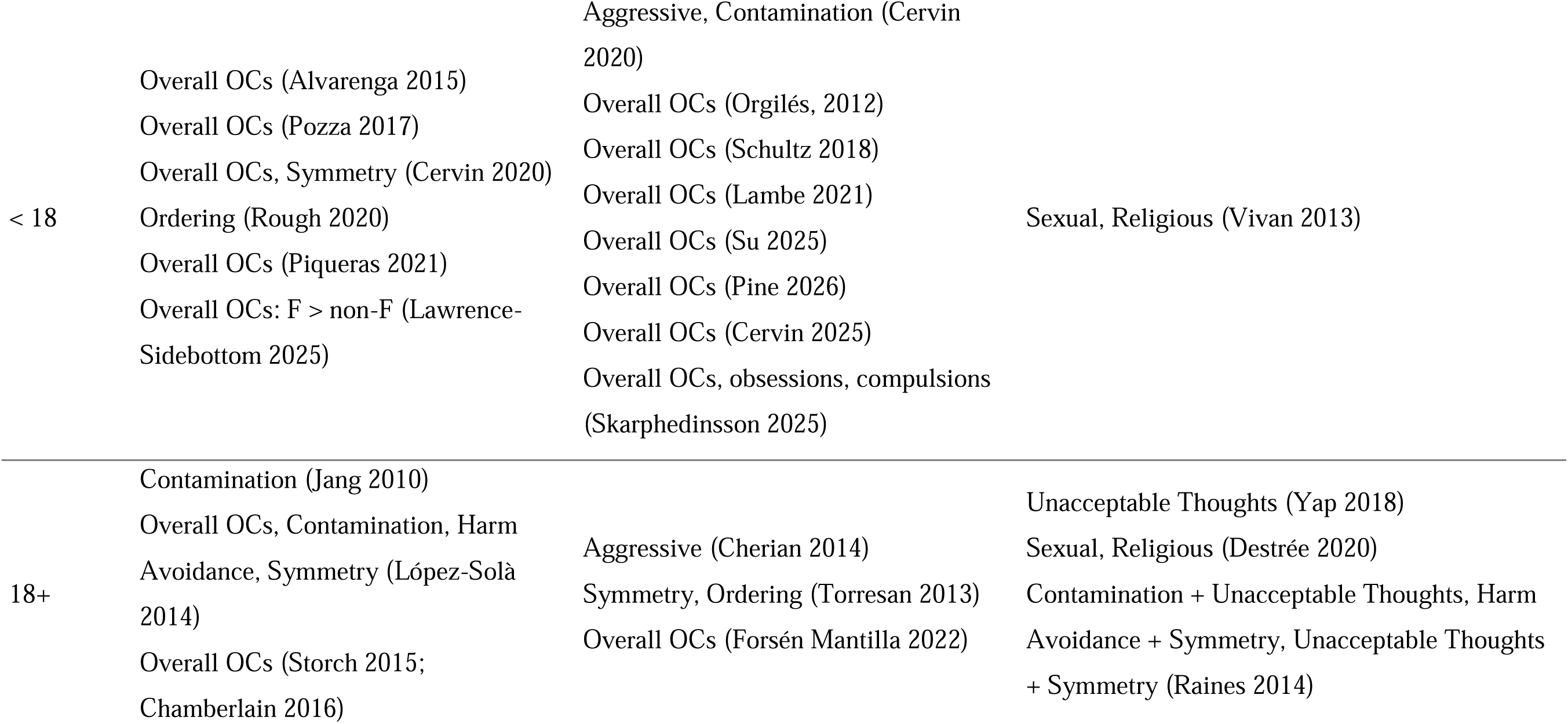
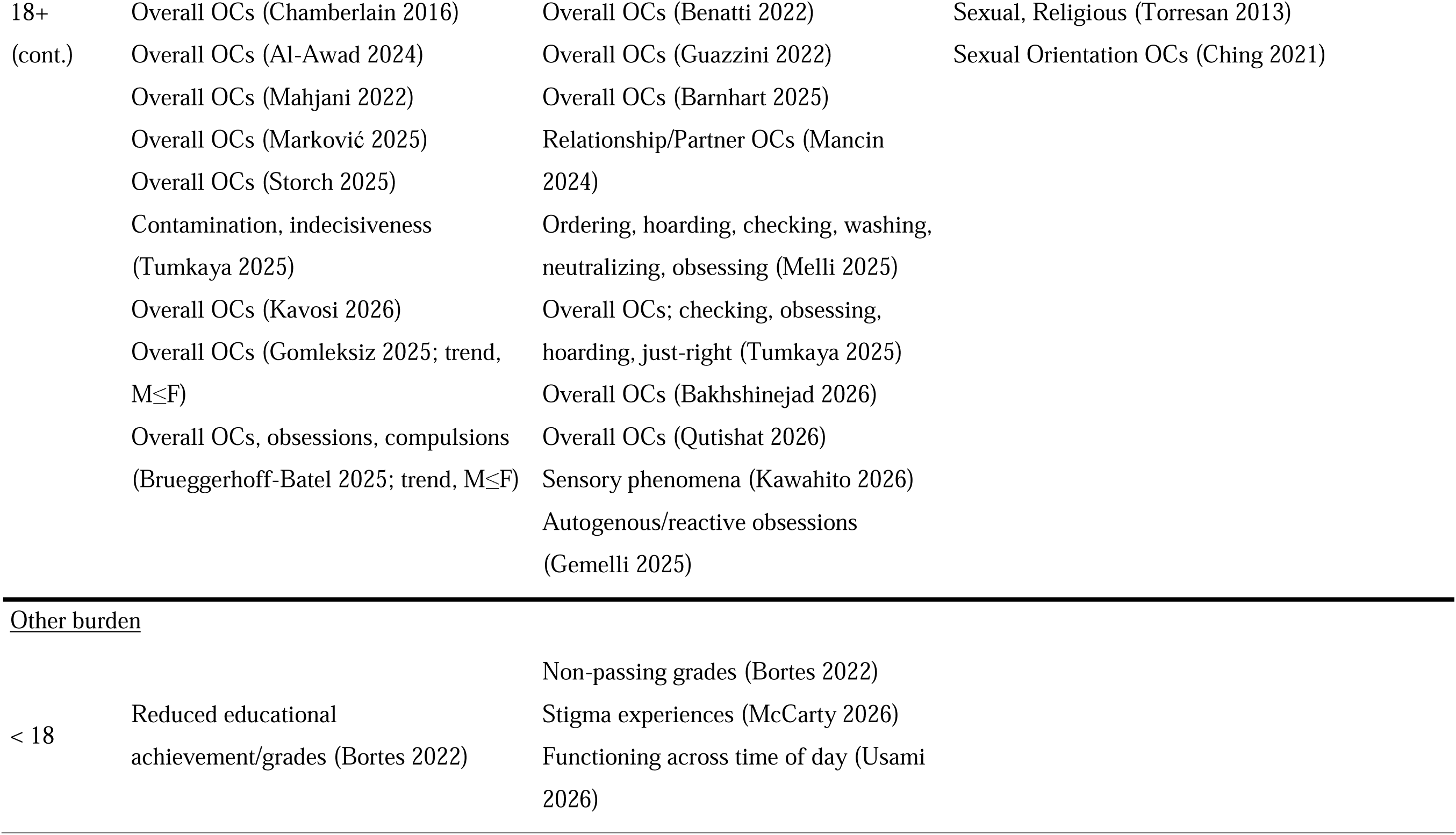
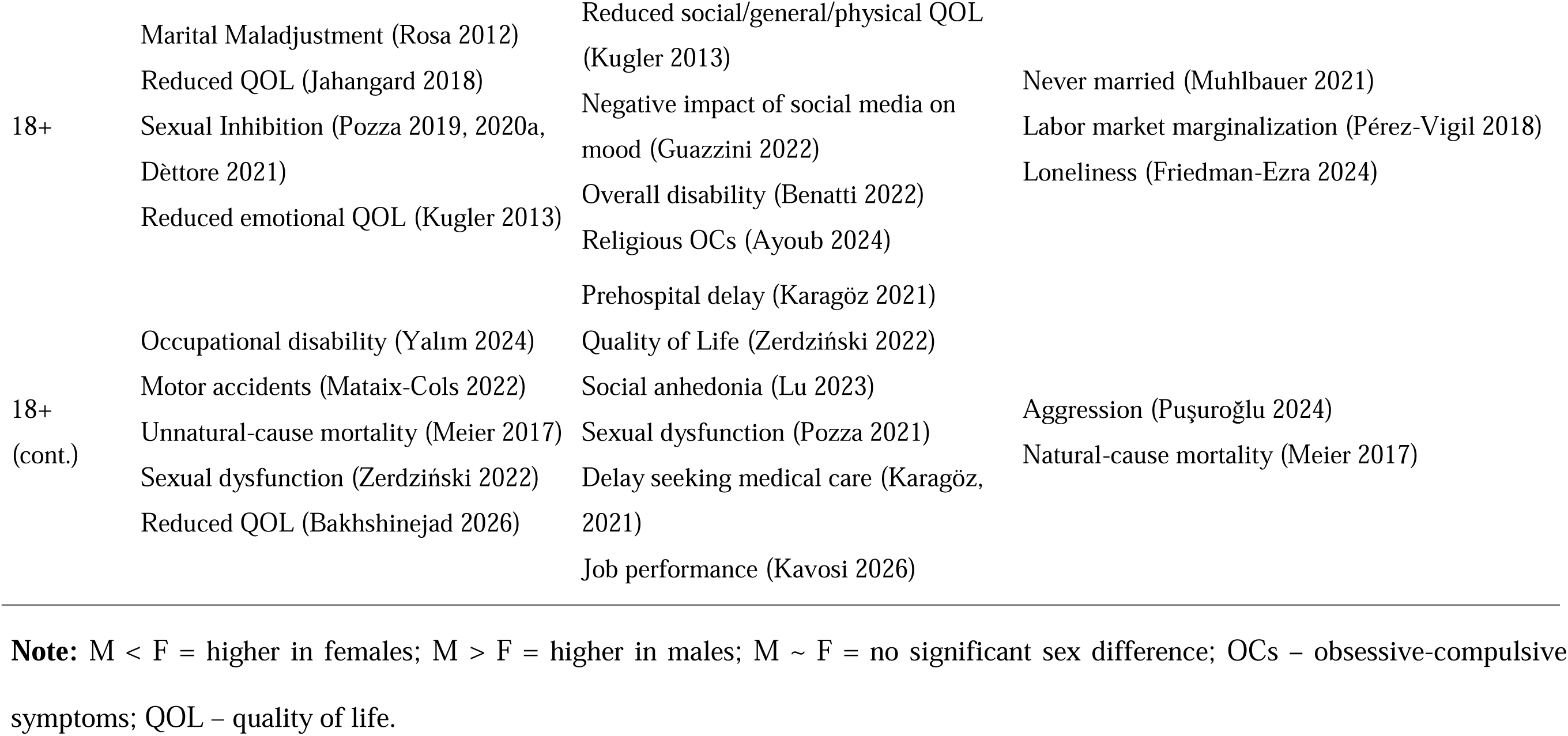
Gender differences in symptoms and other burden in individuals with Obsessive-Compulsive Disorder.

When interpreting symptom patterns, it is important to consider how symptoms were assessed. In studies spanning all ages, all 5 used clinician-administered measures. In children/adolescents, 10 studies used clinician- or interview-administered measures and 12 used self- or parental reports; in adults, 21 used clinician- or interview-administered measures, 28 used self-reports, and 3 used both. For overall symptom severity, self- and parental-report studies yielded more variable estimates across age groups: among studies reporting standardized mean differences, F−M effects ranged from d = −0.10 to +0.12 in children/adolescents and from approximately null to d = +0.44 in adults, with additional studies reporting nonsignificant or unstandardized effects. Clinician-administered assessments in children/adolescents showed either greater severity in females or no gender difference, whereas adult clinical assessments showed a more consistent tendency toward greater severity in females. Two studies in adults (Hauschildt 2019; Storch 2017) compared self-reports and clinical assessments using the Yale-Brown Obsessive-Compulsive Scale, YBOCS (Goodman, Price, Rasmussen, Mazure, Delgado, et al., 1989; Goodman, Price, Rasmussen, Mazure, Fleischmann, et al., 1989). In both studies, self-reported YBOCS scores were lower than clinician ratings; in Hauschildt (2019), this discrepancy decreased with repeated assessments. The correspondence between self- and clinician ratings did not differ by gender. One study of children and adolescents with OCD (ages 6–18; Skarphedinsson, 2021) found that parental reports predicted clinical diagnosis substantially better than youth self-reports (AUC_parent_ = 0.89 [95% CI, 0.85–0.92] versus AUC_youth_ = 0.66 [95% CI, 0.57–0.74]). In the same study, girls self-reported higher overall symptom severity than boys, whereas no significant gender difference was observed in parental reports.

Overall OCD symptom severity was often, but not uniformly, higher in females than in males across clinical and community samples; significant male-higher effects were not observed, although some null studies showed small numerical differences in that direction (Table A2). In samples with clinically significant OCD symptoms, seven studies reported significantly higher overall severity in females than in males (three using self-report measures and four using clinician-administered assessments), whereas six studies found no significant gender difference (two using self- or parent-report measures and four using clinician-administered assessments). One additional clinical study showed a female-higher trend (Brueggerhoff-Batel, 2025; p = .07), and one found a significant association with sex but did not report the coding needed to determine direction (Huang, 2025). In community or nonclinical samples, six studies reported significantly higher overall symptom severity in females (five using self- or parent-report measures and one using a clinician-administered assessment), whereas seven studies—all using self- or parent-report measures—found no significant gender differences.

The prevalence and severity of specific OCD symptom domains differed across age groups (Table 1). Some gender differences were more apparent in children and adolescents: girls showed higher prevalence or severity of symmetry (β = −0.99; Cervin, 2020), symmetry/hoarding (β = −0.095, p < 0.001; Højgaard, 2017), hoarding, checking, and counting symptoms (Pine et al., 2026), and ordering symptoms (t = 2.13, p = 0.03; Rough, 2020). Pine et al. (2026) also found a greater total number of OCD symptoms in girls than in boys (t = 3.02, p = 0.003). None of the studies reported a symptom domain as significantly more prevalent or severe in boys. These patterns were less consistent in adults: symmetry/ordering findings ranged from no gender difference (Yap, 2018; Tumkaya, 2025) to greater severity in women (Cohen’s d = 0.23–0.53; Torresan, 2013; López-Solà, 2014), whereas checking/counting findings were null or occasionally reversed, with Cherian et al. (2014) reporting greater prevalence in men (M:F OR = 2.15 for checking and 2.25 for counting). Conversely, gender differences in contamination/washing and sexual/religious symptoms were generally absent or inconsistent in children but were more consistently observed in adults. Among adults, contamination/washing symptoms were more frequent in women (F:M OR = 1.56–1.70, where reported; Torresan, 2013; Cherian, 2014) and more severe (Cohen’s d = 0.29–0.40; Torresan, 2013; López-Solà, 2014), with additional female-higher findings reported by Matsunaga (2010) and Jang (2010). Sexual/religious and sexual-orientation symptoms were more frequent in men (F:M OR ≈ 0.35– 0.70; Williams, 2011; Cherian, 2014; Torresan, 2013) and were also more severe in several studies (Cohen’s d = 0.16–0.43; β = 0.90; Torresan, 2013; Destrée, 2020; Williams, 2015). Findings for the remaining symptom domains were largely null, inconsistent, or too sparsely reported to establish a clear gender pattern. Evidence for gender differences in compulsion-only OCD subtypes was mixed (Rodgers, 2015).

Lawrence-Sidebottom et al. (2025) was the only study to examine gender-nonconforming adolescents directly; gender-nonconforming youth were more common among those with elevated than non-elevated OC symptoms (8.2% vs. 4.0%, p = 0.001).

Overall patterns suggest that gendered symptom profiles may vary across development (see Table 1 and Section 3.3.2), although differences in sampling (e.g., help-seeking vs. population-based samples) and assessment methods (e.g., self-/parent-report vs. clinician-administered measures) may contribute to these patterns. Direct tests of developmental changes in symptom presentation and co-occurrence from childhood through adulthood are lacking. Reflecting the possibility that symptom presentation may also vary across later life stages, Klenfeldt et al. (2014) called for assessment tools tailored to geriatric OCD, including measures sensitive to checking symptoms in older men and women.

Another classification approach considers underlying motivational factors, such as incompleteness or “not-just-right” experiences and harm avoidance, to gain insight into the driving forces behind OCD symptoms (Ecker & Gönner, 2008; Summerfeldt et al., 2014). Three studies examined gender differences in these motivational or related sensory phenomena. Among adolescents, Ravid (2013) identified age-by-gender interactions for both the presence (β = −0.21, p < 0.01) and severity (β = −0.29, p < 0.01) of “not-just-right” experiences, which were more prominent in younger females and older males. In adults, Sibrava (2016) found no gender difference in the likelihood of high versus low incompleteness (M:F OR = 1.05, p = 0.43), and Kawahito et al. (2026) found numerically greater sensory-phenomena severity in females, but the difference was not significant (β = 1.38, p = 0.12). Note that “not-just-right” experiences have been reported more frequently in OCD patients with comorbid tic disorders and/or autism spectrum disorder (da Silva Prado et al., 2008; Holzer et al., 1994; Neal & Cavanna, 2013), conditions that are themselves diagnosed more often in males (see Section 3.3). More careful analyses accounting for comorbid autism-spectrum and tic disorders are needed to clarify these relationships (see Section 3.4).

Data-driven studies suggest limited gender differences in OCD symptom organization in childhood, with potentially greater divergence in later developmental stages. In pediatric samples, Kenezloi (2018) found no gender differences in the distribution of symptom subtypes (ages 10–18). In youth from the Philadelphia Neurodevelopmental Cohort (ages 12–21), network analyses identified dimension-specific differences, with arranging/ordering more central in males and cleaning/washing more central in females (Wang, 2021). In adults, several studies suggested more pronounced gender differences in symptom organization. Raines (2014) found stronger intercorrelations among several symptom dimensions in men, whereas symptom associations in women were less uniformly interconnected. Shkalim (2017) likewise found gender differences in how OCD symptoms related to fear- and distress-related processes. Latent-class analyses also identified gender differences in autogenous versus reactive symptom profiles (Atli, 2014; Batmaz, 2016), although findings for compulsion-only subtypes were mixed (Rodgers, 2015). Taken together, these findings are consistent with the possibility that gender differences in OCD symptom organization become more differentiated across development, with adult women showing a less uniformly interconnected and potentially more heterogeneous symptom structure; however, direct longitudinal tests of this developmental pattern are lacking.

The autogenous-reactive (AO-RO) model of obsessions and compulsions (Lee & Kwon, 2003; Lee et al., 2005) distinguishes internally oriented autogenous symptoms from more externally focused reactive symptoms. Autogenous symptoms include forbidden thoughts and mental rituals, whereas reactive symptoms include many contamination and harm-related concerns. Using data-driven subtyping, Atli (2014) and Batmaz (2016) found autogenous obsessions to be more common in males and reactive obsessions to be more common in females (Atli: F:M OR = 0.24 for autogenous and 3.76 for reactive obsessions; Batmaz: M:F OR = 0.31 for autogenous vs. reactive obsessions). These findings are consistent with the symptom-specific patterns summarized above, including greater contamination symptoms in females and greater sexual/religious symptoms in males. However, Gemelli et al. (2025) found no significant sex differences in the severity of either autogenous (β = 0.03, p = 0.73) or reactive obsessions (β = −0.02, p = 0.86).

All of these results rely on participants being able to report their symptoms accurately, which may depend in part on insight into their symptoms. Insight also has important implications for treatment engagement and symptom management. Three studies during the review period examined gender differences in insight among adults with OCD using different instruments (Shimshoni, 2011; Cherian, 2014; Guillén-Font, 2021). Two reported better insight in women (Shimshoni, 2011: z = 2.43–2.65, p = 0.004–0.014; Guillén-Font, 2021: F = 1.34 vs. M = 1.95, p = 0.05), whereas Cherian (2014) found no significant gender difference (M:F OR = 0.68, p = 0.44). More research is needed to clarify gender differences in insight and their implications for studies relying on self-report. Insight should also be considered in relation to predominant symptom profiles, as autogenous obsessions, which are more common in men (Batmaz, 2016), have been linked to poorer insight (Tolin et al., 2001).

Further work is needed to characterize heterogeneity in OCD symptom presentation and organization, including the role of insight, and to determine how these features vary by gender across development. Current findings suggest that in childhood, girls with OCD may present with a broader symptom burden and greater severity in selected domains, whereas in adults gender differences may be expressed more through divergence in how symptoms cluster and relate to one another. However, only two of eight data-driven studies included pediatric or adolescent samples, and direct developmental tests are lacking. Gender comparisons may therefore be particularly useful not for defining fixed male and female OCD symptom profiles, but as a strategy for revealing heterogeneity that may otherwise be obscured within the broader OCD phenotype. Moving from comparisons of individual symptoms toward analyses of symptom structure and developmental trajectories may help identify more mechanistically informative phenotypes and, ultimately, clinically meaningful subtypes.

#### 3.2.2. Additional Burden on Patients and Families

OCD significantly impairs well-being and quality of life for patients and their families (Markarian et al., 2010). Forty-five studies during the review period (1 spanning all ages, 5 child/adolescent, and 39 adult) examined gender-related aspects of this burden; 42 included direct gender comparisons, while 3 single-gender studies examined female-specific intergenerational burden (Uguz, 2014, 2015) or stigma and help-seeking among men with OCD (Shmidova, 2026). Overall disability may be similar in men and women (Benatti, 2022), but specific domains vary by gender and age (Table 1, Table A2.2). Educational achievement was more adversely affected in girls than boys with OCD (β = −0.08 vs. −0.03; Bortes, 2022), although gender differences were not observed for non-passing grades or broader daily functioning (Usami, 2026). In adults, men were more likely to remain unmarried (59.1% vs. 46.3%; Muhlbauer, 2021) and reported greater loneliness (Friedman-Ezra, 2024) and physical aggression (Puşuroğlu, 2024), whereas women showed greater marital-role maladjustment (β = 0.38; Rosa, 2012). Social anhedonia (Lu, 2023), relationship/partner-related OCs (Mancin, 2024), and perceived effects of social media on mood (Guazzini, 2022) did not differ by gender. Several studies suggested poorer quality of life in women, including overall or vitality-related quality of life (Jahangard, 2018; Bakhshinejad et al., 2026) and emotional quality of life (Kugler, 2013), although other quality-of-life domains were null (Kugler, 2013; Zerdziński, 2022).

Sexual functioning also showed gender-related differences, although findings varied. Sexual dysfunction was more prevalent in women in some samples (F:M OR = 12.43, Ghassemzadeh, 2017; F:M OR = 2.9, Zerdziński, 2022), whereas Pozza et al. (2021) found no significant gender differences in sexual excitation or inhibition and no gender moderation of OCD-related sexual response. Two other Pozza studies (2019, 2020) reported qualitatively higher sexual inhibition in women (β = 1.12 and 1.10, respectively; p = 0.053 in both studies), and Dèttore et al. (2021) found gender moderation of associations among attachment characteristics, contamination symptoms, and sexual excitation/inhibition. Compulsive sexual behavior was more prevalent in men than women with OCD, consistent with population norms (Snaychuk, 2022).

OCD is also linked to reduced professional functioning in both men and women. In a large Swedish sample (N = 173,443), OCD-related labor-market marginalization—including prolonged sickness absence, unemployment, and disability pension—was somewhat greater in men (HR = 3.6 vs. 2.9; Pérez-Vigil, 2018). This pattern should be considered in the broader labor-market context, in which men—particularly White men—often occupy more advantaged positions, so OCD-related impairment may involve a greater loss of relative advantage (Wadsworth et al., 2020). In contrast, occupational dysfunction was substantially more common in women in a Turkish clinical sample (F:M OR = 8.11; Yalım, 2024), whereas job performance did not differ by gender among nurses with OCS (p = .23; Kavosi, 2026).

Gender-related differences also extend to caregivers. Female caregivers reported greater subjective burden (Siu, 2011; Vikas, 2011) and greater stress and anxiety (Futh, 2012). Family accommodation may also vary by patient gender. In children, accommodation increased with symptom severity in boys (β = 0.11) but not girls (β = −0.04; DuBois, 2025), whereas Skarphedinsson et al. (2025) found no overall association with child gender. Female patients were also more likely to involve others in OCD-related behaviors (F:M OR = 4.12; Yanagisawa, 2015). In adults, family accommodation was higher for women in two studies (Liao, 2021; Kelley, 2024), qualitatively higher but nonsignificant in another (d = 0.41, p = 0.051; Shrinivasa, 2020), and null in Kuru et al. (2023). Whether these differences reflect cultural or family-role expectations, symptom presentation, or other factors remains unclear.

OCD is associated with adverse physical-health outcomes. Contamination symptoms have been linked to delayed medical care, with no evidence that this association differed by gender (Karagöz, 2021). Among individuals with OCD, estimated excess natural-cause mortality was higher in men (MRR = 1.69 vs. 1.37), whereas estimated excess unnatural-cause mortality was higher in women (MRR = 3.15 vs. 2.23; Meier, 2016). Consistent with this pattern, OCD increased motor-accident risk in women (HR = 1.2–1.3) but not men (Mataix-Cols, 2022). Offspring of mothers with OCD also showed lower birth weight, shorter gestational age, and elevated inflammatory markers (Uguz, 2014, 2015), indicating potential intergenerational burden.

Individuals with OCD often face poor public understanding, limited support, and stigma (Ponzini & Steinman, 2022). Among youth with OCD, experienced stigma did not differ between girls and boys (d = 0.04; McCarty, 2026). In a qualitative study of men with OCD, masculine norms, anticipated and enacted stigma, selective disclosure, and mistrust of psychiatric care emerged as barriers to help-seeking (Shmidova, 2026). Recognition of OCD varied widely across countries and symptom profiles (∼25%–85%). Women were more likely to correctly recognize OCD in some studies (OR = 2.33; Shahwan, 2026; Chong, 2016), whereas others found no gender differences (Coles, 2013; García-Soriano, 2017; Stewart, 2019). Men more often interpreted OCD as perfectionism (OR = 1.76; Stewart, 2019), whereas women expressed greater willingness to help individuals with OCD (η² = 0.10–0.11; García-Soriano, 2017).

Overall, gender differences in OCD burden appear to be domain-specific rather than uniformly greater in one gender. Further work is needed to determine how gender shapes educational, occupational, relational, sexual, physical-health, and family burden across development, and how these differences interact with symptom presentation and social context. Greater public understanding of OCD and its diverse manifestations may also reduce stigma and improve support for individuals with OCD and their caregiver.

#### 3.2.3. Summary of Key Considerations

Methodological Concerns:

- Differences exist between studies using self-report measures and those employing clinician-administered assessments. Results based on self- and parental reports were more variable, particularly in pediatric samples, where discrepancies between informants may also affect observed gender differences. Limited evidence suggests that insight itself may vary by gender, raising the possibility that symptom reporting contributes to some of these differences. Studies directly comparing informants and assessment methods across genders are needed.
- The threshold distinguishing subclinical from clinically significant OCD symptoms may also influence observed gender patterns. Inclusion of subthreshold symptoms may help determine whether gender differences extend across the symptom continuum; this issue is discussed further under Epidemiology.
- Traditional studies generally compare predefined symptom domains, whereas data-driven approaches can examine how symptoms co-occur and organize. Only two of eight such studies included pediatric or adolescent samples, leaving developmental changes in symptom organization largely untested.
- As with symptom presentation, gender differences in OCD burden appear to be domain-specific rather than uniformly greater in one gender. Educational, occupational, relational, sexual, physical-health, and family burden may show different patterns, suggesting that overall disability scores can obscure clinically meaningful heterogeneity. Future studies should therefore assess burden across multiple functional domains and consider these patterns in relation to the broader distress profile, symptom presentation, and social context.

Unanswered Questions:

- How do gender differences in insight and symptom reporting contribute to differences observed across self-, parent-, and clinician-administered assessments?
- How does OCD symptom presentation and organization vary by gender across development, including puberty and later life?
- Do gender differences observed in clinical OCD also characterize subthreshold symptoms in the general population?
- Do gender differences in OCD burden reflect distinct profiles of distress and functional impairment across educational, occupational, relational, sexual, physical-health, and family domains, and how are these profiles related to symptom presentation and comorbidity?
- How do gender differences in symptom presentation, healthcare-seeking, and social context contribute to differences in the broader burden of OCD?

Hypotheses for Future Research:

- Gender may help identify mechanistically meaningful heterogeneity in OCD symptom organization.
- Gender-related symptom differences may change across development, from broader symptom burden in girls to greater divergence in adult symptom organization.
- Gender differences in contamination/washing and sexual/religious symptoms may become more pronounced after puberty.
- Gender differences in insight may contribute to discrepancies between self- and clinician-rated symptoms.
- Gender differences in OCD burden may be domain-specific and contribute to broader distress phenotypes.

### 3.3. Epidemiology

#### Highlights from Section 3.3 Epidemiology

<u>Incidence and Prevalence</u>: OCD is more common in women than in men in adults but not in children; early-onset OCD diagnosis is more common in men.

<u>Risk factors:</u> Men may be more susceptible to epigenetic and familial risks, and to prenatal and early life stresses. In women, OCD may more often be linked to reproductive events (puberty, menses, peripartum, and menopause) and to stressful and traumatic life events.

<u>Key concerns:</u> Transgender and nonbinary individuals remain substantially understudied in epidemiological studies. Most estimates of the prevalence of OCD in children and adolescence disproportionally rely on self- or parental reports. OCD symptoms can be difficult to distinguish from comorbid conditions (especially in children).

<u>Future studies:</u> More work is needed to describe the prevalence of OCD subtypes and the evolution of subthreshold symptoms across age groups, including in older individuals and following trauma (especially in highly traumatized populations).

#### 3.3.1. Incidence and Prevalence of OCD

Recent reviews and meta-analyses of epidemiological studies (Fawcett et al., 2020; Guo et al., 2016; Mathes et al., 2019) have reported that OCD affects 1 – 5% of the general population and is more common among females in late adolescence and adulthood, but may be more common among males in childhood (Benatti 2022). Results from 80 epidemiological studies identified during the review period (22 from children/adolescents, 51 from adults, and 7 from pooled samples; Table A3.1) are largely consistent with these generalizations. They also raise a number of methodological and conceptual concerns that may affect estimates of overall incidence/prevalence and gender^2^ ratios.

*3.3.1.1. Incidence* is defined as the number of individuals in a population who newly develop OCD over a given period of time. Ten studies (3 spanning broad age ranges, 4 focused on children/adolescents, 2 on adults in the general population, and 1 on active-duty military personnel) examined gender differences in OCD incidence across Northern European, Taiwanese, U.S., Brazilian, and multinational samples, using national or administrative health records or clinical interviews. Among studies reporting annual incidence, estimates ranged very broadly, from approximately 0.008% to 0.12% in females and from 0.012% to 0.08% in males, with some variation across age groups (Figure 3A). Importantly, studies based on health records capture OCD that presents to clinical care and is diagnosed and therefore may be influenced by differences in help-seeking behaviors and diagnostic practices across time, age groups, and geographical locations.

**Figure 3.** Female-to-male odds ratios in incidence and prevalence of OCD in pediatric and adult samples. F – female, M – male.

Several studies also indicated that diagnosed OCD incidence has changed over time. Rintala (2017) and Ask (2020) reported increasing incidence over successive birth cohorts or calendar years in Finland and Norway, respectively, which they attributed partly to increased recognition and diagnosis. Danish registry studies reported broadly consistent increases in diagnosed OCD across childhood and adolescence (Dalsgaard, 2019; Steinhausen, 2019). In contrast, Veldhuis (2011) reported decreasing OCD incidence in males but not females in the Netherlands from 1996 to 2007 (RR M = 0.56, p < 0.001; RR F ∼ 1). These differences may reflect variation in age groups, geographical location, or diagnostic practices.

Despite this heterogeneity, the age-related pattern of gender differences was relatively consistent across studies. OCD incidence was generally higher among boys in childhood and higher among females in samples extending into adolescence and adulthood (Rintala 2017; Dalsgaard 2019; Steinhausen 2019; Huang 2014). Three studies directly examined changes in gender ratios across age groups in Northern Europe (Rintala, 2017; Dalsgaard, 2019) and Taiwan (Huang, 2014). In Northern European samples, incidence was higher in boys before approximately age 10 and became higher in females starting at approximately age 14. In Taiwan, the shift from male to female predominance occurred later, around ages 25–35, and female predominance attenuated after age 64.

Among 1.36 million active-duty service members, women showed a 1.73-fold higher incidence of first-time OCD diagnosis than men (Russell, 2022), suggesting stronger female predominance in incident OCD diagnosis in the military than in the general population.

*3.3.1.2. Prevalence* is the proportion of individuals in the general population who have OCD, including both new and chronic cases, over a specified period or at a particular point in time. The prevalence of OCD/OCS in children and adolescents was evaluated by ten studies, but only two relied on assessments by trained clinicians. Nineteen studies evaluated the prevalence of OCD/OCS in adults; fifteen relied on clinical interviews, one asked participants to self-report whether they had been formally diagnosed with OCD, one relied on computerized self-reports supervised by trained research personnel, and two used self-report screening measures. Two studies evaluated five-year period prevalence across adolescent and adult age groups using retrospective medical-record data. All but one study compared the prevalence of OCD/OCS between males and females. Smoller et al. (2026) instead compared cisgender and gender-minority individuals across age groups, reporting approximately fivefold higher five-year OCD prevalence among transgender and gender-nonconforming individuals than among cisgender individuals (GNC:cis OR = 5.52, 95% CI [3.60, 8.44]; trans:cis OR = 4.94, 95% CI [3.18, 7.67]). Because only one study examined gender-minority groups, the discussion of prevalence focuses primarily on comparisons between males and females.

Self-reports and parental reports are more likely to be subject to systematic gender-related reporting biases [see Section 3.2.1; (Rosenman et al., 2011)]. The greater reliance on self- and parent-report measures in studies of children than in studies of adults may help explain the highly inconsistent pediatric findings (Figure 3B, Table A3.1). Both studies using clinician-administered interviews reported higher prevalence in girls. Mohammadi et al. (2021) reported F:M OR = 1.27 [95% CI, 1.10–1.48], whereas Vivan et al. (2014) reported the strongest female bias in adolescents (F:M OR = 3.36 for OCD and OR = 1.95 for past-month OCS). Overall, prevalence findings in children and adolescents paralleled incidence trends across age and gender groups: OCD/OCS prevalence was higher among boys in younger age groups and among girls after puberty.

In adults, most studies reported higher OCD/OCS prevalence among women, although F:M ORs varied widely, ranging from 0.6 to 5.0 (Figure 3B, Table A3). Twelve-month OCD prevalence was evaluated most commonly. Five reports assessing lifetime prevalence produced F:M OR estimates closer to 1, possibly reflecting the contribution of childhood OCD histories (Subramaniam, 2012, 2019, 2020; Carta, 2020; Liu, 2025). One study using population-weighted EHR records estimated similar recorded OCD prevalence in females and males (M = F = 0.4%, substantially lower than typically reported in the literature; Mortier, 2026). One study assessed obsessions and compulsions separately and found a stronger female bias in the prevalence of compulsions than obsessions (F:M OR = 1.32 and 1.18, respectively; Lee, 2021). Conversely, several studies found OCS, subthreshold OCD, or likely OCD to be more prevalent among men (F:M OR = 0.6–0.9; Fineberg, 2013; Subramaniam, 2020; Chen, 2025). The DSM uses relatively subjective and environmentally sensitive criteria to distinguish subthreshold symptoms from clinical diagnoses. Comparing gender differences below and above DSM-based diagnostic thresholds may therefore provide additional insight.

Only two studies examined geriatric samples, providing limited evidence of female predominance (F:M OR = 1.0–3.5; Klenfeldt, 2014; Canuto, 2017). More research is needed in this age group.

Several studies also examined OCD/OCS prevalence in selected populations that may not be representative of the general population but may provide insight into environmental and contextual influences on OCD/OCS and associated gender differences. Twelve studies examined male–female differences in self-reported OCD/OCS among college and university students. Seven found no significant difference in overall prevalence, four reported higher prevalence in women, and one reported higher prevalence in men. Female predominance appeared primarily in medical-student samples using the OCI-R, a preliminary pattern requiring further investigation. Some studies also found symptom-specific patterns, including greater subthreshold OCD in men (Jaisoorya, 2017) and dimension-specific differences (Ashraf, 2017). In homeless women from Canada, OCD prevalence was substantially higher than that reported in general-population samples: current OCD was present in 19% and lifetime OCD in 62% of participants (Strehlau, 2012). This may reflect impaired social functioning associated with OCD (Section 3.2.2), effects of the multiple stresses associated with homelessness on risk for OCD (Section 3.3.2), or both.

*Sex differences in help-seeking* may also influence prevalence estimates based on clinical populations. Among adults seeking help in primary-care settings, women tended to predominate (F:M OR = 0.98–1.96; Table A3.1). These individuals often reported anxiety or depression as their primary concern before receiving an OCD diagnosis. In a psychiatric emergency department, gender differences varied across the year: female predominance was significant only from July through September (F:M OR = 2.56), whereas differences during other periods were not statistically significant (Afifi, 2026).

Among individuals receiving care from OCD-specialty services, a tentative geographic pattern emerged. Female representation was higher in studies conducted in the United States/Brazil or multinationally (F:M OR = 1.1–1.7; Medeiros, 2017; Dell’Osso, 2017). A recent Mexican specialty-clinic sample showed approximately equal representation of women and men (48.9% female; Brueggerhoff-Batel, 2025). Male representation was higher in Indian studies of adults, children, and mixed-age samples (F:M OR = 0.4–0.7; Cherian, 2014; Algin, 2018; Deepthi, 2018; Tripathi, 2018; Sharma, 2019). Sharma et al. (2019) suggested that, in the developing world, males may receive preferential care and more prompt clinical attention than females. However, only two studies examined pediatric help-seeking samples, and additional studies are needed to clarify this pattern.

Overall, prevalence findings broadly parallel the age-dependent pattern observed for incidence: OCD diagnoses may be more common among boys in childhood but among girls and women from adolescence onward. OCS appear more evenly distributed between males and females, with some studies suggesting that subthreshold OCS may be more prevalent among males. These patterns should be interpreted cautiously given differences in assessment methods and potential cultural and help-seeking biases.

#### 3.3.2. Onset, progression, and contributing factors

##### 3.3.2.1. Onset of OCD symptoms and diagnosis

There is no consensus as to how to define the onset of OCD (e.g. as emergence of distressing symptoms or when the diagnostic criteria were first met) or how best to assess it (using retrospective self-reports, parental reports, or contemporary medical records). Each of these approaches is associated with its own limitations. Twenty-four reports from the review period (6 in children/adolescents, 17 in adults, and 1 spanning adolescents and adults) examined gender differences in age-of-onset patterns, and six examined progression from symptom onset to diagnosis or subsequent diagnostic progression (Table A3.2).

There is a general consensus that the onset of OCD occurs earlier in males than in females (Mathes et al., 2019). Many researchers argue that early-onset OCD (EO-OCD) may form a distinct subgroup, characterized by unique clinical and familial attributes, higher male prevalence, high tic [∼ 30 – 40%, (Ferrao et al., 2013; Franklin et al., 2012)] and ADHD comorbidity [∼ 25 –30%, (Geller et al., 1996; Masi et al., 2010)], and elevated familial risk (Eichstedt & Arnold, 2001; Geller et al., 1998; Geller et al., 2003). In contrast, adult-onset OCD is more prevalent in women and more frequently co-occurs with depression [≥ 25% (Sharma et al., 2021)]. This pattern was generally supported by studies from the review period. However, Stein et al. (2025) found no evidence of sex differences in retrospectively reported age of onset in a multinational adult sample. The proportion of women in adult samples tends to be higher than in pediatric samples, and early-onset OCD is more common in males (Figure 3B, Table A3.2). Narayanaswamy et al. (2012) relied on a combination of parental and patient reports regarding the onset of distressing symptoms and found a significantly earlier onset in males (Cohen’s d = 0.62, p < 0.001). Torresan et al. (2013) also found that onset of “worst ever” symptoms and associated help-seeking also occur earlier in adult men (∼ 27 – 28 years of age) than in women (∼ 30 – 31 years of age).

Data-driven derivation of age-of-onset subgroups produced different outcomes for distressing symptoms and DSM diagnosis of OCD (Albert, 2015; Anholt, 2014; Nakatani, 2011). These studies suggest that distressing symptoms emerge in three distinct phases: early childhood (prior to ages 9 – 12), adolescence/early adulthood (from ages 10 – 12 to 18 – 23), and during adulthood (after age 24). Symptoms reaching a severity level that satisfies DSM criteria tend to manifest in two waves: before age 20 and after ages 20 – 26. This suggests that some children with early-onset symptoms may experience subthreshold obsessive-compulsive patterns for an extended period before progressing to clinically significant levels justifying a DSM diagnosis. The onset of distressing symptoms is more common in boys in both childhood and adolescence. This could imply that boys may endure subthreshold symptoms longer than girls before manifesting clinically significant OCD. However, two studies (Coles, 2011; Thompson, 2020) relying on retrospective self-reports found that adult women recalled a longer symptom phase— the period from onset of distressing symptoms to meeting diagnostic criteria—than men (approximately 6–8 vs. 4–5 years), whereas one study found no gender difference in time from symptom onset to diagnosis (Ziegler, 2021).

This contradiction could be a consequence of recall bias. For instance, women may have better insight into their past symptoms (Section 3.2.1); alternatively, men with very early onset of OCs may have been too young at the time to remember or to be able to identify their behaviors as symptomatic, or men may have more autogenous symptoms that are more difficult to identify retrospectively. However, another explanation is possible. Castagnini et al. (2016) found that both an adolescent diagnosis of OCD and an adolescent diagnosis of an anxiety disorder (i.e., phobia, panic disorder, generalized anxiety disorder) predicted the need for treatment in adulthood, similarly in men and women. Anxiety symptoms and diagnoses become more common in girls than in boys beginning from middle childhood (Altemus et al., 2014; Lewinsohn et al., 1998). Thus, it is possible that adult OCD emerges from other forms of anxiety more often in females, explaining their longer recalled duration of the pre-diagnostic symptom phase. Barzilay et al. (2019a) found that subthreshold OCs are more prevalent in girls (OR = 1.3), a pattern that becomes significant following puberty (female gender x puberty: p = 0.038). This could be due to emergence of subthreshold OCD symptoms from anxiety in prepubescent girls (Castagnini, 2016) or due to hormonal changes related to reproductive cycle events (see below). Interestingly, Kong (2026) reported that OC symptoms in youth may evolve into schizophrenia-spectrum disorder within a three-year follow-up, with this progression observed more frequently in males than females (50% vs. 11.8%), although the small number of baseline OCD-spectrum cases (N = 27) limited statistical power (F:M OR = 0.15 [95% CI, 0.01–1.23], p = .065). This further emphasizes that distressing symptoms in adolescents might be either more difficult to diagnose using current standards, or more dynamic than in adults.

Including older individuals may provide additional insights. Two studies (Frydman, 2014; Sharma, 2015) found a 3:1 female predominance among individuals who developed distressing obsessions and compulsions after age 40. Interestingly, when PTSD diagnosis and history of pregnancy were included in the model, these variables (rather than gender) become significant predictors of late-onset OCD. This suggests that later-life onset of OCD in women is closely linked to life stressors.

Different types of OCD symptoms may emerge at different ages in males and females. Kichuk et al. (2013) found that female gender was associated with later onset of forbidden thoughts but with earlier onset of symmetry symptoms. Grover et al. (2018) found a male bias in adolescent onset of hoarding and symmetry symptoms (F:M OR = 0.26, p < 0.01), and (at a trend level) of aggressive thoughts and of sexual and religious obsessions (F:M OR = 0.67); this male bias became more pronounced in adult onset of these symptoms (F:M OR = 0.19 and F:M OR = 0.4, respectively). In contrast, onset of contamination symptoms before age 18 was significantly more likely in girls (F:M OR = 16.7); this female bias extended to adulthood, though it became less pronounced (F:M OR = 4.2). In contrast, other studies found that contamination symptoms were similarly reported by both boys and girls in child samples (Cervin, 2020; Schultz, 2018). No identified study used symptom subtypes in data-driven identification of symptom onset subgroups; such an approach might help to clarify this discrepancy.

##### 3.3.2.2. Biosocial Vulnerabilities

Biological and social factors contribute to the emergence and persistence of obsessive-compulsive symptoms. Whether their roles differ by sex or gender, and whether sex and gender themselves function as risk or protective factors, remains underexplored. Seventy-one papers from the review period offer relevant data (12 in mixed-age samples, 7 in children/adolescents, and 52 in adults; Table A3.2).

*3.3.2.2.1. Prenatal and perinatal factors* may increase vulnerability to OCD, and their effects may differ by sex, although specific patterns require replication. Fevang et al. (2016) found similarly increased odds of clinically elevated OCS among boys (OR=2.5) and girls (OR=2.7) born extremely prematurely, which is more common in boys (Peelen et al., 2016), or with extremely low birth weight, which is more common in girls (Beijers et al., 2022; Meadows et al., 2005). In another large Danish cohort, premature birth and low birth weight were associated with OCD in females (HR=1.46 and 1.32, respectively), but not significantly in males, although direct sex differences were not tested (Larsen et al., 2021). Another study found that birth from August to November was associated with OCD in males, but not females (Cheng, 2014). Prenatal exposure to temperature, sunlight, and viral infections may contribute to OCD risk (McTigue & O’Callaghan, 2000; Vinkhuyzen et al., 2016). Collectively, the findings suggest that prenatal and perinatal vulnerabilities may differ by sex and exposure.

*3.3.2.2.2. Family and social context.* OCD symptoms have a strong familial component, including genetic vulnerability (Section 3.5) and shared environmental factors. In female university students, fathers’ hygiene-related disgust and OCS predicted daughters’ contamination symptoms, whereas mothers’ ordering symptoms predicted daughters’ ordering symptoms (β = 0.23 and 0.21; Taberner et al., 2012). In another university sample, gender predicted responsibility attitudes (β = 0.16–0.19, p < 0.01; direction not reported), and perceived parental overprotection predicted stronger responsibility attitudes—a proposed cognitive risk factor for OCD (Hacıömeroğlu, 2008) and responsibility attitudes partially mediated the association between perceived maternal overprotection and OCS (Sobel z=2.61, p < 0.01). These relationships were adjusted for gender but were not examined separately in women and men (Hacıömeroğlu, 2014).

Family and social resources may also be protective. Girls reported greater reliance on family and social resources, whereas boys relied more on personal competence (Hjemdal, 2011). Family cohesion and adaptability were more strongly negatively associated with OCS in women than in men (cohesion: F r = −0.25, M r = −0.16, p = 0.037; adaptability: F r = −0.28, M r = −0.19, p = 0.034). Family adaptability buffered the association between anger suppression and OCS in women, whereas family cohesion buffered the association between anger proneness and OCS in men (Liu, 2017). Finally, child OCD status interacted with supportive parent–child behavior in predicting paternal oxytocin (β = 0.11, p = 0.008), but the effect did not survive FDR correction (pFDR = 0.18); the maternal effect was nonsignificant (β = −0.06, p = 0.23, pFDR = 0.77; Mora-Jensen, 2025).

*3.3.2.2.3. Early life stress.* In individuals with increased familial risk for OCD and other psychiatric disorders, greater adverse childhood experience (ACE) severity was associated with earlier onset of psychiatric illness in males, but not females (M HR=1.13, p<0.001; F HR=0.94, p=0.19; Someshwar, 2020). However, other comorbid symptoms, such as ADHD, which are more common in males, were not included as covariates and could have contributed to this association (Section 3.4.3.5).

*3.3.2.2.4. Stressful life events (SLEs)* can accelerate the progression from subclinical OCS to clinical OCD and may differentially impact specific symptom dimensions. Female patients reported SLEs preceding OCD onset more often than male patients (unadjusted F ORs = 1.50 [95% CI, 1.01–2.30] and 2.20 [95% CI, 1.40–3.40]; Real, 2011; Rosso, 2012), although the association in Real (2011) was attenuated after adjustment (OR = 1.20 [95% CI, 0.74–2.07]). It is possible that female gender-associated factors (e.g., later age of onset, higher predominance of contamination symptoms, greater exposure to stressful events), and not female gender itself, modulates the relationship between stressful events and OCD.

SLEs may impact males and females differently. Current stress was more strongly associated with OCS in men (F OR = 0.33 [95% CI, 0.18–0.63]; OCD sample: OR = 0.39 [0.21– 0.75]; Ashraf et al., 2017). Although death-, relationship-, and illness-related SLEs were similarly prevalent across sexes (ds = 0.001–0.054), relationship termination accelerated progression from OCS to OCD in both sexes, whereas death of a significant other did so only in men (β = −0.31, p < 0.001; Destrée, 2020). Relationship stress was associated with sexual/religious OCS in men (β = 0.90, p = 0.03), but the sex interaction was nonsignificant (β = −0.49, p = 0.069); death of a loved one was associated with hoarding symptoms in both sexes. Women reported higher OCS in an occupational-stress sample (d = 0.24; Forresi, 2022). Bullying predicted OCD similarly in females and males (OR = 1.31 versus 1.32), although associations with later OCS were numerically stronger in females and direct sex differences were not tested (Pol-Fuster, 2025). Minority stress may represent another relevant social stressor. Among sexual- and gender-minority adults with OCD, rejection sensitivity was associated with greater OCS in sexual minorities (β = 0.13, p = 0.02), while gender-minority stress showed symptom-specific associations with responsibility/harm, unacceptable thoughts, and symmetry symptoms (Pinciotti, 2026). These patterns require careful replication.

More extreme forms of life stress, traumatic stressful events (TSEs), involve experiencing or witnessing events that pose an immediate threat of serious harm. TSEs have been linked to higher OCS rates (Barzilay, 2019b). Women with childhood trauma had higher OCS than men (OR = 1.47; d = 0.27), but the study did not establish that childhood trauma was a stronger predictor in women (Jin, 2024). Childhood-trauma scores were higher in participants with OCD than controls in both sexes. OCD–control effect sizes were consistently larger in women than men (total trauma: d = 0.92 versus 0.50), but direct sex differences were not tested (Lochner, 2026). Among trauma-exposed youth, OCD was more prevalent in females than males (OR = 3.04 [95% CI, 2.17–4.28]) and in gender-minority than non-gender-minority youth (OR = 2.78 [1.74–4.44]; Pinciotti, 2025). However, the network linking emotional abuse and physical neglect with emotional distress and OCS did not differ by sex (Huang, 2025). Impacts of trauma on OCs may also be modulated by trauma type. Women may be less often exposed to non-sexual TSEs but more susceptible to their negative consequences; rates of OCS increased more with each additional TSE in women than in men. Rates of OCS were similarly associated in men and women after physical assaults, but women showed higher OCS rates in association with non-assaultive TSEs. Among individuals with traumatic experiences, the presence of OC symptoms was also associated with psychotic symptoms; however, it remains unclear whether comorbidities (Section 3.4) contribute to the observed gender differences, as gender differences in psychotic symptom prevalence were not examined in Barzilay (2019b).

Males and females were similarly susceptible to the development of OCS after sexual TSEs (Barzilay, 2019b), but females are more often exposed to such events (Snyder, 2000; Vidal-Ribas et al., 2015). Young women aged 16 – 19 are 4 times more likely than the general population to be victims of rape, attempted rape, or sexual assault (Spence, 1997). This may contribute to the higher rates of OCD in general, and contamination OCs in particular, in women (Section 3.2.1). If this pattern is further confirmed, then it might be advisable to offer preventive OCD treatment to victims of trauma, especially sexual trauma.

Six studies specifically linked a history of unwanted sexual experiences to contamination symptoms (Peles, 2014; Waller, 2014; Ishikawa, 2015; Kennedy, 2017; Krause, 2021; Tang, 2022). Female undergraduate students’ recall of unwanted sexual experiences triggered a stronger sense of being contaminated (Cohen’s d = 2.02, p < 0.001) and an increased urge to wash (Cohen’s d = 0.44 p < 0.03) in women who had experienced rape/attempted rape as compared with those who experienced verbal or visual sexual assault (Ishikawa, 2015). In both female (Waller, 2014; Tang, 2022) and male (Kennedy, 2017) students, imagining themselves as initiators of a non-consensual kiss led to mental contamination symptoms and to cleansing behaviors (Cohen’s ds > 0.53 p < 0.001). Similarly, mental contamination symptoms in female students were triggered by imagining being a perpetrator in a sexual-harassment scenario (Cohen’s d = 1.6–1.9 p < 0.001; Krause, 2021).

In sexually abused women (Peles, 2014), rates of OCD were strongly associated with a history of opioid addiction (OR = 4.53, p < 0.01); the most common symptoms were contamination obsessions and cleaning compulsions. This relationship may be OCD-specific, as rates of PTSD and dissociation were higher in sexually abused women with no history of opioid addiction. Among women with a history of opioid addiction, other comorbidities and adverse life events were significantly elevated among those with OCD vs. those without; it is possible that self-medicating with opioids helped with PTSD and dissociative symptoms but exacerbated OCD.

Elevated OCD rates have also been linked to stress associated with injury or impairing illness. For example, male veterans with traumatic foot injuries show a markedly elevated prevalence of OCD (14.4%; Taghva, 2017). Similarly, Mazza et al. (2020) found elevated psychiatric symptoms, including OCS, in COVID-19 survivors. OCS were more severe in women (d ≈ 0.43, p = 0.001) and in individuals with a pre-existing psychiatric history (gender × psychiatric-history interactions were not tested). Baseline inflammatory markers were higher in men (ps ≤ 0.007); OCI scores were nominally positively associated with MLR (r = 0.115, p = 0.042), with similar trend-level associations for NLR and SII, but these associations did not survive correction for multiple comparisons. Inflammatory-marker × gender interactions on OCS severity were not tested.

The COVID-19 pandemic prompted extensive investigation of obsessive–compulsive symptoms (OCS), with sex differences varying by study design and population. Cross-sectional studies, largely in general and student samples, often reported higher prevalence or severity of OCS in women, broadly consistent with non-pandemic patterns, with effects ranging from small to large (e.g., Cohen’s d ≈ 0.24–1.76; McKune, 2021; Ahmed, 2021ab; Othman, 2021; Ghaderi, 2025). In contrast, some cross-sectional studies reported higher OCS burden in men— particularly in general population samples from Saudi Arabia, Korea, and student samples from China—though effects were typically small to moderate (Cohen’s d ≈ 0.20–0.40; AlHusseini, 2021; Seo, 2025), and many studies showed null or domain-specific differences (Table A3.2). Methodological heterogeneity likely contributes to variability across findings: only Othman (2021) explicitly controlled for severity of depression, few studies adjusted for baseline symptom burden, and none systematically accounted for other comorbid psychiatric symptoms (e.g., anxiety) or resilience factors (e.g., family support).

Longitudinal studies provide a more consistent picture. Across nine studies spanning student, general population, and clinical OCD samples, overall pandemic-related changes in obsessive–compulsive symptoms (OCS) were broadly similar in men and women, with most sex × time effects small or absent (η² ≤ 0.023; Meda, 2021; Meza, 2024; Silverman, 2024; Schuurmans, 2026). Notable exceptions were context- and phase-specific: Chinese student samples showed stronger OCS increases in men across pandemic waves (β ≈ 0.38–0.67; Ji, 2020); one general population study reported greater increases in contamination-related fears in men (OR ≈ 1.7; Samuels, 2021); and in clinical OCD samples, women exhibited slightly greater symptom worsening during the pandemic (Cohen’s d ≈ 0.34–0.41; Højgaard, 2021; Bostan, 2025), accompanied by a sharper post-pandemic symptom decline in men (Cohen’s d ≈ 0.34; Jelinek, 2021). Overall, longitudinal evidence suggests broadly parallel OCS trajectories following stress across sexes, with limited support for generalized sex-specific effects.

#### 3.3.3. Course of OCD

Eight papers from the review period compared longitudinal trends in OCD symptoms in males and females. Two adult studies found no gender differences in remission rates (Marcks, 2011; Fineberg, 2013). Some longitudinal studies identified sex-differentiated symptom trajectories: in adolescents, boys more often exhibited high but remitting symptoms, whereas girls showed greater increases within a moderate but escalating trajectory (Jagannathan, 2024); in adults, men more frequently displayed episodic courses, whereas women were more likely to show chronic courses (Geiger, 2024). Other studies found no significant gender differences in chronic versus remitting course in adults (Visser, 2014; Zhang, 2025) or children (Mancebo, 2014). Goldberg et al. (2015), however, contrasted the course of OCD preceded by stressful life events (SLE-OCD) with OCD unrelated to an identified stressor (non-SLE-OCD). A chronic course of SLE-OCD was negatively associated with familiality in both men and women, whereas a chronic course of non-SLE-OCD was more strongly negatively associated with familiality in men. These findings suggest that gender differences in OCD course may be more evident for non-stress-related OCD, whereas stress-related OCD may follow a more similar course in men and women.

*3.3.4. Reproductive system events and conditions* and their role in OCD were examined during the review period by seventy-three studies (Table A3.3). In a subset of women, OCD symptoms had their onset in or were exacerbated during pregnancy and postpartum (Goodman, 2014). Guglielmi et al. (2014) extended these findings to a wider range of reproductive cycle events. In a study involving 542 women from the U.S. and the Netherlands, 26.5% reported the onset of obsessive-compulsive symptoms during different reproductive phases: menarche (13%), pregnancy (5.1%), postpartum (4.7%), or menopause (3.7%). These findings align with data showing that the impact of puberty on OCD symptoms is more pronounced in girls than in boys (Barzilay, 2019). Guglielmi et al. (2014) also noted that around one-third of women with OCD experience symptom exacerbation during the premenstrual period, pregnancy, postpartum, and/or menopause. This result is largely aligned with prior reports that severity of OCs elevates during premenstrual period in approximately half of cycling women with OCD (Vulink et al., 2006). Moreira (2012) found that women who experience heightened OCD symptoms with their menstrual cycle tended to exhibit more severe autogenous symptoms, such as aggressive, sexual, or religious obsessions. They also tended to experience adult-onset OCD and higher levels of anxiety, depression, and thoughts of suicide. These findings suggest that female reproductive hormones play a significant role in the emergence and exacerbation of OCD symptoms.

Mulligan et al. (2019) discovered a negative correlation between checking symptoms and an EEG measure of the brain’s capacity to rapidly detect and evaluate errors (error-related negativity) during the luteal phase of the menstrual cycle, when progesterone levels peak, but not during the follicular phase, when estradiol levels peak. This suggests that menstrual-cycle phase may modulate the brain correlates of checking behaviors in women. In contrast, women with polycystic ovary syndrome, which is characterized by dysregulation of estrogen and deficiency in progesterone, reported higher levels of obsessive-compulsive symptoms (Cohen’s d = 0.10, p = 0.04; Li, 2017) and were more frequently diagnosed with OCD (OR = 1.8, p < 0.01; Tay, 2020). Women who relied on assisted reproductive technology to conceive were also at a higher risk for OCD (OR = 1.8; Song, 2024). More focused research and a deeper understanding of the role of sex hormones in the onset and persistence of obsessive-compulsive symptoms are needed (see also Section 3.6).

*3.3.4.1. Pregnancy and the postpartum period* are periods of major hormonal shifts and of increased emotional and physical stress. OCS during this period reduce the quality of parenting experience and of interactions with the baby (d = 0.59–1.47; Challacombe, 2016). Prior systematic reviews (Appendix A1) have estimated that OCD has a prevalence of 0–5.2% in the peripartum period, and that approximately 15% of women with OCD report onset of their first clinically significant symptoms during the peripartum period. Since 2010, 67 studies have further confirmed that the peripartum period is a time of significant risk for the development or exacerbation of obsessive-compulsive symptoms (see Table A3.3). A prospective longitudinal study (Samuels, 2026) reported an 8.4% incidence of OCD between mid-pregnancy and six months postpartum [95% CI, 5–14%], with clinically meaningful increases in OCS occurring more frequently postpartum than antepartum. Cross-sectional studies also indicate higher prevalence during postpartum (median antepartum prevalence of 2.9% [range = 2–15%] with postpartum prevalence of 5.5% [range = 3.6–15%]). Sociocultural differences may contribute to the substantial variability in prevalence estimates: from 2.0% (UK: Howard, 2018) and 2.9% (Canada: Fairbrother, 2016) to 4.9% (Turkey: Uguz, 2019) and 6.1% (Malta: Buhagiar, 2024), and all the way up to 15% (Croatia: Nakić Radoš, 2025). For instance, differences in likelihood of discontinuing pharmacotherapy for OCD during pregnancy across cultures could contribute to this heterogeneity.

Interestingly, fathers also reported elevated OCS during the peripartum period (Coelho, 2014). In contrast to women, men demonstrated higher prevalence of OCS antenatally (prevalence = 3.4%) than postpartum (prevalence = 1.8%). OCD symptoms in fathers tend to emerge when mothers experience OCD symptoms (‘sympathy OCD’). Antenatal OCD in men tended to be comorbid with bipolar symptoms; postpartum OCD tended to be comorbid with both bipolar and unipolar depression. Much more research is needed to verify and clarify these patterns in men, which were described in only one study.

Shokuhi et al. (2020) developed a scale that assesses postpartum distress; scores loaded onto two distinct factors, with obsessive-compulsive symptoms separating from general distress, indicating that peripartum OCS are distinct from the general distress commonly experienced by new parents (AUC = 0.97). Fairbrother et al. (2023) demonstrated that a broadly used self-report-based instrument, the Dimensional Obsessive-Compulsive Scale, DOCS (Abramowitz et al., 2010), can serve as a highly accurate screener (AUC = 0.9), especially if administered during late antepartum or between 4 and 6 months postpartum. More recent studies further supported the validity of brief and disorder-specific assessments of peripartum OCS, including the OCI-4 (AUC = 0.75 antepartum and 0.91 at six months postpartum; Abramowitz, 2025b), the French version of the Perinatal Obsessive-Compulsive Scale (AUC = 0.90; Lord, 2026), and the Y-BOCS during pregnancy (d = 1.90 for women with versus without OCD; Rast, 2025). Peripartum OC symptoms predominantly manifest as baby-related intrusive thoughts and compulsions (Barret, 2015; Fairbrother, 2023); however, increased contamination, aggressive, and somatic obsessions and checking, cleaning, and repeating compulsions are also common (Miller, 2015). Peripartum OCS often coincide with other neuropsychiatric symptoms, such as depression (Malamela, 2019; Abramowitz, 2010; Miller, 2015; Altemus, 2012; Fairbrother, 2018; Howard, 2018), generalized anxiety (Malamela, 2019; Miller, 2015; Abramowitz, 2010), panic disorder, and social anxiety (House, 2013).

Integration of findings across studies with different methodologies suggests that OCS may particularly spike immediately following the birth (day 1), subside somewhat during the following weeks (Yakut, 2019), and follow heterogeneous trajectories thereafter. Some studies suggest a gradual increase by six months followed by relative stability (Miller, 2013, 2015; House, 2013; Abramowitz, 2010; Fairbrother, 2018), whereas others found little overall change (Abramowitz, 2025a) or a modest decrease from pregnancy to postpartum (Rector, 2026). One study found that symptom presentation may evolve over the postpartum period, with hoarding symptoms evolving into washing compulsions, and neutralizing symptoms evolving into avoidance (Miller, 2024). Coping skills, such as humor, were linked to a larger decrease of OCS from day 1 to week two (Yakut, 2019). The increase of OCS from week two to month six was more pronounced in mothers without comorbid depression (this progression could potentially be linked to accumulated sleep deprivation); mothers with comorbid depression had more severe OCS that largely remained constant between week two and month six.

Risk factors for the development of OCD during the peripartum period include higher maternal age (House, 2013; Malamela, 2019; Miller, 2013), prior pregnancy-related complications or losses (House, 2013; Malamela, 2019), long gestation (Malamela, 2019), cesarean vs. vaginal delivery (House, 2013), preexisting anxiety, depression, ADHD, obsessive-compulsive beliefs, and history of sexual trauma (Miller, 2015; House, 2016; Fairbrother, 2018; Fairbrother, 2023; Babinski, 2025; Hamlett, 2025; Rector, 2026; Samuels, 2026), ongoing family conflict (Xie, 2021b), and a history of substance use or dependence (Prevett, 2017; Osnes, 2020). The association between antenatal obsessive beliefs and postpartum changes in OCS may also be moderated by genetic vulnerability (Rector, 2026). Factors that are likely to be associated with elevated perinatal stress, such as being unmarried, having fewer previous children (Miller, 2015), and insomnia and poor sleep quality (Fairbrother, 2018; Osnes, 2020) positively correlate with the severity of postpartum OCD. More recently, lower socioeconomic resources and unplanned pregnancy were also associated with increased risk of incident peripartum OCD (ORs = 4.14– 5.92; Samuels, 2026). Despite evidence that the COVID-19 pandemic triggered significant stress in the general population, an online survey study of pregnant women (N = 3,346) prior to and during the pandemic did not show any significant worsening of pregnancy-related OCD symptoms (Xie, 2020).

Reports of the effects of pregnancy on women with preexisting OCD are highly heterogeneous. In a small study by Yakut et al. (2019, N = 37), 8.1% of OCD patients reported worsening symptoms and 51% reported decreased OCD severity during pregnancy. In another small study by Uguz et al. (2011, N = 52), 32.7% of OCD patients reported worsening symptoms, 53.8% reported unchanged symptoms, and 13.5% reported decreased OCD symptoms. This heterogeneity may be linked not only to patients’ resilience skills (Yakut, 2019; see above) but also to symptom profiles and sociocultural environment. Additionally, it is possible that the decrease in OCD severity experienced by some patient subgroups relates to hormonal changes during pregnancy (see Section 3.6.3). Patients with contamination and symmetry/ordering symptoms were more likely, and patients with religious obsessions were less likely, to report worsening of OCs during the peripartum period (Uguz, 2011). Guglielmi et al. (2014) contrasted women with OCD from the U.S. (N = 352) and from the Netherlands (N = 190). During pregnancy, OCD symptoms worsened in 35.3% of individuals with OCD in the U.S. and 26.5% in the Netherlands, compared to OCD symptom improvement in 8.2% in the U.S. and 26.5% in the Netherlands. Postpartum, OCD symptoms worsened in comparable proportions of individuals with OCD in the U.S. and Netherlands (45.7% and 49.2%, respectively), with exacerbations most commonly occurring after a first pregnancy. More recent longitudinal findings further support this heterogeneity. Among women with preexisting OCD, 70.7% followed a stable-low OCS trajectory from pregnancy through 18 months postpartum, whereas 29.3% showed elevated symptoms during pregnancy followed by postpartum recovery; contamination symptoms showed additional heterogeneity, including a stable-high trajectory in 23.2% of women (Levinson, 2025).

Preexisting OCD has been associated with increased risk for several adverse perinatal outcomes, including preterm birth and low birth weight (de la Cruz, 2023), as well as neuropsychiatric complications during the peripartum period, including postpartum depression (OR = 1.36; Johansen, 2020) and worsening anxiety symptoms (Furtado, 2019). However, these associations may not extend to all medical outcomes; OCD was not independently associated with cardiovascular events during delivery after accounting for other psychiatric diagnoses (Parekh, 2025). Such risks may partly reflect transdiagnostic effects of psychiatric illness more broadly. Kimmel et al. (2021) reported reduced high-frequency heart rate variability (HF-HRV) in pregnant women with a history of OCD, suggesting diminished parasympathetic cardiac control during pregnancy and potential vulnerability to dysregulated stress physiology. Consistent with this pattern, postpartum OCD symptoms have been associated with elevated cortisol levels and higher self-reported stress (Lord, 2010), and mothers with a history of OCD have been found to be at increased risk for negative affect toward their infants (Fairbrother, 2022; Gibbs, 2023). Higher levels of postpartum baby-care OC behaviors have also been associated with lower maternal satisfaction (Öztürkler, 2025).

The peripartum period presents both challenges and opportunities for treatment (Table A7). Challacombe et al. (2017) found intensive home-based CBT to be more effective than clinic-based treatments (d = 1.9). The burden of postpartum OCD may be reduced or even prevented by providing psychoeducation to expecting parents (Timpano, 2011; Steinman, 2025). This represents an important opportunity for prevention.

Although research on peripartum OCD has grown substantially in recent years, this increased knowledge has not yet fully translated into clinical practice. OCD remains largely absent from commonly used obstetric educational resources (Lennon, 2025).

#### 3.3.5. Summary of Key Considerations

Methodological concerns:

- Prevalence estimates of OCD in children and adolescents disproportionately rely on self- or parental reports (six out of seven studies), while adult studies rarely do so (only one out of 12). This complicates comparison of prevalence rates across age groups and genders.
- Diagnosing OCD in young children is challenging, as ritualistic behaviors can be part of normal development, and OCD symptoms can be mistaken for typical developmental phases (Nakatani et al., 2011). Symptoms can also overlap with other conditions that are more prevalent in boys (Coskun et al., 2012). The reliance on self- and parental reports may introduce gendered biases in estimates of incidence, prevalence, progression, and risk factors for pediatric OCD (see also Section 3.2.1)
- Using retrospective reports to determine age of onset can introduce recall bias, which may vary by gender. Other approaches, such as event history calendars, have been effective in improving data accuracy and completeness in large-scale surveys (Drasch & Matthes, 2013).
- OCD frequently co-occurs with various other conditions that can affect its presentation and progression. There are significant inconsistencies across studies in accounting for these comorbidities, likely contributing to heterogeneity in findings. More systematic characterization of comorbidities and their impact is recommended (see also Section 3.4).
- Studies of peripartum OCD vary substantially in the instruments used and timing of assessment. Screening accuracy may also vary across pregnancy and postpartum, potentially contributing to variability in estimates of prevalence and symptom progression.

Unanswered Questions:

- Can small changes in the DSM-based cutoff levels for clinical significance alter our understanding of gendered patterns in incidence, prevalence, and age of onset?
- Do incidence or prevalence of different symptom subtypes vary by gender?
- How common is OCD in older populations? Are there any unique characteristics in OCD in this age group?
- Are there any early predictive biomarkers for OCD (e.g., elevated blood pressure, resting heart rate at earlier ages), and do they differ across genders or sexes?
- What drives the differences between men and women in impacts of traumatic experiences on OCD – variations in types of trauma, subjective experiences, or biological responses?
- How do minority stress, discrimination, and identity-related trauma influence OCD risk and symptom expression in sexual- and gender-minority populations?
- How do peripartum experiences, social and biological, impact OCD in men?

Hypotheses for future research:

- Cultural and regional factors may influence diagnostic patterns; for instance, men are more likely to be diagnosed with OCD in Eastern cultures than in Western cultures.
- Antecedents of clinically significant symptoms may differ between genders: in boys, clinical OCS are more likely to emerge from subthreshold OCS; in girls, clinical OCS are more likely to evolve from non-specific anxiety symptoms.
- During puberty, subthreshold OCS either progress to clinical OCS or resolve; progression is more likely in girls. This period may represent an opportunity for intervention.
- Contamination OCS are linked to unwanted sexual experiences in both men and women; the higher prevalence of contamination OCS in women may be due to the substantially elevated rates of adverse sexual experiences in women, especially at ages 16 – 19. An elevated prevalence of contamination OCD in other groups with elevated rates of adverse sexual experiences is predicted.

### 3.4. Comorbid and Co-occurring Conditions

#### Highlights from Section 3.3 Comorbid and co-occurring conditions

<u>Overall patterns</u>: Individuals with OCD are more predisposed to other psychiatric conditions than the general population, with largely similar gender ratios. OCD disproportionately increases risk for some substance-use and impulse-control problems in women and for social anxiety disorder in men. Gender ratios in “pure OCD” remain inconsistent.

<u>Patterns by condition:</u> Suicidality, eating disorders, and compulsive buying generally show a female bias in OCD; schizophrenia, psychosis, externalizing disorders, and alcohol use show a male bias. Bipolar disorder, hoarding, and personality disorders are largely gender-balanced, though findings for bipolar disorder and hoarding are inconsistent. Compared with the general population, female biases in anxiety, depression, PTSD, and trichotillomania/skin picking, and male biases in ADHD, impulse-control disorders, and SUD, are often attenuated in OCD. Tic/Tourette findings remain inconsistent. Other psychopathologies may reduce the male bias in childhood OCD and female bias in adult OCD. Some medical conditions may increase obsessive–compulsive symptom expression through stress-related or reactive mechanisms rather than sex-specific risk.

<u>Key concerns:</u> Assessment methods are highly heterogeneous, primary versus co-primary conditions are often unclear, and gender ratios are rarely interpreted relative to those in the general population or other psychiatric disorders.

<u>Topics for future study:</u> Standardized, low-burden assessment of comorbidities; prospective study of developmental trajectories alongside OCD symptoms; and testing how OCD–comorbidity interactions affect symptom severity, overall burden, and treatment outcomes.

Obsessive-compulsive disorder (OCD) often presents with co-occurring psychiatric symptoms and diagnoses, contributing to its clinical complexity. In a cross-sectional study of 955 adults with OCD in Brazil, only 7.7% had “pure” OCD without comorbid diagnoses (Torres 2013). In contrast, two large-scale registry studies reported lower comorbidity rates, ranging from 54% to 73% (Huang 2014; Rintala 2017). Methodological differences, such as direct interviews versus medical record reviews, as well as regional factors, may account for these discrepancies. These factors could also contribute to conflicting estimates regarding the impact of gender^3^ on the presence of co-occurring conditions in both adults and children with OCD.

In children, most studies do not report significant gender differences in the presence of at least one comorbid diagnosis (F:M OR = 0.71–1.43; Huang, 2014; Vivan, 2014; Arildskov, 2016; Ortiz, 2016). However, some studies report a significant male bias (F:M OR = 0.09; p < 0.01; Farrell, 2012), while others report a small but significant female bias (F:M OR = 1.22, p = 0.01; Rintala, 2017). Girls were more likely to show an internalizing rather than neurodevelopmental comorbidity pattern (F:M OR = 3.66; Ortiz, 2016), whereas boys were more likely to have comorbid neurodevelopmental disorders (F:M OR = 0.35; Rintala, 2017) and multiple comorbidities (F:M OR = 0.42; Arildskov, 2016).

In adults, findings are similarly mixed. Two studies from Brazil suggested a male bias in current comorbidity (F:M OR = 0.60–0.63; Torres, 2013; Torresan, 2013), while two other studies, from Taiwan and Italy, noted a female bias in lifetime comorbidity (F:M OR = 1.29–2; Huang 2014; Benatti 2020). One study, which used latent class analysis to identify specific clusters of comorbidities with OCD identified a specific subgroup cluster dominated by females, which included MDD (prevalence 21%), Dysthymia (13%), Manic Episode (8%), Hypomanic Episode (8%), Panic Disorder (21%), Agoraphobia (17%), Social Phobia (38%), PTSD (8%), Alcohol Dependence (8%), Drug Dependence (4%), Psychosis (13%), GAD (55%), Specific Phobia (33%) Body Dysmorphia (8%), Tics (8%) and Hair-Pulling (13%), however the small sample size may limit the precision and generalizability of these results (total n=257, cluster n=24, Brakoulias, 2025). These inconsistencies highlight the need for further research across diverse and representative populations, and the importance of accounting for methodological biases.

This section explores gender differences in the prevalence and presentation of specific comorbid symptoms and diagnoses in OCD based on results from 178 studies identified during the review period (56 in children/adolescents, 114 in adults, and 8 in both; Table A4.1). The observed gender effects on co-occurrence or comorbidity could be driven by both biological and social factors. Their interpretation depends on the research or clinical goal. Clinically, recognizing gender-specific comorbidity patterns is crucial for optimizing diagnostic and treatment practices. From a pathophysiological perspective, deviations from general population patterns are more informative than simple descriptions of comorbidity patterns. For instance, since depression is more common in girls and women in the general population, its equal prevalence among men and women with OCD would signify a noteworthy deviation, warranting further investigation. This section is structured to facilitate these comparisons (see Table 2).

**Table 2.**
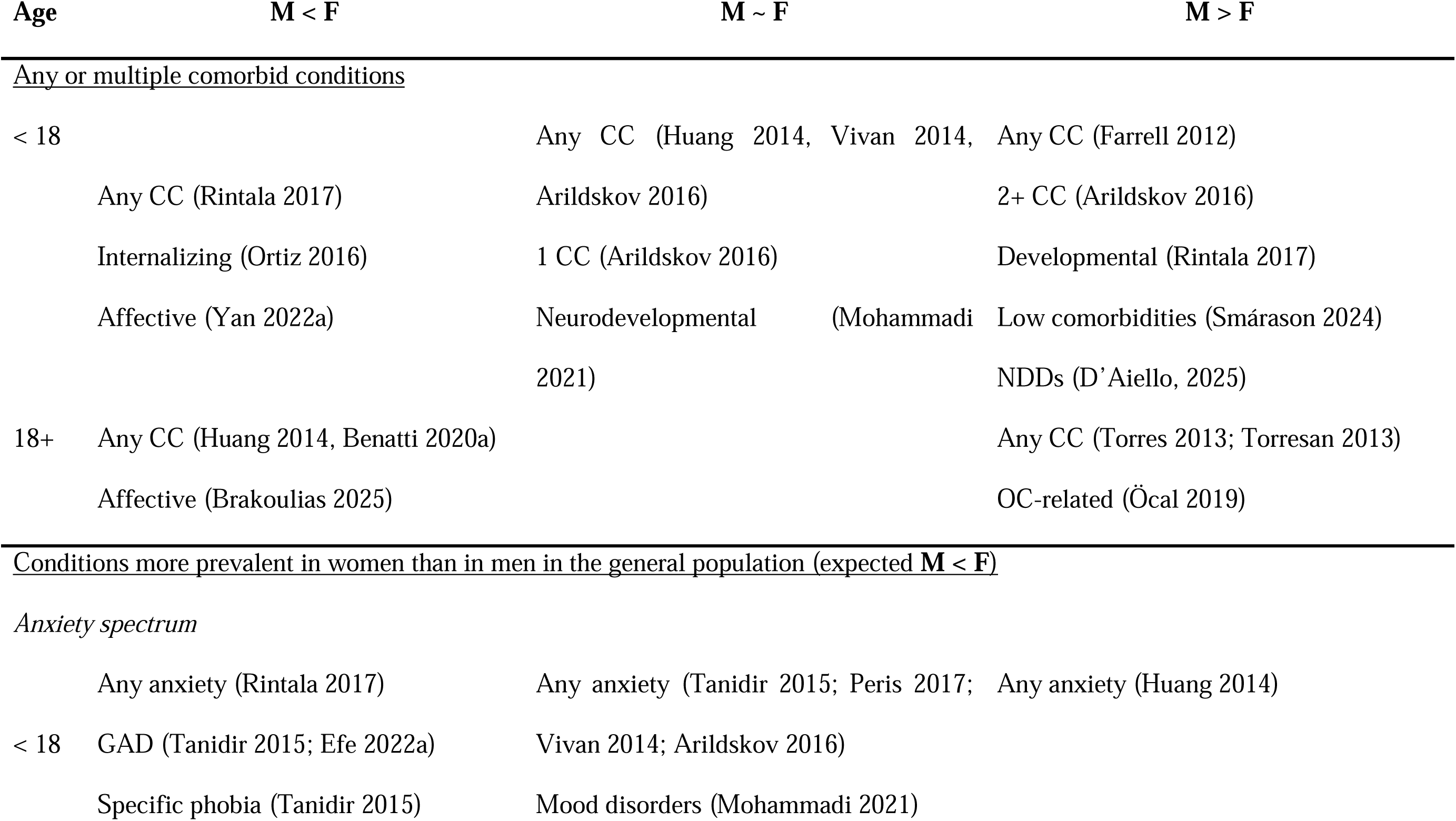
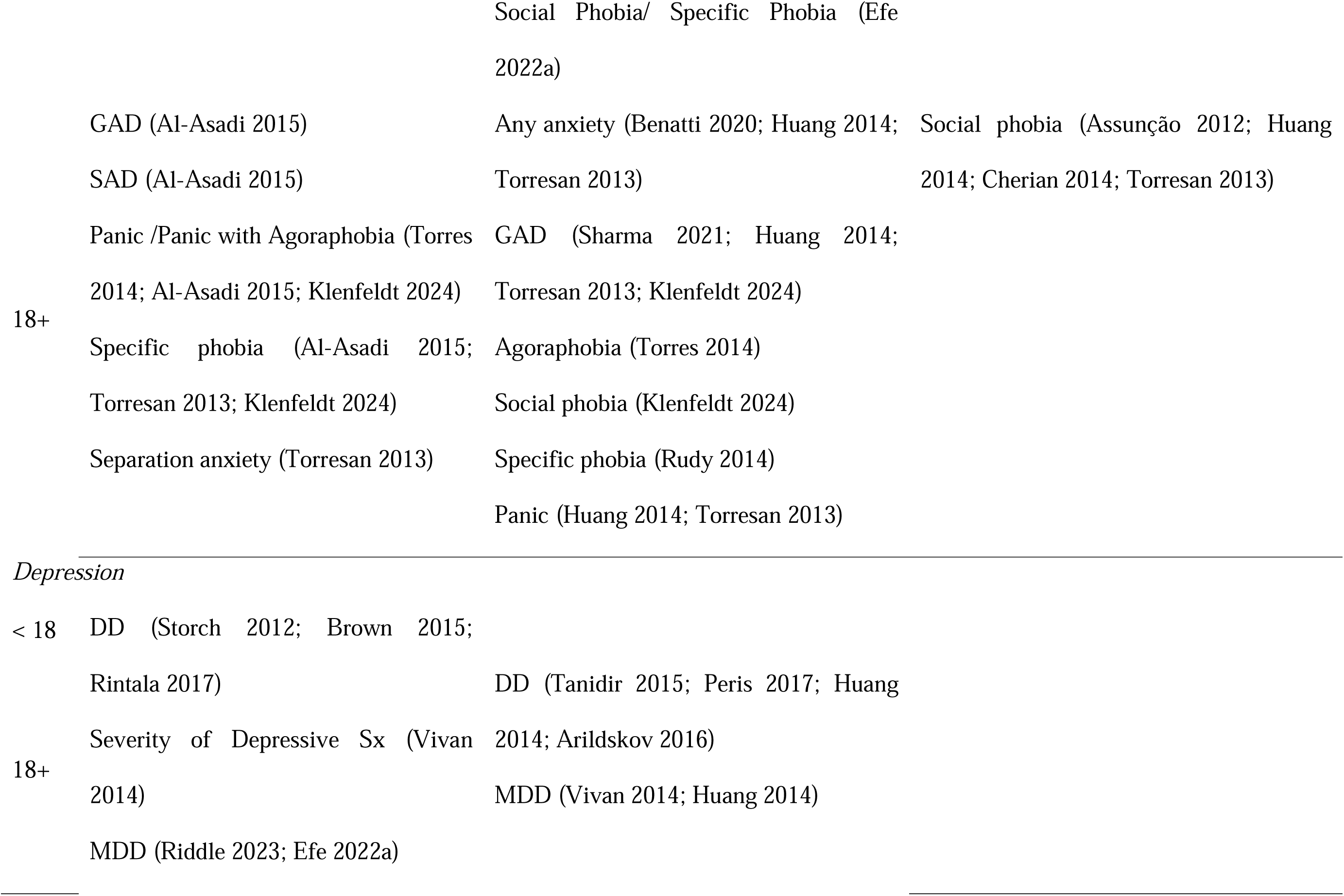
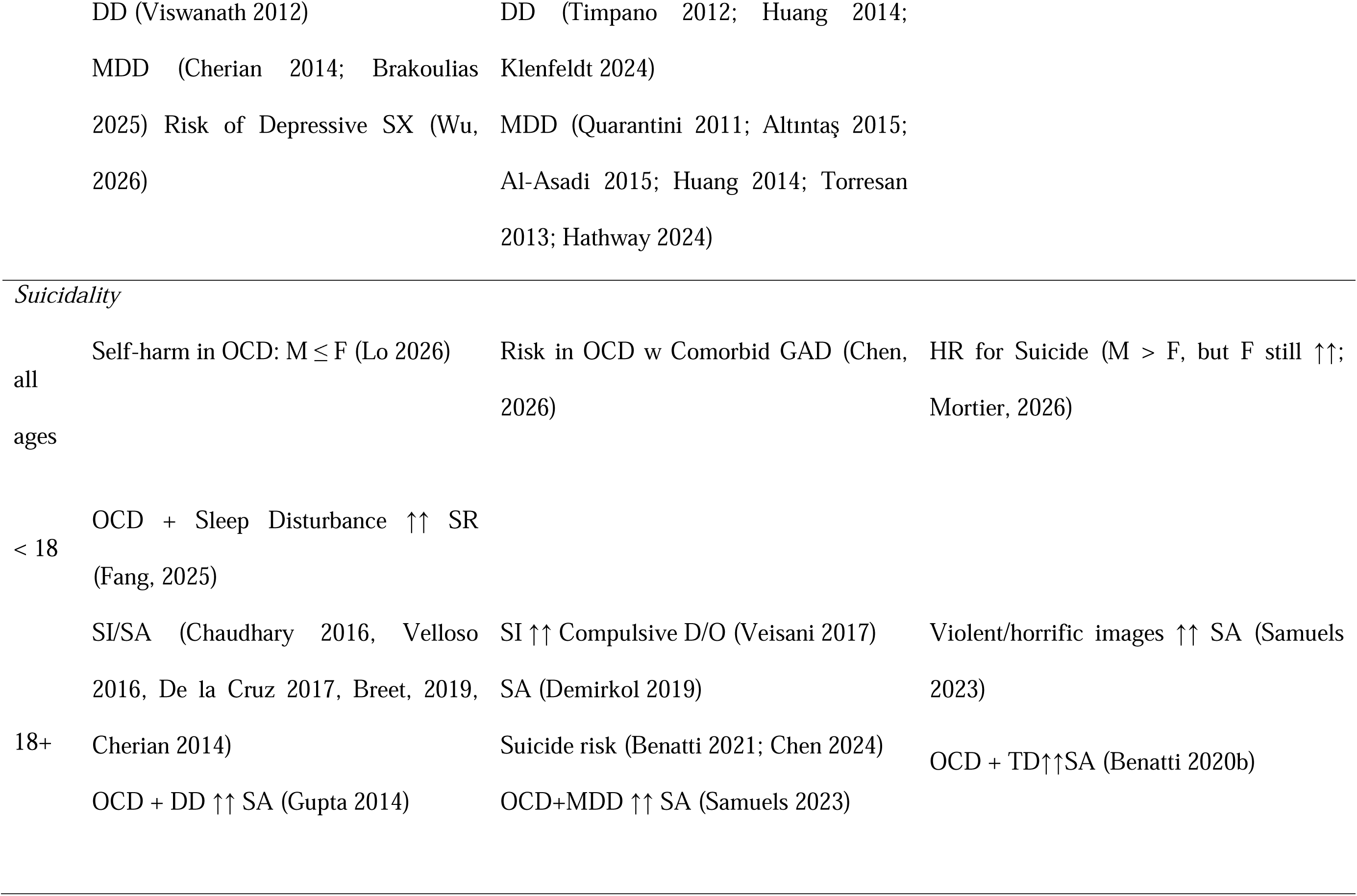
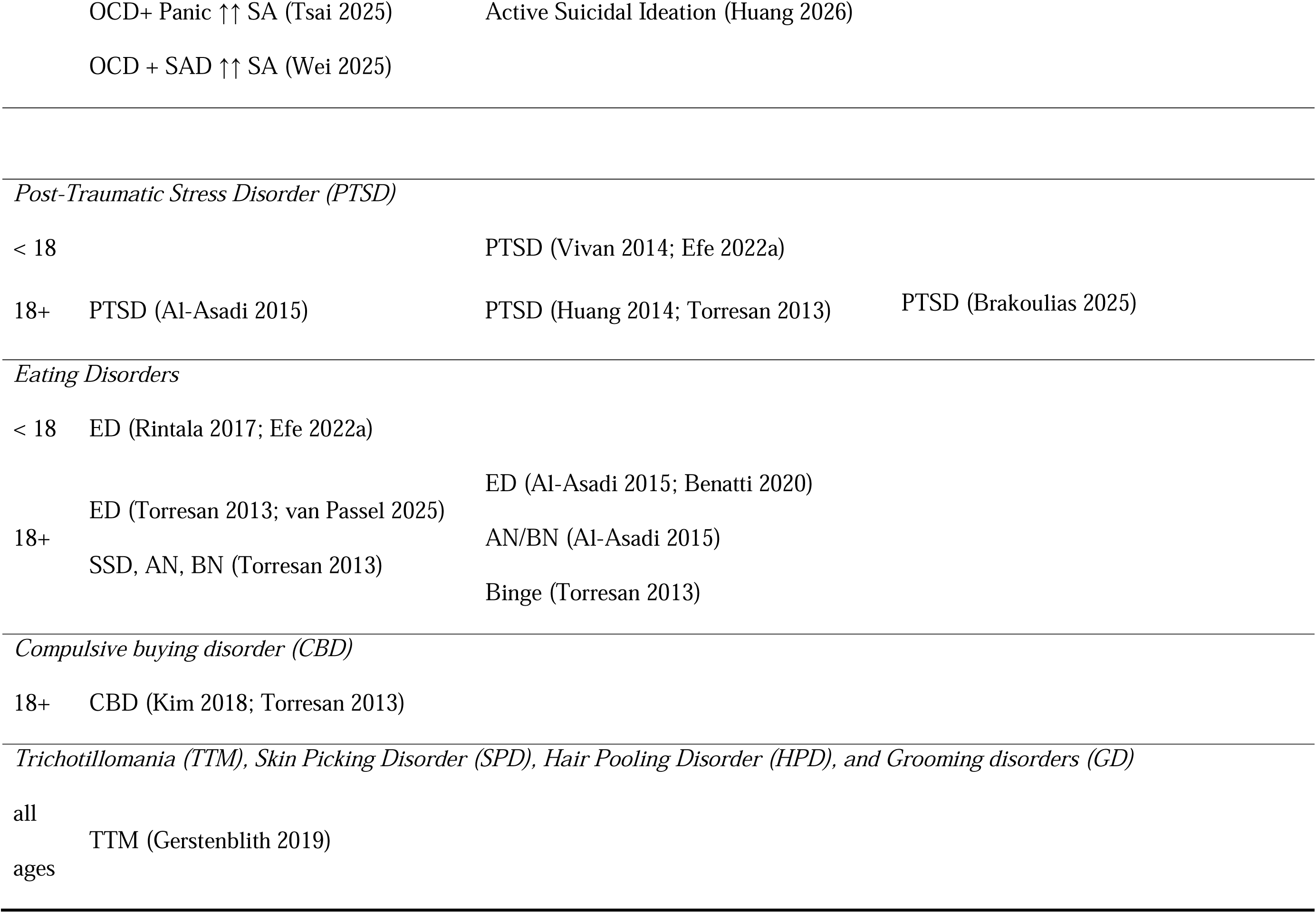
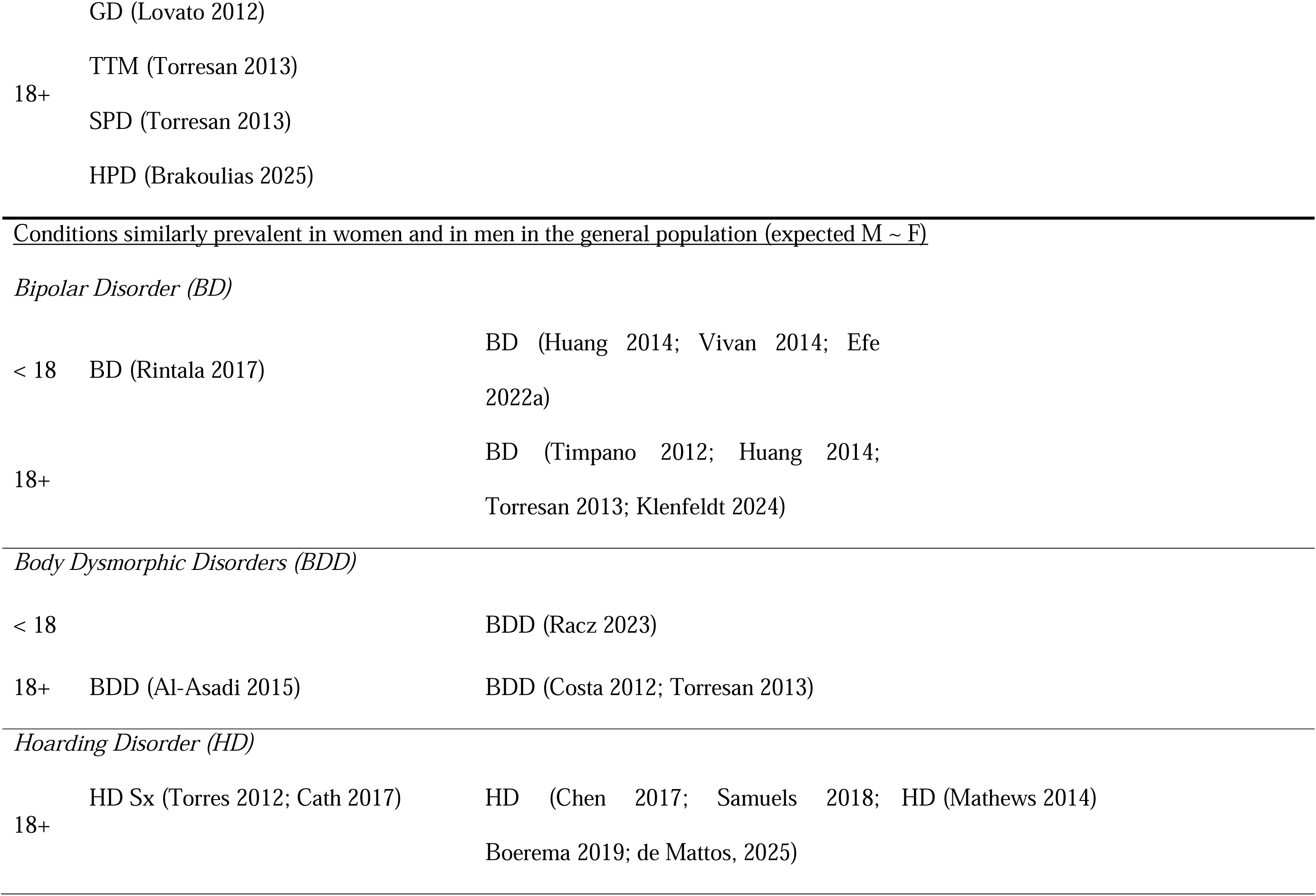
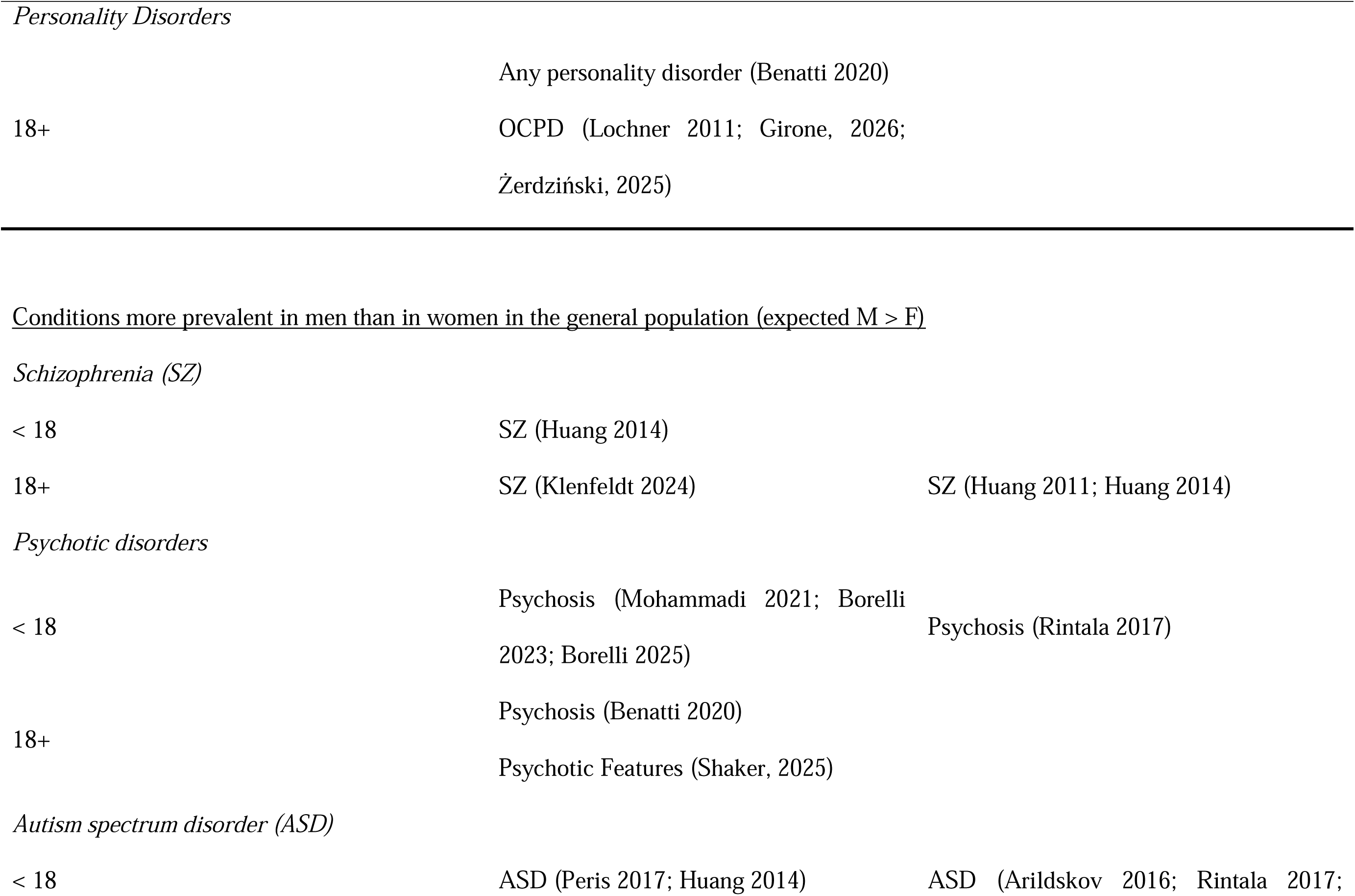
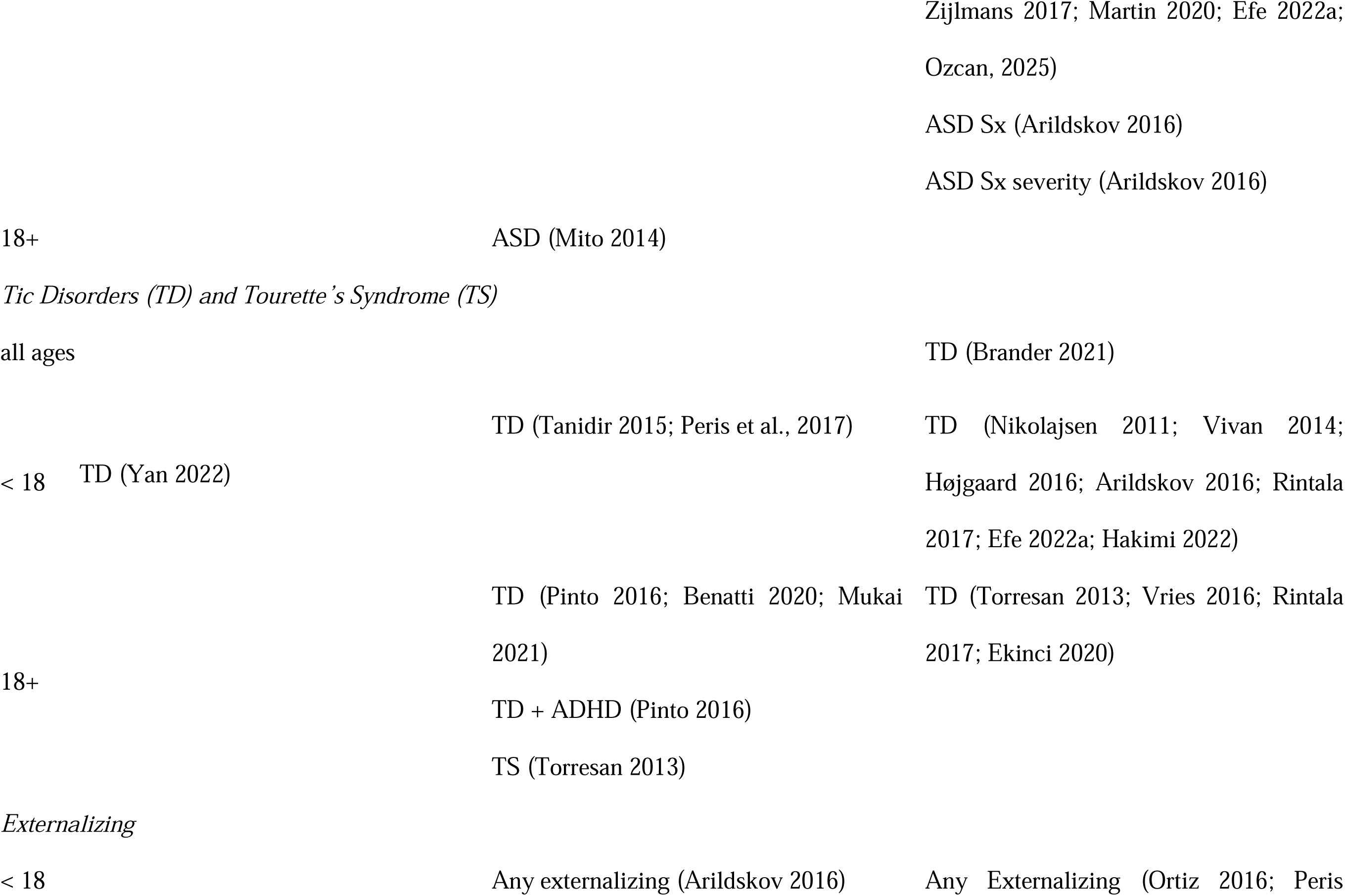
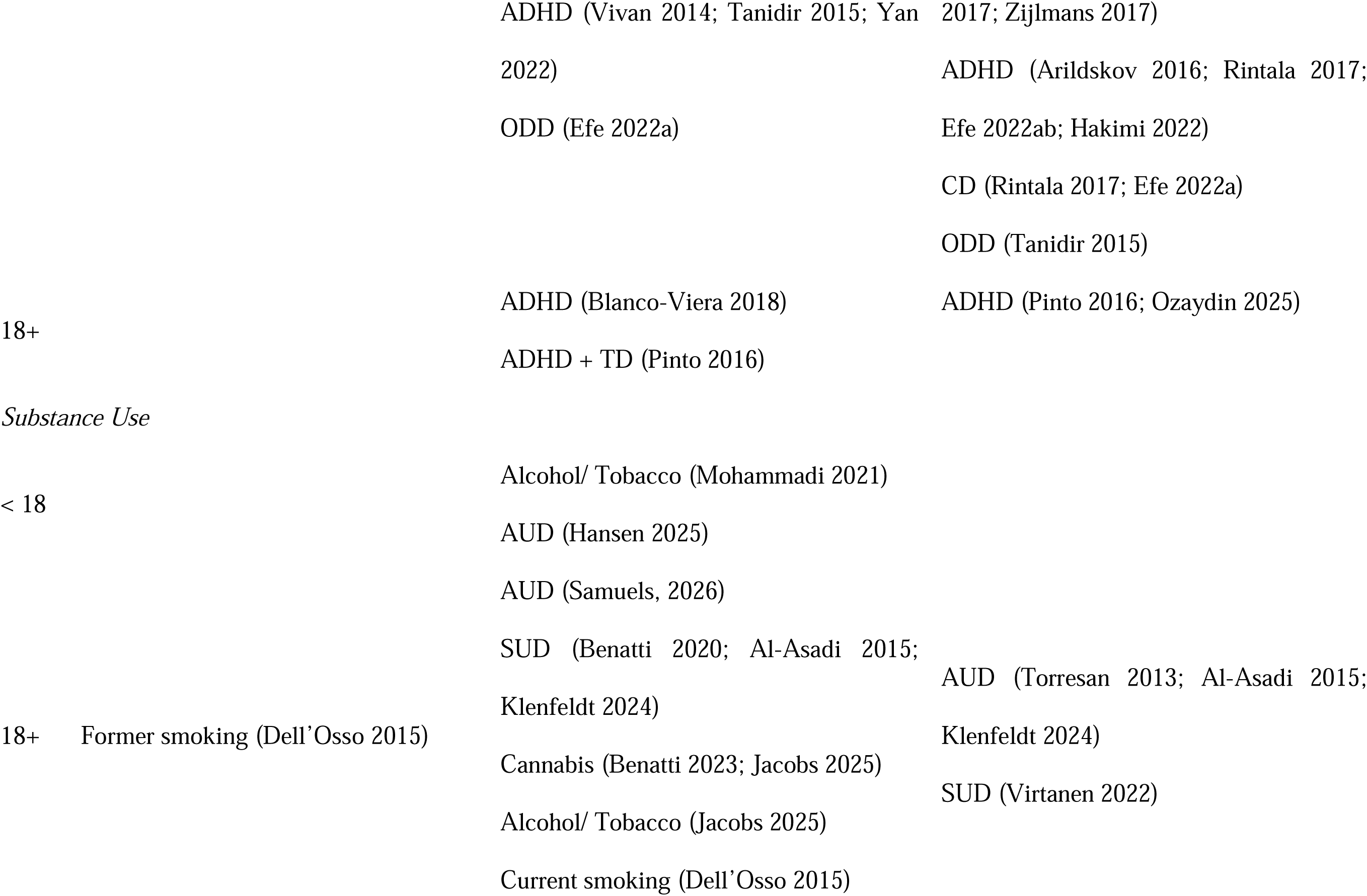
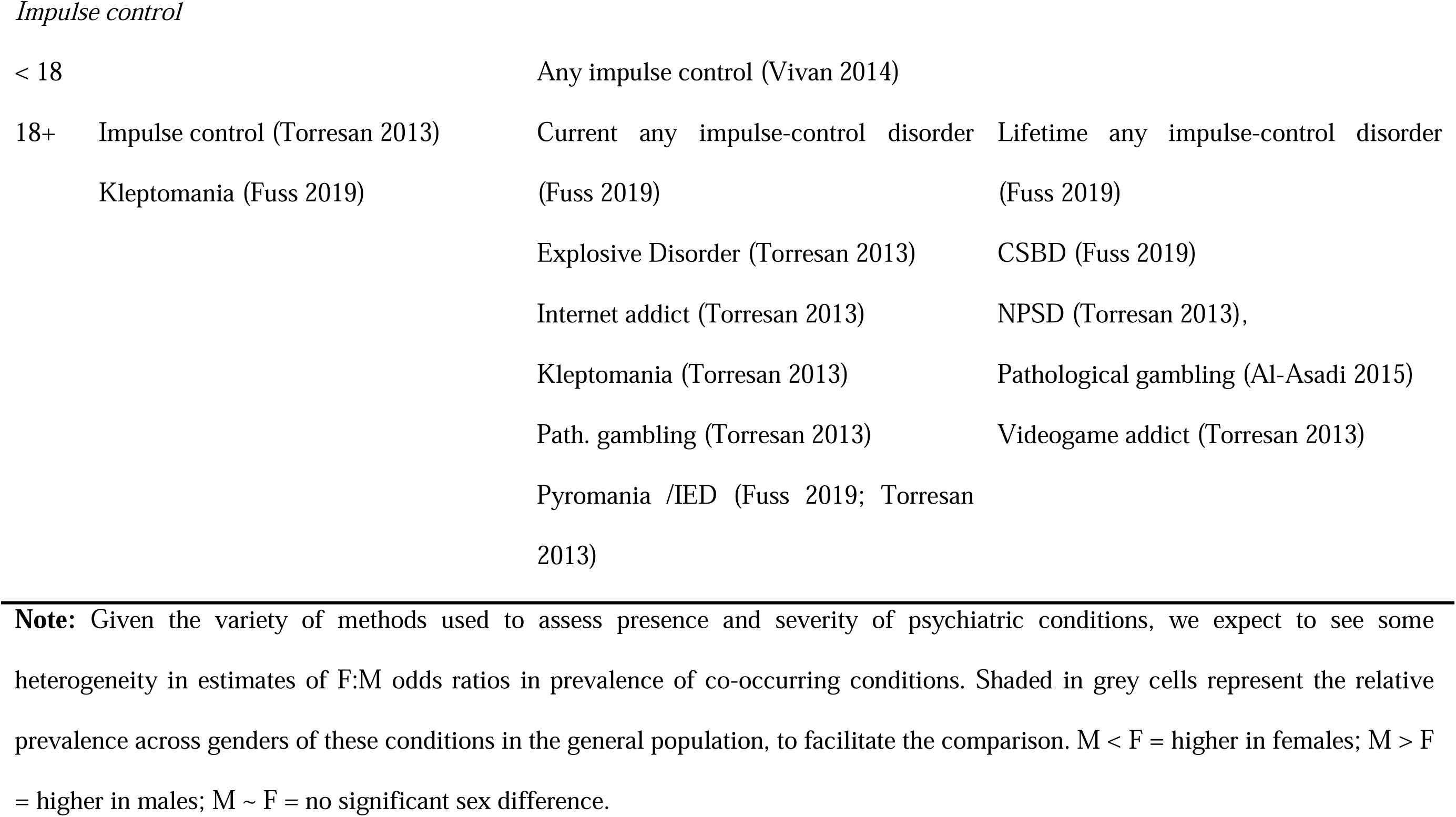
Gender differences in prevalence of comorbid and cooccurring conditions in OCD as compared to that in the general population.

Of note, comorbidity is reported in two different ways in the literature. Most studies retrieved by our search described individuals with OCD and reported the occurrence of other conditions. But others took the converse approach, describing individuals with another condition and reporting the occurrence of OCD within this group. Both types of studies are described here (Table A4.1, Table 2, Table 3).

**Table 3.**
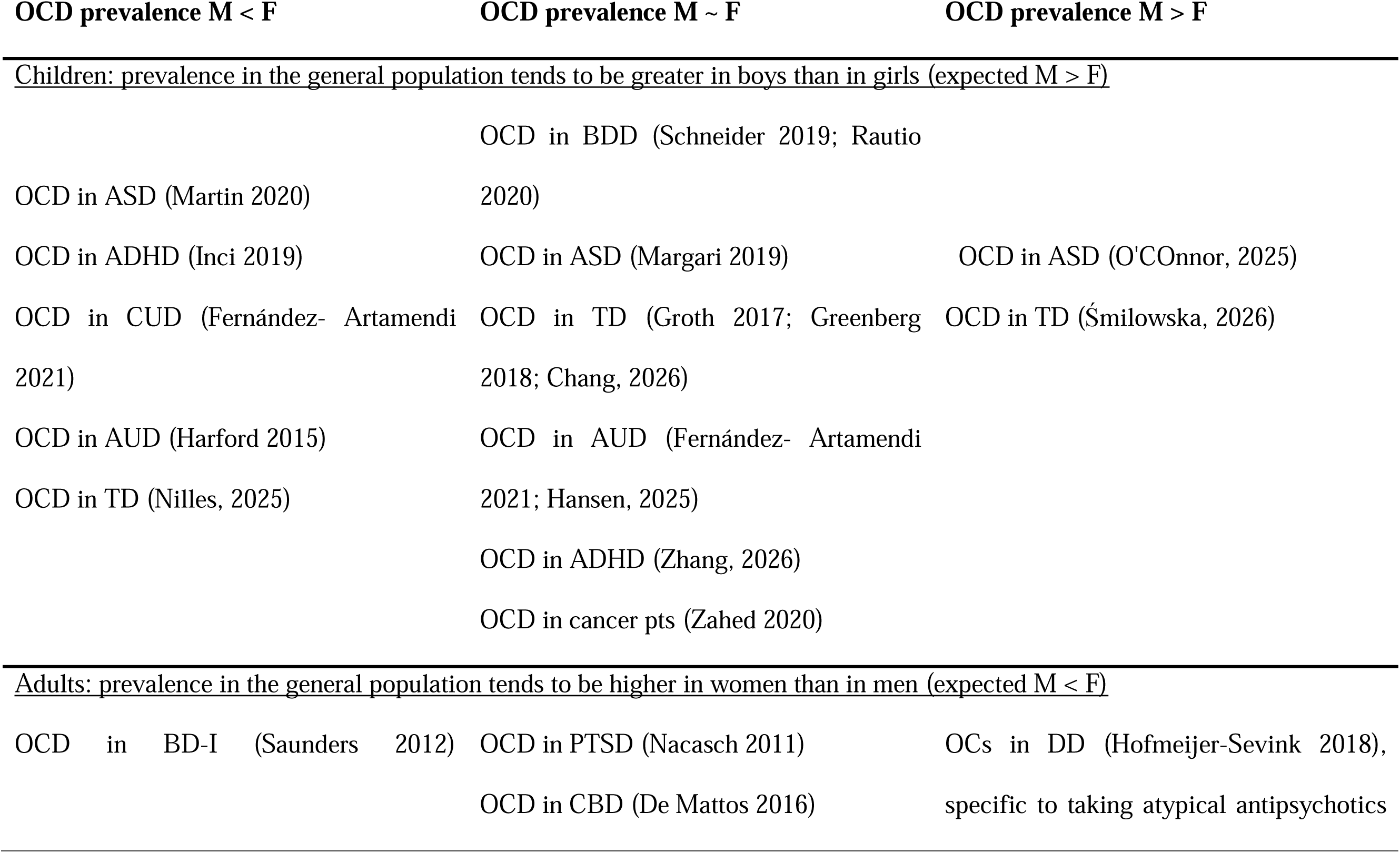
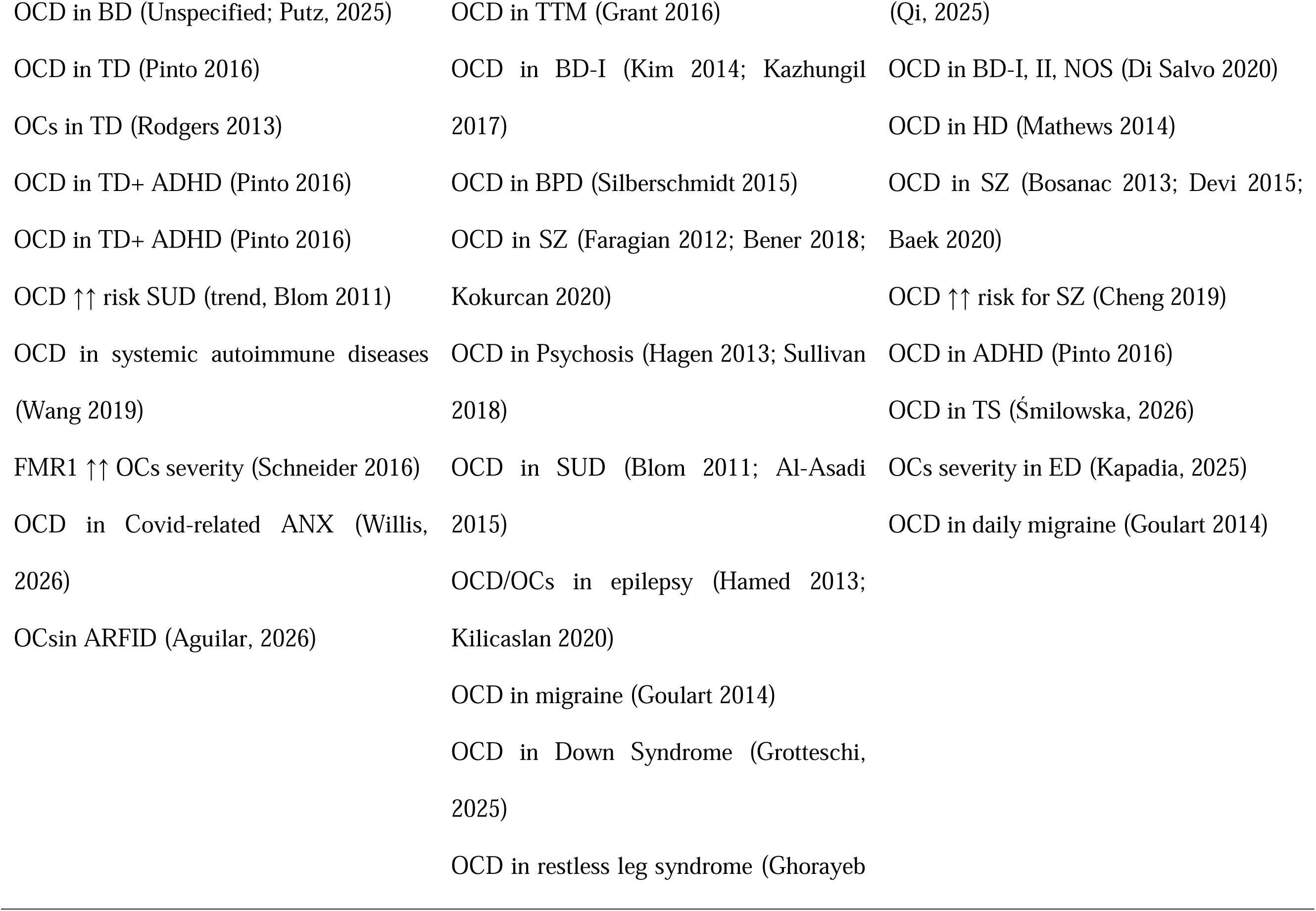
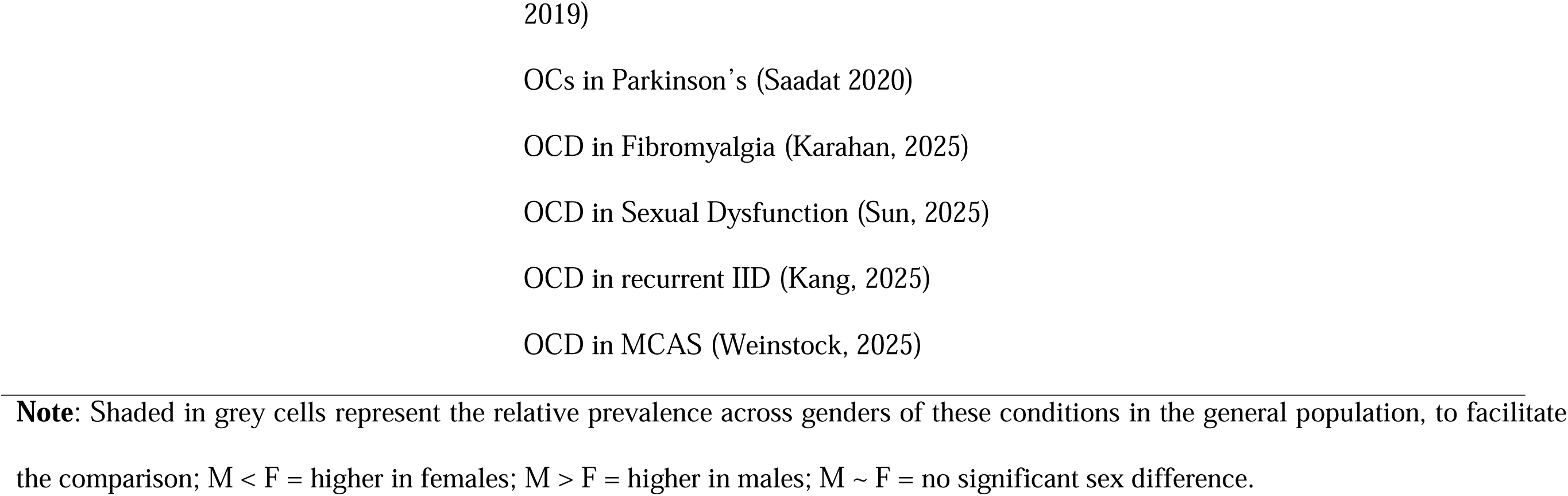
Gender differences in prevalence of OCD in other Psychiatric and general medical conditions as compared to that in the general population.

#### 3.4.1. Psychiatric Conditions with Higher Prevalence in Women in the General Population

*3.4.1.1. Anxiety Disorders* are more common in women in the general population (Bekker & van Mens-Verhulst, 2007); this includes generalized anxiety disorder (GAD) (Grenier et al., 2019), panic disorder (Sheikh et al., 2002), and social anxiety disorder (Ohayon & Schatzberg, 2010). Most individuals with OCD experience anxiety alongside their intrusive thoughts (Bartz & Hollander, 2006; Citkowska-Kisielewska et al., 2019; Montgomery, 1993). However, the relationship between comorbid anxiety disorders, OCD, and gender is complex and not fully understood. These patterns may differ in pediatric and adult samples.

In pediatric OCD, most studies report equal rates of any comorbid anxiety disorders in boys and girls (Peris, 2017, N = 332; Tanidir, 2015, N = 110; Vivan, 2014; Arildskov, 2016). However, some contradictory findings exist. Rintala et al. (2017) reported a female bias in the prevalence of anxiety disorders in a large sample aged 3–25 years old (N = 3372), while Huang et al. (2014) found a male bias in a smaller sample of children aged 6–17 years old (N = 48). Generalized anxiety disorder (F:M OR = 2.4, p = 0.08) and specific phobias (F:M OR = 2.75, p = 0.03) tend to be more common in girls with OCD (Tanidir, 2015), with a similar female-directional trend for GAD reported by Efe (2022a). In a sample of 457 adolescents with OCD in Finland, the prevalence of separation anxiety, PTSD, panic disorder, social phobia, and specific phobias did not differ between males and females (Efe, 2022a).

In adults, rates of any comorbid anxiety disorder are similar in men and women with OCD (F:M OR = 0.77–1.21; Benatti, 2020; Huang, 2014; Torresan, 2013). However, there are possible differences in specific diagnoses. Studies that relied on clinical interviews and medical records reported comorbid social anxiety disorder to be more prevalent in men with OCD (F:M OR = 0.21–0.73, p < 0.05; Assunção, 2012; Huang, 2014; Cherian, 2014; Torresan, 2013); however, one online study found it to be more common in women (F:M OR = 1.75, p < 0.01; Al-Asadi, 2015) and another found no difference between the sexes (Klenfeldt, 2024). Panic disorder with and without agoraphobia was reported to be more common in women with OCD (F:M OR = 1.47–4.15, p < 0.05; Torres, 2014; Klenfeldt, 2024). Reports are inconsistent about GAD (F:M OR = 1.95, p < 0.01 in Al-Asadi, 2015, N = 13,414; F:M OR = 0.92, p = 0.54 in Sharma, 2021, N = 867; no gender modification found in OCD for BAI scores in Kim, 2022 and Klenfeldt, 2024) and specific phobias (F:M OR = 1.58, p < 0.01 in Al-Asadi, 2015, N = 13,414; F:M OR = 0.61, p = 0.42 in Rudy, 2014, N = 196). A clustering analysis in 257 individuals with OCD identified a specific female-predominant sub-cluster (n = 24, 13% male proportion) and another, larger sub-cluster with approximately equal gender distribution (Brakoulias, 2025). In aggregate, these findings suggest that social anxiety disorder may be relatively more common in adult men than women with OCD. If confirmed, this trend may help explain why men with OCD are more likely to remain unmarried (see Section 3.2.2).

*3.4.1.2. Depression*, including major depressive disorder (MDD), is diagnosed about twice as often in women as in men in the general population (Klein et al., 2013; Salari et al., 2020) and is more common in adults over 18 years of age (Kessler et al., 2005; Kessler et al., 2012). Reports on the prevalence of comorbid depression in OCD are mixed. Among children, some studies indicate a significant female bias in comorbid depression prevalence (F:M OR = 2.0–4.1, p < 0.01; Storch, 2012; Rintala, 2017), while others report only a non-significant trend toward female bias (F:M OR = 1.2–1.94, p > 0.10; Brown, 2015; Tanidir, 2015; Peris, 2017). Two studies of adolescents with OCD or MDD found higher prevalence of the co-occurring disorder (F:M OR = 1.58–3.8; Vivan, 2014; Riddle, 2023) and significantly higher depression severity in women (Cohen’s d = 0.30, p = 0.03; Vivan, 2014). In adults with OCD, three studies reported significantly greater comorbid depression or depressive symptoms in females (Viswanath, 2012; Cherian, 2014; Wu, 2026) or a trend toward such a bias (F:M OR = 1.4, p = 0.2; Altıntaş, 2015), while six others found no significant gender effect (F:M OR ≅ 1.1, p > 0.10; Quarantini, 2011; Timpano, 2012; Al-Asadi, 2015; Kim, 2022; Hathway, 2024; Klenfeldt, 2024). In a latent class analysis of comorbidities in OCD which found six clusters of characteristics, one cluster characterized by high comorbidity with MDD and anxiety-spectrum disorders, and hair pulling, was also predominantly female (Brakoulias, 2025).

Conversely, in adults with a diagnosis of depression, obsessive-compulsive symptoms are more common in men (F:M OR = 0.36, p < 0.01; Hofmeijer-Sevink, 2018). In patients taking atypical antipsychotics to treat their MDD, comorbid OCD was reported more often in men (Qi, 2025), suggesting that treatment context may contribute to the observed gender difference rather than MDD itself. Overall, findings in OCD are mixed, although several studies suggest a female bias in comorbid depression, which may be more pronounced in younger individuals. Conversely, the greater prevalence of OCS among adult men with depression raises the possibility that the relationship between OCD and depression may differ by gender. This hypothesis needs to be tested in larger, balanced samples.

*3.4.1.3. Suicide* attempts in the general population are more common in women; however, suicide completion rates are 2–3 times higher in men (Turecki et al., 2019). OCD is associated with increased risk of suicidal ideation and behavior. Comorbid depression further amplifies this risk, particularly in women (de la Cruz, 2017; Chaudhary, 2016; Gupta, 2014), as do comorbid eating disorders in women (Spinner, 2026). Additional comorbidities linked to elevated suicide risk include panic disorder (Tsai, 2025) and ADHD (Chen, 2024), whereas Tourette syndrome (Benatti, 2020) and autism spectrum disorders (Chen, 2024) appear to confer greater risk in men.

Although several studies report no significant gender differences in suicide risk among individuals with OCD (Demirkol, 2019; Benatti, 2021; Chen, 2024; Chen, 2026; Huang, 2026), others indicate that women are more likely than men to experience suicidal ideation or attempt suicide (F:M OR = 1.45–19.33, p < 0.01; Chaudhary, 2016; Velloso, 2016; de la Cruz, 2017; Breet, 2019; Wei, 2025). Self-harm also showed a female-directional association in OCD (Lo, 2026). One study reported a female-specific association between sleep disturbance and suicide risk in adolescents with OCD (Fang, 2025). Another study reported significantly elevated serum estradiol in OCD patients with active suicidal ideation in both males and females (Huang, 2026), although whether this association differs by gender remains unclear. One study examining suicide and self-harm in Spanish patients found that, while overall the association of any mental disorder with suicide was higher in females than males (HR = 35.6 vs. 12.2), the disorder-specific association for OCD was stronger in males (HR = 17.4) than females (HR = 8.9; Mortier, 2026). Thus, although suicidal ideation and attempts frequently show a female bias in OCD, the risk of death by suicide may be greater in men, broadly paralleling patterns observed in the general population. In some studies, OCD may further amplify these sex differences, although this is not observed consistently. These sex differences may partly reflect differences in comorbid depression and other risk factors among women and men with OCD.

*3.4.1.4. Post-traumatic stress disorder (PTSD)* tends to be more common in women in both civilian and military populations (Lehavot et al., 2018; Schein et al., 2021). The co-occurrence of PTSD and OCD is understudied. One study in adults with OCD replicated the female bias in the prevalence of PTSD (F:M OR = 2.58, p < 0.01; Al-Asadi, 2015). Other studies in children and adults found a similarly low prevalence of PTSD in both genders (F:M OR = 0.26–1.25, p > 0.20; Vivan, 2014; Efe, 2022a; Huang, 2014; Torresan, 2013). In contrast, one latent class analysis identified a male-predominant PTSD class, although this difference was not statistically significant (Brakoulias, 2025). In a small study from Israel of individuals with PTSD, 41% of participants were also diagnosed as having current OCD, with no significant differences between genders (Nacasch, 2011; N = 44). Similarly, OCD prevalence did not differ significantly by gender among military personnel with PTSD (Walter, 2022).

*3.4.1.5. Eating disorders (EDs)* include anorexia nervosa (AN), bulimia nervosa (BN), binge eating disorder (BED), orthorexia, and related conditions. AN and BN often involve binge eating and purging. In the general population, AN and BN are more common in women, while subthreshold BED is more frequent in men (Hoek, 2006; Striegel-Moore & Bulik, 2007; Striegel Moore et al., 2009). The prevalence of "any binge eating" is similar across genders (F:M OR ≈ 1.2).

Eleven studies examined sex/gender differences in the co-occurrence, associations, or treatment response of OCD/OCS and EDs. ED symptoms tend to be elevated in both men and women with OCD (Bang, 2020), and the severity of obsessive-compulsive symptoms (OCS) correlates with the severity of ED symptoms similarly in both genders in adolescents (Schneider, 2018). In pediatric OCD samples, eating disorders (ED) appear to be more prevalent in girls (F:M OR = 5.66, p < 0.01; Rintala, 2017). One study, examining the associations of polygenic risk scores (PRS) for AN with OCD characteristics in adolescents, found that only girls showed a significant association between AN PRS and OCD perfectionism (Aicoboaie, 2025). However, severity patterns may differ by sex: one study reported that male adolescents with ED exhibit greater obsessive–compulsive symptom severity (p = 0.012; Riva, 2023). In adults, findings are mixed: EDs, and specifically AN and BN, were similarly prevalent in men and women in some studies (Al-Asadi, 2015; Benatti, 2020), whereas other studies found EDs to be more prevalent in women with OCD (Torresan, 2013; van Passel, 2025). One study, examining OCD symptom dimensions in ED patients, found that gender minority patients exhibited the most severe OCD symptoms, followed by males, then females (Kapadia, 2025), suggesting that EDs might have a greater exacerbating effect on symptoms in males. No study during the review period investigated whether the reversal of the gender differences in prevalence of EDs in the general population around the clinical cut-off levels is also observed in individuals with OCD. Interestingly, Strobel (2019) reported that in AN and BN patients undergoing ED-focused inpatient treatment, obsessive-compulsive symptom severity decreased in both men (Cohen’s d = 0.59, p < 0.001) and women (Cohen’s d = 0.37, p < 0.001), with a stronger treatment response observed in men (ΔOCI-R × M η² = 0.02, p < 0.05).

Seven studies in children/adolescents, eleven in adults, and two in mixed-age samples report significant associations between OCD/OCS and eating disorders in women, although the specific obsessive–compulsive symptom profiles vary across eating-disorder subtypes. In girls with EDs—particularly the binge/purge subtype of anorexia nervosa—obsessive–compulsive symptoms are more severe than in those without EDs (Brooks, 2014; Blachno, 2016; Lewis, 2019; Tyszkiewicz-Nwafor, 2021; De Mey, 2025; Elm, 2026). In adult women, OCD severity correlates positively with ED severity, with higher symptom scores observed in patients with ED compared with healthy controls (Amianto, 2022; Halls, 2023; Wilson, 2025), and a study in both adults and adolescents found body-focused repetitive behaviors in ED patients significantly correlate with worsening OCs (Barber, 2025). One study found that the COVID-19 pandemic did not significantly impact OCD severity in AN patients, however, OCs were indicated to have strong associations with AN in a network analysis (Collatoni, 2025). Moreover, treatment-related reductions in OCD symptoms are accompanied by decreases in ED severity (Lee, 2018; 2020; Stender, 2025; Monaco, 2025). Impulse-related symptoms on the Padua Inventory (Burns et al., 1996) are higher in individuals with AN binge/purge subtype and BN compared to those with AN without binge eating (Claes, 2021). Neutralizing symptoms are notably elevated in women with AN (Cohen’s d = 0.47, p = 0.053; Bang, 2020). A latent class analysis in a female sample with anorexia nervosa identified a subtype characterized by high compulsivity and increased OCD comorbidity (OR = 5.6 [95% CI, 1.77–17.71], p < 0.01; Lavender, 2014). Women with eating disorders who engage in non-suicidal self-injury also exhibit elevated obsessive– compulsive symptoms (Claes, 2021). Together, these findings point to shared compulsive and impulsive traits across OCD and EDs, underscoring the importance of integrated approaches to understanding and treating these conditions.

Avoidant food intake disorder (ARFID) has been generally thought to be more prevalent in males (Becker et al., 2020; Forman et al., 2014; Keery et al., 2019; Peebles et al., 2006), although there are some contradictions. A recent study indicated no sex bias (Sader et al., 2023), and one study in adults in the UK and USA found an increased female prevalence of ARFID (Brownlow et al., 2026), while another in Australian students found a small but significant increase in the ratio of females to males with possible ARFID diagnosis (1.7:1; (Van Buuren et al., 2023)). One study during the review period examined sex differences in OCS among older adolescent and adult ARFID patients; females were found to have significantly higher OCI-R scores and higher scores on the obsessing, ordering, neutralizing, and hoarding subscales, while washing and checking did not differ by sex (Aguilar, 2026), indicating a potential exacerbating factor for OCD symptoms in female patients with ARFID.

*3.4.1.6. Compulsive buying disorder* is more common in females in the general population (Maraz et al., 2016). Two studies in adults with OCD found that compulsive buying disorder was more common in women than men (F:M OR = 2.56–2.8, p < 0.01; Torresan, 2013; Kim, 2018). In individuals with a diagnosis of compulsive buying, there was a non-significant trend toward a male bias in prevalence of OCD (F:M OR = 0.49, p = 0.20; De Mattos, 2016).

*3.4.1.7. Trichotillomania (TTM), excoriation disorder (SPD), and other grooming disorders* are about three times more common in females than males in the general population (Chamberlain et al., 2007; Wilhelm et al., 1999). Five studies during the review period examined gender differences in the co-occurrence of OCD and TTM, SPD, or other grooming disorders. Among individuals with OCD or a family history of OCD, a smaller female bias is observed for TTM (F:M OR = 1.08–1.46, p < 0.05; Torresan, 2013; Gerstenblith, 2019), SPD (F:M OR = 2.61, p < 0.01; Torresan, 2013), and grooming disorders more generally (F:M OR = 2.27, p < 0.01; Lovato, 2012). A female bias in hair pulling was also observed in a latent class analysis of adults with OCD (Brakoulias, 2025). This may suggest that the female bias for TTM is attenuated in individuals with OCD, which should be tested in future studies. In adults diagnosed with TTM, OCD is similarly prevalent in men and women (Grant, 2016).

Overall, many disorders that are more common in women generally are also more prevalent in women with OCD. However, the presence of OCD may attenuate or modify some of these expected gender differences, including for social phobia, trichotillomania, and depression. While OCD raises the risk of suicidal ideation and attempts in both genders, suicidal ideation and attempts tend to show a female bias, whereas the risk of death by suicide may be greater in men; in some studies, OCD may further amplify these sex differences, although this is not observed consistently. Gender differences in eating disorder prevalence in OCD remain unclear, especially around the clinical cut-off levels. OCS severity correlates with ED severity in both genders, but men show greater OCS reduction after inpatient anorexia treatment. Boys, men, and gender minority individuals may also show greater OCS severity in some ED samples. Reports on gender effects on comorbid anxiety disorders in OCD are highly variable. PTSD co-occurrence with OCD is understudied and likely underestimated.

#### 3.4.2. Psychiatric Conditions with Similar Prevalence in Women and Men in the General Population

*3.4.2.1. Bipolar disorder (BD)* has similar prevalence in men and women in the general population, though onset may occur later in women, with symptom presentation differing by gender (Kennedy et al., 2005; Kessing, 2004; Parial, 2015; Suominen et al., 2009). Twelve studies during the review period examined gender differences in the co-occurrence of OCD and BD. Among individuals with OCD, studies report mixed findings. In children with OCD, BD shows a significant female bias (F:M OR = 1.8, p = 0.003, N = 3372; Rintala, 2017), while three studies found similar prevalence in boys and girls (Huang, 2014; Vivan, 2014; Efe, 2022a). Adult studies generally report similar prevalence in men and women (Timpano, 2012; Huang, 2014; Torresan, 2013; Klenfeldt, 2024). In BD-I, a large study noted a significant female bias (F:M OR = 2.5, p = 0.027, N = 736; Saunders, 2012), while smaller studies reported non-significant trends (F:M OR = 1.7, p = 0.28, N ≤ 174; Kazhungil, 2017; Kim, 2014). Another study, which examined BD without specifying subtypes, found a higher incidence of OCD comorbidity in women (OR = 2.24, p = 0.003; Putz, 2025). Conversely, a large-scale study of individuals with any BD diagnosis (BD-I, BD-II, or unspecified BD) found a significant male bias for OCD (F:M OR = 0.61, p < 0.01; Di Salvo, 2020). These findings suggest potential links between OCD and BD-I in women, while findings across broader BD samples remain inconsistent, and indicate that studies most likely need to distinguish between different subtypes of BD for a true understanding of gender effects.

*3.4.2.2. Body dysmorphic disorder (BDD)* occurs equally in men and women in the general population (Koran et al., 2008; Taqui et al., 2008) and was examined in relation to OCD/OCS in 10 studies during the review period. BDD is similarly prevalent in men and women among adults with OCD (F:M OR = 0.83–0.94; Costa, 2012; Torresan, 2013). However, one online study reported a higher prevalence of BDD among women with clinically significant self-reported obsessive–compulsive symptoms (F:M OR = 2.9, p < 0.01; Al-Asadi, 2015). In contrast, another study found similar BDD prevalence across sexes among adolescents with and without OCD (F:M OR = 0.87, p = 1.00; Racz, 2023). In children, OCD symptoms correlate similarly with BDD interference/avoidance symptoms in both genders, but other BDD symptoms correlate more strongly in girls (Spearman ρ: girls = 0.32, boys = 0.26; Fisher p = 0.038; Schneider, 2018). OCD prevalence in children with BDD appears similar across genders (Schneider, 2019; Rautio, 2022). A twin study in women found significant genetic overlap between OCD and BDD (polychoric correlation = 0.34; Monzani, 2012). In young adult women, OCD significantly increases BDD risk (OR = 7.2, p < 0.01), a pattern not observed in adolescents (Enander, 2018). Another study indicates that BDD and muscle dysmorphia specifically may be a risk factor for elevated OCS in young adult men (Çınaroğlu, 2025).

*3.4.2.3. Hoarding disorder (HD)* occurs equally in men and women in the general population (Postlethwaite et al., 2019). Seven studies during the review period examined gender differences in HD or HD symptoms in relation to OCD. Among adults with OCD, most studies also report similar HD comorbidity rates across genders (Chen, 2017; Samuels, 2018; Boerema, 2019; de Mattos, 2025), though contradictory reports exist. A twin study found higher HD prevalence in men with OCD (F:M OR = 0.5, p < 0.01; Mathews, 2014), whereas another study found that elevated HD symptoms were more common in women with OCD (F:M OR = 1.41–2.13, p < 0.01; Torres, 2012; Cath, 2017). In individuals with HD, comorbid OCD is more common in men (F:M OR = 0.59, p < 0.01; Mathews, 2014). Women with comorbid OCD and HD are more likely to report dysfunctional maternal care (Chen, 2017) and greater executive function difficulties (Samuels, 2018). Thus, while the gender balance of HD comorbidity in OCD remains unclear, converging evidence suggests that women may be more severely affected by this comorbidity.

*3.4.2.4. Personality disorders*, including borderline personality disorder (BPD) and obsessive-compulsive personality disorder (OCPD), have similar prevalence in men and women in the general population (Clemente et al., 2022; Friborg et al., 2013; Silberschmidt et al., 2015). Five studies during the review period examined gender differences in personality disorders in relation to OCD. In OCD, studies report equal prevalence of OCPD and other personality disorders in men and women with OCD (Lochner, 2011; Benatti, 2020, Girone, 2026; Żerdziński, 2025). In individuals with borderline personality disorder, OCD comorbidity rates are also similar across genders (Silberschmidt, 2015; Efe, 2022a).

In summary, conditions with similar prevalence across genders in the general population generally show comparable gender balance in individuals with OCD, though findings for bipolar and hoarding disorders are inconsistent. Emerging evidence suggests that gender differences may become more pronounced in some comorbid conditions, with women showing greater burden in bipolar, hoarding, and body dysmorphic disorders in some studies, although findings are not consistent across the literature.

#### 3.4.3. Psychiatric Conditions with Higher Prevalence in Men in the General Population

*3.4.3.1. Schizophrenia* is more common in men in the general population (Ochoa et al., 2012). Seventeen studies during the review period examined gender differences in the co-occurrence or association of OCD/OCS and schizophrenia. A history of OCD significantly increases the risk of developing schizophrenia (HR = 30.2, p < 0.001), with men at greater risk than women (F:M HR = 0.63, p < 0.01; Cheng, 2019). Among individuals with OCD, adult men show a higher prevalence of comorbid schizophrenia and schizotypal traits (F:M OR = 0.09–0.67, p < 0.05; Huang, 2011; Huang, 2014), mirroring general population trends, while one long-term follow-up study found no significant gender difference in schizophrenia-related diagnoses (Klenfeldt, 2024). However, in children, schizophrenia prevalence is similar across genders (F:M OR = 1.25, p > 0.20; Huang, 2014). One study found an association between polygenic risk scores for schizophrenia that was specifically associated with the OCD Perfectionism symptom subset in female adolescents (Aicoboaie, 2025). In adults with schizophrenia, findings on gender differences in OCD comorbidity are mixed. Two studies reported a higher prevalence in men (F:M OR = 0.22–0.49; Devi, 2015; Baek, 2020), while Temircan (2021) also reported a male-directional estimate but was classified as showing no significant gender difference in the appendix. Five found roughly equal prevalence (F:M OR = 0.73–1.23; Faragian, 2012; Bener, 2018; Kokurcan, 2020; Shoaib, 2023; Jelsma, 2025), and one noted a trend toward female bias (F:M OR = 1.23 [95% CI, 0.94–1.60]; Bosanac, 2013). Taha et al. (2024) additionally found no significant sex difference in obsessive–compulsive symptom severity among adults with schizophrenia (M = 19.8 ± 8.7, F = 18.3 ± 9.4; d = 0.17, p = 0.29). Furthermore, while clozapine, a unique antipsychotic drug used to treat refractory schizophrenia, is associated with increased risk of developing comorbid OCD compared to haloperidol, this risk was significant for both sexes (M OR = 3.07, p = 0.003; F OR = 2.46, p = 0.047; Park, 2021). In a large all-ages OCD cohort, males were more likely to subsequently receive a schizophrenia diagnosis (HR = 1.23 [95% CI, 1.12–1.35]; Chen, 2023).

*3.4.3.2. Psychotic symptoms* may appear outside of a formal diagnosis of schizophrenia. In the general population, rates of psychosis are higher in men than in women (Castillejos et al., 2018). Nine studies during the review period examined gender differences in psychotic disorders or symptoms in relation to OCD. In children and young adults with OCD, male patients have been reported to be more likely to have a comorbid psychotic disorder in one study (F:M OR = 0.71, p < 0.01; Rintala, 2017), while others suggest approximately equal vulnerability to psychosis among the sexes (OR = 1.10–1.15, p > 0.10; Borelli, 2023; Mohammadi, 2021; Borelli, 2025). Among adults with OCD, psychosis was found to be similarly present in men and women (F:M OR = 1.03; Benatti, 2020; Shaker, 2025), and clinical high-risk psychosis patients of both sexes showed equal prevalence of comorbid OCD (West, 2025). Sullivan et al. (2018) examined psychotic patients and found that men were more likely to exhibit OCD-like symptoms than women, with stronger differences in younger and older age groups (age < 44 F:M OR = 0.36– 0.57; age > 55 F:M OR = 0.49). On the other hand, Hagen et al. (2013) examined young patients with first-episode psychosis (age = 23 ± 8) and found a non-significant trend toward female bias (F:M OR = 1.93, p = 0.11).

*3.4.3.3. Autism spectrum disorder (ASD)* is nearly four times more common in men than women in the general population (Giarelli et al., 2010; Loomes et al., 2017; Nordahl, 2023), though diagnostic practices and tools may underestimate autism in women (Brugha et al., 2016). Thirteen studies during the review period examined gender differences in the co-occurrence of OCD/OCS and ASD. Among children with OCD, most studies find that ASD symptoms and diagnosis are more common in boys (F:M OR = 0.2–0.4, p < 0.01; Arildskov, 2016; Martin, 2020; Rintala, 2017; Zijlmans, 2017; Efe, 2022; Ozcan, 2025), although two studies found similar ASD prevalence in boys and girls with OCD (Peris, 2017; Huang, 2014). Boys with OCD and ASD show greater symptom severity for both conditions (Arildskov, 2016). Among adults, a smaller study suggested only a non-significant trend toward higher ASD prevalence in men with OCD (F:M OR = 0.6, p = 0.25; Mito, 2014).

Research on OCD prevalence in individuals with ASD reports inconsistent patterns. For example, Margari et al. (2019) found a non-significant trend toward higher OCD prevalence in boys with ASD (F:M OR = 0.14, p = 0.08, N = 164), attributing the underdiagnosis in girls to socially acceptable restrictive interests and behaviors, while another found comorbid OCD to be more common in boys with a primary diagnosis of ASD (O’Connor, 2025). Conversely, Martin et al. (2020) reported a higher prevalence of OCD among girls with ASD in a UK pediatric sample (F:M OR = 1.7, p < 0.001, N = 7,922), and Rødgaard (2021) reported a higher proportion of girls with OCD in a Danish ASD cohort (F:M OR = 1.39, p < 0.001). Similarly, a latent class analysis of comorbidity patterns in patients with OCD identified a specific OCD–ASD cluster that was more prevalent in females (Smárason, 2024).

*3.4.3.4. Tic disorders (TD)*, including *Tourette syndrome (TS)*, are up to four times more common in boys than girls in the general population (Gagnon et al., 2024; Knight et al., 2012). Twenty-six studies during the review period examined gender differences in the co-occurrence or association of OCD/OCS and TD/TS. Eleven studies similarly found TD comorbidity to be significantly more common in males, both in youth (F:M OR = 0.10–0.55, p < 0.01; Nikolajsen, 2011; Vivan, 2014; Højgaard, 2016; Arildskov, 2016; Rintala, 2017; Efe, 2022a; Hakimi, 2022) and adults (F:M OR = 0.2–0.69, p < 0.05; Torresan, 2013; Vries, 2016; Rintala, 2017; Ekinci, 2020; Brander, 2021 [all ages]). However, other studies reported similar prevalence across sexes in children with OCD (F:M OR = 0.51–1.09, p ≥ 0.17; Peris, 2017; Tanidir, 2015; Yan, 2022) and in adults (F:M OR = 0.82–1.47; Pinto, 2016; Benatti, 2020; Mukai, 2021). Three studies likewise reported comparable OCD prevalence between sexes among adolescents with tic disorders (OR = 0.86–1.06; Greenberg, 2018; Girgis, 2022; Chang, 2026). Tourette syndrome was more common in men with OCD only at a trend level (F:M OR = 0.72, p = 0.19; Torresan, 2013), and the presence of comorbid ADHD further increased the male predominance (Pinto, 2016). In patients with TDs, comorbid OCD was reported to be more common in women than in men (Pinto, 2016; Dy-Hollins, 2024; Nilles, 2025). Rodgers et al. (2013) identified a subgroup of TD patients with elevated OCD symptoms; this class only included women, but their small sample size (N = 80) limited conclusions. Another study found that non–tic-related impairment in adolescents with tic disorders predicted greater OCD severity in females (Larsh, 2023), and that compulsions predicted tic onset specifically in girls (Openneer, 2022). Interestingly, one study that reported an increased male bias of OCD in TS also found that the M:F ratio among patients with TS and OCD was substantially lower than among those with TS without OCD (2– 3:1 vs. 5–6:1; Śmilowska, 2026).

*3.4.3.5. Externalizing disorders* such as attention deficit and hyperactivity disorder (ADHD), conduct disorder (CD), and oppositional defiant disorder (ODD) are more prevalent in boys in the general population (Bauermeister et al., 2007; Maughan et al., 2004; Waschbusch & Willoughby, 2008). Nineteen studies during the review period examined gender differences in externalizing disorders in relation to OCD/OCS. Similarly, among children with OCD, most studies report a male bias in prevalence of CD (F:M OR = 0.7, p = 0.04; Rintala, 2017; Efe, 2022a), ODD (F:M OR = 0.3, p = 0.036; Tanidir, 2015), or any externalizing disorder (F:M OR = 0.24–0.60, p < 0.08; Peris, 2017; Zijlmans, 2017; Ortiz, 2016), although Arildskov (2016) found no gender difference in any externalizing disorder and Efe (2022a) found no gender difference in ODD. One study, which pooled several neurodevelopmental disorders including ADHD, ASD, tic disorders, and learning disorders, also found a significantly higher male prevalence in an OCD population (D’Aiello, 2025). Several studies report a significant male bias in ADHD comorbidity among patients with OCD (F:M OR = 0.04–0.43, p < 0.01; Rintala, 2017; Arildskov, 2016; Efe, 2022a; Efe, 2022b; Hakimi, 2022), whereas others observe no significant gender difference (F:M OR = 0.50–0.70; Tanidir, 2015; Vivan, 2014; Yan, 2022). Using latent class analysis, Zijlmans et al. (2017) identified a subgroup of children with elevated OCD symptoms characterized by high rates of ADHD and ODD comorbidity; this subgroup was predominantly male (F:M OR = 0.2, p < 0.005). Interestingly, studies examining ADHD cohorts suggest the reverse pattern: OCD appears to be more prevalent among girls with ADHD (F:M OR = 1.7, p = 0.04; Inci, 2019), although obsessive–compulsive symptom severity may be greater in boys (d = 0.24, p = 0.03; De Rossi, 2022), and another study found similar OCD prevalence in boys and girls with ADHD (Zhang, 2026).

Fewer studies have examined these conditions in adults, but those that have shown similar patterns. ADHD is more common in men than in women (Faheem et al., 2022). In adult OCD patients, comorbid ADHD either appears similarly across genders (F:M OR = 0.9, p = 0.96; Blanco-Viera, 2018) or shows a male bias (F:M OR = 0.43, p < 0.01; Pinto, 2016; Ozaydin, 2025). In adult ADHD patients, OCD is more common in men at a trend level (F:M OR = 0.70; Pinto, 2016).

*3.4.3.5. Substance use disorders (SUDs)* are more common in men in the general population, but women with SUDs often present with a more vulnerable clinical profile (Fonseca et al., 2021; Higgins et al., 2015; Lee, 2024). Fifteen studies during the review period examined gender differences in substance use or SUDs in relation to OCD/OCS. OCD increases SUD risk, with a stronger effect observed in women (OR = 7.52 [95% CI, 3.11–18.18]) than in men (OR = 3.32 [95% CI, 1.67–6.61]; Blom, 2011). However, gender differences in SUD prevalence among adults with OCD are mixed: several studies found comparable prevalence (Al-Asadi, 2015; Benatti, 2020; Klenfeldt, 2024), whereas a large registry study found a male bias (Virtanen, 2022). Among youth, alcohol/tobacco use and AUD were similarly prevalent across genders (Mohammadi, 2021; Hansen, 2025).

Differences may exist in specific substance use patterns. Cigarette smoking, while overall less prevalent in OCD patients than in the general population (Abramovitch et al., 2015; Abramovitch et al., 2014), showed no gender difference for current smoking, while former smoking showed a female bias (Dell’Osso, 2015). Conversely, alcohol use disorder (AUD) is more common in men with OCD than in women (F:M OR = 0.31–0.35, p < 0.01; Torresan, 2013; Al-Asadi, 2015; Klenfeldt, 2024), with one exception in a study of Finnish OCD patients where prevalence of AUD in OCD patients was equal between the sexes (p = 0.14; Samuels, 2026). Unexpectedly, one study in OCD patients with alcohol and drug use indicated that alcohol use predicted lower subsequent OCD symptom severity, and greater OCD severity predicted lower subsequent drug use, only in females (du Mortier, 2025). Cannabis use was similarly prevalent across genders in two studies (Benatti, 2023; Jacobs, 2025), and alcohol and tobacco use were also similar across genders in Jacobs (2025). In contrast, the large Virtanen (2022) study identified substantial substance-specific gender differences, including a male bias in overall substance misuse and a female bias in sedative-related disorders.

Among adults with SUD or AUD, OCD prevalence is comparable between genders (F:M OR = 0.93–0.96; Blom, 2011; Al-Asadi, 2015). However, in adolescents with SUD, a female bias in OCD prevalence has been reported. For instance, adolescents with AUD show a significantly higher OCD prevalence in girls in one study (F:M OR = 2.7, p < 0.001; Harford, 2015) and a trend in the same direction in another (F:M OR = 1.8, p = 0.16; Fernández-Artamendi, 2021). Another study in Danish adolescents indicated a similar prevalence of AUD among boys and girls with OCD (p = 0.14; Hansen, 2025), indicating a need for further inquiry into this area to determine if there is truly gender bias. Similarly, cannabis use disorder in adolescents has been linked to higher OCD prevalence in girls (F:M = 6.9, p < 0.01; Fernández-Artamendi, 2021).

*3.4.3.6. Impulse control disorders (ICDs)* are more common in boys and men in the general population (Leclercq & Corvol, 2024). Seven studies during the review period examined gender differences in ICDs or related behavioral addictions in relation to OCD/OCS. In children with OCD, prevalence of any impulse control disorder is similar in boys and girls (F:M OR = 0.77, p > 0.20; Vivan, 2014). Among adults with OCD, one study reported higher lifetime ICD prevalence — including compulsive sexual behavior disorder (CSBD), pyromania, kleptomania, and intermittent explosive disorder (IED)—in men (F:M OR = 0.26–0.57; Fuss, 2019), while current ICD prevalence was similar between genders and current kleptomania showed a female-directional trend. In contrast, another study found a higher prevalence of ICDs in women (F:M OR = 1.34, p < 0.05), though the prevalence of specific ICDs did not significantly differ between genders (Torresan, 2013).

Pathological gambling in adults with OCD is predominantly observed in men (F:M OR = 0.05–0.08; Al-Asadi, 2015; Torresan, 2013). However, among individuals with pathological gambling, OCD prevalence is similar across genders (Al-Asadi, 2015).

Internet addiction is more common in men in the general population (J Kuss et al., 2014). OCD appears to increase the risk of internet addiction similarly in both sexes (men: d = 0.66, p < 0.001; women: d = 0.93, p < 0.001; Potembska, 2019). In addition, women seeking treatment for problematic internet use were more likely to have comorbid OCD (p = 0.016; Machado, 2022). Addiction to video games is more prevalent in men with OCD than in women (F:M OR = 0.16, p < 0.001; Torresan, 2013), reflecting general population trends. Additionally, among men in addiction treatment centers, internet addiction severity is positively correlated with OCD symptom severity (d = 0.62, p < 0.001; Wölfling, 2013).

Overall, psychiatric conditions more prevalent in men in the general population, such as schizophrenia, psychotic disorders, ASD, externalizing disorders, and impulse control disorders, also tend to show a male bias among OCD patients. However, certain conditions, such as SUD and some substance-use outcomes, may show a stronger relative association with OCD in women, despite male-biased or null prevalence findings for several specific substances. Results related to comorbid tic disorders and Tourette syndrome remain inconsistent and warrant more research.

#### 3.4.5. Prevalence of OCD in individuals with other primary diagnoses

OCD is more common in women than in men overall, but early-onset OCD diagnosis is more common in men (see Section 3.3.1). However, as Table 3 illustrates, in individuals with other psychopathologies as the primary condition, these overall prevalence patterns may be altered. For instance, in children with ADHD, ASD, AUD, cannabis use disorder, and tic disorders, girls are more likely to have an OCD diagnosis than boys in at least some studies. On the other hand, in adults with diagnoses of depression, hoarding disorder, schizophrenia, or ADHD, men are more likely to have comorbid OCD in some studies. Additionally, daily migraine is linked to OCD more strongly in men than in women. Other conditions show either no gender difference or inconsistent patterns across studies. Why these psychopathologies are associated with OCD differently in men and women remains unclear.

#### 3.4.6. OCD and Physical Health

Psychiatric disorders, including OCD, frequently co-occur with general medical conditions (De Hert, Cohen, et al., 2011; De Hert, Correll, et al., 2011; De Hert et al., 2009; Scott et al., 2007). During the review period, 43 studies (9 pediatric, 32 adult, and 2 all-ages/pooled samples) examined whether the prevalence or clinical expression of these medical conditions differed by sex among individuals with OCD (Table 3, Table A4.2). Seventeen studies focused on sex differences in physical health conditions associated with OCD, while 26 examined sex differences in OCD prevalence or symptom severity in the context of medical conditions. Characterizing these co-occurrences clarifies the added medical burden associated with OCD and may identify shared or interacting mechanisms underlying their co-expression. Across studies, women with OCD were more likely than men to report general medical conditions (e.g., Aguglia, 2017; Holmberg, 2024), although sex differences varied by condition. Some patterns mirrored sex differences observed in the general population, whereas others diverged.

*3.4.6.1. Autoimmune diseases* are substantially more prevalent in women than men in the general population: F:M OR ≈2–4:1 (Fairweather et al., 2008). In OCD, however, this female predominance was attenuated and varied across conditions. Rheumatic fever was only modestly more common in women than men with OCD before correction for multiple comparisons (Westwell-Roper, 2019), whereas Sjögren’s syndrome occurred at similarly elevated rates in men and women with OCD relative to the general population (Chang, 2021a). In pediatric OCD, IgA deficiency showed a male-specific association, with elevated odds in boys but not girls (Williams et al., 2019). In contrast, a clinical OCD sample found autoimmune conditions substantially more common in women than men (10.5% vs. 1.1%; Holmberg, 2024). Population-based and clinical studies of systemic autoimmune disease and juvenile idiopathic arthritis reported no statistically significant sex differences in OCD risk or prevalence (Wang, 2019; Li, 2022). While mast cell activation syndrome overall increased OCS severity, these findings did not differ between males and females (Weinstock, 2025). Together, these findings suggest that autoimmune–OCD associations are condition-specific and may involve sex-dependent mechanisms.

*3.4.6.2. Vascular and cardiometabolic conditions.* Risk for vascular disorders was similarly elevated in men and women with OCD relative to the general population (HR ≈ 1.37; Isomura, 2016). Consistent with this, Holmberg et al. (2024) found no significant sex difference in vascular or cardiovascular conditions among adults with OCD. In contrast, risk for vascular dementia was elevated only in women with OCD (HRF = 4.76 [95% CI, 1.54–14.74]; Chen, 2021b). Cardiometabolic conditions and hypertension were approximately 1.6 and 2.6 times, respectively, more common in men than women with OCD (Holmberg, 2024), whereas elevated heart rate was similarly associated with OCD in both sexes (Jüres, 2024). In men, elevated resting heart rate and blood pressure at age 18 predicted later OCD diagnosis (Latvala et al., 2016); whether this reflects enduring physiological vulnerability or situational anxiety remains unclear, and comparable longitudinal data in women are lacking.

*3.4.6.3. Neurological conditions.* Across neurological conditions, sex differences in OCD prevalence and expression often diverged from general-population patterns, in which OCD is typically more prevalent in women (see Section 3.3.1). In adults with temporal lobe or idiopathic epilepsy, OCD prevalence was similar in men and women (Kilicaslan, 2020; Hamed, 2013). In contrast, OCD prevalence was elevated and male-predominant in migraine, particularly in daily migraine, with substantially higher predictive positive values in men than women (PPV\(_M\) = 29.9 vs. PPV\(_F\) = 3.2; Goulart, 2014; Viticchi, 2022), despite the fact that the female predominance of migraine among individuals with OCD mirrored general-population patterns (Holmberg, 2024). In Parkinson’s disease, obsessive–compulsive symptom severity was similarly elevated in men and women (Saadat, 2020). In Down syndrome, OCD comorbidity was similar between male and female patients (Grotteschi, 2025). OCD severity was also similar between male and female fibromyalgia patients (Karahan, 2025). Together, these departures from the usual female predominance in adult OCD suggest condition-specific patterns that may reflect differences in the clinical or neurobiological context of neurological illness. Consistent with this interpretation, OCD symptoms decreased following effective migraine treatment in both men and women (Viticchi, 2022). Notably, however, a large population-based study reported a markedly elevated risk of Alzheimer’s disease exclusively in men with OCD (Chen, 2021b), underscoring the gender- and condition-specific nature of OCD–neurological disease associations.

*3.4.6.4. Cancer.* Among actively treated pediatric cancer patients, elevated OCs were highly prevalent and comparable across sexes (boys: 20.8%; girls: 21.1%), ranking among the most common psychiatric symptoms during treatment (Zahed, 2020). In contrast, a large registry-based study of childhood cancer survivors assessed in adolescence and young adulthood reported a low prevalence of diagnosed OCD (0.34%), with a higher proportion of cases in females than males (female vs. male RR = 1.39; DeWitt, 2025), consistent with the postpubertal female predominance of OCD in the general population rather than evidence of a cancer-specific reversal in gender distribution. Similarly, among older adult cancer patients, women reported greater OCD symptom severity than men (İnci, 2024). Together, these findings suggest that OC symptoms may be relatively common during active pediatric cancer treatment without a sex difference, whereas diagnosed OCD in survivorship and OC symptom severity in older adults show a female bias.

*3.4.6.5. Other conditions.* Several conditions were examined in only one or two studies, limiting the strength of inferences. Across these conditions, gender effects varied considerably. Sleep problems generally showed no sex differences in children with OCD (Ivarsson, 2015; Nabinger de Diaz, 2019), whereas elimination disorders were more common in boys with OCD, and bladder symptoms have also been reported in a female-only clinical sample (Mohammadi, 2021; Westwell-Roper, 2022; Rezaeimehr, 2022). In contrast, OCD comorbidity showed no female bias in cohorts with restless legs syndrome, cerebral palsy, and juvenile idiopathic arthritis, diverging from the female predominance observed in adult OCD in general (Ghorayeb, 2019; Mazurie, 2024; Li, 2022; Bhatnagar, 2025; Kang, 2025). OC symptom severity was more strongly associated with misophonia severity in women than in men (Herdi, 2024). Associations between OCD/OCS and weight or obesity showed no clear sex difference (Isomura, 2016; Husky, 2018; Ma, 2026). Stuttering severity, which occurs at a ratio of approximately 2:1 in boys vs. girls (Briley et al., 2022; Yairi et al., 1996) was positively associated with OCS severity in a male-only sample of adolescents with stuttering (Taherifard, 2025).

Overall, associations between OCD and physical health conditions showed substantial heterogeneity in their sex-specific patterns. Some preserved the gender distribution expected in the general population, whereas others showed attenuated, absent, or occasionally reversed differences. These findings suggest that sex differences in OCD–medical comorbidity are condition-specific rather than reflecting a uniform effect of physical illness on OCD vulnerability.

#### 3.4.7. Summary of Key Considerations

Methodological Concerns:

- Comorbidity measurements from self-report (or parent report), clinical assessments, and medical records are unlikely to align.
- Most studies are cross-sectional; more attention needs to be given to developmental trajectories. For instance, while autism and ADHD comorbidities show a strong male bias in many pediatric OCD samples, this pattern is less consistent in adults, suggesting that sex differences in diagnosis or clinical recognition may change across development.
- Only a few studies focused on the specific symptoms or other characteristics of comorbid conditions, and how these might interact with OCD.
- Many physical health studies relied on registry, claims, or diagnostic-record data, which may under-ascertain OCD and provide limited information about symptom severity, clinical presentation, or sex differences in diagnostic thresholds.

Unanswered Questions:

- What drives differences in gender distribution in co-occurring conditions in the presence of OCD, compared to the same conditions absent OCD comorbidity (e.g., stronger relative associations with some substance-use outcomes in women or higher prevalence of social anxiety disorder in men)?
- When OCD co-occurs with medical conditions, under what circumstances are typical sex differences preserved, attenuated, or reversed, and what does this imply about shared vulnerability versus disorder-specific mechanisms?
- Do OCD and comorbidities interact to impact symptom severity and dimensions differently in males and females?
- Are there differences in the temporal relationship between OCD and comorbidities across genders? Does OCD differentially predict subsequent comorbidity in males and females, do other conditions differentially predict subsequent OCD, or do gender differences arise from shared underlying risk factors, such as neuroticism or stress? How can these differences be leveraged in treatment selection?

Hypotheses for Future Research:

- OCD disproportionately increases risk for social anxiety disorder in men and for some substance-use problems (excluding alcohol) in women.
- OCD comorbid with neurodevelopmental disorders (e.g., tic disorders, ADHD, ASD) is phenomenologically different across gender and age groups.
- OCD comorbid with trauma- and stress-related conditions, including anxiety, depression, and PTSD, is phenomenologically different across gender and age groups.
- When OCD co-occurs with general medical conditions, obsessive–compulsive symptoms are more likely to reflect stress-related or reactive symptom expression that attenuates as the underlying medical condition improves, with weaker sex differences than those observed in primary OCD.

### 3.5 Human Genetics and Family Risk

#### Highlights from Section 3.5 Human Genetics and Family Risk

<u>State of knowledge and key concerns:</u> Few genome-wide molecular studies have examined associations between genetic or molecular features and OCD risk across sexes. Family-based designs (e.g. family, twin and parental age studies) may provide evidence of potential sex differences, but generally cannot identify the specific genetic or molecular factors underlying these differences.

<u>Genome-wide molecular studies (only eight in over 15 years</u>): Findings suggest that some genetic and molecular factors may differentially influence OCD in men and women, although evidence remains limited.

<u>Family-based designs</u>: Family and twin studies generally suggest similar transmission of OCD risk across sex of proband and relatives, though there may be subtle differences based on specific symptom dimensions and subtypes. Findings regarding the roles of paternal versus maternal age are inconsistent. Studies also suggest higher rates of autoimmune conditions in biological mothers of children with OCD than among women in the general population.

<u>Future studies:</u> More and larger genome-wide molecular studies of risk for OCD are needed, with analyses conducted separately in males and females. Studies should also examine whether genetic contributions differ between child-onset and adult-onset OCD. Genome-wide molecular analyses of different OCD subtypes (based on OCS, comorbidity profile, or stress-relatedness) should be performed in males versus females.

Human genetics research holds great promise for better understanding gender differences observed in epidemiological and clinical studies of OCD, by identifying specific underlying genetic causes that can be further probed in mechanistic research. During the review period, 101 human genetic studies were identified examining sex^4^ and gender differences in OCD (see Table A5); these include family, twin, candidate gene, genome-wide, and parental age studies, among others.

#### 3.5.1 Family studies

Decades of studies have demonstrated the familial transmission of OCD. During the review period, 16 registry and clinical studies examined whether familial risk varies by sex or gender. Twelve studies specifically examined sex- or gender-related patterns in familial transmission of OCD risk. Steinhausen et al. (2013) examined 2,057 pediatric onset OCD cases, first-degree relatives, and 6,055 matched controls, showing that both paternal and maternal OCD were risk factors for OCD in the probands (father OR = 34.04, p < 0.01; mother OR = 11.21 p < 0.002). Mataix-Cols et al. (2013) examined 24,768 individuals with OCD, relatives, and matched controls, finding no differences in the familial effects by gender of the probands or their relatives. Kendler et al. (2023) examined 2,413,128 offspring from 4 different family types (intact, not-lived-with biological fathers, lived-with stepfathers, and adoptive) to estimate sources of parent-offspring resemblance. For OCD, the best-estimate correlation for genes plus rearing was 0.19 (95% CI 0.17-0.21) for mothers and 0.19 (95% CI 0.17-0.22) for fathers. The correlation attributable to genetic effects only was 0.26 (95% CI 0.08-0.44) for mothers and 0.17 (95% CI 0.10-0.24) for fathers. The rearing-only correlation for fathers with 0.04 (−0.10-0.19), with insufficient data to estimate this effect for mothers. This suggests similar maternal and paternal transmission of OCD risk.

Two studies have further examined whether parental psychiatric risk differs according to the sex of the offspring. Zhou et al. (2023) found that parental general and specific psychopathology factors showed similar associations with OCD in both male and female offspring in a sample of 2,947,703 individuals. Similarly, Hsu et al. (2024) reported comparable elevations in parental mental disorders among male and female probands with OCD in a cohort of 69,819 patients with schizophrenia, OCD, and both conditions. Together, these studies indicate that parental psychiatric risk does not differ meaningfully by offspring sex in OCD.

Two studies examined whether familial risk differed by OCD subtype and sex. Brander et al. (2021) compared 1257 individuals with tic-related OCD and 20,975 with non-tic related OCD and showed that overall the risk of OCD in relatives of individuals with tic-related OCD was greater than risk of OCD in relatives with non-tic-related OCD (HR in full siblings 10.63 [7.92 – 14.27], vs 4.52. [4.06 – 5.02], p<.0001). Given that tic-related OCD is more common in males, they also performed gender stratified analyses looking at all combinations of siblings; the hazard ratio was higher for tic-related OCD across all combinations, suggesting greater familiality. Interestingly, the risk of OCD was also higher in male:male pairs than other combinations in relatives of non-tic related OCD compared to controls. Balachander et al. (2021) examined 330 individuals with OCD from 153 families to assess familial patterns of symptom dimensions and found that sex-concordant pairs showed greater concordance irrespective of relationship type.

Four studies examined sex- or parent-specific familial associations involving co-occurring medical conditions, including maternal autoimmune disease and other prenatal exposures. Murphy et al. (2010) examined the biological mothers of 107 children with OCD and tics, finding that 17.8% had autoimmune conditions compared to 5% in the general population. In a large Danish population-based registry study, He et al. (2022) found that maternal autoimmune disease diagnosed before or during pregnancy was associated with increased OCD risk (HR, 1.42; 95% CI, 1.24-1.63). Similarly, Zhang et al. (2023) found that prenatal exposure to maternal (but not paternal) infections was linked to higher OCD risk in a Swedish registry study, although this association was not significant in sibling analyses. Jo et al. (2026) found that prenatal exposure to pregestational and gestational diabetes was not associated with risk of OCD in offspring in a South Korean registry study.

Finally, five studies examined OCD risk among relatives of individuals with other medical and psychiatric conditions. Lin et al. (2022) compared 40,462 first-degree relatives of individuals with lupus erythematosus (SLE) with 80,536 matched controls and found that female first-degree relatives had higher rates of OCD than males (female RR 1.46, 95% CI 1.17-1.82; male RR 1.00, 95% 0.48-2.07). This difference may suggest sex-specific shared mechanisms or that female relatives are more sensitive to the stress of having a family member with SLE. Pol-Fuster at al. (2025) examined risk of OCD in relatives of individuals with severe infections (1,811,364) compared to those without (3,105,534), showing elevated rates of OCD in mothers compared to fathers, maternal half-siblings compared to paternal half-siblings, and aunts compared to uncles, suggesting possible maternal or sex specific effects. Li et al. (2026) compared 20,362 individuals with bipolar disorder to 203,620 matched controls, showing elevated rates of OCD in both mothers and fathers compared to controls. The risk of OCD was higher in fathers of offspring with mania-onset bipolar disorder than depressive or unspecified bipolar disorder, and the risk of OCD was higher in mothers of offspring with depressive-onset bipolar disorder than the other two groups, suggesting potentially different maternal and paternal patterns of familial risk. Hsu et al. (2026) examined 153,091 first degree relatives of individuals with panic disorder and 612,364 matched controls, finding a higher rate of OCD in females (adjusted RR 1.95, 95% CI 1.70-2.23) compared to males (adjusted RR 1.56, 95% CI 1.37-1.77). Sackl-Pammer et al. (2015) conducted a small study of 30 parents of children with separation anxiety disorder and found higher rates of lifetime OCD among fathers, but not mothers, compared to controls.

#### 3.5.2 Twin studies

Twin studies are a powerful approach for understanding the distinct roles of genetic and environmental factors in the development of OCD. Of the seven twin studies that examined sex differences, five (Bolhuis, 2014, Pinto, 2016, Lopez-Sola, 2014, Zohao, 2014, Krebs, 2015) reported no sex differences in the role of genetic factors in obsessive-compulsive symptoms and behaviors, consistent with the family studies reviewed above. One study by Moore et al. (2010) examined 517 adolescent twins and found that both obsessions/incompleteness and numbers/luck dimensions were significantly more heritable in males only (60% and 65%, respectively). Anckarsater et al. (2011) examined 17,220 same-sex twins and identified some potential differences in the correlation of compulsiveness (identical F = 0.22, identical M = 0.36, fraternal F = 0.16, fraternal M = 0.09). Thus, there may be some subtle differences in heritability between sexes when specific symptom dimensions are considered.

#### 3.5.3 Candidate gene studies

Candidate gene studies focus on examining a particular genetic variant (or set of variants) that is hypothesized to be associated with OCD. These studies are limited by the polygenic nature of OCD and the fact that common variants tend to have small effect sizes. Most candidate gene studies in psychiatry have not been replicated in large-scale genome-wide studies (Border et al., 2019; Farrell et al., 2015), so these results must be interpreted with skepticism. In fact, the National Institute of Mental Health recommends that researchers abandon candidate gene studies in favor of well powered hypothesis-free genome association studies (Workgroup). Despite this guidance, our review found that the vast majority of genetic studies during the review period (45 out of 101) examining sex used a candidate gene approach. These studies examined a variety of candidate genes (including *ESR1 ESR2, 5-HTTLPR, COMT, GRIN2B, SLC1A1, SLC6A4, SLC18A1, GAD1, GAD2, HTR1A, HT2A, HTR1B, HTR3B, DRD2, DRD4, DAT, NPSR1, MAO-A, DLGAPI, EFNA5, MTHFR, SAPAP3, SLITRK1, PBX1, LMX1A, RYR3, OXTR, FKBP5*). For completeness, the findings are summarized in Table A5. However, to reiterate, such studies have a poor track record of replication.

#### 3.5.4 Genome-wide molecular studies

Genome-wide molecular studies examine associations between molecular features across the genome and OCD risk. These include genome-wide association studies (GWAS), copy number variant (CNV) analyses, DNA sequencing, methylation-wide association studies, and RNA sequencing. Only eight genome-wide molecular studies examining sex or gender impacts were identified during the review period, highlighting a need for more research in this area.

Most studies focused on common genetic variation. Khramtsova et al. (2018) conducted a GWAS meta-analysis of 9,870 individuals with OCD and found a strong genetic correlation between male and female OCD, with similar heritability estimates. No sex-specific, genome-wide significant single nucleotide polymorphisms were identified, but there were significant gene-based associations that emerged only in females in two genes (GRID2 female p = 1.07E-07, male p = 7.23E-01; GRP135 female p = 1.55E-06, male p = 7.04E-01). Oh et al. (2022) used GWAS summary data to assess genetic overlap between reproductive behaviors and OCD, they reported significant positive genetic correlations between OCD and age at first sexual intercourse in both males and females (p<0.005), and between age at first birth in females only (p<0.005). Kutzner et al. (2022) examined polygenic scores for OCS in 547 adolescents and found a negative association with cannabis use at age 17, a finding that replicated among males of European ancestry but did not extend to other groups in sensitivity analyses. Aicoboaie et al. (2025) examined 4,729 individuals of European ancestry in the Philadelphia Neurodevelopmental cohort and did not find associations between OCD polygenic scores and mental health phenotypes, neurocognitive measures or cortical phenotypes in males or females.

Two studies examined copy-number variants. Lionel et al. (2014) examined CNVs in 64,114 individuals with neurodevelopmental disorders, including OCD. They found that CNV disruption of the ASTN2/TRIM32 locus was significantly associated with neurodevelopmental disorders in males, but not females, with OCD among the co-occurring disorders. Halvorsen et al. (2025) examined rare CNVs in 2,248 OCD cases and 3,608 unaffected controls from Sweden and Norway. They found no detectable sex difference in CNV burden, although effect size estimates were generally larger among males.

In a methylome-wide association study of OCD including 414 affected adults and 384 controls, Hoffler et al. (2026) identified sex specific differentially methylated positions and regions. Most male-specific differentially methylated regions showed significant sex-by-diagnosis interactions, supporting sex-specific methylation associations. Sex-specific analyses also identified genes previously associated with OCD or related phenotypes, including *AKAP12* and *PIWIL1*. In a small RNA sequencing study, Sakurai et al. (2025) compared 14 patients with OCD and 14 patients with co-occurring OCD and neurodevelopmental disorders. They identified several sex-specific differentially expressed genes, including the cytokines *IL17A* and *IL11*, suggesting a possible sex-specific shift towards inflammatory processes in the co-occurring group.

Overall, these findings suggest a potential role for genetic and molecular factors that may differentially influence OCD in males and females, offering insight into mechanisms underlying observed sex and gender differences in its presentation.

#### 3.5.5 Parental age studies

Past research suggests that parental age at a child’s birth impacts risk for a range of neuropsychiatric disorders in offspring (de Kluiver et al., 2017). During the review period, six studies examined the association between parental age and risk of OCD, yielding different findings regarding the role of maternal and paternal age at birth. Wu et al. (2012) conducted a cohort study of 711 adults in China and found that OCD risk increased with paternal age, but not with maternal age, and was higher in male offspring. Biria et al. (2019) conducted a small cross-sectional study of 85 individuals with SCZ treated with clozapine and found that younger parental age was associated with OCD risk. In contrast, Chudal et al. (2017) conducted a nested case-control study of 1,358 individuals with OCD, 1195 individuals with chronic tic disorders, and matched controls and found that the risk of OCD was statistically increased in offspring of older mothers and not fathers.

Janecka et al. (2019) conducted a large birth cohort study in Denmark (n=1,490,745) and found that the risk of OCD was statistically increased only in the offspring of older mothers; but gender specific analyses suggested that paternal age effects were also present, in the male offspring only. Consistent with this, Yilmaz also conducted a large registry study in Denmark (n=1,154,067) and found that older maternal age was associated with risk of OCD in male offspring, but not female offspring. Although paternal age was not associated with OCD risk in the overall sample, older paternal age was associated with increased risk in male offspring, though the sex difference was nonsignificant. Finally, Wang et al. (2022) examined a Taiwan registry sample (n=7,264,788) and found that both older paternal and maternal age were associated with increased risk of OCD in offspring.

Overall, this work suggests associations between parental age and OCD risk, but maternal and paternal effects vary across studies and may be more pronounced in male offspring.

#### 3.5.6 Other studies

Six additional studies examined other sex- or gender-related familial, molecular, and cellular risk factors for OCD. Two studies focused on family-related risk factors for OCD. Zhang et al. (2022) examined 3438 middle school students in China and found that both paternal and maternal warmth was associated with lower OCD scores, whereas parental punitive behaviors, father overprotection, and mother over-involvement were significantly associated with higher OCD scores. In a large Swedish registry study, Salvatore et al. (2024) found that divorced females had higher familial genetic risk scores for OCD than males.

Schneider et al. (2016) reported that female carriers of a premutation in the gene FMR1, who carry an expansion in the X-chromosome gene implicated in Fragile X syndrome that is not sufficient to produce the full syndrome, had higher OCD symptom burden compared to controls, whereas male carriers did not. Similarly, Loesch et al. (2021) found that female FMR1 premutation carriers had greater symptom progression than male carriers.

Kang et al. (2021) examined cellular aging markers in 235 patients with OCD and 234 controls. Post-hoc analyses revealed that both males and females with OCD had significantly lower mitochondrial DNA copy numbers compared with controls (p-values<0.001), while only females had shorter telomere lengths (p=0.02), highlighting a role of aging-associated molecular mechanisms in OCD. Kohlrausch et al. (2025) also examined telomere length in 105 patients with OCD and 105 controls, finding that both male and female patients had shorter telomere length compared to controls. Duration of illness was also correlated with telomere length in the entire sample and females only, but this was not significant after adjustment for age.

#### 3.5.7 Single gender studies

Thirteen single-gender studies were identified in the review period. While it is hard to draw conclusions about the impact of gender from these studies due to the inability to statistically test sex or gender effects, these are described in Table A5 for completeness and consistency with the other sections.

#### 3.5.8 Summary of key consideration

Methodological concerns:

- The majority of genetic studies used candidate gene approaches, which the field has moved away from given that such studies have generally lacked replication in subsequent genome-wide molecular studies.
- There is a need for genome-wide molecular studies in larger cohorts to systematically test for sex and gender differences, to provide insight into whether genetic and molecular factors may have a differential impact by sex or gender.
- Large sample sizes are key to identifying specific high-confidence genetic and molecular risk factors in genome-wide approaches, which becomes even more imperative when testing for sex and gender differences.

Unanswered Questions:

- Will larger genome-wide molecular studies offer any evidence of sex and gender differences in genetic and molecular predispositions to OCD?
- Do environmental factors, including significant life stresses and reproductive life events, interact with genetic predispositions differently in men and women?
- Does maternal or paternal age contribute differentially to OCD risk in offspring?
- Are there any genetic and molecular markers that can predict treatment response differently in men and women?

Hypotheses for future research:

- Childhood-onset OCD has a stronger genetic component than adult-onset OCD.
- Genetic liability for childhood-onset OCD differs between girls and boys, with potentially greater genetic liability among affected girls as observed in other childhood neurodevelopmental disorders.
- OCD comprises multiple genetic subtypes, with at least one subtype more prevalent in males and characterized by greater genetic overlap with neurodevelopmental disorders.

##### Highlights from Section 3.6 Altered neurocognitive and neurobiological functioning in OCD

<u>Self-report and behavior-based measures</u>: Most neurocognitive measures show little or no gender difference, and observed differences are often limited to specific aspects of a cognitive domain. In women, deficits have been reported across a broader range of domains, including metacognitive, emotional, visuospatial, sensorimotor, and reward-related processes; in men, difficulties are more concentrated in goal-directed control and decision-making. Emerging evidence suggests that different neurocognitive profiles may occur in both genders but differ in their relative prevalence.

<u>Neuroimaging measures:</u> Some studies report sex-associated differences in specific brain regions or circuits, but most measures show substantial overlap between males and females. More recent work suggests that sex may influence the relative prevalence of distinct neuroanatomical patterns rather than produce a uniform male–female contrast. Symptom profiles and stressful life events may contribute to these differences.

<u>Hormonal, growth factor, and peripheral biomarker measures</u>: Findings across hormonal, immune, metabolic, and blood–brain barrier-related biomarkers are heterogeneous, with limited evidence for consistent gender differences in mean levels. Gender differences are more often found in associations with symptoms, stress exposure, or other clinical characteristics.

<u>Key concerns:</u> Most studies are not systematic, with a narrow focus and small samples. Almost no studies have assessed both sex and gender.

<u>Future studies:</u> Progress will require integrative, longitudinal designs combining clinical, behavioral, neuroimaging, and biological measures, with explicit modeling of sex, gender-associated factors, and clinical and neurocognitive heterogeneity.

### 3.6. Altered neurocognitive and neurobiological functioning in OCD

Research on neurocognitive and neurobiological function in OCD has the potential to unravel the mechanisms underlying clinical differences between male and female patients and to identify sex- or gender-specific endophenotypes. These differences likely result from a complex interplay between various sex- and gender-related processes. Sex and gender differences in cognitive processing styles, brain structure and connectivity, and various neurotransmitter systems may influence OCD symptoms differently in men and women. Neuroactive sex steroids such as estrogen, progesterone, and testosterone may contribute to sex-specific patterns in symptomatology. Social experiences and adherence to gender norms may also shape how individuals experience and cope with their OCD symptoms. Thus, biological sex, gender, and associated social experiences may differentially affect cognition and biology.

Research on altered neurocognitive and neurobiological functioning in OCD has expanded rapidly over the past decade (Bandelow et al., 2017; Benzina et al., 2016), in an effort to identify underlying pathophysiological mechanisms and candidate targets for intervention. Studies of neurocognitive endophenotypes of OCD aim to make sense of the disorder’s heterogeneity. Although several recent studies have examined substantially larger samples, many studies remain too small to rigorously examine how sex or gender might contribute to neurocognitive diversity in OCD. We identified 46 studies published during the review period that directly compared neurocognitive or neurobiological functioning in males and females with adequate statistical power (2 in pooled-age samples, 9 in children/adolescents, and 35 in adults). Ten additional studies examined underpowered samples (Table A6). Eighteen studies focused exclusively on males or females.

#### 3.6.1. Neurocognitive functioning

Twenty-three papers investigated gender differences in neurocognitive functioning in individuals with OCD or OCS, utilizing self-reports or behavioral tasks (1 in a pooled sample, 4 in children/adolescents, and 18 in adults; Table A6). Most examined gender effects only in secondary or exploratory analyses.

Seven papers examined cognitive and metacognitive processes associated with OCD or OCS. Esbjørn et al. (2013) found that girls reported higher severity of subclinical OCS than boys, which indirectly contributed to higher metacognitive-belief scores in girls (M → OCS: β = −0.20, p < .001; OCS → metacognitive beliefs: β = 0.30, p < 0.001). Barcaccia et al. (2026) found that concurrent associations between obsessive beliefs and OCS were similar in boys and girls across three assessments (M: r = 0.33–0.37; F: r = 0.29–0.31), whereas obsessive beliefs prospectively predicted later OCS in boys (β = 0.20, p < 0.05) but not girls. Gülüm (2014) found that repetitive thinking was associated with OCs in both women (β = 0.28, p < 0.001) and men (β = 0.22, p < 0.001), whereas perceived locus of control was associated with OCs in women (β = 0.16, p < 0.001) but not men (β = −0.03, p > 0.05). Attachment anxiety mediated the association between repetitive thinking and OCs in both women (Sobel = 5.3, p < 0.001) and men (Sobel = 3.2, p < 0.001), but mediated the association between locus of control and OCs only in women (Sobel = 3.6, p < 0.001; M: p > 0.10). Cognitive fusion and experiential avoidance were similar in men and women with OCD (Xiong, 2021; cognitive fusion: Cohen’s d = −0.25, p = 0.18; experiential avoidance: d = 0.02, p = 0.93). Ding et al. (2025) likewise found no significant gender differences in the structure (p = 0.053) or overall strength (p = 0.09) of the network linking intolerance of uncertainty and OCS. Yong et al. (2026) found higher responsibility/threat-estimation beliefs in men than women across two waves (d = 0.18–0.22, ps < 0.004), whereas conformity and religiosity were higher in women (d = 0.06 and 0.13, respectively, ps < 0.004). Vivas et al. (2019) found that neuroticism was positively associated with OCS in both men and women, whereas agreeableness was negatively associated with OCS only in women (β = −0.32, p = 0.007). Overall, most cognitive vulnerabilities were similar across genders, with gender differences limited to selected associations with OCS.

Six papers examined gender differences in memory and basic information-processing functions. Martoni et al. (2015) found impaired visuospatial working memory in women with OCD (Cohen’s d ≥ 0.70), whereas men with OCD performed similarly to healthy men and women; visuospatial storage and recognition did not differ by gender (p > 0.10). Steinman et al. (2016; see also Ahmari, 2016) similarly found impaired prepulse inhibition (PPI), a measure of sensorimotor gating, in women with OCD compared with healthy women (Cohen’s d = 0.57, p = 0.039), whereas men with OCD performed similarly to healthy men and women. Bednarek et al. (2024) found impaired executive attention in OCD, but the magnitude of impairment did not differ between men and women (gender × OCD, p > 0.10). Shankar et al. (2026) found better working-memory performance in men than women at the 25th–75th percentiles of performance (β = 2.04–4.16, p = 0.01 to < 0.001), whereas no gender differences were observed among the lowest- or highest-performing individuals. Attention and visuospatial memory were also similar in men and women (p > 0.05). Puialto et al. (2026), in a mean 10.6-year follow-up, found a significant age × sex interaction for a composite of working memory and organizational strategy: with increasing age, women showed poorer performance relative to men (β = −0.056, 95% CI [−0.104, −0.008], p < 0.05). In contrast, neither the sex effect nor the age × sex interaction was significant for non-verbal memory. Thus, most basic attentional and visuospatial-memory measures were similar across genders, with differences limited to specific aspects of working memory, organizational strategy, and sensorimotor gating.

Five papers examined gender differences in emotional and reward processing. Shaw et al. (2021) found that affect intolerance was associated with OCD symptoms in both boys (β = 0.86, p < 0.001) and girls (β = 0.79, p < 0.001), although direct gender differences were not examined. Cougle et al. (2013) found no significant gender differences in distress intolerance among adults with OCS. Vivas et al. (2019) found greater sensitivity to emotional valence in women with high versus low OCS (Cohen’s d = 0.52, p < 0.001), whereas no corresponding difference was observed in men. Bezahler et al. (2024) found poorer emotion regulation among sexual-minority individuals with OCD than among cisgender heterosexual men and women with OCD (Cohen’s d = −0.25 to −0.30, p = 0.011–0.028), together with lower distress tolerance (d = 0.28, p =0.015). Wagner et al. (2025) found that lower social reward responsiveness was associated with OCS in both men (edge = −0.11 [95% CrI, −0.22 to −0.01]) and women (−0.09 [95% CrI, −0.15 to −0.02]). Additional associations differed by gender: lower responsiveness to hobbies was associated with OCS in men (−0.12 [95% CrI, −0.23 to −0.02]), whereas greater goal responsiveness was associated with OCS in women (0.08 [95% CrI, 0.01–0.14]). Overall, broad affective vulnerability was similar across genders, with differences emerging in specific aspects of emotional valuation and reward processing.

Six papers examined gender differences in executive control and decision-making. Cillo et al. (2019) found persistent risk taking, reflecting reduced adjustment to changing conditions, in men but not women with OCD (Cohen’s d = 0.41–0.47, p < 0.05). Cougle et al. (2013) found that goal persistence was negatively associated with OCS in men but not women, an effect driven primarily by obsession severity (M: β = −0.42, p < 0.01; F: n.s.; gender × obsessions: β = 0.21, p < 0.02). Ma et al. (2021) found increased decision thresholds and reduced evidence-accumulation rates in men with OCD relative to healthy men (decision threshold: p < 0.05; drift rate: p < 0.01), whereas women with OCD did not differ from healthy women. Eyüpoğlu et al. (2026) found no gender differences in cognitive flexibility among adolescents with OCD (failure to maintain set: βstd = −0.00, p = 0.99; categories completed: βstd = 0.06, p = 0.55; CCTT-2 completion time: βstd = −.07, p = 0.51). Shankar et al. (2026) found better planning performance in men at the 50th–95th percentiles (p = 0.01–0.05), but not at the 5th or 25th percentiles; measures of cognitive flexibility were similar in men and women across performance levels (p > 0.05). Whitehead et al. (2016) found that the relationship between OCD, depression, and goal-directed difficulties differed by gender: comorbid OCD/depression was associated with greater difficulties than depression alone in men (effect estimate = 1.17, p < 0.05), whereas in women, goal-directed difficulties were greatest in those with probable OCD alone. Overall, male difficulties were more consistently observed in goal persistence, risk adjustment, and evidence accumulation. In contrast, cognitive flexibility was largely similar across genders, and planning showed a male advantage at some levels of performance.

Across these studies, most neurocognitive measures showed little or no gender difference, and when differences were found, they were usually limited to specific aspects of a cognitive domain rather than the domain as a whole. For example, females performed worse on visuospatial working-memory errors and strategy use, but were comparable to males in visuospatial storage capacity and recognition; in a longitudinal study, women also showed greater age-related decline in a working-memory/organizational-strategy composite, whereas non-verbal memory did not differ by sex. One broader pattern nevertheless emerged: differences involving females spanned a wider range of cognitive domains, including metacognitive, emotional, visuospatial, working-memory/organizational, sensorimotor, and reward-related processes, whereas differences involving males were more concentrated in goal-directed control and decision-making. More recently, one study began to reframe the question by identifying neurocognitive profiles within OCD rather than searching for a single male or female cognitive phenotype. Mısır et al. (2025) identified two profiles—a globally impaired and a cognitively intact profile—and found that women were more frequently represented in the globally impaired profile (p = 0.015). These findings raise two possibilities. One is that a broadly impaired cognitive profile, more common in women, produces isolated deficits across multiple cognitive domains, with different studies capturing different manifestations of the same underlying impairment. Alternatively, OCD may include several more domain-specific cognitive profiles, with women represented across a wider range of these profiles and men more concentrated in profiles involving goal-directed control and decision-making.

#### 3.6.2. Neuroimaging

Neuroimaging studies have revealed aberrant brain structure and activity in specific regions and white matter tracts in OCD (Friedlander & Desrocher, 2006; Haghshomar et al., 2023; Saxena & Rauch, 2022). These findings implicate particular neural circuits, such as the cortico-striato-thalamo-cortical (CSTC) circuit, and large-scale networks, such as the default mode and salience networks, in the development of OCD symptoms. Limited evidence suggests that the functioning of these networks may differ in males and females with OCD (Ruigrok et al., 2014).

During the review period, seventeen studies (1 in a pooled sample, 4 in children/adolescents, and 12 in adults) examined gender differences in brain structure or function associated with OCD or OCS (Table A6). Eleven studies focused primarily on structural brain characteristics, and one additional study integrated structural and functional measures in a multimodal analysis (Xu, 2024). Four studies included functional MRI measures (Suñol, 2020; Ma, 2022; Xu, 2024; Li, 2026), with Xu et al. (2024) integrating structural and functional measures in a multimodal analysis. Two additional studies examined electrophysiological brain function using ERP (Hanna, 2026) or resting-state EEG connectivity (Mitiureva, 2024). Thus, although functional studies are now somewhat better represented than in the earlier literature, explicit examination of sex or gender effects remains uncommon relative to the large neuroimaging literature in OCD and represents an important knowledge gap.

Four studies examined gender effects on brain structure or function in children and adolescents. Suñol et al. (2018) found no gender differences in associations between OCS dimensions and gray- or white-matter volume. In a follow-up resting-state study, however, Suñol et al. (2020) found that connectivity within CSTC circuitry related to hoarding symptoms differently by gender: connectivity between the left mediodorsal thalamus and subthalamic nucleus was more strongly negatively associated with hoarding in boys than girls (M: r = −0.82; F: r = −0.34; p = 0.04), whereas mediodorsal thalamus–posterior cingulate connectivity was more strongly negatively associated with hoarding in older girls than in other children (older F: r = −0.51; others: r = −0.11; p = 0.004). Hanna et al. (2026) found no gender differences across ERP measures of error and response processing during a flanker task (all p > 0.05). In contrast, Li et al. (2026) found multiple diagnosis × gender interactions in adolescents with OCD. Compared with same-gender controls, boys with OCD showed lower left-insula activity (Cohen’s d = 0.75, pFDR = 0.02) and lower insula connectivity with the supramarginal gyrus and dorsolateral prefrontal cortex (d = 1.08–1.12, pFDR < 0.001), as well as reduced dynamic insula connectivity with the orbitofrontal cortex and precuneus (d = 0.73–0.83, pFDR = 0.01– 0.03). Girls with OCD instead showed higher right mid-cingulate activity (d = 0.88, pFDR = 0.02) and greater mid-cingulate connectivity with the precentral gyrus and precuneus (d = 0.85– 1.04, pFDR = 0.004–0.02). These neural measures were also related to symptom severity in a gender-specific manner (F: mid-cingulate activity–CY-BOCS total, ρ = 0.80, p < 0.001; M: insula–supramarginal connectivity–CY-BOCS total, ρ = −0.40, p = 0.04). Thus, pediatric findings range from no detectable gender differences to marked differences in the neural circuits associated with OCD, with the strongest recent evidence suggesting different functional patterns in boys and girls rather than a simple difference in the magnitude of the same abnormality.

A large pooled analysis across childhood and adulthood further showed that gender effects on thalamic structure differed by age. Weeland et al. (2022) found diagnosis × gender × age interactions across several thalamic subregions. Gender differences were most pronounced before age 12 (Cohen’s d = −0.32 to −0.42, p = 0.006–0.045), were not significant during adolescence (p > 0.10), and were small and limited to the anterior thalamus in adults (d = −0.07, p = 0.03).

Two studies investigated gender differences in corpus callosum structure in adults with OCD. Fractional anisotropy (FA) was more reduced in women with OCD than in men with OCD, relative to healthy individuals (diagnosis × gender: p = 0.036; Hawco, 2016). Corpus callosum thickness was increased in unmedicated men with OCD relative to healthy men (p = 0.032), whereas women with OCD were comparable to healthy men and women (diagnosis × corpus callosum: p =0.043; Park, 2011). Progesterone levels in women and testosterone levels in men have been associated with corpus callosum volume (Chavarria et al., 2014). For example, in women, total corpus callosum volume increases from the ovulatory to the luteal phase of the menstrual cycle (Khan, 2012). The corpus callosum is critical for coordinating activity across the hemispheres and contributes to movement control, cognition, and vision (Hinkley et al., 2012). Follow-up studies should examine whether sex hormones contribute to the observed gender differences in corpus callosum structure in OCD.

Braber et al. (2013) examined relationships between subthreshold OCS and gray-matter volume in the general population and found different associations in men and women. Higher OCS were associated with larger left middle temporal gyrus volume in men but not women (p <0.001). In the right middle temporal gyrus, volume was greater in men with high than low OCS, whereas the opposite pattern was observed in women (p < 0.001). Right precuneus volume also showed a gender-dependent association with OCS, with the lowest volume in men with high OCS and higher volumes in women with high OCS and men with low OCS (p < 0.001). These contrasting patterns require replication. Given the middle temporal gyrus’s involvement in visual perception and multimodal sensory integration (Gröhn et al., 2022; Jagiellowicz et al., 2011), future studies should investigate sex and gender differences in the impact of OCS on sensory processing.

Among other adult studies, findings again suggested substantial overlap between genders, with differences limited to specific brain features. Mitiureva et al. (2024) found no gender-dependent effects on resting-state EEG connectivity (F = 0.44, p = 0.71). Yi et al. (2026) similarly found no diagnosis × gender effects across most morphometric measures, but identified lower right paracentral-lobule cortical complexity in women than men with OCD (Cohen’s d = −0.71, p < 0.001). Ma et al. (2022) found higher right parahippocampal activity in women than men with OCD (F = 28.48, p < 0.001) and gender-dependent patterns of functional connectivity (OCD × gender: Wilks’ Λ =0.196, F = 4.60, p < 0.001).

Three recent studies indicate that gender differences in OCD neuroanatomy depend on latent subtype or factor expression rather than reflecting a uniform male–female contrast. Across studies, the largest gender effects were observed in gray-matter–dominated components (F1/F2 in Han, 2023; IC1 in Xu, 2024; GMV-defined subtypes in Wen, 2025), whereas multimodal or white-matter–weighted components showed little or no gender differentiation (Xu, 2024). These patterns frequently involved cortico-striato-insular-temporal regions, with males and females differing in the relative expression of particular neuroanatomical patterns. In two studies, patterns characterized by reduced gray-matter volume in the vmPFC, insula, cingulate, and temporal cortices were more common in women with OCD (Han, 2023, factor 2; Wen, 2025, subtype 2). Men were more frequently represented in patterns characterized by increased striatal/thalamic and frontotemporal gray matter (Han, 2023, factor 1; Wen, 2025, GMV1), and showed stronger expression of a widespread cortical gray-matter component in Xu et al. (2024, IC1). Notably, structural neuroimaging meta-analyses of trauma and PTSD report reduced gray matter volume in the ventromedial prefrontal cortex, insula, cingulate cortex, and temporal cortices, suggesting disruption of cortical–affective regulatory systems (Kühn & Gallinat, 2013; Li et al., 2014). The overlap between this trauma-associated pattern and the cortical reduction subtype observed in women with OCD raises the possibility of shared stress-sensitive neurobiological mechanisms; this needs to be investigated further.

Consistent with this interpretation, Real et al. (2016) reported that women with OCD were more likely than men to endorse stressful life events (SLEs) preceding illness onset (F:M OR ∼3; see Section 3.3.2.2.4) and demonstrated that SLE exposure moderated gray-matter volume patterns. Individuals with OCD who did not report SLEs showed increased gray-matter volume in the central tegmental tract (p < 0.05), whereas those with SLEs exhibited increased gray-matter volume in the right anterior cerebellum (p < 0.0005). Given the higher prevalence of SLE exposure among women, these findings suggest that stress-related modulation of regional gray-matter patterns may contribute to gender-linked expression of structural heterogeneity in OCD, without implying gender-specific effects in the absence of environmental context.

Further support for this interpretation comes from symptom clustering studies. Contrasting three data-driven OCD subtypes defined by symptom profiles, Reess et al. (2018) found that gender distributions differed across subtypes (p = 0.035), with a cluster characterized by elevated contamination symptoms predominantly female, consistent with prior symptom-severity findings (see Section 3.2.1). While men showed larger hippocampal volumes overall (controlling for intracranial volume; η² = 0.06, p = 0.035), the contamination-dominant subtype—composed of approximately 94% women—exhibited hippocampal volumes comparable to healthy controls. In contrast, two subtypes with more balanced gender distributions differed significantly from controls: individuals with intrusive-thought-dominant OCD showed larger hippocampal volumes, whereas those with predominant ordering/checking symptoms showed smaller volumes.

Together, these findings do not support a single, uniform gender effect on the neural correlates of OCD. Some brain measures show little or no gender difference, whereas others show gender-specific associations with OCD that are restricted to particular regions or circuits. Data-driven studies further suggest that males and females may be represented differently across neuroanatomical patterns, while symptom profiles and environmental exposures such as stressful life events may help determine which patterns are expressed. Larger, systematically designed studies are needed to distinguish direct effects of sex or gender from differences associated with symptom profiles, environmental exposures, hormonal factors, and other sources of clinical heterogeneity.

#### 3.6.3. Hormonal, growth factor, and peripheral biomarker measures

Fifteen studies (1 in children/adolescents and 14 in adults) have examined sex- or gender-related effects in peripheral biomarkers in OCD, including hormones, growth factors, immune, and metabolic markers (Table A6). Overall, findings are heterogeneous, and evidence for consistent gender^5^ effects in OCD is limited.

The hypothalamic–pituitary–adrenal (HPA) axis is the most frequently studied stress-response system. In children and adolescents, Bilgiç (2021) reported higher cortisol levels in boys with OCD than in male controls (Cohen’s d = 0.63, p = 0.046), whereas girls with OCD did not differ from female controls. ACTH levels were also elevated in boys with OCD (d = 0.37, p = 0.04), with a similar-sized but nonsignificant effect in girls (d = 0.39, p = 0.106). In adults, Kawano et al. (2013) found elevated salivary α-amylase in both women (F = 11.32, p < 0.01) and men (F = 9.87, p < 0.01) with OCD relative to same-gender controls, whereas salivary cortisol did not differ between OCD and control groups in either gender. In a general-population sample, Melia et al. (2019) found a significant gender difference in the association between cortisol awakening response and OCS (OCI-R × gender: β = −0.32, p = 0.02): the association was positive in men (β = 0.23, p = 0.06) and negative in women, with similar interactions for ordering and checking symptoms. Morning cortisol was also positively associated with the number of stressful life events in men (r = 0.30, p = 0.01) but not women. Labad et al. (2021) similarly reported gender-dependent associations between diurnal cortisol regulation and symptom dimensions, including ordering/symmetry severity (ordering × gender: β = −0.68, p = 0.015). Overall, gender differences were more consistently observed in associations between cortisol regulation and symptoms or stress exposure than in mean cortisol levels themselves. These findings are consistent with known sex differences in HPA-axis regulation (Heck & Handa, 2019), potentially modulated by gonadal hormones, although their role in OCD vulnerability remains unclear.

Erbay et al. (2015) examined neurosteroid and gonadal hormone levels in adults with OCD. Cortisol was elevated relative to same-gender controls in both women (Δ = 4.76, p = 0.008) and men (Δ = 6.98, p = 0.005). DHEA was higher in women with OCD than in female controls (Δ = 6.76, p = 0.009), whereas a comparable male OCD–control comparison was not significant or not reported. Among gonadal hormones, testosterone was lower in men with OCD than in male controls (Δ = −89.20, p < 0.047). Other reported differences in DHEA-S and progesterone involved cross-gender comparisons and therefore do not provide clear evidence of OCD-related gender effects. Thus, this study provides some evidence for shared cortisol elevation but more limited evidence for gender-specific differences in other neurosteroids and gonadal hormones.

Evidence for gender differences in growth factors and neurotrophins is limited. In children and adolescents, Bilgiç et al. (2021) found higher BDNF levels in both boys and girls with OCD relative to same-gender controls, with a larger effect in boys (M: Cohen’s d = 0.88, p =0.005; F: d = 0.37, p =0.002). In contrast, GDNF, NGF, and NTF3 showed little or no gender-related variation (d < 0.15, p >0.19). In adults, Fontenelle et al. (2012) found no gender differences in BDNF, NGF, or GDNF (z = −1.69, p =0.91), and Mina et al. (2023) likewise found no significant difference in BDNF between men and women with OCD (p =0.26). Thus, current evidence provides little support for consistent gender differences in growth-factor or neurotrophin levels, aside from a larger pediatric BDNF effect in boys.

Findings for oxytocin suggest gender-dependent associations rather than a consistent difference in mean levels. Marazziti et al. (2015) found no significant difference in plasma oxytocin between men and women with OCD (Cohen’s d = 0.22, p =0.46), but comparisons with same-gender controls differed by gender: oxytocin was lower in men with OCD than in healthy men (d = 1.72, p <0.001) and higher in women with OCD than in healthy women (d = 0.73, p =0.025). Oxytocin was also associated with fearful-avoidant (r =0.35, p =0.05) and dismissing attachment (r =0.38, p =0.04) in men with OCD but not women. A later study likewise found no significant difference in oxytocin between men and women with OCD (M = 5.81, F = 6.93, p >0.10; Marazziti, 2023b). Oxytocin is sometimes described as a “bonding hormone” (Heinrichs et al., 2009); oxytocin levels may increase in response to stress and may help regulate stress responses and facilitate seeking comfort from others (Neumann, 2002, 2008). Interpersonal connectedness is complexly affected in OCD. Individuals with OCD are often aware that their symptoms place a burden on people in their lives and may overcompensate by being especially accommodating or “nice” (Hyman & Pedrick, 2010). At the same time, men with OCD are more likely to remain single (Muhlberger, 2021; Section 3.2.2) and to have comorbid social phobia (Section 3.4.1.1). Targeted studies are needed to test whether gender differences in oxytocin– attachment relationships contribute to these differences in interpersonal functioning among individuals with OCD.

Studies of immune, metabolic, nutritional, and blood–brain barrier (BBB)-related markers have reported selective gender effects. Sarmin et al. (2024) found elevated IL-6 in both men and women with OCD relative to same-gender controls (M: p < 0.001; F: p < 0.001), whereas IL-1β was elevated only in men (M: p = 0.006; F: p = 0.14); however, men and women with OCD did not differ directly on either marker (p > 0.10). Kılıç et al. (2022) found substantially higher serum zonulin in women than men with OCD (Cohen’s d = 1.69, p < 0.001), whereas claudin-5 showed no significant gender difference (d = 0.30, p < 0.10). O’Neill et al. (2016) found no gender differences in Glx, creatine, choline, or GABA-related metabolite levels (p > 0.05). Vitamin D levels also did not differ between men and women with OCD, but associations with symptom dimensions differed by gender: in men, lower vitamin D was associated with greater interference and distress from obsessions and more time spent on compulsions (r = −0.35 to −0.38, p = 0.017–0.028), whereas in women it was more strongly associated with compulsion severity, resistance and control of compulsions, and poorer insight (r = −0.61 to −0.77, p = 0.005–0.048; Marazziti, 2023a). Finally, Algin et al. (2025) found higher vitamin B12 (β = 96.1, p < 0.001) and folate (β = 1.09, p = 0.023) and lower homocysteine (β = −4.97, p < 0.001) in women than men with OCD; higher folate in women was independently replicated by Marazziti et al. (2025; Z = 2.37, p = 0.018). Overall, direct gender differences were inconsistent across peripheral biomarkers, but when present they were often marker-specific or reflected different associations with particular symptom dimensions. In an all-female adolescent sample, Okumuş et al. (2026) identified a distinct plasma metabolomic profile in OCD relative to healthy controls, including lower linoleic acid and higher trihexosylceramide and 7α,12α-dihydroxy-cholestene-3-one; because the sample included only females, it is not clear whether these patterns generalize to males.

In summary, studies of hormones, growth factors, immune, metabolic, and blood–brain barrier-related biomarkers provide limited evidence for consistent gender differences in mean biomarker levels. More often, gender differences emerge in associations between these biomarkers and OCD symptoms, stress exposure, or other clinical characteristics. These effects are generally specific to particular biomarkers or symptom dimensions. Larger longitudinal studies are needed to determine whether such differences represent vulnerability factors, consequences of OCD or associated stress, or processes that modify the expression of the disorder.

#### 3.6.4. Summary of Key Considerations

Methodological concerns:

- Despite substantial growth in neuroimaging research on OCD, evidence on sex- and gender-related differences in neural correlates remains limited, with most studies examining these effects only secondarily and few explicitly modeling clinical heterogeneity.
- Neurocognitive and neuroimaging assessments in men and women with OCD are typically conducted at a single time point, implicitly assuming stability across hormonal states in menstruating participants, despite evidence that brain structure and function vary across the menstrual cycle (Hidalgo-Lopez & Pletzer, 2019; Holländer et al., 2005; Kheloui et al., 2021; Souza et al., 2012). In depression research, accounting for hormonal fluctuations has yielded important insights (Gordon et al., 2015; Harald & Gordon, 2012; Payne et al., 2009), highlighting the need for similar approaches in OCD.
- Recent studies have begun to move beyond comparisons of individual measures in males and females by identifying neurocognitive or neurobiological profiles within OCD and examining whether their prevalence differs by gender. However, such studies remain rare. Broader data-driven approaches integrating clinical, cognitive, hormonal, and neuroimaging measures are needed to characterize these profiles and identify factors associated with their differential representation in males and females.
- As discussed in Sections 3.2.1 and 3.3.1, reliance on self-report measures of obsessive– compulsive symptoms—particularly in pediatric samples—may introduce substantial bias. Neuroimaging studies should preferentially employ clinician-administered instruments when examining brain–symptom relationships.
- Across studies, there remains an almost complete absence of work that distinguishes sex and gender as separable constructs, or that includes transgender and nonbinary individuals.

Unanswered Questions:

- Do the diverse cognitive deficits reported in women reflect a broadly impaired cognitive profile that is more common in women, or a greater diversity of domain-specific cognitive profiles in women than in men? What factors determine why particular neurocognitive or neurobiological profiles are more common in one gender than another?
- Are neurocognitive profiles in individuals with OCD temporally stable (trait-level) or do they fluctuate with changes in severity of symptoms (e.g., in response to therapy)?
- Do reported sex differences in brain structure correspond to differences in brain function and neurocognitive performance, or do they reflect distinct compensatory mechanisms?
- How do hormone fluctuations over the phases of menstrual cycle impact brain functioning in cycling women, and are these fluctuations linked to OCD symptomology?
- How do relationships between gender and stress hormones evolve over development?

Hypotheses for future research:

- OCD may comprise multiple neurocognitive and neurobiological profiles that occur in both males and females but differ in their relative prevalence by sex or gender. Sex- and gender-associated biological, clinical, and environmental factors may help determine which profiles are expressed.
- During puberty, divergent symptom trajectories in boys and girls may reflect divergent trajectories in neurodevelopment. Neural and cognitive trajectories differ in boy and girls are modulated by changing hormonal profiles, as well as potentially by differential gender-associated experiences, and may serve as early biomarkers of the post-pubertal emergence or worsening of obsessive-compulsive symptoms.
- Relationships between HPA-axis regulation and OCD symptoms may differ in males and females and may contribute to differences in symptom presentation and trajectories across development.
- While females with OCD report preceding stressful events more often (Section 3.3.2.2.4), stress exposure may be more strongly associated with cortisol regulation in males than in females.

##### Highlights from Section 3.7. Treatment

<u>State of knowledge:</u> Research on gender differences in OCD treatment remains limited and largely secondary, post hoc, or underpowered.

<u>Treatment response:</u> CBT/ERP outcomes are generally comparable across genders. Pharmacological studies suggest a possible female advantage in fluoxetine response, particularly in adults. Neuromodulation studies show similar average outcomes, although male advantages may emerge in severe/refractory OCD.

<u>Treatment access and selection:</u> Gender differences appear more consistently in pathways to care and treatment selection than in efficacy. Boys may enter care more readily, whereas adult help-seeking findings are mixed. Women are more often prescribed SRIs and psychotherapy; men are more often prescribed antipsychotics. Treatment engagement is generally similar across genders.

<u>Future research:</u> Future research should combine adequately powered randomized controlled trials with real-world analyses of treatment allocation to examine how biological sex and gender interact with symptom dimensions, comorbidities, illness onset, and hormonal state (e.g., menstrual phase, estradiol levels) to shape potentially nonlinear OCD treatment response trajectories.

### 3.7. Treatment

Accounting for gender^6^ differences has been beneficial in many areas of evidence-based precision medicine (Gemmati et al., 2019; Shah & Kornstein, 2021). In OCD, such research is still fragmented at best. During the review period, 51 studies with sufficiently large samples examined gender differences in treatment effectiveness (i.e., at least 50 individuals with OCD per arm, which allows detecting effects of at least a large size); 14 were conducted in children or adolescents, 32 in adults, and 5 included both age groups. Fourteen additional studies tested gender differences in treatment response in underpowered samples (see Table 4, Table A7). Most analyses of gender differences were post hoc or secondary. Other topics included gender differences in help-seeking and treatment utilization (12 studies), side effects (one study), adequacy of medication use (2 studies), and potential mechanisms and markers of treatment response (2 studies). Nine studies looked at effectiveness of treatment only in women, and two looked at prevention of OCD in postpartum women. Most of these studies did not report strong gender differences. Still, some patterns emerged. Given the lack of research and heterogeneity of methods, these patterns should be interpreted as very preliminary.

**Table 4.**
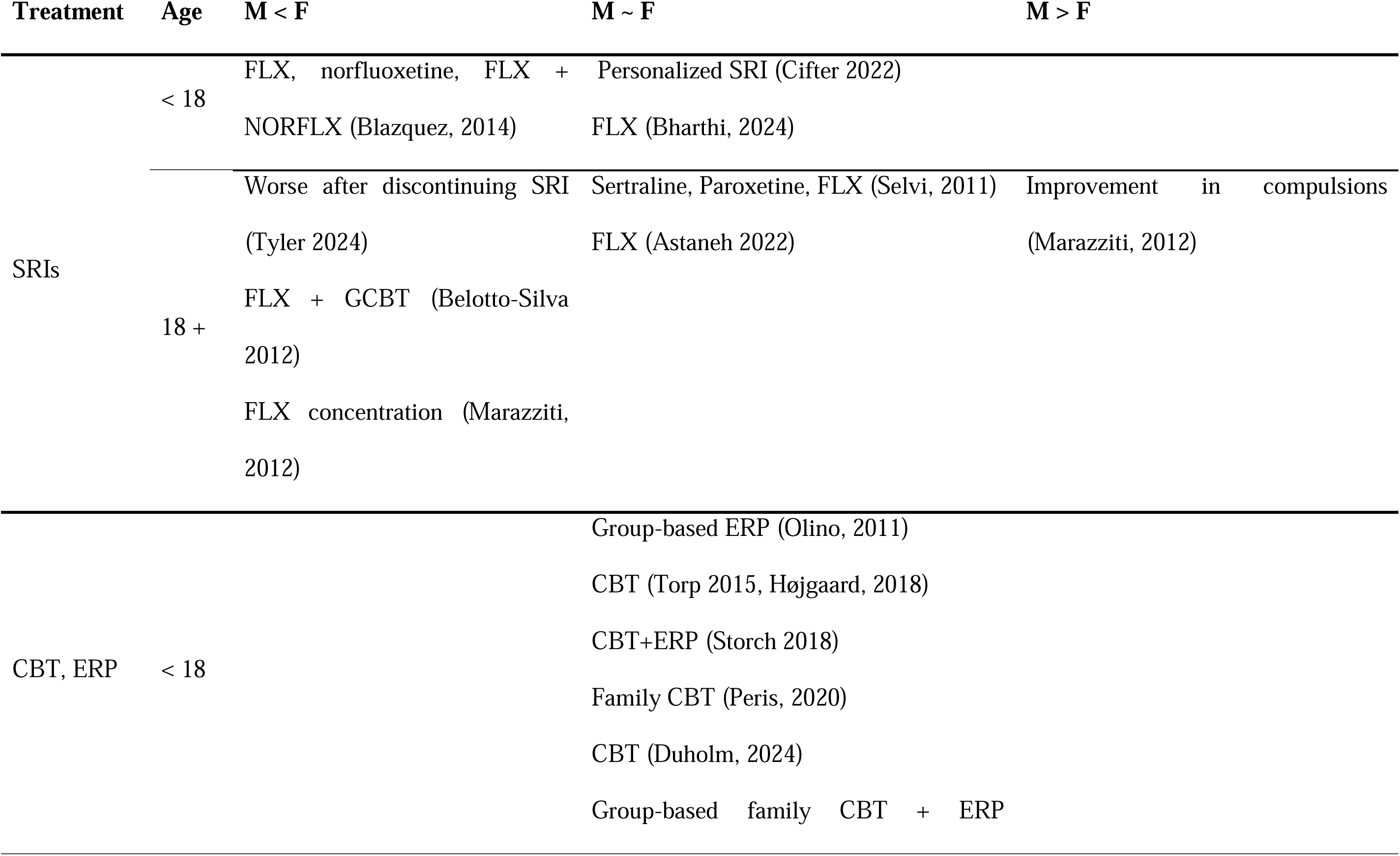
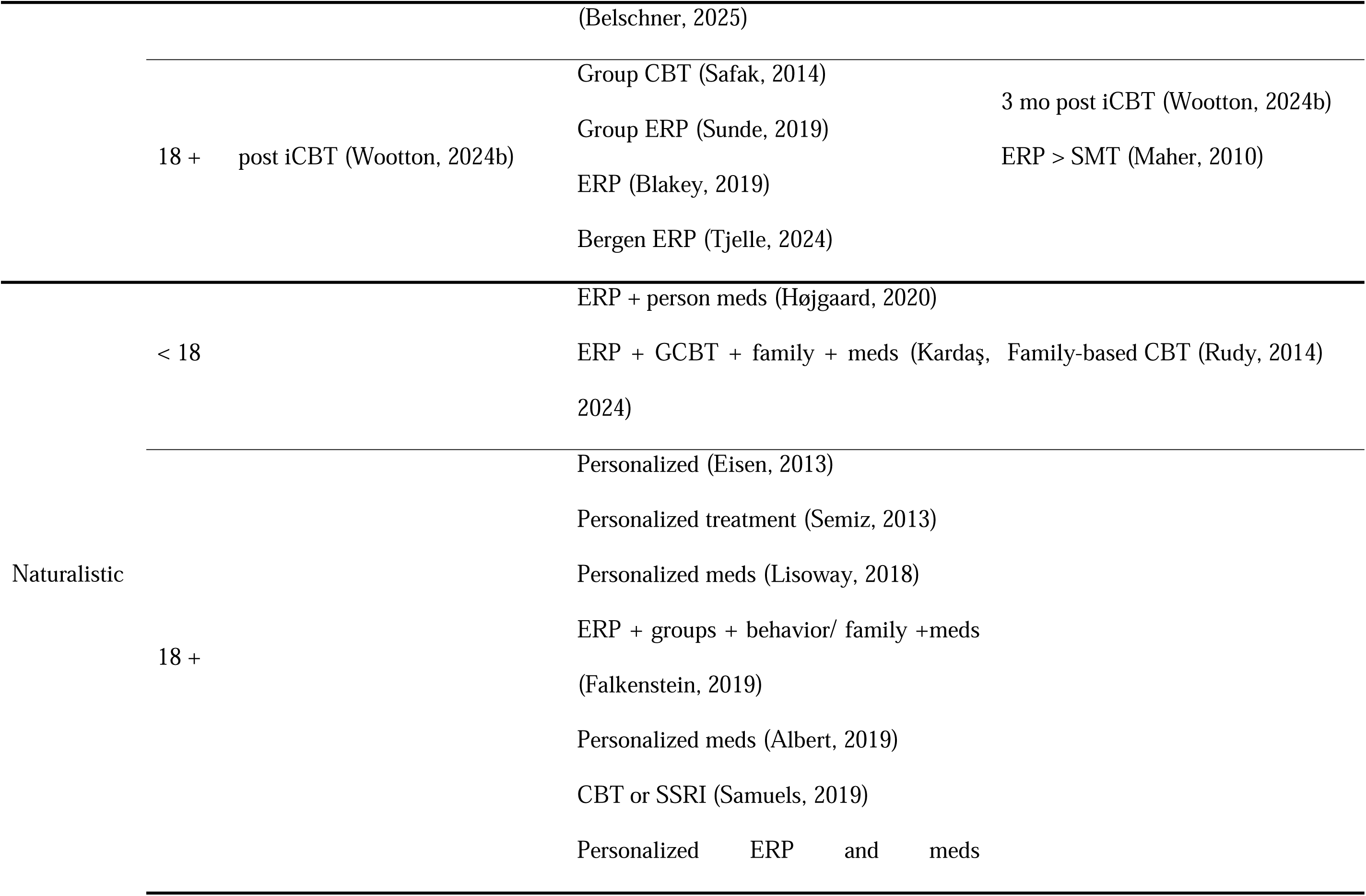
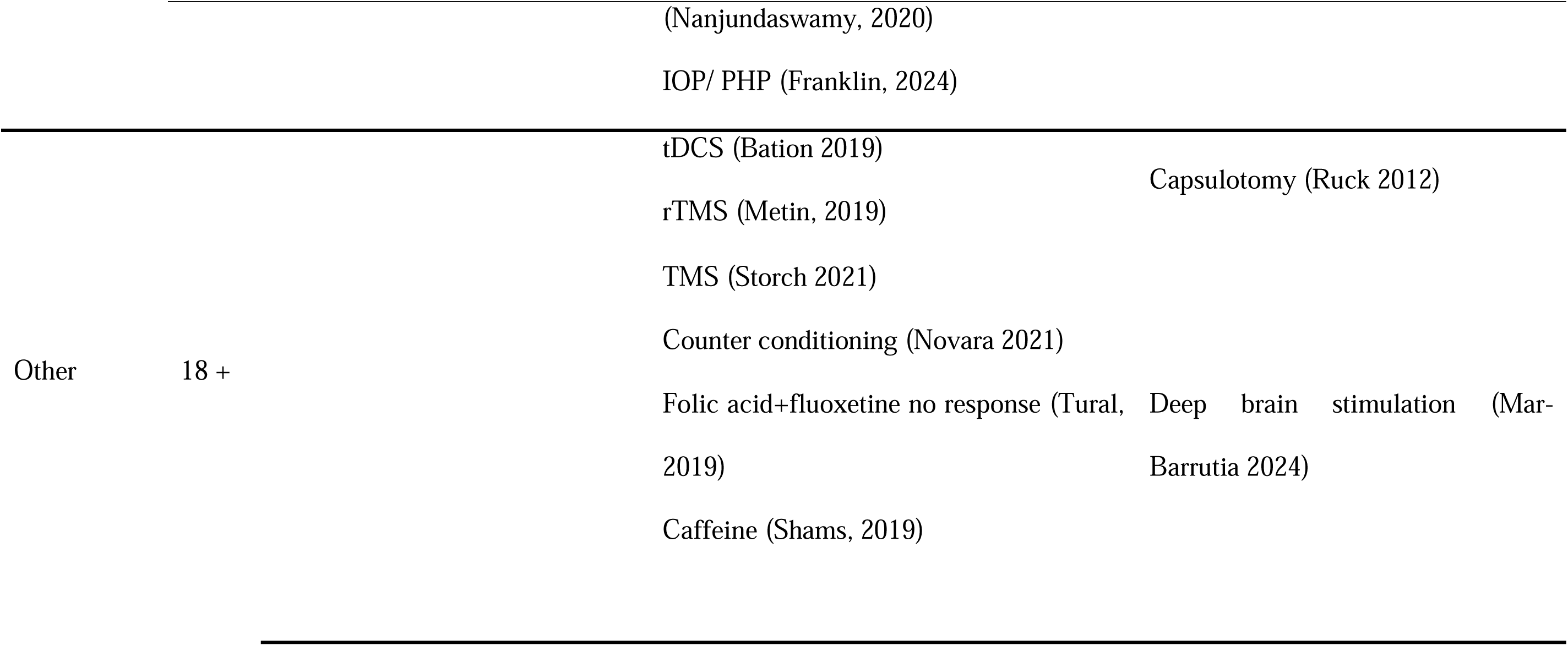
Gender differences in treatment response.

#### 3.7.1. Naturalistic observational studies

Naturalistic observational studies in both pediatric and adult samples generally reported comparable treatment outcomes between males and females (Table 4 and Table A7). However, Rudy et al. (2014) found that an intensive outpatient family-based treatment program in the United States yielded greater improvement in boys than girls (Cohen’s d = 0.69, p <0.01). In contrast, a comparable program conducted in Turkey (Kardaş, 2024) reported similar benefits across genders. These discrepant findings may partly reflect differences in medication status across cohorts, which were inconsistently documented and not incorporated into outcome analyses.

#### 3.7.2. Cognitive behavioral therapy (CBT)

Exposure and response prevention (ERP) is considered the gold-standard cognitive behavioral therapy for OCD. Sixteen studies (seven pediatric and nine adult) have examined gender differences in treatment response within structured CBT/ERP protocols, largely reporting null findings. For instance, a recent large retrospective study of intensive outpatient ERP (N = 712) found no gender difference in responder status (χ²(1) = 0.17, p = 0.68; Swisher, 2026). Interpretation of these results is constrained by several methodological limitations.

First, nearly all studies employed open-label designs, with only one incorporating an active control condition (stress management). This randomized study (Maher et al., 2010) found that the comparative benefit of ERP relative to stress management was significantly greater in men than in women, with a large interaction effect (Cohen’s d ≈ 0.8).

Second, most studies did not consider potential nonlinearities in treatment response trajectories. For example, Wootton et al. (2024b) reported greater symptom improvement in women immediately following internet-based CBT incorporating ERP modules (approximately 7.5% greater improvement; p = .015), whereas at 3-month follow-up the direction reversed, with men showing approximately 7.0% greater improvement (trend level, p = .075). Levy et al. (2024) found that endogenous estradiol levels moderated exposure outcomes among women with OCD: lower estradiol was associated with greater immediate distress reduction during exposure, whereas higher estradiol predicted superior extinction recall following a delay. Together, these findings suggest that biological state may differentially influence immediate versus sustained treatment effects, contributing to nonlinear trajectories of treatment response.

Third, few studies accounted for clinical heterogeneity within treatment samples. Only one study (Højgaard, 2018) adjusted for psychiatric comorbidity and observed a non-significant trend toward greater CBT effectiveness in girls (β_male_ = −0.47, p = 0.11). One pediatric study reported reduced CBT effectiveness among children with prominent contamination/cleaning symptoms (Duholm, 2024), although contamination symptoms over time did not differ by gender. As discussed in Section 3.2, contamination symptoms occur at comparable rates in boys and girls but are more prevalent among adult women. This may suggest that ERP effectiveness may be impacted by gender-associated differences in symptoms presentation. Systematic investigation of the role of patient heterogeneity—both across and within genders—is a critical next step toward developing more effective personalized CBT-based interventions.

#### 3.7.3. Pharmacological treatments

Serotonin reuptake inhibitors (SRIs) are considered first-line pharmacological treatment for obsessive–compulsive disorder. Three pediatric and six adult studies have examined potential gender differences in treatment response. In pediatric samples, findings were mixed. A prospective study suggested greater symptom improvement with fluoxetine in girls than in boys, with moderate effect sizes (Cohen’s d = 0.53–0.58; Blazquez, 2014), whereas a retrospective study based on self-reported outcomes found the opposite trend (Bharthi, 2024). Personalized SRI treatment demonstrated comparable effectiveness across genders, with response rates of 69.5% in girls and 62.8% in boys (Cifter et al., 2022).

In adults, three prospective studies reported stronger treatment effects of fluoxetine in women than in men, with moderate-to-large effect sizes. Consistent with this pattern, Tyler et al. (2024) found that women experienced significantly greater symptom worsening than men following SRI discontinuation while continuing CBT, suggesting greater reliance on pharmacological treatment effects. In contrast, individualized medication strategies showed similar effectiveness across genders (Selvi, 2011). Hasanpour et al. (2019) examined predictors of response to standardized fluvoxamine treatment (12 weeks, 150–300 mg) and identified a significant *age-of-onset × gender* interaction: women with later-onset OCD who were married and had lower compulsion severity showed an 85% probability of treatment response, whereas women with early-onset OCD demonstrated substantially lower response rates (29%).

Taken together, these findings tentatively suggest a female advantage in response to SRIs that appears more evident in adulthood. This pattern may reflect interactions between gender, illness characteristics, and biological state rather than sex or gender alone, and they should be tested in adequately powered studies that explicitly account for comorbidities, symptom dimensions, and hormonal factors (e.g., estradiol levels) to inform personalized treatment selection.

#### 3.7.4. Neuromodulation

Neuromodulation-based interventions, including transcranial magnetic stimulation (TMS), transcranial direct current stimulation (tDCS), and deep brain stimulation (DBS), represent emerging treatment approaches for OCD, particularly in treatment-refractory cases. Seven studies have examined potential gender differences in treatment response following neuromodulation, although several had insufficient power to reliably detect such differences. The two larger deep-TMS studies—Storch et al. (2021; N = 100) and Mudunuru et al. (2025; N = 378)—found no gender differences in average symptom improvement. In contrast, Elvan et al. (2026; N = 60) reported higher dTMS response rates in women than men (74.3% vs. 56.0%), although statistical significance of this difference was not reported. However, when Mudunuru et al. (2025) stratified patients by baseline OCD severity, gender differences emerged: men showed better response than women among patients with severe or extreme OCD (severe: 45.1% vs. 39.6%; extreme: 50.9% vs. 35.7%; p = 0.001). A male advantage was also reported in another small study restricted to patients with severe refractory OCD, in which response to DBS targeting the ventral anterior limb of the internal capsule (vALIC) was higher in men (77%) than in women (33%; Mar-Barrutia, 2024). Interestingly, Mudunuru et al. (2025) also found lower response among menopausal women than among perimenopausal (45.7% vs. 59.7%; d = 0.36, p = 0.031) and reproductive-age women (45.7% vs. 60.7%; d = 0.48, p = 0.020).

Thus, as with CBT/ERP, apparent gender differences in treatment response may depend on patient characteristics that are obscured in analyses of average treatment effects. Future studies should be adequately powered and explicitly examine potential sources of heterogeneity, including baseline severity, comorbidities, symptom dimensions, medication use, and hormonal or reproductive state.

#### 3.7.5 Unconventional treatments

Gold-standard treatments for OCD, including ERP and SRIs, are effective for many patients, but a substantial proportion remain symptomatic or achieve only partial response (Pittenger, 2023). Clinicians have therefore explored a range of augmentation strategies, often empirically combining established treatments with additional agents in an effort to improve response (Van Roessel et al., 2023; Walsh & McDougle, 2004). The potential role of gender in response to such augmentation strategies has been examined in two small studies. Tural et al. (2019; N = 36) found no gender difference in response to folic-acid augmentation of fluoxetine, whereas Shams et al. (2019; N = 40) reported greater symptom improvement in men receiving caffeine.

Psychedelic compounds have recently re-emerged as potential treatments for psychiatric disorders, including OCD. Within this renewed interest, two recent studies examined whether their use or effects differed by gender. In a small pharmacological challenge study (N = 19), Pellegrini et al. (2025) found no significant gender difference in Y-BOCS improvement one week after a 10-mg dose of psilocybin, but men showed greater sustained improvement at four weeks (β = −6.22, p = 0.009). In a larger naturalistic study, men were more likely than women to report psychedelic or related-substance use for OCD (OR = 1.41, 95% CI [1.02–1.90], p = 0.035; Mathai et al., 2026).

At the extremes of the therapeutic spectrum, attempts to improve outcomes have ranged from highly invasive neurosurgical procedures to approaches that move away from conventional medical treatment altogether. Although both ablative neurosurgery and herbal approaches have generated broader treatment literatures (Ayati et al.; Hageman et al., 2021; Kuygun Karcı & Gül Celik, 2020; Maroufi et al., 2026), gender has rarely been examined explicitly. In one small study of capsulotomy for severe refractory OCD, Rück et al. (2012) reported a trend toward poorer Y-BOCS outcomes in women than in men (effect size and p-value were not reported). At the opposite end, Bakhshinejad et al. (2026) evaluated a Dodder–Polypody herbal product added to fluvoxamine and found no gender difference in OCD symptom severity, although quality of life was higher in men (β = 5.96, p = 0.040).

As clinicians continue to search for more effective and creative strategies for patients who do not respond adequately to established treatments, it will be important to examine whether gender contributes to heterogeneity in response and can ultimately inform more personalized treatment selection.

#### 3.7.6. Treatment preferences

Although controlled studies evaluate treatment effects under standardized conditions, recent work has increasingly focused on the real-world processes that determine which treatments patients actually receive, including clinician, patient, and—in pediatric populations— parental preferences, as well as differences in treatment access and utilization.

Observational studies indicate gender differences in pathways to care. In pediatric samples, parents appear more likely to seek professional help for boys than girls once symptoms emerge (Mennies et al., 2020; F:M OR ≈ 0.5), consistent with earlier clinical recognition in males. In adults, findings are mixed. Two studies from Germany found longer delays from symptom onset to recognition, help-seeking, or treatment initiation in men than women (Stengler, 2012; Bey, 2025; d ≈ 0.2–0.4). By contrast, null findings were reported across more geographically diverse settings, including the United States, Italy, and a 10-country sample spanning both high- and low-/middle-income countries (George, 2025; Girone, 2025; Stein, 2025). A community study from Thailand found the opposite pattern, with men more likely than women to have sought professional treatment for elevated OC symptoms (55.6% vs. 27.0%, p = .023; Kaewpila, 2025). As discussed in Section 3.3, sociocultural context may contribute to these differences, although current evidence is too limited to establish a geographic pattern.

Gender differences are also evident in treatment selection and acceptability. In pediatric samples, girls were more likely than boys to receive psychotherapy (OR = 1.17, 95% CI [1.12– 1.22]) and antidepressants (OR = 1.42, 95% CI [1.37–1.48]) after an OCD diagnosis, whereas antipsychotic use was largely comparable (Baweja, 2025). Boys also showed markedly greater acceptance of sensor-assisted CBT technology than girls (Cohen’s d = 1.51), despite similar general technology affinity (Alt, 2025). In adults, males were more frequently prescribed typical and atypical antipsychotics (F:M OR = 0.68–0.86), whereas females were more likely to receive SRIs, non-benzodiazepine anxiolytics, benzodiazepines, and analgesics (Isomura, 2016; N = 10,523; F:M OR = 1.21–1.64). Women also reported stronger preference for psychotherapy over medication (Patel, 2017), and use of internet-based CBT increased more among women during the COVID-19 pandemic (OR = 1.57, 95% CI [1.19–2.06]; Li, 2022). Women were also somewhat more likely to receive virtual partial-hospital treatment and men residential care (φ ≈ 0.14, p = .053; Woodson, 2025). Clinician characteristics may also influence treatment selection: male psychiatrists were more likely than female psychiatrists to refer patients for psychiatric neurosurgery (OR = 6.45, 95% CI [1.08–38.57], p = .041), although attitudes toward these procedures did not otherwise differ by gender (Alqahtani, 2026).

Gender differences in treatment engagement and continuity of care appear limited. In pediatric samples, CBT booster-session attendance did not differ by gender (F:M OR = 0.92, 95% CI [0.35–2.43]; Negreiros, 2019), and a recent family-based CBT study likewise found nearly identical gender distributions among completers and noncompleters (Belschner, 2025). In adults, dropout or completion was also similar across genders in pharmacotherapy (M OR = 1.12; Jakubovski, 2013) and internet-based CBT (Wootton, 2025), although one study reported lower CBT dropout among women (F:M OR = 2.8; Cramer’s V = 0.21; Tabeleão, 2024). No gender differences were evident in psychiatric emergency-room utilization before versus during the COVID-19 pandemic (F:M OR = 0.50 in 2018 and 0.68 in 2020; Sun, 2023). Among patients receiving medication, men and women were also similarly likely to have received an adequate trial, including multiple SRIs and recommended maximum doses (gender coefficients β = 0.18– 0.27; Cohen, 2025).

Together, these findings suggest that treatment allocation in routine care may reflect patient preferences and clinical decision-making processes as much as treatment efficacy itself. Understanding how help-seeking, treatment selection, and engagement shape treatment exposure is therefore essential for linking real-world care patterns with controlled evaluations of CBT and pharmacological interventions.

#### 3.7.7. OCD prevention?

As already described in Section 3.3.4, the burden of postpartum OCD may be reduced or even prevented by providing psychoeducation or targeted CBT interventions to expecting parents (Timpano 2011; Challacombe, 2017; Steinman, 2025). This represents an important opportunity for prevention. It also raises the question of whether targeted intervention during other identifiable vulnerability periods, such as puberty or in the aftermath of trauma (especially sexual trauma), could be of similar benefit (Section 3.3.4, Section 3.3.2.2.4).

#### 3.7.8. Summary of Key Considerations

Methodological concerns:

- Our review did not identify any RCT specifically designed and adequately powered to test gender differences in treatment response. Gender analyses in randomized trials were generally secondary or post hoc.
- When evaluating intervention-related changes in symptom severity, symptoms should be assessed at multiple time points before and after treatment to account for possible fluctuations across phases of the menstrual cycle and for nonlinear treatment-response trajectories. Assessments should also consider potential gender differences in insight, as well as changes in insight that may occur with repeated therapist contact.
- Factors that can influence treatment outcomes are also differentially associated with sex and gender, including comorbidities, age of onset, stress exposure, symptom profile (e.g., autogenous versus reactive obsessions), reproductive-cycle events, and genetic factors. Future studies should account for these sources of heterogeneity and examine whether they modify gender differences in treatment response. Treatment groups should also be evaluated for systematic differences in these characteristics.
- Treatment response may vary over time and may not follow the same trajectory across genders (e.g., immediate versus sustained response). Studies relying solely on a single post-treatment endpoint may therefore miss clinically meaningful gender differences in treatment maintenance or relapse.
- Gender differences in help-seeking, treatment selection, and access may introduce selection bias into treatment studies. Men and women entering a given treatment may therefore differ systematically in illness duration, severity, comorbidities, or prior treatment exposure, complicating interpretation of apparent gender differences in response.

Unanswered Questions:

- What drives gender differences in help-seeking and pathways to care across age groups and sociocultural settings?
- Why are men prescribed antipsychotics more often than women? Are these differences driven by treatment response, patient preferences, prescriber bias, symptom profile, or comorbidities?
- Are the apparent gender-specific advantages in response to particular treatments—such as greater fluoxetine response in women and greater neuromodulation response in men with severe or refractory OCD—robust, and what clinical or biological factors account for them?
- Do comorbidities modify treatment response differently in men and women, and could this help explain inconsistent gender effects across studies?
- Can reproductive or menstrual phase influence treatment response in women with OCD?
- Can clinically significant OCD symptoms be prevented through interventions timed to specific high-risk periods?

Hypotheses for future research:

- Treatment effectiveness may be moderated by age-of-onset × gender interactions; for example, SSRIs may be more effective in women with adult-onset OCD.
- Biological state, including endogenous estradiol levels and reproductive transitions, may influence extinction learning, pharmacological response, and maintenance of treatment gains.
- Comorbidity profiles may modify treatment effectiveness differently in males and females.
- Gender differences in treatment response may become more apparent within clinically severe or refractory subgroups than in average treatment effects.
- Early or preventive interventions may be most effective when timed to periods of increased vulnerability, such as pregnancy or the aftermath of severe trauma.

### 3.8. Animal models of OCD pathophysiology and related processes

#### Highlights from Section 3.8. Animal models of OCD pathophysiology and related processes

<u>Impact of the Search Strategy</u>: The search was designed to identify sex and gender differences in human OCD, not animal studies. Thus, the animal literature identified here is likely incomplete, particularly for relevant work not explicitly framed in relation to OCD.

<u>Possible mechanisms of sex differences in OCD-related pathological behaviors:</u> Identified studies examined estrogen and estrous-cycle effects on OCD-relevant circuits and behaviors; sex differences in dopamine dynamics, glial function, and synaptic proteins, including receptors and scaffolding proteins; and sex-dependent effects of serotonin and stress manipulations.

<u>Caveats:</u> Animal models cannot reproduce the full complexity of human OCD or assess the cognitive symptoms and internal states central to the disorder. Animal behaviors should therefore not be equated with human OCD symptoms. Nevertheless, conserved anatomy and genetics may allow animal studies to provide valuable mechanistic insights.

<u>Future studies:</u> In order to more comprehensively understand the literature on sex differences in animal studies of potential relevance to OCD, an additional scoping review should be undertaken with keywords selected for this purpose, to identify studies not captured here and to better identify gaps in recent research.

While our search strategy (see Section 2) was designed to capture articles about sex and gender effects in individuals with OCD, it also retrieved 63 studies in animal model systems, mostly in mice. These provide an important counterpoint to human literature and an opportunity to more precisely examine mechanistic questions. Rather than discard these studies, therefore, we summarize them here. We note that the coverage of the relevant literature in this section may be sparser than it would be were a search strategy constructed to focus specifically on studies in animals. For example, studies examining sex^7^ effects in potentially relevant processes, such as habit learning (Quinn et al., 2007) or striatal synaptic plasticity (Campanelli et al., 2021), would not be captured by our search if they did not explicitly frame these findings in the context of OCD. We nevertheless believe that this summary of the papers in animal model systems retrieved by our search constitutes a useful component of this scoping review.

How best to conceptualize the relevance of studies in animal model systems to a complex human neuropsychiatric condition such as OCD merits comment. It is unlikely that a mouse can recapitulate the full syndrome of human OCD, given the approximately 100 million years of evolution that separate the two species, the relatively rudimentary nature of the mouse’s frontal cortex, and the language-dependent nature of many OCD symptoms, or at least of their assessment. Even if a mouse could meaningfully recapitulate OCD, it is not clear how we could evaluate it, given how much of the evaluation depends on linguistic self-report of internal states. Invoking ‘a mouse model of OCD’ is therefore simplistic at best, and often misleading. On the other hand, much of the genome and many aspects of brain function are conserved among mammals, including between mice and humans, and so it would be astonishing if studies of fundamental processes in mice did not have something to teach us about the human brain in general, and OCD in particular. Conceptually, there are at least three ways that studies of sex effects in mice may provide insight into relevant sex and gender effects in human OCD: they may examine a brain circuit of clear relevance to OCD; they may examine a process of relevance to OCD; or they may characterize a model system that we have independent reason to believe captures aspects of the pathophysiology of OCD. While we will not critically evaluate each model reviewed here in light of these considerations, they provide a useful frame for the discussion that follows.

What counts as OCD-relevant behavior in mice is a vexed question. Certainly not every instance of anxiety-like behavior, repetitive behavior, or increased grooming in a mouse indicates a parallel to human OCD. Here we follow the literature that we are reviewing, in the spirit of a scoping review: we consider such behaviors to be ‘compulsive-like’ or potentially relevant to OCD when the underlying literature does so, without either defending or critiquing the interpretation. Ultimately the relevance of any particular model system to human OCD is an empirical question and will be determined by future studies testing whether mechanisms that have been elucidated in mice prove to be relevant to in human disease.

A key advantage of an animal model system is the ability to ask causal questions, through direct genetic or other molecular manipulations, circuit disruption or manipulation, or targeted pharmacological interventions. A second advantage is the ability to examine brain function at a much higher resolution than is possible in humans. A third is that experiments can generally be parameterized, controlled, and repeated much more rigorously than in human studies, allowing for more precise conclusions and higher degrees of confidence in findings. Finally, animal models that convincingly capture aspects of OCD pathophysiology could be powerful systems in which to test novel therapeutics without many of the complexities and confounds that plague clinical trials.

Many studies have examined animal models that purport to capture OCD pathophysiology or to probe OCD-relevant structures or processes, but relatively few have examined sex as a moderator. Most studies have examined a single sex (usually males). This began to change in 2014, when the National Institutes of Health (NIH) instituted a new requirement that all NIH-funded studies address sex as a biological variable (Miller et al., 2016). This has proven to be a critical change and has led to a dramatic increase in relevant publications over the years covered by this review. For instance, one recent study showed that in rats, females develop habitual behavior much earlier during training than males (Schoenberg, 2019). In another study, mapping quantitative trait loci (QTL) in mice that relate to prepulse inhibition (PPI) showed significant QTLs on chromosomes 7 and 17 for the acoustic startle response when sex is used as a covariate, suggesting that males and females modulate this OCD-related behavior differently (Samocha, 2010). Additionally, female mice are more likely than males housed under the same conditions to develop abnormal grooming and barbering behaviors (Ratsuki, 2025).

#### 3.8.1. Estrogen

Estrogen can influence both differences between males and females and fluctuations in females due to the estrous cycle, pregnancy, or other hormonal changes. One study of rabbits found that pregnancy upregulates brain activity in orbito-frontal, anterior cingulate, and piriform cortices, among other areas, in conjunction with repetitive, compulsive-like straw-carrying activity (Cano-Ramírez, 2017). This activity is also regulated by dopaminergic circuitry, as administration of either D1 or D2 antagonists reduced straw-carrying bout duration (Hoffman, 2012).

Investigations in a line of mice bred for varying levels of spontaneous compulsive nest-building behavior showed that females exhibit higher levels of nest-building behavior and had higher scores in the marble burying test, which has been suggested as a model of spontaneous compulsive behavior (Mitra, 2017a). Lactating females in the compulsive nest-building group had reduced compulsive-like behaviors compared to non-lactating and nulliparous females of the same genotype. Treatment of these non-lactating and nulliparous females with a D2 antagonist reduced their elevated nest building and marble burying behaviors (Mitra, 2017b); ovariectomy (OVX) exacerbated compulsive-like behaviors and anxiety, while treatment of OVX mice with an E2 agonist, β-estradiol, reduced them (Mitra, 2016). These findings suggest that, at least in this model, states of increased estrogen mitigate compulsive behaviors, via a dopamine-regulated mechanism.

Another set of experiments found that genetic reduction of A-Kinase anchoring protein 13 (AKAP13), an estrogen receptor enhancer expressed in the hypothalamus and pituitary gland (Eddington, 2006), induced abnormal marble-burying and grooming behaviors in female mice but not in males, further suggesting a protective role for elevated estrogen in females (Baig, 2018). Overall, these experiments suggest an important interaction between estrogen and compulsive-like behavior that could explain some of the differences seen between women and men with OCD.

#### 3.8.2. Serotonin

Serotonin dysfunction is commonly hypothesized to be an important contributor to OCD (Hesse et al., 2005; Marazziti et al., 1992; Sinopoli et al., 2017; Soomro et al., 2008; Stengler-Wenzke et al., 2004) primarily because of the therapeutic effect of serotonin reuptake inhibitors (SRIs) in the treatment of OCD patients (Pittenger, 2021; Skapinakis et al., 2016). In one set of genetic studies, introduction of a mutation, Thr276Ala, into the gene for the serotonin transporter (SERT), disrupting its conformation and regulation by phosphorylation, reduced dominance behavior in the tube test in male mice, but not females, and reduced marble-burying behavior in females, but not males (Meinke, 2020, 2022). This supports the relevance of the serotonin system to these plausibly OCD-relevant behaviors and is an elegant demonstration of differential effects of sex despite an identical genetic alteration.

A number of pharmacological studies have probed the serotonin system’s modulation of potentially OCD-relevant behaviors. The 5-HT1A receptor agonist 8-hydroxy-2 (di-*n*-propylamino)-tetralin (8-OH-DPAT) reduces serotonin levels in the frontal cortex (Arora et al., 2013) and has frequently been used to produce repetitive behaviors of potential relevance to OCD. In one study, the 8-OH-DPAT model was examined in older rats that were either experiencing irregular cycles (perhaps analogous to perimenopause in humans) or persistent diestrus (analogous to menopause) to draw parallels to the effects of menopause on OCD. Here, they found that 8-OH-DPAT enhanced perseverative choices in the T-maze only in rats with persistent diestrus, and not in females with irregular cycles or males at a comparable age (Olvera-Hernández, 2013). Administration of a 5-HT1B agonist increased marble-burying behavior in female mice, while reducing nestlet shredding, enhancing sucrose intake, and decreasing overall GABA protein levels (Salloum, 2024).

2,5-dimethoxy-4-iodoamphetamine (DOI), a psychedelic compound that is a selective agonist of serotonin 5-HT2A and 5-HT2C, has been shown to induce motor-like tics and OCD-like behaviors in mice (Canal & Morgan, 2012; Fantegrossi et al., 2010). In one study, investigating the ability of raloxifene to treat behavioral symptoms in mice treated with DOI, researchers found that elevated compulsive grooming phenotypes were equally abated in both sexes by treatment with 10mg/kg raloxifene. However, in regard to ear scratching response, only females showed a decrease with treatment, suggesting that raloxifene may modulate abnormal scratching behaviors differently between sexes in a DOI model (Gorburg, 2025).

Metachlorophenylpiperazine (mCPP), a non-selective 5-HT2c agonist, induces excessive grooming behavior in male and female mice, a commonly used proxy for compulsive-like behavior (da Silva et al., 2022; Graf et al., 2003). After mCPP treatment, females, but not males, show deficits in extinction retention in a contextual fear paradigm, only in the metestrus/diestrus phases of the estrous cycle (Reimer, 2018). These findings suggest that serotonergic signaling interacts with estrogen in the regulation of plausibly OCD-relevant behaviors, with potential implications both for sexual dimorphism and for hormonal modulation of symptoms in women. In another mCPP study in rats, mCPP at 2mg/kg impaired reversal learning in males in a T-Maze task including increased omissions and reaction time. Comparatively, mCPP at 2mg/kg in females impaired both acquisition and reversal learning, with increased omissions, reaction time, and reduced accuracy (Stoppiglia, 2025). These findings suggest sex-dependent effects of mCPP on learning and cognitive flexibility, with broader impairments observed in females.

#### 3.8.3. Dopamine

Dysregulation of the dopaminergic system has long been associated with OCD (Bais et al., 2014; Denys et al., 2004; Hesse et al., 2005; Millet et al., 2003; Olver et al., 2009; Pittenger, 2021; Van Der Wee et al., 2004). Early imaging work in humans demonstrated that baseline dopaminergic function differs between the male and female brain, with women exhibiting higher dopamine synthesis capacity (Laakso et al., 2002; Pohjalainen et al., 1998) and increased dopamine transporter activity (Hsiao et al., 2013; Lee et al., 2015; Williams et al., 2021; Zachry et al., 2021). More recently, measurements of evoked dopamine release in the dorsal striatum of mice showed that females have higher DA release and more rapid recovery compared to males, paralleling the results in humans (Arvidsson, 2014). Similarly, chemogenetic activation of nigrostriatal dopamine projections increased prepulse inhibition in female but not male rats, suggesting sex-dependent effects of striatal dopamine signaling on sensorimotor gating (Casado-Sainz, 2021). In a dopamine β-hydroxylase knockout model, which increases dopamine and reduces norepinephrine levels, behavioral changes were comparable between males and females, although females exhibited enhanced cortisol responses (Lustberg, 2023). Further study of the contributions of these sexually dimorphic dopamine dynamics to relevant behaviors is warranted.

#### 3.8.4. Norepinephrine

Knockout (KO) of Dopamine-β-hydroxylase (*Dbh*), which catalyzes the reaction that converts dopamine to norepinephrine, produces abnormal nest building and marble-burying behaviors in both male and female mice (Lustberg, 2020). Sex-dependent noradrenergic abnormalities have also been reported in Slitrk1-KO mice: although both sexes showed increased noradrenergic fiber density, males exhibited greater increases in locus coeruleus noradrenergic markers and prefrontal norepinephrine levels (Hatayama, 2022). These findings suggest that norepinephrine-related mechanisms may show sex-specific effects, although the literature remains limited.

#### 3.8.5. Synaptic dysregulation

Structural components of excitatory synapses are implicated in the pathophysiology of several neuropsychiatric disorders, including OCD. Manipulating some of these components impacted sexes differently.

The Disks Large Associate Protein 1 (Dlgap1, aka GKAP or SAPAP1), a postsynaptic scaffolding protein (Yao & Yang, 2003), is associated with several neuropsychiatric disorders, including OCD (Mattheisen et al., 2015; Pittenger, 2015; Stewart et al., 2013), autism spectrum disorder (ASD) (Mosca et al., 2017), and schizophrenia (Kajimoto et al., 2003; Kirov et al., 2012; Rasmussen et al., 2017). While both female and male Dlgap1 KO mice showed shorter grooming latencies, only males showed deficits in dig test and social approach behaviors. Interestingly, heterozygous male mice, but not full knockouts, showed significant increases in grooming bouts and grooming duration. This finding has been interpreted as suggesting a male-specific compensatory mechanism that emerges in the complete absence of Dlgap1 (Coba, 2018). In a separate study of Dlgap1 knockout mice, females exhibited longer non-scratching time than males, and fluvoxamine modulated this behavior only in females (Minagawa, 2025). Knockout of related protein SAPAP3, the mouse homolog of DLGAP3, produces compulsive-like behavioral abnormalities and striatal circuit changes (Wan et al., 2014; Wan et al., 2011; Welch et al., 2007), including network dysfunction and decreased power in local field potential oscillations in the lateral orbitofrontal cortex (Lei, 2019) and amygdala hyperexcitability in the intercalated nuclei (Laurent, 2025), but these effects have not been reported to differ between sexes, including in analyses of enhanced self-grooming and head-body twitches (Brownstein, 2024; Gattuso, 2025). A study in juvenile SAPAP3 KO mice suggests no sex differences in the open field, elevated plus, marble burying, or tube dominance tests, although in a test to uncover and consume a sweet treat, only male SAPAP3 KO mice showed deficits (Lazar, 2025). Decreases in locomotion and sociability have also been reported, with equivalent effects in males and females (Gattuso, 2025). One study suggests that SAPAP3 knockout alters grooming syntax differently across sexes, with increased paw grooming observed in females and greater overall grooming time in knockout males compared with control males, whereas differences between knockout and control females were smaller (Wilson, 2024; Wilson, 2025; Wang, 2025). This effect on grooming in males is exacerbated by social isolation stress. In a light-dark box anxiety paradigm, SAPAP3-KO mice show a worsening in anxiety-related behaviors over time, with this anxiety phenotype occurring earlier in females compared to males (8wks vs 11wks of age, Wilson, 2025).

In a study examining the deletion of the carboxyl-terminus (C-tail) of Neuroligin 2 (NL2-StopKI), a scaffolding protein vital to GABAergic synapse formation and stability (Varoqueaux (Ali et al., 2020; Graf et al., 2004; Poulopoulos et al., 2009; Van Zandt et al., 2019; Varoqueaux et al., 2004), while both male and female NL2-StopKI showed an elevated marble burying phenotype and nestlet shredding, this effect was more pronounced in males. Conversely, in the elevated plus maze, although both sexes of NL2-StopKI had significantly higher anxiety levels compared to controls, female NL2-StopKI showed an elevated anxiety phenotype relative to males (Pandey, 2025). This suggests that dysfunction in the NL2 protein may have different degrees of effect on different compulsive-like and anxiety behaviors between the sexes.

Abnormal function of *metabotropic glutamate receptors (mGluRs)* has also been implicated in OCD (Ade et al., 2016; Akkus et al., 2014; Karthik et al., 2020), and administration of ADX88178, an mGlu4 receptor positive allosteric modulator, increased open-arm entries in the elevated plus maze in both male and female mice, without evidence of a sex-specific treatment effect (Kalinichev, 2014). Memantine, an NMDA receptor antagonist, increased nest-building behavior in females 24 hours after treatment at a dose of 10 mg/kg compared with males (Macedo et al., 2024). Similarly, S-ketamine, another NMDA receptor antagonist, reduced marble-burying behavior in female mice at high doses, particularly in those previously identified as prone to more compulsive-like behaviors (Ayub, 2022).

*Kv7.2 channel mutations* are associated with early-onset epileptic encephalopathy and autism (Dirkx et al., 2020; Gamal El-Din et al., 2021; Lauritano et al., 2019; Lehman et al., 2017; Weckhuysen et al., 2012) but may also be relevant to OCD, as some manipulations result in compulsive-like grooming behaviors,. Knocking down Kv7.2 in mice to imitate the more common loss-of-function mutations seen in patients with Kv7.2 dysregulation produced abnormal hyperactive behavior, increased open arm entries in the elevated plus maze (suggestive of reduced anxiety), and enhanced grooming in both sexes but significantly more so in males. Furthermore, Kv7.2 heterozygous males, but not females, showed elevated marble-burying behavior and heightened social dominance (Kim, 2019). The differences in severity of behavioral outcomes between sexes in OCD-related gene manipulations suggest underlying sex-dependent differences in how these mutations affect synaptic function, or sexually dimorphic compensatory mechanisms.

*The CYFIP1 gene*, and the protein it encodes, is necessary for the maintenance and stability of neuron dendritic organization and complexity (Pathania et al., 2014); deletion of this gene in humans is seen in Type I Prader-Willi Syndrome (PWS) (Angulo et al., 2015), which is characterized by food-related and other compulsive behaviors (Dykens, 1999). In mice, *Cyfip1* haploinsufficiency produces significant increases in compulsive behavior in the marble-burying test and trends towards increased head dips in the hole board test. These phenotypes are modulated by genotype of a related gene, *Cyfip2*, which is implicated in binge-eating disorder, in a manner dependent on the sex of both the tested animal and the parent (Kirkpatrick et al., 2017).

Slitrk1, a gene involved in regulating axonal and dendritic growth as well as synapse formation, produces sex-specific phenotypes when knocked out. Although both male and female newborn mice exhibit abnormal vocalizations and increased growth of noradrenergic neurites in the prefrontal cortex, male mice show greater elevations in noradrenaline levels and increased serotonergic varicosities (Hatayama, 2022).

Genome-wide association studies (GWAS) have identified several loci near the protein tyrosine phosphatase receptor type D gene (PTPRD) as genome-wide significant or strongly suggestive for association with OCD. This receptor is involved in cell adhesion and synaptic specification. In PTPRD knockout mice, no sex differences were observed in the open-field, digging, or splash grooming tests. However, sex-specific effects emerged in prepulse inhibition (PPI): knockout females exhibited PPI deficits, whereas males showed increased startle magnitude (Ho, 2023).

Synaptic regulation by interneurons, particularly the cholinergic interneurons (CINs) of the striatum, may also contribute to behavioral phenotypes relevant to OCD. Depletion of CINs produces a distinct behavioral profile in male mice, including reductions in acoustic prepulse inhibition (PPI) and impaired sensorimotor gating. When combined with spatial confinement stress, CIN depletion in males—but not females—leads to increased grooming and digging behaviors, more frequent head-jerking, deficits in cross-modal PPI, and enhanced marble burying (Caddedu, 2023). These findings suggest that striatal cholinergic interneurons may play a role in regulating stress-induced behavioral pathology relevant to OCD.

#### 3.8.6. Glia

Glial abnormalities and inflammation are associated with numerous disorders; their contribution to the pathology of OCD is unclear, although studies in *HoxB8* knockout mice, which exhibit elevated grooming, have suggested a link (Chen et al., 2010; Nagarajan et al., 2018). Depletion of Hoxb8-expressing microglia leads to compulsive grooming phenotypes in both sexes; Tränkner et al. (2019) found this grooming to be significantly more severe in females. Furthermore, OVX in female Hoxb8 KO mice reduced pathological grooming, while administration of estrogen and progesterone in male Hoxb8 KO mice enhanced it. Optogenetic stimulation of Hoxb8 microglia in the dorsomedial striatum or medial prefrontal cortex also produced abnormal grooming behaviors, however, no sex differences were apparent, indicating that direct stimulation might not pick up on the nuances apparent in a genetic depletion model (Nagarajan, 2024). The fact that manipulation of microglia leads to sex differences in pathological behaviors of potential relevance to OCD highlights this as a new area of interest.

#### 3.8.7. Environmental stress

OCD is associated with elevated levels of stress and of cortisol (Adams et al., 2018; Faravelli et al., 2012; K. Gustafsson et al., 2008; Labad et al., 2018; Sousa-Lima et al., 2019). In one study, early-life stress produced compulsive-like behaviors – perseverative nose poking in a lever pressing task – in female but not male rats (Brydges, 2015). Experimental preeclampsia in rats, a form of neonatal stress, induced elevated marble-burying behavior in offspring; this behavior was significantly greater in female rats than in males (Muzyko, 2020). In a model of maternal separation (MS) at two different time points (1-9 post-natal day/early or 10-20 post-natal day/late), several female-specific phenotypes emerged relative to findings from earlier studies in males. In the forced swim test, early MS induced increased immobility in females only, which was accompanied by an increase in spine density on basal dendrites in the female CA1, although late MS increased immobility in both sexes. Late MS also increased marbles buried in the marble burying tests in females only (Kajita, 2025). These studies suggest that early stress may have a more significant impact on OCD-related behaviors in females than in males.

Sleep deprivation, a stressor that can interact with and exacerbate OCD symptoms, can also induce behavioral pathology. In female rats, REM sleep deprivation increased locomotion and center time in the open field, elevated marble-burying behavior, reduced pain thresholds, and decreased immobility time in the forced swim test. Interestingly, crocin—a compound with neuroprotective and anti-inflammatory properties—normalized open-field locomotion, reduced marble burying, restored pain thresholds, and increased immobility time in the forced swim test (Houshyar, 2024). In another study combining REM SD with fear conditioning, compulsive grooming increased in both males and females following REM SD, both alone and in combination with fear conditioning; fear conditioning overall reduced grooming in REM-SD animals. In the marble-burying test, females showed greater marble burying than males across REM-SD conditions (Payamandi, 2025). This suggests that REM SD can affect behavior differently between sexes and may increase some compulsive-like behaviors to a greater extent in females.

Stress is associated with activation of the hypothalamic-pituitary-adrenal (HPA) axis and elevated levels of cortisol (or corticosterone in mice), which acts primarily by binding the glucocorticoid receptor (GR/NR3C1). Knockout of GR/NR3C1 in the hindbrain produces potentially OCD-relevant behaviors: excessive barbering of the fur on the face, back, and stomach and repetitive digging behavior. Both males and females show this behavior, but they show different alterations of the HPA system: females showed elevated apoptosis in the adrenal cortex, while males showed elevated adrenal weight and blood corticosterone (Gannon, 2019). As a group, these studies suggest that stress may have greater impact on OCD-related outcomes in females. Further investigation of the interaction of stress and behavioral pathology relevant to OCD is warranted.

In a study of deep brain stimulation in rats, an effective but invasive procedure sometimes used to treat severe, treatment-resistant OCD in human patients (Mar-Barrutia et al., 2021), stimulation induced improvements in a set-shift operant task were seen to be larger in females than in males, although some caveats were given (small n’s and physical impediment from cable tethering) for this assessment (Sachse, 2026).

#### 3.8.8. Summary of Key Considerations

The search strategy of this scoping review was not specifically designed to identify animal model studies that may be relevant to OCD. Thus, coverage of the relevant literature is likely incomplete. As noted above, work in animal models provides enormous advantages in fine-grained mechanistic analysis, causal manipulations, and rigor. However, establishing a confident relationship between findings in animals and the pathophysiology of a complex human psychiatric disorder such as OCD raises substantial conceptual challenges. We have not attempted to critically evaluate this relationship across the models reviewed above; rather, we view these studies as identifying a series of domains in which further work is needed. While interpretive humility is essential, delineating sex effects in animal model systems may help refine mechanistic hypotheses and identify targets for testing in humans.

## 4. Overall Discussion

The study of sex and gender differences has a complex and often controversial history. Some have argued that these differences form a mosaic of brain and behavior variations (Joel et al., 2015), possibly existing on a continuum (Zhang et al., 2021), while others dismiss the concept of a “gendered brain” all together (Eliot, 2019). A middle-ground perspective suggests that sex and gender differences exist but that their effect sizes are typically small in adults; they may be more pronounced during development (Grissom & Reyes, 2019). In the study of psychopathology, research on sex and gender has largely focused on developmental (e.g., autism, childhood schizophrenia), externalizing (e.g., ADHD, conduct disorder, substance use), internalizing (e.g., depression, anxiety, eating disorders), and personality disorders (Zahn-Waxler et al., 2006). Studies that systematically examine sex and gender effects in OCD remain limited.

Reviews of sex and gender effects in OCD have been published approximately every 10 years. They have evolved over the decades, from initially dismissing the significance of sex effects (Castle et al., 1995) to recognizing evidence of differences in epidemiological, phenomenological, and genetic studies (Lochner et al., 2004; Lochner & Stein, 2001; Mathis et al., 2011). Our scoping review aims to build on this work by systematically analyzing research published from July 2009 to March 2025, starting where Mathis et al. (2011) left off. We have reviewed original studies on sex and gender influences across clinical, epidemiological, neurobiological, genetic, and treatment-related aspects of OCD. We have also summarized meta-analyses and systematic and narrative reviews that were published during the same period (see Appendix 1).

We find a wealth of research but a lack of synthesis. Between July 1^st^, 2009 and May 31^st^, 2026, 1,038 original studies investigated sex and gender effects in various aspects of OCD, including clinical presentation, neurobiology, genetics, and treatment outcomes. While many studies reported negative or contradictory findings, some clear patterns emerged. Despite 72 meta-analyses, 19 systematic reviews, and 47 narrative reviews that addressed sex and gender differences during this period, many of these patterns remain underexplored. We have outlined these patterns and attempted to synthesize evidence to develop some hypotheses for future research.

There has been profound neglect of distinctions between sex and gender in this research literature. Among the 712 human studies reviewed, only four assessed both sex and gender of study participants. Two studies using the same sample (Rautio, 2020; 2022) identified four transgender individuals within a cohort of 172 participants with body dysmorphic disorder. Brooks (2022) identified six transgender or gender-diverse individuals in a general population sample of 594 participants. Bezahler (2022) examined treatment outcomes among sexual minorities (N = 34). Across the broader literature, most studies appear to conflate sex and gender, making no effort to account for participants whose sex assigned at birth and gender identity may not align. Emerging research suggests that misalignment between sex as assigned at birth and experienced gender in gender minorities is associated with chronic stress and ongoing trauma and may uniquely affect the interplay between biological and social factors (Cardona et al., 2022; Pantalone et al., 2020). Ignoring these complexities may limit the interpretability of findings and contribute to inconsistent results.

This scoping review had four key goals: (1) to provide a comprehensive and systematic survey of evidence concerning sex and gender effects in OCD; (2) to assess methodological considerations in this body of research; (3) to identify knowledge gaps; and (4) to propose a roadmap for future hypothesis-driven investigations into sex and gender effects in OCD, incorporating recommendations for precision medicine approaches. In addition to addressing these points in each section separately, we offer an overall summary of the key considerations here.

### 4.1. Summary of Key Considerations

#### 4.1.1. Methodological concerns

- <u>The lack of systematic assessment of biological sex and gender:</u> Most reviewed studies did not ask participants to report both sex assigned at birth and gender identity. Instead, participants were typically asked whether they were men or women (or male or female), and in many studies the method used to determine this classification was not reported. Research on OCD in transgender and nonbinary individuals remains very limited. Eight studies included gender-minority participants, but only five directly examined gender-minority status or gender-minority-related effects. This gap is particularly notable in large epidemiological and genetic studies.
- <u>Variability in Assessment Methods:</u> Findings differ across self-report, parent-report, clinician-administered assessments, and medical records. These discrepancies are particularly important in pediatric samples and may reflect gender differences in insight, reporting, social desirability, and social norms. Heterogeneous assessment methods also complicate comparisons of comorbidity and developmental trajectories. Greater standardization, including direct comparison across informants and assessment methods, is needed, particularly in younger populations.
- <u>Diagnostic Cutoffs:</u> The thresholds used to distinguish subthreshold obsessive–compulsive symptoms from clinically significant OCD may influence observed sex and gender patterns. Several studies found different gender distributions for subthreshold symptoms versus diagnosed OCD, suggesting that conclusions may depend on where clinical cutoffs are placed. However, no identified study directly established or validated different diagnostic cutoffs for males and females. Examining symptom distributions across the full continuum, and testing whether optimal thresholds differ by sex, gender, age, or developmental stage, may improve diagnostic characterization.
- <u>Subtyping into more homogeneous subgroups:</u> Much research has focused on predefined OCD symptom domains, such as checking and contamination. Recent data-driven studies suggest value in examining patterns of symptom co-occurrence and broader clinical and biological profiles. In adults, symptom organization may be more differentiated by gender, with women showing a potentially more heterogeneous symptom structure. Data-driven subtyping can complement traditional clinical categories and may help identify mechanistically meaningful phenotypes that occur in both sexes and genders but differ in their relative prevalence. Such approaches should incorporate comorbid psychiatric and physical conditions, functional burden, and other sources of clinical heterogeneity that may shape individual presentations.
- <u>Accounting for menstrual phase in cycling women:</u> A substantial proportion of women with OCD report symptom fluctuations across the menstrual cycle, with estimates ranging from approximately one-third to one-half. Nevertheless, most studies assess symptoms, cognition, neurobiology, or treatment response at only one or a few time points without accounting for menstrual phase or hormone levels. This implicitly assumes short-term stability across the menstrual cycle, despite evidence that OCD symptoms and relevant neurobiological measures may fluctuate with hormonal state. Such unmeasured variability may obscure both diagnostic effects and interactions with sex- and gender-related factors. Future studies should incorporate menstrual phase and, where feasible, direct measurement of reproductive hormones.

#### 4.1.2. Unanswered Questions

- How do biological sex and gender identity independently and jointly contribute to OCD presentation, burden, neurobiology, and treatment response, and how do these relationships change across development?
- What can large-scale genome-wide, neuroimaging, and multimodal studies reveal about sex- and gender-related mechanisms and heterogeneity in OCD?
- Could sex- and gender–informed approaches improve OCD assessment and diagnosis—for example, by testing different diagnostic thresholds or assessment strategies, or by accounting for reproductive hormonal state? Should assessment also be tailored to developmental and life stages such as childhood, puberty, pregnancy/postpartum, and later life?
- How do comorbid psychiatric and medical conditions interact with sex and gender to shape OCD presentation, burden, neurobiology, and treatment response across development?
- How do gender differences in help-seeking, diagnostic recognition, and pathways to care affect the OCD populations captured in research and, consequently, our understanding of OCD phenomenology and treatment?
- Can effective preventive or early interventions be developed by targeting periods of increased vulnerability, such as early childhood in individuals with high familial risk, puberty, reproductive transitions, or the aftermath of severe trauma?

#### 4.1.3. Hypotheses for future studies

Research on sex and gender differences in OCD can provide valuable insights into the specific needs and challenges faced by men and women, as well as inform approaches for personalized treatment. Additionally, it offers a perspective on the heterogeneity of OCD.

Previous studies have proposed distinct OCD subtypes, including based on age of onset (Poletti et al., 2023; Rosenberg & Keshavan, 1998; Taylor, 2011) or a history of trauma or precipitating stressors (Hühne et al., 2024; Rowsell & Francis, 2015). Early-onset OCD (before puberty) has generally been reported to be more common in boys (Nakatani, 2011; Wang, 2011; Albert, 2015; Grover, 2018). Later-onset OCD (after age 40) has been reported as more frequent in women (Frydman, 2014; Albert, 2015; Sharma, 2015); in one study, however, this difference disappeared after controlling for PTSD diagnosis and a history of pregnancy (Frydman, 2014). Results on gender differences in stress-precipitated OCD are less consistent, with some studies reporting a female bias (Real, 2011; Rosso, 2012) and others finding no difference (Fontenelle et al., 2012). The symptom presentation, course, and structural organization of the brain may differ in stress-related OCD compared with non-stress-related OCD (Goldberg, 2015; Real, 2016).

Synthesizing literature on sex and gender differences allows us to enrich and refine these models. We propose two OCD subtypes (OCD-I and OCD-II), which we detail below focusing on two questions: “*Are males or females differ in susceptibility to this OCD subtype?”* and “*Are risk factors for this OCD subtype more common in males or in females?*” OCD-I and OCD-II may differ in symptom presentation and progression, comorbidity profiles, stress responses, genetic markers, neurocognitive functioning, cortisol regulation, and treatment responses.

Early-onset OCD has a male preponderance and a higher familial risk (Eichstedt & Arnold, 2001; Geller et al., 1998; Geller et al., 2003). We term this form of the condition *OCD-I.* In addition to earlier onset in boys and higher familial or genetic risk, it is characterized by a less robust cortisol regulation system (P. E. Gustafsson et al., 2008) and by frequent comorbidity with neurodevelopmental disorders such as TD, ASD, ADHD, and schizophrenia (Grassi et al., 2022). A male bias in earlier onset is frequently seen in these neurodevelopmental disorders and has been attributed to a female protective effect (or, equivalently, to differential vulnerability in males, who may be more susceptible to prenatal and early-life stressors) (Breach & Lenz, 2022; Jacquemont et al., 2014; Pinares-Garcia et al., 2018). The mechanisms underlying this female protective effect are unclear; one proposed evolutionary explanation is that it may help offset the higher reproductive stress load that females typically experience (Singh et al., 2021).

We suggest that OCD-I may occur more frequently in females than is generally recognized and may even occur at similar rates across men and women, but that its onset is later in girls and women, due to this female protective effect. This would be analogous to the later age of onset of major symptoms in girls and women that is seen in autism and schizophrenia (Bargiela et al., 2016; Selvendra et al., 2022). Later onset in females may occur if and when life stresses – particularly reproductive cycle events – overwhelm the female protective effect (Real, 2011; Rosso, 2012). To experience early onset of OCD-I, girls may need to have higher familial risk for neurodevelopmental difficulties and may therefore have elevated rates of neurodevelopmental comorbidities. Consistent with this idea, our review revealed that in children with neurodevelopmental diagnoses, the male bias in early-onset OCD seen in the general population is reduced, or even reversed to a female bias (see Table 3 and Section 3.4). Similarly, the male bias in the prevalence of ADHD and impulse control disorders seen in the general population is reduced in children and adults with OCD (see Table 2 and Section 3.4).

We propose that a second subtype of OCD, which we term *OCD-II*, is less strongly genetic/familial and tends to onset following stress, including traumatic stress. Because the likelihood of experiencing a significant stressful event increases with age (Benjet et al., 2016; Cohen et al., 2019), OCD-II tends to have a later onset than OCD-I (especially in males, in whom OCD-I onset is particularly early) and is more common in adults. OCD-II is likely to be more common in women, simply because women are more frequently exposed to reproductive and interpersonal stresses and to trauma (especially intimate partner and sexual trauma) in our society (Benjet et al., 2016; Valentine et al., 2019). If this is so, the female bias in OCD-II should attenuate, and may disappear, when males and females are compared at equivalent levels of relevant stress or trauma exposure. Several empirical findings are consistent with this prediction. Sexual trauma and unwanted sexual experiences are far more common in adolescent girls and women (Spence, 1997), but the likelihood of experiencing OCS following these experiences, when they occur, appears to be similar across genders (Barzilay, 2018). OCD comorbid with PTSD shows no gender differences (Nacasch, 2011), and high OCD prevalence has also been reported among heavily traumatized male populations, such as hospitalized veterans (Taghva, 2017). Among trauma-exposed youth, OCD was strongly female-predominant (F:M OR = 3.04; Pinciotti, 2025), but this association was substantially attenuated after adjustment for trauma exposure and related clinical factors, consistent with differential exposure contributing to the female bias. However, sex × trauma interactions and pubertal stage were not examined, leaving the source of the residual female bias unclear.

Strong evidence links unwanted sexual experiences to contamination-related OCD symptoms (Peles, 2014; Waller, 2014; Ishikawa, 2015; Kennedy, 2017; Krause, 2021; Tang, 2022). Contamination symptoms occur at similar rates in boys and girls (Cervin, 2020; Schultz, 2018), but their onset after puberty and before age 18 is significantly more likely in girls (F:M OR = 16.7; Grover, 2018). This female bias continues into adulthood, though it becomes less pronounced (F:M OR = 4.2; Grover, 2018), mirroring gender differences in the prevalence of unwanted sexual experiences across age groups (Benjet et al., 2016; Spence, 1997).

Further systematic investigations are needed to examine how sexual and other types of trauma impact symptoms presentation in males and females and what factors lead to the development of OCD-II—rather than, for example, PTSD or anxiety disorders—following significant stressors. While differential genetic predispositions may play a role, environmental factors and variable use of coping strategies could also contribute. For example, Peles et al. (2014) found that sexually abused women with a history of opioid addiction were more likely to develop OCD, whereas those without opioid addiction exhibited higher rates of PTSD and dissociation. They speculated that opioid use might help mitigate PTSD and dissociative symptoms but increase risk for OCS. Future studies should explore this hypothesis across diverse traumatized populations. Overall, while the course and presentation of OCD-II are likely to be similar in males and females, this subtype may be more common in adolescent girls and adult women, potentially because they are more likely to experience the stressors that trigger OCD-II, although differential susceptibility may also contribute.

Differentiating OCD-I and OCD-II based on the age of onset and history of precipitating stressors may work well in males. However, in females, both OCD-I and OCD-II are more likely to present later than in males (during or after puberty) and be linked to stressful (e.g., reproductive) events, making differentiation more challenging. Greater representation of both OCD-I and OCD-II pathways in adult women could help to explain the higher heterogeneity in cognitive and symptom profiles seen in women with OCD, compared to men (Raines, 2014; Shkalim, 2017; Sections 3.2.1 and 3.6.1). A subset of women experience OCD onset during reproductive events, and one-third to one-half of women with OCD report symptom fluctuations across the phases of the menstrual cycle (Guglielmi et al., 2014; Vulink et al., 2006); reproductive-event-linked onset and menstrual-cycle-related symptom fluctuations might serve as biomarkers for distinguishing OCD-I and OCD-II in women. Detailed examination of natural history, genetic predispositions, and comorbidity profiles in women whose symptoms are linked to reproductive stresses, compared to those without such links, could help address this question. If the two proposed subtypes are validated, differentiating between them will improve diagnosis, prognosis, and treatment selection for males and females with OCD.

### 4.2. Conclusion

Researching sex and gender differences in OCD is not just about uncovering variations in clinical presentation and underlying mechanisms—it is also about advancing a more inclusive and precise understanding of a complex disorder that affects millions of lives. The evidence reviewed here suggests that sex and gender differences in OCD are not uniform, but vary across symptom domains, developmental stages, comorbidities, environmental exposures, biological systems, and pathways to care. Rather than defining fixed male and female forms of OCD, sex and gender may help reveal clinically and mechanistically meaningful heterogeneity within the disorder. Recognizing how biological and social factors impact individuals with OCD will help us develop targeted treatments, identify underexplored risk factors, and ultimately improve outcomes for diverse populations. Progress will require studies that distinguish biological sex from gender, examine developmental and longitudinal trajectories, account for reproductive and hormonal state, and explicitly model clinical heterogeneity, comorbidity, stress and trauma exposure, and treatment selection. Ignoring these differences risks perpetuating gaps in care, particularly for transgender and nonbinary individuals and other populations that remain substantially understudied. Understanding sex and gender effects in OCD is therefore a critical step toward greater equity in mental health research and care, and toward capturing the full complexity of the disorder. The OCD-I/OCD-II framework proposed here provides one testable example of how sex- and gender-related patterns may be used to generate hypotheses about distinct developmental and etiological pathways rather than simply describe average male– female differences. In 2015, the NIH mandated consideration of sex as a biological variable in federally funded research, including the inclusion of both male and female animals in preclinical studies. Consistent with the expected lag between policy implementation and publication, Figure 2 shows an increase in studies reporting sex and gender differences in the years following this policy. We hope this trend will continue, with increasing emphasis on systematic, adequately powered, hypothesis-driven studies capable of translating observed sex and gender differences into improved understanding, diagnosis, prevention, and treatment of OCD.

## Data Availability

All data produced in the present work are contained in the manuscript.

## Declaration of generative AI in scientific writing

During the preparation of this work the authors used ChatGPT in order to improve the readability and language of the manuscript. After using this tool/service, the authors reviewed and edited the content as needed and take full responsibility for the content of the published article.

## Footnotes

1 206 papers were reassigned to a different section of the review over the course of processing.

2 Studies that are reviewed in this section rely on self-reported gender; thus, we use the term ‘gender’ in this section.

3 Studies reviewed in this section generally reported binary male/female categories without clearly distinguishing sex from gender. For consistency with the terminology used in the reviewed literature and throughout this section, we use the term “gender.”

4 Genotyping studies will often confirm reported sex/gender by examining sex chromosomes; since this assesses genotypic sex, rather than self-reported gender, we use the term ‘sex’ in this section.

5 While these studies may have characterized biological sex of study participants, none of them distinguished between participants’ sex and gender. We use term ‘gender’ in this subsection for consistency.

6 Studies that are reviewed in this section rely on self-reported gender; thus, we use the term ‘gender’ in this section.

7 Studies reviewed in this section access animals’ sex; thus, we use the term ‘sex’ in this section.

## References

Abramovitch, A., Pizzagalli, D. A., Geller, D. A., Reuman, L., & Wilhelm, S. (2015). Cigarette smoking in obsessive-compulsive disorder and unaffected parents of OCD patients. European psychiatry, 30(1), 137–144.

Abramovitch, A., Pizzagalli, D. A., Reuman, L., & Wilhelm, S. (2014). Anhedonia in obsessive-compulsive disorder: beyond comorbid depression. Psychiatry research, 216(2), 223–229.

Abramowitz, J. S., Deacon, B. J., Olatunji, B. O., Wheaton, M. G., Berman, N. C., Losardo, D., . . . Adams, T. (2010). Assessment of obsessive-compulsive symptom dimensions: development and evaluation of the Dimensional Obsessive-Compulsive Scale. Psychological assessment, 22(1), 180.

Adams, T. G., Kelmendi, B., Brake, C. A., Gruner, P., Badour, C. L., & Pittenger, C. (2018). The role of stress in the pathogenesis and maintenance of obsessive-compulsive disorder. Chronic Stress, 2, 2470547018758043.

Ade, K. K., Wan, Y., Hamann, H. C., O’Hare, J. K., Guo, W., Quian, A., . . . Wetsel, W. C. (2016). Increased metabotropic glutamate receptor 5 signaling underlies obsessive-compulsive disorder-like behavioral and striatal circuit abnormalities in mice. Biological Psychiatry, 80(7), 522–533.

Akkus, F., Terbeck, S., Ametamey, S. M., Rufer, M., Treyer, V., Burger, C., . . . Buck, A. (2014). Metabotropic glutamate receptor 5 binding in patients with obsessive-compulsive disorder. International journal of neuropsychopharmacology, 17(12), 1915–1922.

Ali, H., Marth, L., & Krueger-Burg, D. (2020). Neuroligin-2 as a central organizer of inhibitory synapses in health and disease. Science Signaling, 13(663), eabd8379.

Altemus, M., Sarvaiya, N., & Epperson, C. N. (2014). Sex differences in anxiety and depression clinical perspectives. Frontiers in neuroendocrinology, 35(3), 320–330.

Angulo, M., Butler, M., & Cataletto, M. (2015). Prader-Willi syndrome: a review of clinical, genetic, and endocrine findings. Journal of endocrinological investigation, 38, 1249–1263.

Arksey, H., & O’Malley, L. (2005). Scoping studies: towards a methodological framework. International journal of social research methodology, 8(1), 19–32.

Arora, T., Bhowmik, M., Khanam, R., & Vohora, D. (2013). Oxcarbazepine and fluoxetine protect against mouse models of obsessive compulsive disorder through modulation of cortical serotonin and CREB pathway. Behavioural Brain Research, 247, 146–152.

Ayati, Z., Sarris, J., & Chang, D. Herbal Medicines and Phytochemicals for Obsessive Compulsive Disorder (OCD).

Bais, M., Figee, M., & Denys, D. (2014). Neuromodulation in obsessive-compulsive disorder. Psychiatric Clinics, 37(3), 393–413.

Bandelow, B., Baldwin, D., Abelli, M., Bolea-Alamanac, B., Bourin, M., Chamberlain, S. R., . . . Fineberg, N. (2017). Biological markers for anxiety disorders, OCD and PTSD: A consensus statement. Part II: Neurochemistry, neurophysiology and neurocognition. The World Journal of Biological Psychiatry, 18(3), 162–214.

Bargiela, S., Steward, R., & Mandy, W. (2016). The experiences of late-diagnosed women with autism spectrum conditions: An investigation of the female autism phenotype. Journal of autism and developmental disorders, 46, 3281–3294.

Bartz, J. A., & Hollander, E. (2006). Is obsessive–compulsive disorder an anxiety disorder? Progress in Neuro-Psychopharmacology and Biological Psychiatry, 30(3), 338–352.

Bauermeister, J. J., Shrout, P. E., Chávez, L., Rubio-Stipec, M., Ramírez, R., Padilla, L., . . . Canino, G. (2007). ADHD and gender: are risks and sequela of ADHD the same for boys and girls? Journal of Child Psychology and Psychiatry, 48(8), 831–839.

Becker, K. R., Breithaupt, L., Lawson, E. A., Eddy, K. T., & Thomas, J. J. (2020). Co-occurrence of avoidant/restrictive food intake disorder and traditional eating psychopathology. Journal of the American Academy of Child & Adolescent Psychiatry, 59(2), 209–212.

Beijers, L., van Loo, H. M., Romeijn, J.-W., Lamers, F., Schoevers, R. A., & Wardenaar, K. J. (2022). Investigating data-driven biological subtypes of psychiatric disorders using specification-curve analysis. Psychological Medicine, 52(6), 1089–1100.

Bekker, M. H., & van Mens-Verhulst, J. (2007). Anxiety disorders: sex differences in prevalence, degree, and background, but gender-neutral treatment. Gender medicine, 4, S178–S193.

Benjet, C., Bromet, E., Karam, E. G., Kessler, R. C., McLaughlin, K. A., Ruscio, A. M., . . . Hill, E. (2016). The epidemiology of traumatic event exposure worldwide: results from the World Mental Health Survey Consortium. Psychological Medicine, 46(2), 327–343.

Benzina, N., Mallet, L., Burguière, E., N’diaye, K., & Pelissolo, A. (2016). Cognitive dysfunction in obsessive-compulsive disorder. Current psychiatry reports, 18, 1–11.

Bloch, M. H., Landeros-Weisenberger, A., Rosario, M. C., Pittenger, C., & Leckman, J. F. (2008). Meta-analysis of the symptom structure of obsessive-compulsive disorder. American Journal of Psychiatry, 165(12), 1532–1542.

Border, R., Johnson, E. C., Evans, L. M., Smolen, A., Berley, N., Sullivan, P. F., & Keller, M. C. (2019). No support for historical candidate gene or candidate gene-by-interaction hypotheses for major depression across multiple large samples. American Journal of Psychiatry, 176(5), 376–387.

Breach, M. R., & Lenz, K. M. (2022). Sex differences in neurodevelopmental disorders: a key role for the immune system. In Sex Differences in Brain Function and Dysfunction (pp. 165-206). Springer.

Briley, P. M., Merlo, S., & Ellis, C. (2022). Sex differences in childhood stuttering and coexisting developmental disorders. Journal of Developmental and Physical Disabilities, 34(3), 505–527.

Brownlow, G., Flack, R., Burton-Murray, H., Palsson, O., & Aziz, I. (2026). The prevalence and burden of avoidant/restrictive food intake disorder (ARFID) symptoms in the adult general population of the UK and USA. International journal of eating disorders, 59(2), 362–370.

Brugha, T. S., Spiers, N., Bankart, J., Cooper, S.-A., McManus, S., Scott, F. J., . . . Tyrer, F. (2016). Epidemiology of autism in adults across age groups and ability levels. The British Journal of Psychiatry, 209(6), 498–503.

Buccheri, R. K., & Sharifi, C. (2017). Critical appraisal tools and reporting guidelines for evidence-based practice. Worldviews on Evidence-Based Nursing, 14(6), 463–472.

Burns, G. L., Keortge, S. G., Formea, G. M., & Sternberger, L. G. (1996). Revision of the Padua Inventory of obsessive compulsive disorder symptoms: Distinctions between worry, obsessions, and compulsions. Behaviour research and therapy, 34(2), 163–173.

Campanelli, F., Laricchiuta, D., Natale, G., Marino, G., Calabrese, V., Picconi, B., . . . Ghiglieri, V. (2021). Long-term shaping of corticostriatal synaptic activity by acute fasting. International Journal of Molecular Sciences, 22(4), 1916.

Canal, C. E., & Morgan, D. (2012). Head-twitch response in rodents induced by the hallucinogen 2, 5-dimethoxy-4-iodoamphetamine: a comprehensive history, a re-evaluation of mechanisms, and its utility as a model. Drug testing and analysis, 4(7-8), 556-576.

Cardona, N. D., Madigan, R. J., & Sauer-Zavala, S. (2022). How minority stress becomes traumatic invalidation: An emotion-focused conceptualization of minority stress in sexual and gender minority people. Clinical Psychology: Science and Practice, 29(2), 185.

Castillejos, M. d. C., Martín-Pérez, C., & Moreno-Küstner, B. (2018). A systematic review and meta-analysis of the incidence of psychotic disorders: the distribution of rates and the influence of gender, urbanicity, immigration and socio-economic level. Psychological Medicine, 48(13), 2101–2115.

Castle, D. J., Deale, A., & Marks, I. M. (1995). Gender differences in obsessive compulsive disorder. Australian & New Zealand Journal of Psychiatry, 29(1), 114–117.

Chamberlain, S. R., Menzies, L., Sahakian, B. J., & Fineberg, N. A. (2007). Lifting the veil on trichotillomania. American Journal of Psychiatry, 164(4), 568–574.

Chavarria, M. C., Sánchez, F. J., Chou, Y.-Y., Thompson, P. M., & Luders, E. (2014). Puberty in the corpus callosum. Neuroscience, 265, 1–8.

Chen, S.-K., Tvrdik, P., Peden, E., Cho, S., Wu, S., Spangrude, G., & Capecchi, M. R. (2010). Hematopoietic origin of pathological grooming in Hoxb8 mutant mice. Cell, 141(5), 775–785.

Cherian, A. V., Narayanaswamy, J. C., Viswanath, B., Guru, N., George, C. M., Math, S. B., . . . Reddy, Y. J. (2014). Gender differences in obsessive-compulsive disorder: findings from a large Indian sample. Asian journal of psychiatry, 9, 17–21.

Citkowska-Kisielewska, A., Rutkowski, K., Sobański, J. A., Dembińska, E., & Mielimąka, M. (2019). Anxiety symptoms in obsessive-compulsive disorder and generalized anxiety disorder. Psychiatria Polska, 53(4), 845–864.

Clemente, M. J., Silva, A. S. M., Pedro, M. O. P., Paiva, H. S., Périco, C. d. A. M., Torales, J., . . . Castaldelli-Maia, J. M. (2022). A meta-analysis and meta-regression analysis of the global prevalence of obsessive-compulsive personality disorder. Heliyon, 8(7).

Cohen, S., Murphy, M. L., & Prather, A. A. (2019). Ten surprising facts about stressful life events and disease risk. Annual review of psychology, 70(1), 577–597.

Coskun, M., Zoroglu, S., & Ozturk, M. (2012). Phenomenology, psychiatric comorbidity and family history in referred preschool children with obsessive-compulsive disorder. Child and Adolescent Psychiatry and Mental Health, 6, 1–9.

da Silva, J. F., Taguchi, L. M., da Silva Leite, E., & de Oliveira, A. R. (2022). Meta-chlorophenylpiperazine-induced behavioral changes in obsessive-compulsive disorder research: A systematic review of rodent studies. Neuroscience, 507, 125–138.

da Silva Prado, H., do Rosário, M. C., Lee, J., Hounie, A. G., Shavitt, R. G., & Miguel, E. C. (2008). Sensory phenomena in obsessive-compulsive disorder and tic disorders: a review of the literature. CNS spectrums, 13(5), 425–432.

De Hert, M., Cohen, D., Bobes, J., Cetkovich-Bakmas, M., Leucht, S., Ndetei, D. M., . . . Moeller, H.-J. (2011). Physical illness in patients with severe mental disorders. II. Barriers to care, monitoring and treatment guidelines, plus recommendations at the system and individual level. World psychiatry, 10(2), 138.

De Hert, M., Correll, C. U., Bobes, J., Cetkovich-Bakmas, M., Cohen, D., Asai, I., . . . Ndetei, D. M. (2011). Physical illness in patients with severe mental disorders. I. Prevalence, impact of medications and disparities in health care. World psychiatry, 10(1), 52.

De Hert, M., Dekker, J., Wood, D., Kahl, K., Holt, R., & Möller, H.-J. (2009). Cardiovascular disease and diabetes in people with severe mental illness position statement from the European Psychiatric Association (EPA), supported by the European Association for the Study of Diabetes (EASD) and the European Society of Cardiology (ESC). European psychiatry, 24(6), 412–424.

de Kluiver, H., Buizer-Voskamp, J. E., Dolan, C. V., & Boomsma, D. I. (2017). Paternal age and psychiatric disorders: A review. American Journal of Medical Genetics Part B: Neuropsychiatric Genetics, 174(3), 202–213.

Denys, D., Zohar, J., & Westenberg, H. (2004). The role of dopamine in obsessive-compulsive disorder: preclinical and clinical evidence. J Clin Psychiatry, 65(Suppl 14), 11–17.

Diniz, J. B., Rosario-Campos, M. C., Shavitt, R. G., Curi, M., Hounie, A. G., Brotto, S. A., & Miguel, E. C. (2004). Impact of age at onset and duration of illness on the expression of comorbidities in obsessive-compulsive disorder. The Journal of clinical psychiatry.

Dirkx, N., Miceli, F., Taglialatela, M., & Weckhuysen, S. (2020). The role of Kv7. 2 in neurodevelopment: insights and gaps in our understanding. Frontiers in Physiology, 11, 570588.

Drasch, K., & Matthes, B. (2013). Improving retrospective life course data by combining modularized self-reports and event history calendars: experiences from a large scale survey. Quality & Quantity, 47(2), 817–838.

Dykens, E. M. (1999). Prader-Willi syndrome: Toward a behavioral phenotype.

Ecker, W., & Gönner, S. (2008). Incompleteness and harm avoidance in OCD symptom dimensions. Behaviour research and therapy, 46(8), 895–904.

Eichstedt, J. A., & Arnold, S. L. (2001). Childhood-onset obsessive-compulsive disorder: a tic-related subtype of OCD? Clinical psychology review, 21(1), 137–157.

Eliot, L. (2019). Neurosexism: the myth that men and women have different brains. Nature, 566(7745), 453–455.

Faheem, M., Akram, W., Akram, H., Khan, M. A., Siddiqui, F. A., & Majeed, I. (2022). Gender-based differences in prevalence and effects of ADHD in adults: A systematic review. Asian journal of psychiatry, 75, 103205.

Fairweather, D., Frisancho-Kiss, S., & Rose, N. R. (2008). Sex differences in autoimmune disease from a pathological perspective. The American journal of pathology, 173(3), 600–609.

Fantegrossi, W. E., Simoneau, J., Cohen, M., Zimmerman, S., Henson, C., Rice, K., & Woods, J. (2010). Interaction of 5-HT2A and 5-HT2C receptors in R (−)-2, 5-dimethoxy-4-iodoamphetamine-elicited head twitch behavior in mice. The Journal of pharmacology and experimental therapeutics, 335(3), 728-734.

Faravelli, C., Sauro, C. L., Godini, L., Lelli, L., Benni, L., Pietrini, F., . . . Ricca, V. (2012). Childhood stressful events, HPA axis and anxiety disorders. World journal of psychiatry, 2(1), 13.

Farrell, M., Werge, T., Sklar, P., Owen, M. J., Ophoff, R., O’Donovan, M. C., . . . Sullivan, P. F. (2015). Evaluating historical candidate genes for schizophrenia. Mol Psychiatry, 20(5), 555–562.

Fawcett, E. J., Power, H., & Fawcett, J. M. (2020). Women are at greater risk of OCD than men: a meta-analytic review of OCD prevalence worldwide. The Journal of clinical psychiatry, 81(4), 13075.

Ferrao, Y., De Alvarenga, P. G., Hounie, A. G., De Mathis, M. A., De Rosario, M. C., & Miguel, E. C. (2013). The phenomenology of obsessive-compulsive symptoms in Tourette syndrome. In Tourette syndrome (pp. 45–64). Oxford University Press, New York.

Fonseca, F., Robles-Martínez, M., Tirado-Muñoz, J., Alías-Ferri, M., Mestre-Pintó, J.-I., Coratu, A. M., & Torrens, M. (2021). A gender perspective of addictive disorders. Current addiction reports, 8, 89–99.

Fontenelle, L. F., Cocchi, L., Harrison, B. J., Shavitt, R. G., do Rosário, M. C., Ferrão, Y. A., . . . Pantelis, C. (2012). Towards a post-traumatic subtype of obsessive–compulsive disorder. Journal of anxiety disorders, 26(2), 377–383.

Forman, S. F., McKenzie, N., Hehn, R., Monge, M. C., Kapphahn, C. J., Mammel, K. A., . . . Romano, M. (2014). Predictors of outcome at 1 year in adolescents with DSM-5 restrictive eating disorders: report of the national eating disorders quality improvement collaborative. Journal of Adolescent Health, 55(6), 750–756.

Franceschini, A., & Fattore, L. (2021). Gender-specific approach in psychiatric diseases: because sex matters. European journal of pharmacology, 896, 173895.

Franklin, M. E., Harrison, J. P., & Benavides, K. L. (2012). Obsessive-compulsive and tic-related disorders. Child and Adolescent Psychiatric Clinics, 21(3), 555–571.

Friborg, O., Martinussen, M., Kaiser, S., Øvergård, K. T., & Rosenvinge, J. H. (2013). Comorbidity of personality disorders in anxiety disorders: A meta-analysis of 30 years of research. Journal of Affective Disorders, 145(2), 143--155. 10.1016/j.jad.2012.07.004, pmid = 22999891

Friedlander, L., & Desrocher, M. (2006). Neuroimaging studies of obsessive–compulsive disorder in adults and children. Clinical psychology review, 26(1), 32–49.

Gagnon, M., Singer, I., Morand-Beaulieu, S., O’Connor, K. P., Gauthier, B., Woods, D. W., . . . Leclerc, J. B. (2024). Sex Differences in Youth with Chronic Tic Disorder and Tourette Syndrome: Evaluation of Tic Severity, Psychological Profiles, and Quality of Life. Journal of Clinical Medicine, 13(9), 2477.

Gamal El-Din, T. M., Lantin, T., Tschumi, C. W., Juarez, B., Quinlan, M., Hayano, J. H., . . . Catterall, W. A. (2021). Autism-associated mutations in KV7 channels induce gating pore current. Proceedings of the National Academy of Sciences, 118(45), e2112666118.

Geller, D. A., Biederman, J., Griffin, S., Jones, J., & Lefkowitz, T. R. (1996). Comorbidity of juvenile obsessive-compulsive disorder with disruptive behavior disorders. Journal of the American Academy of Child & Adolescent Psychiatry, 35(12), 1637–1646.

Geller, D. A., Biederman, J., Jones, J., Shapiro, S., Schwartz, S., & Park, K. S. (1998). Obsessive-compulsive disorder in children and adolescents: a review. Harvard review of psychiatry, 5(5), 260–273.

Geller, D. A., Coffey, B., Faraone, S., Hagermoser, L., Zaman, N. K., Farrell, C. L., . . . Biederman, J. (2003). Does comorbid attention-deficit/hyperactivity disorder impact the clinical expression of pediatric obsessive-compulsive disorder? CNS spectrums, 8(4), 259–264.

Gemmati, D., Varani, K., Bramanti, B., Piva, R., Bonaccorsi, G., Trentini, A., . . . Bellini, T. (2019). “Bridging the gap” everything that could have been avoided if we had applied gender medicine, pharmacogenetics and personalized medicine in the gender-omics and sex-omics era. International Journal of Molecular Sciences, 21(1), 296.

Giarelli, E., Wiggins, L. D., Rice, C. E., Levy, S. E., Kirby, R. S., Pinto-Martin, J., & Mandell, D. (2010). Sex differences in the evaluation and diagnosis of autism spectrum disorders among children. Disability and health journal, 3(2), 107–116.

Goodman, W. K., Price, L. H., Rasmussen, S. A., Mazure, C., Delgado, P., Heninger, G. R., & Charney, D. S. (1989). The yale-brown obsessive compulsive scale: II. Validity. Archives of general psychiatry, 46(11), 1012–1016.

Goodman, W. K., Price, L. H., Rasmussen, S. A., Mazure, C., Fleischmann, R. L., Hill, C. L., . . . Charney, D. S. (1989). The Yale-Brown obsessive compulsive scale: I. Development, use, and reliability. Archives of general psychiatry, 46(11), 1006–1011.

Gordon, J. L., Girdler, S. S., Meltzer-Brody, S. E., Stika, C. S., Thurston, R. C., Clark, C. T., . . . Wisner, K. L. (2015). Ovarian hormone fluctuation, neurosteroids, and HPA axis dysregulation in perimenopausal depression: a novel heuristic model. American Journal of Psychiatry, 172(3), 227–236.

Graf, E. R., Zhang, X., Jin, S.-X., Linhoff, M. W., & Craig, A. M. (2004). Neurexins induce differentiation of GABA and glutamate postsynaptic specializations via neuroligins. Cell, 119(7), 1013–1026.

Graf, M., Kantor, S., Anheuer, Z. E., Modos, E. A., & Bagdy, G. (2003). m-CPP-induced self-grooming is mediated by 5-HT2C receptors. Behavioural brain research, 142(1-2), 175–179.

Grassi, G., Cecchelli, C., Mazzocato, G., & Vignozzi, L. (2022). Early onset obsessive–compulsive disorder: the biological and clinical phenotype. CNS spectrums, 27(4), 421–427.

Grenier, S., Payette, M. C., Gunther, B., Askari, S., Desjardins, F. F., Raymond, B., & Berbiche, D. (2019). Association of age and gender with anxiety disorders in older adults: A systematic review and meta-analysis. International journal of geriatric psychiatry, 34(3), 397–407.

Grissom, N. M., & Reyes, T. M. (2019). Let’s call the whole thing off: evaluating gender and sex differences in executive function. Neuropsychopharmacology, 44(1), 86–96.

Gröhn, C., Norgren, E., & Eriksson, L. (2022). A systematic review of the neural correlates of multisensory integration in schizophrenia. Schizophrenia Research: Cognition, 27, 100219.

Guglielmi, V., Vulink, N. C., Denys, D., Wang, Y., Samuels, J. F., & Nestadt, G. (2014). Obsessive– compulsive disorder and female reproductive cycle events: Results from the OCD and reproduction collaborative study. Depression and anxiety, 31(12), 979–987.

Guo, X., Meng, Z., Huang, G., Fan, J., Zhou, W., Ling, W., . . . Su, L. (2016). Meta-analysis of the prevalence of anxiety disorders in mainland China from 2000 to 2015. Scientific reports, 6(1), 1–15.

Gustafsson, K., Lindfors, P., Aronsson, G., & Lundberg, U. (2008). Relationships between self-rating of recovery from work and morning salivary cortisol. Journal of occupational health, 50(1), 24–30.

Gustafsson, P. E., Gustafsson, P. A., Ivarsson, T., & Nelson, N. (2008). Diurnal cortisol levels and cortisol response in youths with obsessive-compulsive disorder. Neuropsychobiology, 57(1-2), 14–21.

Hacıömeroğlu, A. B. (2008). Perceived parental rearing behaviors, responsibility attitudes and life events as predictors of obsessive compulsive symptomatology: Test of a cognitive model.

Hageman, S. B., van Rooijen, G., Bergfeld, I. O., Schirmbeck, F., de Koning, P., Schuurman, P. R., & Denys, D. (2021). Deep brain stimulation versus ablative surgery for treatment-refractory obsessive-compulsive disorder: a meta-analysis. Acta Psychiatrica Scandinavica, 143(4), 307–318.

Haghshomar, M., Mirghaderi, S. P., Shobeiri, P., James, A., & Zarei, M. (2023). White matter abnormalities in paediatric obsessive–compulsive disorder: a systematic review of diffusion tensor imaging studies. Brain Imaging and Behavior, 17(3), 343–366.

Harald, B., & Gordon, P. (2012). Meta-review of depressive subtyping models. Journal of affective disorders, 139(2), 126–140.

Heck, A. L., & Handa, R. J. (2019). Sex differences in the hypothalamic–pituitary–adrenal axis’ response to stress: an important role for gonadal hormones. Neuropsychopharmacology, 44(1), 45–58.

Heinrichs, M., von Dawans, B., & Domes, G. (2009). Oxytocin, vasopressin, and human social behavior. Frontiers in neuroendocrinology, 30(4), 548–557.

Hesse, S., Müller, U., Lincke, T., Barthel, H., Villmann, T., Angermeyer, M. C., . . . Stengler-Wenzke, K. (2005). Serotonin and dopamine transporter imaging in patients with obsessive–compulsive disorder. Psychiatry Research: Neuroimaging, 140(1), 63–72.

Hidalgo-Lopez, E., & Pletzer, B. (2019). Individual differences in the effect of menstrual cycle on basal ganglia inhibitory control. Scientific Reports, 9(1), 1–11.

Higgins, S. T., Kurti, A. N., Redner, R., White, T. J., Gaalema, D. E., Roberts, M. E., . . . Stanton, C. A. (2015). A literature review on prevalence of gender differences and intersections with other vulnerabilities to tobacco use in the United States, 2004–2014. Preventive medicine, 80, 89-100.

Hinkley, L. B., Marco, E. J., Findlay, A. M., Honma, S., Jeremy, R. J., Strominger, Z., . . . Paul, L. K. (2012). The role of corpus callosum development in functional connectivity and cognitive processing.

Hoek, H. W. (2006). Incidence, prevalence and mortality of anorexia nervosa and other eating disorders. Current opinion in psychiatry, 19(4), 389–394.

Holdcroft, A. (2007). Gender bias in research: how does it affect evidence based medicine? In (Vol. 100, pp. 2–3): SAGE Publications Sage UK: London, England.

Holländer, A., Hausmann, M., Hamm, J. P., & Corballis, M. C. (2005). Sex hormonal modulation of hemispheric asymmetries in the attentional blink. Journal of the International Neuropsychological Society, 11(3), 263–272.

Holzer, J. C., McDougle, C. J., Boyarsky, B. K., Price, L. H., Goodman, W. K., Baer, L., & Leckman, J. F. (1994). Obsessive–compulsive disorder with and without a chronic tic disorder: A comparison of symptoms in 70 patients. The British Journal of Psychiatry, 164(4), 469–473.

Hsiao, M.-C., Lin, K.-J., Liu, C.-Y., & Schatz, D. B. (2013). The interaction between dopamine transporter function, gender differences, and possible laterality in depression. Psychiatry Research: Neuroimaging, 211(1), 72–77.

Hühne, V., dos Santos-Ribeiro, S., Moreira-de-Oliveira, M. E., de Menezes, G. B., & Fontenelle, L. (2024). Towards the correlates of Stressful Life Events as precipitants of OCD: a systematic review and metanalysis. CNS spectrums, 1-30.

Hyman, B. M., & Pedrick, C. (2010). The OCD workbook: Your guide to breaking free from obsessive-compulsive disorder. New Harbinger Publications.

J Kuss, D., D Griffiths, M., Karila, L., & Billieux, J. (2014). Internet addiction: A systematic review of epidemiological research for the last decade. Current pharmaceutical design, 20(25), 4026–4052.

Jacquemont, S., Coe, B. P., Hersch, M., Duyzend, M. H., Krumm, N., Bergmann, S., . . . Eichler, E. E. (2014). A higher mutational burden in females supports a “female protective model” in neurodevelopmental disorders. The American Journal of Human Genetics, 94(3), 415–425.

Jagiellowicz, J., Xu, X., Aron, A., Aron, E., Cao, G., Feng, T., & Weng, X. (2011). The trait of sensory processing sensitivity and neural responses to changes in visual scenes. Social cognitive and affective neuroscience, 6(1), 38–47.

Joel, D., Berman, Z., Tavor, I., Wexler, N., Gaber, O., Stein, Y., . . . Margulies, D. S. (2015). Sex beyond the genitalia: The human brain mosaic. Proceedings of the National Academy of Sciences, 112(50), 15468–15473.

Kajimoto, Y., Shirakawa, O., Lin, X.-H., Hashimoto, T., Kitamura, N., Murakami, N., . . . Maeda, K. (2003). Synapse-associated protein 90/postsynaptic density-95-associated protein (SAPAP) is expressed differentially in phencyclidine-treated rats and is increased in the nucleus accumbens of patients with schizophrenia. Neuropsychopharmacology, 28(10), 1831–1839.

Karthik, S., Sharma, L. P., & Narayanaswamy, J. C. (2020). Investigating the role of glutamate in obsessive-compulsive disorder: current perspectives. Neuropsychiatric disease and treatment, 1003-1013.

Keery, H., LeMay-Russell, S., Barnes, T. L., Eckhardt, S., Peterson, C. B., Lesser, J., . . . Le Grange, D. (2019). Attributes of children and adolescents with avoidant/restrictive food intake disorder. Journal of eating disorders, 7(1), 31.

Kennedy, N., Boydell, J., Kalidindi, S., Fearon, P., Jones, P. B., van Os, J., & Murray, R. M. (2005). Gender differences in incidence and age at onset of mania and bipolar disorder over a 35-year period in Camberwell, England. American Journal of Psychiatry, 162(2), 257–262.

Kessing, L. V. (2004). Gender differences in the phenomenology of bipolar disorder. Bipolar Disorders, 6(5), 421–425.

Kessler, R. C., Berglund, P., Demler, O., Jin, R., Merikangas, K. R., & Walters, E. E. (2005). Lifetime prevalence and age-of-onset distributions of DSM-IV disorders in the National Comorbidity Survey Replication. Archives of general psychiatry, 62(6), 593–602.

Kessler, R. C., Petukhova, M., Sampson, N. A., Zaslavsky, A. M., & Wittchen, H. U. (2012). Twelve-month and lifetime prevalence and lifetime morbid risk of anxiety and mood disorders in the United States. International journal of methods in psychiatric research, 21(3), 169–184.

Khan, I. N. (2012). Sex differences and the effects of sex hormones on the structure of the corpus callosum University of Nottingham].

Kheloui, S., Brouillard, A., Rossi, M., Marin, M.-F., Mendrek, A., Paquette, D., & Juster, R.-P. (2021). Exploring the sex and gender correlates of cognitive sex differences. Acta psychologica, 221, 103452.

Kirkpatrick, S. L., Goldberg, L. R., Yazdani, N., Babbs, R. K., Wu, J., Reed, E. R., . . . Luttik, K. P. (2017). Cytoplasmic FMR1-interacting protein 2 is a major genetic factor underlying binge eating. Biological Psychiatry, 81(9), 757–769.

Kirov, G., Pocklington, A., Holmans, P., Ivanov, D., Ikeda, M., Ruderfer, D., . . . Georgieva, L. (2012). De novo CNV analysis implicates specific abnormalities of postsynaptic signalling complexes in the pathogenesis of schizophrenia. Molecular Psychiatry, 17(2), 142–153.

Klein, D. N., Glenn, C. R., Kosty, D. B., Seeley, J. R., Rohde, P., & Lewinsohn, P. M. (2013). Predictors of first lifetime onset of major depressive disorder in young adulthood. Journal of abnormal psychology, 122(1), 1.

Knight, T., Steeves, T., Day, L., Lowerison, M., Jette, N., & Pringsheim, T. (2012). Prevalence of tic disorders: a systematic review and meta-analysis. Pediatric neurology, 47(2), 77–90.

Koran, L. M., Abujaoude, E., Large, M. D., & Serpe, R. T. (2008). The prevalence of body dysmorphic disorder in the United States adult population. CNS spectrums, 13(4), 316–322.

Kühn, S., & Gallinat, J. (2013). Gray matter correlates of posttraumatic stress disorder: a quantitative meta-analysis. Biological Psychiatry, 73(1), 70–74.

Kuygun Karcı, C., & Gül Celik, G. (2020). Nutritional and herbal supplements in the treatment of obsessive compulsive disorder. General psychiatry, 33(2), e100159.

Laakso, A., Vilkman, H., Örgen Bergman, J., Haaparanta, M., Solin, O., Syvälahti, E., . . . Hietala, J. (2002). Sex differences in striatal presynaptic dopamine synthesis capacity in healthy subjects. Biological Psychiatry, 52(7), 759–763.

Labad, J., Menchon, J. M., Alonso, P., Segalas, C., Jimenez, S., Jaurrieta, N., . . . Vallejo, J. (2008). Gender differences in obsessive–compulsive symptom dimensions. Depression and anxiety, 25(10), 832–838.

Labad, J., Soria, V., Salvat-Pujol, N., Segalàs, C., Real, E., Urretavizcaya, M., . . . Jiménez-Murcia, S. (2018). Hypothalamic-pituitary-adrenal axis activity in the comorbidity between obsessive-compulsive disorder and major depression. Psychoneuroendocrinology, 93, 20–28.

Lauritano, A., Moutton, S., Longobardi, E., Tran Mau-Them, F., Laudati, G., Nappi, P., . . . Jouan, T. (2019). A novel homozygous KCNQ3 loss-of-function variant causes non-syndromic intellectual disability and neonatal-onset pharmacodependent epilepsy. Epilepsia Open, 4(3), 464–475.

Leclercq, V., & Corvol, J.-C. (2024). Impulse control disorder: Review on clinical, pharmacologic, and genetic risk factors. Revue Neurologique.

Lee, H.-J., & Kwon, S.-M. (2003). Two different types of obsession: autogenous obsessions and reactive obsessions. Behaviour research and therapy, 41(1), 11–29.

Lee, H. J., Kwon, S. M., Kwon, J. S., & Telch, M. J. (2005). Testing the autogenous–reactive model of obsessions. Depression and anxiety, 21(3), 118–129.

Lee, H. K. (2024). Sex/Gender Differences in Addictive Disorders. In Sex/Gender-Specific Medicine in Clinical Areas (pp. 381-388). Springer.

Lee, J. J., Ham, J. H., Lee, P. H., & Sohn, Y. H. (2015). Gender differences in age-related striatal dopamine depletion in Parkinson’s disease. Journal of movement disorders, 8(3), 130.

Lehavot, K., Katon, J. G., Chen, J. A., Fortney, J. C., & Simpson, T. L. (2018). Post-traumatic stress disorder by gender and veteran status. American Journal of Preventive Medicine, 54(1), e1–e9.

Lehman, A., Thouta, S., Mancini, G. M., Naidu, S., van Slegtenhorst, M., McWalter, K., . . . Adam, S. (2017). Loss-of-function and gain-of-function mutations in KCNQ5 cause intellectual disability or epileptic encephalopathy. The American Journal of Human Genetics, 101(1), 65–74.

Lewinsohn, P. M., Gotlib, I. H., Lewinsohn, M., Seeley, J. R., & Allen, N. B. (1998). Gender differences in anxiety disorders and anxiety symptoms in adolescents. Journal of abnormal psychology, 107(1), 109.

Li, L., Wu, M., Liao, Y., Ouyang, L., Du, M., Lei, D., . . . Gong, Q. (2014). Grey matter reduction associated with posttraumatic stress disorder and traumatic stress. Neuroscience & Biobehavioral Reviews, 43, 163–172.

Lindqvist, A., Sendén, M. G., & Renström, E. A. (2021). What is gender, anyway: a review of the options for operationalising gender. Psychology & sexuality, 12(4), 332–344.

Lochner, C., Hemmings, S. M., Kinnear, C. J., Moolman-Smook, J. C., Corfield, V. A., Knowles, J. A., . . . Stein, D. J. (2004). Gender in obsessive–compulsive disorder: clinical and genetic findings. European Neuropsychopharmacology, 14(2), 105–113.

Lochner, C., & Stein, D. J. (2001). Gender in obsessive-compulsive disorder and obsessive-compulsive spectrum disorders. Archives of Women’s Mental Health, 4(1), 19–26.

Lochner, C., & Stein, D. J. (2003). Heterogeneity of obsessive-compulsive disorder: a literature review. Harvard review of psychiatry, 11(3), 113–132.

Long, H. A., French, D. P., & Brooks, J. M. (2020). Optimising the value of the critical appraisal skills programme (CASP) tool for quality appraisal in qualitative evidence synthesis. Research Methods in Medicine & Health Sciences, 1(1), 31–42.

Loomes, R., Hull, L., & Mandy, W. P. L. (2017). What is the male-to-female ratio in autism spectrum disorder? A systematic review and meta-analysis. Journal of the American Academy of Child & Adolescent Psychiatry, 56(6), 466–474.

Mar-Barrutia, L., Real, E., Segalas, C., Bertolin, S., Menchon, J. M., & Alonso, P. (2021). Deep brain stimulation for obsessive-compulsive disorder: A systematic review of worldwide experience after 20 years. World journal of psychiatry, 11(9), 659.

Maraz, A., Griffiths, M. D., & Demetrovics, Z. (2016). The prevalence of compulsive buying: a meta-analysis. Addiction, 111(3), 408–419.

Marazziti, D., Hollander, E., Lensi, P., Ravagli, S., & Cassano, G. B. (1992). Peripheral markers of serotonin and dopamine function in obsessive-compulsive disorder. Psychiatry research, 42(1), 41–51.

Markarian, Y., Larson, M. J., Aldea, M. A., Baldwin, S. A., Good, D., Berkeljon, A., . . . McKay, D. (2010). Multiple pathways to functional impairment in obsessive–compulsive disorder. Clinical psychology review, 30(1), 78–88.

Maroufi, S. F., Fallahi, M. S., Kotecha, R., Fariselli, L., Gorgulho, A., Levivier, M., . . . Suh, J. H. (2026). Stereotactic Radiosurgery Capsulotomy for Intractable Severe Obsessive-Compulsive Disorder: A Systematic Review, Meta-Analysis, and International Stereotactic Radiosurgery Society Practice Guidelines. Neurosurgery, 99(2), 265–276.

Masi, G., Millepiedi, S., Perugi, G., Pfanner, C., Berloffa, S., Pari, C., . . . Akiskal, H. S. (2010). A naturalistic exploratory study of the impact of demographic, phenotypic and comorbid features in pediatric obsessive-compulsive disorder. Psychopathology, 43(2), 69–78.

Mathes, B. M., Morabito, D. M., & Schmidt, N. B. (2019). Epidemiological and clinical gender differences in OCD. Current psychiatry reports, 21(5), 1–7.

Mathis, M. A. d., Alvarenga, P. d., Funaro, G., Torresan, R. C., Moraes, I., Torres, A. R., . . . Hounie, A. G. (2011). Gender differences in obsessive-compulsive disorder: a literature review. Brazilian Journal of Psychiatry, 33(4), 390–399.

Mattheisen, M., Samuels, J. F., Wang, Y., Greenberg, B. D., Fyer, A. J., McCracken, J. T., . . . Grados, M. A. (2015). Genome-wide association study in obsessive-compulsive disorder: results from the OCGAS. Molecular Psychiatry, 20(3), 337–344.

Maughan, B., Rowe, R., Messer, J., Goodman, R., & Meltzer, H. (2004). Conduct disorder and oppositional defiant disorder in a national sample: developmental epidemiology. Journal of Child Psychology and Psychiatry, 45(3), 609–621.

Meadows, S. O., Land, K. C., & Lamb, V. L. (2005). Assessing Gilligan vs. Sommers: Gender-specific trends in child and youth well-being in the United States, 1985-2001. Social Indicators Research, 1-52.

Mendrek, A. (2015). Is it important to consider sex and gender in neurocognitive studies? Frontiers in psychiatry, 6, 83.

Miller, L. R., Marks, C., Becker, J. B., Hurn, P. D., Chen, W.-J., Woodruff, T., . . . Wetherington, C. L. (2016). Considering sex as a biological variable in preclinical research. The FASEB Journal, 31(1), 29.

Millet, B., Chabane, N., Delorme, R., Leboyer, M., Leroy, S., Poirier, M. F., . . . Loo, H. (2003). Association between the dopamine receptor D4 (DRD4) gene and obsessive-compulsive disorder. American Journal of Medical Genetics Part B: Neuropsychiatric Genetics, 116(1), 55–59.

Montgomery, S. A. (1993). Obsessive compulsive disorder is not an anxiety disorder. International clinical psychopharmacology, 8, 57–62.

Mosca, E., Bersanelli, M., Gnocchi, M., Moscatelli, M., Castellani, G., Milanesi, L., & Mezzelani, A. (2017). Network diffusion-based prioritization of autism risk genes identifies significantly connected gene modules. Frontiers in Genetics, 8, 129.

Murray, C. J., Lopez, A. D., & Organization, W. H. (1996). The global burden of disease: a comprehensive assessment of mortality and disability from diseases, injuries, and risk factors in 1990 and projected to 2020: summary. World Health Organization.

Nagarajan, N., Jones, B. W., West, P. J., Marc, R., & Capecchi, M. (2018). Corticostriatal circuit defects in Hoxb8 mutant mice. Molecular Psychiatry, 23(9), 1868–1877.

Nakatani, E., Krebs, G., Micali, N., Turner, C., Heyman, I., & Mataix-Cols, D. (2011). Children with very early onset obsessive-compulsive disorder: Clinical features and treatment outcome. Journal of Child Psychology and Psychiatry, 52(12), 1261–1268.

Neal, M., & Cavanna, A. E. (2013). “Not just right experiences” in patients with Tourette syndrome: complex motor tics or compulsions? Psychiatry research, 210(2), 559–563.

Neumann, I. D. (2002). Involvement of the brain oxytocin system in stress coping: interactions with the hypothalamo-pituitary-adrenal axis. Progress in brain research, 139, 147–162.

Neumann, I. D. (2008). Brain oxytocin: a key regulator of emotional and social behaviours in both females and males. Journal of neuroendocrinology, 20(6), 858–865.

NIH, N. I. o. H. (2015). Consideration of sex as a biological variable in NIH-funded Research, 2015.. https://grants.nih.gov/grants/guide/notice-files/NOT-OD-15-102.html

Nordahl, C. W. (2023). Why do we need sex-balanced studies of autism? Autism Research, 16(9), 1662–1669.

Ochoa, S., Usall, J., Cobo, J., Labad, X., & Kulkarni, J. (2012). Gender differences in schizophrenia and first-episode psychosis: A comprehensive literature review. Schizophrenia research and treatment, 2012(1), 916198.

Ohayon, M. M., & Schatzberg, A. F. (2010). Social phobia and depression: prevalence and comorbidity. Journal of Psychosomatic Research, 68(3), 235–243.

Olver, J. S., O’Keefe, G., Jones, G. R., Burrows, G. D., Tochon-Danguy, H. J., Ackermann, U., . . . Norman, T. R. (2009). Dopamine D1 receptor binding in the striatum of patients with obsessive– compulsive disorder. Journal of affective disorders, 114(1-3), 321–326.

Page, M. J., McKenzie, J. E., Bossuyt, P. M., Boutron, I., Hoffmann, T. C., Mulrow, C. D., . . . Brennan, S. E. (2021). The PRISMA 2020 statement: an updated guideline for reporting systematic reviews. International journal of surgery, 88, 105906.

Pantalone, D. W., Gorman, K. R., Pereida, E. T., & Valentine, S. E. (2020). Trauma among sexual and gender minority populations. Rothblum,(Ed.), The Oxford handbook of sexual and gender minority mental health, 59-71.

Parial, S. (2015). Bipolar disorder in women. Indian Journal of Psychiatry, 57(Suppl 2), S252–S263.

Pathania, M., Davenport, E., Muir, J., Sheehan, D., López-Doménech, G., & Kittler, J. (2014). The autism and schizophrenia associated gene CYFIP1 is critical for the maintenance of dendritic complexity and the stabilization of mature spines. Translational Psychiatry, 4(3), e374–e374.

Payne, J. L., Palmer, J. T., & Joffe, H. (2009). A reproductive subtype of depression: conceptualizing models and moving toward etiology. Harvard review of psychiatry, 17(2), 72–86.

Peebles, R., Wilson, J. L., & Lock, J. D. (2006). How do children with eating disorders differ from adolescents with eating disorders at initial evaluation? Journal of Adolescent Health, 39(6), 800–805.

Peelen, M. J., Kazemier, B. M., Ravelli, A. C., De Groot, C. J., Van Der Post, J. A., Mol, B. W., . . . Kok, M. (2016). Impact of fetal gender on the risk of preterm birth, a national cohort study. Acta obstetricia et gynecologica Scandinavica, 95(9), 1034–1041.

Pinares-Garcia, P., Stratikopoulos, M., Zagato, A., Loke, H., & Lee, J. (2018). Sex: a significant risk factor for neurodevelopmental and neurodegenerative disorders. Brain Sciences, 8(8), 154.

Pittenger, C. (2015). Animal models of Tourette syndrome and obsessive-compulsive disorder. In Movement Disorders (pp. 747-764). Elsevier.

Pittenger, C. (2021). Pharmacotherapeutic strategies and new targets in OCD. The neurobiology and treatment of OCD: Accelerating progress, 331-384.

Pittenger, C. (2023). The Pharmacological Treatment of Obsessive-Compulsive Disorder. Psychiatric Clinics, 46(1), 107–119.

Pohjalainen, T., Rinne, J. O., Någren, K., SyvÄlahti, E., & Hietala, J. (1998). Sex differences in the striatal dopamine D2 receptor binding characteristics in vivo. American Journal of Psychiatry, 155(6), 768–773.

Poletti, M., Gebhardt, E., Pelizza, L., Preti, A., & Raballo, A. (2023). Neurodevelopmental antecedents and sensory phenomena in obsessive compulsive disorder: a systematic review supporting a phenomenological-developmental model. Psychopathology, 56(4), 295–305.

Postlethwaite, A., Kellett, S., & Mataix-Cols, D. (2019). Prevalence of hoarding disorder: A systematic review and meta-analysis. Journal of affective disorders, 256, 309–316.

Poulopoulos, A., Aramuni, G., Meyer, G., Soykan, T., Hoon, M., Papadopoulos, T., . . . Harvey, K. (2009). Neuroligin 2 drives postsynaptic assembly at perisomatic inhibitory synapses through gephyrin and collybistin. Neuron, 63(5), 628–642.

Quinn, J. J., Hitchcott, P. K., Umeda, E. A., Arnold, A. P., & Taylor, J. R. (2007). Sex chromosome complement regulates habit formation. Nature neuroscience, 10(11), 1398–1400.

Rasmussen, A. H., Rasmussen, H. B., & Silahtaroglu, A. (2017). The DLGAP family: neuronal expression, function and role in brain disorders. Molecular brain, 10, 1–13.

Reid, J. E., Laws, K. R., Drummond, L., Vismara, M., Grancini, B., Mpavaenda, D., & Fineberg, N. A. (2021). Cognitive behavioural therapy with exposure and response prevention in the treatment of obsessive-compulsive disorder: A systematic review and meta-analysis of randomised controlled trials. Comprehensive psychiatry, 106, 152223.

Rosenberg, D. R., & Keshavan, M. S. (1998). Toward a neurodevelopmental model of obsessive– compulsive disorder. Biological Psychiatry, 43(9), 623–640.

Rosenman, R., Tennekoon, V., & Hill, L. G. (2011). Measuring bias in self-reported data. International journal of behavioural & healthcare research, 2(4), 320.

Rowsell, M., & Francis, S. E. (2015). OCD subtypes: Which, if any, are valid? Clinical Psychology: Science and Practice, 22(4), 414.

Ruigrok, A. N., Salimi-Khorshidi, G., Lai, M.-C., Baron-Cohen, S., Lombardo, M. V., Tait, R. J., & Suckling, J. (2014). A meta-analysis of sex differences in human brain structure. Neuroscience & Biobehavioral Reviews, 39, 34–50.

Sader, M., Harris, H. A., Waiter, G. D., Jackson, M. C., Voortman, T., Jansen, P. W., & Williams, J. H. (2023). Prevalence and characterization of avoidant restrictive food intake disorder in a pediatric population. JAACAP open, 1(2), 116–127.

Salari, N., Hosseinian-Far, A., Jalali, R., Vaisi-Raygani, A., Rasoulpoor, S., Mohammadi, M., . . . Khaledi-Paveh, B. (2020). Prevalence of stress, anxiety, depression among the general population during the COVID-19 pandemic: a systematic review and meta-analysis. Globalization and health, 16, 1–11.

Saxena, S., Funk, M., & Chisholm, D. (2015). Comprehensive mental health action plan 2013–2020. EMHJ-Eastern Mediterranean Health Journal, 21(7), 461–463.

Saxena, S., & Rauch, S. L. (2022). Functional neuroimaging and the neuroanatomy of obsessive-compulsive disorder. Obsessive-Compulsive Disorder and Tourette’s Syndrome, 159-182.

Schein, J., Houle, C., Urganus, A., Cloutier, M., Patterson-Lomba, O., Wang, Y., . . . Lefebvre, P. (2021). Prevalence of post-traumatic stress disorder in the United States: a systematic literature review. Current medical research and opinion, 37(12), 2151–2161.

Scott, K. M., Bruffaerts, R., Tsang, A., Ormel, J., Alonso, J., Angermeyer, M. C., . . . De Graaf, R. (2007). Depression–anxiety relationships with chronic physical conditions: results from the World Mental Health Surveys. Journal of affective disorders, 103(1-3), 113–120.

Selles, R. R., Storch, E. A., & Lewin, A. B. (2014). Variations in symptom prevalence and clinical correlates in younger versus older youth with obsessive–compulsive disorder. Child Psychiatry & Human Development, 45(6), 666–674.

Selvendra, A., Toh, W. L., Neill, E., Tan, E. J., Rossell, S. L., Morgan, V. A., & Castle, D. J. (2022). Age of onset by sex in schizophrenia: Proximal and distal characteristics. Journal of psychiatric research, 151, 454–460.

Shah, B. M., & Kornstein, S. G. (2021). Mental health: Sex and gender evidence in depression, generalized anxiety disorder, and schizophrenia. How Sex and Gender Impact Clinical Practice, 153-169.

Sharma, E., Sharma, L. P., Balachander, S., Lin, B., Manohar, H., Khanna, P., . . . Au, A. C. L. (2021). Comorbidities in obsessive-compulsive disorder across the lifespan: a systematic review and meta-analysis. Frontiers in psychiatry, 12, 703701.

Sheikh, J. I., Leskin, G. A., & Klein, D. F. (2002). Gender differences in panic disorder: findings from the National Comorbidity Survey. American Journal of Psychiatry, 159(1), 55–58.

Silberschmidt, A., Lee, S., Zanarini, M., & Schulz, S. C. (2015). Gender Differences in Borderline Personality Disorder: Results From a Multinational, Clinical Trial Sample. J Pers Disord, 29(6), 828---838. 10.1521/pedi\_2014\_28\_175

Singh, R. S., Singh, K. K., & Singh, S. M. (2021). Origin of sex-biased mental disorders: an evolutionary perspective. Journal of Molecular Evolution, 89, 195–213.

Sinopoli, V. M., Burton, C. L., Kronenberg, S., & Arnold, P. D. (2017). A review of the role of serotonin system genes in obsessive-compulsive disorder. Neuroscience & Biobehavioral Reviews, 80, 372–381.

Skapinakis, P., Caldwell, D. M., Hollingworth, W., Bryden, P., Fineberg, N. A., Salkovskis, P., . . . Churchill, R. (2016). Pharmacological and psychotherapeutic interventions for management of obsessive-compulsive disorder in adults: a systematic review and network meta-analysis. The Lancet Psychiatry, 3(8), 730–739.

Snyder, H. N. (2000). Sexual Assault of Young Children as Reported to Law Enforcement: Victim, Incident, and Offender Characteristics. A NIBRS Statistical Report.

Soomro, G. M., Altman, D. G., Rajagopal, S., & Browne, M. O. (2008). Selective serotonin re-uptake inhibitors (SSRIs) versus placebo for obsessive compulsive disorder (OCD). Cochrane database of systematic reviews(1).

Sousa-Lima, J., Moreira, P. S., Raposo-Lima, C., Sousa, N., & Morgado, P. (2019). Relationship between obsessive compulsive disorder and cortisol: Systematic review and meta-analysis. European Neuropsychopharmacology, 29(11), 1185–1198.

Souza, E. G. V., Ramos, M. G., Hara, C., Stumpf, B. P., & Rocha, F. L. (2012). Neuropsychological performance and menstrual cycle: a literature review. Trends in Psychiatry and Psychotherapy, 34(1), 5–12.

Spence, S. H. (1997). Structure of anxiety symptoms among children: a confirmatory factor-analytic study. Journal of abnormal psychology, 106(2), 280.

Stengler-Wenzke, K., Müller, U., Angermeyer, M. C., Sabri, O., & Hesse, S. (2004). Reduced serotonin transporter–availability in obsessive–compulsive disorder (OCD). European Archives of Psychiatry and Clinical Neuroscience, 254, 252–255.

Stewart, S. E., Yu, D., Scharf, J. M., Neale, B. M., Fagerness, J. A., Mathews, C. A., . . . Osiecki, L. (2013). Genome-wide association study of obsessive-compulsive disorder. Molecular Psychiatry, 18(7), 788–798.

Striegel-Moore, R. H., & Bulik, C. M. (2007). Risk factors for eating disorders. American psychologist, 62(3), 181.

Striegel-Moore, R. H., Rosselli, F., Perrin, N., DeBar, L., Wilson, G. T., May, A., & Kraemer, H. C. (2009). Gender difference in the prevalence of eating disorder symptoms. International Journal of Eating Disorders, 42(5), 471–474.

Summerfeldt, L. J., Kloosterman, P. H., Antony, M. M., & Swinson, R. P. (2014). Examining an obsessive-compulsive core dimensions model: Structural validity of harm avoidance and incompleteness. Journal of Obsessive-Compulsive and Related Disorders, 3(2), 83–94.

Suominen, K., Mantere, O., Valtonen, H., Arvilommi, P., Leppämäki, S., & Isometsä, E. (2009). Gender differences in bipolar disorder type I and II. Acta Psychiatrica Scandinavica, 120(6), 464–473.

Taqui, A. M., Shaikh, M., Gowani, S. A., Shahid, F., Khan, A., Tayyeb, S. M., . . . Shamsi, A. (2008). Body Dysmorphic Disorder: gender differences and prevalence in a Pakistani medical student population. BMC psychiatry, 8, 1–10.

Taylor, S. (2011). Early versus late onset obsessive–compulsive disorder: evidence for distinct subtypes. Clinical psychology review, 31(7), 1083–1100.

Tolin, D. F., Abramowitz, J. S., Kozak, M. J., & Foa, E. B. (2001). Fixity of belief, perceptual aberration, and magical ideation in obsessive–compulsive disorder. Journal of anxiety disorders, 15(6), 501–510.

Transue, B. (2019). APA Style 7th edition.

Turecki, G., Brent, D. A., Gunnell, D., O’Connor, R. C., Oquendo, M. A., Pirkis, J., & Stanley, B. H. (2019). Suicide and suicide risk. Nature reviews Disease primers, 5(1), 74.

Valentine, S. E., Marques, L., Wang, Y., Ahles, E. M., De Silva, L. D., & Alegría, M. (2019). Gender differences in exposure to potentially traumatic events and diagnosis of posttraumatic stress disorder (PTSD) by racial and ethnic group. General hospital psychiatry, 61, 60–68.

Van Buuren, L., Fleming, C. A. K., Hay, P., Bussey, K., Trompeter, N., Lonergan, A., & Mitchison, D. (2023). The prevalence and burden of avoidant/restrictive food intake disorder (ARFID) in a general adolescent population. Journal of eating disorders, 11(1), 104.

Van Der Wee, N. J., Stevens, H., Hardeman, J. A., Mandl, R. C., Denys, D. A., van Megen, H. J., . . . Westenberg, H. M. (2004). Enhanced dopamine transporter density in psychotropic-naive patients with obsessive-compulsive disorder shown by [123I] β-CIT SPECT. American Journal of Psychiatry, 161(12), 2201–2206.

Van Roessel, P. J., Grassi, G., Aboujaoude, E. N., Menchon, J. M., Van Ameringen, M., & Rodríguez, C. I. (2023). Treatment-resistant OCD: Pharmacotherapies in adults. Comprehensive psychiatry, 120, 152352.

Van Zandt, M., Weiss, E., Almyasheva, A., Lipior, S., Maisel, S., & Naegele, J. (2019). Adeno-associated viral overexpression of neuroligin 2 in the mouse hippocampus enhances GABAergic synapses and impairs hippocampal-dependent behaviors. Behavioural brain research, 362, 7–20.

Varoqueaux, F., Jamain, S., & Brose, N. (2004). Neuroligin 2 is exclusively localized to inhibitory synapses. European journal of cell biology, 83(9), 449–456.

Vidal-Ribas, P., Stringaris, A., Rück, C., Serlachius, E., Lichtenstein, P., & Mataix-Cols, D. (2015). Are stressful life events causally related to the severity of obsessive-compulsive symptoms? A monozygotic twin difference study. European psychiatry, 30(2), 309–316.

Vulink, N. C., Denys, D., Bus, L., & Westenberg, H. G. (2006). Female hormones affect symptom severity in obsessive–compulsive disorder. International clinical psychopharmacology, 21(3), 171–175.

Wadsworth, L. P., Potluri, S., Schreck, M., & Hernandez-Vallant, A. (2020). Measurement and impacts of intersectionality on obsessive-compulsive disorder symptoms across intensive treatment. American Journal of Orthopsychiatry, 90(4), 445.

Walsh, K. H., & McDougle, C. J. (2004). Pharmacological augmentation strategies for treatment-resistant obsessive-compulsive disorder. Expert Opinion on Pharmacotherapy, 5(10), 2059–2067.

Wan, Y., Ade, K. K., Caffall, Z., Ozlu, M. I., Eroglu, C., Feng, G., & Calakos, N. (2014). Circuit-selective striatal synaptic dysfunction in the Sapap3 knockout mouse model of obsessive-compulsive disorder. Biological Psychiatry, 75(8), 623–630.

Wan, Y., Feng, G., & Calakos, N. (2011). Sapap3 deletion causes mGluR5-dependent silencing of AMPAR synapses. Journal of Neuroscience, 31(46), 16685–16691.

Waschbusch, D. A., & Willoughby, M. T. (2008). Parent and teacher ratings on the IOWA Conners Rating Scale. Journal of Psychopathology and Behavioral Assessment, 30, 180–192.

Weckhuysen, S., Mandelstam, S., Suls, A., Audenaert, D., Deconinck, T., Claes, L. R., . . . Yordanova, I. (2012). KCNQ2 encephalopathy: emerging phenotype of a neonatal epileptic encephalopathy. Annals of neurology, 71(1), 15–25.

Welch, J. M., Lu, J., Rodriguiz, R. M., Trotta, N. C., Peca, J., Ding, J.-D., . . . Luo, J. (2007). Cortico-striatal synaptic defects and OCD-like behaviours in Sapap3-mutant mice. Nature, 448(7156), 894–900.

Wilhelm, S., Keuthen, N. J., Deckersbach, T., Engelhard, I. M., Forker, A. E., Baer, L., . . . Jenike, M. A. (1999). Self-injurious skin picking: clinical characteristics and comorbidity. Journal of clinical psychiatry, 60(7), 454–459.

Williams, O. O., Coppolino, M., George, S. R., & Perreault, M. L. (2021). Sex differences in dopamine receptors and relevance to neuropsychiatric disorders. Brain Sciences, 11(9), 1199.

Workgroup, N. G. NAMHC Genomics Workgroup: Research Recommendations Summary. Retrieved 8/31/2024 from https://www.nimh.nih.gov/about/advisory-boards-and-groups/namhc/reports/namhc-genomics-workgroup-research-recommendations-summary

Yairi, E., Ambrose, N., & Cox, N. (1996). Genetics of stuttering: A critical review. Journal of Speech, Language, and Hearing Research, 39(4), 771–784.

Yao, Y.-L., & Yang, W.-M. (2003). The metastasis-associated proteins 1 and 2 form distinct protein complexes with histone deacetylase activity. Journal of Biological Chemistry, 278(43), 42560–42568.

Zachry, J. E., Nolan, S. O., Brady, L. J., Kelly, S. J., Siciliano, C. A., & Calipari, E. S. (2021). Sex differences in dopamine release regulation in the striatum. Neuropsychopharmacology, 46(3), 491–499.

Zahn-Waxler, C., Crick, N. R., Shirtcliff, E. A., & Woods, K. E. (2006). The origins and development of psychopathology in females and males.

Zhang, Y., Luo, Q., Huang, C.-C., Lo, C.-Y. Z., Langley, C., Desrivières, S., . . . Bokde, A. L. (2021). The Human Brain Is Best Described as Being on a Female/Male Continuum: Evidence from a Neuroimaging Connectivity Study. Cerebral Cortex, 31(6), 3021–3033.

