## Appendices for "Sex and gender differences in OCD: A scoping review and a two subtypes hypothesis"

**for**

**Sex or gender differences in OCD:**

**A scoping review of literature**

**July 1, 2009 – June 7, 2026**

**Appendix A.1.** Meta-analyses, systematic reviews, narrative reviews, and other integrative papers published between July 1, 2009, and June 7, 2026.

A.1.1. Topics addressed by prior reviews and meta-analyses.

| **#** | **Author Date** | **Symptoms & Burden**  **(N = 24)** | **Incidence & Prevalence**  **(N = 15)** | **Risks, Onset, Progression & Course**  **(N = 49)** | **Comorbidities**  **(N = 28)** | **Genetics**  **(N = 21)** | **Neurocognitive & Neurobiological functioning**  **(N = 13)** | **Treatment**  **(N = 37)** | **Animal models**  **(N = 7)** |
| --- | --- | --- | --- | --- | --- | --- | --- | --- | --- |
| **Meta analyses** | | | | | | | | | |
| 1 | Speisman 2011 |  |  | x |  |  |  |  |  |
| 2 | Taylor 2011 |  |  | x |  |  |  |  |  |
| 3 | Russell 2012 |  |  | x |  |  |  |  |  |
| 4 | Friborg 2013 |  |  |  | x |  |  |  |  |
| 5 | Olatunji 2013 |  |  |  |  |  |  | x |  |
| 6 | Stewart 2013 |  |  |  |  | x |  |  |  |
| 7 | Taylor 2013 |  |  |  |  | x |  |  |  |
| 8 | Goodman 2014 |  |  | x |  |  |  |  |  |
| 9 | Sharma 2014 |  |  |  |  |  |  | x |  |
| 10 | Swets 2014 |  |  |  | x |  |  |  |  |
| 11 | Thompson-Hollands 2014 |  |  |  |  |  |  | x |  |
| 12 | Mak 2015 |  |  |  |  | x |  |  |  |
| 13 | Marchesi 2015 |  |  |  |  |  |  | x |  |
| 14 | Wang 2015 |  |  |  |  | x |  |  |  |
| 15 | Coluccia 2016 | x |  |  |  |  |  |  |  |
| 16 | Guo 2016 |  | x |  |  |  |  |  |  |
| 17 | Moser 2016 |  |  |  |  |  | x |  |  |
| 18 | Taylor 2016 |  |  |  |  | x |  |  |  |
| 19 | Vieira 2016 | x |  |  |  |  |  |  |  |
| 20 | Coluccia 2017 | x |  |  |  |  |  |  |  |
| 21 | Lijster 2017 |  |  | x |  |  |  |  |  |
| 22 | Brutocao 2018 |  |  | x |  |  |  |  |  |
| 23 | Hsu 2018 |  |  |  | x |  |  | x |  |
| 24 | Taylor 2018 |  |  |  |  | x |  |  |  |
| 25 | Lai 2019 |  |  |  | x |  |  |  |  |
| 26 | Pozza 2019 | x |  |  |  |  |  |  |  |
| 27 | Viswasam 2019 |  |  | x |  |  |  |  |  |
| 28 | Brakoulias 2020 |  |  |  |  |  |  | x |  |
| 29 | Crow 2020 | x |  |  |  |  |  |  |  |
| 30 | Eser 2020 |  |  |  | x |  |  |  |  |
| 31 | Fawcett 2020 |  | x |  |  |  |  |  |  |
| 32 | Kumar 2020 |  |  |  |  | x |  |  |  |
| 33 | Leeuwen 2020 |  |  |  |  |  | x |  |  |
| 34 | Mattina 2020 |  |  |  |  | x |  |  |  |
| 35 | Stevens 2020 |  |  |  | x |  |  |  |  |
| 36 | Stewart 2020 |  |  |  |  |  |  | x |  |
| 37 | Pozza 2021 |  |  |  | x |  |  |  |  |
| 38 | Sharma 2021 |  | x |  | x |  |  |  |  |
| 39 | Burton 2022 |  |  |  |  |  |  |  |  |
| 40 | Handy 2022 |  |  |  | x |  |  |  |  |
| 41 | Hao 2022 |  |  |  |  |  | x |  |  |
| 42 | Mar 2022 |  |  |  |  | x |  |  |  |
| 43 | Melkonian 2022 |  |  | x |  |  |  | x |  |
| 44 | Rosa-Alcázar 2022 |  |  |  |  |  |  | x |  |
| 45 | Shang 2022 |  |  |  |  | x |  |  |  |
| 46 | Solmi 2022 |  | x |  |  |  |  |  |  |
| 47 | Jalalifar 2023 |  | x |  |  |  |  |  |  |
| 48 | Li 2023 |  |  |  |  |  | x |  |  |
| 49 | Mendez 2023 |  |  |  |  |  |  | x |  |
| 50 | Mitchell 2023 |  |  | x |  |  |  |  |  |
| 51 | de Pablo 2023 |  |  |  |  |  |  | x |  |
| 52 | Aujla 2024 |  |  | x |  |  |  |  |  |
| 53 | Aymerich 2024 | x |  |  | x |  |  | x |  |
| 54 | Cohen 2024 |  |  |  |  |  |  | x |  |
| 55 | Heydarikhayat 2024 |  | x |  |  |  |  |  |  |
| 56 | Mitchell 2024 |  |  |  |  |  |  | x |  |
| 57 | Yucens 2024 | x |  |  |  |  |  |  |  |
| 58 | Zhang 2024 |  |  |  | x |  |  | x |  |
| 59 | Zisler 2024 |  |  |  |  |  |  | x |  |
| 60 | Hühne 2025 |  |  | x |  |  |  |  |  |
| 61 | Cohen 2025a |  |  |  |  |  |  | x |  |
| 62 | Cohen 2025b |  |  |  |  |  |  | x |  |
| 63 | Cohen 2025c |  |  |  |  |  |  | x |  |
| 64 | Kępińska 2025 |  |  | x |  |  |  |  |  |
| 65 | Lamothe 2025 |  |  |  |  |  |  | x |  |
| 66 | Li 2025 |  |  |  |  |  | x |  |  |
| 67 | Maroufi 2026 |  |  |  |  |  |  | x |  |
| 68 | Pellegrini 2025 |  |  |  |  |  |  |  |  |
| 69 | Socratous 2026 | x |  |  |  |  |  |  |  |
| 70 | Somer 2025 | x |  |  |  |  |  |  |  |
| 71 | Thavarajasingam 2026 |  |  |  |  |  |  | x |  |
| 72 | Wan 2025 |  |  |  |  |  | x |  |  |
| **Systematic/Scoping Reviews** | | | | | | | | | |
| 73 | Mathis 2011 | x |  | x | x | x |  |  |  |
| 74 | McGuinness 2011 |  |  | x |  |  |  |  |  |
| 75 | Subramaniam 2014 | x |  | x | x | x |  |  |  |
| 76 | Ma 2015 |  |  |  |  |  |  |  |  |
| 77 | Kluiver 2016 |  |  | x |  | x |  |  |  |
| 78 | Kern 2017 |  |  | x |  |  |  |  |  |
| 79 | Green 2021 |  |  | x |  |  |  |  |  |
| 80 | Velurajah 2021 | x |  |  |  |  |  |  |  |
| 81 | Walker 2021 |  |  | x |  |  |  |  |  |
| 82 | Westby 2021 |  |  | x |  |  |  |  |  |
| 83 | da Silva 2022 |  |  |  |  |  |  |  | x |
| 84 | Liveri 2023 |  |  | x | x |  |  |  |  |
| 85 | Albanese 2026 |  |  |  |  |  |  |  |  |
| 86 | Baldini 2025 |  |  | x |  |  |  |  |  |
| 87 | JXC Ke 2025 |  |  | x |  |  |  |  |  |
| 98 | Mojgani 2026 |  |  |  |  |  |  |  |  |
| 89 | Naderi 2025 |  |  |  |  |  | x |  |  |
| 90 | Perich 2025 |  |  | x |  |  |  |  |  |
| 91 | Sharifi 2025 |  |  |  |  |  |  |  |  |
| 92 | Taylor 2011 | x | x | x | x | x | x | x |  |
| **Narrative reviews** | | | | | | | | | |
| 93 | Stewart 2012 |  | x | x |  |  |  |  |  |
| 94 | Kokras 2014 |  |  |  |  |  |  |  | x |
| 95 | Meltzer-Brody 2014 |  |  | x |  |  |  | x |  |
| 96 | Namouz-Haddad 2014 |  |  | x |  |  |  | x |  |
| 97 | Sharma 2015 |  |  | x |  |  |  | x |  |
| 98 | Uguz 2015 |  |  |  |  |  |  | x |  |
| 99 | Bandelow 2016 |  |  |  |  |  | x |  |  |
| 100 | Mattina 2016 |  |  | x |  | x | x |  |  |
| 101 | Karpinski 2017 |  |  |  |  |  | x |  |  |
| 102 | Sinopoli 2017 |  |  |  |  | x |  |  |  |
| 103 | Uguz 2018 |  |  |  |  |  |  | x |  |
| 104 | Burchi 2019 |  |  | x |  |  |  |  |  |
| 105 | Challacombe 2019 | x |  | x |  |  |  |  |  |
| 106 | Mathes 2019 | x | x | x | x |  |  | x |  |
| 107 | Piontek 2019 | x |  |  |  |  |  |  |  |
| 108 | Sharma 2019 | x |  |  |  |  |  |  |  |
| 109 | Escobar 2020 | x |  | x | x |  |  |  | x |
| 110 | Mattina 2020 | x | x | x | x | x | x | x |  |
| 111 | Vasiliadis 2020 |  | x |  |  |  |  |  |  |
| 112 | Wendt 2020 |  |  |  |  | x |  |  |  |
| 113 | Franceschini 2021 | x | x | x | x | x | x | x | x |
| 114 | Garris 2021 |  |  |  | x |  |  |  |  |
| 115 | Mitra 2021 |  |  |  |  |  |  |  | x |
| 116 | Naguy 2021 | x | x | x | x |  |  | x |  |
| 117 | Reale 2021 |  |  |  |  |  |  | x |  |
| 118 | Sharma 2021a |  |  | x |  |  |  | x |  |
| 119 | Sharma 2021b |  | x |  | x |  |  |  | x |
| 120 | Hudepohl 2022 |  |  | x |  |  |  |  |  |
| 121 | Katz 2022 |  |  | x | x |  |  | x |  |
| 122 | Wang 2022 |  |  |  |  | x |  |  |  |
| 123 | Baláž 2023 |  |  |  |  |  |  |  |  |
| 124 | Schuyler 2023 |  | x |  | x |  |  | x |  |
| 125 | Grandinetti 2024 | x |  | x |  |  |  |  |  |
| 126 | Vismara 2024 | x |  | x |  |  |  |  |  |
| 127 | Hipke, 2024 |  |  | x |  |  |  |  |  |
| 128 | Zarowski 2024 |  |  | x |  |  |  |  |  |
| 129 | d’Aversa 2026b |  |  |  |  | x |  |  |  |
| 130 | de la Iglesia-Larrad 2025 | x |  |  |  |  |  |  |  |
| 131 | Dolfi 2025 |  |  |  | x |  |  |  |  |
| 132 | Etyemez 2025 |  |  | x |  |  |  |  |  |
| 133 | Fairbrother 2025 |  |  |  |  |  |  |  |  |
| 134 | Lacroix 2026 |  |  |  | x |  |  |  |  |
| 135 | Pinciotti 2025 |  |  |  |  |  |  | x |  |
| 136 | Puszcz 2025 |  |  |  | x |  |  |  |  |
| 137 | Scholten 2026 |  |  | x |  |  |  |  |  |
| 138 | Syed 2025 |  |  |  |  |  |  |  | x |
| **Conceptual and integrative reviews** | | | | | | | | | |
| 139 | Kloft 2018 |  |  | x | x |  |  |  |  |
| 140 | Vervoort 2021 |  |  | x |  |  |  |  |  |
| 141 | Moreira-de-Oliveira 2025 |  |  | x |  |  |  |  |  |

A.1.2. Summary of results reported by the prior reviews and meta-analyses.

| **Topic** | **Consensus** | | **Hypotheses** |
| --- | --- | --- | --- |
|  | **Meta analyses (N = 38),**  **Systematic reviews (N = 8)** | **Narrative reviews (N = 25)** |  |
| **Symptoms & Burden** | Juvenile. **M ~ F**: Sx  **M ≤ F:** Academic burden  Adults. **M > F**: sexual-religious and aggressive Sx; olfactory impairment; overall QoL (but see [26])); social impairment, single; family accommodation  **M < F**: contamination/cleaning.; OCS severity in trauma-exposed  **M:** OCD↑↑erectile dysfunction | Juvenile Sx: **M ~ F**.  Adult**. M > F**: broad symptom presentation; sexual, aggressive, checking, symmetry, ordering Sx; sexual obsessions; sleep problems.  **M < F:** 1-2 symptoms; contamination; sexual disfunction (but ED in M is common); insight; suicide risk; married with children; | Broader Sx + burden in males and more specific in females. |
| **Comorbidities** | Juvenile. **M > F**: ASD, tic, ADHD  Adults. **M > F**: tic, SUD, internet addiction;  **M < F**: ANX, MDD, ED, impulse control d/o. | Juvenile **M > F**: ODD, ADHD, TD, tic disorder.  Adult. **M > F**: tic, AUD, compulsive drug use, compulsive sexual behavior, compulsive internet use, sexual dysfunction.  **M < F:** ANX, MDD, ED, AN, BN, BP, ASD, TTN, specific phobia, skin picking and compulsive buying, impulse control d/o, self-injury, atopic dermatitis, progression to compulsive drug use; hoarding in ASD + OCD. | Externalizing d/o in males and internalizing d/o in females |
| **Incidence & Prevalence** | Juvenile. **M > F**; community samples less consistent  Adult/lifetime. **M < F**.  Older: **M < F**. | Juvenile Clinical. **M > F**; subclinical: unclear; possible recent trends toward F > M?  Adults: Clinical **M < F**; subclinical: unclear.  Treatment seeking: **M < F** | More common in juvenile boys and adult women |
| **Risks, Onset, Progression, & Course** | **M > F:** juvenile onset, longer duration, chronic course, sensitivity to several neurotoxins, familiar TS > familiar TD (inconsistent).  **M < F**: adult onset.  M ~ F: SLEs pre-onset ↑↑ OCD  **F:** OCS ↑↑reproductive cycle events; PP ↑↑ contamination & aggressive Sx; PCOS↑↑OCD.  M: OCS ↑↑ perinatal partners (Ante > PP)  Methodology concerns: retrospective reports | **M > F:** juvenile onset (more recent report weaker effects); family history; chronic course; PANDA.  **M < F:** adult onset; childhood Sx --> adult Dx; episodic course; SLE-OCD; sexual abuse in childhood.  **F:** aggressive, sexual, religious Sx↑↑ episodic course, contamination Sx ↑↑ chronic course; reproductive cycle events ↑↑ OCS; premenstrual phase ↑↑ OCS; menopause may ↑ or ↓ OCS, with new onset reported in a minority; PP ↑↑ aggression toward the baby, PP ↑↑ OCS in 1/3 of OCD mothers; stillbirth ↑↑ OCS in mothers; maternal OCD ↑ pre-eclampsia, preterm birth, CS, low birth weight, neonatal complications. OCS  PP OCD risks: primiparity, early postpartum, prior MD, OCPD, AVPD, SSD; obstetric complication; OCD-related beliefs; familial mood d/o, SUD. PP OCD clinical concerns: safety of treatments, safety of the baby  In students, COVID ↑↑ OCS; severity during COVID: **M < F**. | SLE x estrogens & progestogens ↑↑ OCD |
| **Genetics** | Candidate gene studies of certain polymorphisms show some sex differences.  BDNF Val66Met ↑↑ risk for OCD in **F** only  Paternal age ↑↑ risk for OCD, possibly through epigenetic alternations, in a small # of studies | Familial/twin studies: OCD in **M** with earlier onset higher familial loading, but twin studies show small to no gender differences  Candidate gene studies: specific polymorphisms show some differences with some inconsistencies  GWAS: **M~F** heritability estimates  Epigenetics: sex-stratified methylome analyses identified opposite-sex DMRs and significant sex × methylation interactions, suggesting potentially sex-dimorphic epigenetic architecture.  F: PP OCD ↑↑ methylation at *TTC9B* and *MS4A7* (preliminary)  Genetic risk: **F:** SNPs linked to PCOS → risk for OCD (but SNPs linked to OCD do not predict risk for PCOS) | Δ estradiol ↑↑ Δ OCS - through transcriptional modulation of associated genes |
| **Neurocognitive & Neurobiological functioning** | Juvenile. **M ~ F**: ERN elevation in OCD. **M > F**: ↓ left MOG GM volume  Adult. **M ~ F:** attachment anxiety, ERN ↑↑ OCS; OCD ↓ peripheral BDNF; rs-fMRI: IPL/cingulate/cerebellum/MFG–IFG ↑↑ OCD; IPL/cingulate/cerebellum/MFG–IFG; insula/PreCG–PostCG/caudate ↓↓ OCD; SSRT ↑↑ OCD | Among opposite sex twin pairs. **M > F:** Right temporal gyrus volume.  **M < F:** the right precuneus volume.  Juvenile. **M, OCD < HC:** the pituitary volume.  Adults. **M > F:** OCD-related changes in 5-HT synthesis; perinatal trauma or early life brain injury ↑↑ OCD (inconsistent)  **M, OCD > HC:** midsagittal corpus callosum area volume; cortisol level; **OCD < HC:** testosterone.  **F, OCD < HC:** total white matter; **OCD ~ HC:** progesterone; **OCD > HC:** DHEA. | Estrogen ↑↓OCD;  Estrogen & progesterone → serotonergic, dopaminergic, & glutamatergic neurotransmission → OCS |
| **Treatment** | **M > F:** family based (VERY small literature); accuracy of early non-improvement for predicting later antidepressant nonresponse  **M ~ F:** CBT/ SSRI, pediatric; SSRI efficacy/response in adults; DBS response in severe/refractory OCD; remission in adults; naturalistic clinical progress in adults. Extinction (fear) > Extinction (disgust)  **M ≤ F:** SSRI/ clomipramine; adults and children; dropout from iCBT  **F:** CBT/SSRI effective in P/PP; capsulotomy complete response may be higher with greater female representation. | **M ≥ F**: CBT.  **M < F:** treatment-seeking & health care usage; SSRI response: decrease and subsequent remission of Sx in adults.  **F:** PP OCD: use SSRI/CBT, no guidelines for the lactation period. | Treatment response is a function of comorbidities |
| **Animal models** | **M < F:** Binge Eating – high responders, enhanced palatable chow consumption; activity-based anorexia (inconsistent); mCPP ↑ OCD-like responses (limited evidence)  **M = F:** Marble burying in mice selected for high burying  **M > F:** Spontaneous alternation perseveration in 8-OH-DPAT model, F perseveration varies with estrous cycle | **M < F**: Amphetamine induced dopamine release, locomotion  **M = F:** Compulsive Lever Pressing; Deer mice, stereotyped behaviors  **M > F**: KOR sensitivity, F blunted inhibition of dopamine release, ArKO mice, enhanced stereotyped grooming, hyperlocomotion, decreased PPI  **F**: Neonatal activation of D2 receptor ↑↑ amphetamine behavioral sensitization; KOR attenuates cocaine-induced hyperlocomotion in estradiol-dependent manner  **M**: Repeat D2 agonism ↑↑ locomotor sensitization/compulsive-like behaviors, KOR facilitates inhibitory control of D2 receptor | estrogen and estrous cycle → behavior |

**Appendix A.2.** Studies on Symptoms and burden of OCD

A.2.1. Studies of gender differences in OCD symptom presentation.

| **First Auth, Year** | | **N**  **(F)** | | **Age mean**±**SD (range)** | **Pop** | **Meas.** | **Gender effects (F vs. M)** | |
| --- | --- | --- | --- | --- | --- | --- | --- | --- |
|  |  |  |  |  |  |  | **Stats** | **Direction** |
| *all ages* | | | | | | | | |
| Zhang 2013^c^ | | 512  (231) | | 28 ± NA  (8-77) | OCD  (CHN) | YBOCS CL | Presence. Contam/Wash ~ F: β = 0.47, p < 0.001. | contam  M < F |
| Algin  2018 | | 400  (165) | | 27 ± 10  8-63 | OCD  (BGD) | YBOCS CL | Presence. M:F OR: Obsess: Contam = 2.79 [95%CI, 1.74-4.49], Sexual = 0.43 [95%CI, 0.26-0.73], misc = 0.51 [95%CI, 0.29-0.90]; Compuls: clean = 3.35 [95%CI, 2.05-5.46], Order = 1.59 [95%CI, 1.03-2.45], check = 0.51 [95%CI, 0.34-0.77], repeat = 0.61 [95%CI, 0.38-0.96]. | sex obs/misc obs /check/repeat: M > F  contam/order: M < F |
| Nanthakumar 2025 | | 1001 (569) | | 35 ± 13  (9–82) | OCD  (BRA) | YBOCS, DYBOCS | Prevalence. Health-related OCS: F:M χ² = 0.12, p = 0.73 | health-related OCS:  M ~ F |
| *children/adolescents* | | | | | | | | |
| Orgilés  2012 | | 2522  (1286) | | NA  (8-17) | STU  (ESP) | *SCAS* | Severity. M = 4.5 ± 3.3 F = 4.9 ± 3.4, d = 0.12 p > 0.10 | severity  M ~ F |
| Fernández  2013 | | 383  (167) | | 14 ± 2  (8-17) | OCD  (GBR) | CY-BOCS  CL | Presence. Sexual obs M:F OR = 1.2 ([95%CI= 0.7 ‒ 1.8], χ2 = 0.33 p = 0.56). | sexual M ~ F |
| Vivan  2013 | | 75  (59) | | 15 ± 1  (NA) | students  (BRA) | DYBOCS | Presence. Aggress, Sex/Relig, Symm, Contam, Misc: M:F OR < 1.14 p > 0.23  Severity. Sex/Relig, M ≥ F: d =0.47 p = 0.10; Aggress, Symm, Contam, Misc: d<0.39 p> 0.16 | severity  sexual/relig M ≥ F |
| Alvarenga  2015 | | 9937  (4729) | | 9 ± 2  (6-12) | GEN  (BRA) | DYBOCS | Severity. Controlling for age and OCS in FDRs: OCS ~ F β = 0.08 p = 0.001; Symm/Aggress/Sexual/Relig/Contam ~ M β < 0.1 p = 0.45. | severity  M < F |
| Højgaard  2017 | | 854 (401) | | 12  (7-18) | OCD  (USA  EURO) | CY-BOCS | Presence. Factor scores ~ M: Harm/Sex β = 0.007 p > 0.05, Symm/Hoard β = -0.095, p < 0.001, Contam/Wash β = -0.020, p > 0.05. | symm/hoard M < F  harm/sex/contam  M ~ F |
| Pozza  2017 | | 1408  (823) | | 12 ± 2  (8 – 17) | GEN  (world) | *OCI-CV* | Severity. M < F Fs(1,1406) = 20.81 – 66.43 ps<0.001 | all dimensions  M < F |
| Wu  2018^t^ | | 215 (92) | | 12 ± 3  (7-17) | OCD  (USA) | CY-BOCS CL | Presence. Relig OCS: F χ^2^(1,215) = 0 p = 0.98. | relig  M ~ F |
| Schultz  2018 | | 57  (39) | | 14 ± 2  (7-17) | OCD  (DNK) | *OBQ-CV* | Severity. OBQ-CV subscales ~ F ps > 0.05, statistics not reported | all dimensions  M ~ F |
| Cervin 2020 | | 120  (75) | | 13 ± 3  (7 – 17) | OCD  (SWE) | DYBOCS | Presence. F%: Aggrs = 0.64, Symm = 0.71, Contam = 0.54, p = 0.35  Severity by types. Aggrs ~ M: β= 0.33 [95%CI = -0.71 ‒ 1.38]; Symm ~ M: β = -0.99 [95%CI = -0.92 ‒ -0.07]; Contam ~ M: β= 0.67 [95%CI = -1.04 ‒ 1.29]. | overall /symm M < F aggrs/contam  M ~ F |
| Rough  2020 | | 755  (431) | | 14 ± 3  (7 – 18) | OCD  (USA) | *OCI-CV* | Severity. Order ~ F: t(1,753) = 2.13 p = 0.03 | order  M < F |
| Piqueras 2021 | | 1499 (745) | | 13 ± 3 (8-18) | schools (ESP) | *DWDS* | Severity. OCD F – M: F = 1.71 vs. M = 1.64, d = 0.12, p < 0.05 | overall severity  M < F |
| Cervin 2025 | | 248 (128) | | 12 ± 3 | OCD  DEN, NOR, SWE | CY-BOCS | Severity. OCS ~ M: β = −1.17, 95% CI [−2.72, 0.39], p = 0.14. | overall severity  M ~ F |
| Su  2025 | | 147  (69) | | NA  (6-16) | OCD  (CHI) | CBCL-OCS | Severity. M = 4.7 ± 2.4, F = 4.4 ± 2.5; Cohen’s d = 0.10 t = 0.61, p = 0.54 | severity M ~ F |
| Pine  2026 | | 101 (46) | | 13 ± 3  (7–17) | help- seeking OCD (USA) | CY-BOCS-II | Presence. # of symptoms: F > M t(84.22) = 3.02, p=0.003; checking, hoarding, and counting symptoms F > M.  Severity. CY-BOCS-II total ~ sex, p > 0.05. Sex × age-group interactions for symptom count and severity: ps > 0.05. | M < F count/content  M ~ F severity |
| *adults* | | | | | | | | |
| Matsunaga 2010 | | 343  (201) | | 30 ± 9  (15 - 68) | OCD  (JPN) | GAF, YBOCS | Predominant. F%: Contam/Wash (CW) = 0.82; Hoard = 0 (vs. CW Fisher p<0.01); (3) Symm/Order = 0.40 (vs. CW χ2 = 30.5, p< 0.01); (4) Forbid thoughts/Check = 0.42 (vs. CW χ2 = 30.2, p< 0.01) | contam  M < F |
| Siev  2011 | | 147 (111) | | 37 ± 14  (18+) | OCD  (USA) | *OCI-R, PIOS* | Presence. F%: scrupulosity = 0.7, no scrupulosity = 0.8 (χ2 *p* = 0.17). | scrupulosity  M ~ F |
| Starcevic 2011 | | 124 (76) | | 45 ± 16  (18+) | OCD  (NZL) | YBOCS | Presence. Avoidance ~ F: β = 1.34, *p* = 0.54 | avoidance  M ~ F |
| Jang  2010 | | 288  (102) | | 25 ± 7  (18+) | OCD, HC  (KOR) | YBOCS CL | Severity. Contam/Wash F > M z = -2.91 p = 0.004. | contam  M < F |
| Shimshoni  2011 | | 60  (24) | | 33 ± 12  (18-62) | OCD  (ISR) | DSM-IV  BABS | Insight. DSM-IV criteria mean rank: M = 34.0, F = 25.25, z = 2.43 p = 0.014; BABS impression mean rank: M = 25.86, F = 37.45, z = 2.65 p = 0.004. | insight  M < F |
| Williams 2011 | | 431 (220) | | 35 ± 13  (18+) | OCD  (USA) | YBOCS | Presence. Lifetime history of obsessions about sexual orientation: M:F OR = 1.9 ([95%CI=1.02- 3.55], χ2 = 4.19 p = 0.048) | sex-orient  M > F |
| Torresan 2013^c, e^ | | 858 (504) | | 35 ± 12 (18-77) | OCD  (BRA) | YBOCS, DYBOCS | Presence. F:M OR & χ2 p-value: Sex/relig = 0.7 p = 0.01; Aggres = 1.5 p = 0.009; Contam = 1.7 p = 0.001; Hoard = 1.4 p = 0.03; Symm/Order = 1.0 p = 0.94.  Severity. F - M d: YBOCS total = 0.35 p < 0.001; YBOCS obs = 0.29 p < 0.001; YBOCS contam = 0.39 p < 0.001; OCS ‘worse ever’ = 0.2 p = 0.05; OCS current = 0.29 p = 0.001  Severity by types. F - M d & M-W p-value: Sex/relig = -0.16 p = 0.02; Aggres = 0.19 p = 0.005; Contam = 0.29 p < 0.001; Hoard = 0.17 p = 0.006; Symm/Order = 0.23 p = 0.001. | Presence: aggress/  contam/ hoard: M < F  symm/order: M ~ F  sex/relig: M > F  Severity:  overall/aggress/contam/hoard/symm-order: M < F  sex/relig: M > F |
| Cherian 2014^c, e^ | | 545 (213) | | 30 ± 12  (18+) | OCD  (IND) | YBOCS | Presence. M:F OR & χ2 /p-value: Sex = 2.87 p < 0.001; Relig =2.03 p = 0.001; doubts = 2.57 p < 0.001; Contam = 0.64 p = 0.017; Aggres = 1.45 p = 0.062; Check = 2.15 p < 0.001; Count = 2.25 p = 0.012; Repeat = 2.34 p < 0.0001; Wash/Clean = 0.55 p = 0.002; Insight = 0.68 p = 0.44.  Severity. F – M d: YBOCS total = 0.18 p = 0.04. | sex/relig/doubt/check/ count/repeat: M > F  aggrs/insight M ~ F  overall severity/ contam/wash M < F |
| López-Solà 2014 | | 110  (50) | | 30 ± 10  (18+) | OCD  (ESP) | *DOCS* | Severity. F – M d & p-value: DOCS-tot = 0.44 p = 0.03, Contam = 0.40 p = 0.04, Resp for harm = 0.46 p = 0.03, Unacc thoughts = 0.12 p = 0.55, Symm = 0.53 p =0.01. | overall/contam/ resp/symm M < F  unacc M ~ F |
| Storch 2015 | | 954 (555) | | 36 ± 13 (18-82) | OCD  (BRA) | YBOCS,  CGI-S | Severity. F – M d: YBOCS total = 0.34 p < 0.01, YBOCS obs = 0.26 p < 0.01, YBOCS comp = 0.36 p < 0.01, CGI-S = 0.18 p < 0.01. | overall severity  M < F |
| Williams  2015 | | 1176  (299) | | 26 ± 9  (18+) | GEN  (world) | *Study-specific* | Presence. Worry About Sexuality (LGBT) M – F d = 0.43 t(1,235)=2.7 p =0.004. | sexual orient  M > F |
| Chamberlain 2016 | | 515 (174) | | 22 ± 4 (18-29) | gamble (GBR) | *Padua Inventory* | Presence. Padua total ~ M: β = - 0.10 p = 0.02. | overall severity  M < F |
| Yap  2018 | | 306  (220) | | 33 ± 12  (19-78) | GEN  (AUS) | *DOCS* | Severity. Contam~ F: β = 0.03 t = 0.47 p > 0.10, Resp for harm ~ F: β = 0.12 t = 2.34 p < 0.05, Unacc thoughts ~ F: β = -0.10 t = 2.13 p < 0.05, Symm ~ F: β = 0.05 t = 0.97 p > 0.10, overall sever ~ F: β = 0.03 t = 0.59 p > 0.10 | unacc M > F  contam/symm/ overall M ~ F  harm M < F |
| Citkowska-Kisielewska 2020 | | 262  (183) | | 33 ± 8 (18-58) | OCD,  GAD  (POL) | *SCL-90-R* | Presence. Check: OCD-F = 0.72 vs. GAD-F = 0.54 p =0.001, OCD-M = 0.76 vs. GAD-M = 0.70 p > 0.05; Obses/Repeat/Agress/Comps/Relig/Count/SlowComp: Dx x F p > 0.05. Count: CogDys-M = 0.38 vs. noCogDys-M = 0.16 p = 0.047, CogDys-F = 0.18 vs. noCogDys-F = 0.25 p > 0.05; Check/Obses/Repeat/Agress/Comps/Relig/Slow Comp: CogDys x F p > 0.05. | check  OCD-F ~ OCD-M ~ GAD-M > GAD-F |
| Destrée  2020 | | 954  (548) | | 34 ± 13  (18+) | OCD  (BRA) | DYBOCS | Severity. Sex/Relig ~ M β = 0.90 p =0.03. | Sex/Relig  M > F |
| Ching  2021 | | 592 (413) | | 19 ± 2  (18+) | STU (USA) | *SORT, HS, DPSS-R-12* | Sx content. Sexual-orientation OC (SORT): M = 7.06 ± 8.12 F = 5.23 ± 6.47; Disgust sensitivity (DPSS-R-12): M = 14.4 ± 4.8 F = 13.4 ± 5.4 p < 0.05; Disgust propensity (DPSS-R-12): M = 16.4 ± 4.5 F = 17.3 ± 4.3 p < 0.05 | Sex orient OCS:  M > F  Disgust: M < F |
| Guillén-Font 2021 | | 81  (41) | | 45 ± 12  (18+) | OCD  (ESP) | YBOCS  item 11 | Insight impairment. F =1.34 ± 1.2 M = 1.95 ± 1.5, p = 0.05 | Insight  M < F |
| Forsén Mantilla 2022 | | 500 (376) | | 33 ± 12 (18–71) | GEN  (SWE) | *OCI-R* | Severity. F = 16.0 ± 10.8; M = 14.3 ± 11.5; Cohen’s d = 0.16; p = 0.123 | overall severity  M ~ F |
| Benatti  2022^B^ | | 491 (297) | | 39 ± 13  (18+) | OCD  (11 countr.) | YBOCS, MADRS, *SDS* | Severity. YBOCS ~F β = 0.03 p = 0.56. | overall severity  M ~ F |
| Guazzini 2022^B^ | | 660  (469) | | 25 ± 7  (18+) | GEN  (ITA) | *VOCI* | Severity. VOCI: M = 46.2 ± 28.6 F = 48.9 ± 27.4. | overall severity  M ~ F |
| Kim  2022 | | 398 (218) | | 36 ± 11  (18+) | OCD, ANX (KOR) | *Padua, BDI, BAI* | Severity. Correspondence analysis (CA) for severity levels, r: M = 0.86-0.97. F = 0.89-0.94. CA for OC-beliefs – distress coupling: for threat, reponseb, uncertainty M > F; | OCS: M ~ F  OC-beliefs ↑↑distress:  M > F |
| Pascual-Vera  2022 | | 39  (14) | | 33 ± 12 (18–62) | OCD  (ESP) | ADIS-5-L, YBOCS*, QUIT* | Presence. OCS + body + illness-related intrusive thoughts: F:M OR = 0.97 [95%CI, 0.23-4.15] | OCS + illness + body:  M ~ F |
| Al-Awad 2024 | | 886  (453) | | med=22  (16–32) | med STU  (SAU) | *OCI-R* | Severity. F = 21 (IQR 12–34), M = 17 (IQR 10–30), Wilcoxon p < 0.01; OCI-R ~ F=1.86 [95%CI, 0.13-3.50]; OCI-R ~ F x living alone = 10.10 [95%CI, 3.93 – 16.27] | overall severity  M < F |
| Ayoub  2024 | | 50  (31) | | NA  (18-64) | OCD-religious  (LBN) | *ASOC, SIBS* | Severity. M vs. F: U = 232.0, p = 0.21 | overall severity  M ~ F |
| Barnhart 2025 | | 509 (390) | | 20 ± 3  (18+) | STU (USA) | *SOCDS* | Severity. SOCDS ~ F = -0.053 p = 0.19. | overall severity  M ~ F |
| Brueggerhoff-Batel 2025 | | 45  (22) | | 31 IQR 19  (18–70) | OCD (MEX) | *YBOCS* | Severity. Obses: M=7.0, F=9.5, U=177, p=.086; compul: M=8.0, F=9.5, U=174, p=0.07; total: M=15.0, F=20.0, U=172, p=0.07. Severity-category distribution ~ sex: p=0.24. | overall, obsses, compl M ≤ F |
| Lawrence-Sidebottom 2025 | | 2151  (1309) | | 15 ± 1 (13–17) | digital health  (USA) | *DOCS* | Presence. Elevated OCS: F = 72.2% vs non-elevated OCS F = 55.5%, χ²(1)=54.5, p<0.001; gender non-conforming: 8.2% vs 4.0%, χ²(1)=15.91, p=0.001. Severity. During care, F > non-F: β=4.55, t(525)=3.27, p=0.001; symptom-dimension severity F > non-F: β=1.42, t(504)=3.80, p<0.001. | overall OCS:  F > non-F |
| Melli  2025 | | 393 (285) | | 23 ± 5 18–67 | GEN  (ITA) | *OC-CDS* | Severity. OCD was gender effects non-significant with all p-values > .05 for ordering, hoarding, checking, washing, neutralizing, obsessing | M ~ F severity |
| Huang  2025 | | 211 (102) | | 31 ± 8 (18-53) | OCD  (CHN) | YBOCS | Severity. YBOCS ~ gender: r=.165, FDR-adjusted p<0.05. Sex coding/direction NR. | N/A |
| Gomleksiz 2025 | | 882 (645) | | 21 ± 3 (17–45) | STU (TUR) | *OCI-R* | Severity. F=25 (0–72), M=22 (0–72); U=69,899.00, p=.051. | M ≤ F severity |
| Marković 2025 | | 963 (482) | | 42 ± 13 (18-65) | GEN  (SER) | *OCI-4* | Severity. OCS ~ F: r = 0.14, p < 0.01; adj β = 0.16, p < 0.01. | M < F severity |
| Storch 2025 | 1805 (1246) | | 31 ± NA (18–88) | | OCD (USA) | *Spanish YBOCS-II* | Severity. F = 20.74 ± 8.76, M = 19.05 ± 8.70; Cohen’s d = 0.19, p < 0.001. | M < F severity |
| Tumkaya 2025 | 460 (256) | | 37 ± 15  (18+) | | GEN (TUR) | VOCI | Severity. F−M d: contam = 0.26 p=0.005; indecis. = 0.28, p=0.003; check/obsess/hoard/just-right/total: \|d\| ≤ 0.12, p ≥ 0.21. | M < F contam/indec  M ~ F: overall/check/ obsess /hoard/just-right |
| Bakhshinejad 2026^B^ | 51 (34) | | 37 ± 10 (18–70) | | OCD  (IRN) | YBOCS | Severity. OCS ~ M: β = −0.59 [95% CI, −2.52–1.33], p=0.54. | M ~ F severity |
| Kavosi  2026^B^ | 340 (264) | | NA  (18+) | | nurses (IRN) | *Padua* | Severity. F > M: t = 2.38, p = 0.018. | M < F severity |
| Qutishat  2026 | 175 (123) | | 29 ± 4 (20–49) | | nurses (OMN) | *OCI-R* | Severity. F = 44.8 ± 18.1, M = 40.2 ± 16.0; F−M d = 0.26, reported p = 0.091 | M ~ F severity |
| Motivation behind symptoms | | | | | | | | |
| Ravid  2013 | 152 (87) | | 15 ± 1  (14-17) | | schools (USA) | *NJRE-YV CL* | Presence. NJRE checklist score ~ Age x F: β= -0.21 p < 0.01.  Severity. NJRE severity score ~ Age x F: β= -0.29 p < 0.01. | NJRE: older M > F  younger M < F |
| Sibrava 2016 | 307 (168) | | 40 ± 13 (19+) | | OCD  (USA) | *Study specific* | Severity. High INC vs. low INC: F:M OR = 1.05 [95% CI, 0.62–1.80], p = 0.89 | incompleteness  M ~ F |
| Kawahito 2026 | 30 (21) | | 32 ± 15 (15–65) | | OCD (JAP) | USP-SPS | Sensory phenomena severity. F vs M: β = 1.38, SE=0.87, t = 1.60, p = 0.12, BC [95% CI, −0.10, 3.15]. | sensory phen M ~ F |
| Latent classes of symptoms | | | | | | | | |
| Atli  2014 | 193  (133) | | 29 ± 10 (16–60) | | OCD  (TUR) | YBOCS CL | Presence. Reactive obs F:M OR = 3.76 [95%CI, 1.91–7.41]; Autogenous F:M OR = 0.24 [95%CI, 0.10–0.60]; Reactive compulsive F:M OR = 0.43 [95%CI, 0.18–1.00]. | autogenous obs M > F  reactive obs: M < F |
| Raines 2014 | 297 (187) | | 32 ± 10  (18-70) | | GEN  (USA) | *DOCS* | Relations among Sx. Pearlson r_M_: Contam ~ Resp ~ Unacc Thought ~ Symm > 0.35 p <0.001; Pearlson r_F_: Contam ~ Resp, Contam ~ Symm, Resp ~ Unacc Thought > 0.28 p <0.001. Pearlson r_M_ vs r_F_ & Wald χ2 p-value: Contam ~ Unacc Thought r_M_ = 0.39 p < 0.001, r_F_ = -0.05 p < 0.50, χ^2^(1)= 10.06 p = 0.002; Resp ~ Symm r_M_ = 0.83 p < 0.001, r_F_ = 0.10 p < 0.17, χ^2^(1)= 26.2 p<0.001, Unacc ~ Symm r_M_=0.44 p<0.001, r_F_=0.08 p < 0.27, χ^2^(1) = 4.3 p < 0.05.  Severity. Mean factor scores M - F: Wald χ^2^(4) =8.61 p =0.07. | correlations:  contam & unacc thought /resp & symm / unacc & symm  M > F |
| Rodgers  2015 | 4547 → 591 | | 20 ‒ 50 | | GEN  (CHE) | *SCL-90-R →* SPIKE | Presence. M:F OR: Pure comp. vs obsess.-comp. = 1.14 [95%CI, 0.44-2.98]. | Pure comp.  M ~ F |
|  | 9829 →1500 | | 20 ‒ 41 | | GEN  (CHE) | *SCL-90-R*  *→* SPIKE | Presence. M:F OR: Pure comp. vs obsess.-comp. = 0.38 [95%CI, 0.16-0.88]. | Pure comp.  M > F |
|  | 5535 →3720 | | 15 ‒ 66 | | GEN  (CHE) | DIGS | Presence. M:F OR: Pure comp. vs obsess.-comp. = 1.71 [95%CI, 0.39-7.58]. | Pure comp.  M ~ F |
| Batmaz  2016t | 180  (109) | | 34 ± 12  (18+) | | OCD  (TUR) | CGI-S | Presence. Autogenous vs. Reactive obs. M:F OR = 0.31 ([95%CI = 0.16 – 0.60], χ^2^ (1) = 12.3 p < 0.001). | autogenous obs  M > F |
| Shkalim  2017 | 233  (133) | | 41 ± 14  (18 – 77) | | Patients  (ISR) | MMPI-2/ *MOCI* | Relations among Sx. Pearlson r_M_: OCD ~ distress d/o = 0.35 p < 0.05, OCD ~ fear d/o= 0.42 p < 0.05, z = -0.64 p =0.52; Pearlson r_F_: OCD ~distress d/o = 0.24 p <0.05, OCD ~ fear d/o = 0.45 p < 0.05, z = -2.07 p < 0.05. CFA: noninvariant across genders ΔꞳ^2^(12) = 21.43 p<0.05, CFA M: unacceptable fit; CFA F: OCD Sx similar loading to distress/fear d/o. | relations of OCD to fear and distress  M ≠ F |
| Kenezloi  2018 | 325  (187) | | 15 ± 4  (10 – 18) | | OCD  (HUN) | CY-BOCS CL | Presence. Obsessive, Compulsive, Cleaning, Sexual M~F (stats not reported) | Sx subtypes M~F: |
| Wang  2021 | 6063  (3330) | | NA  (12-21) | | GEN  (USA) | comp. interview | Interconnectedness (centrality) of symptom. M: OCD-arranging/ordering β=1.8 (3^d^ most important); F: OCD-cleaning/washing β=2.05 (3^d^ most important); “*The network for female youth had a higher global strength than that for males*.” (no stats) | centrality: M arranging/ordering  F cleaning/washing |
| Gemelli 2025 | 61 (52) | | 22 ± 5 | | STU (USA) | ROII | Severity. AO ~ sex (coding not reported): β=0.03, t=0.35, p=0.73; RO ~ sex: β=−0.02, t=−0.17, p=0.86. | Severity AO/RO  M ~ F |
| Assessments | | | | | | | | |
| Allison  2011 | 428F | | 30 ±5  (18+) | | PP  (USA) | *PP distress, OCI-R* | Severity. PDM-OCD ~OCI-R: β = 0.46 p < 0.001 | PDM-OCD↑↑OCI-R |
| Lord  2011 | 168F | | 31 ±5  (18+) | | 67 preg  95 PP | *POCS,*  YBOCS | Severity. POCS = 21.1±8.9, YBOCS = 17.3±10.7, d = 0.39 p < 0.001. | PP OCD severity  POCS >YBOCS |
| Lydsdottir  2013 | 2411F | | 29 ±5  (16+) | | preg w16  (ISL) | *EPDS,* MINI+ | Presence. EPDS ≥ 12: 17.6% OCD | EPDS↑↑OCD |
| Storch  2017 | 67  (33) | | 32 ±12  (18 - 67) | | OCD  (USA) | YBOCS  Self/clinic | Severity. YBOCS-clinician ~– YBOCS-self x F: β= -0.63 p > 0.05 | Self - clinician  M ~ F |
| Hellberg  2018 | 169  (73) | | 20 ±2  (18+) | | students  (USA) | BIS, ATQ, *FMPS, OCI-R* | Severity. OCI-R: Overcontrolled personality (high FMPS + low BIS) = 19.9, Undercontrolled personality (high BIS + low ATQ) = 11.6, Resilient personality (high ATQ, low BIS&FMPS) = 10.1, F>11.4 p <0.0001; OCI-R x cluster x F: F(1, 163) = 2.35, p = 0.26 | Personality → OCD:  M ~ F. |
| Hauschildt  2019 | 128  (86) | | 39 ±11  (18+) | | OCD  (DEU) | YBOCS  Self/clinic | Severity. YBOCS-self – YBOCS-clinician ~ F: p > 0.67 | Self - clinician  M ~ F |
| Lambe 2021 | 350 (175) | | 13 ± 3 (6–21) | | OCD (CAN/USA) | TOCS  parental | Severity. TOCS total/Sx count/subscales ~ F: r ≈ −0.01 to −0.10 p > 0.1 | severity:  M ~ F |
| Skarphedinsson 2021 | 267 (117) | | 12 ± 3 (6–18) | | psychiatric  (SWE) | CBCL OCD, YSR OCD | Severity. Parental reports (CBCL): M = 2.87 ± 2.87, F = 3.31 ± 2.83, p > 0.1.  Youth self-reports (YSR): M = 3.79 ± 2.99, F = 5.30 ± 3.61, p < 0.01 | parental report: M ~ F  self-report: M < F |
| Mahjani 2022 | 1010 (658) | | 22 ± 7  (18+) | | OCD (SWE) | *OCI-R*  *(swedish)* | Severity. OCI-R-swe F = 23.3 ± 13.9; M = 20.9 ± 12.8; p < 0.01. | severity:  M < F |
| Mercan  2023 | 312 (204) | | 21 ± 2  (18–30) | | students  (TUR) | *BALCI*  *self* | Psychometric properties. Measurement invariance (configural, metric, scalar, strict) across genders | psychometrics:  M~F |
| Zhou  2024 | 2221 (1090) | | 6 ± 1  (4-8) | | GEN  (GBR) | *PAS*  *parental* | Psychometric properties. Measurement invariance across genders of OCD subscale. | psychometrics:  M~F |
| Hemade 2025 | 392 (278) | | 21 ± 4  (18+) | | GEN (LBN) | OBQ-9 | Psychometric properties. Configural, metric, and scalar invariance supported across sex. OBQ-9 scores: M=31.8 ± 9.9, F=32.7 ± 10.5, t(390)=−0.76, p=0.45. | psychometrics:  M~F |
| Pinciotti 2025 | 2982 (1892) | | 31±12 (7–86) | | OCD/OCS 13 countries | YBOCS-II  CY-BOCS-II, CGI-S | Severity benchmarks. Non-/subclinical: F = 0–14; M = 0–13; Mild: F = 15–21; M = 14–21; Moderate: F = 22–34; M = 22–37; Severe: F = ≥35; M = ≥38. | Severe cutoff F < M |
| Van Doren 2025 | 292 (233) | | 18 ± 2  NA | | GEN (USA) | *OCI-R* | Psychometric properties. OCI-R showed strict measurement invariance across gender (configural, metric, scalar, residual); statistics NR in main article. | psychometrics:  M~F |
| Hussain 2026 | 237 (179) | | 237 (179) | | school (PAK) | RCADS | Psychometric properties. OCD subscale α=.64; gender was not associated with RCADS OCD classification, χ²=4.02, p=0.13. | psychometrics:  M~F |

A.2.2. Studies of gender effects on additional OCD burden.

| **First Auth, Year** | **N**  **(F)** | **Age mean**±**SD (range)** | | **Pop** | **Meas.** | | **Gender effects (F vs. M)** | | |
| --- | --- | --- | --- | --- | --- | --- | --- | --- | --- |
|  |  |  |  |  |  |  | **Stats** | | **Direction** |
| Burden on patients | | | | | | | | | |
| *all ages* | | | | | | | | | |
| Meier  2016 | 10155  (5935) | wave 1:  0-47  10 waves | | GEN  (DNK) | Danish CRS/PCR/ NPR | | Mortality in OCD. Natural causes MRR: OCD-M = 1.69 [95%CI, 1.19-2.30], OCD-F =1.37 [95%, 0.94-1.93]; Unnatural causes MRR: OCD-M = 2.23 [95%CI, 1.49-3.20], OCD-F =3.15 [95%, 1.87-4.94]. | | Natural: M > F  Unnatural: M < F |
| *children/adolescents* | | | | | | | | | |
| Bortes 2022 | 266664  (130665) | (15-16) | | GEN  (SWE) | medical records | | Educational achievement. Grades ~ OCD: β_F_ = −0.08 p < 0.01, β_M_ = −0.03 p < 0.01; Grades ~OCD x F p = 0.046; Non-passing grade ~ OCD: F OR = 1.38 p < 0.01, M OR = 1.29 p < 0.01; Non-passing grade ~ OCD x F p = 0.64; | | Grades↓↑ OCD: F < M  Non pass↓↑ OCD: F ~M |
| McCarty 2026 | 238 (116) | 13 ± 3  (7-17) | | OCD (10 countries) | YBOCS, CY-BOCS | | Stigma experiences M~F. Stigma did not vary between males and females (t_pooled_ = 0.33, p = .740, d_pooled_ = 0.04) | | M ~ F stigma; stigma experiences |
| Usami 2026 | 136 (66) | 12 ± 2  (6–15) | | OCD (JPN) | QCD parental | | Functioning/overall behavior. Morning, school, after-school, evening, night, and overall functioning ~ F: β = −0.13 to 0.13, all p > 0.05. | | functioning across time of day M ~ F |
| *adults* | | | | | | | | | |
| Rosa  2012 | 815  (475) | 35 ± 14  (18 ‒ 60) | | OCD  (BRA) | *SAS*,  *SF-36*, | | Social maladjustment. Marital role ~ F: β = 0.38 p < 0.001. | | marital maladjst  M < F |
| Kugler  2013 | 102  (49) | 29 ± 11  (18 – 79) | | OCD  (USA) | YBOCS,  *MOS-36* | | Severity. QoL-emotional~ F: t(100)=-2.07 p < 0.05. QoL-social/general/physical ~ F: p > 0.05. | | emotion: M > F  soc/gen/physic: M ~ F |
| Uguz  2014 | 37F | 27 ± 5  (18+) | | OCD  HC  (TUR) | Serum TNF-α concentr. | | Impact of OCD in mothers on newborn babies. OCD – HC mothers d p-value: Serum TNF-α (a marker for chronic inflammation) = 2.0 z = 2.09 p = 0.036. | | OCD mother → chronic inflammation baby |
| Uguz  2015 | 63F | 29 ± 5  (18+) | | OCD  HC  (TUR) | Medical records | | Impact of OCD in mothers on newborn babies. OCD – HC mothers d p-value: Birth weight = 0.85 t = -3.37 p = 0.001; Gestation age = 0.65 z = -2.49 p = 0.013. | | OCD mother → low birth weight baby |
| Ghassemzadeh  2017 | 56  (36) | 34 ± 8  (18+) | | OCD IRN | SCID-IV, IIEF | | Presence of sexual dysfunction in OCD. F:M OR = 12.43 [95%CI, 3.37-45.9]. | | Sex dysf: M < F |
| Jahangard  2018 | 200  (123) | 31 ± 6  (18-65) | | OCD, HC  (IRN) | *SF-36,*  *HAM-D,*  *HAM-A* | | Severity. QoL, controlling for depression and anxiety, Overall/Vitality ~ OCD x F: $\eta_{p}^{2}$ > 0.06, F (1,195) ≥ 4.05 p < 0.05; Physical/Health/Social/Emotional/Mental ~ OCD x F: $\eta_{p}^{2}$ < 0.015, F (1,195) ≤ 3.12 p > 0.05. | | QoL overall/ vitality: HC > OCD-M > OCD-F |
| Pérez-Vigil  2018 | 173,443 | NA  (16-64) | | OCD, GEN  (SWE) | Records multiple population and health registries and databases | | Labor market marginalization, controlled for education, rurality, family composition, region, somatic disorders, OCD vs GEN HR. Disability pension: M = 19.8 [95% CI=17.8-21.9], F = 14.4 [95%CI = 13.3-15.7]; Sickness absence ⩾90 days: M = 4.0 [95% CI=3.8-4.3], F = 2.7 [95%CI = 2.6-2.8]; Unemployment ⩾180 days: M = 1.8 [95% CI=1.6-1.9], F = 1.7[95%CI = 1.6-1.8]; Any type of market marginalization: M = 3.6 [95% CI=3.4-3.8], F = 2.9[95%CI = 2.8-3.0].  Labor market marginalization, controlled for education, rurality, family composition, region, somatic disorders, OCD vs no OCD siblings HR. Disability pension: M = 14.9 [95% CI= 9.9-22.5], F =12.0 [95%CI = 8.7-16.6]; Sickness absence ⩾90 days: M = 4.4 [95% CI= 3.6-5.3], F = 2.8 [95%CI = 2.4-3.2]; Unemployment ⩾180 days: M = 1.9 [95% CI= 1.6-2.4], F = 1.7 [95%CI = 1.4-2.1]; Any type of market marginalization: M = 3.6 [95% CI= 3.1-4.2], F = 2.7[95%CI = 2.4-3.0]. | | Labor market marginalization  M > F |
| Pozza  2019 | 72 (27) | 35 ± 10 (18-58) | | OCD  (ITA) | *OCI-R,*  *SIS/SES* | | Presence. Inhibition due to threat of performance failure ~ F, when controlled for contam: β= 1.12 p = 0.053. | | sexual inhibition  M $\leq$ F |
| Pozza  2020 | 72  (27) | 35 ± 10 (18-58) | | OCD  (ITA) | *OCI-R,*  *SIS/SES* | | Severity of sexual dysfunction in OCD. SIS x M: β = 1.1 p = 0.053 | | sexual inhibition  M < F |
| Pozza  2021 | 144  (54) | 35 ± 10 (18-58) | | OCD  HC  (ITA) | *OCI-R,*  *SIS/SES* | | Severity of sexual dysfunction in OCD. SES/SIS M vs F t-tests: OCD range t (70) = -1.87‒1.40, p=0.16‒0.94; HC range t (70) = -0.35‒2.00, p = 0.05‒0.72). SES/SIS OCD vs HC t-tests: range t (142) =-5.91 ‒ -2.66 p < 0.01. | | sexual response  OCD-M ~ OCD-F < HC-M ~ HC-F |
| Dèttore 2021 | 72  (27) | 35±10  (18-58) | | OCD  (ITA) | *OCI-R*,  *SIS/SES,*  *ASQ* | | Severity of sexual dysfunction in OCD. SE~M: β =-3.24 p =0.022, SES~M x confidence: β =0.06 p=0.02, SE ~ M x discomfort with closeness: β = 0.05 p=0.03; SI-perform ~M x cont. = 0.69 p =0.03; SI-conseq ~M = 3.53 p = 0.01, SI-conseq ~M x confidence = -0.075 p =0.002; SI-conseq ~M x preoccupation with relationships = -104 p <0.001, SI-conseq ~M x relationship as secondary =0.059 p = 0.023, SI-conseq ~Mx cont. = 0.963 p =0.001. | | SI conseq M > F  SE, SI-perform M < F  F: cont ↑↓SI conseq;  M: cont ↑↑ SI-perform, SI-conseq |
| Muhlbauer  2021 | 913  (NA) | NA  (18+) | | OCD  (BRA) | SCID-I | | Marital status. F: single = 46.3%, married = 44.4%, divorced=9.3%; M: single = 59.1%, married = 36.5%, divorced=4.4%; χ^2^ (2) = 17.8 p < 0.001. | | never married  M > F |
| Guazzini 2022 ^S^ | 660  (469) | 25 ± 7  (18+) | | GEN  (ITA) | *VOCI, SM items* | | Perceived impact of social media on mood. M = 2.8 ± 0.8 F = 2.9 ± 0.8 p = 0.27 | | impact soc media on mood: M ~ F |
| Benatti  2022^S^ | 491 (297) | 39 ± 13  (18+) | | OCD  (11 countr.) | YBOCS, MADRS, *SDS* | | Disability. SDS ~F β = 0.02 p = 0.72 | | overall severity  M ~ F |
| Snaychuk  2022 | 1001 (551) | 35 ± 13  (18+) | | OCD  BRA | Interview  YBOCS | | Presence in OCD of Compulsive Sexual Behavior (CSB). OCDnOCSB M=40.7%, F=59.3%, OCDwCSB M=75%, F=25%, χ2 = 16.73, p<0.001 | | CSB in OCD: M > F |
| Karagöz 2021 | 139 (44) | med = 55  (18+) | | STEMI  (TUR) | *PI-WSUR* | | Prehospital delay. F:M OR = 1.34 [95%CI, 0.63–2.83], p = 0.43. | | contam↑↑ prehospital delay: M ~ F |
| Mataix-Cols  2022 | 23126 (13491) | NA  (18–65) | | OCD (SWE) | medical records | | Motor accidents, OCD HR. Serious transport accidents: F = 1.2 [95%CI 1.13–1.28], M = 0.98 [95%CI, 0.91–1.06]; Motor vehicle accidents: F = 1.2 [95%CI, 1.09–1.31], M = 0.92 [95%CI, 0.84–1.02]; Traffic-related convictions: F = 1.26 [95%CI, 0.81–1.96], M = 1.10 [95%CI, 0.88–1.38]. | | F: OCD↑↑ motor accidents |
| Zerdziński  2022 | 75  (40) | 45 ± NA  (24–73) | | OCD, HC  (POL) | *OHQ-23,*  *ASEX* | | Severity. Happiness / QoL: F vs M p > 0.60  Prevalence. %Sexual dysfunction (ASEX): F = 0.78 M = 0.54, F:M OR = 2.9 | | QoL: M ~ F  Sex dysf: M < F |
| Lu  2023 | 293 (138) | 24 ± 7  (18+) | | OCD (CHN) | *ACIPS* | | Social anhedonia. M = 66.8 ± 14.1; F = 67.3 ± 13.8; t = −0.28, p = 0.78. | | Soc Anh:  M ~ F |
| Friedman-Ezra 2024 | 363 (204) | 37 ± 11 (18–79) | | OCD (NLD) | *DJGLS,* YBOCS | | Loneliness. %Severe: M = 0.0.3 F = 0.0.27; %Moderate M = 0.52 F = 0.41; %None: M = 0.18 F = 0.33; χ²(2) = 9.98, p < 0.01. | | loneliness:  M > F |
| Mancin 2024 | 211 (150) | 30 ± 12 (19–67) | | GEN  (ITA) | *ROCI, PROCSI* | | Relationship/partner OCD. ROCI ~ F β = −0.05 p 0.94; PROCSI~ F β = -0.06 p = 0.39 | | relationship/partner OCD: M ~ F |
| Puşuroğlu  2024 | 87  (63) | 33 ± 10  (18–59) | | OCD  (TUR) | YBOCS, *BPAQ* | | Physical aggression. M = 21.29; F = 18.17; Mann–Whitney U p = 0.038 | | agress: M > F |
| Yalım  2024 | 183 (99) | 33 ± 9 (18–65) | | OCD  (TUR) | YBOCS*, LIFE-BASE* | | Occupational dysfunction. F:M OR= 8.11 [95%CI,4.17–15.79], p <0.001; adj. F: M OR = 6.63 [95%CI, 3.27–13.61]. | | occupational dysf.:  M < F |
| Shmidova 2026 | 11M | 25 ± NA (18–35) | OCD  (RUS) | | | *interviews* | | Stigma/help-seeking, emergent themes. making sense of OCD; disclosure and recognition; coping and control; social reactions and support. Cross-cutting themes included anticipated/enacted stigma, selective disclosure, masculine norms discouraging help-seeking, mistrust of psychiatry, and different stigma experiences by diagnostic status—self-identified men sought validation, whereas clinically diagnosed men described greater stigma tied to formal labeling. | M: stigma/help-seeking burden |
| Kavosi 2026^S^ | 340 (264) | NA  (18+ ) | | nurses (IRN) | survey | | Job performance. F vs. M: t = 1.21, p = 0.23. | | M ~ F job performance |
| Bakhshinejad 2026^S^ | 51 (34) | 37 ± 10 (18–70) | | OCD  (IRN) | YBOCS | | Quality of life. M vs F: β = 5.96 [95% CI, 0.27–11.67], p = 0.04. | | M > F quality of life |
| Burden on caregivers | | | | | | | | | |
| *children/adolescents* | | | | | | | | | |
| DuBois  2025 | 61  (38) | 12 ± 2 (7–17) | OCD, parents  (USA) | | | *CAS-P OCD, FASA, CBCL* | | Cross-sectional. OCS ~ Family accommodation (FA) patient: M β = 0.11 p = 0.02, F β = −0.04 p = 0.46; OCS ~ FA x M: β = −0.15, p = 0.01. | M: OCS ↑↑FA  F: OCS ↑↓FA |
| Skarphedinsson 2025 | 238 (114) | 13 ± 3  (7–17) | OCD parents (DNK/ NOR/SWE) | | | *CY-BOCS FAS*  *COIS-R* | | Cross-sectional. OCS severity ~ gender, r: CY-BOCS total r = 0.029; obsess = 0.030; compul. = 0.024, all p > 0.05. Family accommodation ~ gender: r = −0.036, p > 0.05. | M ~ F: FA |
| *adults* |  |  |  | | |  | |  |  |
| Vikas  2011 | 32 /32  (NA) | 30 ±11  (18-60) | | OCD/ CARE  (IND) | *WHOQOL-Bref-Hindi, FAS* | | Social maladjustment in OCD. Marital role ~ F: β = 0.73 p < 0.01. Burden on caregivers. Subjective burden ~ F: β = 0.37 p < 0.05. | | marital maladj  OCD M < F;  subj burden  CARE M < F |
| Siu  2011 | 77  (41) | 48 ± 14  (18-79) | | CARE  (CHN) | YBOCS,  GAF | | Burden on caregivers. Subjective burden ~ F: U(77) = 567 p = 0.038. | | subj burden  M < F |
| Futh  2012 | 70  (41) | 48 ± 6  (18+) | parents  (GBR) | | | *DASS-21*,  *FAS* | | Cross-sectional, mothers-fathers d: Stress = 0.38 z = 1.95 p = 0.05; Anxiety = 0.29 z = 2.10 p = 0.035; Depression = 0.39 z = 1.47 p = 0.17; FA = 0.34 t(68) = 1.38 p = 0.17. | stress/anx M < F  depress/FA M ~ F |
| Yanagisawa  2015 | 256  (143) | 30 ± 9  (18+) | OCD  (JAP) | | | interviews,  2 clinicians | | Burden on caregivers. Involved behavior by OCD pts F: M OR = 4.12 [95%CI, 2.21-7.67], p<0.001. | Involved behav.  M < F |
| Shrinivasa  2020 | 100  (43) | 28 ± 9  (18+) | OCD  (IND) | | | *FAS*  WSAS | | Cross-sectional. Family accommodation (FA) ~ F patient d = 0.41 p = 0.051. FA ~ F patient β = 0.14 p = 0.21, when controlled for relations with caregiver (spouse > parent β = 0.20 p =0.023), social adjustment (β = 0.43 p< 0.001), and contam (β = 0.21 p< 0.01). | FA  M ≤ F |
| Liao  2021 | 109/91 (55/48) | 31 ± 10 /  43 ± 10  (18+) | OCD/  CARE  (CHN) | | | YBOCS,  *FAS-IR,*  *FAD-12* | | Cross sectional. Family accommodation (FA) ~ F patient median: M = 11, F = 14 z = 1.968 p = 0.05. | FA  M < F |
| Kuru  2023 | 92  (69) | 32 ± 12 (18–68) | OCD  (TUR) | | | *FAS-PV* | | Cross sectional. Family accommodation (FA) ~ F patient: M = 22.8 ± 17.7; F = 24.3 ± 14.5; t(90) = −0.42, p = 0.68. | FA  M ~ F |
| Kelley  2024 | 315 (159) | 30 ± 10  (18+) | OCD, IRT (USA) | | | *FAS-PV* | | Cross sectional. Family accommodation (FA) ~ F patient: F = 22.2 ± 17.9; M = 16.9 ± 15.1; t(306.2) = 2.85, p < 0.01 | FA  M < F |
| Perspectives in general population | | | | | | | | | |
| Coles  2013 | 577  (322) | 18+ | | GEN  (USA) | AKS, vignettes | | Public perception. Recognized OCD: M = 33.1% F = 33.9% n.s.; Recommend therapy: M = 89.2% F = 89.7% n.s., Therapy is helpful: M = 89.0% F = 90.3% n.s. | | recognition/ perception M ~ F |
| Chong  2016 | 3006  (1506) | 41 ± NA  18-65 | | GEN  (SGP) | vignettes | | Public perception. Recognized OCD F:M OR =2.9% [95%CI, 1.5-5.6] | | recognition M < F |
| García-Soriano  2017 | 102  (51) | 17 ± 1  (16-19) | | students  (ISP) | vignettes, Q-8-C | | Public perception. Recognized: order-OCD M = 86.3% F = 82.4% n.s., aggres-OCD M = 23.5% F = 29.4% n.s.; Recommend psychologist/psychiatrist: order-OCD M = 62.7/19.6%% F= 78.4/15.7% n.s., aggres-OCD M = 51/41.2% F = 47.1/49 n.s.  Stigma. Want to help: M = 5.85±0.29 F = 7.26±0.30 η^2^ = 0.10 p = 0.001, Want to help x aggress-OCD: M = 5.74±0.33 F = 7.40±0.34 η^2^ = 0.11 p = 0.001. | | recognition  M ~ F  want to help  M < F |
| Stewart  2019 | 806  (427) | 49 ± 18  (18-93) | | GEN  (USA) | *open-end questions* | | Public perception, M:F OR. Defined OCD: perfectionism = 1.76 p < 0.05, compulsions = 1.1 p > 0.10, don’t know = 1.31 p >0.10; Recognized OCD =0.86 p > 0.10, OCD is not just obsessive-compulsive behaviors = 1.24 p > 0.10, OCD is more extreme = 1.27 p >0.10 | | Recognition M ~ F  OCD = perfectionism  M > F |
| Shahwan 2026 | 3002 (1534) | NA  (18–67) | | GEN (SGP) | vignettes | | Public perception. Correct recognition of OCD ~ F: OR = 2.33 [95% CI, 1.27–4.25], p = 0.006 | | M < F recognize OCD |

**Abbreviations used in Tables A2.1 - A2.2.**

**Instruments: ACIPS –** Anticipatory and Consummatory Interpersonal Pleasure Scale**;^1^ ADIS-5-L –** Anxiety and Related Disorders Interview Schedule for DSM-5, Lifetime version;^2^ **AKS -** Anxiety Knowledge Survey (study specific); **ASEX** – Arizona Sexual Experience Scale;^3^ **ASOC** – Revised Arabic Scale of Obsession Compulsion;^4^ **ASQ** - The Attachment Style Questionnaire;^5^ **ATQ** - the adult temperament questionnaire-short form;^6^ **BABS** impression - Brown Assessment of Beliefs Scale;^7^ **BAI** – Beck Anxiety Inventory;^8^ **BALCI** – (Turkish) Obsessive–Compulsive Symptoms Scale (study specific version of **^9^**); **BDI** – Beck Depression Inventory [version NR in Kim et al., 2022]; **BIS** - The Barratt Impulsiveness Scale-11;^6, 10^ **BPAQ** – Buss–Perry Aggression Questionnaire;^11^ **CAS-P OCD** – Child Accommodation Scale – Parent version (OCD-specific);^12^ **CBCL/** **CBCL-OCS** Child Behavior Checklist (OCS);^13^ **Child Sheehan Disability Scale**^;14^ **CGI-S** - Clinical Global Impressions–Severity scale;^15^ **COIS-R** – Child Obsessive-Compulsive Impact Scale–Revised;^16^ **CY-BOCS** - the Children’s Yale-Brown Obsessive-Compulsive Scale;^17^ **CY-BOCS CL** - the Children’s Yale-Brown Obsessive-Compulsive checklist;^17^ **CY-BOCS-II** - Children’s Yale-Brown Obsessive-Compulsive Scale Second Edition;^18^ **Danish CRS** - the Danish Register of Causes of Death;^19^ **Danish PCR** - the Danish Psychiatric Central Register;^20^ **Danish NPR** – the Danish National Patient Register;^21^ **DASS-21**- the short‐form version of the Depression Anxiety Stress Scales;^22^ **DIGS** – Diagnostic Interview for Genetic Studies;^23^ **DJGLS** – de Jong-Gierveld 11-item Loneliness Scale;^24^ [ref].**DOCS** - Dimensional Obsessive-Compulsive Scale;^25^ **DPSS-R**-12 – Disgust Propensity and Sensitivity Scale – Revised (12-item);^26^ **DSM-IV** - Diagnostic and Statistical Manual of Mental Disorders, fourth edition;^27^ **DWDS** - DetectaWeb-Distress Scale;^28^ **DYBOCS** - The Dimensional Yale–Brown Obsessive–Compulsive Scale;^29^ **EPDS** - the Edinburgh Postnatal Depression Scale;^30^ **FAD-12** - the Family Assessment Device-12 Item Version (study specific version of ^31^); **FAS** - Family Accommodation Scale;^32^ **FAS-PV** – Family Accommodation Scale for Obsessive-Compulsive Disorder–Patient Version;^33^ **FASA** – Family Accommodation Scale – Anxiety;^32^ **FAS-IR** - the Family Accommodation Scale for OCD Interviewer-Rated;^34^ **FMPS** - The Frost Multidimensional Perfectionism Scale;^35^ **GAF** - the Global Assessment of Functioning scale;^27^ **HAM-A** - The Hamilton Anxiety Scale;^36^ **HAM-D** – The Hamilton Depression Scale;^37^ **HS** – Heterosexism Scale;^38^ **IIEF** – International Index of Erectile Function;^39^ **LIFE-BASE** – Longitudinal Interval Follow-up Evaluation – Baseline;^40^ **MADRS** – Montgomery–Åsberg Depression Rating Scale;^41^ **MINI+** - the Mini-International Neuropsychiatric Interview-Plus;^42^ **MMPI-2** - Minnesota Multiphasic Personality Inventory-2;^43^ **MOCI** - the Maudsley Obsessional–Compulsive Inventory;^44^ **MOS-36** - RAND MOS-36 Item Short Form Health Survey;^45^ **NJRE-YV CL** - Youth Version of the Not Just Right Experiences Questionnaire--Revised^46^ checklist (study specific); **OBQ-46** – Obsessive Beliefs Questionnaire-46;^47^ **OBQ-CV** - a child version of the Obsessive Beliefs Questionnaire;^48^ **OBQ-9** - Obsessive Beliefs Questionnaire;^49^ **OC-CDS** – Obsessive–Compulsive Core Dimensions Scale;^50^ **OCI-4** – four-item Obsessive-Compulsive Inventory;^51^ **OCI-CV** – Obsessive-Compulsive Inventory, child version;^52^; **OCI-R** – Obsessive Compulsive Inventory (revised);^53^ **OHQ-23 -** Oxford Happiness Questionnaire;^54^ **Padua Inventory;**^9^ **PI-WSUR** – Padua Inventory–Washington State University Revision;^55^ **PAS –** Preschool Anxiety Scale;^56^ **PDM/ PDM-OCD** – the Postpartum Distress Measure (includes OCD-related symptom items);^57^ **PIOS** – the Penn Inventory of Scrupulosity;^58^ **POCS** - Perinatal-specific OCD symptom scale;^59^ **PROCSI** - Partner-Related Obsessive–Compulsive Symptoms Inventory;^60^ **Q-8-C** - Attribution Questionnaire–Child version (8-item);^61^ **QCD** – Questionnaire–Children with Difficulties;^62^ **QUIT** – Questionnaire of Unwanted Intrusive Thoughts;^63^ **RCADS** – Revised Child Anxiety and Depression Scale;^64^ **ROCI –** Relationship Obsessive Compulsive Inventory;^65^ **ROII –** Revised Obsessional Intrusion Inventory;^66^ **SAS** - Social Adjustment Scale;^67^ **SCAS** - Spence Children’s Anxiety Scale;^56^ **SCID-I/IV** - Structured Clinical Interview for DSM Axis I Disorders^68^; **SCL-90-R** – Symptom Checklist-90-Revised;^69^ **SDS** – Sheehan Disability Scale;^70^ **SF-36** – the 36-Item Short Form Health Survey;^45^ **SIBS** – Spiritual Involvement and Beliefs Scale;^71^ **SIS/SES** – Sexual Inhibition Scale/Sexual Excitation Scale;^72^ **SI-conseq** – Sexual inhibition due to the threat of performance consequences subscale;^72^ **SI-perform** - Sexual inhibition due to the threat of performance failure subscale;^72^ **SOCDS –** Short Obsessive Compulsive Disorder Screener;^73^ **SORT** – Sexual Orientation Obsessive–Compulsive Scale;^74^ **SPIKE** – Screening Psychopathology Interview;^75^ **TOCS** – Toronto Obsessive–Compulsive Scale;^76^ **USP-SPS** – University of São Paulo Sensory Phenomena Scale;^77^ **VOCI –** Vancouver Obsessional Compulsive Inventory;^78^ **WHOQOL-BREF** - World Health Organization Quality of Life instrument, brief version;^79, 80^ **WHOQOL-Bref-Hindi** - Hindi Version of ^79, 80^; **WSAS** -Work and Social Adjustment Scale;^81^ **YBOCS** – the Yale-Brown Obsessive Compulsive Scale;^82, 83^ **YBOCS CL** - the Yale-Brown Obsessive Compulsive Scale, check list;^82, 83^ **YBOCS-II** - Yale-Brown Obsessive Compulsive Scale–Second Edition;^84^ **YSR** – Youth Self-Report.^85^

**Other abbreviations: 95%CI** - 95% Confidence Interval; **Adj.** – Adjusted (e.g., adjusted OR); **Aggress.** – Aggressive obsessions; **ANX** – anxiety; **AO** – autogenous obsessions; **BC** – bias-corrected; **CA** – Correspondence analysis; **CARE** – caregivers; **CFA** – Confirmatory factor analysis; **Check.** – Checking compulsions; **CL** – Checklist; **Clinic.** - Clinical assessments; **CogDys** – Cognitive Dysfunction; **Contam.** – Contamination obsessions; **Count.** – Counting compulsions; **CSB** – compulsive sexual behavior; **d** – effect size Cohen’s d; **Depress.** – Depression; **Dx** – Diagnosis; **FA** – Family Accommodation; **FDR** – false discovery rate; **FDRs** - first-degree relatives; **Fetus neurodev.** - Fetal neurodevelopment; **Forbid thoughts** - Forbidden thoughts; **GAD** – generalized anxiety disorder; **GEN –** general population; **HC** – Healthy controls; **Hoard** – hoarding obsessions and compulsions; **HR** – Hazard ratio; **INC** – incompleteness; **Involved behav.** - Involved behaviors; **IQR** – Interquartile range; **IRT** – intensive residential treatment; **LGBT** - lesbian, gay, bisexual, and transgender; **Marital maladj** - marital maladjustment; **med** – Median; **Misc.** – Miscellaneous obsessions; **MRR** - mortality rate ratio; **NA** – not available; **NR** – not reported; **n.s.** – Not significant; **Obs.** – obsessions; **OCD** – obsessive compulsive disorder; **OCS** – obsessions and compulsions; **OR** – Odds ratio; **Order.** – ordering compulsions; **Physic** – Physical; **PP** – Postpartum; **PPV** – Positive predictive value; **Preg** – Pregnancy; **QoL** – Quality of Life; **Relig.** – Religious obsessions; **Repeat.** - Repetitive behaviors; **Resp. for Harm.** – Responsibility for Harm; **RO** – reactive obsessions; **SE** – standard error; **Serum TNF-α concentr.** - Serum concentrations of tumor necrosis factor-α; **Sex.** – sexual obsessions; **Soc.** – social; **STEMI** – ST-elevation myocardial infarction; **STU** – students; **subj burden** – subjectively reported burden; **Sx** – Symptoms; **Symm.** – symmetry obsessions; **Unacc thoughts** – obsessions with unacceptable thoughts; **Vascular compl.** - Vascular complaints; **Wash** – washing compulsions; **M** – males; **F** – females.

**Appendix 3** Papers that addressed gender differences in epidemiology of obsessive-compulsive disorder.

Table A3.1. Gender differences in incidence and prevalence of OCD.

| **First Auth, Year** | **N** | **Age range** | **Pop** | **Sampling** | **Instr.** | **Assessors** | **Meas.** | **Gender effects** | | | | | | | | | | | | | |
| --- | --- | --- | --- | --- | --- | --- | --- | --- | --- | --- | --- | --- | --- | --- | --- | --- | --- | --- | --- | --- | --- |
|  |  |  |  |  |  |  |  | **Stat: % F, M** | | **Stat. test** | | | | | **p-value** | | **F:M OR** | | | | |
| Incidence | | | | | | | | | | | | | | | | | | | | | |
| *all ages* | | | | | | | | | | | | | | | | | | | | | |
| Veldhuis  2011 | 577085 | 0+ | primary h/c, (NLD) | Integrated Primary Care Information database, 1996-2007 | patient's history records | medical records | 1^st^ time OCD | F = 0.20 [95%CI, 0.12-0.23]  M = 0.12 [95%CI, 0.10-0.14] | | | | | | p < 0.01 | | | 1.77 | | | | |
| Huang  2014 | 1000000 | 6+ | inpatients,  outpatients  (TWN) | general population registry-based prospective cohort design, 2000-2008 | ICD-9-CM | medical records: ambulatory,  hospital,  prescriptions. | 1^st^ time OCD ≤6 | F = 0.028  M = 0.027 | | [95%CI, 1.10 – 1.20] | | | | | | | 1.2 | | | | |
|  |  |  |  |  |  |  | 1^st^ time OCD age 6-10 | F = 0.008  M = 0.014 | | [95%CI, 0.34-0.91] | | | | | | | 0.6 | | | | |
|  |  |  |  |  |  |  | 1^st^ time OCD age 11-17 | F = 0.016  M =0.026 | | [95%CI, 0.45-0.83] | | | | | | | 0.6 | | | | |
|  |  |  |  |  |  |  | 1^st^ time OCD age 18-24 | F = 0.034  M =0.046 | | [95%CI, 0.59-0.91] | | | | | | | 0.8 | | | | |
|  |  |  |  |  |  |  | 1^st^ time OCD age 25-34 | F = 0.035  M = 0.036 | | [95%CI, 0.77-1.11] | | | | | | | 1.0 | | | | |
|  |  |  |  |  |  |  | 1^st^ time OCD age 35-44 | F = 0.036  M = 0.026 | | [95%CI, 1.11-1.67] | | | | | | | 1.4 | | | | |
|  |  |  |  |  |  |  | 1^st^ time OCD age 45-64 | F = 0.031  M = 0.022 | | [95%CI, 1.11-1.67] | | | | | | | 1.4 | | | | |
|  |  |  |  |  |  |  | 1^st^ time OCD age > 64 | F = 0.020  M = 0.019 | | [95%CI, 0.77-1.43] | | | | | | | 1.1 | | | | |
|  |  |  |  |  |  |  | current OCD ≥ 6 | F = 0.060  M = 0.070 | | [95%CI, 0.83-0.91] | | | | | | | 0.8 | | | | |
|  |  |  |  |  |  |  | current OCD age 6-10 | F = 0.012  M = 0.018 | | [95%CI, 0.43-1.0] | | | | | | | 1.0 | | | | |
|  |  |  |  |  |  |  | current OCD age 11-17 | F = 0.028  M = 0.049 | | [95%CI, 0.43-0.71] | | | | | | | 0.6 | | | | |
|  |  |  |  |  |  |  | current OCD age 18-24 | F = 0.068  M = 0.123 | | [95%CI, 0.48-0.63] | | | | | | | 0.6 | | | | |
|  |  |  |  |  |  |  | current OCD age 25-34 | F = 0.078  M = 0.106 | | [95%CI, 0.67-0.83] | | | | | | | 0.7 | | | | |
|  |  |  |  |  |  |  | current OCD age 35-44 | F = 0.089  M = 0.079 | | [95%CI, 1.0-1.25] | | | | | | | 1.1 | | | | |
|  |  |  |  |  |  |  | current OCD age 45-64 | F = 0.065  M = 0.055 | | [95%CI, 1.0-1.25] | | | | | | | 1.2 | | | | |
|  |  |  |  |  |  |  | current OCD age > 64 | F = 0.041  M = 0.041 | | [95%CI, 0.77-1.25] | | | | | | | 1.0 | | | | |
| Teixeira  2022 | 10463672 | 0+  (53±NA) | GEN  (USA) | insurance claims 2018–2020 | ICD-10 | medical records | 1st time OCD | | not reported | [95%CI, 0.94–1.60] | | | | | | | | | | | 1.2 |
| *children/adolescents* | | | | | | | | | | | | | | | | | | | | | |
| Rintala  2017 | 139017 | 10, 15 | live births 1987–2001 (FIN) | general population registry-based cohort design, 1987–2001 | hospital discharge register | medical records from specialty clinics | 1^st^ time OCD by 10y | born 1987-89: F = 0.01 M = 0.02 | | | | | | p < 0.01 | | | 0.43 | | | | |
|  |  |  |  |  |  |  |  | born 1999-01: F = 0.06 M = 0.08 | | | | | | p < 0.01 | | | 0.69 | | | | |
|  |  |  |  |  |  |  | 1^st^ time OCD by 15y | born 1987-89: F = 0.09 M = 0.12 | | | | | | p < 0.01 | | | 0.69 | | | | |
|  |  |  |  |  |  |  |  | born 1999-01: F = 0.28 M = 0.24 | | | | | | p > 0.10 | | | 1.18 | | | | |
| Dalsgaard 2019 | 1300000 | 0-18 | liveborn singletons 1995- 2016  (DNK) | The Danish Civil Registration System | registers  medical codes | medical records | 1^st^ time OCD | F = 0.96 [95%CI, 0.92-1]  M = 0.63 [95%CI, 0.56-0.72]  “*before age 10: equal, ages 10 – 18: increases faster in girls, by age 18: higher in girls*” | | | | | | | | | 1.5 | | | | |
| Steinhausen  2019 | 67347 | 0-18 | GEN  (DNK) | nationwide Danish registries, born 1995, longitudinal until 18 | registers  medical codes | medical records | 1^st^ time OCD | F = 0.0063  M = 0.0046 | | χ2 = 9.18 p=0.002, *“incidence rate is higher in females starting age 14”* | | | | | | | 1.4 | | | | |
| Ask  2020 | total population | 3-17 | GEN  (NOR) | database for reimbursement of health expenses, 2008-2015 | registers  medical codes | medical records | 1^st^ time OCD | F: ‘2010’ = 0.0008, ‘2011’ = 0.0008, ‘2012’ = 0.0009, ‘2013’ = 0.001, ‘2014’ = 0.0012, ‘2015’ = 0.0012;  M: ‘2010’ = 0.00055, ‘2011’ = 0.0006, ‘2012’ = 0.0007, ‘2013’ = 0.0007, ‘2014’ = 0.0008, ‘2015’ = 0.0008 | | | | | | | | | 1.3 – 1.5 | | | | |
| *adults* |  |  |  |  |  |  |  |  | | | | | | | | |  | | | | |
| Guimarães  2023 | 1560 (880) →  1244 (721) | 21±2  →  26±2 | GEN (BRA) | 1993 Pelotas Birth Cohort | MINI → MINI-Plus | trained prsnl,  interviews | new OCD at FU | | F = 0.026  M = 0.009 | [95%CI, 1.05–7.47] p=0.039 | | | | | | | | | 2.8 | | |
| Stein  2025 [A3.2] | 26136 | (18+) | GEN  (woldwide) | multistage clustered-area probability household sampling | CIDI 3.0,  YBOCS | clinical interviews | 1st time OCD | | [95%CI, 1.00–1.60], χ2=5.3 p = 0.02 | | | | | | | | | | | 1.30 | |
|  |  |  |  |  |  |  | YBOCS ≥16 | | [95%CI, 0.70–1.60], χ2=0.1 p > 0.05. | | | | | | | | | | | 1.10 | |
|  |  |  |  |  |  |  | 12-m persistence | | [95%CI, 0.60–1.50], χ2==0.1 p > 0.05 | | | | | | | | | | | 0.90 | |
| Prevalence in general population | | | | | | | | | | | | | | | | | | | | | |
| Smoller  2026 | 12504 (cis=8336  gnc =2125  trans=2043) | 12 + | GEN  (USA) | medical records  2019-2023 | registers  medical codes | medical records | 5-year EHR OCD  (age adj.) | | cis= 0.49  gnc =2.0  trans=2.3 | GNC:cis = 5.52 [95% CI, 3.60–8.44]  trans:cis = 4.94 [95% CI, 3.18–7.67]  gnc:trans =1.12 [95% CI, 0.74–1.67] | | | | | | | | | | | |
| Mortier  2026 | 645,571 (340,317) | 10 + | GEN  (ESP) | Database review / diagnostic codes→ weighted population estimates | registers  medical codes | medical records | recorded OCD  (2014–2019) | | F = 0.4  M = 0.4 | no formal tests | | | | | | | | 1.0 | | | |
| *children/adolescents* | | | | | | | | | | | | | | | | | | | | | |
| Nikolajsen 2011 | 2522 | 8-17 | students  (ESP) | school-based sampling, 15 schools | SCAS | SR | current clinic OCs | F = 4.9  M = 4.5 | | NA, *“n.s.”* | | | | | | | 1.1 | | | | |
| Orgilés  2012 [A3.1] |  |  |  |  |  |  |  |  |  |  |  |  |  |  |  |  |  |  |  |  |  |
| Shams  2011 | 909 | 14-18 | students  (IRN) | clustered, random, two stage sampling | MOCI → SCL-90-R | SR | 1-w clinic OCs | F = 6.5  M = 4.7 | | χ2 = 0.432 | | | | | p=0.51 | | 1.4 | | | | |
| Canals  2012 | 2023 | 9-13 | students  (ESP) | two step sampling, | LOI-CV →MINI-KID &CSI-4 parent | SR | current OCD | F = 1.9  M = 1.6 | | χ2 = 0.02 | | | | | p = 0.89 | | 1.2 | | | | |
|  |  |  |  |  |  |  | current subOCD | F = 3.8  M = 7.6 | | χ2 = 9.43 | | | | | p = 0.002 | | 0.5 | | | | |
|  |  |  |  |  |  |  | current OCs | F = 4.7  M = 4.8 | | χ2 = 0.01 | | | | | p = 0.92 | | 1.0 | | | | |
| Vivan 2014 | 2323 | 14 ‒ 17 | students  (BRA) | school-based cluster two stage sampling  (7 high schools) | OCI-R→ SCID-TCI, KSADS-P, YBOCS | SR→  clinicians, interviews | 1-mo OCs | F = 24.8  M = 14.4 | | z = 4.25 | | | | | p < 0.001 | | 1.95 | | | | |
|  |  |  |  |  |  |  | current OCD | F = 4.9  M = 1.4 | | z = 6.17 | | | | | p < 0.001 | | 3.36 | | | | |
| Jaisoorya, 2015 | 7380 | 12-18 | students  (IND) | school-based sampling | CIS-R | SR | current OCD | F = 0.5  M =1.1 | | χ2 = 7.98 | | | | | p = 0.005 | | 0.5 | | | | |
| Zijlmans  2017 (com) | 4632 | 8-11 | students  (NLD) | subsampling, Generation R prospective cohort | SOCS | parental reports | current OCs | F = 4.3  M = 4.8 | | χ2 = 0.67 | | | | | p=0.42 | | 0.9 | | | | |
| Mohammadi  2021 | 29858 | 6-18 | Gen  (IRN) | Multistage cluster random sampling | K-SADS-PL | trained prsnl,  interviews | OCD | | F = 3.8%  M = 3.1 | [95% CI 1.10–1.48] | | | | | | | | | 1.27 | | |
| Dissanayake  2024 | 8927 | 6-17 | GEN  (CAN) | representative  community based  Spit for Science study | CBCL/6–18, SR | parental reports | current OCD | | NA | | | | | | | p =0.15 | | | 0.65 | | |
| Khan  2021 | 1783 | 15-19 | students  (SAU) | random sampling (preexisting Dx excluded) | PHQ-9, YBOCS | SR | subOCs  (8-15) | F = 36.6  M = 37.3 | | χ2 = 6.75 | | | | | p = 0.02 | | 1.0 | | | | |
|  |  |  |  |  |  |  | modrt OCs  (16-23) | F = 21.9  M = 15.8 | | χ2 = 0.10 | | | | | p = 0.75 | | 1.4 | | | | |
|  |  |  |  |  |  |  | severe OCs  (24-31) | F = 6.4  M = 3.5 | | χ2 =10.93 | | | | | p < 0.001 | | 1.8 | | | | |
|  |  |  |  |  |  |  | extr OCs  (32-40) | F = 0.96  M = 1.05 | | χ2 = 7.97 | | | | | p = 0.005 | | 1.1 | | | | |
|  |  |  |  |  |  |  | clinic OCs  (16-40) | F = 29.4  M = 20.3 | | χ2 = 0.03 | | | | | p = 0.85 | | 1.4 | | | | |
| *adults* | | | | | | | | | | | | | | | | | | | | | |
| Subramaniam  2012 | 6166 | 18 ‒ 70 | GEN  (GRC) | stratified sampling, 2009-2010 | CIDI 3.0 | trained prsnl,  interviews | life  OCD | F = 3.2  M = 2.8 | | χ2 not reported | | | | | p = 0.59 | | 1.1 | | | | |
| Adam  2012 | 4181 | 18 ‒ 65 | GEN  (DEU) | representative, two step sampling, 1997 | CID-S, DIA-X/ M-CIDI | clinicians, interviews | 12-mo  OCD | F = 0.9  M = 0.6 | | [95%CI,  0.7-3.3] | | | | | p > 0.10 | | 1.6 | | | | |
| Skapinakis  2013, 2019 | 4894 | 18 ‒ 70 | GEN  (GRC) | representative sampling, 2009-2010 | CIS-R | computerized SR supervised by trained prsnl | 1 wk  OCD | F = 2.10  M =1.3 | | χ2 = 5.06 | | | | | p = 0.025 | | 1.7 | | | | |
|  |  |  |  |  |  |  | 1 wk  OCD | F = 3.4  M = 2.1 | | χ2 = 7.63 | | | | | p = 0.006 | | 1.6 | | | | |
| Fineberg  2013 | 591→ 335 | 20/21 → 49/50 | GEN  (SWE) | two-step sampling, Zurich Cohort Study of Young Adults, 7w | SCL-90-R,  SPIKE, interviews | clinicians, interviews | 12-mo OCs | F = 8.8  M =13.7 | | z = 3.44 | | | | | p < 0.001 | | 0.6 | | | | |
|  |  |  |  |  |  |  | 12-mo subOCD | F = 9.3  M =10.1 | | z = 0.87 | | | | | p = 0.38 | | 0.9 | | | | |
|  |  |  |  |  |  |  | 12-mo OCD | F = 5.3  M = 1.7 | | z = 4.16 | | | | | p < 0.001 | | 3.2 | | | | |
| Karg  2014 | 5653 | 18+ | noninstit.  (USA) | multistage cluster sampling, 2008–2012 | modified SCID I | clinicians, interviews | est. 12-mo OCD | F = 0.5  M = 0.1 | | χ2 = 236283 | | | | | p < 0.001 | | 5.0 | | | | |
| Rodgers  2015 | 4547 → 591 | 20 ‒ 50 | GEN  (CHE) | two step sampling,  2010–2012 | SCL-90-R → SPIKE | clinicians, interviews | 7 years OCD | 7.8 | | χ2= 1.83 | | | | | p = 0.18 | | 1.9 | | | | |
|  | 9829 → 1500 | 20 ‒ 41 | GEN  (CHE) | two step sampling,  1979–2008 | SCL-90-R  →SPIKE | clinicians, interviews | 12-mo OCD | 5.8 | | [95%CI, 0.5-1.5] | | | | | | | 0.8 | | | | |
|  | 5535 → 3720 | 15 ‒ 66 | GEN  (CHE) | two step sampling,  2003-2006 | DIGS | clinicians, interviews | 12-mo  OCD | 1.3 | | [95%CI, 0.94-3.17] | | | | | | | 1.7 | | | | |
| Hajebi  2018 | 7886 | 15 ‒ 64 | GEN  (IRN) | representative sampling, 2011 | CIDI 2.1 | clinicians, interviews | 12-mo OCD | F = 6.8  M = 3.4 | | χ2= 44.36 | | | | | p < 0.001 | | 2.0 | | | | |
| Osland  2018 | 25097 | 15+ | GEN  (CAN) | representative sampling, 2012 | SR of lifetime clinician-given OCD Dx | | est. 12-mo OCD | F = 1.04 [95%CI, 0.75-1.25]  M = 0.81 [95%CI, 0.60-1.03] | | | | | | | | | 1.3 | | | | |
| Shirzadi  2019 | 2102 →  822→ 139 | 20 ‒ 73 | GEN  (IRN) | two step sampling, Kermanshah county | GHQ-28 →SCID I&II | clinicians, interviews | est. 12-mo OCD | F = 5.9  M = 3.4 | | χ2= 7.56 | | | | | p = 0.006 | | 1.7 | | | | |
| Subramaniam  2019 | 6126 | 18+ | GEN  (SGP) | stratified sampling of 3 main ethnic groups, 2016-2018 | CIDI 3.0 | trained prsnl, computer-assist interviews | 12-mo OCD | F = 4.1  M = 3.1 | | χ2 = 4.35 | | | | | p = 0.037 | | 1.8 | | | | |
|  |  |  |  |  |  |  | life OCD | F = 3.7  M = 2.1 | | χ2 = 14.13 | | | | | p = 0.0002 | | 1.3 | | | | |
| Subramaniam  2020 | 6126 | 18+ | GEN  (SGP) | stratified sampling of 3 main ethnic groups, 2016 | CIDI 3.0, YBOCS, SF-12 | trained prsnl,  interviews | 12-mo OCD | F = 3.7  M = 2.1 | | [95%CI,  1.1-2.9] | | | | | p = 0.016 | | 1.8 | | | | |
|  |  |  |  |  |  |  | life OCD | F = 4.1  M = 3.0 | | [95%CI,  1.0-2.2] | | | | | p = 0.084 | | 1.0 | | | | |
|  |  |  |  |  |  |  | 12-mo subOCs | F = 4.0  M = 5.0 | | [95%CI,  0.56-1.11] | | | | | p > 0.10 | | 0.8 | | | | |
| Carta  2020 | 2267  (1254) | 18+ | GEN  (ITA) | stratified random sampling 6 regions | SCID, MDQ, BDS, SF-12 | clinicians, interviews | life OCD | F = 2.0  M = 1.8 | | [95%CI, 0.6-2.2] | | | | | | | 1.2 | | | | |
| Lee  2021 | 12532 | 18-60 | GEN  (KOR) | nationwide community sample  KECA 2006-201 | K-CIDI,  DSM-IV | clinicians, interviews | obss | F = 2.93  M = 2.45 | | χ2 = 2.86 | | p = 0.09 | | | | | 1.18 | | | | |
|  |  |  |  |  |  |  | comps | F = 5.71  M = 4.18 | | χ2 = 12.92 | | p =0.0003 | | | | | 1.32 | | | | |
| Chen  2025 | 1572  (852) | 18+  81%≤30 | online  (CHN) | online snowball sampling | chinese OCI-R | SR | current likely OCD | F = 27.3  M = 34.0 | | [95%CI, 0.59–0.91] χ² = 8.23  p = 0.004 | | | | | | | 0.73 | | | | |
| Marković 2025 [A3.1] | 963 (482) | 18–65 | GEN (SER) | market-research panel; weighted representative sample | OCI-4 | SR | screen-positive OCS | F = 25.2  M = 18.0 | | [95% CI, 1.13–2.08] | | | | | | | 1.53 | | | | |
| Liu  2025 | 28140 (15603) | 18+ | GEN (CHN) | multistage clustered-area probability sampling | CIDI 3.0/ DSM-IV | trained prsnl,  interviews | life OCD | F = 2.39  M = 2.47 | | [95% CI, 0.76–1.23], p > 0.05 | | | | | | | 0.97 | | | | |
|  |  |  |  |  |  |  | 12-mo subOCs | F = 1.61  M = 1.65 | | [95% CI, 0.72–1.31], p > 0.05 | | | | | | | 0.97 | | | | |
| *older adults* | | | | | | | | | | | | | | | | | | | | | |
| Klenfeldt  2014 | 900 | 70 M/F,  78-92F | nondement.  (SWE) | subsampling, Swedish Population Register | SCID | trained nurse,  interviews | 1-mo OCs, age 70 | F = 25.4  M = 24.6 | | χ2 = 0.05 | | | | | p= 0.83 | | 1.0 | | | | |
|  |  |  |  |  |  |  | 1-mo OCs, age 78-92 | F = 22 | | - | | | | | - | | - | | | | |
|  |  |  |  |  |  |  | 1-mo OCD, age 70 | F = 4.5  M = 1.3 | | χ2 = 4.24 | | | | | p=0.04 | | 3.5 | | | | |
|  |  |  |  |  |  |  | 1-mo OCD, age 78-92 | F = 2.3 | | - | | | | | - | | - | | | | |
| Canuto  2017 | 3142 | 65-84 | GEN  (EU, ISR) | random sampling | CIDI65+ | trained prsnl, interviews | 12-mo OCD | F = 0.8  M = 0.8 | | F(1,20)=0.0 | | | | | p = 0.91 | | 1.0 | | | | |
| *by age groups* | | | | | | | | | | | | | | | | | | | | | |
| Alvarenga 2015 [A3.1] | 9937 | 6 ‒ 12 | students  (BRA) | school-based sampling | FHS, DY-BOCS | parental reports | $\geq$1 current OCs age 6-12 | β_M_ = 0.08 [95%CI, 0.08-1.52] p = 0.001 | | | | | | | | | 0.89 | | | | |
|  |  |  |  |  |  |  | $\geq$1 current OCs, age 6 | F $\cong$ 3.1  M $\cong$ 5 | | NA | | | | | NA | | 0.62 | | | | |
|  |  |  |  |  |  |  | $\geq$1 current OCs, age 7 | F $\cong$ 5.9  M $\cong$ 7.9 | |  |  |  |  |  |  |  | 0.75 | | | | |
|  |  |  |  |  |  |  | $\geq$1 current OCs, age 8 | F $\cong$ 5  M $\cong$ 9.5 | |  |  |  |  |  |  |  | 0.53 | | | | |
|  |  |  |  |  |  |  | $\geq$1 current OCs, age 9 | F $\cong$ 7  M $\cong$ 9.2 | |  |  |  |  |  |  |  | 0.76 | | | | |
|  |  |  |  |  |  |  | $\geq$1 current OCs, age 10 | F $\cong$ 7.5  M $\cong$ 9.9 | |  |  |  |  |  |  |  | 0.76 | | | | |
|  |  |  |  |  |  |  | $\geq$1 current OCs, age 11 | F $\cong$ 5  M $\cong$ 10.5 | |  |  |  |  |  |  |  | 0.48 | | | | |
|  |  |  |  |  |  |  | $\geq$1 current OCs, age 12 | F $\cong$ 9  M $\cong$ 9 | |  |  |  |  |  |  |  | 1.0 | | | | |
| Cath  2017 | 15194 | 15-97 | twins & family  (NLD) | Netherlands Twin Register | Padua ABBR | SR | provisional OCD | F vs M: OR: 0.99, z = −2.14, p = 0.032  “*primarily driven by 65 and older group*” | | | | | | | | | | | | | |
| Help-seeking | | | | | | | | | | | | | | | | | | | | | |
| *general clinics* | | | | | | | | | | | | | | | | | | | | | |
| Ghuloum  2011 | 1660 | 18-65 | primary h/c, (QAT) | random sampling,  12 clinics | DSM-based, study-spec | SR, supervised by trained nurses | current OCD | F = 4.5  M = 2.3 | | χ2 = 5.57 | | | | | p = 0.018 | | 1.96 | | | | |
| Lihua  2014 | 1576 | 16+ | general hospitals  (CHN) | random sampling, stratified by hospital type | CIDI 3.0 | trained prsnl | 30-d OCD,  age ≥ 16 | F = 2.84 [95%CI, 1.70–3.98]  M = 1.04 [95%CI, 0.32–1.76] | | | | | | p < 0.05 | | | 2.73 | | | | |
|  |  |  |  |  |  |  | 30-d OCD,  age 16-35 | F = 3.52 [95%CI, 1.38–5.66]  M = 0 | | | | | | - | | | inf | | | | |
|  |  |  |  |  |  |  | 30-d OCD,  age 36-54 | F = 2.26 [95%CI, 0.48–4.04]  M = 1.16 [95%CI, 0.56–1.76] | | | | | | p > 0.05 | | | 2.17 | | | | |
|  |  |  |  |  |  |  | 30-d OCD,  age ≥ 56 | F = 3.08 [95%CI, 0.98–5.48]  M = 1.94 [95%CI, 0.26–3.62] | | | | | | p > 0.05 | | | 1.59 | | | | |
|  |  |  |  |  |  |  | 12-mo OCD,  age ≥ 16 | F = 3.09 [95%CI, 1.90–4.28]  M = 1.56 [95%CI, 0.68–2.44] | | | | | | p < 0.05 | | | 1.98 | | | | |
|  |  |  |  |  |  |  | 12-mo OCD,  age 16-35 | F = 4.93 [95%CI, 2.41–7.45]  M = 4.38 [95%CI, 1.85–6.91] | | | | | | p > 0.05 | | | 1.13 | | | | |
|  |  |  |  |  |  |  | 12-mo OCD,  age 36-54 | F = 2.64 [95%CI, 0.71–4.57]  M = 1.55 [95%CI, 0.79–2.31] | | | | | | p > 0.05 | | | 1.46 | | | | |
|  |  |  |  |  |  |  | 12-mo OCD,  age ≥ 56 | F = 1.54 [95%CI, 0.04–3.04]  M = 1.94 [95%CI, 0.26–3.62] | | | | | | p > 0.05 | | | 0.79 | | | | |
|  |  |  |  |  |  |  | life OCD,  age ≥ 16 | F = 5.44 [95%CI, 3.88–7.00]  M = 2.61 [95%CI, 1.48–3.74] | | | | | | p < 0.05 | | | 2.08 | | | | |
|  |  |  |  |  |  |  | life OCD,  age 16-35 | F = 6.69 [95%CI, 3.78–9.60]  M = 5.58 [95%CI, 2.74–8.42] | | | | | | p > 0.05 | | | 1.20 | | | | |
|  |  |  |  |  |  |  | life OCD,  age 36-54 | F = 6.42 [95%CI, 3.47–8.37]  M = 1.55 [95%CI, 0.79–2.31] | | | | | | p < 0.01 | | | 4.14 | | | | |
|  |  |  |  |  |  |  | life OCD,  age ≥ 56 | F = 3.08 [95%CI, 0.98–5.48]  M = 2.33 [95%CI, 0.49–4.71] | | | | | | p > 0.05 | | | 1.32 | | | | |
| Bener  2015 | 1475 | 16-65 | primary h/c, (QAT) | random sampling,  13 clinics | WHO‑CIDI | psych nurses, interviews | current OCD | F = 13.5  M = 7.8 | | χ2 = 12.24 | | | | | p < 0.001 | | 1.86 | | | | |
| Sundquist  2017 | 5397675 | 18+ | primary h/c, (SWE) | Primary Care, Specialist Outpatient Care, Hospital Discharge records | registers  medical codes | medical records | current OCD | NA | | [95%CI, 1.38-1.46], p < 0.01 | | | | | | | 1.42 | | | | |
| Snorrason  2019 | 498  (275) | 18-74 | behavioral health  (USA) | consecutive patients at Behavioral Health Partial Program, 7/2016 - 7/2017 | MINI | clinicians, interviews | current OCD | F = 18.0  M = 20.0 | | χ2 =0.32 | | | P = 0.57 | | | | 0.9 | | | | |
| Jabeen  2020 | 59220 | 18+ | primary h/c, (PAK) | 11 public & 5 private hospitals | hospital Dx code | medical records | current OCD | F = 4.1  M = 4.2 | | χ2 = 0.37 | | | | | p = 0.61 | | 0.98 | | | | |
| Wazir  2023 | 231  (108) | 18-89 | psychiatric h/c, (PAK) | Community based | medical records | clinicians, interviews | current OCD | | F = 0.045  M = 0.057 | | [95%CI, 0.30–2.01], χ² = 0.18 p = 0.67 | | | | | | | | 0.76 | | |
| Alsaweer  2024 | 614  (506) | 18–65 | primary h/c, (BHR) | snowball sampling | MOCI  YBOCS | SR | current OCs | | “no significant sex difference (p = 0.260–0.997) | | | | | | | | | | | | |
| *psychiatric emergency department* | | | | | | | | | | | | | | | | | | | | | |
| Afifi  2026 | 27292  (13965?) | 18+ | psychiatric ED, (SAU) | retrospective chart review, one clinic, Jan–Dec 2024 | medical records | clinicians, interviews | ED presentation with OCD | | Jan-Mar:  F = 0.60%  M = 0.56% | [95%CI, 0.58–1.95], χ² = 0.04 p = 0.844 | | | | | | | | 1.06 | | | |
|  |  |  |  |  |  |  |  |  | Apr-Jun:  F = 0.87%  M = 0.44% | [95%CI, 0.94–4.12], χ² = 3.36 p = 0.067 | | | | | | | | 1.97 | | | |
|  |  |  |  |  |  |  |  |  | Jul-Sept:  F = 2.29%  M = 0.90% | [95%CI, 1.33–4.95], χ² = 8.45 p = 0.004 | | | | | | | | 2.56 | | | |
|  |  |  |  |  |  |  |  |  | Oct-Dec:  F = 0.39%  M = 0.42% | [95%CI, 0.53–1.62], χ² = 0.06 p = 0.803 | | | | | | | | 0.93 | | | |
| *OCD specialty clinics* | | | | | | | | | | | | | | | | | | | | | |
| *all ages* | | | | | | | | | | | | | | | | | | | | | |
| Algin  2018 [A3.1] | 400 | 8-63 | OCD  (IND) | consecutive patients at OCD clinic over 2 yrs | SCID-I, YBOCS | clinicians, interviews | 12-mo OCD | F = 41.2% | | z = 5.200 | | | | | p < 0.001 | | 0.7 | | | | |
| *children/adolescents* | | | | | | | | | | | | | | | | | | | | | |
| Deepthi  2018 | 451 | 17 | OCD  (IND) | retrospective chart review, one clinic, 1998-2014 | unstructured  interviews,  YBOCS | clinicians, interviews | life OCD | F = 27.9% | | Z = 4.672 | | | | | p < 0.001 | | 0.4 | | | | |
| Sharma  2019 | 173 | 5-17 | OCD  (IND) | consecutive patients,  6 clinics | YBOCS | clinicians, interviews | current OCD | NA | | z = 4.025 | | | | | p < 0.001 | | 0.5 | | | | |
| *adults* | | | | | | | | | | | | | | | | | | | | | |
| Cherian  2014 [A3.1] | 545 | 18+ | OCD  (IND) | consecutive patients at OCD clinic over 5 yrs | MINI, YBOCS | clinicians, interviews | 12-mo OCD | F = 39.1% | | z = 5.10 | | | | | p < 0.001 | | 0.6 | | | | |
| Medeiros  2017 | 236 | 18-72 | OCD  (USA, | OCD clinics, MA RI | SCID-I, YBOCS, DY-BOCS | clinicians, interviews | life OCD | F = 57.6 | | z = 2.34 | | | | | p = 0.02 | | 1.4 | | | | |
|  | 951 | 18-72 | BRA) | 8 clinics from 5 states |  |  |  | F = 58.3 | | z = 5.12 | | | | | p < 0.001 | | 1.3 | | | | |
| Dell’Osso 2017 | 416 | 18+ | OCD  (world) | consecutive outpatients,  multiple clinics | SCID I&II,  YBOCS | clinicians, interviews | current OCD | F = 57.5 | | z = 2.86 | | | | | p = 0.004 | | 1.4 | | | | |
|  |  | 18-42 |  |  |  |  |  | F = 52.1 | | z = 0.58 | | | | | p = 0.56 | | 1.1 | | | | |
|  |  | 42+ |  |  |  |  |  | F = 63.1 | | z = 3.70 | | | | | p = 0.0002 | | 1.7 | | | | |
| Tripathi  2018 | 945 | 18-72 | OCD  (IND) | 16 medical centers over 1 yr | SCID-5-RV, YBOCS | clinicians, interviews | 12-mo OCD | F = 40.3 | | z = 5.20 | | | | | p < 0.001 | | 0.7 | | | | |
| Brueggerhoff-Batel 2025 [A3.1] | 45 | 18-70 | OCD  (MEX) | consecutive patients at OCD clinic, Dec 2024–Apr 2025 | YBOCS | clinicians, interviews | current OCD | F = 48.9 | | Z = -0.149 | | | | | p = 0.881 | | 1.0 | | | | |
| Incidence and Prevalence in sub-populations | | | | | | | | | | | | | | | | | | | | | |
| Sulkowski  2011 | 358 | 17-27 | psych/edu STU (USA) | anonymous survey during class | OCI-R | SR | clinic OCs (OCI-R>27) | F = 5.5  M = 5.9 | | χ2 = 0.048 | | | | | p = 0.83 | | 0.9 | | | | |
| Strehlau  2012 | 193F | 19-57 | homeless (CAN) | homeless outreach, drop-ins, and shelters | DSM-IV  interviews | clinicians, interviews | current OCD | F = 19 | | - | | | | | - | | - | | | | |
|  |  |  |  |  |  |  | life OCD | F = 62 | |  |  |  |  |  |  |  |  |  |  |  |  |
| Desouky  2015 | 1024F | 15-17 | students  (SAU) | multistage cluster sampling | OCDS | SR | clinic OCs | F = 23.1 | | - | | | | | - | | - | | | | |
| Jaisoorya 2017 | 4926 | 18 -25 | college  (IND) | college based sampling | CIS-R | SR | current OCD | F = 3.2  M = 3.5 | | χ2 = 0.20 | | | | | p = 0.56 | | 0.9 | | | | |
|  |  |  |  |  |  |  | current subOCD | F = 7.7  M = 9.9 | | χ2 = 6.7539 | | | | | p = 0.02 | | 0.8 | | | | |
| Ashraf  2017 [A3.2] | 377  (196) | 18-46 | college  (PAK) | college based sampling,  two universities | Padua | SR | current  subOCs | F = 43.4  M = 50.3 | | χ2 = 1.80 | | | | | p > 0.10 | | 0.8 | | | | |
|  |  |  |  |  |  |  | current  contam | F = 48.5  M =38.7 | | χ2 = 3.67 | | | | | p < 0.05 | | 1.5 | | | | |
|  |  |  |  |  |  |  | current  harm | F = 50.5  M =39.8 | | χ2 = 4.37 | | | | | p < 0.05 | | 1.6 | | | | |
|  |  |  |  |  |  |  | current  impulses | F = 30.1  M = 63.0 | | χ2 = 40.97 | | | | | p < 0.001 | | 0.3 | | | | |
| Asghar  2020 | 1172  (492) | 18+ | med students  (PAK) | convenience sampling, med schools | YBOCS | SR | current OCD | F = 7.7  M =11.9 | | z = 1.7 | | | | | P = 0.086 | | 0.7 | | | | |
| Batchelder 2021 | 290M | 18–65 | Gay and bisexual males, history of sex abuse  (USA) | HIV prevention and mental health programs | MINI CTQ-SF | clinicians, interviews | current OCD | | Bi-s =7.7  Gay = 11.9 | | χ2 = 0.49 | | | | | p < 0.05 | | | G:Bi  1.46 | | |
| Russell  2022 | 1.36 M | 20-40 | active military  (USA) | Defense Medical Epidemiological Database (1999–2018) | ICD-10 codes | health records | incidence (per 1,000) | | F = 0.52  M = 0.30 | | χ² = 10 166 | | | | | p < 0.001 | | | 1.73 | | |
| Faruki  2023 | 319430 (198400) | 6–24 | suicidal inpatient/mood d/o | stratified probability sampling | ICD-10 codes | health records | current OCD | | GD= 6.2  noGD = 2.9 | | [95% CI, 1.51–1.94], p<0.001 | | | | | | | | GD:  1.71 | | |
| Li  2023a | 1248  (640) | 18 + | students  (CHN) | college based sampling,  three universities | SCL-90 | SR | current OCD | | F: M OR = 0.75 | | [95%CI, 0.57–0.98], p < 0.05 | | | | | | | | 0.75 | | |
| Pei  2023 | 7493  (3791) | 12–18 | students  (CHN) | two-step school-based sampling | MINI-KID + DSM-IV | clinicians, interviews | current OCD | | F = 10  M = 8.1 | | [95%CI, 1.08–1.47], p = 0.005 | | | | | | | | 1.26 | | |
| Almadani  2024 | 371  (137) | 18+ | med students  (SAU) | school-based sampling | OCI-R | SR | current likely OCD | | F = 43.8  M = 23.5 | | χ² = 16.2, p < 0.001 | | | | | | | | 2.5 | | |
| Barzallo Farfán  2024 | 270 (145) | 18-30 | medical students, (PER) | convenience sampling, med schools | YBOCS, KNOW-P-COVID-19 | SR | current OCs | | F = 15.9  M = 10.4 | | χ2 = 1.73 | | | | | p = 0.19 | | | 1.61 | | |
| Farahbakhsh  2024 | 1282 (693) | 20–60 | empl. med school, (IRN) | convenience sampling, med school | CIDI | trained prsnl | 12m OCD | | F = 4.2  M = 3.6 | | χ² = 0.32 | | | | | p=0.568 | | | 1.05 | | |
| Nasirpour  2024 | 4453  (2813) | 18–65 | public employ.  (IRN) | Employees’ Health Cohort Study | CIDI-2.1,  ERI | Clinicians,  interviews | Current OCD | | F = 2.9  M = 2.2 | | χ² = 2.09 | | | | | p = 0.15 | | | 1.34 | | |
| Asma  2025 | 640  (391) | 17-22 | med students  (IND) | convenience sampling, med school | OCI-R | SR | current likely OCD | | F = 35.0  M = 19.7 | | [95%CI, 1.51–3.20], χ² = 17.41 p < 0.001 | | | | | | | | 2.2 | | |
| Assil  2025 | 190  (106) | 18 + | students  (SAU) | school-based sampling | Arabic OCI-R | SR | current likely OCD | | F= 45.3  M = 36.9 | | [95%CI, 0.79–2.54], p = 0.3 | | | | | | | | 1.41 | | |
| Faghihzadeh  2025 | 131  (87) | 24 ± 4 | med students  (IRN) | convenience sampling, med school | SCL-90-R | SR | current OCs | | F = 20.7  M = 31.8 | | [95% CI, 0.25–1.27], χ² = 1.96  p = 0.16 | | | | | | | | 0.56 | | |
| Madoro  2025 | 370  (203) | 19–35 | med/health students  (ETH) | systematic random sampling, med school | OCI-R | SR | current OCs | | F = 52.7  M = 41.9 | | Adjusted [95% CI, 1.09–2.18], χ² = 4.28 p < 0.05. | | | | | | | | 1.33 | | |
| Picard  2025 | 10F | 18-25 | Ballet  (CAN/USA) | qualitative structured interviews | thematic analysis | trained prsnl | OCs themes | | 40% reported OCs | | | | | | | | | | | | |
| Shaheen  2026 | 1850  (980) |  | med  students (EGY) |  | OCI-R | SR | current OCs | | OR = 1.35 [95% CI, 1.13–1.63], p = .001; adj. OR = 1.25 [95% CI, 1.03–1.51], p = .023. | | | | | | | | | | | 1.35 | |

Table A3.2. Gender differences in onset, progression, and contributing factors.

| **First Auth, Year** | **N**  **(F)** | | **Age mean**±**SD (range)** | | | **Pop** | | | **Meas.** | | | **Gender effects (F/M)** | | | | |
| --- | --- | --- | --- | --- | --- | --- | --- | --- | --- | --- | --- | --- | --- | --- | --- | --- |
|  |  |  |  |  |  |  |  |  |  |  |  | **Stats** | | | **Direction** | |
| Age of onset (AO) | | | | | | | | | | | | | | | | |
| Memis  2019 | | 74  (47) | | 15 ± 2  (<18)  31 ± 7  (18+) | | | OCD  (TUR) | | | Pts repts. | | | Retrospective. M:F: EO = 1.07, LO = 0.36, LO:EO OR = 0.34, χ^2^ = 4.8 p = 0.02 | | | (AO≥18) : (EO<18) OR: M < F |
| *children/adolescents* | | | | | | | | | | | | | | | | |
| Nakatani  2011 | 365  (151) | | 14 ± 3  (6-18) | | | OCD  (GBR) | | | parent/ pts repts | | | distress Sx, 2 subgr: median split with robustness check. %F: EO (AO ≤ 9) = 0.41 z-test p = 0.03, LO (13 < AO ≤ 17) = 0.42 z-test p = 0.02. | | | AO ≤ 17: M > F | |
| *adults* |  | |  | | |  | | |  | | |  | | |  | |
| Wang  2011 | 602  (263) | | 30 ± 11  (8-65) | | | OCD  (CHN) | | | 2 clinic interviews | | | distress Sx, 2 subgr: median split. %F: EO (AO ≤ 18) = 0.33, LO (AO > 18) = 0.53, t(2) =6.4 p = 0.042. | | | AO ≤ 18 M > F  AO >18 M ~ F | |
| De Luca  2011 | 196  (110) | | 39 ± 12  (18+ ) | | | OCD  (CAN) | | | SCID-I/P | | | NA, 2 subgr: mixture analysis. %F: EO (AO ≤ 15) = 0.54, LO (AO > 15) = 0.60, p = 0.47. | | | AO ≤ 15: M ~ F  AO > 15: M < F | |
| Narayanaswamy  2012 | 545  (213) | | 29 ± 11  (18+ ) | | | OCD  (IND) | | | parent/ pts repts | | | distss Sx. F-M d = 0.62 U = 23453 p <0.001. | | | AO  M < F | |
| Kichuk  2013 | 955  (555) | | 36 ± 13  (18 – 65) | | | OCD  (BRA) | | | DY-BOCS  pts repts | | | distress Sx, by primary Sx. AO ~ Primary Sx x F: F(3,511) = 2.6 p = 0.051; Forbid thght M < F, contam/hoard M ~ F, Sym M > F. | | | forbid thght M < F, contam/hoard M ~ F, symm M > F | |
| Anholt  2014 | 377  (215) | | 36 ± 11  (18+ ) | | | OCD  (NOR) | | | SCID-I | | | DSM, 2 subgr: mixture analysis. %F: EO (AO < 20) = 0.59, LO (AO ≥ 20) = 0.53, χ^2^ =1.2 p = 0.28. | | | AO >,< 20: M ~ F | |
| Frydman  2014 | 983  (562) | | med = 32  (25 – 44) | | | OCD  (BRA) | | | SCID-I | | | distress Sx 3 subgr: *a priory*. %F: EO (AO ≤ 16) = 0.48, IO (16 < AO < 40) = 0.58, LO (AO ≥ 40) = 0.73, χ^2^ (2) =16.2 p <0.001. Risk for LO. LO~F: β = 0.5 [95CI%, 0.9 – 2.7], controlling for OCs, PTSD after age 40/no history of PTSD, pregn. | | | AO ≥ 40: M < F | |
| Albert  2015 | 483  (232) | | 34 ± 12  (18+ ) | | | OCD  (ITA) | | | clinic interview/parent repts | | | distress Sx, 3 subgr: mixture analysis. %F: EO (AO ≤ 9) = 0.44, IO (10 ≤ AO ≤ 23) = 0.46, LO (AO ≥ 24) = 0.58, χ^2^ =4.3 p = 0.10. DSM/DSM, 2 subgr: mixture analysis. %F: EO (AO ≤ 26) = 0.44, LO (AO ≥ 26) = 0.61, χ^2^ =9.3 p = 0.002 | | | AO < 23-26: M > F  AO ≥ 23-26: M < F | |
| Sharma  2015 | 802  (305) | | 29 ± 10  (N/A) | | | OCD  (IND) | | | medical records | | | NA, 3 subgr: *a priory*. %F: EO (AO < 18) = 0.27, IO (18 ≤ AO ≤ 39) = 0.42, LO (AO ≥ 40) = 0.74, χ^2^ =42 p < 0.001. F OR: IO vs. LO = 0.27 [95%CI, 0.13 – 0.56], controls: LSE/ aggres/ hoard; EO vs. LO = 0.21 [95%CI, 0.09 – 0.49], controls: LSE/aggres/sex/repeat/ socil phob/ psychos; EO vs IO = 0.58 [95%, 0.41 – 0.82], controls: for sex/repeat/tic/ hoard/ YBOCS. | | | AO ≤ 39 M > F  AO ≥ 40 M < F | |
| Dell’Osso  2016 | 431  (249) | | 43 ± 13  (18+ ) | | | OCD  (world) | | | SCID I, II | | | distress Sx, 3 subgr: *a priory*. %F: EO (AO ≤ 12) = 0.52 p, IO (13 ≤ AO ≤ 17) = 0.50, LO (AO ≥ 18) = 0.67, Fisher’s Exact test p =0.004. | | | AO ≥ 18: M < F | |
| Kang  2017 | 615  (217) | | 30 ± 11  19-65 | | | OCD, HC  (KOR) | | | family/pts repts | | | Retrospective. M:F: EO = 2.94, LO = 1.11, LO:EO M:F = 0.38 z = 5.6 p < 0.001 | | | (AO≥18) : (EO<18) OR:  M < F | |
| Grover  2018^s^ | 345  (149) | | 30 ± 11  (18+ ) | | | OCD  (IND) | | | YBOCS CL | | | NA, 2 Sx subgr: *a priory*. %F: EO (AO ≤ 18) = 0.25, LO (AO >18) = 0.50, χ^2^ =16.2 p < 0.001. By primary Sx. M OR EO-OCD: hoard/sym = 3.81 [95%CI, 1.17–12.45], contam = 0.06 [95%CI, <0.01,0.48], aggress/sex/relig = 1.47 [95%CI, 0.56–3.84]; M OR LO-OCD: hoard/ sym = 5.29 [95%CI, 2.78–10.09], contam = 0.24 [95%CI, 0.19–0.46], aggress/sex/relig = 2.48 [95%CI, 1.49–4.12]. | | | AO ≤ 18: M > F AO >18: M ~ F | |
| Girone  2024 | 284 (137) | | 39 ± 14  (18–69) | | | OCD (ITA) | | | SCID-5 YBOCS | | | Retrospective. M:F EO = 1.86 (% F=0.35), LO = 0.74 (% F=0.58), LO:EO M:F = 0.40; χ²(1) = 14.1, p < 0.001. | | | EO: M > F  AO: M < F | |
| Stein  2025 [A3.1] | 26136 | | (18+) | | | GEN  (woldwide) | | | CIDI 3.0,  YBOCS | | | Retrospective. “There is no evidence of sex differences in age of onset.” Stats not reported. | | | M ~ F | |
| Progression from onset to Dx | | | | | | | | | | | | | | | | |
| Coles  2011 | 199  (140) | | 34 ± 11  (18-64) | | | OCD  (USA) | | | Study-specific | | | Retrospective. Sx phase: F = 5.83±6.37, M = 3.88±5.09, t(182) = 1.97 p =0.05. ***F x onset – NA.*** | | | Sx phase  M ≤ F | |
| Torresan  2013 [A3.2] | 858 (504) | | 35 ± 12 (18-77) | | | OCD  (BRA) | | | YBOCS, DY-BOCS | | | Cross-sectional. Worse ever Sx, age: M = 27.7 ± 11.6 F = 30.5 ± 12.4 p < 0.001, Help-seeking, age: M = 27.9 ± 11.6 F = 31.2 ± 12.5 p < 0.001. | | | Onset worse Sx/help  M < F | |
| Thompson  2020 | 954  (545) | | 35 ± 13  (18+ ) | | | OCD  (BRA) | | | YONHQ,  YBOCS,  DY-BOCS | | | Retrospective. Sx phase, median: M = 5 F = 8 z = 4.89 p < 0.001; Poisson regr: M β = 0.16 [95%CI, 0.11-0.21] Wald = 45.9 p < 0.001, controlling for age, symptom type, symptom severity, agoraphobia, GAD, Tic, Family history. | | | Sx phase  M < F | |
| Ziegler  2021 | 100  (57) | | 40 ± 14  (18-82) | | | OCD  (DEU) | | | OCI-R, GAF, USS | | | Retrospective. Onset → Dx: short (< 7) F=21, M=11; long (≥7): F=33, M=29. F:M OR (long vs short) = 0.60 [95% CI 0.25–1.44] | | | Onset → Dx: M ~ F | |
| Pellegrini  2026 | 220 (101) | | 35 ± 12  (18+ ) | | | OCD  (ITA) | | | SCID-5, YBOCS | | | Retrospective. Untreated illness duration (DUI), %males: DUI ≤ 42m = 50.9 DUI > 42m = 53.7 p = 0.69. | | | DUI: M ~ F | |
| Kong  2026 | 884  (547) | | 15± NA  (12–17) | | | psych inpat (CHN) | | | med records | | | Retrospective longitudinal EHR (2+ admis & 3+ years f/u): Baseline OCD-spect: M = 10; F = 17. F/u OCD-spect: M = 40% (4/10); F = 70.6% (12/17); F:M OR = 3.42 [95% CI, 0.54–25.29] p = 0.22. F/u SZ-spect: M = 50% (5/10); F = 11.8% (2/17); F:M OR = 0.15 [exact 95% CI, 0.01–1.23] p = 0.065 | | | OCs → SZ-spect:  M ≥ F | |
| Puberty | | | | | | | | | | | | | | | | |
| Voltas  2014 | w1 1514  (794)  w2 562  (308)/  w3 242  (147) | | w1 10 ± 1  (8-12)/  w3 14 ±1  (11-15) | | | GEN,  (ESP) | | | LOI-CV,  YI-4, CDI,  SCARE | | | Longitudinal. Wave 1: order/check/pollution: F=7.33 ± 5.4 M = 6.76 ± 5.2 t = -2.08 p = 0.037; superstition/ mental comps: F = 2.31 ± 3.0 M = 2.65 ± 3.4 t = 1.99 p = 0.046. Progression. ***“all symptoms decreased over time with no differences between sexes” – stats NA.*** | | | subOCs ↓  from 10 to 14  M ~ F | |
| Castagnini  2016 | 6043  (2496) | | baseline  12 ± 3  (6-16) | | | Psych concerns  (DNK) | | | Dx codes | | | Medical records. OCD incidence in kids: IRR M/F = 1.1 [95%CI, 0.8-1.5], OCD in kids → need for treatment in adult: OCD RR = 5.4 [95%CI, 4.0-7.1] M/F RR= 0.7[95%CI, 0.4-1.2]; Dx kids →Dx in adults: OCD→OCD OR = 43.3 [95%CI, 19.6-95.6]; but note Phob/panic/anx → OCD OR = 6.6 [95%CI, 2.0 – 21.9] & Phob/panic/anx incidence in kids RR = 3.3 [95%CI 2.5-4.3] M/F RR = 0.5 [95%CI, 0.3-0.8]. | | | OCD kids → treatm in adults M ~ F Phob/panic/anx in kids M < F & →OCD in adults | |
| Barzilay  2019a | 6598  (3567) | | 16 ± 3  (11-21) | | | GEN  (USA) | | | GOASSESS, K-SADS,  C-GAS | | | Longitudinal. SubOCs, %F: At least 1 OCs = 0.59 no OCs = 0.51, F OR = 1.3 [95%CI, 1.2 – 1.5] Wald(1) = 25.97 p < 0.001; puberty (Pb) F x Pb M < F Wald(1) = 4.3 p = 0.038.  SubOCs subtypes, Bad thoughts: F OR=1.16 [95%CI, 1.03-1.31] W(1)=5.6 p=0.02, Pb OR = 1.14 [95%CI, 1.01-1.28] W(1)=4.5 p = 0.03, F x Pb W(1) = 0.09 p = 0.77; Repeat/Check: F OR = 1.27 [95%CI, 1.13-1.43] W(1) = 16.14 p <0.001, Pb OR = 1.2 [95%CI, 1.07-1.34] W(1) = 9.15 p = 0.002, F x Pb W(1) = 0.73 p = 0.39; Sym: F OR = 1.42 [95%CI, 1.24-1.63] W(1) = 25.46 p <0.001, Pb OR = 1.02 [95%CI, 0.9-1.17] W(1) = 0.12 p = 0.73, F x Pb W(1) = 1.49 p = 0.22; Clean/Contam: F OR = 1.41 [95%CI, 1.19-1.67] W(1) = 15.31 p < 0.001, Pb OR = 1.13 [95%CI, 0.96-1.33] W(1) = 2.0 p = 0.16, F x Pb W(1) = 0.40 p = 0.53; Hoard: F OR = 1.31 [95%CI, 1.06-1.62] W(1) = 6.17 p = 0.01, Pb OR = 1.18 [95%CI, 0.95-1.45] W(1) = 2.31 p = 013, F x Pb M ≤ F W(1) = 3.21p = 0.07. SubOCs in kids → lifetime Dx, OR: OCD = 11.7, Depress = 4.3, psychos = 3.3, Suicide Id = 4.4 ps < 0.001; ***OR x F – NA.*** | | | subOCs  F x puberty  M < F,  subOCs types  F x puberty  M ~ F | |
| de Barros  2021 | 1877  (822) | | 13 ± 2  (9–18) | | | GEN  (BRA) | | | DAWBA,  CBCL | | | Longitudinal, 3 years. Latent factor-based OCs ~ F: β=0.007 [95%CI, -0.081, 0.096]. | | | ΔOCs in teens  M ~ F | |
| Biosocial contributing factors and early markers  *prenatal and perinatal stresses* | | | | | | | | | | | | | | | | |
| Cheng  2014 | 566044  (292207) | | | | N/A  (14-49) | | | GEN  (TWN) | | | National database | | | Longitudinal. Season of birth (SOB) ~ OCD: Mar – Jul OR = 0.85[95%CI, 0.74-0.96]; Aug – Nov OR = 1.19 [95%CI, 1.05 – 1.36]; only in M: SOB ~ OCD x M p = 0.03 | | SOB → OCD  M > F |
| Fevang  2016 | 216  (111), 1767  (944) | | | | newborn/  2/5/11 yo | | | EP/ ELBW,  controls  (NOR) | | | Medical records, SCARED,  SOCD | | | Longitudinal. OCD severity ≥ 95% based on the control group EP/ ELBW OR: M = 2.5 [95%CI, 1.8 – 5.2], F = 2.7 [95%CI, 1.4 – 5.4] | | premature / low weight newborns  M ~ F |
| Larsen  2021 | 1167043  (568313) | | 6 - 28 | | | GEN  (DEN) | | | Medical records  ICD-8/10 | | | Longitudinal national cohort (1989–2010 births, follow-up to 2016), Risk factors (OCD HR): Premature birth (<33w): F = 1.46 [95%CI, 1.15–1.83] M = 1.12 [95%CI, 0.85–1.48]; Low birth weight (<1500 g): F = 1.32 [95%CI, 1.01–1.74] M = 1.08 [95%CI, 0.80–1.46]; Multiple birth F = 1.51 [95%CI, 1.29–1.77] M = 1.26 [95%CI, 1.04–1.52]; Congenital malformations F = 1.72 [95%CI, 1.10–2.69] M = 1.40 [95%CI, 0.90–2.18]; Ante genitourinary infection: F = 1.39 [95%CI, 1.16–1.67] M = 1.20 [95%CI, 1.01–1.43]; maternal age >35 y: F = 1.24 [95%CI, 1.09–1.41] M = 1.10 [95%CI, 0.95–1.26]; paternal age > 40: F = 1.22 [95%CI, 1.08–1.39] M = 1.13 [95%CI, 0.97–1.31]. | | | M ≤ F: Ante/PP complic. ↑↑ OCD | |
| *resilience* |  | |  | | |  | | |  | | |  | | |  | |
| Hjemdal  2011 | 307  (140) | | 16 ± 1  (14–18) | | | school  (NOR) | | | OCI-R,  READ | | | Cross-sectional. Resilience ~ F: Pers. comp. = -0.19 p < 0.001, Soc. Comp. = 0.05 p > 0.05, Struc style = 0.06 p > 0.05, Soc. res. = 0.19 p < 0.001, Fam. coh. = 0.15 p < 0.01; Resilience ~ OCI-R: Pers. comp. = -0.22 p < 0.015, Soc. Comp. = -0.17 p < 0.015, Struc style = -0.08 p > 0.10, Soc. res. = -0.24 p < 0.015, Fam. = -0.21 p < 0.015; OCI-R ~ F = 0.15 p < 0.01. | | | pers comp M > F  soc comp/ struc style: M ~ F  soc res/fam M < F | |
| *family* | | | | | | | | | | | | | | | | |
| Taberner  2012 | 184F/  180mo/  174fa | | 22 ± 3  (18-35)/  50 ± 7  (39-74) | | | student/  parents  (ESP) | | | Padua,  DS | | | Cross-sectional. Disgust/OCs in parents ~ Contam daughter: hygiene fathers β = 0.23 p < 0.01; ordering mothers β = 0.21 p < 0.01. | | | father hygiene → contam daughters  mother ordering → contam daughters | |
| Hacıömeroğlu 2014 | 300  (147) | | 20 ± 2  (18+ ) | | | students  (TUR) | | | PADUA,  RAS,  s-EMBU | | | Cross-sectional. [Perceived mother overprotection (mOP) → OCs] is mediated by Resp: mOP →OCs β = 0.23 p < 0.01, mOP →Resp β = 0.35 p < 0.001, Resp →OCs β = 0.18 p < 0.01. | | | overprotect mother → OCs | |
| Liu  2017 | 2008  (880) | | 20 ± 1  (18+ ) | | | students  (CHN) | | | SCL-90, STAXI-2,  FACES-II | | | Cross-sectional. Family dynamic (FD), M < F: fam cohes (F(1903)=35.76 p <0.001), fam adapt (F(1928)=11.84 p=0.001), anger/trait (F(1984)=6.59 p = 0.01), anger-in (F(1939) = 5.89 p = 0.015); M > F: anger-out (F(1961) = 4.62 p = 0.032.  FD ~ OCs: fam cohes M = -0.16 F = -0.25, z = 2.09 p = 0.037, fam adap M = -0.19 F = -0.28, z = 2.12 p = 0.034.  [Anger →OCs] mediated by FD: Anger x fam cohes M = -0.36 t = -2.54 p <0.01; Anger-in x fam adapt M = -0.14 t = -1.10 p > 0.05, F = -0.30 t = -2.24 p < 0.05; Anger-in x fam cohes M = 0.27 t = 1.82 p > 0.05, F = 0.31 t = 1.91p > 0.05. | | | fam cohes/adapt  M < F,  fam cohes/adapt ↑↓ OCs M < F,  [anger-in →OCs] ↑↓ fam adap M < F,  [anger →OCs] ↑↓ fam cohes M > F | |
| Krebs  2018 | 2616→  1579 | | t1=15 → t2 =17 | | | Genesis 1219  (GBR) | | | *SCAS,* negative sanctions | | | Longitudinal. OCs (t2) ~ parental punitive discipline (t1), while controlling for OCs at t1 and stressful event at t2: maternal β = 0.02 p = 0.49, paternal β = 0.02 p = 0.59 | | | maternal/paternal punitive discipline ≠> OCS | |
| Someshwar  2020 | 509  (220) | | 39 ± 13 (18+ ) | | | fam. risk (IND) | | | MINI,  ACE-IQ | | | Retrospective. Earlier psychiatric onset ~ ACE severity × sex: M HR=1.13, 95% CI [1.07, 1.20], p<.001; F HR=0.94 [0.86, 1.03], p=.19. | | | familial risk + ACE → EO M > F | |
| Mora-Jensen 2025 | 155  (79) | | 13 ± 3 (8–17) | | | OCD, HC (DNK) | | | CYBOCS,  salivary OT, TEC-M | | | Cross-sectional. Child OCD × supportive interaction ~ parental OT: father β = 0.11 p = 0.008 p_FDR_=0.18; mother β = -0.06 p = 0.23 p_FDR_ = 0.77 | | | Parental OT↑↑support child x OCD: M > F | |
| *stressful life events (SLE): trauma, illness, etc.* | | | | | | | | | | | | | | | | |
| Real  2011 | 412  (174) | | 34 ± 12  (18+ ) | | | OCD  (ESP) | | | YBOCS,  SCID-II,  interview | | | Retrospective. SLE-preceding OCD: F OR = 1.5 [95%CI, 1.01-2.3]; F OR = 1.2 [95%CI, 0.74 – 2.07], while controlling for age, YBOCS, complicated birth, age of onset, contam, fam history. | | | SLE-preceding OCD  M ≤ F | |
| Rosso  2012 | 329  (160) | | 35 ± 12  (18+ ) | | | OCD  (ITA) | | | YBOCS  SCID-I, II | | | Retrospective. SLE-preceding OCD: F OR = 2.2 [95%CI, 1.4–3.4]. | | | SLE-preceding OCD  M < F | |
| Peles  2014 | 116F | | 38 ± 10  (NA) | | | sex abused  (ISR) | | | LEIQ,  YBOCS | | | Cross-sectional. Prevalence of OCD in sex abuse victims: frm opioid use = 0.66, no opioid use =0.30, OR = 4.47 [95%CI, 2.0–10.0]. | | | F:  sex abuse + opioid use ↑ OCD | |
| Ishikawa  2015 | 148F | | 19 ± 2  (18–28) | | | students  (JPN) | | | MCR,  unwanted sexual exp. recall task | | | Symptom provocation. Mental Contam (dirtiness, wash urge, internal/external neg feel) ↑ over Time, T, in response to Event, E, (verbal/visual assault, forcible touching, (attempted) rape). T: F(4,141) = 22.08 p < 0.001, E: F(12,429) = 1.96 p < 0.05, E x T: F(12,429) = 3.03 p < 0.001; dirtiness↑ T: F(1,144) = 50.13 p < 0.001 η^2^ = 0.26, E: F(12,429) = 1.96 p < 0.05 η^2^ = 0.07, E x T: F(12,429) = 3.03 p < 0.001 η^2^ =0.10; urge to wash↑ E x T: F(3,144) = 2.79 p < 0.03 η^2^ = 0.07, Δ urge to wash↑~ (attempted) rape p = 0.02; internal neg feel↑ T: F(1,144) = 60.42 p < 0.001 η^2^ = 0.30; external neg feel↑ T: F(1,144) = 49.12 p < 0.001 η^2^ = 0.26. | | | F:  unwanted sexual exp recall ~ contam ↑ | |
| Vidal-Ribas  2015 | 22084  (12524) | | 34 ± 8  (28-54) | | | twin registry  (SWE) | | | LSC-R, study-spec OCs | | | Cross-sectional. Prevalence M-F, in monozygotic twins, ps<0.00001: Abuse/ family disruption d = -0.14, Loss d = -0.12, Sex abuse d = -0.56, Non-sexual assault d = 0.40. | | | assault  sexual M < F  non-sexual M > F | |
| Waller  2014 | 80F | | 24 ± 10  (15-60) | | | students  (AUS) | | | non-cons. “dirty kiss” task, OCI-R; DASS, MCR | | | Symptom provocation. "dirty-kiss" perpetrator role → feeling of dirtiness, urge to wash, negative emotions: ƞ>0.47, p <0.001  Symptom mitigation. physical washing < mental washing: d >= 1.5 p <0.001. | | | F: non-consensual kiss →mental contam  physical wash < mental wash | |
| Ashraf  2017 [A3.1] | 377 (196) | | 27 ± 6 (18-46) | | | GEN  PAK | | | Padua, PSS | | | Cross-sectional. In high stress F:M OR = 0.33 [95%CI, 0.18-0.63]. High stress in OCD F:M OR = 0.39 [95%CI, 0.21-0.75]. | | | OCs↑↑stress: M > F | |
| Taghva  2017 | 215M | | 52 ± 8 (29–92) | | | veterans/ancle inj  (IRN) | | | SCL-90-R | | | Cross-sectional. <rate> OCs ~ Disability rate: “< 30%” = 1.4± 0.7, “≥30%” = 1.1±0.7 p =0.027; OCs ~ limb disability: “No” = 1.0 ± 0.7, “Yes” = 1.2 ± 0.6 p= 0.005; OCs ~ hospitalization due to injury: “No” = 1.3 ± 0.7, “Yes” = 1.5 ± 0.7 p= 0.05; OCs ~ hospitalization due other reasons: “No” = 1.3 ± 0.7, “Yes” = 1.6 ± 0.6 p= 0.044. | | | M:  foot injury ↑↑ OCS  disabil rate ↑↓ OCS | |
| Kennedy  2017 | 60M | | med=20  18 - 40 | | | students  (AUS) | | | non-cons. “dirty kiss” task, study-spec | | | Symptom provocation. Non-consensual > consensual: mental contam ↑ d > 0.53, p <0.001; choose a cleansing product as a gift↑ OR = 4.5 p = 0.024 | | | M: non-consensual kiss →mental cont.→ cleansing | |
| Barzilay  2019b | 7054  (3809) | | 16 ± 3  (11-21) | | | GEN  (USA) | | | DSM, interview | | | Retrospective. OCs ~ Trauma exposure x F: Wald (1) = 7.93 p = 0.005 F > M; OCs ~ non-assault trauma exposure x F: Wald(1) = 5.20 p = 0.023 F > M; OCs ~ assault trauma exposure x F: p > 0.05 F ~ M | | | trauma exposure  non-assault M < F  assault M ~ F | |
| An  2019 | 3160F | | 18-74 | | | GEN  (KOR) | | | K-CIDI | | | Lifetime prevalence OCD IPV OR. Physical IPV = 8 [95%CI, 1.6-26.3], Sexual IPV = 27.8 [95%CI, 5.0-107.3, any IPV = 10.6 [2.7-31.3]- the highest among psychiatric conditions. | | | F: IPV ↑↑ OCD | |
| Destrée  2020 [A3.2] | 954  (548) | | 34 ± 13  (18+ ) | | | OCD  (BRA) | | | YBOCS, DY-BOCS  YONHQ | | | Retrospective. SLE prevalence: Death SLE F-M d = 0.054 p = 0.094, R/ship SLE d = 0.001 p =0.99, illness SLE d = 0.02 p = 0.53; speed subclinic →clinic OCD: Death SLE x M β = -0.31 p < 0.001; speed subclinic →clinic OCD, by Sx dimensions: Sex/Relig ~ M β = 0.90 p =0.03, Sex/Relig ~ R/ship SLE x M β = -0.49 p = 0.069. | | | OCD ~ Death SLE  M > F | |
| Krause  2021 | 149F | | 23±5 (19–46) | | | students (CAN) | | | sexual-harassment imagery task | | | Symptom provocation. Sexual-harassment role → mental contam η² ≈0 .40–0.47 p < 0.001, disgust η² ≈ 0.30–0.40 p < 0.001, distress η² ≈0.20–0.30; p < .001, Washing urge η² ≈0 .18–0.25; p <0 .001, avoidance urge η² ≈ .20; p < 0.001, shame/guilt/fear η² ≈0 .25–0.35 p < 0.001. | | | F: harassment imagery → mental contamination | |
| Samuels  2021 | 2117  (1137) | | 46 ± 16  (18–89) | | | GEN  (USA) | | | DY-BOCS,  OCI-R,  spec phob | | | Cross-sectional + retrospective. Prevalence, F:M OR: contam obses = 1.70, p <0 .001, adj for age, edu, resid = 1.23 [95%CI, 0.98–1.54]; contam phob = 2.41, p < 0.001. | | | contam obses: M ~ F  contam phob: M < F | |
| Forresi  2022 | 1676 (959) | | 47 ± 9  (25–65) | | | occup. stress clinic  (ITA) | | | SCL-90  work stress CL | | | Cross-sectional: OCs severity: F = 2.21 ± 0.93 M = 1.96 ± 0.91, t = –4.94, Cohen’s d = 0.24, p < 0.001. | | | bulling ↑↑ OCs:  M < F | |
| Tang  2022 | 148F | | 21 ± 2  (17–27) | | | students  (CHN) | | | non-cons. “dirty kiss” task, MCR-C, PI-C-C | | | Symptom provocation. Non-consensual > consensual: mental contamination ↑ d ≈ 1.00, p < .001 (ANOVA: F(1,148)=37.86, η²=.20). Immoral > moral: mental contamination ↑ d ≈ 0.35, p = .010 (F(1,148)=4.56, η²=.03). Hygiene behaviors during the break: Fisher’s exact p = 0.819; contamination ~ washing: logistic regressions p = 0.50–0.95. | | | F: non-consensual kiss/ immoral →mental cont. | |
| Shafighi  2023 | 300  (166) | | 42 ± 9  (18–65) | | | post COVID 3–18 mo  (IRN) | | | OCI-R, DASS-21, PCL-5, PSQI | | | Prevalence. %OCs: F = 0.77 M = 0.64 F:M OR = 1.79 [95% CI, 1.13–2.84] p = 0.01. | | | OCs: M < F | |
| Jin  2024 | 27671  (13966) | | 20 ± 2  (15 – 26) | | | GEN +CT (CHN) | | | DOCS-SF, CTQ-SF | | | Cross-sectional: Within child trauma (CT): F:M OR = 1.47 [95%CI, 1.39 – 1.55]; Cohen’s d = 0.27; p < 0.001 | | | Child trauma↑↑ OCD: M < F | |
| Pol-Fuster  2025 | 16030 twins  (8 712) | | 15 → 24 | | | twin registry  (SWE) | | | OCI-R, BOCS, OBVQ-R, med records | | | Longitudinal twin registry study. %OCD: F = 0.01 M = 0.007. F:M OR = 1.43 [0.98–2.09], p ≈ 0.07; Bulling →OCD: F OR = 1.31 [1.17–1.46]; M OR = 1.32 [1.15–1.52] | | | OCD: M ~ F  Bulling →OCD:  M ~ F | |
| Huang  2025 | 410  (193) | | 30 ± 8 (16–65) | | | OCD  (CHN) | | | MINI, YBOCS; CTQ, OCI-R, OBQ | | | Cross-sectional + retrospective. Emotional abuse/physical neglect ↔ emotional distress ↔ OCD symptom dimensions, (M – F): network structure = 0.24, p = 0.14; global-strength = 0.25, p = 0.40; individual edges and node centralities all BH-adjusted p > 0.05. | | | Emot abuse/physical neglect ↔ emot dist ↔ OCs: M ~ F | |
| Lochner  2026 | 524  (294) | | NA  (18–52) | | | HC, OCD  (BRA, IND, NLD, ZAF, USA) | | | YBOCS,  DY-BOCS,  CTQ | | | Cross-sectional + retrospective. Total trauma: F: OCD = 43.19 ± 15.68, HC = 32.01 ± 6.93, d = 0.92 p < 0.001; M: OCD = 40.75 ± 12.76 versus HC = 34.96 ± 10.18, d = 0.50 p < 0.001.  Emotional abuse: F: OCD = 10.93 ± 5.15 versus HC = 6.85 ± 2.37, d = 1.02 p < 0.001; M: OCD = 9.72 ± 4.58 versus HC = 7.33 ± 3.06, d = 0.61 p < 0.001.  Physical abuse: F: OCD = 7.34 ± 3.98 versus HC = 5.79 ± 1.46, d = 0.52 p < 0.001; M: OCD = 7.14 ± 3.33 versus HC = 6.55 ± 2.68, d = 0.19 p = 0.137.  Sexual abuse: F: OCD = 7.01 ± 4.36 versus HC = 5.57 ± 1.77, d = 0.43 p < 0.001; M: OCD = 6.16 ± 2.64 versus HC = 5.58 ± 2.44, d = 0.23 p = 0.145.  Emotional neglect: F: OCD = 10.83 ± 4.78 versus HC = 7.76 ± 3.22, d = 0.75 p < 0.001; M: OCD = 10.72 ± 4.01 versus HC = 8.89 ± 3.63, d = 0.48 p < 0.001.  Physical neglect: F: OCD = 7.09 ± 2.79 versus HC = 6.03 ± 1.65, d = 0.46 p < 0.001; M: OCD = 7.01 ± 2.59 versus HC = 6.61 ± 2.45, d = 0.16 p = 0.210. | | | OCD: total trauma, PA, EN, PN: F ~ M  EA, SA: F > M | |
| Pinciotti  2025ab | 2066  (1190) | | 15 ± 3 (8–20) | | | trauma registry  (USA) | | | TX-CTRN: MINI-KID, CAPS-CA-5, TESI-C | | | Cross-sectional baseline analysis of longitudinal registry. OCD prevalence: F = 14.9% M = 5.4% F:M OR = 3.04 [95% CI, 2.17–4.28]. Gender-minority identity: OCD = 13.9% no OCD = 5.5%; OCD OR = 2.78 [95% CI, 1.74–4.44]. | | | OCD in trauma-exposed youth: F > M, gender minority > non-gender minority | |
| Pol-Fuster  2025 | 16030 twins (8799) | | 15 → 18 → 24 | | | CATSS cohort (SWE) | | | OBVQ-R; YBOCS, OCI-R, med records | | | Longitudinal, registry based. Prevalence: F = 0.99% M = 0.72% F:M OR = 1.38 [95% CI, 0.98–1.95], Fisher p = .072  Per 1-SD greater bullying victimization at 15, OCD diagnosis: F OR = 1.31 [95% CI, 1.17–1.46]; M OR = 1.32 [95% CI, 1.15–1.52].  OCS at 18 ~ bulling victimization at 15: F β = 0.15 [95% CI, 0.11–0.18]; M β = 0.11 [95% CI, 0.07–0.15]. OCS at 24 ~ bulling victimization at 15: F β = 0.14 [95% CI, 0.07–0.20]; M β = 0.09 [0.04–0.14]. | | | OCD: M ~ F  bulling victim → OCs: F ≥ M | |
| Pinciotti  2026 | 463  (353 AFAB) | | 27± 8  (18–72) | | | OCD  SGM (USA) | | | OBQ-SF; DOCS; YBOCS-II-SR | | | OCS ~ minority stress, β. SM: rejection sensitivity = 0.13, p = 0.02, discrimination = 0.11, p = 0.07. GM: n.s.  OCS dims ~ minority stress, β. GM: responsibility/harm ~ identity-based physical assault = 0.22 p=0.03, unacceptable thoughts~ identity-based physical assault = 0.26, p = 0.01, symmetry ~ internalized stigma = 0.22 p = 0.03; symmetry ~ identity-based sexual assault = −0.28 p = 0.01. | | | SM ≠ GM OCS ~ minority stress | |
| *COVID-19 pandemic* | | | | | | | | | | | | | | | | |
| Littman  2020 | 65  (38) | | NA  (18+ ) | | | OCD (ISR) | | | study-spec | | | Retrospective: COVID-19. ΔOCs > 0: 12F/9M, ΔOCs = 0: 14F/7M, ΔOCs < 0: 12F/11M, Fisher’s Exact test p = 0.60 | | | Δ OCs  M ~ F | |
| Mazza  2020 | 402  (137) | | 58 ± 13  (18-87) | | | COVID  patients  (ITA) | | | PCL-5,  OCI | | | Cross-sectional: COVID-19. OCs F-M d = 0.43 t = 3.64 p < 0.001; OCI ~ F, while controlling for prior psych Dx, hospitalization, β = 0.20 F = 9.29 p = 0.0025. Inflam MLR M – F d = 0.40 t = 3.32 p < 0.001; OCI ~ MLR r = 0.12 p = 0.04; OCI ~ MLR x F- NA. | | | post COVID inflam  M > F; post COVID inflame ~ OCI;  OCI M < F | |
| Ji  2020 | 13478  (8816) | | 21 ±3  (17-50) | | | student  (CHN) | | | YBOCS | | | Longitudinal. YBOCS ~ M: wave 1 β = 0.5 p < 0.001, wave 2 β = 0.67 p <0.001, wave 3 β =0.38 p < 0.001. | | | YBOCS lockdowns  M > F | |
| McKune  2021 | 280  (145) | | NA  (8 – 18) | | | K-12  (USA) | | | KAP,  study-spec | | | Cross-sectional: COVID-19. Prevalence of Risk for OCD M = 0.26, F = 0.38, F OR=1.76 [95%CI, 1.03-3.03]. | | | risk for OCD  M < F | |
| AlHusseini 2021 \| 2186 (1322) \| NA (18+ ) \| GEN (SAU) \| OCI-R \| Cross-sectional: COVID-19. Prevalence of OCD symptoms during COVID pandemic M = 0.67, F = 0.59, M OR= 1.42 [95%CI, 1.19 – 1.70]. \| likely OCD M > F; Ahmed 2021a \| 524 (301) \| NA (18+ ) \| GEN (EGY) \| YBOCS \| Cross-sectional: COVID-19. Prevalence of OCD symptoms, elevated: health care workers (HCW) = 0.29, non HCW = 0.28; moderate/severe: HCW = 0.03, non HCW = 0.065, p = 0.047; OCD ~ F β = 1.16 [95%CI, 0.06-2.27]. \| elevated OCs M < F; Ahmed 2021b \| 182 (98) \| 47 ± 17 (18–80) \| COVID patients (EGY) \| PSQI, PCL-5, SCL-90 OCS \| Cross-sectional: COVID-19. Prevalence of OCD symptoms during COVID pandemic F = 0.204 M = 0.19, F:M OR = 1.134 [95%CI, 0.44 – 2.94], p = 0.79 \| likely OCD M < F; Alateeq 2021 \| 2909 (2149) \| 30 ± 11 (18+ ) \| GEN (SAU) \| BOCS, PSS \| Cross-sectional: COVID-19. Incidence of OCD symptoms during COVID pandemic. Obs: F = 0.603 M = 0.569 F:M OR = 0.87 [95%CI, 0.44 – 2.94], p = 0.76. Comps: F = 0.47, M = 0.44, F:M OR = 1.11 [95% CI, 0.92 – 1.34], p = 0.28. \| new OCD M ~ F; Carmi 2021 \| 113 (57) → 90 (43) \| 34± 11 (8–73) \| OCD (ISR) \| CGI-I, COVID obs \| Longitudinal: worsening of OCs 2 &6 mo after COVID-19 lockdown. Δ OCs at 2 mo: F = 56.1%; M = 43.9%; χ²(1) = 1.54, p = 0.21; Δ OCs at 6 mo: F = 39.5%; M = 36.2%; χ²(1) = 0.08, p = 0.77; COVID-19 as a new obsession (2-mo): F = 42.1%; M = 39.2%; χ²(1) = 0.08, p = 0.77 \| COVID↑↑Δ OCs: M ~ F; Di Consiglio 2021 \| 454 (386) \| 25 ± 4 (19–54) \| student (ITA) \| SCL-90-R \| Cross-sectional, multi-wave study. OCs severity F: 1–10 w of lockdown = 53.74 ± 11.75; 3–13 mo pre = 48.70 ± 11.62; 5–7 mo post = 47.97 ± 10.01; 9–12 mo post = 49.31 ± 10.65; η² = 0.027, p = 0.015. M: 1–10 w of lockdown = 50.12 ± 10.88; 3–13 mo pre = 47.80 ± 10.91; 5–7 mo post = 48.66 ± 10.45; 9–12 mo post = 48.89 ± 10.24; η² = 0.032, p = 0.098. \| COVID↑↑Δ OCs: M < F; Ferreira 2021 \| 420 (203) \| med = 53 (IQR=23) \| phys (PRT) \| OCI-R \| Cross-sectional: COVID-19. Severe OCs: F = 18.2% vs M = 17.8% χ2(1) = 0.01 p=0.92. OCs: M = 18.7 ± 12.4 F = 17.6 ± 11.3, Cohen’s d = 0.09, p = 0.36; \| COVID↑↑ OCs: M ~ F; Fontenelle 2021 \| 829 (436) \| 39 ± 13 (18-82) \| MTurk (USA) \| DOCS \| Cross-sectional: COVID-19 (current + retrospect). OCs current (controlling for pre): β_M_= –0.17 (0.07), p = 0.03; β_M_ = –1.15, p = 0.012 \| COVID↑↑ OCs: M < F; Højgaard 2021 \| 201 (132) \| 40 ± 2 (18+ ) \| GEN (DEN) \| YBOCS SR \| Cross-sectional: retrospective Δ in OCs dues to COVID-19. F = 2.76, M = 1.61, Cohen’s d ≈ 0.37 t(199) = 2.55, p = 0.012 \| COVID↑↑Δ OCs: M < F; Jelinek 2021 \| 179 (135) \| 40 ± 12 (18-80) \| OCD (GER) \| YBOCS \| Longitudinal: ΔOCs following lockdown 3-mo FU. M = –2.57 p = 0.02, F = –0.17, p = 0.03; Cohen’s d = 0.34 p = 0.02. \| ↓OCs: M > F; Othman 2021 \| 386 (293) \| 31 ± 11 (18+ ) \| GEN (LBN) \| YBOCS \| Cross-sectional: COVID-19. OCs F-M Cohen’s d = 1.26 t = 5.23 p < 0.001; hygienic prevention practices F-M Cohen’s d = 0.49 t = 2.03 p < 0.05. \| OCs/hygienic M < F; Samuels 2021 \| 2117 (1137) \| 46 ± 16 (18 - 89) \| GEN (USA) \| CIS, DY-BOCS OCI-R \| Longitudinal: pre-during COVID-19. COVID prevalence of contam obs M: F OR = 1.7 [95%CI, 1.39-2.1]; prevalence of contam phobias M:F OR = 1.7 [95%CI, 1.39-2.1]; pre-post Δ OCI-R from 0-9 to 21+ M:F OR = 1.79 [95%CI, 0.73-4.37]; pre-post Δ OCI-R exclude contam from 0-9 to 16+ M:F OR = 1.64 [95%CI, 0.72-3.77]. \| contam M > F Δ OCs M ~ F; Liao 2021 \| 110 (53) \| 30 ± 9 (18-60) \| OCD (CHN) \| OCI-R \| Longitudinal: pre-during -1 year f/u COVID-19: Δ OCI-R ~ COVID x F = 1.51, p = 0.034 \| Δ OCs M < F; Meda 2021 \| 358 (286) \| 21 ± 2 (18-30) \| student (ITA) \| OCI-R \| Longitudinal: pre-during-post COVID-19 lockdown. Δ OCI-R ~ lockdown x F = -3.3 p = 0.41 \| Δ OCs M ~ F; Arora 2022 \| 250 (198) \| 21 ± 1.3 (18–25) \| med student (IND) \| YBOCS, FCV-19S \| Cross-sectional: COVID-19. Prevalence of OCD symptoms during COVID pandemic F = 0.14, M = 0.174, F:M OR = 0.76 [95% CI, 0.34 – 1.72], p = 0.13. \| OCs M ~ F; Benatti 2022 \| 116 (53) \| 40 ± 13 (18+) \| OCD (ITA) \| clinical interview \| Cross-sectional: worsening of OCs during COVID-19. Obsessions: Past recurring F = 0.46, M = 0.79, p = 0.033; New F = 0.46, M = 0.32, p = 0.341. Compulsions: Past recurring F = 0.42, M = 0.63, p = 0.196; New F = 0.42, M = 0.26, p = 0.320. \| Δ past obs: M < F; Δ new obs/comp: M ~ F:; Brooks 2022 \| 594 (385) \| 35 ± 15 (18–78) \| GEN (USA) \| SCARED-A \| Cross-sectional: OCs severity during COVID-19. OCs: F = 7.84 ± 3.46; M = 6.91 ± 3.21; d = 0.28, p = 0.04. OCs ~ distress, r: F = 0.42, p < 0.001; M = 0.37, p < 0.001; F vs M z = 0.71, p = 0.48. OCs ~ depr, r: F = 0.44, p < 0.001; M = 0.41, p < 0.001; F vs M z = 0.36, p = 0.72. OCs ~ anx, r: F = 0.77, p < 0.001; M = 0.75, p < 0.001; F vs M z = 0.28, p = 0.78. OCs ~ Hopelessness, r: F = 0.38, p < 0.001; M = 0.35, p < 0.001; F vs M z = 0.34, p = 0.73. \| OCs: M < F OCs ~ distress, depr, anx, hopeless: M ~ F; Pacitti 2022 \| 20241 (16309) \| 39 ± 13 (18–65) \| GEN (ITA) \| DOCS \| Cross-sectional: clinically sign. OCs (DOCS ≥ 30) during COVID-19. F = 52% M = 39%, F:M OR = 1.69 [95%CI, 1.55–1.84] p < 0.001. \| High OCs: M < F; Radhakrishnan 2022 \| ~71% of nonfed EDs \| 0-17 \| help-seeking (USA) \| medical records \| 3 cross-sectional waves: 2020, 2021, 2022. OCD-related visit ratio (ref = 2019): **2020 (Mar-Jan)**: F = 1.81 [95%CI, 1.67–1.96] M = 1.37 [95%CI, 1.26–1.49] F:M OR = 1.20 [95%CI, 1.10–1.31) ↑ (p < .001]; **2021 (Jan)**: F = 1.39 [95%CI, 1.30–1.49]; M = 1.03 [95%CI, 0.96–1.11]; F:M OR = 1.20 [95%CI, 1.12–1.29] p < .001; **2022 (Jan):** F = 1.24 [95%CI, 0.94–1.63] M = 0.76 [95%CI, 0.58–1.00] F:M OR = 1.31 [95%CI, 0.98–1.74] ns. \| COVID ↑↑OCD: M < F; Taner 2022 \| 7880 (3271) \| med 10 → 11 \| help-seeking (TUR) \| medical records \| 2 cross-sectional waves: (2019, 2020). Relative prevalence post vs pre OR F = 1.19 [95% CI, 0.94–1.50]; M = 1.99 [95% CI, 1.57–2.52]; F:M OR = 0.60 [95% CI, 0.43–0.83], p < .01. \| OCD: M > F; Dehghani 2023 \| 320 (208) \| 34 ± 9 (18–65) \| help-seeking (IRN) \| FCV-19S, PI-WSUR \| Cross-sectional: OCs severity during COVID-19. Fear of COVID-19: F = 17.29 ± 5.81; M = 15.92 ± 5.60; d = 0.24, p = 0.002. Washing compulsions: F = 6.75 ± 4.24; M = 5.91 ± 3.89; d = 0.21, p = 0.03. Impulses to harm self/others: F = 2.41 ± 1.09; M = 3.18 ± 1.16; d = 0.69, p = 0.004. Other OCD dimensions: ns (p > 0.05). \| Imp. to harm: M > F Wash: M < F; Jensen 2023 \| 186824 (115831) \| 34 ± 9 (18–65) \| GEN (NOR) \| ICPC-2 codes \| Longitudinal; pre-pandemic vs pandemic help seeking, Δ consults rates, %. M = 0 F = 2, Cohen’s d = 0.02 p > 0.99 \| Δ consults rates: M ~ F; Audet 2024 \| 653 (343) \| 43 ± 12 (18+ ) \| GEN (CAN) \| VOCI-C, CSS, FSQ-MV \| Cross-sectional: OCs severity during COVID-19. Severity of contamination symptoms during COVID pandemic: F > M d = 0.54, p < 0.001 (mean & SD not reported); Feared self-perceptions → Contamination: F = 0.49 p < 0.001; M = 0.47 p < 0.001, F vs M ΔCFI < 0.01, p > 0.05; COVID stress → Contamination: F = 0.37, p < 0.001; M = 0.35, p < 0.001; F vs M ΔCFI < 0.01, p > 0.05. \| Contm: M < F Fear/stress → contm M ~ F; Gümüşsoy, 2024 \| 726F \| 25 ± 1 (20–76) \| GEN (TUR) \| Fear of COVID-19 scale; BSI \| Cross-sectional: OCs severity during COVID-19, Pearlson r. Fear of COVID-19 ~ OCS = 0.43 p < 0.001 \| F: Fear of COVID-19 ↑↑ OCS; Köktaş 2024 \| 528 (329) \| 21 ± 2 (18–25) \| STU (TUR) \| VOCI \| Cross-sectional: OCs severity during COVID-19. Contam: F = 14.64 ± 12.05; M = 14.62 ± 9.51; d = 0.00, p < 0.05; Controlling: F = 8.31 ± 6.31; M = 7.05 ± 4.74; d = 0.22, p < 0.05; Obsess: F = 16.64 ± 11.85; M = 14.88 ± 9.49; d = 0.16, p = 0.003; Accumul.: F = 9.30 ± 7.31; M = 8.41 ± 5.85; d = 0.13, p = 0.003; Just-feeling-right: F = 16.29 ± 12.06; M = 13.45 ± 8.93; d = 0.27, p < 0.05; Indecision: F = 8.76 ± 6.23; M = 7.11 ± 4.86; d = 0.29, p < 0.05. \| OCs: M < F; Li 2024 \| 4246 (2978) \| 38 ± 9 (18+ ) \| health care (CHN) \| SCL-90-R, online \| Cross-sectional: provisional OCD during COVID-19: F:M OR = 1.69 [95%CI,1.19–2.40]. \| OCD in health care: M < F; Ghaderi 2025 \| 593 (324) \| 41 ± 12 (11–68) \| GEN (IRN) \| OCI-R, online \| Cross-sectional: provisional OCs during COVID-19: F = 78%; M = 67% F:M OR = 1.76 [95% CI, 1.22–2.55] χ² = 9.28 = p = 0.002 \| provisional OCs: M < F; Seo 2025 \| Population based \| 20+ \| GEN (KOR) \| \| Monthly OCD incidence/100,000. M: pre-pandemic = 0.45 [95% CI, 0.42–0.47], pandemic = 0.55 [95% CI, 0.51–0.59]; IRR = 1.23 [95% CI, 1.13–1.35], p < 0.001. F: pre-pandemic = 0.30 [95% CI, 0.29–0.32], pandemic = 0.47 [95% CI, 0.44–0.51]; IRR = 1.55 [95% CI, 1.42–1.70], p < 0.001. ΔOCD incidence level at February 2020 (/100,000): M = 0.038 p = 0.34 F = 0.065 p = 0.018 Δ monthly OCD incidence slope after February 2020 (/100,000/month) β: M = 0.007 p=0.042 F = 0.011 p<0.001 \| OCD incidence: M>F COVID ↑↑ OCD incidence: M < F COVID ↑↑ monthly ΔOCD: M ≤ F; Schuurmans 2026 \| 1214 (666) \| 56 ± 14 (18+ ) \| GEN (DEU) \| OCI-R, online \| Prospective longitudinal, six assmnt over 48 months. “Gender had no significant effect on OCs trajectory” \| COVID~OCs: M ~ F; Meza 2024 \| 75 (55) → 64 (54) \| 27 ± 10 (18–63) \| students (USA) \| OCI-R, DOCS, FOCI-B \| Longitudinal; pre-pandemic vs pandemic: time x gender, η²: OCI-R = 0.001 p = 0.717; DOCS = 0.001 p = 0.687; FOCI-B= 0.023 p = 0.120. \| Δ OCs: M ~ F; Pellegrini 2024 \| 20720 (14720) \| 40 ± 14 (18–80) \| GEN (ITA) \| OCI-R \| Cross-sectional: provisional OCD (OCI-R ≥ 21): F = 12.3 %, M = 8.8 %; F:M OR = 1.45 (1.30–1.63), p < .001. New OCD during COVID: F:M OR = 1.35 [1.21–1.51], p < .001 \| High OCs: M < F New OCD: M < F; Silverman 2024 \| 524 (370) \| 20 ± 1 (18–25) \| Students (USA) \| DOCS \| Cross-sectional cohorts across three waves (2016–2021). Severity /Δ Severity (3 × 2 × 4 ANOVA), η²: OCs × F = 0.002 p = 0.177 (F ≥ M), ΔOCs × F = 0.006 p = 0.053 (F ≥ M), contam × F = 0.002 p = 0.252, Δcontam × F = 0.008 p = 0.118 (F ≥ M), unaccep thghts × F = 0.004 p = 0.134 (F ≥ M); Δunacc.thghts × F = 0.009 p = 0.085, η² = (F ≥ M), symm × F < 0.001 p = 0.501, Δsymm × F = 0.006 p = 0.224, harm × F = 0.001 p = 0.441, Δharm × F = 0.004 p = 0.308. \| COVID↑↑ ΔOCs: F ≥ M; Triantafillou 2024 \| 130 (73) \| 49 ± 18 (18–85) \| GEN (GRC) \| DOCS (C1) \| Longitudinal; 2020, 2022: DOCS 2022 x F = 1.19 ± 0.50, β_std_ = –0.13, p = .019 \| COVID OCS; M < F; Bostan 2025 \| 200 (134) \| 33 ± 11 (18–65) \| OCD, HC (TUR) \| YBOCS, PATCQ, CCPMS \| Cross-sectional, case–control design, retrospective pre–during COVID-19. OCs F = 27.16 ± 8.41; M = 23.91 ± 7.63; d = 0.41, p = 0.04. Δ OCs: F = 3.21 ± 4.11; M = 2.94 ± 4.02; d = 0.07, p = 0.69. Avoidance behavior: F = 10.42 ± 3.13; M = 9.91 ± 2.84; d = 0.17, p = 0.33. Compliance: F = 37.29 ± 5.81; M = 36.02 ± 5.34; d = 0.23, p = 0.21. \| OCs: M < F Δ OCs, avoid, comply: M ~ F | | | | | | | | | | | | | | | | |
| Course | | | | | | | | | | | | | | | | |
| Marcks  2011 | 113  (65) | | 35 ± 12  (18+) | | | OCD  (USA) | | | SCID,  LIFE-UP | | | Longitudinal, 15 years. Remission: F HR = 1.16 [95%CI, 0.58-2.33], Wald = 0.19 p = 0.67, controlling for marital status, comorbid MDD, SSRI use. | | | remission  M ~ F | |
| Fineberg  2013 | 591  (299) | | wave 1:  19-20 | | | GEN  (CHE) | | | SCL-90-R,  SPIKE | | | Longitudinal, 30 years in 7 waves. Remission: M HR = 0.80 [95%CI, 0.50-1.29]; Remission age: M = 37 [95%CI, 34-40] F = 35 [95%CI, 33-38] p = 0.41. | | | remission  M ~ F | |
| Visser  2014 | 379  (214) | | 36 ± 11 (18-77) | | | OCD  (NLD) | | | YBOCS-SC,  EuroQol,  SCID-I/P | | | Retrospective, 2 years. %F: Chronic = 0.59 non-Chronic = 0.52, OR = 1.31 [95%CI, 0.86-1.98] p = 0.21, controlling for onset, severity, and type of OCD, illness burden, comorbid, anx, depress, Big-5, family history | | | chronic  M ~ F | |
| Mancebo  2014 | 60 (20) | | 11 ± 3  (6-18) | | | OCD (USA) | | | Y-LIFE,  CY-BOCS | | | Longitudinal, 3 years. %F: Chronic = 0.68, Remitting = 0.71 p = 0.84. | | | Course  M ~ F | |
| Goldberg  2015 | 449  (198) | | 34 ± 11  (17-73) | | | OCD  (ESP) | | | SCID-CV,  PSLE,  FHRDC | | | Prevalence. SLE: M < F χ^2^(1) = 9.9 p = 0.002; Familial load (FL): M ~ F χ^2^(1) = 1.97 p = 0.16. Retrospective, 5 years. Chronic vs. intermittent/episodic ~ SLE, FL, F: FL β = -0.18 p = 0.04 OR = 0.83 [95%CI, 0.70-0.98], FL x F β = 0.26 p = 0.015 OR = 1.30 [95%CI, 0.91-1.84], FL x SLE β = -0.37 p = 0.056 OR = 0.69 [95%CI, 0.47-1] | | | chronic ↑↓ FL  nonSLE: M > F  SLE M ~ F | |
| Geiger  2024 | 213  (114) | | 38 ± 11 (18–79) | | | OCD (NLD) | | | SCID, YBOCS | | | 6-year naturalistic follow-up: Trajectory, F:M OR: Episodic vs Chronic = 0.34 [95%CI, 0.12–0.96], p = 0.041; Remitted vs Chronic = 0.63 [95%CI, 0.16–2.41], p = 0.50; Episodic vs Remitted = 0.54 [95%CI, 0.14–2.13], p = 0.38. | | | chronic: M < F  remission: M ~ F  episodic: M > F | |
| Jagannathan  2024 | 3335  (1779) | | 15 ± 1  (14–18) | | | GEN  (USA) | | | RCADS-OCD, CASI, DTS | | | Longitudinal (4 waves, 2013–2017): Trajectories, M:F OR: High-but-Remitting : Low-and-Stable = 0.64 (β_M_ = −0.45 p = 0.01); Moderate-but-Escalating : Low-and-Stable = 0.86 (β = -0.15 p = 0.45). ΔOCs in Moderate-but-Escalating: wave 7 x F: β_M_ = 0.45, p < 0.05 (d ≈0.30). | | | high-but-Remit:M> F; Moder-excal:  ΔOCs: M < F | |
| Zhang  2025 | w1: 3535 (2319) → w5: 2604 | | 18 ± 1 | | | student  (CHN) | | | OCI-R, CTQ-SF, PSS-10 | | | 5-wave longitudinal cohort, 2019–2022. *OCI-R ≥21*: w1 F = 29.7%, M = 33.3% F:M OR = 0.85 [95% CI, 0.67–1.07]. *OCI-R*: F ≈ M at all five waves, all p > 0.05. *Longitudinal trajectory x F:* Initial OCS level: β = -0.045 p > 0.05; Linear change: β = -0.076 p > 0.05; Quadratic change: β = 0.056 p > .05. | | | prevalence, severity, and three-year trajectory: M ~ F | |

Table A3.3. OCD and reproductive events (Females only).

| **First Auth, Year** | **N** | **age**  **mean±SD**  **(range)** | | **Pop** | **Meas.** | **Gender effects (F/M)** | |
| --- | --- | --- | --- | --- | --- | --- | --- |
|  |  |  |  |  |  | **Stats** | **Direction** |
| Reproductive events | | | | | | | |
| Moreira  2012 | 455 | 34 ± 13  (18+ ) | | OCD  (BRA) | YBOCS,  DY-BOCS,  BDI/BAI | Course, Menstr. phases. %: PMOCS-same = 0.503, PMOCS-worse = 0.497, p > 0.10; Aggres: PMOCS-same = 0.67, PMOCS-worse = 0.77 p = 0.02; Sex/relig: PMOCS-same = 0.45, PMOCS-worse = 0.62 p < 0.001; Misc: PMOCS-same = 0.86, PMOCS-worse = 0.92 p = 0.4, Sym/Order/Contam/Hoard: PMOCS-same ~ PMOCS-worse p > 0.27. AO Sx: PMOCS-same = 12.5 PMOCS-worse = 12.3 p = 0.72, AO OCD: PMOCS-same = 20.9 PMOCS-worse = 24.4 p = 0.001; Chronic/wax/wane: PMOCS-same ~ PMOCS-worse p > 0.59; BAI: PMOCS-same = 15.1 PMOCS-worse = 20.7 p < 0.001; BDI: PMOCS-same = 15.9 PMOCS-worse = 19.7 p = 0.001. Other factors. Hypothyr: PMOCS-same = 0.04, PMOCS-worse = 0.09 p = 0.04, Suicide Id: PMOCS-same = 0.28, PMOCS-worse = 0.47 p < 0.001; Suicide Att: PMOCS-same = 0.10, PMOCS-worse = 0.18 p =0.03; Mood stab meds hist: PMOCS-same = 0.07, PMOCS-worse = 0.15 p = 0.02; SSRI cur: PMOCS-same = 0.39, PMOCS-worse = 0.50 p = 0.02. | aggres/sex/relig,  AO-OCD, anx/depress,  suicide id/attm,  meds/hypothyr  PMOCS-worse > PMOCS-same |
| Guglielmi  2014 | 542 | 45 ± 15  (18+) | | OCD  (USA,  NLD) | YBOCS, study-spec | Course. Onset, % of the sample: Menarche = 0.13, pregn = 0.051, PP = 0.047, menopause = 0.037. Worse, % of the sample: premenstr = 0.38, pregn = 0.33, PP = 0.47, menopause = 0.33; ↑Sx 1^st^ pregn ~↑Sx 2^nd^ pregn: OR = 10.82 [95%CI, 4.48-26.16]; ↑Sx 1^st^ PP ~↑Sx 2^nd^ PP: OR = 6.86 [95%CI, 3.27-14.36]. | menses, pregn, PP, menopause → Sx in F |
| Li  2017 | 213 | NA  (20+) | | GEN  (CHN) | SCL-90 | Severity. OCs: Polycystic ovary syndrome (PCOS) =1.5 ± 0.4, no PCOS = 1.4 ± 0.3, d = 0.10 p = 0.037. | PCOS ↑↑ OCs |
| Mulligan  2019 | 40 | 21 ± 3  (18+) | | GEN  (USA) | IDAS-II | Course, Menstr. phases. Check Sx: follicular (*fol*) = 6.1 ± 2.9, luteal (*lut*) = 6.2 ± 2.5 t(40) = -0.19 p = 0.85; Check Sx ↑↑ progesterone (P4): fol r(38) = 0.06 p = 0.72, lut r(38) = 0.14 p = 0.39; Check Sx ↑↑ estradiol (E2): fol r(38) = 0.03 p = 0.87, lut r(38) = -0.10 p = 0.53; Check Sx ↑↓ ΔERN: fol r(40) = -0.23 p = 0.20, lut r(40) = -0.47 p < 0.01. | F: check Sx  fol ~ lut  check Sx↑↓ ΔERN  fol < lut |
| Tay  2020 | 8670 | 23 ± 2  (19-26) | | GEN  (AUS) | ACES,  SR Dx | Prevalence. Polycystic ovary syndrome (PCOS) ↑↑ OCD: OR = 1.8 [95%CI, 1.2-2.5]. | PCOS ↑↑ OCD |
| Song  2024 | 213 | (18+) | | reprod. treat  (CHN) | SCL-90 | Risk (crossectional). Assisted reproductive technology (ART) ↑↑ OCD: OR = 1.8 [95%CI, 1.0–3.2] | ART↑↑ OCD |
| Incidence/Prevalence Ante/PP | | | | | | | |
| Chaudron  2010 | 24→16  25±6 | Ante 27w+  /PP 1m | | GEN  (USA) | EPDS,  YBOCS | Prevalence. %OCD**:** Ante = 0.29 post = 0.25. Severity: Ante = 8.5±5.1 PP 1mo= 14.4±7.9 Cohen’s d = 0.89 p < 0.01. | OCS  ante < PP |
| Miller  2015 | 461  33 ± 5 | PP: 2w  6m | | GEN  (USA) | PHQ-9, STAI, YBOCS | Prevalence of Obs. %Contam: 2w = 0.16, 6m = 0.18, %aggres: 2w = 0.12, 6m = 0.17; %somat: 2w = 0.05, 6m = 0.05; Prevalence of Coms. %Check: 2w = 0.09, 6m = 0.12, %clean: 2w = 0.07, 6m = 0.08, %repeat: 2w = 0.04, 6m = 0.05.  Associate factors. %Married: OCD=0.79, subOCs=0.92, noOCs = 0.89%, p=0.039; # kids: OCD=1.3±0.5, subOCs = 1.7±0.8, noOCs = 1.5±0.8, p = 0.02; Race, Edu, Income, Age NS.  Comorbid. %Depress: OCD = 0.54, SubOCs = 0.24, noOCs = 0.11, p < 0.001; %Anx: OCD = 0.26, SubOCs = 0.08, noOCs = 0.02, p<0.001. | OCs by type  2 wks < 6 mo;  OCD↑↓married/# kids;  Depress/anx  OCD>subOCS>noOCD |
| Samuels  2026 | 256  →199 | Ante: 20–24w PP: 6m | | GEN  (USA) | MINI, YBOCS, OBQ-44, PASS, PSS | Incidence. 8.4 [95% CI, 5–14] between Ante 20–24 w and PP 6 m.  Change OCs ΔYBOCS ≥5. Ante → PP = 25%; Ante = 5%, PP = 20%  Risk factors. ***Psychological:*** OBQ-44 d = 0.74, p = 0.02; PASS d = 1.35, p < 0.001; PSS d = 0.66, p = 0.04.  ***Socioeconomic:*** no bachelor’s degree OR = 4.14, p = 0.047; unmarried OR = 5.52, p = 0.01; income <$50,000 OR = 4.96, p = 0.03; public assistance OR = 5.92, p = 0.01; unplanned pregnancy OR = 4.42, p = 0.02; ≥1 socioeconomic risk factor OR = 4.72, p = 0.02. | OCD ↑↑ PP < Ante  Risks: low socio-economic resources, obs beliefs, perceived stress |
| Barrett 2016 | 94  30 ± 4 | Ante: 13-40w  PP: 1-9m | | GEN  (GBR) | YBOCS, PTBC, RAS, RIQ | Severity. YBOCS: noPregn = 6.82, Ante = 9.04, PP = 11.0, p = 0.02. Prevalence. %Mild: noPregn = 0.30, Ante = 0.31%, PP = 0.34; %Moderate: noPregn = 0, Ante= 0.15, PP = 0.11; %Severe noPregn = 0, Ante = 0, PP = 0.06; PP vs noPregn: t(66) = −3.09, p = 0.003.  Distress. Ante = 44.35±27.9, PP = 65.1±25.3, t(40.07) = −3.34, p = 0.002. Intrusive Resp. Baby-related vs. Other: Ante t(51)= −1.51, p= 0.14, PP t(64) = −4.43, p < 0.001. | prevalence/severity  noPregn < Ante < PP;  distress Ante < PP  Baby-related Ante *~ PP*  PP: baby related > other |
| Fairbrother 2016 | 310  34 ± 6 | Ante: 28±6w  PP:0-3m | | GEN  (CAN) | OCI-R | Prevalence. %OCD**:** Ante = 0.029 [95%CI, 0.01-0.048], PP = 0.036 [95%CI, 0.015-0.057], Peri = 0.017 [95%CI, 0.017-0.061]. Onset. %OCD**:** Ante= 0.01 [95%CI, 0.001-0.021], PP = 0.029 [95%CI, 0.01-0.048]. | prevalence/ onset  Ante ~ PP |
| Uguz  2019 | 1482 | PP: 1d | | GEN  (TUR) | SCID-I | Prevalence**.** PP = 0.049, noPreg = 0.021, OR 2.31 [95%CI, 1.07-5.02], p = 0.03. | OC:  noPreg < PP |
| Osnes  2019 | 1563  31 ± 5 | Ante: 17w, PP: 8w | | GEN, (NOR) | MINI,  SCL-A, | Prevalence. Ante w17 = 0.025, PP w8 = 0.042, p = 0.008, PP OR = 1.72 [95%CI, 1.15-2.57]. | OCD:  Ante < PP |
| Malemela  2019 | 206  28±6 | Ante  w4-40 | | GEN  (ZAF) | OCI-R,  CAS, PI-WSUR,  EPDS,  PRAS | Prevalence. %OCD = 0.39. Sx severity. OCD-noOCD Cohen’s d p-value: Depress = 0.32 p = 0.027, Anger = 0.27 p = 0.061, Anx = 0.66 p = 0.001, Contam = 1.19 p = 0.001. OCI-R ~Associated factors. β, p-value: Age = 0.20 p < 0.01, gestation wks = 0.17 p <0.01, past complic = 0.17 p < 0.01, depress = 0.28 p < 0.01, Anger = 0.14 p = 0.10, anx = 0.24 p < 0.01. | Ante↑↑OCD;  Depress/anger/anx/contam  OCD > no OCD;  age/gestation time/previous complic/depress/anx ↑↑OCs |
| Shokuhi  2020 | 160  30 ± 5 | PP  NA | | GEN  (IRN) | PDM | Construct. PP distress, % var expl, α: General distress (GD) = 0.37, α = 0.60, Obsessive-Compulsive (OC) = 0.14, α = 0.62. Concur valid: GD↑↑anx = 0.53, GD↑↑depress = 0.42, GD↑↑stress = 0.46; OC↑↑anx =0.42, OC↑↑depress=0.40, OC↑↑stress = 0.56. | PP distress:  General & OS |
| Burton  2021 | 5  26-40 | PP 5 yrs | | OCD  (GBR) | semistruct  interviews | Common OC themes: (1) Hyper-responsibility, (2) Intrusive imagery, (3) Guilt/Shame, (4) Eventual Coping and acceptance |  |
| Beck  2022 | 43  NA | Ante/PP  NA | | OCs  (USA) | Blogs, content analysis | Common OC themes: (1) Intrusive Thoughts Take Hold, (2) Desperate to Protect, (3) Paralyzed by Shame and Fear, (4) Journey to Recovery. | Ante/PP↑↑Intrusive thghts, shame/fear, protect |
| Kawoos  2022 | 95 | Ante/PP | | GEN  (IND) | YBOCS | Prevalence. OCs severity post COVID: Ante = 8.38 ± 5.8 vs PP = 5.83 ± 5.8; t(93) = 2.14, p = 0.035 | OCs post COVID-19:  Ante > PP |
| Phillips  2022 | 232  18–47 | PP: 2-6m | | GEN  (USA) | OCI-R, pOCDS, infant-feeding | Prevalence. %(OCI-R > 21)=0.14. PP OCI-R ~Associated factors. Breastfeeding problems β =0.3, p≈.05; breastfeeding ≥2 mo RR = 0.85, [95% CI, 0.64–1.11]; repeatedly washing baby dishes RR = 2.37 [95% CI, 1.55–3.64], counting/organizing stored milk RR = 2.71, [95% CI, 1.20–6.12], worry about accidentally harming baby RR = 2.01 [95% CI, 1.19–3.41]; depressive symptoms OR = 7.73, [95% CI, 3.39–17.62, state anxiety r = .45, p<.001. | PP OCD ↑↑infant-care and feeding obsessions/compulsions |
| Reagu  2022 | 261  20–45 | Ante: ~38w | | GEN  (QAT) | YBOCS, study spec. | Prevalence. Severity. Ante = 3.4 ± 5.3, %YBOCS ≥ 8 = 0.21, %YBOCS ≥16 = 0.057, %YBOCS ≥25 = 1 case. Predictors of OCD: COVID impact on breastfeeding = 4.33, [95 %CI, 0.40–8.26] p = .031, r = .25, OCD symptoms ↑ vs pre-pandemic: 9 %→21 % | Ante OCD ↑↑ Covid |
| Vasileiou  2022 | 30  32 ± 5 | Ante:  23 ± 9w | | OCs  (GRC) | MINI, YBOCS, OBQ, Leyton Trait Scale, HADS | Prevalence, %OCs. Aggres = 0.73, contam = 0.53; wash =0.50, check = 0.43. Associated factors. OCP traits (Leyton ≥ 12), OR: symm = 7.4 [95% CI, 1.2–44.9] p = .020, wash = 7.6 [95% CI 1.5–37.4] p = .011, check = 0.44 [95% CI, 0.11–1.75], contam = 1.7 [95% CI, 0.44–6.6], aggr = 1.0 [95% CI, 0.26–3.8]; OBQ (Responsibility/Threat), OR: contam = 4.4 [95% CI 1.1–17.9] p =0 .036, wash = 4.2 [1.0–17.3], p =0 .040; OBQ (Importance/Control), OR: aggres = 4.3 [1.1–17.3] p =0 .039, HADS-D (Depression), OR: check = 5.0 [95% CI, 1.2–20.5], p = 0.027; total OCS severity ↑ (β ≈ 0.44, p =0 .004); HADS-A (Anxiety), OR: contam = 6.6 [95% CI, 1.6–27.2] p = 0.011, wash = 5.1 [95% CI, 1.2–21.4] p =0 .025. | Ante ↑↑ aggres, contam, check  OCP, Resp, Control, Depr, Anx ↑↑ Ante OCs |
| Garcia  2023 | 10  30- 35 | PP | | OCs  (USA) | semistruct  interviews | Common OC themes: (1) Obses. – safety, cleanliness (2) Compl. – checking, cleaning, reassurance/info-seeking (3) Emotion – fear, panic, anxiety, guilt/shame, suicidality, anger. | Ante/PP↑↑check, contam, reassure, emotion distress |
| Buhagiar 2024 | 350  30 ± 5 | Ante/PP < 6m | | GEN  (MLT) | OCI-R | Prevalence. %OCD = 0.39 | Ante/PP↑↑Likely OCD: |
| Nakić Radoš  2025 | 397  32 ± 5 | Ante: 16-41w, PP:  6-12w | | GEN  (HRV) | YBOCS  EPDS, ASI, IPIP-50, BRS | Prevalence. %OCD Ante = 0.15 PP = 0.15, Ante/PP = 0.098, Severity. Ante = 3.81±4.24 PP = 3.23±4.67 Cohen’s d = 0.14, p < 0.001. %OCD–PPD comorb. Ante = 0.33 OR = 2.6, p < .01, PP = 0.43 OR = 3.2, p < 0.001. Predictors of OCD, β. Ante: depr = 0.25 p <0.001, psych concerns = 0.21 p <0.001, neuro = 0.19, p = 0.001, resil = –0.13, p = 0.007. | Ante/PP↑↑Likely OCD  OCs: Ante > PP  Depr: Ante < PP |
| Pre-existing OCD: Changes Ante/PP | | | | | | | |
| Uguz 2011 | 52  27 ± 5 | Ante: 30±8w | | OCD  (TUR) | SCID-I, YBOCS | Progression. %ΔOCs: “ΔOCs > 0” = 0.32, “ΔOCs = 0” = 054, *“*ΔOCs < 0” = 0.14.  Associated factors, ΔOCs > 0 OR, p-value. History of MDD = 5.3 [95%CI, 1.4 – 20.4], p = 0.02, MDD at preg = 14.2 [95%CI, 1.5-133.8] p = 0.01, Anx Disd at Preg = 4.1 [95%CI, 1.2-14.1] p = 0.03. Obs, ΔOCs > 0 OR. Contam = 9.5 [95%CI, 1.1 – 79.8] p = 0.02, Sym = 4.0 [95%CI, 1.2-13.6] p = 0.036, Relig = 0.07 [95%CI, 0.004-1.3] p = 0.02; Comp, ΔOCs > 0 OR. Clean = 8.4 [95%CI, 1.0-70.8] p = 0.04, Order = 4.1 [95%CI, 1.2-14.1] p = 0.03, Aggres, Sexual, Hoard, somatic, check, repeat, count: OR $\cong$ 1 p > 0.05. | hist of MDD/ preg MDD & Anx ↑↑ ΔOCs > 0;  contam/clean/sym/order  ↑↑ ΔOCs > 0;  relig ↑↓ ΔOCs > 0 |
| Yakut 2019 | 37  28 ± 6 | PP: 1d, 6-8 w | | OCD  (TUR) | YBOCS, COPE, TEMPS-A | Progression. PP obsess: *1 day* =13.0 ± 1.7, *6 wks* = 9.4 ± 4.0; PP compuls: 1 day = 12.0 ± 2.3 6w=8.5 ± 4.8p; PP total: 1d = 25.0 ± 3.8, 6w = 17.9 ± 8.7 p < 0.001; ΔOCs 1d →6w, %: “ΔOCs < -25%” = 0.51, “ΔOCs > 25%” = 0.08, “ΔOCs ~ 0” = 0.41.  Coping Strategies. COPE-humor: “ΔOCs < -25%” = 8.1 ± 2.9, “ΔOCs ≥ 0” = 6.1 ±2.4, p=0.029, COPE-suppression of competing activities/: “ΔOCs < -25%” = 11.0 ± 1.4, “ΔOCs ≥ 0” = 9.3±2.1, p=0.014; other COPE subscales p > 0.11.  Associated factors. Hyperthymic tempt: “ΔOCs < -25%” = 9.9 ± 4.5, “ΔOCs ≥ 0” = 5.5 ± 4.1, p = 0.004; Depressive, Cyclothymic, Irritable, and Anxious (p > 0.05) | PP Severity:  1 day > 6 wks  PP coping strategies  humor/focus ↑↓ OCs  hyperthymic ↑↓ OCs |
| Chatterje  2021 | 19  26 ± 6 | Ante/PP | | OCD  (USA) | SCID-I/P, YBOCS, LIFE, PSR | Progression. Mean distress: Ante/PP = 3.12 ± 0.88; non-Ante/PP = 3.13 ± 0.89; Δ = −0.008 ± 0.76; p = 0.90. Clinical exacerbation (PSR ≥ 4): Ante/PP = 33.4%; non-Ante/PP = 32.0%; OR = 1.06 [0.80–1.42], p = 0.70. Remission (PSR ≤ 2): Ante/PP = 28.8%; non-Ante/PP = 30.5%; OR = 0.92 [0.67–1.27], p = 0.61. | OCs: Ante/PP ~ non- Ante/PP |
| Fairbrother  2024 | 97  31 ± 5 | Ante 37w/PP: 9w, 22w | | OCD  (CAN) | SCID-5 | Timing of onset. By phase: Ante=0.17, PP = 0.83 χ² = 29.3, p < 0.01. By weeks: Ante OCs = 22 ± 11 w, Ante OCD = 23 ± 9 w; PP OCs = 3.5 ± 7.2 w, PP OCD = 5.2 ± 6.8 w. | onset: Ante < PP |
| Levinson 2025 | 164  30 ± 5 | Ante: 25w; PP: 18m | | OCD  (USA) | SCID-V screen OCI-4 | Progression. OCs: Recovering = 29.3%; intercept = 9.64, slope = −0.53, p < 0.01;  Stable-low = 70.7%; intercept = 4.10, slope = −0.009, p > 0.05  Cont.: Recovering = 8.5%; intercept = 3.45, slope = −0.51, p < 0.01; Stable-low = 68.3%; intercept = 0.92, slope = −0.10, p < 0.01; Stable-high = 23.2%; intercept = 2.50, slope = 0.03, p > 0.05 | |
| Factors associated with changes in OCs, Ante/PP | | | | | | | |
| Abramowitz  2010 | 52  30 ± 5 | PP: < 12m | | ≥1 PP intrusion  (USA) | YBOCS, EPDS, STAI,  PHQ | Progression. OCs ~ PP time: obsess r = 0.13 p=0.36, compuls r= 0.14 p = 0.34; PP 0-3m vs > PP 3m p > 0.53. Associated factors. depress ↑↑ obsess = 0.25 p < 0.05, depress ↑↑ compuls = 0.38 p < 0.05, anx ↑↑ obsess = 0.31 p < 0.05, anx ↑↑ compuls = 0.37 p < 0.05; OCs severity: depress group = 5.3 ± 3.0, not depress group = 2.6 ± 1.6, p < 0.001. | OCs PP ~ time  depress/anx ↑↑ OCs |
| Altemus 2012 | 229  32 ± 1 | Ante:  1^st^ - 3d trimester  PP: 0-12m | | help-seeking (USA) | medical record | Prevalence, in MDD, Ante onset vs PP onset. History of OCD: OR = 0.61 [95%CI, 0.14 -2.60] p =0.50; current Intrusive Violent Thoughts: OR = 0.22 [95%CI, 0.08-0.62] p < 0.01; current other OCD Sx: OR = 0.31 [95%CI, 0.11-0.89] p = 0.03;  Prevalence. In Ante Intrusive Violent Thoughts: history of OCD = 0.12, history of subOCD = 0.08; in Ante other OCD Sx: history of OCD = 0.30, history of subOCD = 0.20. | PP Depress ↑↑ PP Intrusive Violent Thoughts/ other OCs |
| House 2016 | 208  33 ± 4 | Ante: 1^st^, 2^nd^, 3d trimester  PP: 0-6w,  7-24w,  25-52w | | OCD,  noOCD,  (USA) | SCID, YBOCS | Progression. OCs severity in OCD: Ante 1^st^ tri = 10.9 ± 8.9, 2^nd^ tri = 8.4 ± 9.3, 3d tri = 9.03 ± 8.3; PP “0-6w” =10.3 ± 10.1, “7-24w” = 10.1 ± 8.6, “25-52w” = 9.6 ± 7.6.  Associated factors. OCs ~ age: Comp r = − 0.25 p < 0.0001, Obses r = − 0.20, p = 0.002, overall r = − 0.24, p<0.001; Preg Loss: OCD OR = 0.48 [95%CI, 0.23-0.99] χ2 = 4.08, p = 0.04; Comorbidity: Panic Disorder OR = NA *z* = 2.89, p = 0.004, Social Anxiety OR = NA *z*= 2.88, p=0.004; BPD, depress, schiz, PTSD, GAD, SUD p > 0.10; OCs ~ Cesarian (vs Vaginal): β = 5.4, p = 0.04. | OCs Ante ~ PP  age /panic d/o, social anxiety/ Cesarian ↑↑ OCS  Preg Loss:  no OCD > OCD |
| Miller  2013 | 461→  329 | PP: 2w, 6m | | GEN  (ISL) | PHQ-9,  YBOCS | Prevalence. 2w PP = 0.11, 6m PP = 0.106. Risk factors. Age: OCD = 31.2 ± 4.9 HC = 33.9 ± 4.9 d = 0.55 p = 0.03, Education, %college: OCD = 0.79 HC = 0.87 p = 0.02; %Never married: OCD = 0.19 HC = 0.06 p < 0.01; Psych history %, Anxiety: OCD = 0.09 HC = 0.04, Depression/anxiety: OCD = 0.04 HC = 0.0, Bipolar: OCD = 0.02 HC = 0.002, OCD: OCD = 0.04 HC = 0, relation. counseling: OCD = 0 HC = 0.02; p = 0.02. Progression. 2w OCD → 6m OCD: R^2^ = 0.20 p < 0.01. | PP: Prevalence↑  Age, edu, married, prior anxiety, depress, bipolar, relation counseling → PP OCD; PP OCD persist |
| Miller  2015 | 461→  331  33 ± 5 | PP: 2w, 6m | | GEN  (USA) | YBOCS, PHQ-9, STAI | Associated factors. 2w PP: OCs (Depress-) = 1.3 ± 2.9, OCs (Depress+) = 4.0 ± 5.0, p < 0.01; Obsess (Depress-) = 0.7 ± 1.7, Obsess (Depress+) = 2.0 ± 3.2, p < 0.01; Comps (Depress-) = 0.5 ± 1.4, Comps (Depress+) = 1.1 ± 2.4, p = 0.02. 6m PP: OCs (Depress-) = 1.7 ± 3.3, OCs (Depress+) = 3.3 ± 4.5, p < 0.01; Obsess (Depress-) = 1.2 ± 2.0, Obsess (Depress+) = 2.2 ± 2.8, p < 0.01; Comps (Depress-) = 0.6 ± 1.6, Comps (Depress+) = 1.3 ± 2.6, p<0.01. | PP:  Depress ↑↑ OCD |
| Prevatt 2017 | 100  33 ± 5 | PP: <3m | | GEN  (CAN) | YBOCS, SASSI-3 | Associated factors. OCs (Subst abuse-) = 4.35 ± 5.95, OCs (Subst abuse +) =7.21 ± 7.71 p = 0.002; OCs (Subst depend-) = 4.51 ± 5.95, OCs (Subst depend+) =8.80 ± 8.46, p = 0.03 | Subst abuse/depend ↑↑  PP OCs |
| Fairbrother 2018 | 100  32 ± 4 | Ante: 36w  PP: 4w, 12w | | GEN  (CAN) | OBQ-44, PPII, BDI-II, MAF, PSQI | Risk factors. Ante OC beliefs →PP obsess: 4w = 0.34 p < 0.01, 12w = 0.36 p <0.001.  Ante OC beliefs → PP Accidental harm subscales (var expl r^2^), at 4w: Time = 0.13 p < 0.01, Distress = 0.31 p<0.01, Parenting = 0.11 p < 0.01, Life = 0.08 p < 0.01, Distract = 0.16 p<0.01, Behav responses = 0.23 p<0.01; at 12w: Time = 0.09 p < 0.01, Distress = 0.15 p < 0.01, Parenting = 0.08 p < 0.05, Life = 0.10 p < 0.01, Distract = 0.18 p < 0.001, Behav responses = 0.17 p < 0.01. Ante OC beliefs → PP Intentional harm subscales (var expl r^2^), at 4w: Time = 0.13 p < 0.05, Distress = 0.18 p < 0.01, Parenting = 0.08 p < 0.01, Behav responses = 0.16 p < 0.01; at 12w: Distress = 0.50 p < 0.01, Parenting = 0.36 p < 0.05, Behav responses = 0.41 p < 0.05. Associated factors, with PP OCs (Pearlson r). Poor Sleep Quality: 4w = 0.34 (p<0.01), 12w = 0.18 (NS); fatigue: 4w = 0.16 (NS), 12w = 0.44 (p<0.001); depress: 4w = 0.47 (p<0.001), 12w = 0.67 (p<0.001)**.** | Ante OC beliefs ↑↑PP intrusive thoughts;  PP 4w:  OCs ↑↓ sleep quality  OCs ↑↑ depress  PP 12w:  OCs ↑↑ fatigue/ depress |
| Howard 2018 | 545  32 ± NA  (16-48) | Ante: NA | | GEN  (GBR) | Whooley (W), EPDS | Prevalence. OCD = 0.02 [95%CI, 0.01-0.04]. Prevalence by pre-screened groups. OCD (W+) = 0.05 [95%CI, 0.033-0.07], OCD (W-) = 0.02 [95%CI,0.004-0.04], OCD (EPDS+) = 0.03 [95%CI, 0.01-0.05], OCD (EPDS-) = 0.02 [95%CI, 0.01-0.04]. | Ante depress ↑↑ OCD |
| Osnes  2020 | 530  31 ± 4 | Ante: 17w PP: 8w | | GEN, (NOR) | SCL-A, MINI, BIS | Prevalence, in insomnia, %. Ante 17w: OCs (Ins+) = 0.038, OCs (Ins-) = 0.038, OR =1 p = 1.0; PP 8w: OCs (Ins+) = 0.085, OCs (Ins-) = 0.033 OR = 2.3 [95%CI, 1.2 – 6.4] p = 0.03. | insomnia ≠ Ante OCs  insomnia ↑↑ PP OCs |
| Xie  2021a | 3346  28 ± 5 | Ante: 16±5w | | GEN  (CHN) | SCL-90-R | Severity. OCs (pre-COVID) = 1.41 ± 0.37, OCs (COVID) =1.43 ± 0.39, p = 0.25. | OCs (pre-COVID) ~  OCs (COVID) |
| Xie  2021b | 3346  28 ± 5 | Ante: 16±5w | | GEN  (CHN) | SCL-90-R, PSQI, FES | Associated factors, with Ante OCs (Pearlson r). Family cohesion ↓ (r = –0.24 to –0.41, p < .001); family conflict ↑ (r = 0.20 – 0.33, p < .001). | fam cohesion↑↓Ante OCs  fam conflict↑↑ Ante OCs |
| Grekin  2021 | 271  33 ± 5 | Ante → PP 4w, 8w, 12w | | GEN  (USA) | PCL-5, OCI-R, IDAS-GD, W-DEQ-A, MOS-SS, PES, PLDS | Associated factors, with PP OCs (Pearlson r). **4w:** PTSS =0.52 p< 0.01; depress = 0.58 p < 0.001; fear of childbirth = 0.38 p <0.01; pers support = -0.25 p < 0.01; obj. birth stress = 0.28 p < 0.01.**8w:** PTSS = 0.60 p < 0.001; depress = 0.63 p < 0.001; fear of childbirth = 0.35 p < 0.01; obj. birth stress = 0.41 p < 0.001; pers. support = –0.21 p < 0.05.**12w**: PTSS = 0.57 p < 0.001; depress = 0.55 p < 0.001; fear of childbirth = 0.31 p < 0.01; obj. birth stress = 0.22 p < 0.05. | PP OCs ↑↑ PTSS, depress, fear ,birth stress |
| Kimmel  2021 | 126  32 ± 4 | Ante 38w | | GEN  (SWE) | MINI, PPG-HRV | Associated factors, with Ante OCD: HRV 38 w: OCD = 4.75 ± 1.05 non-OCD = 5.73 ± 1.28 Cohen’s d = 0.77 p = 0.004. | HRV Ante:  OCD < non-OCD |
| Thompson  2021 | 369  30 ± 4 | PP <12mo | | GEN  (USA) | DOCS, COVID-distress | Associated factors, with PP OCs (v.s. non pregrn), η²: OCs = 0.02 p =.022 (d ≈ 0.29);  contam. = 0.03 p =.001 (d ≈ 0.35); resp for harm =0.05 p < .001 (d ≈ 0.46);  unaccep thghts < 0.001p =0.70; symm=0.001 p =0.51. OCs ~ COVID-distress × PP = 0.06 p =0.007, 95% CI [0.02–0.10]. | PP↑↑OCs  PP OCs ↑↑ COVID-distress |
| Mahaffey  2022 | 4451  31 ± 5 | Ante | | GEN  (USA) | SCID-5 OCD screen OCI-R; | Associated factors, with Ante OCs: pandemic-related stress = 0.17 p < 0.001; maternal age = -0.24 p < 0.001, 1^st^ pregnancy = -0.14 p < 0.01. | Ante OCs ↑↑ COVID stress  Ante OCs ↑↓age/1t pregn |
| Miller  2022 | Ante 276  PP=221  31 ± 5 | Ante: 28w-32w  PP: 6w-8w | | GEN  (USA) | MINI, IMAS | Prevalence Ante = 6.2 PP = 14  Associated factors. Pearlson r. Total maladjustment=0.15 p <0.05; family role = 0.21 p <0.01. | Ante < PP  PP OCs ↑↓family role |
| Hinds  2023 | 392  NA | Ante: 37w | | GEN (IRL) | YBOCS, interview | Prevalence. %OCD (>13 YBOCS) = 0.10, Risk for OCs severity, U. ante med Dx = −3.55, p < 0.001, current med treatment = −3.58, p < 0.001, ante medication history = −2.99, p = 0.003, relationship strain = 3.70, p < 0.001. | PP OCs ↑↑ante med Dx, med treat, medicat hist, relation strain |
| Miller  2023 | Ante=251  PP=216  31 ± 5 | Ant:e 28-32w,  PP: 6-8w | | GEN  (USA) | MINI, IMAS | Prevalence. Current OCD: Ante= 4.4%, PP = 3.2%.  Associated factors: LCA of intern Sx: OCD loads on anxious–fearful (social anxiety + agoraphobia + phobias + OCD) and mood-reactivity (mania + OCD), but not distress. | Ante > PP  OCD↑↑ anxious–fearful, mood-reactivity |
| Sun  2023 | 284  33 ± 4 | PP: 42 ± 7 d | | GEN  (CHN) | MINI,  SCL-90 | Associated factors, OR. High stress = 6.26 [95% CI, 1.35–29.09], p = 0.019, Baby female sex= 5.95 [95% CI, 1.35, 25.0], p = 0.022. | Stress/baby F sex ↑↑OCD |
| Drake  2024 | 167  31 ± 5 | PP: 3d, 4w, 12w | | GEN (USA) | IDAS-II, PTBC, PCL-C, TLEQ, PRAMS. | Prevalence. OCD: check =0.08, order = 0.06, wash =0.10. Risk factors. *PTSD → OCD & PP intrusive, r:* 4w: preoccup = 0.45 p < 0.01, neutr = 0.21 p = 0.01, check = 0.49 p < 0.01, order = 0.38 p < 0.01, wash = 0.31 p < 0.01, obses = 0.51 p < 0.01. 12w: preoccup = 0.32 p < 0.001, neutr = 0.29 p = 0.002, check = 0.46 p < 0.001, order = 0.39 p < 0.01, wash = 0.32 p < 0.01, obses = 0.46 p < 0.01. *PTSD → OCD, controlling for deprs, β:* 4w: Check = 0.33 p < 0.01; obses = 0.33 p < 0.01. 12w: check = 0.40 p < 0.01; obses = 0.34 p = 0.01. | PTSD↑↑check/obsess |
| Miller  2024 | 270  31 ± 6 | Ante  PP: 4w, 8w, 12 w | | GEN  (USA) | PCL-5 OCI-R | Progression & Associated factors, ante → PP (4w → 8w → 12w): hoard → wash = 0.15 (p<.05); neutr → avoid = 0.14 (p<.05); hyperarousal → dysphoria = 0.14 (p<.05); neg cogn in PTSD → order = −0.14 (p<.05). | neg cogn in PTSD → order  neutr → avoid |
| Wang  2024 | 122  38 ± 3 | Ante:28w  PP: 6w | | 35+  (CHN) | YBOCS, SAS, EPDS, SOC-13 | Associated factors, OR OCs. Pregnancy history = 1.82 [95%, 1.15–2.89], p = 0.008; complications = 1.67 [95%, 1.23–2.26], p < 0.001; fetal health = 1.75 [95%, 1.25–2.47], p <0 .001; SAS = 1.89 [95%, 1.06–3.37], p = 0.027; EPDS = 1.67 [95%, 1.10–2.53], p = 0.012; SOC-13 = 1.27 [95%, 1.07–1.52], p = 0.002. | anx, depres, obstetric risk↑↑Ante/PP OCs  resilience ↑↓ Ante/PP OCs |
| Abramowitz  2025a | 256  33 ± 4 | Ante: 20w  PP:6w, 6m | | GEN  (USA) | MINI, OCI-12, OBQ | Progression (Ante 20w→ PP 8w /PP12w, Cohen’s d): OCI-12 =0.04/0.06, p >0.10; subscales =0.01-0.05 /0.02-0.10 p > 0.10.  Associated factors: Ante responsibility/threat beliefs → PP 6m OCD: OR = 1.04, p < 0.01; PP 6m OCs severity: β = 0.38, p < 0.01; obsessing: β = 0.31, p < 0.01. Ante perfectionism/certainty beliefs → ordering: PP 6w β = 0.28, p < 0.01; PP 6m β = 0.31, p < 0.01. Prior OCD → PP 6m OCD: OR = 6.40, p < 0.0001. | OCs: Ante ~ PP  Ante resp/threat ↑↑ PP OCD/OCs; perfect/cert ↑↑ PP order; prior OCD ↑↑ PP OCD |
| Babinski  2025 | 488377  18–45 | PP: 6w, 12m | | GEN/  ADHD  (USA) | EHR, Dx codes | Associated factors, OCD. 6-w PP OCD: ADHD = 0.1%, no ADHD = 0.1%. 12-mo PP OCD: ADHD = 3.1%, no ADHD = 0.3%; 12-mo PP OCD: OR = 10.63 [95% CI, 9.52–11.87]. | ADHD ↑↑  PP OCD at 12 m |
| Ceylan  2025 | 265  28 ± 5 | Ante: 35w; PP: 38d | | GEN  (TUR) | PSI, OCS-mothers | Prevalence. PP baby-care OC behaviors (score >9) = 68.3%. Associated factors. Ante pregnancy complaints → PP baby-care OC behaviors: β_std_ = 0.14, p = 0.026, adj R² = 0.015 | Ante complaints ↑↑ PP baby-care OC behaviors |
| Hamlett  2025 | 32 ± 5 | PP ≤ 6m | | GEN  (USA) | LEC-5, PCL-5, BSI | Associated factors, OCs severity: ST > no ST, t(1276) = −3.53, p < 0.001.  Network: ABE–OC edge differed between ST and no-ST: ST r_partial_ = 0.04, no-ST r_partial_ = −0.03, p = 0.01; difference disappeared after accounting for prior mental-health history. | History of sexual trauma ↑↑ PP OCs |
| Rector  2026 | 196  34 ± 4 | Ante: 29w, PP =8w | | GEN  (CAN) | OCI-R; OBQ-44; PSS; salivary 5-HTTLPR, BDNF | Severity. OCI-R: Ante = 8.18 ± 4.25; PP = 7.14 ± 6.71 d = −0.19 p = 0.007  Risks. Ante stress ~ Ante obsessive beliefs: β = 0.47 p < 0.05; Ante obsessive beliefs → PP OCS change: β = −0.42 p < 0.05.  Genetic variant (HTTLPR, BDNF) x Ante obsessive beliefs → PP OCS change: neither variant, n = 44, β = −0.16 p > 0.10, one variant, n = 90, β = 0.46 p > 0.10, both 5-HTTLPR-s and BDNF-Met variants, n = 62, β = −0.59 p < 0.05. | OCS: Ante > PP  Ante obsessive beliefs ↑↓ PP OCS change (only in HTTLPR, BDNF variant) |
| Yolcu  2026 | 334  29 ± 5 | PP: 2–8w | | GEN  (TUR) | VBS,  OCS-mothers | Associated factors. Vulnerable baby perception → PP baby-care OC behaviors: βstd = 0.14, p = 0.007, ΔR² = 0.020. Younger age (≤25y): d = 0.36, p = 0.003; lower education: η² = 0.09, p < 0.001; lower income: η² = 0.03, p = 0.009; younger age at first motherhood (≤25y): d = 0.50, p < 0.001; postnatal hospitalization: d = 0.38, p = 0.01 | Vulnerable baby perception  ↑↑ PP baby-care OC behaviors |
| OCD as a Risk Factor for Associated Conditions, Ante/PP | | | | | | | |
| Lord  2011 | 18  32 ± 5 | PP: 17±7w | | OCD,  HC  (CAN) | CPT, sal cortisol, STAI, subj stress | Risk for PP stress reactivity. Induced stress → distress (OCD > noOCD η_p_^2^, p-value): Subj stress = 0.41 p = 0.006; Anxiety = 0.47 p = 0.002; Cortisol Release =0.53 p = 0.001. | PP OCD ↑↑ stress reactivity |
| Challacombe  2016 | 73  34 ± 5 | PP: 6m | | OCD, HC (GBR) | PSSS, GRIMS, MSES | Risk for aberrant parenting experience, OCD vs HC. Parental SE d = 0.96 p < 0.0001; Soc support d = 0.82 p <0.001; marital satisfy d = 1.11 p = 0.009; poor relation build OR = 5.6 p = 0.018; enjoy nappy d = 0.59 p = 0.002; enjoy feed d = 0.65 p p = 0.015; enjoy play d = 0.97 p 0.011. | Parenting experience:  OCD < HC |
| Furtado 2019 | 35  31 ± 6 | Ante: 27w+  PP: 2w, 6w | | Anx d/o  (CAN) | HAM-A,  OCI-R | Risk for PP anxiety. Ante OCs → ΔAnx (PP 6w – Ante), mean rank/sum of ranks: OCs (ΔAnx > 0) = 23.0/391, OCs (ΔAnx ≤ 0) =13.3/239, U = 68, z = -2.8, p = 0.004. | Ante OCs ↑↑ PP ΔAnx |
| Johansen  2020 | 336,552 | Ante: 2m PP: 6n, 12m | | GEN  (USA) | med records | Risk for PP depression. OCD, OR. 2 m = 7.72 [(%%CI, 6.75-8.83]; 12 m = 5.47 [95%CI, 4.86-6.16]. | Ante OCD ↑↑ PP depress |
| Nasiri  2021 | 13792544 | Nationwide Inpatient Sample, 1999–2015 | | GEN  USA | medical records | Prevalence: OCD in childbirth = 2.4%; (in 1999: 0.7%; in 2015: 5.6%)  Risk Ante OCD for mothers: Gestational hypertension = 1.50 [95%CI, 1.14–1.98], p = 0.004; Preeclampsia = 1.22 [95%CI, 1.05–1.41], p = 0.007; Premature rupture of membranes = 1.32 [95%CI, 1.15–1.52], p = 0.0001; Cesarean delivery = 1.08 [95%CI, 1.01–1.17], p = 0.026; Instrumental delivery = 1.17 [95%CI, 1.01–1.35], p = 0.032; Induction of labor = 0.92 [95%CI, 0.86–0.99], p = 0.026; VTE = 1.55 [95%CI, 1.02–2.37], p = 0.041 Risk Ante OCs for children. Preterm birth = 1.31 [95%CI, 1.17–1.46], p < 0.001; Small for gestational age = 1.08 [95%CI, 0.88–1.31]; Intrauterine fetal death = 1.09 [95%CI, 0.75–1.58]. | Ante OCD: 1999 <2015  Ante OCD ↑↑ PP complic  Ante OCD ↑↑ Preterm birth |
| Fairbrother  2022 | 388  33 ± 5 | PP: ≤ 9mo | | GEN  (CAN) | PPII, SCID-5, PBQ | Prevalence. OCD = 0.17 [95% CI, 0.14–0.21], UITs of infant harm = 0.44 [95%CI, 0.39–0.50]. Risk. UITs of infant harm → Infant aggression: 2.6% vs 3.1%, RR = 0.84 [95% CI, 0.25–2.87]. OCD → Infant aggression: 1.9% vs 3.5%, RR = 0.52 [95%CI, 0.07–2.00].  . | UIT/OCD ⊥ Infant aggression |
| Gibbs  2023 | 125 Ante → 96 PP  18–35 | Ante: ≥27w PP: ≈27w | | Rural  (USA) | YBOCS IBQ-R-SF | Risk Ante OCs for infant neg. affectivity: OC×SER β = 0.02 p = 0.39  Risk PP OCs for infant neg. affectivity: OC×SER β = -0.33 p = 0.02 | low SER:  PP OCs ↑ infant neg. affect |
| de la Cruz  2023 | 1431251  31 ± 5 | PP | | OCD,  GEN  (SWE) | ICD-10 codes, registry | Risk for adverse perinatal outcomes. OCD > noOCD, adjusted OR: Preterm birth = 1.25 [95% CI, 1.15–1.35], p < 0.001; Cesarean delivery = 1.24 [95% CI, 1.19–1.30], p < 0.001;  Induction of labor = 1.11 [95% CI, 1.05–1.18], p < 0.001; Low birth weight (<2500 g) = 1.13 [95% CI, 1.02–1.26], p = 0.02; Neonatal intensive care admission = 1.17 [95% CI, 1.10–1.25], p < 0.001; Apgar <7 at 5 min = 1.12 [95% CI, 1.02–1.23], p = 0.02. | adverse perinatal outcomes:  OCD > noOCD |
| Öztürkler  2025 | 350  19-45 | PP: 4-8w | | GEN  (TUR) | OCB-PPBC, BSES-SF | Risk for maternal dissatisfaction. OCB ~ poorer maternal satisfaction, r: late-preterm =0.311 p<0.001; term=0.206 p=0.006. Adjusted OCB ~ poorer maternal satisfaction, β: late-preterm =0.168 p=0.018 term=0.21 p=0.003. | OCs ↑↓ maternal satisfaction |
| Parekh  2025 | 667502  all ages | delivery | | GEN  (USA) | med records | Risk for cardiovascular event during delivery. OCD versus no OCD, aIRR = 1.26 [95% CI, 0.69–2.32]. After controlling for other psych Dx: aIRR = 1.01 [0.57–1.78] | OCD not a risk for cardiovascular |
| OCs in Fathers, Ante/PP | | | | | | | |
| Coelho  2014 | fathers 726 →  707 | Ante:  28-34w  PP:  30-60d | | GEN  (BRA) | MINI | Prevalence. %OCD: Ante = 0.034 PP = 0.018. Comorbid. Ante: bipolar mixed RR=7.8 [95%CI, 2.0-30.2], manic episodes RR=15.8 [95%CI, 2.5-99.0], PP: bipolar mixed RR = 17.4 [95%CI, 3.1-96.2], unipolar depress RR = 28.2 [95%CI, 6.7-119.4]. Progression. Bipolar mixed ante → OCD PP RR=10.9 [95%CI, 2.8-42.9]. Concordant OCD, mothers & fathers. Ante RR = 6.1 [95%CI, 1.8-21.2], PP RR = 5.9 [95%CI, 1.2-28.4]. | Fathers OCD:  Ante > PP  CC: Ante = bipolar,  PP = bipolar/unipolar  OCD fathers↑↑mothers |
| Assessments/education | | | | | | | |
| Pereira  2022 | 212  33 ± 7 | PP: 3-6m | | GEN  (PRT) | POCS, DIPD-PP | Prevalence. %OCD: 0.033; %OCs: ≥ 1 OCs = 0.76; ≥ 1 obsess=0.74; ≥ 1 comp.=0.42; CFA fit: χ²/df = 2.30, CFI = 0.93, TLI = 0.91, RMSEA = 0.086. Reliability: α = 0.91 (total), 0.82–0.92 (subscales). Convergent validity: r = .27–.42 with PDSS/PASS. Criterion: OCD > Dep/Anx p < .01. AUC = 0.97, cut-off = 20 (Sens 85.7 %, Spec 100 %). | POCS valid for ppOCD |
| Fairbrother  2023 | 574  33 ± 5 | Ante: 37w, PP: 9w, 21w | | GEN, (CAN) | DOCS, EPDS/EPDS-3A, SCID-5 | Prevalence. %OCD: Ante = 0.036, PP-2mo = 0.106, PP-5mo = 0.058. Screening accuracy:  DOCS AUC: Ante = 0.90 [95%CI, 0.80–1.00], PP-2mo = 0.81 [95%CI, 0.74–0.88], PP-5mo = 0.88 [95%CI, 0.80–0.96]; EPDS-Full AUC: Ante = 0.71 [95%CI, 0.54–0.88], PP-2mo = 0.79 [95%CI, 0.72–0.85], PP-5mo = 0.78 [95%CI, 0.66–0.90]; EPDS-3A AUC: Ante = 0.73 [95%CI, 0.55–0.91], PP-2mo = 0.79 [95%CI, 0.72–0.86], PP-5mo = 0.80 [95%CI, 0.68–0.92]. Best-performing subscales. Unacc thghts: Ante AUC = 0.86, J = 0.54, LR⁺ = 3.84; PP-5mo AUC = 0.85, J = 0.63, LR⁺ = 5.85; Resp/Harm: Ante AUC = 0.81, J = 0.53, LR⁺ = 5.82; PP-5mo AUC = 0.83, J = 0.47, LR⁺ = 2.81. | best screener: DOCS (unacc thghts)  best timing: late ANTE and 4-6 mo PP |
| Abramowitz  2025b | 255  33 ± 4 | Ante: 20w, 34w; PP: 6w, 6m | | GEN  (USA) | OCI-4, MINI, YBOCS | Psychometrics: Test-retest = 0.75–0.84. Convergent with YBOCS = 0.61–0.70, p < 0.004. OCD vs non-OCD AUC: Ante 20w = 0.75 [95% CI, 0.67–0.83]; PP 6m = 0.91 [95% CI, 0.86–0.96]. OCD vs GAD AUC: Ante 20w = 0.69 [95% CI, 0.57–0.81]; PP 6m = 0.79 [95% CI, 0.65–0.92]. Cutoff ≥3, Ante 20w/PP 6m: sensitivity = 0.62/0.79; specificity = 0.78/0.87. | OCI-4 screen: valid/reliable screen; cutoff ≥3; accuracy PP > Ante |
| Lennon  2025 | NA | OBGYN education (USA) | | | content analysis | OCD-related themes: Total= 7 commonly used resources; 3,222 total items; 36 mental health.  OCD-related = 3 items (~8%) | |
| Lord  2026 | 340  31 ± 5 | Ante: 35w PP: 5w | GEN  (FRA) | | POCS-French | Psychometrics: Cronbach's α = 0.90, single factor 57% of variance; convergent with OCI-R = 0.62 p <0.001, OCD vs non-OCD AUC = 0.90. | POCS-French: valid/reliable screen |
| Rast  2025 | 256  33 ± NA | Ante: 20-24w | GEN  (USA) | | YBOCS, MINI, OCI-R | Psychometrics: YBOCS total = 6.66 ± 7.96 (range 0–32). Cronbach's α = .95; obsessions α = .90; compulsions α = .91. Convergent with OCI-R = 0.61, p < 0.001. MINI-OCD = 16.60 ± 6.92 versus no OCD = 4.23 ± 6.07; t = −10.37, p < 0.001, d = 1.90. CFA fit: CFI = 0.99, TLI = 0.99 and SRMR = 0.026–0.027, RMSEA = 0.105–0.106. | YBOCS: valid/reliable |

Table A3.1. Gender differences in incidence and prevalence of OCD.

| **First Auth, Year** | **N** | **Age range** | **Pop** | **Sampling** | **Instr.** | **Assessors** | **Meas.** | | **Gender effects** | | | | | | | | | | | |
| --- | --- | --- | --- | --- | --- | --- | --- | --- | --- | --- | --- | --- | --- | --- | --- | --- | --- | --- | --- | --- |
|  |  |  |  |  |  |  |  |  | **Stat: % F, M** | **Stat. test** | | | | | | **p-value** | | **F:M OR** | | |
| Incidence | | | | | | | | | | | | | | | | | | | | |
| *all ages* | | | | | | | | | | | | | | | | | | | | |
| Veldhuis  2011 | 577085 | 0+ | primary h/c, (NLD) | Integrated Primary Care Information database, 1996-2007 | patient's history records | medical records | 1^st^ time OCD | | F = 0.20 [95%CI, 0.12-0.23]  M = 0.12 [95%CI, 0.10-0.14] | | | | | | p < 0.01 | | | 1.77 | | |
| Huang  2014 | 1000000 | 6+ | inpatients,  outpatients  (TWN) | general population registry-based prospective cohort design, 2000-2008 | ICD-9-CM | medical records: ambulatory,  hospital,  prescriptions. | 1^st^ time OCD ≤6 | | F = 0.028  M = 0.027 | [95%CI, 1.10 – 1.20] | | | | | | | | 1.2 | | |
|  |  |  |  |  |  |  | 1^st^ time OCD age 6-10 | | F = 0.008  M = 0.014 | [95%CI, 0.34-0.91] | | | | | | | | 0.6 | | |
|  |  |  |  |  |  |  | 1^st^ time OCD age 11-17 | | F = 0.016  M =0.026 | [95%CI, 0.45-0.83] | | | | | | | | 0.6 | | |
|  |  |  |  |  |  |  | 1^st^ time OCD age 18-24 | | F = 0.034  M =0.046 | [95%CI, 0.59-0.91] | | | | | | | | 0.8 | | |
|  |  |  |  |  |  |  | 1^st^ time OCD age 25-34 | | F = 0.035  M = 0.036 | [95%CI, 0.77-1.11] | | | | | | | | 1.0 | | |
|  |  |  |  |  |  |  | 1^st^ time OCD age 35-44 | | F = 0.036  M = 0.026 | [95%CI, 1.11-1.67] | | | | | | | | 1.4 | | |
|  |  |  |  |  |  |  | 1^st^ time OCD age 45-64 | | F = 0.031  M = 0.022 | [95%CI, 1.11-1.67] | | | | | | | | 1.4 | | |
|  |  |  |  |  |  |  | 1^st^ time OCD age > 64 | | F = 0.020  M = 0.019 | [95%CI, 0.77-1.43] | | | | | | | | 1.1 | | |
|  |  |  |  |  |  |  | current OCD ≥ 6 | | F = 0.060  M = 0.070 | [95%CI, 0.83-0.91] | | | | | | | | 0.8 | | |
|  |  |  |  |  |  |  | current OCD age 6-10 | | F = 0.012  M = 0.018 | [95%CI, 0.43-1.0] | | | | | | | | 1.0 | | |
|  |  |  |  |  |  |  | current OCD age 11-17 | | F = 0.028  M = 0.049 | [95%CI, 0.43-0.71] | | | | | | | | 0.6 | | |
|  |  |  |  |  |  |  | current OCD age 18-24 | | F = 0.068  M = 0.123 | [95%CI, 0.48-0.63] | | | | | | | | 0.6 | | |
|  |  |  |  |  |  |  | current OCD age 25-34 | | F = 0.078  M = 0.106 | [95%CI, 0.67-0.83] | | | | | | | | 0.7 | | |
|  |  |  |  |  |  |  | current OCD age 35-44 | | F = 0.089  M = 0.079 | [95%CI, 1.0-1.25] | | | | | | | | 1.1 | | |
|  |  |  |  |  |  |  | current OCD age 45-64 | | F = 0.065  M = 0.055 | [95%CI, 1.0-1.25] | | | | | | | | 1.2 | | |
|  |  |  |  |  |  |  | current OCD age > 64 | | F = 0.041  M = 0.041 | [95%CI, 0.77-1.25] | | | | | | | | 1.0 | | |
| Teixeira  2022 | 10463672 | 0+  (53±NA) | GEN  (USA) | insurance claims 2018–2020 | ICD-10 | medical records | 1st time OCD | not reported | | | [95%CI, 0.94–1.60] | | | | | | | | | 1.2 |
| *children/adolescents* | | | | | | | | | | | | | | | | | | | | |
| Rintala  2017 | 139017 | 10, 15 | live births 1987–2001 (FIN) | general population registry-based cohort design, 1987–2001 | hospital discharge register | medical records from specialty clinics | 1^st^ time OCD by 10y | | born 1987-89: F = 0.01 M = 0.02 | | | | | | p < 0.01 | | | 0.43 | | |
|  |  |  |  |  |  |  |  |  | born 1999-01: F = 0.06 M = 0.08 | | | | | | p < 0.01 | | | 0.69 | | |
|  |  |  |  |  |  |  | 1^st^ time OCD by 15y | | born 1987-89: F = 0.09 M = 0.12 | | | | | | p < 0.01 | | | 0.69 | | |
|  |  |  |  |  |  |  |  |  | born 1999-01: F = 0.28 M = 0.24 | | | | | | p > 0.10 | | | 1.18 | | |
| Dalsgaard 2019 | 1300000 | 0-18 | liveborn singletons 1995- 2016  (DNK) | The Danish Civil Registration System | ICD-10DCR | medical records | 1^st^ time OCD | | F = 0.96 [95%CI, 0.92-1]  M = 0.63 [95%CI, 0.56-0.72]  “*before age 10: equal, ages 10 – 18: increases faster in girls, by age 18: higher in girls*” | | | | | | | | | 1.5 | | |
| Steinhausen  2019 | 67347 | 0-18 | GEN  (DNK) | nationwide Danish registries, born 1995, longitudinal until 18 | registers  medical codes | medical records | 1^st^ time OCD | | F = 0.0063  M = 0.0046 | χ2 = 9.18 p=0.002, *“**incidence rate is higher in females starting age 14”* | | | | | | | | 1.4 | | |
| Ask  2020 | total population | 3-17 | GEN  (NOR) | database for reimbursement of health expenses, 2008-2015 | registers  medical codes | medical records | 1^st^ time OCD | | F: ‘2010’ = 0.0008, ‘2011’ = 0.0008, ‘2012’ = 0.0009, ‘2013’ = 0.001, ‘2014’ = 0.0012, ‘2015’ = 0.0012;  M: ‘2010’ = 0.00055, ‘2011’ = 0.0006, ‘2012’ = 0.0007, ‘2013’ = 0.0007, ‘2014’ = 0.0008, ‘2015’ = 0.0008 | | | | | | | | | 1.3 – 1.5 | | |
| *adults* |  |  |  |  |  |  |  | |  | | | | | | | | |  | | |
| Guimarães  2023 | 1560 (880) →  1244 (721) | 21±2  →  26±2 | GEN (BRA) | 1993 Pelotas Birth Cohort | MINI → MINI-Plus | trained prsnl,  interviews | new OCD at FU | F = 0.026  M = 0.009 | | [95%CI, 1.05–7.47] p=0.039 | | | | | | | | | 2.8 | |
| Prevalence in general population | | | | | | | | | | | | | | | | | | | | |
| *children/adolescents* | | | | | | | | | | | | | | | | | | | | |
| Nikolajsen 2011 | 2522 | 8-17 | students  (ESP) | school-based sampling, 15 schools | SCAS | SR | current clinic OCS | | F = 4.9  M = 4.5 | NA, *“n.s.”* | | | | | | | | 1.1 | | |
| Orgilés  2012 |  |  |  |  |  |  |  |  |  |  |  |  |  |  |  |  |  |  |  |  |
| Shams  2011 | 909 | 14-18 | students  (IRN) | clustered, random, two stage sampling | MOCI → SCL-90-R | SR | 1-w clinic OCS | | F = 6.5  M = 4.7 | χ2 = 0.432 | | | | | | p=0.51 | | 1.4 | | |
| Canals  2012 | 2023 | 9-13 | students  (ESP) | two step sampling, | LOI-CV →MINI-kid &CSI-4 parent | SR | current OCD | | F = 1.9  M = 1.6 | χ2 = 0.02 | | | | | | p = 0.89 | | 1.2 | | |
|  |  |  |  |  |  |  | current subOCD | | F = 3.8  M = 7.6 | χ2 = 9.43 | | | | | | p = 0.002 | | 0.5 | | |
|  |  |  |  |  |  |  | current OCS | | F = 4.7  M = 4.8 | χ2 = 0.01 | | | | | | p = 0.92 | | 1.0 | | |
| Vivan 2014 | 2323 | 14 ‒ 17 | students  (BRA) | school-based cluster two stage sampling  (7 high schools) | OCI-R→ SCID-TCI, KSADS-P, YBOCS | SR→  clinicians, interviews | 1-mo OCS | | F = 24.8  M = 14.4 | z = 4.25 | | | | | | p < 0.001 | | 3.36 | | |
|  |  |  |  |  |  |  | current OCD | | F = 4.9  M = 1.4 | z = 6.17 | | | | | | p < 0.001 | | 1.95 | | |
| Jaisoorya, 2015 | 7380 | 12-18 | students  (IND) | school-based sampling | CIS-R | SR | current OCD | | F = 0.5  M =1.1 | χ2 = 7.98 | | | | | | p = 0.005 | | 0.5 | | |
| Zijlmans  2017 (com) | 4632 | 8-11 | students  (NLD) | subsampling, Generation R prospective cohort | SOCS | parental reports | current OCS | | F = 4.3  M = 4.8 | χ2 = 0.67 | | | | | | p=0.42 | | 0.9 | | |
| Mohammadi  2021 | 29858 | 6-18 | Gen  (IRN) | Multistage cluster random sampling | K-SADS-PL | trained prsnl,  interviews | OCD | F = 3.8%  M = 3.1 | | | [95% CI 1.10–1.48] | | | | | | | | 1.27 | |
| Dissanayake  2024 | 8927 | 6-17 | GEN  (CAN) | representative  community based  Spit for Science study | CBCL/6–18, SR | parental reports | current OCD | NA | | | | | | | | | p =0.15 | | 0.65 | |
| Khan  2021 | 1783 | 15-19 | students  (SAU) | random sampling (preexisting Dx excluded) | PHQ-9, YBOCS | SR | subOCS  (8-15) | | F = 36.6  M = 37.3 | χ2 = 6.75 | | | | | | p = 0.02 | | 1.0 | | |
|  |  |  |  |  |  |  | modrt OCS  (16-23) | | F = 21.9  M = 15.8 | χ2 = 0.10 | | | | | | p = 0.75 | | 1.4 | | |
|  |  |  |  |  |  |  | severe OCS  (24-31) | | F = 6.4  M = 3.5 | χ2 =10.93 | | | | | | p < 0.001 | | 1.8 | | |
|  |  |  |  |  |  |  | extr OCS  (32-40) | | F = 0.96  M = 1.05 | χ2 = 7.97 | | | | | | p = 0.005 | | 1.1 | | |
|  |  |  |  |  |  |  | clinic OCS  (16-40) | | F = 29.4  M = 20.3 | χ2 = 0.03 | | | | | | p = 0.85 | | 1.4 | | |
| *adults* | | | | | | | | | | | | | | | | | | | | |
| Subramaniam  2012 | 6166 | 18 ‒ 70 | GEN  (GRC) | stratified sampling, 2009-2010 | CIDI 3.0 | trained prsnl,  interviews | life  OCD | | F = 3.2  M = 2.8 | χ2 not reported | | | | | | p = 0.59 | | 1.1 | | |
| Adam  2012 | 4181 | 18 ‒ 65 | GEN  (DEU) | representative, two step sampling, 1997 | CID-S, DIA-X/ M-CIDI | clinicians, interviews | 12-mo  OCD | | F = 0.9  M = 0.6 | [95%CI,  0.7-3.3] | | | | | | p > 0.10 | | 1.6 | | |
| Skapinakis  2013, 2019 | 4894 | 18 ‒ 70 | GEN  (GRC) | representative sampling, 2009-2010 | CIS-R | computerized SR supervised by trained prsnl | 1 wk  OCD | | F = 2.10  M =1.3 | χ2 = 5.06 | | | | | | p = 0.025 | | 1.7 | | |
|  |  |  |  |  |  |  | 1 wk  OCD | | F = 3.4  M = 2.1 | χ2 = 7.63 | | | | | | p = 0.006 | | 1.6 | | |
| Fineberg  2013 | 591→ 335 | 20/21 → 49/50 | GEN  (SWE) | two-step sampling, Zurich Cohort Study of Young Adults, 7w | SCL-90-R,  SPIKE, interviews | clinicians, interviews | 12-mo OCS | | F = 8.8  M =13.7 | z = 3.44 | | | | | | p < 0.001 | | 0.6 | | |
|  |  |  |  |  |  |  | 12-mo subOCD | | F = 9.3  M =10.1 | z = 0.87 | | | | | | p = 0.38 | | 0.9 | | |
|  |  |  |  |  |  |  | 12-mo OCD | | F = 5.3  M = 1.7 | z = 4.16 | | | | | | p < 0.001 | | 3.2 | | |
| Karg  2014 | 5653 | 18+ | noninstit.  (USA) | multistage cluster sampling, 2008–2012 | modified SCID I | clinicians, interviews | est. 12-mo OCD | | F = 0.5  M = 0.1 | χ2 = 236283 | | | | | | p < 0.001 | | 5.0 | | |
| Rodgers  2015 | 4547 → 591 | 20 ‒ 50 | GEN  (CHE) | two step sampling,  2010–2012 | SCL-90-R → SPIKE | clinicians, interviews | 7 years OCD | | 7.8 | χ2= 1.83 | | | | | | p = 0.18 | | 1.9 | | |
|  | 9829 → 1500 | 20 ‒ 41 | GEN  (CHE) | two step sampling,  1979–2008 | SCL-90-R  →SPIKE | clinicians, interviews | 12-mo OCD | | 5.8 | [95%CI, 0.5-1.5] | | | | | | | | 0.8 | | |
|  | 5535 → 3720 | 15 ‒ 66 | GEN  (CHE) | two step sampling,  2003-2006 | DIGS | clinicians, interviews | 12-mo  OCD | | 1.3 | [95%CI, 0.94-3.17] | | | | | | | | 1.7 | | |
| Hajebi  2018 | 7886 | 15 ‒ 64 | GEN  (IRN) | representative sampling, 2011 | CIDI 2.1 | clinicians, interviews | 12-mo OCD | | F = 6.8  M = 3.4 | χ2= 44.36 | | | | | | p < 0.001 | | 2.0 | | |
| Osland  2018 | 25097 | 15+ | GEN  (CAN) | representative sampling, 2012 | SR of lifetime clinician-given OCD Dx | | est. 12-mo OCD | | F = 1.04 [95%CI, 0.75-1.25]  M = 0.81 [95%CI, 0.60-1.03] | | | | | | | | | 1.3 | | |
| Shirzadi  2019 | 2102 →  822→ 139 | 20 ‒ 73 | GEN  (IRN) | two step sampling, Kermanshah county | GHQ-28 →SCID I&II | clinicians, interviews | est. 12-mo OCD | | F = 5.9  M = 3.4 | χ2= 7.56 | | | | | | p = 0.006 | | 1.7 | | |
| Subramaniam  2019 | 6126 | 18+ | GEN  (SGP) | stratified sampling of 3 main ethnic groups, 2016-2018 | CIDI 3.0 | trained prsnl, computer-assist interviews | 12-mo OCD | | F = 4.1  M = 3.1 | χ2 = 4.35 | | | | | | p = 0.037 | | 1.8 | | |
|  |  |  |  |  |  |  | life OCD | | F = 3.7  M = 2.1 | χ2 = 14.13 | | | | | | p = 0.0002 | | 1.3 | | |
| Subramaniam  2020 | 6126 | 18+ | GEN  (SGP) | stratified sampling of 3 main ethnic groups, 2016 | CIDI 3.0, YBOCS, SF-12 | trained prsnl,  interviews | 12-mo OCD | | F = 3.7  M = 2.1 | [95%CI,  1.1-2.9] | | | | | | p = 0.016 | | 1.8 | | |
|  |  |  |  |  |  |  | life OCD | | F = 4.1  M = 3.0 | [95%CI,  1.0-2.2] | | | | | | p = 0.084 | | 1.0 | | |
|  |  |  |  |  |  |  | 12-mo subOCS | | F = 4.0  M = 5.0 | [95%CI,  0.56-1.11] | | | | | | p > 0.10 | | 0.8 | | |
| Carta  2020 | 2267  (1254) | 18+ | GEN  (ITA) | stratified random sampling 6 regions | SCID, MDQ, BDS, SF-12 | clinicians, interviews | life OCD | | F = 2.0  M = 1.8 | [95%CI, 0.6-2.2] | | | | | | | | 1.2 | | |
| Lee  2021 | 12532 | 18-60 | GEN  (KOR) | nationwide community sample  KECA 2006-201 | K-CIDI,  DSM-IV | clinicians, interviews | obss | | F = 2.93  M = 2.45 | χ2 = 2.86 | | | p = 0.09 | | | | | 1.18 | | |
|  |  |  |  |  |  |  | comps | | F = 5.71  M = 4.18 | χ2 = 12.92 | | | p =0.0003 | | | | | 1.32 | | |
| *older adults* | | | | | | | | | | | | | | | | | | | | |
| Klenfeldt  2014 | 900 | 70 M/F,  78-92F | nondement.  (SWE) | subsampling, Swedish Population Register | SCID | trained nurse,  interviews | 1-mo OCS, age 70 | | F = 25.4  M = 24.6 | χ2 = 0.05 | | | | | | p= 0.83 | | 1.0 | | |
|  |  |  |  |  |  |  | 1-mo OCS, age 78-92 | | F = 22 | - | | | | | | - | | - | | |
|  |  |  |  |  |  |  | 1-mo OCD, age 70 | | F = 4.5  M = 1.3 | χ2 = 4.24 | | | | | | p=0.04 | | 3.5 | | |
|  |  |  |  |  |  |  | 1-mo OCD, age 78-92 | | F = 2.3 | - | | | | | | - | | - | | |
| Canuto  2018 | 3142 | 65-84 | GEN  (EU, ISR) | random sampling | CIDI65+ | trained prsnl, interviews | 12-mo OCD | | F = 0.8  M = 0.8 | F(1,20)=0.0 | | | | | | p = 0.91 | | 1.0 | | |
| *by age groups* | | | | | | | | | | | | | | | | | | | | |
| Alvarenga 2015 | 9937 | 6 ‒ 12 | students  (BRA) | school-based sampling | FHS, DYBOCS | parental reports | $\geq$1 current OCS age 6-12 | | β_M_ = 0.08 [95%CI, 0.08-1.52] p = 0.001 | | | | | | | | | 0.89 | | |
|  |  |  |  |  |  |  | $\geq$1 current OCS, age 6 | | F $\cong$ 3.1  M $\cong$ 5 | NA | | | | | | NA | | 0.62 | | |
|  |  |  |  |  |  |  | $\geq$1 current OCS, age 7 | | F $\cong$ 5.9  M $\cong$ 7.9 |  |  |  |  |  |  |  |  | 0.75 | | |
|  |  |  |  |  |  |  | $\geq$1 current OCS, age 8 | | F $\cong$ 5  M $\cong$ 9.5 |  |  |  |  |  |  |  |  | 0.53 | | |
|  |  |  |  |  |  |  | $\geq$1 current OCS, age 9 | | F $\cong$ 7  M $\cong$ 9.2 |  |  |  |  |  |  |  |  | 0.76 | | |
|  |  |  |  |  |  |  | $\geq$1 current OCS, age 10 | | F $\cong$ 7.5  M $\cong$ 9.9 |  |  |  |  |  |  |  |  | 0.76 | | |
|  |  |  |  |  |  |  | $\geq$1 current OCS, age 11 | | F $\cong$ 5  M $\cong$ 10.5 |  |  |  |  |  |  |  |  | 0.48 | | |
|  |  |  |  |  |  |  | $\geq$1 current OCS, age 12 | | F $\cong$ 9  M $\cong$ 9 |  |  |  |  |  |  |  |  | 1.0 | | |
| Cath  2017 | 15194 | 15-97 | twins & family  (NLD) | Netherlands Twin Register | Padua ABBR | SR | provisional OCD | | F vs M: OR: 0.99, z = −2.14, p = 0.032  “*primarily driven by 65 and older group*” | | | | | | | | | | | |
| Help-seeking | | | | | | | | | | | | | | | | | | | | |
| *general clinics* | | | | | | | | | | | | | | | | | | | | |
| Ghuloum  2011 | 1660 | 18-65 | primary h/c, (QAT) | random sampling,  12 clinics | DSM-based, study-spec | SR, supervised by trained nurses | current OCD | | F = 4.5  M = 2.3 | χ2 = 5.57 | | | | | | p = 0.018 | | 1.96 | | |
| Lihua  2014 | 1576 | 16+ | general hospitals  (CHN) | random sampling, stratified by hospital type | CIDI 3.0 | trained prsnl | 30-d OCD,  age ≥ 16 | | F = 2.84 [95%CI, 1.70–3.98]  M = 1.04 [95%CI, 0.32–1.76] | | | | | | p < 0.05 | | | 2.73 | | |
|  |  |  |  |  |  |  | 30-d OCD,  age 16-35 | | F = 3.52 [95%CI, 1.38–5.66]  M = 0 | | | | | | - | | | inf | | |
|  |  |  |  |  |  |  | 30-d OCD,  age 36-54 | | F = 2.26 [95%CI, 0.48–4.04]  M = 1.16 [95%CI, 0.56–1.76] | | | | | | p > 0.05 | | | 2.17 | | |
|  |  |  |  |  |  |  | 30-d OCD,  age ≥ 56 | | F = 3.08 [95%CI, 0.98–5.48]  M = 1.94 [95%CI, 0.26–3.62] | | | | | | p > 0.05 | | | 1.59 | | |
|  |  |  |  |  |  |  | 12-mo OCD,  age ≥ 16 | | F = 3.09 [95%CI, 1.90–4.28]  M = 1.56 [95%CI, 0.68–2.44] | | | | | | p < 0.05 | | | 1.98 | | |
|  |  |  |  |  |  |  | 12-mo OCD,  age 16-35 | | F = 4.93 [95%CI, 2.41–7.45]  M = 4.38 [95%CI, 1.85–6.91] | | | | | | p > 0.05 | | | 1.13 | | |
|  |  |  |  |  |  |  | 12-mo OCD,  age 36-54 | | F = 2.64 [95%CI, 0.71–4.57]  M = 1.55 [95%CI, 0.79–2.31] | | | | | | p > 0.05 | | | 1.46 | | |
|  |  |  |  |  |  |  | 12-mo OCD,  age ≥ 56 | | F = 1.54 [95%CI, 0.04–3.04]  M = 1.94 [95%CI, 0.26–3.62] | | | | | | p > 0.05 | | | 0.79 | | |
|  |  |  |  |  |  |  | life OCD,  age ≥ 16 | | F = 5.44 [95%CI, 3.88–7.00]  M = 2.61 [95%CI, 1.48–3.74] | | | | | | p < 0.05 | | | 2.08 | | |
|  |  |  |  |  |  |  | life OCD,  age 16-35 | | F = 6.69 [95%CI, 3.78–9.60]  M = 5.58 [95%CI, 2.74–8.42] | | | | | | p > 0.05 | | | 1.20 | | |
|  |  |  |  |  |  |  | life OCD,  age 36-54 | | F = 6.42 [95%CI, 3.47–8.37]  M = 1.55 [95%CI, 0.79–2.31] | | | | | | p < 0.01 | | | 4.14 | | |
|  |  |  |  |  |  |  | life OCD,  age ≥ 56 | | F = 3.08 [95%CI, 0.98–5.48]  M = 2.33 [95%CI, 0.49–4.71] | | | | | | p > 0.05 | | | 1.32 | | |
| Bener  2015 | 1475 | 16-65 | primary h/c, (QAT) | random sampling,  13 clinics | WHO‑CIDI | psych nurses, interviews | current OCD | | F = 13.5  M = 7.8 | χ2 = 12.24 | | | | | | p < 0.001 | | 1.86 | | |
| Sundquist  2017 | 5397675 | 18+ | primary h/c, (SWE) | Primary Care, Specialist Outpatient Care, Hospital Discharge records | registers  medical codes | medical records | current OCD | | NA | [95%CI, 1.38-1.46], p < 0.01 | | | | | | | | 1.42 | | |
| Snorrason  2019 | 498  (275) | 18-74 | behavioral health  (USA) | consecutive patients at Behavioral Health Partial Program, 7/2016 - 7/2017 | MINI | clinicians, interviews | current OCD | | F = 18.0  M = 20.0 | χ2 =0.32 | | | | P = 0.57 | | | | 0.9 | | |
| Jabeen  2020 | 59220 | 18+ | primary h/c, (PAK) | 11 public & 5 private hospitals | hospital Dx code | medical records | current OCD | | F = 4.1  M = 4.2 | χ2 = 0.37 | | | | | | p = 0.61 | | 0.98 | | |
| Wazir  2023 | 231  (108) | 18-89 | Psychiatric h/c, (PAK) | Community based | medical records | clinicians, interviews | current OCD | F = 0.045  M = 0.057 | | | | [95%CI, 0.30–2.01], χ² = 0.18 p = 0.67 | | | | | | | 0.76 | |
| Alsaweer  2024 | 614  (506) | 18–65 | primary h/c, (BHR) | snowball sampling | MOCI  YBOCS | SR | current OCS | “ no significant sex difference (p = 0.260–0.997) | | | | | | | | | | | | |
| *OCD specialty clinics* | | | | | | | | | | | | | | | | | | | | |
| *all ages* | | | | | | | | | | | | | | | | | | | | |
| Algin  2018 | 400 | 8-63 | OCD  (IND) | consecutive patients at OCD clinic over 2 yrs | SCID-I, YBOCS | clinicians, interviews | 12-mo OCD | | F = 41.2% | z = 5.200 | | | | | | p < 0.001 | | 0.7 | | |
| *children/adolescents* | | | | | | | | | | | | | | | | | | | | |
| Deepthi  2018 | 451 | 17 | OCD  (IND) | retrospective chart review, one clinic, 1998-2014 | unstructured  interviews,  YBOCS | clinicians, interviews | life OCD | | F = 27.9% | Z = 4.672 | | | | | | p < 0.001 | | 0.4 | | |
| Sharma  2019 | 173 | 5-17 | OCD  (IND) | consecutive patients,  6 clinics | YBOCS | clinicians, interviews | current OCD | | NA | z = 4.025 | | | | | | p < 0.001 | | 0.5 | | |
| *adults* | | | | | | | | | | | | | | | | | | | | |
| Cherian  2014 | 545 | 18+ | OCD  (IND) | consecutive patients at OCD clinic over 5 yrs | MINI, YBOCS | clinicians, interviews | 12-mo OCD | | F = 39.1% | z = 5.10 | | | | | | p < 0.001 | | 0.6 | | |
| Medeiros  2017 | 236 | 18-72 | OCD  (USA, | OCD clinics, MA RI | SCID-I, YBOCS, DYBOCS | clinicians, interviews | life OCD | | F = 57.6 | z = 2.34 | | | | | | p = 0.02 | | 1.4 | | |
|  | 951 | 18-72 | BRA) | 8 clinics from 5 states |  |  |  |  | F = 58.3 | z = 5.12 | | | | | | p < 0.001 | | 1.3 | | |
| Dell’Osso 2017 | 416 | 18+ | OCD  (world) | consecutive outpatients,  multiple clinics | SCID I&II,  YBOCS | clinicians, interviews | current OCD | | F = 57.5 | z = 2.86 | | | | | | p = 0.004 | | 1.4 | | |
|  |  | 18-42 |  |  |  |  |  |  | F = 52.1 | z = 0.58 | | | | | | p = 0.56 | | 1.1 | | |
|  |  | 42+ |  |  |  |  |  |  | F = 63.1 | z = 3.70 | | | | | | p = 0.0002 | | 1.7 | | |
| Tripathi  2018 | 945 | 18-72 | OCD  (IND) | 16 medical centers over 1 yr | SCID-5-RV, YBOCS | clinicians, interviews | 12-mo OCD | | F = 40.3 | z = 5.20 | | | | | | p < 0.001 | | 0.7 | | |
| Incidence and Prevalence in sub-populations | | | | | | | | | | | | | | | | | | | | |
| Sulkowski  2011 | 358 | 17-27 | psych/edu STU (USA) | anonymous survey during class | OCI-R | SR | clinic OCS (OCI-R>27) | | F = 5.5  M = 5.9 | χ2 = 0.048 | | | | | | p = 0.83 | | 0.9 | | |
| Strehlau  2012 | 193F | 19-57 | homeless (CAN) | homeless outreach, drop-ins, and shelters | DSM-IV  interviews | clinicians, interviews | current OCD | | F = 19 | - | | | | | | - | | - | | |
|  |  |  |  |  |  |  | life OCD | | F = 62 |  |  |  |  |  |  |  |  |  |  |  |
| Desouky  2015 | 1024F | 15-17 | students  (SAU) | multistage cluster sampling | OCDS | SR | clinic OCS | | F = 23.1 | - | | | | | | - | | - | | |
| Jaisoorya 2017 | 4926 | 18 -25 | college  (IND) | college based sampling | CIS-R | SR | current OCD | | F = 3.2  M = 3.5 | χ2 = 0.20 | | | | | | p = 0.56 | | 0.9 | | |
|  |  |  |  |  |  |  | current subOCD | | F = 7.7  M = 9.9 | χ2 = 6.7539 | | | | | | p = 0.02 | | 0.8 | | |
| Ashraf  2017 | 377  (196) | 18-46 | college  (PAK) | college based sampling,  two universities | Padua | SR | current  subOCS | | F = 43.4  M = 50.3 | χ2 = 1.80 | | | | | | p > 0.10 | | 0.8 | | |
|  |  |  |  |  |  |  | current  contam | | F = 48.5  M =38.7 | χ2 = 3.67 | | | | | | p < 0.05 | | 1.5 | | |
|  |  |  |  |  |  |  | current  harm | | F = 50.5  M =39.8 | χ2 = 4.37 | | | | | | p < 0.05 | | 1.6 | | |
|  |  |  |  |  |  |  | current  impulses | | F = 30.1  M = 63.0 | χ2 = 40.97 | | | | | | p < 0.001 | | 0.3 | | |
| Asghar  2020 | 1172  (492) | 18+ | med students  (PAK) | convenience sampling, med schools | YBOCS | SR | current OCD | | F = 7.7  M =11.9 | z = 1.7 | | | | | | P = 0.086 | | 0.7 | | |
| Batchelder 2021 | 290M | 18–65 | Gay and bisexual males, history of sex abuse  (USA) | HIV prevention and mental health programs | MINI CTQ-SF | clinicians, interviews | current OCD | Bi-s =7.7  Gay = 11.9 | | | | χ2 = 0.49 | | | | | p < 0.05 | | G:Bi  1.46 | |
| Russell  2022 | 1.36 M | 20-40 | active military  (USA) | Defense Medical Epidemiological Database (1999–2018) | ICD-10 codes | health records | incidence (per 1,000) | F = 0.52  M = 0.30 | | | | χ² = 10 166 | | | | | p < 0.001 | | 1.73 | |
| Faruki  2023 | 319430 (198400) | 6–24 | suicidal inpatient/mood d/o | stratified probability sampling | ICD-10 codes | health records | current OCD | GD= 6.2  noGD = 2.9 | | | | [95% CI, 1.51–1.94], p<0.001 | | | | | | | GD:  1.71 | |
| Li  2023a | 1248  (640) | 18 + | Students  (CHN) | college based sampling,  three universities | SCL-90 | SR | current OCD | F: M OR = 0.75 | | | | [95%CI, 0.57–0.98], p < 0.05 | | | | | | | 0.75 | |
| Pei  2023 | 7493  (3791) | 12–18 | students  (CHN) | two-step school-based sampling | MINI-KID + DSM-IV | clinicians, interviews | current OCD | F = 10%  M = 8.1% | | | | [95%CI, 1.08–1.47], p = 0.005 | | | | | | | 1.26 | |
| Almadani  2024 | 371  (137) | 18+ | med students  (SAU) | school-based sampling | OCI-R | SR | current likely OCD | F = 43.8  M = 23.5 | | | | χ² = 16.2, p < 0.001 | | | | | | | 2.5 | |
| Barzallo-Farfán  2024 | 270 (145) | 18-30 | medical students, (PER) | convenience sampling, med schools | YBOCS, KNOW-P-COVID-19 | SR | current OCS | F = 15.9  M = 10.4 | | | | χ2 = 1.73 | | | | | p = 0.19 | | 1.61 | |
| Farahbakhsh  2024 | 1282 (693) | 20–60 | empl. med school, (IRN) | convenience sampling, med school | CIDI | trained prsnl | 12m OCD | F = 4.2  M = 3.6 | | | | χ² = 0.32 | | | | | p=0.568 | | 1.05 | |
| Nasirpour  2024 | 4453  (2813) | 18–65 | public employ.  (IRN) | Employees’ Health Cohort Study | CIDI-2.1,  ERI | Clinicians,  interviews | Current OCD | F = 2.9  M = 2.2 | | | | χ² = 2.09 | | | | | p = 0.15 | | 1.34 | |

Table A3.2. Gender differences in onset, progression, and contributing factors.

| **First Auth, Year** | **N**  **(F)** | | **Age mean**±**SD (range)** | | | **Pop** | | | **Meas.** | | | **Gender effects (F/M)** | | | | |
| --- | --- | --- | --- | --- | --- | --- | --- | --- | --- | --- | --- | --- | --- | --- | --- | --- |
|  |  |  |  |  |  |  |  |  |  |  |  | **Stats** | | | **Direction** | |
| Age of onset (AO) | | | | | | | | | | | | | | | | |
| Memis  2019 | | 74  (47) | | 15 ± 2  (<18)  31 ± 7  (18+) | | | OCD  (TUR) | | | Pts repts. | | | Retrospective. M:F: EO = 1.07, LO = 0.36, LO:EO OR = 0.34, χ^2^ = 4.8 p = 0.02 | | | (AO≥18) : (EO<18) OR: M < F |
| *children/adolescents* | | | | | | | | | | | | | | | | |
| Nakatani  2011 | 365  (151) | | 14 ± 3  (6-18) | | | OCD  (GBR) | | | parent/ pts repts | | | distress Sx, 2 subgr: median split with robustness check. %F: EO (AO ≤ 9) = 0.41 z-test p = 0.03, LO (13 < AO ≤ 17) = 0.42 z-test p = 0.02. | | | AO ≤ 17: M > F | |
| *adults* |  | |  | | |  | | |  | | |  | | |  | |
| Wang  2011 | 602  (263) | | 30 ± 11  (8-65) | | | OCD  (CHN) | | | 2 clinic interviews | | | distress Sx, 2 subgr: median split. %F: EO (AO ≤ 18) = 0.33, LO (AO > 18) = 0.53, t(2) =6.4 p = 0.042. | | | AO ≤ 18 M > F  AO >18 M ~ F | |
| De Luca  2011 | 196  (110) | | 39 ± 12  (18+) | | | OCD  (CAN) | | | SCID-I/P | | | NA, 2 subgr: mixture analysis. %F: EO (AO ≤ 15) = 0.54, LO (AO > 15) = 0.60, p = 0.47. | | | AO ≤ 15: M ~ F  AO > 15: M < F | |
| Narayanaswamy  2012 | 545  (213) | | 29 ± 11  (18+) | | | OCD  (IND) | | | parent/ pts repts | | | distss Sx. F-M d = 0.62 U = 23453 p <0.001. | | | AO  M < F | |
| Kichuk  2013 | 955  (555) | | 36 ± 13  (18 – 65) | | | OCD  (BRA) | | | DYBOCS  pts repts | | | distress Sx, by primary Sx. AO ~ Primary Sx x F: F(3,511) = 2.6 p = 0.051; Forbid thght M < F, contam/hoard M ~ F, Sym M > F. | | | forbid thght M < F, contam/hoard M ~ F, symm M > F | |
| Anholt  2014 | 377  (215) | | 36 ± 11  (18+) | | | OCD  (NOR) | | | SCID-I | | | DSM, 2 subgr: mixture analysis. %F: EO (AO < 20) = 0.59, LO (AO ≥ 20) = 0.53, χ^2^ =1.2 p = 0.28. | | | AO >,< 20: M ~ F | |
| Frydman  2014 | 983  (562) | | med = 32  (25 – 44) | | | OCD  (BRA) | | | SCID-I | | | distress Sx 3 subgr: *a priory*. %F: EO (AO ≤ 16) = 0.48, IO (16 < AO < 40) = 0.58, LO (AO ≥ 40) = 0.73, χ^2^ (2) =16.2 p <0.001. Risk for LO. LO~F: β = 0.5 [95CI%, 0.9 – 2.7], controlling for OCS, PTSD after age 40/no history of PTSD, pregn. | | | AO ≥ 40: M < F | |
| Albert  2015 | 483  (232) | | 34 ± 12  (18+) | | | OCD  (ITA) | | | clinic interview/parent repts | | | distress Sx, 3 subgr: mixture analysis. %F: EO (AO ≤ 9) = 0.44, IO (10 ≤ AO ≤ 23) = 0.46, LO (AO ≥ 24) = 0.58, χ^2^ =4.3 p = 0.10. DSM/DSM, 2 subgr: mixture analysis. %F: EO (AO ≤ 26) = 0.44, LO (AO ≥ 26) = 0.61, χ^2^ =9.3 p = 0.002 | | | AO < 23-26: M > F  AO ≥ 23-26: M < F | |
| Sharma  2015 | 802  (305) | | 29 ± 10  (N/A) | | | OCD  (IND) | | | medical records | | | NA, 3 subgr: *a priory*. %F: EO (AO < 18) = 0.27, IO (18 ≤ AO ≤ 39) = 0.42, LO (AO ≥ 40) = 0.74, χ^2^ =42 p < 0.001. F OR: IO vs. LO = 0.27 [95%CI, 0.13 – 0.56], controls: LSE/ aggres/ hoard; EO vs. LO = 0.21 [95%CI, 0.09 – 0.49], controls: LSE/aggres/sex/repeat/ socil phob/ psychos; EO vs IO = 0.58 [95%, 0.41 – 0.82], controls: for sex/repeat/tic/ hoard/ YBOCS. | | | AO ≤ 39 M > F  AO ≥ 40 M < F | |
| Dell’Osso  2016 | 431  (249) | | 43 ± 13  (18+) | | | OCD  (world) | | | SCID I, II | | | distress Sx, 3 subgr: *a priory*. %F: EO (AO ≤ 12) = 0.52 p, IO (13 ≤ AO ≤ 17) = 0.50, LO (AO ≥ 18) = 0.67, Fisher’s Exact test p =0.004. | | | AO ≥ 18: M < F | |
| Kang  2017 | 615  (217) | | 30 ± 11  19-65 | | | OCD, HC  (KOR) | | | family/pts repts | | | Retrospective. M:F: EO = 2.94, LO = 1.11, LO:EO M:F = 0.38 z = 5.6 p < 0.001 | | | (AO≥18) : (EO<18) OR:  M < F | |
| Grover  2018^s^ | 345  (149) | | 30 ± 11  (18+) | | | OCD  (IND) | | | YBOCS CL | | | NA, 2 Sx subgr: *a priory*. %F: EO (AO ≤ 18) = 0.25, LO (AO >18) = 0.50, χ^2^ =16.2 p < 0.001. By primary Sx. M OR EO-OCD: hoard/sym = 3.81 [95%CI, 1.17–12.45], contam = 0.06 [95%CI, <0.01,0.48], aggress/sex/relig = 1.47 [95%CI, 0.56–3.84]; M OR LO-OCD: hoard/ sym = 5.29 [95%CI, 2.78–10.09], contam = 0.24 [95%CI, 0.19–0.46], aggress/sex/relig = 2.48 [95%CI, 1.49–4.12]. | | | AO ≤ 18: M > F AO >18: M ~ F | |
| Girone  2024 | 284 (137) | | 39 ± 14  (18–69) | | | OCD (ITA) | | | SCID-5 YBOCS | | | Retrospective. M:F EO = 1.86 (% F=0.35), LO = 0.74 (% F=0.58), LO:EO M:F = 0.40; χ²(1) = 14.1, p < .001. | | | EO: M > F  AO: M < F | |
| Progression from onset to Dx | | | | | | | | | | | | | | | | |
| Coles  2011 | 199  (140) | | 34 ± 11  (18-64) | | | OCD  (USA) | | | Study-specific | | | Retrospective. Sx phase: F = 5.83±6.37, M = 3.88±5.09, t(182) = 1.97 p =0.05. ***F x onset – NA.*** | | | Sx phase  M ≤ F | |
| Torresan  2013 | 858 (504) | | 35 ± 12 (18-77) | | | OCD  (BRA) | | | YBOCS, DYBOCS | | | Cross-sectional. Worse ever Sx, age: M = 27.7 ± 11.6 F = 30.5 ± 12.4 p < 0.001, Help-seeking, age: M = 27.9 ± 11.6 F = 31.2 ± 12.5 p < 0.001. | | | Onset worse Sx/help  M < F | |
| Thompson  2020 | 954  (545) | | 35 ± 13  (18+ ) | | | OCD  (BRA) | | | YONHQ,  YBOCS,  DYBOCS | | | Retrospective. Sx phase, median: M = 5 F = 8 z = 4.89 p < 0.001; Poisson regr: M β = 0.16 [95%CI, 0.11-0.21] Wald = 45.9 p < 0.001, controlling for age, symptom type, symptom severity, agoraphobia, GAD, Tic, Family history. | | | Sx phase  M < F | |
| Ziegler  2021 | 100  (57) | | 40 ± 14  (18-82) | | | OCD  (DEU) | | | OCI-R, GAF, USS | | | Retrospective. Onset → Dx: short (< 7) F=21, M=11; long (≥7): F=33, M=29. F:M OR (long vs short) = 0.60 [95% CI 0.25–1.44] | | | Onset → Dx: M ~ F | |
| Puberty | | | | | | | | | | | | | | | | |
| Voltas  2014 | w1 1514  (794)  w2 562  (308)/  w3 242  (147) | | w1 10 ± 1  (8-12)/  w3 14 ±1  (11-15) | | | GEN,  (ESP) | | | LOI-CV,  YI-4, CDI,  SCARE | | | Longitudinal. Wave 1: order/check/pollution: F=7.33 ± 5.4 M = 6.76 ± 5.2 t = -2.08 p = 0.037; superstition/ mental comps: F = 2.31 ± 3.0 M = 2.65 ± 3.4 t = 1.99 p = 0.046. Progression. ***“all symptoms decreased over time with no differences between sexes” – stats NA.*** | | | subOCS ↓  from 10 to 14  M ~ F | |
| Castagnini  2016 | 6043  (2496) | | baseline  12 ± 3  (6-16) | | | Psych concerns  (DNK) | | | Dx codes | | | Medical records. OCD incidence in kids: IRR M/F = 1.1 [95%CI, 0.8-1.5], OCD in kids → need for treatment in adult: OCD RR = 5.4 [95%CI, 4.0-7.1] M/F RR= 0.7[95%CI, 0.4-1.2]; Dx kids →Dx in adults: OCD→OCD OR = 43.3 [95%CI, 19.6-95.6]; but note Phob/panic/anx → OCD OR = 6.6 [95%CI, 2.0 – 21.9] & Phob/panic/anx incidence in kids RR = 3.3 [95%CI 2.5-4.3] M/F RR = 0.5 [95%CI, 0.3-0.8]. | | | OCD kids → treatm in adults M ~ F Phob/panic/anx in kids M < F & →OCD in adults | |
| Barzilay  2019a | 6598  (3567) | | 16 ± 3  (11-21) | | | GEN  (USA) | | | GOASSESS, K-SADS,  C-GAS | | | Longitudinal. SubOCS, %F: At least 1 OCS = 0.59 no OCS = 0.51, F OR = 1.3 [95%CI, 1.2 – 1.5] Wald(1) = 25.97 p < 0.001; puberty (Pb) F x Pb M < F Wald(1) = 4.3 p = 0.038.  SubOCS subtypes, Bad thoughts: F OR=1.16 [95%CI, 1.03-1.31] W(1)=5.6 p=0.02, Pb OR = 1.14 [95%CI, 1.01-1.28] W(1)=4.5 p = 0.03, F x Pb W(1) = 0.09 p = 0.77; Repeat/Check: F OR = 1.27 [95%CI, 1.13-1.43] W(1) = 16.14 p <0.001, Pb OR = 1.2 [95%CI, 1.07-1.34] W(1) = 9.15 p = 0.002, F x Pb W(1) = 0.73 p = 0.39; Sym: F OR = 1.42 [95%CI, 1.24-1.63] W(1) = 25.46 p <0.001, Pb OR = 1.02 [95%CI, 0.9-1.17] W(1) = 0.12 p = 0.73, F x Pb W(1) = 1.49 p = 0.22; Clean/Contam: F OR = 1.41 [95%CI, 1.19-1.67] W(1) = 15.31 p < 0.001, Pb OR = 1.13 [95%CI, 0.96-1.33] W(1) = 2.0 p = 0.16, F x Pb W(1) = 0.40 p = 0.53; Hoard: F OR = 1.31 [95%CI, 1.06-1.62] W(1) = 6.17 p = 0.01, Pb OR = 1.18 [95%CI, 0.95-1.45] W(1) = 2.31 p = 013, F x Pb M ≤ F W(1) = 3.21p = 0.07. SubOCS in kids → lifetime Dx, OR: OCD = 11.7, Depress = 4.3, psychos = 3.3, Suicide Id = 4.4 ps < 0.001; ***OR x F – NA.*** | | | subOCS  F x puberty  M < F,  subOCS types  F x puberty  M ~ F | |
| de Barros  2021 | 1877  (822) | | 13 ± 2  (9–18) | | | GEN  (BRA) | | | DAWBA,  CBCL | | | Longitudinal, 3 years. Latent factor-based OCS ~ F: β=0.007 [95%CI, -0.081, 0.096]. | | | ΔOCS in teens  M ~ F | |
| Biosocial contributing factors and early markers  *biological* | | | | | | | | | | | | | | | | |
| Cheng  2014 | 566044  (292207) | | | | N/A  (14-49) | | | GEN  (TWN) | | | National database | | | Longitudinal. Season of birth (SOB) ~ OCD: Mar – Jul OR = 0.85[95%CI, 0.74-0.96]; Aug – Nov OR = 1.19 [95%CI, 1.05 – 1.36]; only in M: SOB ~ OCD x M p = 0.03 | | SOB → OCD  M > F |
| Fevang  2016 | 216  (111), 1767  (944) | | | | newborn/  2/5/11 yo | | | EP/ ELBW,  controls  (NOR) | | | Medical records, SCARED,  SOCD | | | Longitudinal. OCD severity ≥ 95% based on the control group EP/ ELBW OR: M = 2.5 [95%CI, 1.8 – 5.2], F = 2.7 [95%CI, 1.4 – 5.4] | | premature / low weight newborns  M ~ F |
| *resilience* |  | |  | | |  | | |  | | |  | | |  | |
| Hjemdal  2011 | 307  (140) | | 16 ± 1  (14–18) | | | school  (NOR) | | | OCI-R,  READ | | | Cross-sectional. Resilience ~ F: Pers. comp. = -0.19 p < 0.001, Soc. Comp. = 0.05 p > 0.05, Struc style = 0.06 p > 0.05, Soc. res. = 0.19 p < 0.001, Fam. coh. = 0.15 p < 0.01; Resilience ~ OCI-R: Pers. comp. = -0.22 p < 0.015, Soc. Comp. = -0.17 p < 0.015, Struc style = -0.08 p > 0.10, Soc. res. = -0.24 p < 0.015, Fam. = -0.21 p < 0.015; OCI-R ~ F = 0.15 p < 0.01. | | | pers comp M > F  soc comp/ struc style: M ~ F  soc res/fam M < F | |
| *family* | | | | | | | | | | | | | | | | |
| Taberner  2012 | 184F/  180mo/  174fa | | 22 ± 3  (18-35)/  50 ± 7  (39-74) | | | student/  parents  (ESP) | | | Padua,  DS | | | Cross-sectional. Disgust/OCS in parents ~ Contam daughter: hygiene fathers β = 0.23 p < 0.01; ordering mothers β = 0.21 p < 0.01. | | | father hygiene → contam daughters  mother ordering → contam daughters | |
| Hacıömeroğlu 2014 | 300  (147) | | 20 ± 2  (18+) | | | students  (TUR) | | | PADUA,  RAS,  s-EMBU | | | Cross-sectional. [Perceived mother overprotection (mOP) → OCS] is mediated by Resp: mOP →OCS β = 0.23 p < 0.01, mOP →Resp β = 0.35 p < 0.001, Resp →OCS β = 0.18 p < 0.01. | | | overprotect mother → OCS | |
| Liu  2017 | 2008  (880) | | 20 ± 1  (18+) | | | students  (CHN) | | | SCL-90, STAXI-2,  FACES-II | | | Cross-sectional. Family dynamic (FD), M < F: fam cohes (F(1903)=35.76 p <0.001), fam adapt (F(1928)=11.84 p=0.001), anger/trait (F(1984)=6.59 p = 0.01), anger-in (F(1939) = 5.89 p = 0.015); M > F: anger-out (F(1961) = 4.62 p = 0.032.  FD ~ OCS: fam cohes M = -0.16 F = -0.25, z = 2.09 p = 0.037, fam adap M = -0.19 F = -0.28, z = 2.12 p = 0.034.  [Anger →OCS] mediated by FD: Anger x fam cohes M = -0.36 t = -2.54 p <0.01; Anger-in x fam adapt M = -0.14 t = -1.10 p > 0.05, F = -0.30 t = -2.24 p < 0.05; Anger-in x fam cohes M = 0.27 t = 1.82 p > 0.05, F = 0.31 t = 1.91p > 0.05. | | | fam cohes/adapt  M < F,  fam cohes/adapt ↑↓ OCS M < F,  [anger-in →OCS] ↑↓ fam adap M < F,  [anger →OCS] ↑↓ fam cohes M > F | |
| Krebs  2018 | 2616→  1579 | | t1=15 → t2 =17 | | | Genesis 1219  (GBR) | | | *SCAS,* negative sanctions | | | Longitudinal. OCS (t2) ~ parental punitive discipline (t1), while controlling for OCS at t1 and stressful event at t2: maternal β = 0.02 p = 0.49, paternal β = 0.02 p = 0.59 | | | maternal/paternal punitive discipline ≠> OCS | |
| Someshwar  2020 | 509  (220) | | 32 ± 14  (18+) | | | fam. risk (IND) | | | MINI,  ACE-IQ | | | Retrospective. Earlier age of onset (EO) ~ adverse childhood experience (ACE) x M: ACE:M HR = 1.13 [95%CI, 1.07-1.20], ACE: F HR = 0.94 [95%CI, 0.86-1.03]. | | | familial risk + ACE → EO M > F | |
| *stressful life events (SLE): trauma, illness, etc.* | | | | | | | | | | | | | | | | |
| Real  2011 | 412  (174) | | 34 ± 12  (18+) | | | OCD  (ESP) | | | YBOCS,  SCID-II,  interview | | | Retrospective. SLE-preceding OCD: F OR = 1.5 [95%CI, 1.01-2.3]; F OR = 1.2 [95%CI, 0.74 – 2.07], while controlling for age, YBOCS, complicated birth, age of onset, contam, fam history. | | | SLE-preceding OCD  M ≤ F | |
| Rosso  2012 | 329  (160) | | 35 ± 12  (18+) | | | OCD  (ITA) | | | YBOCS  SCID-I, II | | | Retrospective. SLE-preceding OCD: F OR = 2.2 [95%CI, 1.4–3.4]. | | | SLE-preceding OCD  M < F | |
| Peles  2014 | 116F | | 38 ± 10  (NA) | | | sex abused  (ISR) | | | LEIQ,  YBOCS | | | Cross-sectional. Prevalence of OCD in sex abuse victims: frm opioid use = 0.66, no opioid use =0.30, OR = 4.47 [95%CI, 2.0–10.0]. | | | F:  sex abuse + opioid use ↑ OCD | |
| Ishikawa  2015 | 148F | | 19 ± 2  (18–28) | | | students  (JPN) | | | MCR,  unwanted sexual exp. recall task | | | Symptom provocation. Mental Contam (dirtiness, wash urge, internal/external neg feel) ↑ over Time, T, in response to Event, E, (verbal/visual assault, forcible touching, (attempted) rape). T: F(4,141) = 22.08 p < 0.001, E: F(12,429) = 1.96 p < 0.05, E x T: F(12,429) = 3.03 p < 0.001; dirtiness↑ T: F(1,144) = 50.13 p < 0.001 η^2^ = 0.26, E: F(12,429) = 1.96 p < 0.05 η^2^ = 0.07, E x T: F(12,429) = 3.03 p < 0.001 η^2^ =0.10; urge to wash↑ E x T: F(3,144) = 2.79 p < 0.03 η^2^ = 0.07, Δ urge to wash↑~ (attempted) rape p = 0.02; internal neg feel↑ T: F(1,144) = 60.42 p < 0.001 η^2^ = 0.30; external neg feel↑ T: F(1,144) = 49.12 p < 0.001 η^2^ = 0.26. | | | F:  unwanted sexual exp recall ~ contam ↑ | |
| Vidal-Ribas  2015 | 22084  (12524) | | 34 ± 8  (28-54) | | | twin registry  (SWE) | | | LSC-R, study-spec OCS | | | Cross-sectional. Prevalence M-F, in monozygotic twins, ps<0.00001: Abuse/ family disruption d = -0.14, Loss d = -0.12, Sex abuse d = -0.56, Non-sexual assault d = 0.40. | | | assault  sexual M < F  non-sexual M > F | |
| Waller  2014 | 80F | | 24 ± 10  (15-60) | | | students  (AUS) | | | non-cons. “dirty kiss” task, OCI-R; DASS, MCR | | | Symptom provocation. "dirty-kiss" perpetrator role → feeling of dirtiness, urge to wash, negative emotions: ƞ>0.47, p <0.001  Symptom mitigation. physical washing < mental washing: d >= 1.5 p <0.001. | | | F: non-consensual kiss →mental contam  physical wash < mental wash | |
| Ashraf  2017 (A3.1) | 377 (196) | | 27 ± 6 (18-46) | | | GEN  PAK | | | Padua, PSS | | | Cross-sectional. In high stress F:M OR = 0.33 [95%CI, 0.18-0.63]. High stress in OCD F:M OR = 0.39 [95%CI, 0.21-0.75]. | | | OCS↑↑stress: M > F | |
| Taghva  2017 | 215M | | 52 ± 8 (29–92) | | | veterans/ancle inj  (IRN) | | | SCL-90-R | | | Cross-sectional. <rate> OCS ~ Disability rate: “< 30%” = 1.4± 0.7, “≥30%” = 1.1±0.7 p =0.027; OCS ~ limb disability: “No” = 1.0 ± 0.7, “Yes” = 1.2 ± 0.6 p= 0.005; OCS ~ hospitalization due to injury: “No” = 1.3 ± 0.7, “Yes” = 1.5 ± 0.7 p= 0.05; OCS ~ hospitalization due other reasons: “No” = 1.3 ± 0.7, “Yes” = 1.6 ± 0.6 p= 0.044. | | | M:  foot injury ↑↑ OCS  disabil rate ↑↓ OCS | |
| Kennedy  2017 | 60M | | med=20  18 - 40 | | | students  (AUS) | | | non-cons. “dirty kiss” task, study-spec | | | Symptom provocation. Non-consensual > consensual: mental contam ↑ d > 0.53, p <0.001; choose a cleansing product as a gift↑ OR = 4.5 p = 0.024 | | | M: non-consensual kiss →mental cont.→ cleansing | |
| Barzilay  2019b | 7054  (3809) | | 16 ± 3  (11-21) | | | GEN  (USA) | | | DSM, interview | | | Retrospective. OCS ~ Trauma exposure x F: Wald (1) = 7.93 p = 0.005 F > M; OCS ~ non-assault trauma exposure x F: Wald(1) = 5.20 p = 0.023 F > M; OCS ~ assault trauma exposure x F: p > 0.05 F ~ M | | | trauma exposure  non-assault M < F  assault M ~ F | |
| An  2019 | 3160F | | 18-74 | | | GEN  (KOR) | | | K-CIDI | | | Lifetime prevalence OCD IPV OR. Physical IPV = 8 [95%CI, 1.6-26.3], Sexual IPV = 27.8 [95%CI, 5.0-107.3, any IPV = 10.6 [2.7-31.3]- the highest among psychiatric conditions. | | | F: IPV ↑↑ OCD | |
| Destrée  2020 | 954  (548) | | 34 ± 13  (18+) | | | OCD  (BRA) | | | YBOCS, DYBOCS  YONHQ | | | Retrospective. SLE prevalence: Death SLE F-M d = 0.054 p = 0.094, R/ship SLE d = 0.001 p =0.99, illness SLE d = 0.02 p = 0.53; speed subclinic →clinic OCD: Death SLE x M β = -0.31 p < 0.001; speed subclinic →clinic OCD, by Sx dimensions: Sex/Relig ~ M β = 0.90 p =0.03, Sex/Relig ~ R/ship SLE x M β = -0.49 p = 0.069. | | | OCD ~ Death SLE  M > F | |
| Krause  2021 | 149F | | 23±5 (19–46) | | | students (CAN) | | | sexual-harassment imagery task | | | Symptom provocation. Sexual-harassment role → mental contam η² ≈0 .40–0.47 p < 0.001, disgust η² ≈ 0.30–0.40 p < 0.001, distress η² ≈0.20–0.30; p < .001, Washing urge η² ≈0 .18–0.25; p <0 .001, avoidance urge η² ≈ .20; p < 0.001, shame/guilt/fear η² ≈0 .25–0.35 p < 0.001. | | | F: harassment imagery → mental contamination | |
| Larsen  2021 | 1167043  (568313) | | 6 - 28 | | | GEN  (DEN) | | | Medical records  ICD-8/10 | | | Longitudinal national cohort (1989–2010 births, follow-up to 2016), Risk factors (OCD HR): Premature birth (<33w): F = 1.46 [95%CI, 1.15–1.83] M = 1.12 [95%CI, 0.85–1.48]; Low birth weight (<1500 g): F = 1.32 [95%CI, 1.01–1.74] M = 1.08 [95%CI, 0.80–1.46]; Multiple birth F = 1.51 [95%CI, 1.29–1.77] M = 1.26 [95%CI, 1.04–1.52]; Congenital malformations F = 1.72 [95%CI, 1.10–2.69] M = 1.40 [95%CI, 0.90–2.18]; Ante genitourinary infection: F = 1.39 [95%CI, 1.16–1.67] M = 1.20 [95%CI, 1.01–1.43]; maternal age >35 y: F = 1.24 [95%CI, 1.09–1.41] M = 1.10 [95%CI, 0.95–1.26]; paternal age > 40: F = 1.22 [95%CI, 1.08–1.39] M = 1.13 [95%CI, 0.97–1.31]. | | | M ≤ F: Ante/PP complic. ↑↑ OCD | |
| Samuels  2021 | 2117  (1137) | | 46 ± 16  (18–89) | | | GEN  (USA) | | | DYBOCS,  OCI-R,  spec phob | | | Cross-sectional + retrospective. Prevalence, F:M OR: contam obses = 1.70, p <0 .001, adj for age, edu, resid = 1.23 [95%CI, 0.98–1.54]; contam phob = 2.41, p < 0.001. | | | contam obses: M ~ F  contam phob: M < F | |
| Forresi  2022 | 1676 (959) | | 47 ± 9  (25–65) | | | occup. stress clinic  (ITA) | | | SCL-90  work stress CL | | | Cross-sectional: OCS severity: F = 2.21 ± 0.93 M = 1.96 ± 0.91, t = –4.94, Cohen’s d = 0.24, p < 0.001. | | | bulling ↑↑ OCS:  M < F | |
| Tang  2022 | 148F | | 21 ± 2  (17–27) | | | students  (CHN) | | | non-cons. “dirty kiss” task, MCR-C, PI-C-C | | | Symptom provocation. Non-consensual > consensual: mental contamination ↑ d ≈ 1.00, p < .001 (ANOVA: F(1,148)=37.86, η²=.20). Immoral > moral: mental contamination ↑ d ≈ 0.35, p = .010 (F(1,148)=4.56, η²=.03). Hygiene behaviors during the break: Fisher’s exact p = 0.819; contamination ~ washing: logistic regressions p = 0.50–0.95. | | | F: non-consensual kiss/ immoral →mental cont. | |
| Shafighi  2023 | 300  (166) | | 42 ± 9  (18–65) | | | post COVID 3–18 mo  (IRN) | | | OCI-R, DASS-21, PCL-5, PSQI | | | Prevalence. %OCS: F = 0.77 M = 0.64 F:M OR = 1.79 [95% CI, 1.13–2.84] p = 0.01. | | | OCS: M < F | |
| Jin  2024 | 27671  (13966) | | 20 ± 2  (15 – 26) | | | GEN +CT (CHN) | | | DOCS-SF, CTQ-SF | | | Cross-sectional: Within child trauma (CT): F:M OR = 1.47 [95%CI, 1.39 – 1.55]; Cohen’s d = 0.27; p < 0.001 | | | Child trauma↑↑ OCD: M < F | |
| Pol-Fuster  2025 | 16030 twins  (8 712) | | 15 → 24 | | | twin registry  (SWE) | | | OCI-R, BOCS, OBVQ-R, med records | | | Longitudinal twin registry study. %OCD: F = 0.01 M = 0.007. F:M OR = 1.43 [0.98–2.09], p ≈ 0.07; Bulling →OCD: F OR = 1.31 [1.17–1.46]; M OR = 1.32 [1.15–1.52] | | | OCD: M ~ F  Bulling →OCD:  M ~ F | |
| *COVID-19 pandemic* | | | | | | | | | | | | | | | | |
| Littman  2020 | 65  (38) | | NA  (18+) | | | OCD (ISR) | | | study-spec | | | Retrospective: COVID-19. ΔOCS > 0: 12F/9M, ΔOCS = 0: 14F/7M, ΔOCS < 0: 12F/11M, Fisher’s Exact test p = 0.60 | | | Δ OCS  M ~ F | |
| Mazza  2020 | 402  (137) | | 58 ± 13  (18-87) | | | COVID  patients  (ITA) | | | PCL-5,  OCI | | | Cross-sectional: COVID-19. OCS F-M d = 0.43 t = 3.64 p < 0.001; OCI ~ F, while controlling for prior psych Dx, hospitalization, β = 0.20 F = 9.29 p = 0.0025. Inflam MLR M – F d = 0.40 t = 3.32 p < 0.001; OCI ~ MLR r = 0.12 p = 0.04; OCI ~ MLR x F- NA. | | | post COVID inflam  M > F; post COVID inflame ~ OCI;  OCI M < F | |
| Ji  2020 | 13478  (8816) | | 21 ±3  (17-50) | | | student  (CHN) | | | YBOCS | | | Longitudinal. YBOCS ~ M: wave 1 β = 0.5 p < 0.001, wave 2 β = 0.67 p <0.001, wave 3 β =0.38 p < 0.001. | | | YBOCS lockdowns  M > F | |
| McKune  2021 | 280  (145) | | NA  (8 – 18) | | | K-12  (USA) | | | KAP,  study-spec | | | Cross-sectional: COVID-19. Prevalence of Risk for OCD M = 0.26, F = 0.38, F OR=1.76 [95%CI, 1.03-3.03]. | | | risk for OCD  M < F | |
| AlHusseini 2021 \| 2186 (1322) \| NA (18+ ) \| GEN (SAU) \| OCI-R \| Cross-sectional: COVID-19. Prevalence of OCD symptoms during COVID pandemic M = 0.67, F = 0.59, M OR= 1.42 [95%CI, 1.19 – 1.70]. \| likely OCD M > F; Ahmed 2021a \| 524 (301) \| NA (18+ ) \| GEN (EGY) \| YBOCS \| Cross-sectional: COVID-19. Prevalence of OCD symptoms, elevated: health care workers (HCW) = 0.29, non HCW = 0.28; moderate/severe: HCW = 0.03, non HCW = 0.065, p = 0.047; OCD ~ F β = 1.16 [95%CI, 0.06-2.27]. \| elevated OCS M < F; Ahmed 2021b \| 182 (98) \| 47 ± 17 (18–80) \| COVID patients (EGY) \| PSQI, PCL-5, SCL-90 OCS \| Cross-sectional: COVID-19. Prevalence of OCD symptoms during COVID pandemic F = 0.204 M = 0.19, F:M OR = 1.134 [95%CI, 0.44 – 2.94], p = 0.79 \| likely OCD M < F; Alateeq 2021 \| 2909 (2149) \| 30 ± 11 (18+ ) \| GEN (SAU) \| BOCS, PSS \| Cross-sectional: COVID-19. Incidence of OCD symptoms during COVID pandemic. Obs: F = 0.603 M = 0.569 F:M OR = 0.87 [95%CI, 0.44 – 2.94], p = 0.76. Comps: F = 0.47, M = 0.44, F:M OR = 1.11 [95% CI, 0.92 – 1.34], p = 0.28. \| new OCD M ~ F; Carmi 2021 \| 113 (57) → 90 (43) \| 34± 11 (8–73) \| OCD (ISR) \| CGI-I, COVID obs \| Longitudinal: worsening of OCS 2 &6 mo after COVID-19 lockdown. Δ OCS at 2 mo: F = 56.1%; M = 43.9%; χ²(1) = 1.54, p = 0.21; Δ OCS at 6 mo: F = 39.5%; M = 36.2%; χ²(1) = 0.08, p = 0.77; COVID-19 as a new obsession (2-mo): F = 42.1%; M = 39.2%; χ²(1) = 0.08, p = 0.77 \| COVID↑↑Δ OCS: M ~ F; Di Consiglio 2021 \| 454 (386) \| 25 ± 4 (19–54) \| student (ITA) \| SCL-90-R \| Cross-sectional, multi-wave study. OCS severity F: 1–10 w of lockdown = 53.74 ± 11.75; 3–13 mo pre = 48.70 ± 11.62; 5–7 mo post = 47.97 ± 10.01; 9–12 mo post = 49.31 ± 10.65; η² = 0.027, p = 0.015. M: 1–10 w of lockdown = 50.12 ± 10.88; 3–13 mo pre = 47.80 ± 10.91; 5–7 mo post = 48.66 ± 10.45; 9–12 mo post = 48.89 ± 10.24; η² = 0.032, p = 0.098. \| COVID↑↑Δ OCS: M < F; Ferreira 2021 \| 420 (203) \| med = 53 (IQR=23) \| phys (PRT) \| OCI-R \| Cross-sectional: COVID-19. Severe OCS: F = 18.2% vs M = 17.8% χ2(1) = 0.01 p=0.92. OCS: M = 18.7 ± 12.4 F = 17.6 ± 11.3, Cohen’s d = 0.09, p = 0.36; \| COVID↑↑ OCS: M ~ F; Fontenelle 2021 \| 829 (436) \| 39 ± 13 (18-82) \| MTurk (USA) \| DOCS \| Cross-sectional: COVID-19 (current + retrospect). OCS current (controlling for pre): β_M_= –0.17 (0.07), p = 0.03; β_M_ = –1.15, p = 0.012 \| COVID↑↑ OCS: M < F; Højgaard 2021 \| 201 (132) \| 40 ± 2 (18+ ) \| GEN (DEN) \| YBOCS SR \| Cross-sectional: retrospective Δ in OCS dues to COVID-19. F = 2.76, M = 1.61, Cohen’s d ≈ 0.37 t(199) = 2.55, p = 0.012 \| COVID↑↑Δ OCS: M < F; Jelinek 2021 \| 179 (135) \| 40 ± 12 (18-80) \| OCD (GER) \| YBOCS \| Longitudinal: ΔOCS following lockdown 3-mo FU. M = –2.57 p = 0.02, F = –0.17, p = 0.03; Cohen’s d = 0.34 p = 0.02. \| ↓OCS: M > F; Othman 2021 \| 386 (293) \| 31 ± 11 (18+ ) \| GEN (LBN) \| YBOCS \| Cross-sectional: COVID-19. OCS F-M Cohen’s d = 1.26 t = 5.23 p < 0.001; hygienic prevention practices F-M Cohen’s d = 0.49 t = 2.03 p < 0.05. \| OCS/hygienic M < F; Samuels 2021 \| 2117 (1137) \| 46 ± 16 (18 - 89) \| GEN (USA) \| CIS, DYBOCS OCI-R \| Longitudinal: pre-during COVID-19. COVID prevalence of contam obs M: F OR = 1.7 [95%CI, 1.39-2.1]; prevalence of contam phobias M:F OR = 1.7 [95%CI, 1.39-2.1]; pre-post Δ OCI-R from 0-9 to 21+ M:F OR = 1.79 [95%CI, 0.73-4.37]; pre-post Δ OCI-R exclude contam from 0-9 to 16+ M:F OR = 1.64 [95%CI, 0.72-3.77]. \| contam M > F Δ OCS M ~ F; Liao 2021 \| 110 (53) \| 30 ± 9 (18-60) \| OCD (CHN) \| OCI-R \| Longitudinal: pre-during -1 year f/u COVID-19: Δ OCI-R ~ COVID x F = 1.51, p = 0.034 \| Δ OCS M < F; Meda 2021 \| 358 (286) \| 21 ± 2 (18-30) \| student (ITA) \| OCI-R \| Longitudinal: pre-during-post COVID-19 lockdown. Δ OCI-R ~ lockdown x F = -3.3 p = 0.41 \| Δ OCS M ~ F; Arora 2022 \| 250 (198) \| 21 ± 1.3 (18–25) \| med student (IND) \| YBOCS, FCV-19S \| Cross-sectional: COVID-19. Prevalence of OCD symptoms during COVID pandemic F = 0.14, M = 0.174, F:M OR = 0.76 [95% CI, 0.34 – 1.72], p = 0.13. \| OCS M ~ F; Benatti 2022 \| 116 (53) \| 40 ± 13 (18+) \| OCD (ITA) \| clinical interview \| Cross-sectional: worsening of OCS during COVID-19. Obsessions: Past recurring F = 0.46, M = 0.79, p = 0.033; New F = 0.46, M = 0.32, p = 0.341. Compulsions: Past recurring F = 0.42, M = 0.63, p = 0.196; New F = 0.42, M = 0.26, p = 0.320. \| Δ past obs: M < F; Δ new obs/comp: M ~ F:; Brooks 2022 \| 594 (385) \| 35 ± 15 (18–78) \| GEN (USA) \| SCARED-A \| Cross-sectional: OCS severity during COVID-19. OCS: F = 7.84 ± 3.46; M = 6.91 ± 3.21; d = 0.28, p = 0.04. OCS ~ distress, r: F = 0.42, p < 0.001; M = 0.37, p < 0.001; F vs M z = 0.71, p = 0.48. OCS ~ depr, r: F = 0.44, p < 0.001; M = 0.41, p < 0.001; F vs M z = 0.36, p = 0.72. OCS ~ anx, r: F = 0.77, p < 0.001; M = 0.75, p < 0.001; F vs M z = 0.28, p = 0.78. OCS ~ Hopelessness, r: F = 0.38, p < 0.001; M = 0.35, p < 0.001; F vs M z = 0.34, p = 0.73. \| OCS: M < F OCS ~ distress, depr, anx, hopeless: M ~ F; Pacitti 2022 \| 20241 (16309) \| 39 ± 13 (18–65) \| GEN (ITA) \| DOCS \| Cross-sectional: clinically sign. OCS (DOCS ≥ 30) during COVID-19. F = 52% M = 39%, F:M OR = 1.69 [95%CI, 1.55–1.84] p < 0.001. \| High OCS: M < F; Radhakrishnan 2022 \| ~71% of nonfed EDs \| 0-17 \| help-seeking (USA) \| medical records \| 3 cross-sectional waves: 2020, 2021, 2022. OCD-related visit ratio (ref = 2019): **2020 (Mar-Jan)**: F = 1.81 [95%CI, 1.67–1.96] M = 1.37 [95%CI, 1.26–1.49] F:M OR = 1.20 [95%CI, 1.10–1.31) ↑ (p < .001]; **2021 (Jan)**: F = 1.39 [95%CI, 1.30–1.49]; M = 1.03 [95%CI, 0.96–1.11]; F:M OR = 1.20 [95%CI, 1.12–1.29] p < .001; **2022 (Jan):** F = 1.24 [95%CI, 0.94–1.63] M = 0.76 [95%CI, 0.58–1.00] F:M OR = 1.31 [95%CI, 0.98–1.74] ns. \| COVID ↑↑OCD: M < F; Taner 2022 \| 7880 (3271) \| med 10 → 11 \| help-seeking (TUR) \| medical records \| 2 cross-sectional waves: (2019, 2020). Relative prevalence post vs pre OR F = 1.19 [95% CI, 0.94–1.50]; M = 1.99 [95% CI, 1.57–2.52]; F:M OR = 0.60 [95% CI, 0.43–0.83], p < .01. \| OCD: M > F; Dehghani 2023 \| 320 (208) \| 34 ± 9 (18–65) \| help-seeking (IRN) \| FCV-19S, PI-WSUR \| Cross-sectional: OCS severity during COVID-19. Fear of COVID-19: F = 17.29 ± 5.81; M = 15.92 ± 5.60; d = 0.24, p = 0.002. Washing compulsions: F = 6.75 ± 4.24; M = 5.91 ± 3.89; d = 0.21, p = 0.03. Impulses to harm self/others: F = 2.41 ± 1.09; M = 3.18 ± 1.16; d = 0.69, p = 0.004. Other OCD dimensions: ns (p > 0.05). \| Imp. to harm: M > F Wash: M < F; Jensen 2023 \| 186824 (115831) \| 34 ± 9 (18–65) \| GEN (NOR) \| ICPC-2 codes \| Longitudinal; pre-pandemic vs pandemic help seeking, Δ consults rates, %. M = 0 F = 2, Cohen’s d = 0.02 p > 0.99 \| Δ consults rates: M ~ F; Audet 2024 \| 653 (343) \| 43 ± 12 (18+ ) \| GEN (CAN) \| VOCI-C, CSS, FSQ-MV \| Cross-sectional: OCS severity during COVID-19. Severity of contamination symptoms during COVID pandemic: F > M d = 0.54, p < 0.001 (mean & SD not reported); Feared self-perceptions → Contamination: F = 0.49 p < 0.001; M = 0.47 p < 0.001, F vs M ΔCFI < 0.01, p > 0.05; COVID stress → Contamination: F = 0.37, p < 0.001; M = 0.35, p < 0.001; F vs M ΔCFI < 0.01, p > 0.05. \| Contm: M < F Fear/stress → contm M ~ F; Gümüşsoy, 2024 \| 726F \| 25 ± 1 (20–76) \| GEN (TUR) \| Fear of COVID-19 scale; BSI \| Cross-sectional: OCS severity during COVID-19, Pearlson r. Fear of COVID-19 ~ OCS = 0.43 p < 0.001 \| F: Fear of COVID-19 ↑↑ OCS; Köktaş 2024 \| 528 (329) \| 21 ± 2 (18–25) \| STU (TUR) \| VOCI \| Cross-sectional: OCS severity during COVID-19. Contam: F = 14.64 ± 12.05; M = 14.62 ± 9.51; d = 0.00, p < 0.05; Controlling: F = 8.31 ± 6.31; M = 7.05 ± 4.74; d = 0.22, p < 0.05; Obsess: F = 16.64 ± 11.85; M = 14.88 ± 9.49; d = 0.16, p = 0.003; Accumul.: F = 9.30 ± 7.31; M = 8.41 ± 5.85; d = 0.13, p = 0.003; Just-feeling-right: F = 16.29 ± 12.06; M = 13.45 ± 8.93; d = 0.27, p < 0.05; Indecision: F = 8.76 ± 6.23; M = 7.11 ± 4.86; d = 0.29, p < 0.05. \| OCS: M < F; Li 2024 \| 4246 (2978) \| 38 ± 9 (18+ ) \| health care (CHN) \| SCL-90-R, online \| Cross-sectional: provisional OCD: F:M OR = 1.69 [95%CI,1.19–2.40]. \| OCD in health care: M < F; Meza 2024 \| 75 (55) → 64 (54) \| 27 ± 10 (18–63) \| students (USA) \| OCI-R, DOCS, FOCI-B \| Longitudinal; pre-pandemic vs pandemic: time x gender, η²: OCI-R = 0.001 p = 0.717; DOCS = 0.001 p = 0.687; FOCI-B= 0.023 p = 0.120. \| Δ OCS: M ~ F; Pellegrini 2024 \| 20720 (14720) \| 40 ± 14 (18–80) \| GEN (ITA) \| OCI-R \| Cross-sectional: provisional OCD (OCI-R ≥ 21): F = 12.3 %, M = 8.8 %; F:M OR = 1.45 (1.30–1.63), p < .001. New OCD during COVID: F:M OR = 1.35 [1.21–1.51], p < .001 \| High OCS: M < F New OCD: M < F; Silverman 2024 \| 524 (370) \| 20 ± 1 (18–25) \| Students (USA) \| DOCS \| Cross-sectional cohorts across three waves (2016–2021). Severity /Δ Severity (3 × 2 × 4 ANOVA), η²: OCS × F = 0.002 p = 0.177 (F ≥ M), ΔOCS × F = 0.006 p = 0.053 (F ≥ M), contam × F = 0.002 p = 0.252, Δcontam × F = 0.008 p = 0.118 (F ≥ M), unaccep thghts × F = 0.004 p = 0.134 (F ≥ M); Δunacc.thghts × F = 0.009 p = 0.085, η² = (F ≥ M), symm × F < 0.001 p = 0.501, Δsymm × F = 0.006 p = 0.224, harm × F = 0.001 p = 0.441, Δharm × F = 0.004 p = 0.308. \| COVID↑↑ ΔOCS: F ≥ M; Triantafillou 2024 \| 130 (73) \| 49 ± 18 (18–85) \| GEN (GRC) \| DOCS (C1) \| Longitudinal; 2020, 2022: DOCS 2022 x F = 1.19 ± 0.50, β_std_ = –0.13, p = .019 \| COVID OCS; M < F; Bostan 2025 \| 200 (134) \| 33 ± 11 (18–65) \| OCD, HC (TUR) \| YBOCS, PATCQ, CCPMS \| Cross-sectional, case–control design, retrospective pre–during COVID-19. OCS F = 27.16 ± 8.41; M = 23.91 ± 7.63; d = 0.41, p = 0.04. Δ OCS: F = 3.21 ± 4.11; M = 2.94 ± 4.02; d = 0.07, p = 0.69. Avoidance behavior: F = 10.42 ± 3.13; M = 9.91 ± 2.84; d = 0.17, p = 0.33. Compliance: F = 37.29 ± 5.81; M = 36.02 ± 5.34; d = 0.23, p = 0.21. \| OCS: M < F Δ OCS, avoid, comply: M ~ F | | | | | | | | | | | | | | | | |
| Course | | | | | | | | | | | | | | | | |
| Marcks  2011 | 113  (65) | | 35 ± 12  (18+) | | | OCD  (USA) | | | SCID,  LIFE-UP | | | Longitudinal, 15 years. Remission: F HR = 1.16 [95%CI, 0.58-2.33], Wald = 0.19 p = 0.67, controlling for marital status, comorbid MDD, SSRI use. | | | remission  M ~ F | |
| Fineberg  2013 | 591  (299) | | wave 1:  19-20 | | | GEN  (CHE) | | | SCL-90-R,  SPIKE | | | Longitudinal, 30 years in 7 waves. Remission: M HR = 0.80 [95%CI, 0.50-1.29]; Remission age: M = 37 [95%CI, 34-40] F = 35 [95%CI, 33-38] p = 0.41. | | | remission  M ~ F | |
| Visser  2014 | 379  (214) | | 36 ± 11 (18-77) | | | OCD  (NLD) | | | YBOCS-SC,  EuroQol,  SCID-I/P | | | Retrospective, 2 years. %F: Chronic = 0.59 non-Chronic = 0.52, OR = 1.31 [95%CI, 0.86-1.98] p = 0.21, controlling for onset, severity, and type of OCD, illness burden, comorbid, anx, depress, Big-5, family history | | | chronic  M ~ F | |
| Mancebo  2014 | 60 (20) | | 11 ± 3  (6-18) | | | OCD (USA) | | | Y-LIFE,  CY-BOCS | | | Longitudinal, 3 years. %F: Chronic = 0.68, Remitting = 0.71 p = 0.84. | | | Course  M ~ F | |
| Goldberg  2015 | 449  (198) | | 34 ± 11  (17-73) | | | OCD  (ESP) | | | SCID-CV,  PSLE,  FHRDC | | | Prevalence. SLE: M < F χ^2^(1) = 9.9 p = 0.002; Familial load (FL): M ~ F χ^2^(1) = 1.97 p = 0.16. Retrospective, 5 years. Chronic vs. intermittent/episodic ~ SLE, FL, F: FL β = -0.18 p = 0.04 OR = 0.83 [95%CI, 0.70-0.98], FL x F β = 0.26 p = 0.015 OR = 1.30 [95%CI, 0.91-1.84], FL x SLE β = -0.37 p = 0.056 OR = 0.69 [95%CI, 0.47-1] | | | chronic ↑↓ FL  nonSLE: M > F  SLE M ~ F | |
| Geiger  2024 | 213  (114) | | 38 ± 11 (18–79) | | | OCD (NLD) | | | SCID, YBOCS | | | 6-year naturalistic follow-up: Trajectory, F:M OR: Episodic vs Chronic = 0.34 [95%CI, 0.12–0.96], p = 0.041; Remitted vs Chronic = 0.63 [95%CI, 0.16–2.41], p = 0.50; Episodic vs Remitted = 0.54 [95%CI, 0.14–2.13], p = 0.38. | | | chronic: M < F  remission: M ~ F  episodic: M > F | |
| Jagannathan  2024 | 3335  (1779) | | 15 ± 1  (14–18) | | | GEN  (USA) | | | RCADS-OCD, CASI, DTS | | | Longitudinal (4 waves, 2013–2017): Trajectories, M:F OR: High-but-Remitting : Low-and-Stable = 0.64 (β_M_ = −0.45 p = 0.01); Moderate-but-Escalating : Low-and-Stable = 0.86 (β = -0.15 p = 0.45). ΔOCS in Moderate-but-Escalating: wave 7 x F: β_M_ = 0.45, p < 0.05 (d ≈0.30). | | | high-but-Remit:M> F; Moder-excal:  ΔOCS: M < F | |

Table A3.3. OCD and reproductive events (Females only).

| **First Auth, Year** | **N** | **age**  **mean±SD**  **(range)** | **Pop** | **Meas.** | **Gender effects (F/M)** | |
| --- | --- | --- | --- | --- | --- | --- |
|  |  |  |  |  | **Stats** | **Direction** |
| Reproductive events | | | | | | |
| Moreira  2012 | 455 | 34 ± 13  (18+ ) | OCD  (BRA) | YBOCS,  DYBOCS,  BDI/BAI | Course, Menstr. phases. %: PMOCS-same = 0.503, PMOCS-worse = 0.497, p > 0.10; Aggres: PMOCS-same = 0.67, PMOCS-worse = 0.77 p = 0.02; Sex/relig: PMOCS-same = 0.45, PMOCS-worse = 0.62 p < 0.001; Misc: PMOCS-same = 0.86, PMOCS-worse = 0.92 p = 0.4, Sym/Order/Contam/Hoard: PMOCS-same ~ PMOCS-worse p > 0.27. AO Sx: PMOCS-same = 12.5 PMOCS-worse = 12.3 p = 0.72, AO OCD: PMOCS-same = 20.9 PMOCS-worse = 24.4 p = 0.001; Chronic/wax/wane: PMOCS-same ~ PMOCS-worse p > 0.59; BAI: PMOCS-same = 15.1 PMOCS-worse = 20.7 p < 0.001; BDI: PMOCS-same = 15.9 PMOCS-worse = 19.7 p = 0.001. Other factors. Hypothyr: PMOCS-same = 0.04, PMOCS-worse = 0.09 p = 0.04, Suicide Id: PMOCS-same = 0.28, PMOCS-worse = 0.47 p < 0.001; Suicide Att: PMOCS-same = 0.10, PMOCS-worse = 0.18 p =0.03; Mood stab meds hist: PMOCS-same = 0.07, PMOCS-worse = 0.15 p = 0.02; SSRI cur: PMOCS-same = 0.39, PMOCS-worse = 0.50 p = 0.02. | aggres/sex/relig,  AO-OCD, anx/depress,  suicide id/attm,  meds/hypothyr  PMOCS-worse > PMOCS-same |
| Guglielmi  2014 | 542 | 45 ± 15  (18+) | OCD  (USA,  NLD) | YBOCS, study-spec | Course. Onset, % of the sample: Menarche = 0.13, pregn = 0.051, PP = 0.047, menopause = 0.037. Worse, % of the sample: premenstr = 0.38, pregn = 0.33, PP = 0.47, menopause = 0.33; ↑Sx 1^st^ pregn ~↑Sx 2^nd^ pregn: OR = 10.82 [95%CI, 4.48-26.16]; ↑Sx 1^st^ PP ~↑Sx 2^nd^ PP: OR = 6.86 [95%CI, 3.27-14.36]. | menses, pregn, PP, menopause → Sx in F |
| Li  2017 | 213 | NA  (20+) | GEN  (CHN) | SCL-90 | Severity. OCS: Polycystic ovary syndrome (PCOS) =1.5 ± 0.4, no PCOS = 1.4 ± 0.3, d = 0.10 p = 0.037. | PCOS ↑↑ OCS |
| Mulligan  2019 | 40 | 21 ± 3  (18+) | GEN  (USA) | IDAS-II | Course, Menstr. phases. Check Sx: follicular (*fol*) = 6.1 ± 2.9, luteal (*lut*) = 6.2 ± 2.5 t(40) = -0.19 p = 0.85; Check Sx ↑↑ progesterone (P4): fol r(38) = 0.06 p = 0.72, lut r(38) = 0.14 p = 0.39; Check Sx ↑↑ estradiol (E2): fol r(38) = 0.03 p = 0.87, lut r(38) = -0.10 p = 0.53; Check Sx ↑↓ ΔERN: fol r(40) = -0.23 p = 0.20, lut r(40) = -0.47 p < 0.01. | F: check Sx  fol ~ lut  check Sx↑↓ ΔERN  fol < lut |
| Tay  2020 | 8670 | 23 ± 2  (19-26) | GEN  (AUS) | ACES,  SR Dx | Prevalence. Polycystic ovary syndrome (PCOS) ↑↑ OCD: OR = 1.8 [95%CI, 1.2-2.5]. | PCOS ↑↑ OCD |
| Song  2024 | 213 | (18+) | reprod. treat  (CHN) | SCL-90 | Risk (crossectional). Assisted reproductive technology (ART) ↑↑ OCD: OR = 1.8 [95%CI, 1.0–3.2] | ART↑↑ OCD |
| Prevalence Ante/PP | | | | | | |
| Chaudron  2010 | 24→16  25±6 | Ante 27w+  /PP 1m | GEN  (USA) | EPDS,  YBOCS | Prevalence. %OCD**:** Ante = 0.29 post = 0.25. Severity: Ante = 8.5±5.1 PP 1mo= 14.4±7.9 Cohen’s d = 0.89 p < 0.01. | OCS  ante < PP |
| Miller  2015 | 461  33 ± 5 | PP: 2w  6m | GEN  (USA) | PHQ9, STAI, YBOCS | Prevalence of Obs. %Contam: 2w = 0.16, 6m = 0.18, %aggres: 2w = 0.12, 6m = 0.17; %somat: 2w = 0.05, 6m = 0.05; Prevalence of Coms. %Check: 2w = 0.09, 6m = 0.12, %clean: 2w = 0.07, 6m = 0.08, %repeat: 2w = 0.04, 6m = 0.05.  Associate factors. %Married: OCD=0.79, subOCS=0.92, noOCS = 0.89%, p=0.039; # kids: OCD=1.3±0.5, subOCS = 1.7±0.8, noOCS = 1.5±0.8, p = 0.02; Race, Edu, Income, Age NS.  Comorbid. %Depress: OCD = 0.54, SubOCS = 0.24, noOCS = 0.11, p < 0.001; %Anx: OCD = 0.26, SubOCS = 0.08, noOCS = 0.02, p<0.001. | OCS by type  2 wks < 6 mo;  OCD↑↓married/# kids;  Depress/anx  OCD>subOCS>noOCD |
| Barrett 2016 | 94  30 ± 4 | Ante: 13-40w  PP: 1-9m | GEN  (GBR) | YBOCS, PTBC, RAS, RIQ | Severity. YBOCS: noPregn = 6.82, Ante = 9.04, PP = 11.0, p = 0.02. Prevalence. %Mild: noPregn = 0.30, Ante = 0.31%, PP = 0.34; %Moderate: noPregn = 0, Ante= 0.15, PP = 0.11; %Severe noPregn = 0, Ante = 0, PP = 0.06; PP vs noPregn: t(66) = −3.09, p = 0.003.  Distress. Ante = 44.35±27.9, PP = 65.1±25.3, t(40.07) = −3.34, p = 0.002. Intrusive Resp. Baby-related vs. Other: Ante t(51)= −1.51, p= 0.14, PP t(64) = −4.43, p < 0.001. | prevalence/severity  noPregn < Ante < PP;  distress Ante < PP  Baby-related Ante *~ PP*  PP: baby related > other |
| Fairbrother 2016 | 310  34 ± 6 | Ante: 28±6w  PP:0-3m | GEN  (CAN) | OCI-R | Prevalence. %OCD**:** Ante = 0.029 [95%CI, 0.01-0.048], PP = 0.036 [95%CI, 0.015-0.057], Peri = 0.017 [95%CI, 0.017-0.061]. Onset. %OCD**:** Ante= 0.01 [95%CI, 0.001-0.021], PP = 0.029 [95%CI, 0.01-0.048]. | prevalence/ onset  Ante ~ PP |
| Uguz  2019 | 1482 | PP: 1d | GEN  (TUR) | SCID-I | Prevalence**.** PP = 0.049, noPreg = 0.021, OR 2.31 [95%CI, 1.07-5.02], p = 0.03. | OC:  noPreg < PP |
| Osnes  2019 | 1563  31 ± 5 | Ante: 17w, PP: 8w | GEN, (NOR) | MINI,  SCL-A, | Prevalence. Ante w17 = 0.025, PP w8 = 0.042, p = 0.008, PP OR = 1.72 [95%CI, 1.15-2.57]. | OCD:  Ante < PP |
| Malemela  2019 | 206  28±6 | Ante  w4-40 | GEN  (ZAF) | OCI-R,  CAS, PI-WSUR,  EPDS,  PRAS | Prevalence. %OCD = 0.39. Sx severity. OCD-noOCD Cohen’s d p-value: Depress = 0.32 p = 0.027, Anger = 0.27 p = 0.061, Anx = 0.66 p = 0.001, Contam = 1.19 p = 0.001. OCI-R ~Associated factors. β, p-value: Age = 0.20 p < 0.01, gestation wks = 0.17 p <0.01, past complic = 0.17 p < 0.01, depress = 0.28 p < 0.01, Anger = 0.14 p = 0.10, anx = 0.24 p < 0.01. | Ante↑↑OCD;  Depress/anger/anx/contam  OCD > no OCD;  age/gestation time/previous complic/depress/anx ↑↑OCS |
| Shokuhi  2020 | 160  30 ± 5 | PP  NA | GEN  (IRN) | PDM | Construct. PP distress, % var expl, α: General distress (GD) = 0.37, α = 0.60, Obsessive-Compulsive (OC) = 0.14, α = 0.62. Concur valid: GD↑↑anx = 0.53, GD↑↑depress = 0.42, GD↑↑stress = 0.46; OC↑↑anx =0.42, OC↑↑depress=0.40, OC↑↑stress = 0.56. | PP distress:  General & OS |
| Burton  2021 | 5  26-40 | PP 5 yrs | OCD  (GBR) | semistruct  interviews | Common OC themes: (1) Hyper-responsibility, (2) Intrusive imagery, (3) Guilt/Shame, (4) Eventual Coping and acceptance |  |
| Beck  2022 | 43  NA | Ante/PP  NA | OCS  (USA) | Blogs, content analysis | Common OC themes: (1) Intrusive Thoughts Take Hold, (2) Desperate to Protect, (3) Paralyzed by Shame and Fear, (4) Journey to Recovery. | Ante/PP↑↑Intrusive thghts, shame/fear, protect |
| Kawoos  2022 | 95 | Ante/PP | GEN  (IND) | YBOCS | Prevalence. OCS severity post COVID: Ante = 8.38 ± 5.8 vs PP = 5.83 ± 5.8; t(93) = 2.14, p = 0.035 | OCS post COVID-19:  Ante > PP |
| Phillips  2022 | 232  18–47 | PP: 2-6m | GEN  (USA) | OCI-R, pOCDS, infant-feeding | Prevalence. %(OCI-R > 21)=0.14. PP OCI-R ~Associated factors. Breastfeeding problems β =0.3, p≈.05; breastfeeding ≥2 mo RR = 0.85, [95% CI, 0.64–1.11]; repeatedly washing baby dishes RR = 2.37 [95% CI, 1.55–3.64], counting/organizing stored milk RR = 2.71, [95% CI, 1.20–6.12], worry about accidentally harming baby RR = 2.01 [95% CI, 1.19–3.41]; depressive symptoms OR = 7.73, [95% CI, 3.39–17.62, state anxiety r = .45, p<.001. | PP OCD ↑↑infant-care and feeding obsessions/compulsions |
| Reagu  2022 | 261  20–45 | Ante: ~38w | GEN  (QAT) | YBOCS, study spec. | Prevalence. Severity. Ante = 3.4 ± 5.3, %YBOCS ≥ 8 = 0.21, %YBOCS ≥16 = 0.057, %YBOCS ≥25 = 1 case. Predictors of OCD: COVID impact on breastfeeding = 4.33, [95 %CI, 0.40–8.26] p = .031, r = .25, OCD symptoms ↑ vs pre-pandemic: 9 %→21 % | Ante OCD ↑↑ Covid |
| Vasileiou  2022 | 30  32 ± 5 | Ante:  23 ± 9w | OCS  (GRC) | MINI, YBOCS, OBQ, Leyton Trait Scale, HADS | Prevalence, %OCS. Aggres = 0.73, contam = 0.53; wash =0.50, check = 0.43. Associated factors. OCP traits (Leyton ≥ 12), OR: symm = 7.4 [95% CI, 1.2–44.9] p = .020, wash = 7.6 [95% CI 1.5–37.4] p = .011, check = 0.44 [95% CI, 0.11–1.75], contam = 1.7 [95% CI, 0.44–6.6], aggr = 1.0 [95% CI, 0.26–3.8]; OBQ (Responsibility/Threat), OR: contam = 4.4 [95% CI 1.1–17.9] p =0 .036, wash = 4.2 [1.0–17.3], p =0 .040; OBQ (Importance/Control), OR: aggres = 4.3 [1.1–17.3] p =0 .039, HADS-D (Depression), OR: check = 5.0 [95% CI, 1.2–20.5], p = 0.027; total OCS severity ↑ (β ≈ 0.44, p =0 .004); HADS-A (Anxiety), OR: contam = 6.6 [95% CI, 1.6–27.2] p = 0.011, wash = 5.1 [95% CI, 1.2–21.4] p =0 .025. | Ante ↑↑ aggres, contam, check  OCP, Resp, Control, Depr, Anx ↑↑ Ante OCS |
| Garcia  2023 | 10  30- 35 | PP | OCS  (USA) | semistruct  interviews | Common OC themes: (1) Obses. – safety, cleanliness (2) Compl. – checking, cleaning, reassurance/info-seeking (3) Emotion – fear, panic, anxiety, guilt/shame, suicidality, anger. | Ante/PP↑↑check, contam, reassure, emotion distress |
| Buhagiar 2024 | 350  30 ± 5 | Ante/PP < 6m | GEN  (MLT) | OCI-R | Prevalence. %OCD = 0.39 | Ante/PP↑↑Likely OCD: |
| Nakić Radoš  2025 | 397  32 ± 5 | Ante: 16-41w  PP: 6-12w | GEN  (HRV) | YBOCS  EPDS, ASI, IPIP-50, BRS | Prevalence. %OCD Ante = 0.15 PP = 0.15, Ante/PP = 0.098, Severity. Ante = 3.81±4.24 PP = 3.23±4.67 Cohen’s d = 0.14, p < 0.001. %OCD–PPD comorb. Ante = 0.33 OR = 2.6, p < .01, PP = 0.43 OR = 3.2, p < 0.001. Predictors of OCD, β. Ante: depr = 0.25 p <0.001, psych concerns = 0.21 p <0.001, neuro = 0.19, p = 0.001, resil = –0.13, p = 0.007. | Ante/PP↑↑Likely OCD  OCS: Ante > PP  Depr: Ante < PP |
| Pre-existing OCD: Changes Ante/PP | | | | | | |
| Uguz 2011 | 52  27 ± 5 | Ante: 30±8w | OCD  (TUR) | SCID-I, YBOCS | Progression. %ΔOCS: “ΔOCS > 0” = 0.32, “ΔOCS = 0” = 054, *“*ΔOCS < 0” = 0.14.  Associated factors, ΔOCS > 0 OR, p-value. History of MDD = 5.3 [95%CI, 1.4 – 20.4], p = 0.02, MDD at preg = 14.2 [95%CI, 1.5-133.8] p = 0.01, Anx Disd at Preg = 4.1 [95%CI, 1.2-14.1] p = 0.03. Obs, ΔOCS > 0 OR. Contam = 9.5 [95%CI, 1.1 – 79.8] p = 0.02, Sym = 4.0 [95%CI, 1.2-13.6] p = 0.036, Relig = 0.07 [95%CI, 0.004-1.3] p = 0.02; Comp, ΔOCS > 0 OR. Clean = 8.4 [95%CI, 1.0-70.8] p = 0.04, Order = 4.1 [95%CI, 1.2-14.1] p = 0.03, Aggres, Sexual, Hoard, somatic, check, repeat, count: OR $\cong$ 1 p > 0.05. | hist of MDD/ preg MDD & Anx ↑↑ ΔOCS > 0;  contam/clean/sym/order  ↑↑ ΔOCS > 0;  relig ↑↓ ΔOCS > 0 |
| Yakut 2019 | 37  28 ± 6 | PP: 1d, 6-8 w | OCD  (TUR) | YBOCS, COPE, TEMPS-A | Progression. PP obsess: *1 day* =13.0 ± 1.7, *6 wks* = 9.4 ± 4.0; PP compuls: 1 day = 12.0 ± 2.3 6w=8.5 ± 4.8p; PP total: 1d = 25.0 ± 3.8, 6w = 17.9 ± 8.7 p < 0.001; ΔOCS 1d →6w, %: “ΔOCS < -25%” = 0.51, “ΔOCS > 25%” = 0.08, “ΔOCS ~ 0” = 0.41.  Coping Strategies. COPE-humor: “ΔOCS < -25%” = 8.1 ± 2.9, “ΔOCS ≥ 0” = 6.1 ±2.4, p=0.029, COPE-suppression of competing activities/: “ΔOCS < -25%” = 11.0 ± 1.4, “ΔOCS ≥ 0” = 9.3±2.1, p=0.014; other COPE subscales p > 0.11.  Associated factors. Hyperthymic tempt: “ΔOCS < -25%” = 9.9 ± 4.5, “ΔOCS ≥ 0” = 5.5 ± 4.1, p = 0.004; Depressive, Cyclothymic, Irritable, and Anxious (p > 0.05) | PP Severity:  1 day > 6 wks  PP coping strategies  humor/focus ↑↓ OCS  hyperthymic ↑↓ OCS |
| Chatterje  2021 | 19  26 ± 6 | Ante/PP | OCD  (USA) | SCID-I/P, YBOCS, LIFE, PSR | Progression. Mean distress: Ante/PP = 3.12 ± 0.88; non-Ante/PP = 3.13 ± 0.89; Δ = −0.008 ± 0.76; p = 0.90. Clinical exacerbation (PSR ≥ 4): Ante/PP = 33.4%; non-Ante/PP = 32.0%; OR = 1.06 [0.80–1.42], p = 0.70. Remission (PSR ≤ 2): Ante/PP = 28.8%; non-Ante/PP = 30.5%; OR = 0.92 [0.67–1.27], p = 0.61. | OCS: Ante/PP ~ non- Ante/PP |
| Fairbrother  2024 | 97  31 ± 5 | Ante 37w/PP: 9w, 22w | OCD  (CAN) | SCID-5 | Timing of onset. By phase: Ante=0.17, PP = 0.83 χ² = 29.3, p < 0.01. By weeks: Ante OCS = 22 ± 11 w, Ante OCD = 23 ± 9 w; PP OCS = 3.5 ± 7.2 w, PP OCD = 5.2 ± 6.8 w. | onset: Ante < PP |
| Factors associated with changes in OCS, Ante/PP | | | | | | |
| Abramowitz  2010 | 52  30 ± 5 | PP: < 12m | ≥1 PP intrusion  (USA) | YBOCS, EPDS, STAI,  PHQ | Progression. OCS ~ PP time: obsess r = 0.13 p=0.36, compuls r= 0.14 p = 0.34; PP 0-3m vs > PP 3m p > 0.53. Associated factors. depress ↑↑ obsess = 0.25 p < 0.05, depress ↑↑ compuls = 0.38 p < 0.05, anx ↑↑ obsess = 0.31 p < 0.05, anx ↑↑ compuls = 0.37 p < 0.05; OCS severity: depress group = 5.3 ± 3.0, not depress group = 2.6 ± 1.6, p < 0.001. | OCS PP ~ time  depress/anx ↑↑ OCS |
| Altemus 2012 | 229  32 ± 1 | Ante:  1^st^ - 3d trimester  PP: 0-12m | help-seeking (USA) | medical record | Prevalence, in MDD, Ante onset vs PP onset. History of OCD: OR = 0.61 [95%CI, 0.14 -2.60] p =0.50; current Intrusive Violent Thoughts: OR = 0.22 [95%CI, 0.08-0.62] p < 0.01; current other OCD Sx: OR = 0.31 [95%CI, 0.11-0.89] p = 0.03;  Prevalence. In Ante Intrusive Violent Thoughts: history of OCD = 0.12, history of subOCD = 0.08; in Ante other OCD Sx: history of OCD = 0.30, history of subOCD = 0.20. | PP Depress ↑↑ PP Intrusive Violent Thoughts/ other OCS |
| House 2016 | 208  33 ± 4 | Ante: 1^st^, 2^nd^, 3d trimester  PP: 0-6w,  7-24w,  25-52w | OCD,  noOCD,  (USA) | SCID, YBOCS | Progression. OCS severity in OCD: Ante 1^st^ tri = 10.9 ± 8.9, 2^nd^ tri = 8.4 ± 9.3, 3d tri = 9.03 ± 8.3; PP “0-6w” =10.3 ± 10.1, “7-24w” = 10.1 ± 8.6, “25-52w” = 9.6 ± 7.6.  Associated factors. OCS ~ age: Comp r = − 0.25 p < 0.0001, Obses r = − 0.20, p = 0.002, overall r = − 0.24, p<0.001; Preg Loss: OCD OR = 0.48 [95%CI, 0.23-0.99] χ2 = 4.08, p = 0.04; Comorbidity: Panic Disorder OR = NA *z* = 2.89, p = 0.004, Social Anxiety OR = NA *z*= 2.88, p=0.004; BPD, depress, schiz, PTSD, GAD, SUD p > 0.10; OCS ~ Cesarian (vs Vaginal): β = 5.4, p = 0.04. | OCS Ante ~ PP  age /panic d/o, social anxiety/ Cesarian ↑↑ OCS  Preg Loss:  no OCD > OCD |
| Miller  2013 | 461→  329 | PP: 2w, 6m | GEN  (ISL) | PHQ-9,  YBOCS | Prevalence. 2w PP = 0.11, 6m PP = 0.106. Risk factors. Age: OCD = 31.2 ± 4.9 HC = 33.9 ± 4.9 d = 0.55 p = 0.03, Education, %college: OCD = 0.79 HC = 0.87 p = 0.02; %Never married: OCD = 0.19 HC = 0.06 p < 0.01; Psych history %, Anxiety: OCD = 0.09 HC = 0.04, Depression/anxiety: OCD = 0.04 HC = 0.0, Bipolar: OCD = 0.02 HC = 0.002, OCD: OCD = 0.04 HC = 0, relation. counseling: OCD = 0 HC = 0.02; p = 0.02. Progression. 2w OCD → 6m OCD: R^2^ = 0.20 p < 0.01. | PP: Prevalence↑  Age, edu, married, prior anxiety, depress, bipolar, relation counseling → PP OCD; PP OCD persist |
| Miller  2015 | 461→  331  33 ± 5 | PP: 2w, 6m | GEN  (USA) | YBOCS, PHQ-9, STAI | Associated factors. 2w PP: OCS (Depress-) = 1.3 ± 2.9, OCS (Depress+) = 4.0 ± 5.0, p < 0.01; Obsess (Depress-) = 0.7 ± 1.7, Obsess (Depress+) = 2.0 ± 3.2, p < 0.01; Comps (Depress-) = 0.5 ± 1.4, Comps (Depress+) = 1.1 ± 2.4, p = 0.02. 6m PP: OCS (Depress-) = 1.7 ± 3.3, OCS (Depress+) = 3.3 ± 4.5, p < 0.01; Obsess (Depress-) = 1.2 ± 2.0, Obsess (Depress+) = 2.2 ± 2.8, p < 0.01; Comps (Depress-) = 0.6 ± 1.6, Comps (Depress+) = 1.3 ± 2.6, p<0.01. | PP:  Depress ↑↑ OCD |
| Prevatt 2017 | 100  33 ± 5 | PP: <3m | GEN  (CAN) | YBOCS, SASSI-3 | Associated factors. OCS (Subst abuse-) = 4.35 ± 5.95, OCS (Subst abuse +) =7.21 ± 7.71 p = 0.002; OCS (Subst depend-) = 4.51 ± 5.95, OCS (Subst depend+) =8.80 ± 8.46, p = 0.03 | Subst abuse/depend ↑↑  PP OCS |
| Fairbrother 2018 | 100  32 ± 4 | Ante: 36w  PP: 4w, 12w | GEN  (CAN) | OBQ-44, PPII, BDI-II, MAF, PSQI | Risk factors. Ante OC beliefs →PP obsess: 4w = 0.34 p < 0.01, 12w = 0.36 p <0.001.  Ante OC beliefs → PP Accidental harm subscales (var expl r^2^), at 4w: Time = 0.13 p < 0.01, Distress = 0.31 p<0.01, Parenting = 0.11 p < 0.01, Life = 0.08 p < 0.01, Distract = 0.16 p<0.01, Behav responses = 0.23 p<0.01; at 12w: Time = 0.09 p < 0.01, Distress = 0.15 p < 0.01, Parenting = 0.08 p < 0.05, Life = 0.10 p < 0.01, Distract = 0.18 p < 0.001, Behav responses = 0.17 p < 0.01. Ante OC beliefs → PP Intentional harm subscales (var expl r^2^), at 4w: Time = 0.13 p < 0.05, Distress = 0.18 p < 0.01, Parenting = 0.08 p < 0.01, Behav responses = 0.16 p < 0.01; at 12w: Distress = 0.50 p < 0.01, Parenting = 0.36 p < 0.05, Behav responses = 0.41 p < 0.05. Associated factors, with PP OCS (Pearlson r). Poor Sleep Quality: 4w = 0.34 (p<0.01), 12w = 0.18 (NS); fatigue: 4w = 0.16 (NS), 12w = 0.44 (p<0.001); depress: 4w = 0.47 (p<0.001), 12w = 0.67 (p<0.001)**.** | Ante OC beliefs ↑↑PP intrusive thoughts;  PP 4w:  OCS ↑↓ sleep quality  OCS ↑↑ depress  PP 12w:  OCS ↑↑ fatigue/ depress |
| Howard 2018 | 545  32 ± NA  (16-48) | Ante: NA | GEN  (GBR) | Whooley (W), EPDS | Prevalence. OCD = 0.02 [95%CI, 0.01-0.04]. Prevalence by pre-screened groups. OCD (W+) = 0.05 [95%CI, 0.033-0.07], OCD (W-) = 0.02 [95%CI,0.004-0.04], OCD (EPDS+) = 0.03 [95%CI, 0.01-0.05], OCD (EPDS-) = 0.02 [95%CI, 0.01-0.04]. | Ante depress ↑↑ OCD |
| Osnes  2020 | 530  31 ± 4 | Ante: 17w PP: 8w | GEN, (NOR) | SCL-A, MINI, BIS | Prevalence, in insomnia, %. Ante 17w: OCS (Ins+) = 0.038, OCS (Ins-) = 0.038, OR =1 p = 1.0; PP 8w: OCS (Ins+) = 0.085, OCS (Ins-) = 0.033 OR = 2.3 [95%CI, 1.2 – 6.4] p = 0.03. | insomnia ≠ Ante OCS  insomnia ↑↑ PP OCS |
| Xie  2021a | 3346  28 ± 5 | Ante: 16±5w | GEN  (CHN) | SCL-90-R | Severity. OCS (pre-COVID) = 1.41 ± 0.37, OCS (COVID) =1.43 ± 0.39, p = 0.25. | OCS (pre-COVID) ~  OCS (COVID) |
| Xie  2021b | 3346  28 ± 5 | Ante: 16±5w | GEN  (CHN) | SCL-90-R, PSQI, FES | Associated factors, with Ante OCS (Pearlson r). Family cohesion ↓ (r = –0.24 to –0.41, p < .001); family conflict ↑ (r = 0.20 – 0.33, p < .001). | fam cohesion↑↓Ante OCS  fam conflict↑↑ Ante OCS |
| Grekin  2021 | 271  33 ± 5 | Ante → PP 4w, 8w, 12w | GEN  (USA) | PCL-5, OCI-R, IDAS-GD, W-DEQ-A, MOS-SS, PES, PLDS | Associated factors, with PP OCS (Pearlson r). **4w:** PTSS =0.52 p< 0.01; depress = 0.58 p < 0.001; fear of childbirth = 0.38 p <0.01; pers support = -0.25 p < 0.01; obj. birth stress = 0.28 p < 0.01.**8w:** PTSS = 0.60 p < 0.001; depress = 0.63 p < 0.001; fear of childbirth = 0.35 p < 0.01; obj. birth stress = 0.41 p < 0.001; pers. support = –0.21 p < 0.05.**12w**: PTSS = 0.57 p < 0.001; depress = 0.55 p < 0.001; fear of childbirth = 0.31 p < 0.01; obj. birth stress = 0.22 p < 0.05. | PP OCS ↑↑ PTSS, depress, fear ,birth stress |
| Kimmel  2021 | 126  32 ± 4 | Ante 38w | GEN  (SWE) | MINI, PPG-HRV | Associated factors, with Ante OCD: HRV 38 w: OCD = 4.75 ± 1.05 non-OCD = 5.73 ± 1.28 Cohen’s d = 0.77 p = 0.004. | HRV Ante:  OCD < non-OCD |
| Thompson  2021 | 369  30 ± 4 | PP <12mo | GEN  (USA) | DOCS, COVID-distress | Associated factors, with PP OCS (v.s. non pregrn), η²: OCS = 0.02 p =.022 (d ≈ 0.29);  contam. = 0.03 p =.001 (d ≈ 0.35); resp for harm =0.05 p < .001 (d ≈ 0.46);  unaccep thghts < 0.001p =0.70; symm=0.001 p =0.51. OCS ~ COVID-distress × PP = 0.06 p =0.007, 95% CI [0.02–0.10]. | PP↑↑OCS  PP OCS ↑↑ COVID-distress |
| Mahaffey  2022 | 4451  31 ± 5 | Ante | GEN  (USA) | SCID-5 OCD screen OCI-R; | Associated factors, with Ante OCS: pandemic-related stress = 0.17 p < 0.001; maternal age = -0.24 p < 0.001, 1^st^ pregnancy = -0.14 p < 0.01. | Ante OCS ↑↑ COVID stress  Ante OCS ↑↓age/1t pregn |
| Miller  2022 | Ante 276  PP=221  31 ± 5 | Ante: 28w-32w  PP: 6w-8w | GEN  (USA) | MINI, IMAS | Prevalence Ante = 6.2 PP = 14  Associated factors. Pearlson r. Total maladjustment=0.15 p <0.05; family role = 0.21 p <0.01. | Ante < PP  PP OCS ↑↓family role |
| Hinds  2023 | 392  NA | Ante: 37w | GEN (IRL) | YBOCS, interview | Prevalence. %OCD (>13 YBOCS) = 0.10, Risk for OCS severity, U. ante med Dx = −3.55, p < 0.001, current med treatment = −3.58, p < 0.001, ante medication history = −2.99, p = 0.003, relationship strain = 3.70, p < 0.001. | PP OCS ↑↑ante med Dx, med treat, medicat hist, relation strain |
| Miller  2023 | Ante=251  PP=216  31 ± 5 | Ant:e 28-32w,  PP: 6-8w | GEN  (USA) | MINI, IMAS | Prevalence. Current OCD: Ante= 4.4%, PP = 3.2%.  Associated factors: LCA of intern Sx: OCD loads on anxious–fearful (social anxiety + agoraphobia + phobias + OCD) and mood-reactivity (mania + OCD), but not distress. | Ante > PP  OCD↑↑ anxious–fearful, mood-reactivity |
| Sun  2023 | 284  33 ± 4 | PP: 42 ± 7 d | GEN  (CHN) | MINI,  SCL-90 | Associated factors, OR. High stress = 6.26 [95% CI, 1.35–29.09], p = 0.019, Baby female sex= 5.95 [95% CI, 1.35, 25.0], p = 0.022. | Stress/baby F sex ↑↑OCD |
| Drake  2024 | 167  31 ± 5 | PP: 3d, 4w, 12w | GEN (USA) | IDAS-II, PTBC, PCL-C, TLEQ, PRAMS. | Prevalence. OCD: check =0.08, order = 0.06, wash =0.10. Risk factors. *PTSD → OCD & PP intrusive, r:* 4w: preoccup = 0.45 p < 0.01, neutr = 0.21 p = 0.01, check = 0.49 p < 0.01, order = 0.38 p < 0.01, wash = 0.31 p < 0.01, obses = 0.51 p < 0.01. 12w: preoccup = 0.32 p < 0.001, neutr = 0.29 p = 0.002, check = 0.46 p < 0.001, order = 0.39 p < 0.01, wash = 0.32 p < 0.01, obses = 0.46 p < 0.01. *PTSD → OCD, controlling for deprs, β:* 4w: Check = 0.33 p < 0.01; obses = 0.33 p < 0.01. 12w: check = 0.40 p < 0.01; obses = 0.34 p = 0.01. | PTSD↑↑check/obsess |
| Miller  2024 | 270  31 ± 6 | Ante  PP: 4w, 8w, 12 w | GEN  (USA) | PCL-5 (PTSD), OCI-R (OCD), | Progression & Associated factors, ante → PP (4w → 8w → 12w): hoard → wash = 0.15 (p<.05); neutr → avoid = 0.14 (p<.05); hyperarousal → dysphoria = 0.14 (p<.05); neg cogn in PTSD → order = −0.14 (p<.05). | neg cogn in PTSD → order  neutr → avoid |
| Wang  2024 | 122  38 ± 3 | Ante:28w  PP: 6w | 35+  (CHN) | YBOCS, SAS, EPDS, SOC-13 | Associated factors, OR OCS. Pregnancy history = 1.82 [95%, 1.15–2.89], p = 0.008; complications = 1.67 [95%, 1.23–2.26], p < 0.001; fetal health = 1.75 [95%, 1.25–2.47], p <0 .001; SAS = 1.89 [95%, 1.06–3.37], p = 0.027; EPDS = 1.67 [95%, 1.10–2.53], p = 0.012; SOC-13 = 1.27 [95%, 1.07–1.52], p = 0.002. | anx, depres, obstetric risk↑↑Ante/PP OCS  resilience ↑↓ Ante/PP OCS |
| OCD as a Risk Factor for Associated Conditions, Ante/PP | | | | | | |
| Lord  2011 | 18  32 ± 5 | PP: 17±7w | OCD,  HC  (CAN) | CPT, sal cortisol, STAI, subj stress | Risk for PP stress reactivity. Induced stress → distress (OCD > noOCD η_p_^2^, p-value): Subj stress = 0.41 p = 0.006; Anxiety = 0.47 p = 0.002; Cortisol Release =0.53 p = 0.001. | PP OCD ↑↑ stress reactivity |
| Challacombe  2016 | 73  34 ± 5 | PP: 6m | OCD, HC (GBR) | PSSS, GRIMS, MSES | Risk for aberrant parenting experience, OCD vs HC. Parental SE d = 0.96 p < 0.0001; Soc support d = 0.82 p <0.001; marital satisfy d = 1.11 p = 0.009; poor relation build OR = 5.6 p = 0.018; enjoy nappy d = 0.59 p = 0.002; enjoy feed d = 0.65 p p = 0.015; enjoy play d = 0.97 p 0.011. | Parenting experience:  OCD < HC |
| Furtado 2019 | 35  31 ± 6 | Ante: 27w+  PP: 2w, 6w | Anx d/o  (CAN) | HAM-A,  OCI-R | Risk for PP anxiety. Ante OCS → ΔAnx (PP 6w – Ante), mean rank/sum of ranks: OCS (ΔAnx > 0) = 23.0/391, OCS (ΔAnx ≤ 0) =13.3/239, U = 68, z = -2.8, p = 0.004. | Ante OCS ↑↑ PP ΔAnx |
| Johansen  2020 | 336,552 | Ante: 2m PP: 6n, 12m | GEN  (USA) | med records | Risk for PP depression. OCD, OR. 2 m = 7.72 [(%%CI, 6.75-8.83]; 12 m = 5.47 [95%CI, 4.86-6.16]. | Ante OCD ↑↑ PP depress |
| Nasiri  2021 | 13792544 | Nationwide Inpatient Sample, 1999–2015 | GEN  USA | medical records | Prevalence: OCD in childbirth = 2.4%; (in 1999: 0.7%; in 2015: 5.6%)  Risk Ante OCD for mothers: Gestational hypertension = 1.50 [95%CI, 1.14–1.98], p = 0.004; Preeclampsia = 1.22 [95%CI, 1.05–1.41], p = 0.007; Premature rupture of membranes = 1.32 [95%CI, 1.15–1.52], p = 0.0001; Cesarean delivery = 1.08 [95%CI, 1.01–1.17], p = 0.026; Instrumental delivery = 1.17 [95%CI, 1.01–1.35], p = 0.032; Induction of labor = 0.92 [95%CI, 0.86–0.99], p = 0.026; VTE = 1.55 [95%CI, 1.02–2.37], p = 0.041 Risk Ante OCS for children. Preterm birth = 1.31 [95%CI, 1.17–1.46], p < 0.001; Small for gestational age = 1.08 [95%CI, 0.88–1.31]; Intrauterine fetal death = 1.09 [95%CI, 0.75–1.58]. | Ante OCD: 1999 <2015  Ante OCD ↑↑ PP complic  Ante OCD ↑↑ Preterm birth |
| Fairbrother  2022 | 388  33 ± 5 | PP: ≤ 9mo | GEN  (CAN) | PPII, SCID-5, PBQ | Prevalence. OCD = 0.17 [95% CI, 0.14–0.21], UITs of infant harm = 0.44 [95%CI, 0.39–0.50]. Risk. UITs of infant harm → Infant aggression: 2.6% vs 3.1%, RR = 0.84 [95% CI, 0.25–2.87]. OCD → Infant aggression: 1.9% vs 3.5%, RR = 0.52 [95%CI, 0.07–2.00].  . | UIT/OCD ⊥ Infant aggression |
| Gibbs  2023 | 125 Ante → 96 PP  18–35 | Ante: ≥27w PP: ≈27w | Rural  (USA) | YBOCS IBQ-R-SF | Risk Ante OCS for infant neg. affectivity: OC×SER β = 0.02 p = 0.39  Risk PP OCS for infant neg. affectivity: OC×SER β = -0.33 p = 0.02 | low SER:  PP OCS ↑ infant neg. affect |
| de la Cruz  2023 | 1431251  31 ± 5 | PP | OCD,  GEN  (SWE) | ICD-10 codes, registry | Risk for adverse perinatal outcomes. OCD > noOCD, adjusted OR: Preterm birth = 1.25 [95% CI, 1.15–1.35], p < 0.001; Cesarean delivery = 1.24 [95% CI, 1.19–1.30], p < 0.001;  Induction of labor = 1.11 [95% CI, 1.05–1.18], p < 0.001; Low birth weight (<2500 g) = 1.13 [95% CI, 1.02–1.26], p = 0.02; Neonatal intensive care admission = 1.17 [95% CI, 1.10–1.25], p < 0.001; Apgar <7 at 5 min = 1.12 [95% CI, 1.02–1.23], p = 0.02. | adverse perinatal outcomes:  OCD > noOCD |
| OCS in Fathers, Ante/PP | | | | | | |
| Coelho  2014 | fathers 726 →  707 | Ante:  28-34w  PP:  30-60d | GEN  (BRA) | MINI | Prevalence. %OCD: Ante = 0.034 PP = 0.018. Comorbid. Ante: bipolar mixed RR=7.8 [95%CI, 2.0-30.2], manic episodes RR=15.8 [95%CI, 2.5-99.0], PP: bipolar mixed RR = 17.4 [95%CI, 3.1-96.2], unipolar depress RR = 28.2 [95%CI, 6.7-119.4]. Progression. Bipolar mixed ante → OCD PP RR=10.9 [95%CI, 2.8-42.9]. Concordant OCD, mothers & fathers. Ante RR = 6.1 [95%CI, 1.8-21.2], PP RR = 5.9 [95%CI, 1.2-28.4]. | Fathers OCD:  Ante > PP  CC: Ante = bipolar,  PP = bipolar/unipolar  OCD fathers↑↑mothers |
| Assessments | | | | | | |
| Pereira  2022 | 212  33 ± 7 | PP: 3-6m | GEN  (PRT) | POCS, DIPD-PP | Prevalence. %OCD: 0.033; %OCS: ≥ 1 OCS = 0.76; ≥ 1 obsess=0.74; ≥ 1 comp.=0.42; CFA fit: χ²/df = 2.30, CFI = 0.93, TLI = 0.91, RMSEA = 0.086. Reliability: α = 0.91 (total), 0.82–0.92 (subscales). Convergent validity: r = .27–.42 with PDSS/PASS. Criterion: OCD > Dep/Anx p < .01. AUC = 0.97, cut-off = 20 (Sens 85.7 %, Spec 100 %). | POCS valid for ppOCD |
| Fairbrother  2023 | 574  33 ± 5 | Ante: 37w, PP: 9w, 21w | GEN, (CAN) | DOCS, EPDS/EPDS-3A, SCID-5 | Prevalence. %OCD: Ante = 0.036, PP-2mo = 0.106, PP-5mo = 0.058. Screening accuracy:  DOCS AUC: Ante = 0.90 [95%CI, 0.80–1.00], PP-2mo = 0.81 [95%CI, 0.74–0.88], PP-5mo = 0.88 [95%CI, 0.80–0.96]; EPDS-Full AUC: Ante = 0.71 [95%CI, 0.54–0.88], PP-2mo = 0.79 [95%CI, 0.72–0.85], PP-5mo = 0.78 [95%CI, 0.66–0.90]; EPDS-3A AUC: Ante = 0.73 [95%CI, 0.55–0.91], PP-2mo = 0.79 [95%CI, 0.72–0.86], PP-5mo = 0.80 [95%CI, 0.68–0.92]. Best-performing subscales. Unacc thghts: Ante AUC = 0.86, J = 0.54, LR⁺ = 3.84; PP-5mo AUC = 0.85, J = 0.63, LR⁺ = 5.85; Resp/Harm: Ante AUC = 0.81, J = 0.53, LR⁺ = 5.82; PP-5mo AUC = 0.83, J = 0.47, LR⁺ = 2.81. | best screener: DOCS (unacc thghts)  best timing: late ANTE and 4-6 mo PP |

**Abbreviations used in Tables A3.1 -A3.3**

**Instruments:** **ACES** - the Adverse Childhood Experience scale^1^; **ACE-IQ** – the Adverse Childhood Experiences– International Questionnaire^2^; **Anger-in** – *Anger-in* subscale of the State-Trait Anger expression Inventory 2^3^; **Anger-out** - *Anger-out* subscale of the State-Trait Anger expression Inventory 2^3^; **BAI** – the Beck Anxiety Inventory^4^; **BDI** - the Beck Depression Inventory^5^; **BDI-II** - the Beck Depression Inventory–second edition^6^; **BIS** - the Bergen Insomnia Scale^7, 8^; **BOCS** – the Brief Obsessive-Compulsive Scale;^9^ **CAPS-CA-5** – the Clinician-Administered PTSD Scale for DSM-5–Child/Adolescent Version;^10^ **CAS** - Clinical anger scale^11^; **CASI** – the Childhood Anxiety Sensitivity Index;^12^ **CBCL** - the Child Behavior Checklist^13^; **CCPMS** – the COVID-19 Compliance with Prevention Measures Scale;^14^ **CDI** - Children’s depression inventory^15^; **C-GAS** - the Children’s Global Assessment Scale^16^; **CGI-I** – the Clinical Global Impressions–Improvement Scale;^17^ **CIDI** - Composite International Diagnostic Interview^18^; **CIDI65+** - age‐adapted Composite International Diagnostic Interview^19^; **CID-S** - the Composite International Diagnostic–Screener^20^; **CIS** - Coronavirus Impact Scale^21^; **CIS-R** - Clinical Interview Schedule–Revised^22^; **COPE** - Coping Orientation to Problems Experienced^23^; **CPT** - the Cold Pressor Test^24, 25^; **CSI-4** - Child Symptom Inventory – 4^26^; **CSS** – the COVID Stress Scales;^27^ **CTQ**/**CTQ-SF** – the Childhood Trauma Questionnaire/ Short Form;^28, 29^ **CY-BOCS** - the Children’s Yale-Brown Obsessive-Compulsive Scale^30^; **DASS** (**DASS-21**) - the short‐form version of the Depression Anxiety Stress Scales^31^; **DAWBA** - the Developmental and Well-Being Assessment^32^; **DIA-X/M-CIDI** - an updated version of the World Health Organization's CIDI with additional questions to cover DSM-IV and ICD-10^33^ criteria;^34, 35^; **DIGS** – the Diagnostic Interview for Genetic Studies^36^; **DOCS/DOCS-SF** – the Dimensional Obsessive-Compulsive Scale / Short Form;^37, 38^ **DS** - Disgust Scale^39^; **DTS** – the Distress Tolerance Scale;^40^ **DSM** (**DSM-IV**) - Diagnostic and Statistical Manual of Mental Disorders (fourth edition^41^); **DY-BOCS** - The Dimensional Yale–Brown Obsessive–Compulsive Scale^42^; **EPDS** **/EPDS-3A** - Edinburgh postnatal depression scale^43, 44^; **ERI** – the Effort–Reward Imbalance Questionnaire;^45^ **EuroQol** – the Euro Group Quality of Life questionnaire^46^; **FACES-II** - the Family Adaptability and Cohesion Scale, second edition^47^; **FCV-19S** – the Fear of COVID-19 Scale;^48^ **FHRDC** - the Family History Research Diagnostic Criteria;^49^ FL – the Familial load; **FHS** - the (Psychiatric) Family History Screen^50^; **FOCI-B** – the Florida Obsessive-Compulsive Inventory, symptom/checklist portion;^51^ **FSQ-MV** – the Feared Self Questionnaire–Multidimensional Version;^52^ **GAF** – the Global Assessment of Functioning;^53^ **GHQ-28** - General Health Questionnaire 28^54^; **GOASSESS** - a computerized structured interview version of the Kiddie Schedule for Affective Disorders and Schizophrenia^55^; **GRIMS** - Golumbok-Rust Inventory of Marital Satisfaction^56^; **HAM-A** - The Hamilton Anxiety Scale^57^; **ICD-10DCR** - Mental and Behavioral Disorders: Diagnostic Criteria for Research^58^; **ICD-10/ICD-9-CM** - the International Classification of Disease, 9th Revision /10^th^ Revision/, Clinical Modification;^33, 59^ **IDAS-II** - Inventory of Depression and Anxiety Symptoms, the expended version^60^; **ITQ** - Bates Infant Temperament Questionnaire^61^; **KAP** - the knowledge, attitude, and practice^62^; **K-CIDI** - the Korean version of the WHO-Composite International Diagnostic Interview^63^; **KNOW-P-COVID-19** – the Knowledge of COVID-19 Scale;^64^ **K-SADS** - the Kiddie Schedule for Affective Disorders and Schizophrenia^65^; **K-SADS-PL** - Schedule for Affective Disorders and Schizophrenia for School Aged Children: Present and Lifetime Version^65^; **LEIQ** - the Life Events Inventory Questionnaire^66^; **LIFE-UP** - The Longitudinal interval follow-up evaluation—Upjohn^67^; **LOI-CV** – the Leyton Obsessional Inventory–Child Version;^68^ **LSC-R** - a modified version of the women, co-occurring disorders, and violence study version of the life stressor checklist-revised^69^; **MAF** - The Multidimensional Assessment of Fatigue Scale^70^; **MCR/MCR-C** – the Mental Contamination Report / Chinese version;^71-73^ **MINI-KID** - the Mini International Neuropsychiatric Interview for Children and Adolescents^74^; **DIA-X/M-CIDI** - an updated version of the World Health Organization's CIDI with additional questions to cover DSM-IV and ICD-10^33^ criteria.^34^; **MDQ** - the Mood Disorder Questionnaire^75^; **MINI** - the Mini International Neuropsychiatric Interview^76^; **MINI-Plus** – the Mini-International Neuropsychiatric Interview–Plus;^77^ **MIST** - Montreal Imaging Stress Task^78^; **MOCI** - the Maudsley Obsessional–Compulsive Inventory^79^; **MSES** - Maternal Self-Efficacy Scale^80^; **Non-cons. “dirty kiss” task** - non-consensual “dirty kiss” task^81^; **OBQ/OBQ-44/OBQ-SF** – the Obsessive Beliefs Questionnaire / 44-item version / Short Form^82, 83^; **OBVQ-R** – the Revised Olweus Bully/Victim Questionnaire;^84, 85^ **OCDS** – the Obsessive-Compulsive Disorder Scale;^86^ **OCI-4** – the four-item Obsessive-Compulsive Inventory;^87^ **OCI/OCI-R** – the Obsessive-Compulsive Inventory / Revised version^88, 89^; **Padua Inventory** - a measure of obsessive–compulsive phenomena, which permits measurement of both number and severity of symptoms^90^; **PATCQ** – the Perceptions and Attitudes Towards COVID-19 Questionnaire;^14^ **PCL-5** – Post Traumatic Stress Disorder Checklist for DSM-V^91^; **PDM** – the Postpartum Distress Measure^92^; **PHQ** (**PHQ-9**) - Patient Health Questionnaire;^93^ **PI-C-C** – the Padua Inventory contamination-related measure used in the Chinese mental-contamination study;^72^ **PI-WSUR** - the Padua inventory: Washington State University Revision^94^; **POCS** - the Perinatal Obsessive-Compulsive Scale^95^; **PPII** - The Postpartum Intrusions Interview^96^; **PRAS** - Pregnancy-Related Anxiety Scale^97^; **PSQI** - The Pittsburgh Sleep Quality Index^98^; **PSS/PSS-10** – the Perceived Stress Scale / 10-item version;^99, 100^ **PSSS** – Perceived Social Support Scale from family and friends^101^; **PTBC** - Postpartum Thoughts and Behaviours Checklist^102^; **RAS** - Responsibility Attitudes Questionnaire^103^; **RCADS-OCD** – the obsessive-compulsive disorder subscale of the Revised Child Anxiety and Depression Scale;^104^ **READ** - Resilience Scale for Adolescents^105^; **RIQ** - Responsibility Interpretations Questionnaire^103^; **SASSI-3** - Substance Abuse Subtle Screening Inventory^106^; **SCARED/SCARED-A** – the Screen for Child Anxiety Related Emotional Disorders / adult version;^107, 108^; **SCAS** - the Spence Children’s Anxiety Scale^109^; **SCID-I** - the Structured Clinical Interview for DSM-IV^110^; **SCID-5-RV** – the Structured Clinical Interview for DSM-5;^111^ **SCID-CV** - Structured Clinical Interview (clinician version)^112^; **SCID-I/P** - Structured Clinical Interview (patient version)^113^; **SCID-TCI** - Structured Clinical Interview, modules for tic and impulse control disorders^114^; **SCL-90** - Symptom Checklist 90^115^; **SCL-90-R** - Symptom Checklist, revised^116^; **SCL-A** – the anxiety dimension of SCL-25 from the Hopkins Symptom Check List-25 (SCL-25)^117^; **s-EMBU** - An inventory for the assessment of perceived parental rearing practices^118^; **SF-12** - Short Form health survey^119^; **SPIKE** - the Structured Psychopathological Interview and Rating of Social Consequences of Psychic Disturbances for Epidemiology^120^; **SOCS** – Short OCD Screener;^121^ **STAI** - State Trait Anxiety Inventory^122^; **STAXI-2** - the State-Trait Anger Expression Inventory 2^3^; **TEC-M** – the Tangram Emotion Coding Manual;^123^ **TESI-C** – the Traumatic Events Screening Inventory for Children;^124^ **TEMPS-A** - Temperament Evaluation of Memphis, Pisa and San Diego Autoquestionnaire^125^; **USS** – the University Stress Scale;^126^ **VOCI/VOCI-C** – the Vancouver Obsessional Compulsive Inventory / contamination subscale;^127^ **Whooley** (**W**) - depression-screening Whooley questions^128^; **YBOCS** – the Yale-Brown Obsessive Compulsive Scale^129, 130^; **YBOCS/YBOCS-CL/YBOCS-SC/Y-BOCS-II-SR –** the Yale-Brown Obsessive Compulsive Scale / checklist / Second Edition, Self-Report;^129-131^ **YI-4** - Youth’s inventory-4^132^; **Y-LIFE** - the Longitudinal Interval Follow-up Evaluation^67^; **YONHQ** - Yale OCD Natural History Questionnaire.^133^

**Other abbreviations:** **% var expl** – percent of variance explained; **≥1 PP intrusion** – intrusive thoughts present during the postpartum period; **1 wk** – met diagnostic criteria for OCD within 1 week; **12-mo** – met diagnostic criteria for OCD within 12 months; **1st time OCD** – met diagnostic criteria for OCD for the first time in lifetime; **1st tri** – first trimester of pregnancy; **30-d OCD** – met diagnostic criteria for OCD during the last 30 days; **ACC** – anterior cingulate cortex; **ACE** – adverse childhood experience; **Adj.** – adjusted; **AFAB** – assigned female at birth; **Aggres** – obsessions with aggressive thoughts; **Ante** – antenatal period; **Anx** – anxiety; **AO** – age of onset; **AUC** – area under the receiver operating characteristic curve; **BDS** – bipolar spectrum disorder; **Behav responses** – behavioral responses; **Bipolar mixed RR** – recurrence rates in bipolar disorder; **BP** – blood pressure; **BPD** – bipolar disorder; **CFA** – confirmatory factor analysis; **CFI** – comparative fit index; **Check** – checking compulsions; **cis** – cisgender; **Clean** – cleaning compulsions; **Clinic OCs** – obsessions and compulsions above the DSM-based threshold; **Clozap** – clozapine; **Comps.** – compulsions; **Computerized SR supervised by trained prsnl** – self-reports completed on a computer under supervision of trained research personnel; **Concur valid** – concurrent validity; **Contam** – contamination obsessions; **Controls** – control participants; **Count** – counting compulsions; **d** – effect size Cohen’s d; **d/o** – disorder; **Depress** – depression; **Disabil rate** – disability rate; **Distss Sx** – distressing symptoms; **DLPFC** – dorsolateral prefrontal cortex; **DUI** – duration of untreated illness; **Dx** – diagnosis; **E2** – estradiol; **EA** – emotional abuse; **ED** – emergency department; **Edu** – education; **EHR** – electronic health record; **ELBW** – extremely low birth weight; **EN** – emotional neglect; **EO** – early onset of OCD; **EP** – extremely preterm; **Est.** – estimated; **Extr OCs** – extreme obsessions and compulsions; **F** – females; **Fam** – family; **Fam adap** – family adaptability; **Fam cohes** – family cohesion; **Fam history** – family history; **Fam. risk** – familial risk; **FDR** – false discovery rate; **FL** – familial load; **Forbid thght** – forbidden thoughts; **Frm** – former; **FU/f/u** – follow-up; **GAD** – generalized anxiety disorder; **GD** – gender-diverse; **GEN** – general population; **Genesis 1219 (G1219)** – a longitudinal study that aims to examine the interplay between genes and the environment on the development of anxiety and depression over time;^134^ **GM** – gender minority; **gnc/GNC** – gender nonconforming; **HC** – healthy controls; **HCW** – health care worker; **Hoard** – hoarding obsessions and compulsions; **HR** – hazard ratio; **Hyperthymic tempt** – hyperthymic temperament; **Hypothyr** – hypothyroidism; **Inflam MLR** – monocyte-to-lymphocyte ratio inflammatory marker; **INS** – insular cortex; **Ins+/Ins−** – insomnia present/insomnia absent; **Intrusive Resp.** – intrusive thoughts about responsibility; **IO** – intermediate age of onset; **IPV** – intimate partner violence; **IRR** – incidence rate ratio; **ITG** – inferior temporal gyrus; **J** – Youden index; **life OCD** – met DSM criteria for OCD diagnosis during lifetime; **LO** – late age of onset; **LR+** – positive likelihood ratio; **LSE** – stressful life event; **M** – males; **MDD** – major depressive disorder; **Meds** – medications; **Menstr. phases** – menstrual phases; **Mental comps** – mental compulsions; **Mo** – month; **Mood stab meds hist** – history of mood-stabilizing medications; **Modrt OCs** – moderate obsessions and compulsions; **mOP** – perceived mother overprotection; **mPFC** – medial prefrontal cortex; **NA** – information not available/not reported; **Neg feel** – negative feelings; **NHS Foundation Trust** – – a type of hospital which is independent from the Department of Health and run on a not-for-profit basis;^135^ **noPregn** – nonpregnant; **NS** – non-significant; **OBGYN** – obstetrics and gynecology; **OCD** – obsessive-compulsive disorder; **OCD-spect** – obsessive-compulsive spectrum; **OCs** – obsessions and compulsions; **OFC** – orbitofrontal cortex; **Order** – ordering compulsions; **P4** – progesterone; **PA** – physical abuse; **Parent rept.** – parental reports; **Parenting & interact. baby** – perceived quality of parenting and interactions with the baby; **Past complic** – past complications; **Pb** – puberty; **Peri** – perinatal period; **Pers. comp.** – personal competence as a source of resilience; **pFDR** – false-discovery-rate-adjusted p value; **Phob.** – phobia; **PMOCS** – premenstrual manifestation of obsessive-compulsive disorder symptoms; **PN** – physical neglect; **post COVID inflame** – post-COVID inflammation; **PP** – postpartum; **PP depress** – postpartum depression; **PP OCD persist** – persistent postpartum OCD; **Pregn** – pregnancy; **Premenstr** – premenstrual period; **Primary h/c** – primary health care; **Primary Sx** – primary symptoms; **PSLE** – preceding stressful life event; **Psych concerns** – psychiatric concerns; **Psych Dx** – psychiatric diagnosis; **Psych/edu** – students from psychology/education departments; **Psychos** – psychosis; **Pts repts** – patients’ reports; **PTSD** – post-traumatic stress disorder; **R/ship** – relationship; **Regr** – registry; **Relation. counseling** – attended relationship counseling; **Relig** – religious obsessions; **Repeat** – repetition compulsion; **Resp** – responsibility; **Resp/Harm** – responsibility for harm; **rHR** – heart rate at rest; **RMSEA** – root mean square error of approximation; **RR** – risk ratio; **rsfMRI** – resting-state functional magnetic resonance imaging; **SA** – sexual abuse; **Sal cortisol** – salivary cortisol; **Schiz** – schizophrenia; **school** – K–12 students; **schz pts** – schizophrenia patients; **Sex** – sexual obsessions; **Sex abuse** – sexual abuse; **SGM** – sexual and gender minority; **SLE** – stressful life event; **SM** – sexual minority; **SOB** – season of birth; **Soc comp** – social competence as a source of resilience; **Soc res** – social resources as a source of resilience; **Socil phob** – social phobia; **SOCD** – subthreshold obsessive-compulsive disorder; **Somat** – somatic obsessions; **SR** – self-reports; **SRMR** – standardized root mean square residual; **SSRI** – selective serotonin reuptake inhibitor; **STG** – superior temporal gyrus; **Struc style** – structured style as a source of resilience; **Study-spec** – study-specific; **Subgr** – subgroup; **Subj stress** – subjective stress; **subOCD** – subclinical/subthreshold OCD; **subOCs** – subclinical/subthreshold obsessions and compulsions; **Subst abuse** – substance abuse; **Subst depend** – substance dependence; **SUD** – substance use disorder; **Suicide Att** – suicide attempts; **Suicide Id** – suicide ideation; **Sx** – symptoms; **Sx phase** – symptom phase; **Sym** – symmetry obsessions; **SZ-spect** – schizophrenia spectrum; **TLI** – Tucker–Lewis index; **Trained prsnl** – trained personnel; **trans** – transgender; **Treatm in adults** – treatment in adulthood; **Unacc thghts** – unacceptable thoughts; **Unwanted sexual exp recall** – unwanted sexual experience recall; **Var expl R²** – variance explained, R²; **Veterans/ancle inj** – veterans with ankle injury; **W/wks** – weeks; **Yo** – years old; **ΔERN** – change in error-related negativity; **ΔOCD** – change in OCD incidence/prevalence; **ΔOCs** – change in obsessive-compulsive symptoms. **M** – males; **F** – females.

**Appendix 4.** Papers that addressed gender differences in comorbid and co-occurring conditions in individuals with obsessive compulsive disorder.

Table A4.1. Gender differences in comorbid and co-occurring conditions in OCD.

| **First Auth,**  **Year** | **N**  **(F)** | **Age**  **mean±SD**  **(range)** | **Pop** | **Meas** | **Gender effects (F/M)** | |
| --- | --- | --- | --- | --- | --- | --- |
|  |  |  |  |  | **Stats** | **Effect** |
| General/Multiple Comorbid Psychiatric Conditions (CC) | | | | | | |
| *pooled samples* | | | | | | |
| Huang  2014 | 2040 (1054) | NA  6-65+ | OCD  TWN | Registry Analysis | **Comorbidity in OCD, F:M OR.** Child (6 – 17): Any = 0.71 [95%CI, 0.43-1.17]; 1 CC = 0.83 [95%CI, 0.49-1.40]; 2+ CC = 0.45 [95%CI, 0.19-1.05].  Anxiety Spectrum = 0.43 [95%CI, 0.18-1.0]; MDD =1.11 [95%CI, 0.3-5.0]; Depressive disorders = 1.67 [95%CI, 0.91-2.50]; BD = 0 [95%CI, 0-0]; Schz = 1.25 [95%CI, 0.42-3.33]; ASD = 0.63 [95%CI, 0.21-2.0]; TD = 0 [95%CI, 0-0];  ADHD = 0.67 [95%CI, 0.34-1.43]; CD = 2.5 [95%CI, 0.45-10]; AdjD = 0.15 [95%CI,0.02-1.11].  Adult (18+): Any = 1.29 [95%CI, 1.07-1.56], 1 CC = 1.21 [95%CI, 1.00-1.48], 2+ CC = 1.13 [95%CI, 0.81-1.58].  Anxiety Spectrum = 0.83 [95%CI, 0.59-1.19]; GAD = 1.25 [95%CI, 0.91-1.67]; Social phobia = 0.21 [95%CI, 0.05-1.0]; Panic = 0 [95%CI, 0-0]; MDD = 1.43 [95%CI, 0.91-2.0]; Depressive disorders = 1.11 [95%CI, 0.91-1.25]; PTSD = 1.25 [95%CI, 0.21-10.0]; BPD = 1.25 [95%CI, 0.77-2.0]; Schz = 0.67 [95%CI, 0.50-1.0]; AdjD = 1.0 [95%CI, 0.56-2.0]. | Child:  Any CC/ MDD/ DD/ BPD/ SZ/ ASD/ Externalizing: M ~ F  Anx Sp: M > F  Adult:  Any CC: M < F  Anx Sp/ GAD/ Panic/ MDD/ DD/ PTSD/ BPD/ AdjD: M ~ F  Soc ph/ SZ: M > F |
| *children/adolescents* | | | | | | |
| Farrell  2012 | 43  (13) | 11 ± 3  (7-17) | OCD AUS | ADIS-P, CY-BOCS | **Comorbidity in OCD, F:M OR.** Any = 0.09 [95%CI, 0.02-0.38] | Any CC: M > F |
| Vivan  2014 | 75  (59) | 16 ± 1  (14-17) | high school  BRA | SCID-IV, YBOCS, BDI | **Comorbidity in OCD, F:M OR.** Any = 0.99 [95%CI, 0.32-3.01]  Any from Anxiety Spectrum = 0.99 [95%CI, 0.32-3.01]; Generalized anxiety = 0.99 [95%CI, 0.17-5.05]; SPe = 4.71 [95%CI, 0.26-87.03]; Social phobia = 0.53 [95%CI, 0.05 – 6.20]; Panic disorder = 0.26 [95%CI, 0.02-4.38]; MDD = 3.83 [95%CI, 0.56-31.95]; PTSD = 0.26 [95%CI, 0.02-4.38]. BPD = 4.71 [95%CI, 0.26-87.03]. Impulse control disorders = 0.77 [95%CI, 0.21-2.80]; ADHD = 0.51 [95%CI, 0.09-3.07]; Chronic Tic = 0.03 [95%CI, 0.002-0.67]; TD = 0.26 [95%CI, 0.02-4.38].  **Comorbidity in OCD, severity:** depression M < F, d = 0.30 p = 0.03 | Any CC/ anxiety spectrum/ MDD/ BPD/ PTSD/ impulse control / TD: M ~ F  Chronic Tic: M > F  Depression severity: M < F |
| Tanidir  2015 | 110  (62) | 12 ± 3  (6-18) | OCD TUR | K-SADS-PL | **Comorbidity in OCD, F:M OR.** Anxiety Spectrum = 0.66 [95%CI, 0.55-2.59], Spec. ph = 2.75 [95%CI, 1.1 – 6.9], GAD = 2.4 [95%CI, 0.91-6.33]; DD =1.65 [95%CI, 0.72-3.80], Extern = 0.64 [95%CI, 0.28-1.47], ADHD = 0.72 [95%CI, 0.29-1.79], ODD = 0.3 [95%CI, 0.09-1.04], TD = 0.51 [95%CI, 0.2-1.33] | Spec ph / GAD: M ≤ F  Anx/ DD/ Ext/ ADHD/Tic: M ~ F  ODD: M > F |
| Arildskov 2016 | 257 (132) | 13 ± 3  (7-17) | OCD DNK/ NOR/ SWE | K-SADS-PL, CY-BOCS, ASSQ | **Comorbidity in OCD, F:M OR.** Any = 0.83 [95%CI, 0.50 – 1.37]; 1 CC = 1.32 [95%CI, 0.74-2.36], 2+ CC = 0.42 [95%CI, 0.20-0.88]; Anx = 1.32 [95%CI, 0.71-2.45], DD = 2.28 [95%CI, 0.58-9.01], ADHD = 0.04 [95%CI, 0.01-0.29], Exter = 0.46 [95%CI, 0.11-1.89], TD = 0.55 [95%CI, 0.29-1.04], ASD Sx = 0.29 [95%CI, 0.14-0.60] | ADHD/poly CC/Tic/ ASD Sx/ ASD Sx Severity: M > F  Any, 1 CC, Anx/ DD/ Ext: M ~ F |
| Ortiz  2016 | 112  (55) | 15 ± 3  (8-18) | OCD ESP | K-SADS-PL, CY-BOCS | **Comorbidity in OCD, F:M OR.** Any = 1.42 [95%CI, 0.64-3.14]; NDC = 0.57 [95%CI, 0.19-1.73]; IC = 2.1 [95%CI, 0.89-4.95].  IC vs. NDC = 3.66 [95%CI, 1.28-10.44] | IC vs NDC:  M < F |
| Peris  2017 | 332 (148) | 12 ± 3  (7-17) | OCD  USA | ADIS-C, CYBOCS | **Comorbidity in OCD, F:M OR.** Anxiety Spectrum = 1.20 [95%CI, 0.77-1.86], DD = 1.17 [95%CI, 0.59-2.31], Extren. = 0.59 [95%CI, 0.32-1.09], Tic = 1.09 [95%CI, 0.53-2.34], ASD = 0.32 [95%CI, 0.07-1.56]. | Extern: M ≥ F Anx/ DD/Tic/ ASD:  M ~ F |
| Rintala  2017 | 3372 (1793) | 15 ± 4  (3-25) | OCD  FIN | Registry records | **Comorbidity in OCD, F:M OR.** Any =1.22 [95%CI, 1.05-1.42]  Anxiety Spectrum **=** 1.78 [95%CI, 1.55-2.05]; Depression **=** 2.03 [1.76-2.34], p<0.001; Eating Disorder **=** 5.66 [95%CI, 4.14-7.73]. BPD **=** 1.78 [95%CI, 1.22-2.59]. Psychotic Disorder **=** 0.71 [95%CI, 0.58-0.86]; ASD **=** 0.22 [95%CI, 0.17-0.03]; TD **=** 0.23 [95%CI, 0.16-0.34]; ADHD + ADD **=** 0.32 [95%CI, 0.24-0.42]; Conduct Disorder **=** 0.67 [95%CI, 0.46-0.97]; Developmental Disorder = 0.35 [95%CI, 0.28-0.44]; Mental Retardation **=** 0.72 [95%CI, 0.38-1.34] | Any CC/ Anxiety/ Depression/ ED/ BPD: M < F  Retardation M ~ F Psychosis/ ASD/ TD/ Externalizing/ Developmental: M > F |
| Zijlmans  2017 | 4632  (2287) | 8 ± 1  (8-11) | OCD  NLD | SOCS, CBCD, CPRS-R-S | **Comorbidity in OCD, F:M OR.** ASD subtype = 0.41 [95%CI, 0.14-1.24], Externalizing subtype = 0.24 [95%CI, 0.08-0.73]. | ASD subtype/Extern subtype: M > F |
| Schneider  2018 | 3149 (1159) | High School | GEN  AUS | BPD DQ-A; SCA | **OCs ~BDD-OS, Spearman ρ:** F = 0.32 M = 0.26 Fisher p = 0.038  **OCs ~BDD-IA/ED, Spearman ρ:** F = 0.32-0.34 M = 0.30 – 0.32 Fisher p > 0.18 | OCs↑↑BDD-OS: M < F; OCs↑↑BDD-IA/ED:M~F |
| Mohammadi  2021 | 29858  (15242) | (6-9, 10-14, 15-18) | GEN  IRN | K-SADS-PL | **Comorbidity in OCD, F:M OR.** Mood Disorders = 1.19 [95% CI, 0.75-1.9], Psychosis = 1.15 [95% CI, 0.32-4.11], Anxiety Disorders = 1.27 [95% CI, 0.94-1.72], Behavioral Disorders = 0.53 [95% CI, 0.38-0.75], Neurodevelopmental Disorders = 0.50 [95% CI, 0.24-1.05], Elimination Disorders* = 0.50 [95% CI, 0.30-0.83], Smoking and alcohol disorder = 0.62 [95% CI, 0.32-1.19]  *Elimination Disorders = Enuresis and Encopresis | Elimination d/o: M> F Mood/ Psychosis/ Anx., Behav./ Neurodevelop/ Smoking/Alcohol: M~F |
| Efe  2022a | 457 (254) | 14 ± 4  (NA) | OCD  FIN | K-SADS-PL | **Comorbidity in OCD, F:M OR.** ADHD = 0.52 [95% CI, 0.34–0.79]; Depression = 1.74 [95% CI, 1.05–2.87]; Tic disorders = 0.24 [95% CI, 0.10–0.55]; ASD = 0.06 [95% CI, 0.00–1.07]; Fisher exact p = 0.007; Language d/o = 0.26 [95% CI, 0.08–0.83]; Sleep d/o = 0.51 [95% CI, 0.26–0.99].  *Trends*: Conduct disorder = 0.23 [95% CI, 0.05–1.04] p=0.085; GAD = 1.39 [95% CI, 0.94–2.06] p=0.082; Eating disorders = 2.45 [95% CI, 0.92–6.53] p=0.074;  *n.s.:* ODD, Separation Anxiety, PTSD, PD, Social Phobia, Simple Phobia, BPD, SI, BFRB, Intellectual Disability, Dyslexia, Enuresis, Encopresis, Sex Abuse | ADHD/ TD/ ASD/ lang & sleep d/o: M > F  Conduct d/o: M ≥ F  ODD/ SepA, PTSD/ PD/ SP/ Phobia/ BPD/ SI/ BFRB/ physical cond: M ~ F GAD, ED: M ≤ F  MDD: M < F |
| Yan  2022 | 73992  (37099) | NA  (6-16) | GEN  CHN | CBCL, MINI-KID | **Comorbidity in OCD, F:M OR.** Tic ≈ 1.39 [95%CI, 1.02–1.90]; Affective disorders ≈ 1.82 [95%CI, 1.11–2.99]; ADHD ≈ 0.60 [95%CI, 0.33–1.10] | Tic/ Affective: M < F  ADHD: M ~ F |
| Smárason  2024 | 830 (448) | 13 ± 3  (5-19) | OCD  INT | CY-BOCS, SADS-PL, ADIS-CP | **Latent Clusters of comorbidities in OCD:** Cluster 1 (Internalizing, Tic, ADHD) M~F, Cluster 2 (GAD, Social Phobia, Tics, ADHD) M < F, Cluster 3 (Low Comorb) M ~ F, Cluster 4 (Low to moderate comorb) M > F, Cluster 5 (GAD, Social Phobia) M = F, Cluster 6 (Highest comorbidity – GAD, Social Phobia, Tics, ADHD, ASD ) M = F, Cluster 7 (ASD ) M < F; stats not reported. | Low Comorb.: M > F ASD: M < F |
| Aicoboaie  2025 | 4729 (2343) | 14 ± 4 (NA) | AN  OCD  SCZ  USA | PRS genotype K-SADS | **OC-related characteristics by genetic risk for psychopathology (PRS), OR.**  F: AN PRS → perfectionism, OR = 1.12 [1.00–1.25], p = 0.04; SCZ PRS → perfectionism, OR = 1.15 [1.04–1.28], p = 0.01.  M: corresponding AN PRS → perfectionism OR = 1.03 [0.92–1.16], p = 0.61; SCZ PRS → perfectionism OR = 1.03 [0.91–1.15], p = 0.65 | AN/SCZ PRS → perfectionism: F only |
| D’Aiello  2025 | 121  (52) | NA  (6-17) | OCD  ITA | K-SADS | **NDD comorbidity in OCD.** OCD without NDDs: M:F = 25:30; OCD + NDDs: M:F = 44:22; χ²(1) = 5.50, p = 0.015. NDDs included ASD, ADHD, tic disorders, and/or specific learning disorders. | NDDs in OCD: M > F |
| *adults* |  |  |  |  |  |  |
| Dell’Osso  2012 | 389  (227) | 35 ± 13  (18-60) | Psy pts  HC  ITA | OBS-SR, MOODS-SR | **Comorbid OCs, risk.** Sexual obsess. Mood: M = 2.84 [95%CI, 1.11-7.29], F = 8.97 [95%CI, 2.61-30.89]; Panic: M = 1.05 [95%CI, 0.23-4.76], F = 4.93 [95%CI, 1.22-19.85]; SZ: M = 4.52 [95%CI, 1.75-11.68], F = 20.33 [95%CI, 4.97-83.16]. **Comorbid OCs, F:M OR.** Sexual obsess. In mood **=** 4.23 [95%CI, 1.06-16.82], In panic = 9.78 [95%CI, 1.55-61.65], In SZ = 1.42 [95%CI, 0.33-5.99]. | Sexual OCs ↑↑ mood/ panic/ SZ: M < F  Sexual OCs in mood/ panic: M < F  Sex OCs in SZ: M ~ F |
| Timpano  2012 | 605 (363) | 39 ± NA  (17-92) | OCD USA | SCID-P, YBOCS | **Comorbidity in OCD, F:M OR.** DD = 1.27 [95%CI, 0.86-1.88]; BPD = 1.49 [95%CI, 0.84-2.63]. | DD: M ~ F  BPD: M ~ F |
| Cherian  2014 | 545  (213) | 30 ± 12  (18+) | OCD  IND | MINI,  YBOCS | **Comorbidity in OCD, F:M OR.** MDD = 1.64 [95%CI, 1.11-2.44], suicidal risk = 2.13 [95%CI, 1.30-3.45], social phobia = 0.41 [95%CI, 0.22-0.78]. | SP: M > F MDD/ Suicide: M < F |
| Torresan  2013 | 858  (504) | 35 ± 12  (18-77) | OCD  BRA | clin interv  YBOCS | **Comorbidity in OCD, F:M OR. Any** = 0.63 [95%CI, 0.39-1.04].  **Any mood** = 0.93 [95%CI, 0.68-1.25], MDD = 0.96 [95%CI, 0.72-1.29], Dysthimia = 1.38 [95%CI, 0.90-2.11], BPD = 0.85 [95%CI, 0.53-1.36].  AUD = 0.31 [95%CI, 0.19-0.53], other drug related = 0.44 [95%CI, 0.17-1.14].  **Any anxiety** = 0.77 [95%CI, 0.57-1.05], panic/ agro = 1.37 [95%CI, 0.97-1.94],  social phobia = 0.69 [95%CI, 0.52-0.91], SPe = 1.62 [95%CI, 1.21-2.18],  PTSD = 1.25 [95%CI, 0.88-1.77], GAD = 0.87 [95%CI, 0.66-1.16], IA = 1.12 [95%CI, 0.54-2.33], Separation anx. = 1.31 [95%CI, 0.96-1.80].  **Any ED** = 1.79 [95%CI, 1.15-2.77], BDD = 0.83 [95%CI, 0.55-1.26], SSD = 3.24 [95%CI, 1.09-9.66], AN = 5.59 [95%CI, 1.66-18.59], BN = 4.18 [95%CI, 1.42-12.08], Binge = 1.07 [95%CI, 0.66-1.75].  **Any ICD** = 1.34 [95%CI, 1.01-1.77], Explosive = 0.87 [95%CI, 0.53-1.41], Klept. = 0.98 [95%CI, 0.43-2.24], Pyromania = 0.23 [95%CI, 0.02-2.25], PG = 0.70 [95%CI, 0.14-3.49], TTM. = 1.80 [95%CI, 1.01-3.22], CB = 2.56 [95%CI, 1.58-4.13], NPSD = 0.24 [95%CI, 0.11-0.52], Internet =1.13 [95%CI, 0.51-2.52], videogame = 0.16 [95%CI, 0.04-0.56], SPD = 2.61 [95%CI, 1.77-3.85], TD = 0.69 [95% CI, 0.52-0.93], TS = 0.72 [95%CI, 0.45-1.17], ADHD = 0.75 [95%CI, 0.51-1.12]. | SPe, **Any ED,** SSD, AN, BN, **Any ICD,** TTM, CB, SPD M < F  Separation anx: M ≤ F  **Any mood,** MDD, Dysthimia, BPD, other drug related, PTSD, GAD, IA, BDD, Binge, Explosive, Klept., Pyromania, PG, Internet addict, TS, ADHD: M ~ F  **Any, Any anxiety,** panic/ agro: M ≥ F  AUD, social phobia, NPSD, videogame addict, TD: M > F |
| Torres  2013 | 955 (555) | 36 ± 12  (18+) | OCD BRA | SCID-I,  YBOCS | **Comorbidity in OCD, F:M OR.** Any = 0.60 [95%CI, 0.36-1.00]; 2+ CC = 3.46 [95%CI, 1.27-9.42]. | Any CC: M > F |
| Al-Asadi  2015 | 13414 (9440) | 35 ± 13  (18-86) | GEN INTL | e-PASS (online survey) | **Comorbidity in OCD, F:M OR.** GAD = 1.95 [95%CI, 1.33-2.85]; SAD = 1.75 [95%CI, 1.18-2.59]; PD+AGO = 1.37 [95%CI, 0.90-2.10]; SPe = 1.58 [95%CI, 1.07-2.33]; PTSD = 2.58 [95%CI, 1.53-4.37]; BDD = 2.9 [95%CI, 1.42-5.93]; PI = 1.66 [95%CI, 1.13-2.46]; PH = 2.0 [95%CI, 0.98- 4.05]; AUD = 0.35 [95%CI, 0.20-0.61]; PG = 0.05 [95%CI, 0.007-0.41]; SD_ST = 0.37 [95%CI, 0.12-1.15]; SD_CAN =0.5 [95%CI, 0.23-1.11]; MDD/ED/Agrph/somat/AN/BN: n.s. | AUD/PG: M > F  MDD/ ED/ Agrph/ somat/ AN/ BN/ SUD: M ~ F  PD+AGO: M ≤ F  GAD/SAD/SPe/PTSD/ BDD/ PI / PH: M < F |
| Pinto  2016 | 21911 (12005) | 34 ± 8  (19-47) | TD SWE | STAGE | **Comorbidity in OCD, F:M OR.** TD = 0.82 [95%CI, 0.48-1.41], ADHD = 0.43 [95%CI, 0.29-0.65], TD + ADHD = 1.57 [95%CI, 0.51-4.86].  **Comorbid OCD, F:M OR.** In TD = 1.66 [95%CI, 1.01-2.71], in ADHD = 0.70 [95%CI, 0.49 – 1.00], in TD + ADHD = 3.64 [95%CI, 1.15-11.53]. | ADHD: M > F  TD/ TD + ADHD:M ~ F  OCD in TD/ OCD in TD+ ADHD: M < F  OCD in ADHD: M ≥F |
| Öcal  2019 | 90  (64) | 27 ± 8.1  (18-52) | OCD ITA | SCID-I, YBOCS | **Comorbidity in OCD, F:M OR.** OC-related (TTM, SPD, BDD, HD) = 0.31 [95%CI, 0.11-0.90] | OC-related: M > F |
| Benatti  2020a | 229 (113) | 41 ± 14  (18+) | OCD ITA | SCID-5-PD | **Comorbidity in OCD, F:M OR.** Any (lifetime) = 2.0 [95%CI, 1.15-3.48]; 2+ CC = 2.38 [95%CI, 0.93-6.09].Anxiety = 1.21 [95%CI, 0.49-2.59]; Mood = 1.22 [95%CI, 0.70-2.11], ED = 1.03 [95%CI, 0.14-7.42], PersD = 2.07 [95%, 0.19-23.18]; OCPD = 1.34 [95%CI,0.66-2.75]; Psychotic = 1.03 [95%CI, 0.14-7.42]; TD = 1.47 [95%CI, 0.45-4.76]; SUD = 0.95 [95%CI, 0.44-2.08]. | Any CC:  M < F  Anx/ Mood/ ED/ PersD/ Psychotic/ TD/SUD: M ~ F |
| Kim  2022 | 398  (218) | 36 ± 11  (18+) | OCD  (USA) | Padua, BDI, BAI | **Comorbidity in OCD, severity.** BAI M=15.29±9.03, F=18.16±10.82 d = 0.08 p > 0.05; BDI M=15.38±8.59, F=17.83±10.51 d = 0.02 p > 0.05 | Anx./Depres: M ~ F |
| Klenfeldt  2024 | 144  (103) | 36 (19-52) →71 (53-97) | OCD  SWE | clinical exam | **Comorbidities in OCD at ~47 year follow up, F:M OR.** Specific phobia (SPe)= 2.22 [95% CI, 1.05–4.71] p<0.05, Panic anxiety disorder = 4.15 [95% CI, 1.88–9.17] p<0.001; AUD = 0.05 [95% CI, 0.01–0.20] p<0.001  *n.s.:* GAD, Social Phobia, AGO, Depression (Any), BPD-I, Dysthymia, Dementia, SZ-related, Delusional disorder, Paranoia, Other SUD | AUD: M > F  GAD, Social Phobia, AGO, Depress, BPD-I Dysthmia, Dementia, SZ, Delusions, Paranoia, SUD: M ~ F SPe/ Panic Anx.: M < F |
| Brakoulias  2025 | 257  (141) | 39 ± NA  (25-62) | OCD  AUS | YBOCS, MINI | **Latent class analysis of comorbidities in OCD, F:M OR.** Cluster 1 (Low comorb) 1.29 [95% CI, 0.75–2.21]; Cluster 2 (MDD, panic, agroph, phobia, PTSD, anx.): 0.97 [95% CI, 0.58–1.62]; Cluster 3 (PTSD): 0.44 [95% CI, 0.18–1.06]; Cluster 4 (MDD/anxiety spectrum, hair pulling): 6.52 [95% CI, 1.87–22.7]; Cluster 5 (BDD): 0.64 [95% CI, 0.25–1.64]; Cluster 6 (AUD): 0.91 [95% CI, 0.30–2.80] | MDD/anxiety spectrum, hair pulling: M < F  PTSD: M > F |
| *Single gender studies* | | | | | | |
| Taherifard  2025 | 102M | NA  15-17 | Stutter  HC  IRA | BASS, SCAS, SR stuttering | **Comorbid OCS.** In adolescent boys with stuttering, relations with other Sx; Kendall’s τ. ANX τ = 0.58, p < 0.001; body image τ = −0.26, p < 0.001; separation anxiety τ = 0.29, p < 0.001; social phobia τ = 0.38, p < 0.001; panic/agoraphobia τ = 0.434, p < 0.001; stuttering severity τ = 0.15, p = 0.045. | M: OCS ↑↑ ANX /stuttering |
| CC more prevalent in women than in men in the general population (M < F) | | | | | | |
| *Anxiety Spectrum Disorders* | | | | | | |
| *adults* |  |  |  |  |  |  |
| Assunção  2012 | 1001 (569) | 35 ± 13  (18+) | OCD  BRA | SCID-I, YBOCS | **Comorbidity in OCD, F:M OR.** Soc ph = 0.73 [95%CI, 0.56-0.95] | Soc ph: M > F |
| Rudy  2014 | 196 (159) | 20 ± 5  (18-66) | OCD INTL | FOCI, SPS, SIAS | **Comorbidity in OCD, F:M OR.** Spec ph = 0.61 [95%CI, 0.18-2.03]. | Spec ph: M ~ F |
| Torres  2014 | 1001 (569) | 35 ± 13  (18+) | OCD BRA | SCID-I, YBOCS | **Comorbidity in OCD, F:M OR.** PD = 1.47 [95%CI, 1.03-2.11], AG = 1.45 [95%CI, 0.80-2.66], PD+AG = 1.52 [95%CI, 1.1-2.1]. | PD/ PD+AG: M < F  AG: M ~ F |
| Sharma  2021 | 867 (504) | 35 ± 13  (18+) | OCD  BRA | SCID-I, YBOCS | **Comorbidity in OCD, F:M OR.** GAD = 0.92 [95%CI, 0.69-1.22] | GAD: M ~ F |
| Citkowska -Kisielewska  2020^s^ | 186 (140) | 34 ±9  (18-58) | GAD  POL | Sx CL “O” (KO) | **Comorbid OCs, F:M OR.** In GAD**:** checking = 0.52 [95%CI, 0.26-1.06], obsess thghts = 0.72 [95%CI, 0.29-1.78], repet. of actions = 0.81 [95%CI, 0.40-1.62], aggress thghts = 0.83 [95%CI, 0.40-1.69], compul. = 0.71 [95%CI, 0.36-1.42], inappr. thghts = 0.53 [95%CI, 0.26-1.10], counting = 1.14 [95%CI, 0.53-2.48], slow compul. = 0.87 [95%CI, 0.41-1.88]. | In GAD: checking/ inappr. thghts: M ≥ F  obsess thghts/ repet./ aggress thghts/ compul./ counting/ slow compul: M ~ F |
| *Depression (DD)* | | | | | | |
| *children/adolescents* | | | | | | |
| Storch  2012 | 160  (70) | 13 ± 3  (7-20) | OCD  USA | ADIS-IV-P, CYBOCS | **Comorbidity in OCD, F:M OR.** DD = 4.1 [95%CI, 1.6-10.5]. | DD: M < F |
| Brown  2015 | 295 (124) | 15 ± 3  (7-18) | OCDF  GBR | CY-BOC, BDI-Y | **Comorbidity in OCD, F:M OR.** DD = 1.94 [95%CI, 0.79-4.76] ^**^ | DD: M ≤ F |
| Riddle  2023 | 797  (583) | 16 ± 3  (8-20) | MDD  USA | MINI-KID, SCARED, PHQ-9 | **Comorbid OCD, F:M OR.** OCD in MDD = 1.58 [95% CI, 1.13–2.20] | OCD in MDD: M < F |
| Fang  2025 | 526 (396) | 15 ± 1 (13–17) | OCD  CHN | SCL-90, SDS, Zung SR Anxiety Scale, Suicide Ideation SR | **Comorbidities in OCD.** Model: OCS → depression / anxiety / sleep disturbance → suicide risk; model × sex: Δχ²=22.69, Δdf=7, p=0.002.  M: OCS→DEP =0.479, p<0.001; OCS→ANX =0.523, p<0.001; OCS→sleep =.321, p<.001; OCS→suicide risk =.187, p=.021; DEP mediation =.251; ANX mediation n.s.; sleep mediation n.s.  F: OCS→DEP =.419, p<.001; OCS→ANX =.475, p<.001; OCS→sleep =.373, p<.001; OCS→suicide risk =.291, p<.001; DEP mediation =.120; ANX mediation n.s.; sleep mediation =.065. | M&F: OCS ↑↑ suicide risk; mediation: depress M>F; anxiety: M~F (n.s.); sleep: F |
| Bai  2026 | 465 (348) | 16 ± 1 (11–18) | MDD GBR | LOI,  DEQ-A | **Comorbid OCS in MDD, OCS ~ MDD Sx, β.** Self-criticism×F = 0.23 p = 0.044.  Self-criticism → OCS: F β = 0.19, p = 0.003; M β = −0.08, n.s.  Dependency → OCS: F β = 0.19, p = 0.004; M β = 0.25, p = 0.038. | self-criticism → OCS F > M  dependency → OCS  M > F |
| a*dults* |  |  |  |  |  |  |
| Quarantini  2011 | 814 (475) | 35 ±13  (18-65) | OCD  BRA | SCID, YBOCS | **Comorbidity in OCD, F:M OR.** Current MDD = 1.11 [95%CI, 0.82-1.50], lifetime MDD = 1.08 [95%CI**,** 0.81-1.46]. | MDD: M ~ F |
| Viswanath 2012 | 545 (213) | 29 ± 11  (18+) | OCD IND | MINI, YBOCS | **Comorbidity in OCD, F:M OR.** DD = 1.69 [95%CI, 1.16-2.44];  controlling for severity and types of obsessions, DD = 1.5 [95%CI, 1.04-2.37]. | DD: M < F |
| Altintas  2015 | 140  (91) | 33 ± 11 (15-65) | OCD  TUR | SCID, YBOCS | **Comorbidity in OCD, F:M OR.** MDD = 1.48 [95%CI, 0.73-3.0]**.** | MDD: M ~ F |
| Hofmeijer-Sevink 2018 | 2125 (1418) | 45 ± 13 (18-65) | NESDA NLD | YASR-OCS, IDS | **Comorbid OCs, F:M OR.** OCs in DD = 0.36 [95%CI, 0.29-0.44]. | OCS in DD: M > F |
| Hathway  2024 | 243  (179) | 33 ± 12  (18-78) | OCD  USA, AUS, EUR | DIAMOND-OCD, PHQ-9 DPSS-R | **Comorbidity in OCD, F:M OR.** MDD = 0.76 [95% CI: 0.42–1.38] | MDD: M ~ F |
| Qi  2025 | 10265  (6554) | NA  18-65+ | MDD | Database Review | **Comorbid OCD, F:M ROR.** In MDD treated with atypical antipsychotics (AAP) = 0.61 [95% CI, 0.48–0.77], p=3.1×10⁻⁵ | OCD in MDD patients taking AAP: M > F |
| Mishra  2025 | 146  (46) | 32 ± 11 (18–60) | OCD IND | YBOCS, OVIS, HDRS | **Comorbid Sx in OCD, severity.** Depression ~ F β = 4.35 [95% CI, 1.60–7.09], p=0.002. | Depression severity in OCD: M < F |
| Wu  2026 | 1293 (730) | 28 ± 9  (17+) | OCD  CHN | YBOCS, OCI-R, BDI-II | **Comorbidity in OCD.** Depressive Symptoms: Depres-: M=62.1%; Depres+: M = 52.1%, OR=0.43 [95%CI, 0.30, 0.62] p<0.001 | Depres in OCD: M < F |
| *Suicidality* | | | | | | |
| *all ages* | | | | | | |
| Chen  2026 | 1614275 (F:61%) | all ages | GEN GAD  TWN | Database Review (NHIRD) | **Comorbid OCD. GAD → suicide risk.** OCD-: M: HR = 2.37 [95% CI 2.08–2.69]; F: HR = 2.52 [95% CI 2.21–2.87]  OCD+: M: HR = 2.54 [95% CI 1.88–3.43], F: HR = 2.66 [95% CI 1.96–3.61] | OCD+ GAD→ suicide risk: M ~ F |
| Lo  2026 | 5262 (2667) | NA  12+ | GEN  CHI | Database Review | **Comorbidities in OCD.** Self-Harm, HR**:** OCD M = 2.22 [95% CI, 1.51-3.33], F = 4.33 [95% CI, 3.3-5.66]; OCD+SUD M= 86.8 [95% CI, 32.5-231.7]; F=164.5 [68.3-396.2] | Self-Harm in OCD:  M ≤ F |
| Mortier  2026 | 645571 (340317) | NA  10+ | GEN  ESP | Database review | **Comorbidities in OCD.** Non-lethal intentional self-harm. M HR= 6.4 [95%CI, 5.6-7.4], PAF=3.1 [95%CI, 2.7-3.5]; F HR=6.0 [95%CI, 5.3-6.9], PAF=2.3 [95%CI, 2.0-2.6].  Suicide. M HR=17.4 [95%CI, 5.3-56.5], PAF=5.7 [95%CI, 3.4-7.9]; F HR=8.9 [95%CI, 4.9-16.1], PAF=2.2 [95%CI, 0.9-3.5].  Lethality Index, %. M = 6.0 [95%CI, 2.1-9.8]; F=3.5 [95%CI, 0.4-6.5].  Suicide, IR, per 100,000 person-years. M = 44.0 [95%CI, 15.1-72.9]; F = 26.5 [95%CI, 3.2-49.6].  Nonlethal intentional self-harm, IR, per 100,000 person-years. M= 693.1 [95%CI, 569.9-816.2]; F=734.8 [95%CI, 604.6-865.1]  Suicide by Age, HR. M: ‘10-19’ NE, ‘20-44’=2.9 [95%CI, 1.4-6.0], ‘45-64’=36.1 [95%CI, 7.3-178.6], ‘65+’=56.9 [95%CI, 7.8-416.7]; F: ‘10-19’=21.6 [95%CI, 2.7-172.9], ‘20-44’=9.9 [95%CI, 4.3-23.0], ‘45-64’= 10.2 [95%CI, 4.1-25.2], ‘65+’ NE. | suicide in OCD:  M > F  M: Age ↑↑ suicide  F: Age ~ suicide |
| *adults* | | | | | | |
| Gupta  2014 | 130  (60) | 32 ± 9  (18+) | OCD IND | SCID-CV, YBOCS, SSI | **Comorbidity in OCD, F:M OR.** SI+DD = 2.9 [95%CI, 1.12-7.49], SI+DD vs SI-DD = 3.1 [95%CI, 1.21-7.97], SI-DD = 0.94 [95%CI, 0.42-2.08]**.** | DD↑↑SI: M < F |
| Chaudhary  2016 | 50  (20) | NA  (18+) | OCD IND | CSSRS, YBOCS | **Comorbidity in OCD, F:M OR.** SI = 3.5 [95%CI, 1.05-11.66], SA = 19.33 [95%CI, 2.18-171.9] | SI/SA: M < F |
| Velloso  2016 | 548 (296) | 34±11  (18+) | OCD  BRA | SCID-I, YBOCS, study-spec | **Comorbidity in OCD, F:M OR.** SI+SA = 1.52 [95%CI, 1.07-2.16] | SI+SA: M < F |
| De la Cruz  2017 | 36788  (20791) | NA  (18+) | OCD  SWE | Registry records | **Comorbidity in OCD, F:M OR.** Death by suicide = 1.45 [95%CI, 1.14-1.84],  Suicide attempt = 11.8 [95%CI, 10.58-13.16]. | Suicide risk: M < F |
| Veisani  2017 | 763 (423) | 38 ± 14  (15-88) | GEN KOR | GHQ-28, SCID-IV | **Comorbidity in Compulsive d/o.** SI: M = 5.13 [95%CI, 2.02-16.25], F = 6.02 [95%CI, 2.08-20.0] | SI ↑↑ Compulsive:  M ~ F |
| Breet  2019 | 496 (257) | 35 ± 12  (18-75) | OCD USA | SCID, YBOCS | **Comorbidity in OCD, F:M OR.** SA = 2.75 [95%CI, 1.28-5.94]**,** controlling for type and severity of OCs. | SA: M < F |
| Demirkol  2019 | 130  (76) | 36 ± 12  (18+) | OCD TUR | BSIS, YBOCS | **Comorbidity in OCD, F:M OR.** SA = 1.14 [95%CI, 0.30-4.28]. | SA: M ~ F |
| Benatti  2020b | 218  (91) | 36 ± 14  (18+) | OCD ITA | SCID-I, II, study-spec | **Comorbidity in OCD, F:M OR.** OCTD+SA vs OCD-SA = 0.44 [95%CI, 0.17-1.12], OCTD+SA vs OCTD-SA = 0.98 [95%CI, 0.37-2.57]; OCD+SA vs OCD-SA = 0.70 [95%CI, 0.27-1.83]. | TD↑↑SA**:** M ≥ F |
| Benatti  2021 | 409 (235) | 39 ± 14  (18+) | OCD  ITA | SCID-I, II, YBOCS, MINI | **Comorbidity in OCD, F:M OR.** Current suicide risk in OCD, F:M OR = 1.44 [95% CI, 0.71–2.92] | Suicide Risk: M~ F |
| Samuels  2023 | 515 (300) | 36 ± 13  (18-75) | OCD+MDD  USA | SCID-IV, SADS-LA-R, YBOCS, SIDP-IV | **Comorbidities in OCD.** SA in OCD+MDD, F:M OR. SA = 1.18 [95% CI, 0.76–1.83]  **Risk factors for SA in OCD+MDD, F:M OR.** Violent/horrific images: M OR = 4.48 [95%CI 1.82-11.0]; F OR = 1.76 [95%CI 0.89-3.47], p < 0.01 | SA in OCD+MDD:  M ~ F; Violent Images in OCD+MDD ↑↑ SA: M > F |
| Chen  2024 | 56977 (28250) | NA  (18+) | OCD  TWN | med records | **Comorbidity in OCD.** Death by Suicide in OCD, HR. M HR=1.86, [95%CI 1.37–2.53], F HR=1.98, [95%CI, 1.38-2.84]  Suicide risk in OCD + Comorbidities, HR. Schizophrenia: M = 3.50 [95%CI, 2.61–4.68]; F =3.37 [95%CI, 2.34–4.83]; Bipolar Disorder: M = 2.71 [95%CI, 1.94–3.79] F = 3.79 [95%CI, 2.67–5.37]; MDD: M = 5.27 [95%CI, 4.27–6.52]; F = 8.47 [95%CI, 6.66–10.78], AUD: M = 2.16 [95%CI, 1.35–3.45] F = 4.12, [95%CI, 2.40–7.06]; SUD M = 2.64 [95%CI, 1.72–4.06]; F = 3.88 [95%CI, 2.41–6.24]; ASD M = 2.26 [95%CI, 1.12–4.58] F *n.s*.; ADHD F = 2.41 [95%CI, 1.02–5.67] | Suicide Risk: M ~ F  Suicide Risk w Comorbidity: for SZ, BPD, MDD, AUD, SUD: M ~ F  ASD: M > F  ADHD: M < F |
| Fang  2025 | 526 (396) | 15 ± 1 (13–17) | OCD  CHN | SCL-90, SDS, Zung SR Anxiety Scale, Suicide Ideation SR | **Comorbidities in OCD.** Model: OCS → depression / anxiety / sleep disturbance → suicide risk; model × sex: Δχ²=22.69, Δdf=7, p=0.002.  M: OCS→DEP =0.479, p<0.001; OCS→ANX =0.523, p<0.001; OCS→sleep =.321, p<.001; OCS→suicide risk =.187, p=.021; DEP mediation =.251; ANX mediation n.s.; sleep mediation n.s.  F: OCS→DEP =.419, p<.001; OCS→ANX =.475, p<.001; OCS→sleep =.373, p<.001; OCS→suicide risk =.291, p<.001; DEP mediation =.120; ANX mediation n.s.; sleep mediation =.065. | M&F: OCS ↑↑ suicide risk; mediation: depress M>F; anxiety: M~F (n.s.); sleep: F |
| Tsai  2025 | 858685 (333115) | NA  (18+) | Panic HC  TWN | med records | **Comorbid OCD as a risk for** **Suicide, OR.** Pooled Panic + OCD = 2.10 [95%CI 1.64-2.67], Panic =1.78 [95%CI 1.58-2.02];  M: Panic + OCD =1.65 [95%CI 1.11-2.43], Panic = 1.63 [95%CI 1.36-1.95];  F: Panic + OCD=2.60 [95%CI 1.9-3.55], Panic =1.91 [95%CI 1.61-2.26], p < 0.05 | OCD in Panic ↑↑ Suicide M < F |
| Wei  2025 | 15776  (6599) | NA  (18+) | SAD  No-SAD  TWN | med records | **Comorbid OCD as a risk for** **Suicide, OR.** Matched to 1:4 Cohort: SAD: M=2.44 [95%C 1.46-4.09], F=4.53 [95%CI 1.98-10.40]; SAD + OCD: M=1.13 [95%CI 0.46-2.78], F=8.49 [95%CI 2.95-24.43],  Matched to 1:10 Cohort: SAD M=2.17 [95%CI 1.47-3.21], F=3.01 [95%CI 1.6-5.66], SAD + OCD M=1.00 [95%CI 0.44-2.28], F=5.07 [95%CI 2.08-12.33] | In F Only: SAD + OCD ↑↑ Suicide |
| Huang  2026 | 114  (49) | 26 ± 11 (18-65) | OCD  CHI | GAD-7, clinical interview, HAMD, blood markers | **Comorbidities in OCD.** Active suicidal ideation (ASI): overall ASI=28%. OCD−ASI: M=60.5%; OCD+ASI: M=50.0%, p=0.320. F:M OR=0.81 [95%CI, 0.38–1.75] p = 0.60.  Serum Estradiol (E2). F: OCD−ASI=191.87 OCD+ASI =314.82 pg/mL, p=0.001; M: OCD−ASI= 96.87 OCD+ASI = 126.46 pg/mL, p=0.004. | ASI in OCD: M ~ F M& F: E2 ↑ ↑ASI |
| *single gender studies* | | | | | | |
| Spinner  2026 | 80259F | 21 ± 4  (18+) | ED  MULT | Database review | **Comorbidity in OCD+ED.** Suicide Ideation, %. OCD+ = 20%, OCD- = 9.3%, χ^2^(1) = 7.67, p < 0.05 | F: ED+OCD↑↑Suicide Ideation |
| *Post-Traumatic Stress Disorder (PTSD)* | | | | | | |
| *adults* | | | | | | |
| Nacasch  2011 | 44  (5) | 44 ±16  (18+) | PTSD  ISR | MINI | **Comorbid OCD, F:M OR.** In PTSD = 0.22 [95%CI, 0.02-2.2] | OCD in PTSD: M ~ F |
| Walter  2022 | 9447 (2496) | active duty 2006-2013 (USA) | | PTSD | **Comorbid OCD.** In PTSD**:** M=1.0%, F=1.4%, χ^2^=2.12, F:M OR=1.36, [95%CI 0.9-2.05] | OCD in PTSD: M ~ F |
| Santoro  2023 | 333 (189) | 26 ± 7 (18-56) | GEN  ITA | TEC, DES-II, SCL-90 | **Comorbid OCS, severity.** In Trauma M=1.22±0.89, F=1.29±0.90, d = 0.08 p=0.49 | OCS in Trauma: M ~F |
| *Eating Disorders* | | | | | | |
| *all ages* | | | | | | |
| *children/adolescents* | | | | | | |
| Riva  2023 | 42  (28) | NA  (12-17) | ED  ITA | EDI-3, SCL-90R | **OCS in Eating d/o:** OCS M > F p=0.012 (stats not given) | For OCs in ED:  M > F |
| *adults* |  |  |  |  |  |  |
| Hübel  2019 | 155 (70183) | 55 ± 8  (18+) | GEN  GRB | SCID  genetic | *“AN-genomic variation associated with OCD in males, but statistical tests did not confirm this.”* | Genomic correlation: *AN ↑↑ OCD: M ≥ F?* |
| Strobel  2019 | 358 (179) | 28 ± 10  (18+) | ED DEU | EDI, SIAB;  CBT | **Treatment response, Cohen’s d.** M = 0.59 p < 0.001, F = 0.37 p < 0.001, Δ OCI-R x M η^2^ = 0.02 p < 0.05. | T ΔOCs severity in AN/BN: M > F |
| Bang  2020 | 392 (327) | 33 ± 9  (18+) | GEN NOR | EDE-Q, OCI-R | **Comorbid OCD, risk.** ED vs HC, OR: F **=** 2.29 [95% CI, 1.22–4.32]; M = 3.18 [95% CI, 0.34–30.10] | F: ED ↑↑ OCs  M: ED ↑↑ OCs |
| Aguilar  2026 | 3120 (2846) | 31±13 (15–83) | ARFID  USA | Database Analysis (OCI-R) | **Comorbid OCS.** In ARFID: OCI-R M=17.4 ± 13.3, F=21.0 ± 14.4, q=0.0002. Washing: M = 2.0 ± 2.9, F=2.3 ± 3.0, F(q-value)=4.01(0.09); Obsessing: M = 3.7±3.7, F=4.7±3.8, q=0.0004); Ordering: M = 4.1±3.4, F=4.7±3.6, q=0.018; Checking: M=2.6±2.7, F=2.7±2.8, q=0.39; Neutralizing: M=1.7±2.5, F=2.3±2.9, q=0.0004; Hoarding: M=3.3±2.9, F=4.2±3.3, q=0.0004. | In ARFID, OCS M < F  wash/check: M ~ F  obsess/neutr/hoard:  M < F |
| Kapadia  2025 | 5927 (5262) | 34± 13  (18+) | ED  USA | OCI-R | **Comorbid OCD/OCS, F vs M vs gen.-diverse.** In ED: OCD+ gender-diverse=79%, M=65%, F=56%, adjusted ps < 0.05; OCS gender-diverse > M > F across multiple ED groups/dimensions, p<0.001. | OCD/OCS in ED: gender-diverse > M > F |
| van Passel  2025 | 382 (216) | 36 ± 10 (18+) | OCD  NLD | SCID-I, YBOCS | **Comorbidity in OCD.** OCD−ED M=48%, F=52%; OCD+ED M = 9%, F=91%; χ²(1) = 25.72, p = 0.011. OCD+ED had higher YBOCS across follow-up, group p = 0.003; group × time p=0.50. | ED in OCD: M < F; OCD + ED severity↑, course~ |
| *Single gender studies* | | | | | | |
| *all ages* | | | | | | |
| Lee  2020 | 112F | 19 ± 6  (12-45) | ED  USA | EDRC, OCI-R | **Comorbid OCs, severity.** OCs severity ↑↑ ED symptoms: r = 0.43 p < 0.001.  **Comorbid OCs, treatment.** Δ OCs severity ↑↑ Δ ED severity: r = 0.41 p < 0.001 | F: OCs ↑↑ED Sx;  Δ OCs ↑↑ Δ ED |
| Barber  2025 | 95F | Adol: 15±1; Adult: 25±8 | ED  USA | Habit Questionnaire EDE-Q, DOCS, | **Comorbid OCS, associations with Sx.** In ED: associations with BFRBs (β, adj. for ED Sx): any BFRB—clinical = 0.04 p = 0.019; full-spectrum = 0.05, p = 0.019; skin picking full-spectrum = 0.06, p < 0.001; mouth chewing clinical = 0.07 p = 0.007, full-spectrum = 0.06 p< 0.001; hair pulling/nail biting n.s. | F: OCS↑ with BFRBs |
| *children/adolescents* | | | | | | |
| Brooks  2014 | 35F | 15 ± 2  (13-17) | ED, HC  ZAF | EDE-Q, OCI-R | **Comorbid OCs, severity.** ED > HC: d = 1.1 p < 0.001. | F: ED ↑↑ OCs |
| Blachno  2016 | 76F | 15 ± 2  (NA) | AN POL | LOI-CV | **Comorbid OCD, presence.** In AN = 46%.  **Comorbid OCD, impact.** Activity level: OCD > no OCD p = 0.07. | F: AN ↑↑ OCD Risk |
| Lewis  2019 | 209F | 16 ± 2  (NA) | ED ISR | SCID, YBOCS | **Comorbid OCs severity, Cohen’s d.** AN-BP > AN-R, d = 0.48 p < 0.01; N-BP > BN, d = 0.40 p < 0.01 | F OCs severity:  AN-BP > AN-R/ BN |
| De Mey  2025 | 79F | 17 ± 2  (12-22) | AN | RMQ, CY-BOCS, MINI | **Comorbid OCD in AN, presence:** OCD = 21.1%  **Comorbid OCs in AN, presence:** OCS = 95%  **Comorbid OCs in AN, severity.** CY-BOCS Score=10.44±10.6 | F OCs ↑ in AN |
| Tyszkiewicz-Nwafor 2021 | 30F | 16 ± 6  (NA) | AN  POL | EAT-26, YBOCS | **Comorbid OCs in AN, severity.** Acute Stage of AN= 11.69 ± 8.36, After partial weight recovery= 7.19 ± 8.51, HC = 3.53 ± 4.24, p=0.048 | OCD ↑ in AN  OCD ↓ AN recovery |
| Collatoni  2025 | 185F | 19 ± 3 →  18 ± 3 | AN  ITA | EDI-1, SCL-90 | **OC symptoms in AN, pre- →post- COVID.** OC symptoms showed greater network centrality (overall connectivity with other symptoms) in post-COVID-onset than pre-COVID-onset AN (p = 0.005); the OC symptom–interoceptive awareness association was present only post-COVID (p = 0.004). | F: OCS–AN network integration: pre- COVID < post COVID |
| Stender  2025 | t1: 313F t2: 217F | 16 ± 2 (12–29) | AN  GER | SIAB-EX, EDI-2, SCL-90-R, BDI-II | **Comorbid OCS.** In AN, T1→T2. OCS: 1.16±0.82→0.78±0.66, p<.001; OCS–DEP .37→.38; OCS–ANX .34→.35; OCS–IA .19→.22; OCS–SI 0→0. | AN F: weight restoration ↓ OCS, OCS ~ DEP/ANX/IA no change |
| *adults* |  |  |  |  |  |  |
| Lavender  2014 | 116F | 25 ± 11  (18+) | AN USA | SCID-I/P,  PANAS-X | **Comorbid OCD, presence.** In AN subtype: Overregulated OR = 5.6 [95%CI, 1.77-17.71]. | F: Overregulated AN ↑↑ OCD |
| Lee  2018a | 103F | 19 ± 6 (12-45) | ED  USA | EDRC, OCI-R | **Comorbid OCs, severity.** OCs ↑↑ ED Sx Pre-treatment: r = 0.43 p < 0.001 | F: OCs ↑↑ ED Sx |
| Bang  2020 | 79F | 23 ± 6  (18-35) | AN, HC  KOR | EDE-Q, OCI-R | **Comorbid OCs severity, Cohen’s d.** AN vs HC: Neutralizing: d = 0.47 p = 0.053, washing/ checking/ obsessing/ ordering/ hoarding/total d < 0.13 p > 0.5 | F: AN ↑↑ Neutralizing |
| Amianto  2022 | 123F | 22 ± 6  (18+) | ED, HC | SCID,  YBOCS | **Comorbid OCs, severity.** *Total:* AN = 21.20±8.5, HC = 0.31±1.21, d = 4.15 p<0.001; AN+OCD = 31.50 ± 4.75, AN = 18.70 ± 7.27, d = 2.08 p<0.001.  *Obsess*.: AN+OCD = 15.63 ± 2.87, AN = 9.82 ± 4.36, d = 1.58, *p* < 0.001.  *Compuls*.: AN+OCD = 15.88 ± 3.31, AN = 8.88 ± 4.20, d = 1.85, *p* < 0.001 | AN+OCD > AN > HC |
| Bevione  2024 | 112F | 23 ± NA (18-65) | AN, HC  ITA | BIS-11, OCI | **Comorbid OCs** ↑↑ **Cognitive Impulsivity in AN, Pearlson r.** OCI total = 0.449, p < 0.001, obsess = 0.594, p < 0.001; mental neutral = 0.402, p = 0.003, hoard = 0.331, p = 0.016; doubting = 0.325, p = 0.018; order = 0.312, p = 0.023; wash = 0.285, p = 0.039; check = 0.284, p = 0.039. | In AN, Cognitive Impulsivity ↑↑OCs |
| Claes  2021 | 429F | 21 ± 6 (14-45) | ED, BEL | SIQ-TR, PI-R | **Comorbid OCs, severity.** In ED + Non-Suicidal Self-Injury (NSSI), severity. Lifetime NSSI > no NSSI on OC symptoms: Wilks’ λ = 0.798, η² = 0.202, p < 0.001. ED subtype overall: Wilks’ λ = 0.958, η² = 0.021, p = 0.058; impulses: AN-BP > AN-R and BN, F(2,422) = 5.65, p < 0.01. | F: OCs in ED NSSI > no NSSI; OC impulses in ED: AN-BP > AN-R/BN |
| Halls  2023 | 134F | 19 ± 3 →21± 3  (12-27) | AN  GBR | EDE-Q, OCI | **Comorbid OCs in AN, severity.** *Baseline:* AN = 20.15±13.56, HC = 8.45±10.52, d = 0.96. Follow up: AN =20.56±13.56, HC =8.45±10.52, d = 1.00. | OCI Scores: AN > HC |
| Kambanis  2022 | 91F | 18 ± 4  (10-23) | ARFID,  AN, USA | KSADS-PL, PARDI,  EDE-Q | **Comorbid OCD in AN and ARFID, presence.** ARFID=53%, AN=65%, OR=1.04 [95%CI, 0.39-2.8] p=0.938. | Comorbid OCD:  ARFID ~ AN |
| Wilson  2025 | 138F | 23±6  (18–44) | GEN  CAN | SAM, FSQ, OCI-R, EDE-Q | **Comorbid OCS and ED Sx.** SAM–OCS r = 0.58, p < 0.001; EDE-Q–OCS: r = 0.42, p<001. Direct effects on OCS, β: SAM = 0.20 p = 0.03; FSQ-COR = 0.35 p < 0.001; FSQ-CUL/FSQ-MAL n.s. Direct effects on ED symptoms, β: SAM = 0.03 p = 0.02; FSQ-MAL = 0.14 p< 0.001; FSQ-COR/FSQ-CUL n.s | F: OCS ↑↑ ED Sx |
| Elm  2026 | 17F | 30 ± 5  (18-34) | GEN AN  SWE | OCI-R | **Comorbid OCS.** In An **(**AN subdivided into weight restored and non-weight restored; WRAN): OCI-R AN=24 vs HC=10, p=0.010; non-WRAN=25 vs CT=10, p=0.048; WRAN=23 vs non-WRAN=25, p=0.69. | F: AN ↑↑ OCS |
| *Compulsive Buying* | | | | | | |
| *adults* | | | | | | |
| De Mattos 2016 | 171 (151) | 40 ± 13  (18+) | CBD BRA | CBMS, ICD-SCID | **Comorbid OCD, F:M OR.** In CBD = 0.49 [95%CI, 0.16-1.47] | OCD in CBD:  M ~ F |
| Kim  2018 | 993 (563) | 36 ± 13  (18+) | OCD BRA | YBOCS, SCID-IV | **Comorbidity in OCD, F:M OR.** CBD = 2.8 [95%CI, 1.60-4.87] | CBD: M < F |
| *Trichotillomania (TTM), Skin Picking Disorder (SPD), Hair Pooling Disorder (HPD), and Grooming disorders (GD)* | | | | | | |
| *all ages* | | | | | | |
| Gerstenblith 2019 | 2606 (1553) | NA  (5-50+) | OCD- fam  USA | SADS-LA, YBOCS, CYBOCS | **Prevalence in OCD-affected families, F:M OR.** TTM = 1.46 [95%CI, 1.01-2.13] | TTM in OCD-fam:  M < F |
| *adults* | | | | | | |
| Lovato  2012 | 901  (477) | 34 ± 13  (18+) | OCD BRA | SCID-I, YBOCS | **Comorbidity in OCD, F:M OR.** GD = 2.27 [95%CI, 1.49-3.53]; ICD = 0.85 [95%CI, 0.58-1.29]. | GD: M < F  ICD: M ~ F |
| Grant  2016 | 462 (435) | 33 ± 14  (18+) | TTM USA | SCID-I, MGH-HPS | **Comorbid OCD, F:M OR.** In TTM = 0.85 [95%CI, 0.33-2.17] | OCD in TTM: M ~ F |
| CC similarly prevalent in women and men in the general population (M ~ F) | | | | | | |
| *Bipolar Disorder (BPD)* | | | | | | |
| *adults* |  |  |  |  |  |  |
| Saunders  2012 | 736 (460) | 43 ± 12  (18+) | BPD-I USA | DIGS, med records | **Comorbid OCD, F:M OR.** In BPD-I = 2.44 [95%CI, 1.16-5.16].  **Comorbid OCD, characteristics.** Age-at-onset M < F d = 0.33 p < 0.05. | OCD in BPD-I: M < F; onset: M < F |
| Kim  2014 | 174 (106) | 43 ± 13  (18+) | BPD-I  AUS | MINI | **Comorbid OCD, F:M OR.** In BPD-I = 1.66 [95%CI, 0.65-4.26] | OCD in BPD-I: M ~F |
| Kazhungil  2017 | 90  (61) | 40 ± 12  (18+) | BPD-I  IND | SCID-I, YBOCS | **Comorbid OCD, F:M OR.** In BPD-I = 1.7 [95%CI, 0.65-4.26] | OCD in BPD-I: M ~F |
| Di Salvo  2020 | 990 (573) | 49 ± 16  (18+) | BPD ITA | Med records | **Comorbid OCD, F:M OR.** In BPD-I, II, NOS = 0.61 [95%CI, 0.45-0.84]. | OCD in BPD-I, II, NOS: M > F |
| Putz  2025 | 340 (169) | NA  (18-82) | BPD  AUT | SCID, medical records | **Comorbid OCD in BPD.** F:M OR = 2.24, 95% CI = [95% CI, 2.12–41.33], *p* = 0.003 | OCD in BPD: M < F |
| *Body Dysmorphic Disorders (BDD)* | | | | | | |
| *children/adolescents* | | | | | | |
| Schneider  2019 | 162  (69) | 15 ± 1  (NA) | GEN US | BDDQ-A, BIQ-C | **Comorbid OCD, F:M OR.** In BDD = 0.73 [95%CI, 0.37-1.41] | OCD in BDD: M ~ F |
| Rautio  2022 | 172 (136) | 16 ± 2 (10-19) | BDD GBR/ SWE | BDD-YBOCS | **Comorbid OCD, F:M OR.** In BDD = 2.72 [95%CI, 0.15-50.43] | OCD in BDD: M ~ F |
| Racz  2023 | 107  (57) | 12 ± 3  (7-17) | OCD  AUS | ADIS-IV-P  BDDQ-A, CY-BOCS | **Comorbidity in OCD, F:M OR.** BDD = 0.87 [95% CI: 0.41–1.86] | BDD in OCD: M ~F |
| *adults* | | | | | | |
| Costa  2012 | 901 (516) | 34 ± 13  (18+) | OCD BRA | DY-BOCs, SCID-I/P | **Comorbidity in OCD, F:M OR.** BDD = 0.94 [95%CI, 0.63-1.41] | BDD: F ~ M |
| Çınaroğlu  2025 | 71M | 27±5  (18+) | BDD/MD, TUR | OCI-R | **Comorbid OCS, OCI-R.** In BDD/muscle dysmorphia: BDD/MD = 29.2 ± 7.0, HC = 16.9 ± 6.4, p < 0.001, d=1.83. | M: OCS ↑↑ BDD/MD |
| *Single sex* | | | | | | |
| Monzani  2012 | 2148F | 60 ±13  (NA) | Twins  GBR | OCI-R, DCQ, DNA | **Polychoric correlations for OCs and BPD Sx presence**: 0.34 [95%CI, 0.29-0.39], MZ=0.12 [95%CI, 0.15-0.27]; DZ=0.14 [95%CI, 0.05-0.22] | F: Genetic risk for OCs ↑↑gen risk for BPD |
| Enander  2018 | 15377F | NA  (15-28) | Twins  SWE | DCQ, A-TAC, OCI-R | **OCD Impact, F:M OR.** Risk for BDD, age 15 **=** 2.32 [95%CI, 0.3–17.6]; age 18 = 0, ages 20-28 = 7.2 [95%CI, 3.6–14.2]. | F: OCD ↑↑ risk for BDD for age 20-28. |
| *Hoarding Disorder (HD)* | | | | | | |
| *adults* | | | | | | |
| Torres  2012 | 1001 (569) | 35 ± 13  (18+) | OCD BRA | SCID-I,  YBOCS | **Comorbidity in OCD, F:M OR.** HD Sx = 1.41 [95%CI, 1.09-1.81] | HD Sx: M < F |
| Mathews  2014 | 15,377 (9,658) | 41 ± 16  (NA) | Twin NLD | HRS-SR, PI-ABBR | **Comorbidity in OCD, F:M OR.** HD = 0.52 [95%CI, 0.37-0.73]  **Comorbid OCD, F:M OR.** In HD = 0.59 [95%CI, 0.42-0.83] | HD/OCD in HD:  M > F |
| Cath  2017 | 15194  (9714) | NA  (15-97) | Twin NLD | PI-R ABBR | **Comorbidity in OCD, F:M OR.** Provisional HD = 2.13, z = −2.87, p = 0.004. | HD Sx: M < F |
| Chen  2017 | 894  (601) | 40 ± NA  (18-89) | OCD USA | SCID-IV, YBOCS, PBI | **Comorbidity in OCD, F:M OR.** HD = 0.95 [95%CI, 0.71-1.26]  **Comorbidity in OCD, features, Cohen’s d.** HD x Mother control**:** M = 0.02 [95%CI, -0.26 – 0.22] F = 0.20 [95%CI, 0.04-0.37]. | HD: M ~ F  Mother control ↑↑ OCD+HD: M < F |
| Samuels  2018 | 448 (265) | 37 ± 12  (18-75) | OCD USA | SCID-IV, BRIEF-A, HRS-I | **Comorbidity in OCD, F:M OR.** HD = 1.31 [95%CI, 0.86-1.99].  **Comorbidity in OCD, features.** HD x executive function impairment: Initiate/plan/organize/ metacognition F > M d > 0.33 p < 0.05 | HD: M ~ F  HD x Exec fun: M < F |
| Boerema  2019 | 407 (227) | 38 ± 13  (18-79) | OCD NLD | SCID-I/P, YBOCS | **Comorbidity in OCD, F:M OR.** HD = 0.92 [95%CI, 0.56-1.70]. | HD: M ~ F |
| de Mattos  2025 | 1910 (1184) | 40±15 (18–88) | OCD  USA | YBOCS, FOCI | **Comorbidities in OCD.** OCD-pHD: F=63.4%, M=35.6%; OCD+pHD: F=59.3%, M=40.1%; χ²=3.38, p=0.066 | Hoarding in OCD:  M ~ F |
| *Personality Disorders* | | | | | | |
| *adults* | | | | | | |
| Lochner  2011 | 403 (204) | 35 ± 12 (18-75) | OCD  ZAF | SCID-II, YBOCS | **Comorbidity in OCD, F:M OR.** OCPD = 1.2 [95%CI, 0.61-2.35]. | OCPD: M ~ F |
| Silberschmidt  2015 | 770 (559) | 32 ± 11  (18-65) | BPD  Multi | SCID-I, SCL-90-R | **Comorbid OCD, F:M OR.** In BPD =5 [95%CI, 0.28-88.6] | OCD in BPD: M ~ F |
| Girone  2026 | 300 (138) | 38 ± 15  (18+) | OCD  ITA | SCID-5-PD | **Comorbidity in OCD.** OCPD: OCD only: M=54.2%, F=45.8%; OCD+OCPD: M = 56.9%, F = 43.1%; χ² = 1.76, p = 0.184. | OCPD in OCD: M~F |
| Żerdziński  2025 | 78  (42) | 45 ± NA  (18+) | OCD  POL | DSM-5 criteria | **Comorbidity in OCD.** OCPD: M vs F χ^2^ = 0.55 p = 0.46 | OCPD in OCD: M~F |
| CC more prevalent in men than in women in the general population (M > F) | | | | | | |
| *Schizophrenia (SZ)* | | | | | | |
| *all ages* | | | | | | |
| Chen  2023 | 35255  (17368) | 35 ± 15 (10+) | OCD  TWN | med records | **Comorbid OCD, impact.** OCD ↑↑Risk for SZ: M HR: 1.23 [95% CI 1.12–1.35] | SZ in OCD: M > F |
| *adults* | | | | | | |
| Huang  2011 | 44  (19) | 30 ± 14  (18+) | OCD  CHI | YBOCS, ICD-10 | **Comorbidity in OCD, F:M OR.** Schizotypal traits (ST) = 0.43 [95%CI, 0.11-1.76]; SZ Spectrum disorders (SS) = 0.09 [95%CI, 0.01-0.82]. | ST: M ≥ F  SS: M > F |
| Faragian  2012 | 246  (68) | 31 ± 9  (18+) | SZ ISR | SCID-I/P, YBOCS | **Comorbid OCD, F:M OR.** In SZ = 1.06 [95%CI, 0.61-1.86]**.** | OCD in SZ: M ~ F |
| Bosanac  2013 | 1825  (730) | 38 ± 11  (18-65) | SZ  AUS | DIP, MINI, SCID-I | **Comorbid OCD, F:M OR.** In SZ = 1.23 [95%CI, 0.94-1.60]. | OCD in SZ: M ≤ F |
| Devi  2015 | 200  (96) | 33 ± 10  (18+) | SZ  IND | SCID-I, II, YBOCS | **Comorbid OCD, F:M OR.** In SZ = 0.22 [95%CI, 0.12-0.43]**.** | OCD in SZ: M > F |
| Bener  2018 | 396 (230) | 43 ± 14 (18-65) | SZ QAT | WMH-CIDI | **Comorbid OCD, F:M OR.** In SZ = 1.23 [95%CI, 0.77-1.95] | OCD in SZ: M ~ F |
| Cheng  2019 | 10045 (5135) | NA  ~20 - ~40 | OCD CHI | LHID med record | **Comorbid OCD, impact.** OCD ↑↑Risk for SZ: HR = 30.17 [95%CI, 18.88-50.90]; F:M HR = 0.63 [95%CI, 0.45-0.90]. | OCD ↑↑Risk for SZ: M > F |
| Baek  2020 | 320 (155) | 33 ± 9  (18+) | SZ  KOR | clin interv, YBOCS | **Comorbid OCD, F:M OR.** In SZ = 0.49 [95%CI, 0.28-0.87]. | OCD in SZ: M > F |
| Kokurcan  2020 | 300  (90) | 38 ± 10  (18+) | SZ  TUR | DIGS-K, SDS, clin interv | **Comorbid OCD, F:M OR.** OCD in SZ **=** 1.02[95%CI, 0.52-1.99], subOCD in SZ = 0.71 [95%CI, 0.49-2.91]. | OCD/subOCD in SZ: M ~ F |
| Jelsma  2025 | 527  (138) | 33 ± 10  (16-50) | SZ | RAS-25, YBOCS | **Comorbid OCD, F:M OR.** OCD in SZ = 1.10 [95%CI, 0.77–1.58] | OCD in SZ: M ~ F |
| Shoaib  2023 | 139  (63) | 32± 9  (18+) | SZ  PAK | med records | **Comorbid OCs, F:M OR.** OCs in SZ = 0.73 [95%CI, 0.43–1.25], p = 0.365 | OCD in SZ: M~ F |
| Taha  2024 | 174  (58) | NA  (18-60) | SZ  EGY | MCQ-30, YASR-OCS | **Comorbid OCs, severity:** M=19.8 ± 8.7, F=18.3 ± 9.4, d = 0.17, p=0.29 | OCS in SZ: M ~ F |
| Temircan  2021 | 78  (31) | 37 ± 12  (18-65) | SZ  TUR | SCID-I | **Comorbid OCD in SZ, F:M OR.** OCD in SZ = 0.16 [95% CI, 0.06–0.40], χ² = 12.38, p < 0.001 | OCD in SZ: M ~ F |
| Park  2021 | 3875 (1785) | 31 ± 12  (15-60) | SZ  KOR | med records | **Risk for comorbid OCD in SZ.** Clozapine: M = 3.07 [95%CI, 1.47-6.43]; F = 2.46 [95%CI, 1.01-5.99] | Clozapine SZ↑↑ OCD: M~F |
| *Psychotic Disorder* | | | | | | |
| *children/adolescents* | | | | | | |
| Borelli  2023 | 52  (31) | 16 ± 2  (18+) | OCD  ITA | COPER, COGDIS, CY-BOCS, SPI-CY | **Comorbidity in OCD, F:M OR.** Psychosis vulnerability in OCD = 1.10 [95% CI, 0.37–3.29], p = 0.54 | Psychosis in OCD: M~F |
| Borelli  2025 | 340 (176) | 13 ± 3  (7–17) | OCD  NOR  USA | CBCL | **Risk for comorbidity in OCD.** Psychosis, above vs below cutoff: M vs F χ² = 0.93, p = 0.33. | risk for psychosis in OCD: M ~ F |
| *adults* | | | | | | |
| Hagen  2013 | 246 (106) | 23 ± 8  (18+) | PSY  NOR | SCID-I | **Comorbid OCD, F:M OR.** In **1^st^** Ep Psychosis = 1.93 [95%CI, 0.85-4.40] | In **1^st^** Ep Psychosis  M ~ F |
| Sullivan  2018 | 93,483 (53,659) | 51 ± 22  (18+) | PSY UK | med record | **Comorbid OCs, PPV.** In psychosis, by age: **≤24:** M=10 [95%CI, 4.6-22.5], F=NR, **25-34:** M=8.0 [95%CI, 4.0-16.1], F=2.9 [95%CI, 1.4-6.0], **35-44:** M=7.2 [95%CI, 3.2-16.2], F=4.1 [95%CI, 2.3-7.2], **45-54:** M=9.2 [95%CI, 3.7-22.7], F=7.3 [95%CI, 3.9-13.7], **55+:** M=17.0 [95%CI, 6.6-43.7], F=8.3 [95%CI, 4.6-15.0] | In psychosis: M > F |
| Shaker  2025 | 100  (43) | 18 – 55 | OCD  EGY | PANSS | **Comorbid Sx in OCD.** OCD−PF: M=63.3%, F=36.7%; OCD+PF: M=60.0%, F=40.0%; χ² = 0.071, p = 0.791. | Psychotic Sx in OCD: M ~ F |
| West  2025 | 192  (69) | 18 ± 4  (NA) | CHR  USA | med record  SIPS, FOCI | **Comorbid OCD in clinical high risk for psychosis (CHR):** OCD in CHR = 0.88 [95% CI, 0.58–1.33], p = 0.89 | OCD in CHR: M ~ F |
| Autism (ASD) | | | | | | |
| *children/adolescents* | | | | | | |
| Margari  2019 | 159  (59) | 11 ± 5  (NA) | ASD ITA | med record | **Comorbid OCD, F:M OR.** In ASD = 0.15 [95%CI, 0.01-2.69] | In ASD: M ≥ F |
| Martin  2020 | 7922 (2157) | NA  (4-17) | OCD/ASD  GBR | med record | **Comorbidity in OCD, F:M OR.** ASD = 0.53 [95%CI, 0.41-0.68]  **Comorbid OCD, F:M OR.** In ASD = 1.72 [95%CI, 1.37-2.17] | ASD: M > F  In ASD: M < F |
| Rødgaard  2021 | 16126  (4212) | NA  (0-15) | ASD  DEN | med record | **Comorbid OCD, F:M OR.** In ASD = 1.39 [95%CI, 1.20–1.64]. | OCD in ASD: M < F |
| O’Connor  2025 | 111  (50) | 15 ± 2  (NA) | ASD, IRL | retrospective chart review | **Comorbid OC-related disorders.** In ASD M = 23.6%, F = 6.0%, FTM = 0%; M vs F χ² = 5.22 p = 0.02. | OCD in ASD: M > F |
| Ozcan  2025 | 484 (240) | 15 ± 2  (5 – 18) | OCD  GBR | SCQ, SDQ, ChOCI-R | **Comorbidity in OCD.** ASD: OCD-ASD F = 55%, OCD+ASD F = 41.3%), p=0.004 | OCD + ASD: M > F |
| *adults* |  |  |  |  |  |  |
| Mito  2014 | 81  (53) | 28 ± 9  (18+) | OCD JPN | SCID-IV, YBOCS, AQ | **Comorbidity in OCD, F:M OR.** ASD = 0.58 [95%CI, 0.22-1.49] | ASD: M ≥ F |
| Tic Disorders (TD) and Tourette’s Syndrome (TS) | | | | | | |
| *all ages* | | | | | | |
| Brander  2021 | 4063135 (1973849) | NA  (6-46) | GEN  SWE | med records | **Comorbidity in OCD, F:M OR.** Tic-related OCD vs non-tic OCD:  M = 70.96% vs 40.94%; F:M OR = 0.28 [95%CI, 0.25–0.31] | TD: M > F |
| *children/adolescents* | | | | | | |
| Nikolajsen  2011 | 83  (45) | 14± 3  (6-19) | OCD  DNK | med record  CY-BOCS | **Comorbidity in OCD, F:M OR.** TD = 0.17 [95%CI, 0.04-0.88]. | TD: M > F |
| Højgaard  2016 | 269 (132) | 13 ± 3  (7-17) | OCD  NOR/DEN/  SWE | YBOCS, K-SADS-PL | **Comorbidity in OCD, F:M OR.** TD = 0.45 [95%CI, 1.29-3.77] | TD: M > F |
| Groth  2017 | 314  (57) | w1: 12 ± 3  (5-20)  w2: 19 ± 3 (11-26) | TS  DEN | YGTSS, CYBOCS | **Comorbid OCD, F vs M.** w1 /w2 OCD x F: p > 0.45. stats not reported  **Comorbid OCD, severity.** In TS: F x Δ OCs with age: total p = 0.33, obsess p = 0.63, compuls. p = 0.21; stats not reported. | OCD in TD: M ~ F |
| Groth  2018 | 314 (57) | w1: 12 ± 3  (5-20)  w2: 19 ± 3 (11-26) | TS  DEN | YGTSS, CYBOCS | **Comorbid OCs, course.** *“The clinical course did not differ significantly between the sexes (p > 0.05) in any of the analyses.”* (stats not reported) | Δ **OCs** with age in TS: M ~ F |
| Greenberg 2018 | 811  (183) | 19 ± 14  NA | TS USA | TICS  YBOCS | **Comorbid OCD, F:M OR.** In TD = 0.86 [95%CI, 0.62-1.2] | OCD in TD: M ~ F |
| Girgis  2022 | 270  (58) | 10 ± 3  (5-17) | TS  CAN | YGTSS, CY-BOCS | **Comorbid OCD, F:M OR.** TS=1.06, [95%CI, 0.44-2.55], p=0.13 | OCD in TS: M ~ F |
| Hakimi  2022 | 851859  (419139) | 11 ± 4  (5-18) | Multi  USA | med records | **Comorbidity in OCD, F:M OR.** OCD + TD ≈ 0.59 [95%CI 0.42–0.83]; OCD + ADHD ≈ 0.45 [95%CI 0.39–0.52]; OCD + TD + ADHD≈0.20 [95%CI 0.15–0.27] | TD, ADHD, TD + ADHD in OCD: M > F |
| Larsh  2023 | 132  (49) | 16 ± 2  (13–17) | TD  USA | CY-BOCS, mini-CTIM | **Comorbid OCs, features.** *OCs severity:* M = 7.9 ± 6.5, F = 6.8 ± 5.1, d = 0.19, p = 0.8. *Parent-reported non-tic impairment ↑↑ OCs,* *Spearman’s ρ*: F = 0.57, p = 0.024; M = 0.05, p = 0.93. | OCs in TD: M ~ F  non-tic impairment ↑↑ OCs: F |
| Openneer  2022 | 187 (101) | 7 ± 2  (3-10) | GEN  EUR | YGTSS  CY-BOCS | **Comorbid OCs, impact.** OCs as risk for Tic onset: F: Compulsions β = −0.39, explained deviance = 2.65%; obsessions β = −0.01, explained deviance = 0.01%  M: Obsessions β = 0, deviance = 0%; compulsions β = 0.02, deviance = 0.11% | F: Compuls. → Tic Onset |
| Chang  2026 | 39930 (8529) | 13±11  (18+) | GEN  TWN | Database Review | **Risk for comorbid OCD.** In TS: aHR = 5.18 [95%CI, 3.86-6.95] p < 0.001  M:F aHR = 1.24 [95%CI, 0.87-1.78] p = 0.24 | OCD in TS: M ~ F |
| *adults* |  |  |  |  |  |  |
| Rodgers  2013 | 80  (34) | NA  (33-66) | TD CHE | DIGS, EPQ-R | **Comorbid OCs, subtype.** In TD: Class ↑OCs F:M OR = 17.34 [95%CI 0.92-325] | ↑OCs in TD subtype: M < F |
| Vries  2016 | 377 (213) | 36 ± 11  (18+) | OCD NLD | SCID-IV, YBOCS | **Comorbidity in OCD, F:M OR.** TD = 0.64 [95%CI, 0.41-1.00]. | TD: M > F |
| Ekinci  2020 | 210 (138) | 29 ± 10  (18+) | OCD TUR | SCID-I, SCID-II | **Comorbidity in OCD, F:M OR.** TD = **0.18 [95%CI, 0.08-0.43].** | TD: M > F |
| Mukai  2021 | 44  (20) | 32 ± 10  (18+) | GEN  JPN | YBOCS,  YGTSS | **Comorbidity in OCD, F:M OR.** TD ≈ 1.14 [95% CI, 0.24–5.50] | TD M ~ F |
| Dy-Hollins 2024 | 2109 (537) | 21 ± 14  (18+) | TD  USA | YGTSS, YBOCS | **Comorbid OCD, F:M OR.** TD = 1.29 [95%CI, 1.08–1.54], p = 0.01  **Comorbid OCD, features.** Age of OCD onset in TD**:** M = 7.3 ±3.8 yrs, F = 8.1 ± 5.5 yrs; d = 0.17, p = 0.06  **Comorbid OCs, severity.** M = 19.4±9.3, F = 20.3±9.3; d = 0.10, p = 0.40 | OCD in TD: M < F  OCD Onset in TD:  Trend M < F  OCs Severity: M ~ F |
| Nilles  2025 | 227  (82) | 31 ± NA  (18+) | TD  FRA | Registry Review | **Comorbid OCD, F:M OR.** In TS= 2.51 [95%CI, 1.27-4.95], p=0.008; M=14.3%, F=29.5%. | OCD in TS: M < F |
| Śmilowska  2026 | 2594 (NA) | all ages | TD  POL | NHF database | **Comorbid OCD, F vs M.** In TS, M:F TS+OCD <18=2–3:1; ≥18 ≈ 2:1. TS-OCD <18=5–6:1; ≥18 ≈ 6:1. Ratio of M:F prevalence ratios: TS+OCD:TS−OCD M:F ratio ≈0.45 (<18), 0.33 (≥18). | OCD+TS M:F < OCD−TS M:F |
| Externalizing Disorders – Attention Deficit Hyperactivity Disorder (ADHD) and Oppositional Defiant  Disorder (ODD) | | | | | | |
| *children/adolescents* | | | | | | |
| Inci  2019 | 1000 (242) | 11 ± 3  (6-18) | ADHD  TUR | K-SADS-PL | **Comorbid OCD, F:M OR.** In ADHD = 1.78 [95%CI, 1.07 – 2.97]. | OCD in ADHD:  M < F |
| Efe  2022b | 364  (194) | 14 ± 3  (8-18) | OCD  TUR | K-SADS-PL, CYBOCS | **Comorbidity in OCD, F:M OR.** ADHD ≈ 0.39 [95% CI, 0.25–0.61], χ²(1)=18, p<0.001 | ADHD in OCD:  M > F |
| De Rossi  2022 | 714 (108) | 9 ± 3  (NA) | ADHD  ITA | CBCL-OC | **Comorbid OCs, severity.** M=61.44±9.87, F=59.08±9.65, d = 0.24 p=0.03 | OCs Severity in ADHD: M > F |
| Zhang  2026 | 1249 (405) | 10 ± 1  (9 – 10) | ADHD  USA | MRI, KSADS | **Comorbid OCD, F vs M.** In ADHD *“No significant effect of sex on any data analyzed.”* | OCD in ADHD M ~ F |
| *adults* |  |  |  |  |  |  |
| Blanco-Viera 2018 | 955 (555) | 33 ± NA  (18-82) | OCD BRA | SCID‐I | **Comorbidity in OCD, F:M OR.** ADHD = 0.8 [95%CI, 0.55-1.16]. | ADHD: M ~ F |
| Ozaydin  2025 | 159  (NA) | 36 ± 14  (18 – 65) | OCD, TUR | SCID-IV | **Comorbidity in OCD, F:M OR.** ADHD = 3.73 p=0.031 | ADHD in OCD:  M > F |
| Substance Use and Addiction | | | | | | |
| *children/adolescents* | | | | | | |
| Harford  2015 | 19430 (9574) | NA  (12-17) | GEN USA | NHSDA 2000 | **AUD ↑↑ risk OCD, OR.** M = 1.6 [95%CI, 1.3-2.08], F = 2.5 [95%CI, 2.03-2.99].  **Comorbid OCD, F:M OR.** In AUD = 2.2 [95%CI, 1.66-2.98]. | AUD↑↑OCD risk: M < F  OCD in AUD: M < F |
| Fernández- Artamendi 2021 | 684 (317) | 17 ± 1  (14-18) | GEN ESP | ACID-IV, BSI | **Comorbid OCD, F:M OR.** In AUD = 1.8 [95%CI, 0.87-3.55], in CUD = 6.9 [95%CI, 1.82-26.27]. | OCD in AUD: M ~ F  OCD in CUD: M < F |
| Hansen  2025 | 68301  (40807) | 18 ± 1  (15 – 19) | GEN  DEN | DNYS, registers | **Comorbidity in OCD, F vs M.** AUD: AUD+OCD: M=43%, F=57%; AUD−OCD: M=37%, F=63%; χ² = 2.19, p = 0.14. | AUD in OCD: M~F |
| *adults* |  |  |  |  |  |  |
| Blom  2011 | 7076 (3777) | 41 ± NA  (18-64) | GEN NLD | NEMESISCIDI | **OCD ↑↑ risk SUD, OR.** M=3.32 [95%CI, 1.67-6.61], F=7.52 [95%CI, 3.11-18.18].  **Comorbid OCD, F:M OR.** In SUD = 0.96 [95%CI, 0.58-1.6] | OCD ↑↑ risk SUD:  M ≤ F  OCD in SUD: M ~ F |
| Dell’Osso  2015 | 483 (290) | 43 ± 13  (18+) | OCD ITA | YBOCS, clin interv | **Comorbidity in OCD, F:M OR.** Current = 1.3 [95%CI, 0.81- 2.04]; Former = 0.3 [95%CI, 0.21- 0.55] | Smoking current: M ~ F  Former: M < F |
| Fuss  2019 | 539 (279) | 35 ± 12  (18-75) | OCD DEU | SCID-OCSD, YBOCS, CSBD | **Comorbidity in OCD, F:M OR.** Lifetime: Any = 0.26 [95% CI, 0.18-0.39], CSBD **=** 0.27 [95%CI, 0.11- 0.63], Pyromania = 0.13 [95%CI, 0.01- 1.90]; Kleptomania = 0.11 [95%CI, 0.29- 1.14], IED = 0.36 [95%CI, 0.23-0.57], PG = 0.08 [95%CI, 0.005 – 1.51]. Current: Any = 0.78 [95% CI,0.47 – 1.30], CSBD **=** 0.26 [95%CI, 0.08 - 0.77], Pyromania = 0.31 [95%CI, 0.01 – 7.63]; Kleptomania = 3.8 [95%CI, 0.80- 18.10], IED = 0.93 [95%CI, 0.49 – 1.77], PG = NA. | lifetime:  any imps: M > F  current:  kleptoman: M < F, pyroman/IED: M ~ F CSBD: M > F |
| Potembska  2019 | 692 (485) | 21 ± NA  (18-25) | GEN POL | IAT, “O” Sx CL | **Effect of OCD on Risk for addiction, Cohen’s d:** M = 0.66 p < 0.001; F = 0.93 p < 0.001; M vs F = 0.28 p > 0.5. | OCD ↑↑ risk Internet Addiction**:** M ~ F |
| Machado  2022 | 115  (20) | medn = 26  (NA) | PIU  BRA | SPQ, MINI, | **Comorbid OCD, F:M OR.** *In Problematic Internet use (PIU):* = 4.36 [95%CI, 1.54–12.34], p = 0.016 | OCD in PIU M < F |
| Benatti  2023 | 70  (40) | 37 ± 14  (18+) | OCD  ITA | study specific | **Comorbidity in OCD, F:M OR.** *Cannabis Use:* Current = 4.09 [95% CI, 0.79–21.08], Prior = 1.02 [95% CI, 0.33–3.12] | Cannabis Use in OCD: M ~ F |
| Jacobs  2025 | 92757  (58314) | 20 ± 2  (18-24) | GEN  USA | ACHA-NCHA III, ASSIST | **Comorbidity in OCD, aOR.** *Tobacco*: F = 1.12 [95% CI, 1.03-1.22], M = 0.99 [95% CI, 0.82-1.21], Trans/Non-Conforming (TGNC) = 1.24 [95% CI, 1.05-1.48]; F:M OR =1.13 [0.83–1.54]. *Cannabis:* F = 1.13 [95% CI, 1.06=1.23], M = 0.97 [95% CI, 0.81-1.17], TGNC=1.12 [95% CI, 0.95-1.33]; F: M OR = 1.16 [0.83–1.62]. *Alcohol:* F= 1.18 [95% CI, 1.08-1.29], M = 1.07 [95% CI, 0.86-1.33], TGNC = 1.03 [95% CI, 0.81-1.32]; F: M OR = 0.99 [0.73–1.34]. *ED*: F = 2.35 [95% CI, 1.93-2.83], M = 2.35 [95% CI, 1.93-2.83], TGNC = 2.14 [95% CI, 1.85-2.47]; F:M OR = 0.98 [0.81–1.19]. | Tobacco, Cannabis, Alcohol /ED:  M ~ F ~ TGNC |
| Virtanen  2022 | 6304188  (3082748) | 31  18+ | GEN  SWE | ICD-10 | **Comorbidity in OCD, F:M OR.** Any substance misuse = 0.81 [95% CI, 0.80–0.82], Alcohol-related= 0.62 [95% CI, 0.61–0.63]; Acute intoxication = 0.90 [95% CI, 0.89–0.92]; Any Drug-related disorder= 0.96 [95% CI, 0.95–0.97]; Opioid = 0.91 [95% CI, 0.88–0.94]; Cannabis = 0.34 [95% CI, 0.33–0.35]; Sedative = 1.24 [95% CI, 1.22–1.26]; Stimulants = 0.76 [95% CI, 0.74–0.79]; Other substances = 0.75 [95% CI, 0.74–0.77]; Substance-related convictions = 0.26 [95% CI, 0.25–0.27]; Substance-related deaths = 0.38 [95% CI, 0.35–0.41]. | SUD in OCD, SUD convictions/ deaths: M > F;  Sedatives: M < F |
| du Mortier  2025 | 419 (235) | 37 ± 11 (18 – 79),  2 yrs f/u | OCD  NLD | YBOCS alcohol/ drug-use SR | **Effects of comorbidity in OCD.** Alcohol use → OCD Severity (2yrs later): F: β = -2.3, [95% CI, -4.52− -0.09] d = -0.25 p = 0.04; M: β = [95% CI, 1.02− 4.06], d = 0.16 p = 0.24.  OCD Severity → Drug use (2yrs later): F: OR = 0.91 [95% CI, 0.87−0.95] p <0.001; M: OR = 1.0 [95% CI, 0.99−1.08], p = 0.09 | F: alcohol use→later OCS↓; OCS↑→later drug use↓  M: n.s. |
| Samuels  2026 | 1222 (764) | NA  18-82 | OCD  FIN | SADS-LA-R, YBOCS, SCID-IV | **Comorbid AUD in OCD.** AUD+: M=43%, F=57%; AUD−: M=37%, F=63%; χ²=2.19, p=0.14. | AUD in OCD: M ~ F |
| Willis  2026 | 656  (445) | 18-29  24 ± 3 | GEN  USA | CAS, DOCS AAQ-II | **COVID-related anxiety (COV-A) and OCS.** Gender → COV-A: β=0.121, SE=0.043, p=0.005; COV-A → OCS: β=0.471, SE=.033, p<0.001 | COV-A: M < F  COV-A↑↑ OCS |
| *Single gender studies* | | | | | | |
| Semple  2011 | 245MSM | 40 ± 8 (18-61) | HIV+  USA | OCI-R,  clin interv | **Comorbid OCD, impact.** Methamphetamine Use (Days): OCD Cohen’s d = 0.35 p = 0.019. | OCD ↑↑ meth use. |
| Wölfling  2013 | 129M | 31 ± 11 (16-65) | IA  DEU | AICA-S, SCL-90-R | **Comorbidity in OCD**, **impact**. OCs severity: IA vs. noIA: Cohen’s d = 0.62 p < 0.001 | IA ↑↑ OCs severity |

A.4.2. Studies of gender effects on general medical conditions in individuals with OCD.

| **First Auth, Year** | | **N**  **(F)** | | **Age mean**±**SD (range)** | **Pop** | **Meas.** | **Gender effects (F vs. M)** | |
| --- | --- | --- | --- | --- | --- | --- | --- | --- |
|  |  |  |  |  |  |  | **Stats** | **Direction** |
| *all ages* | | | | | | | | |
| Isomura  2016 | | 25415  (9303) | | 22 ± 13  (6+) | OCD  (SWE) | Medical registry | **Presence of OCD.** Vascular compl.**:** F HR = 1.37 [95%CI=1.45‒1.54]; M HR = 1.37 [95%CI=1.32‒1.42]; Type 2 diabetes: F HR = 1.3s1 [95%CI=1.19‒1.44]; M HR = 1.11 [95%CI=0.99‒1.23]; Dyslipidemia: F HR = 1.08 [95%CI=1.01‒1.15]; M HR = 0.99 [95%CI=0.92‒1.06]; Obesity: F HR = 1.64 [95%CI=1.52‒1.77]; M HR = 1.51 [95%CI=1.32‒1.73]. | metabolic M < F  Obesity/ vascular  M ~ F |
| Hsu  2026 | 765455 (382,385) | | | 12+ | FDRs of PD, TWN | NHIRD/ database review | **Risk of OCD in FDRs of PD.** overall aRR = 1.73 [1.58–1.90]; M = 1.56 [1.37–1.77]; F = 1.95 [1.70–2.23] | Risk of OCD in FDRs of PD: M ~ F |
| *In children/adolescents* | | | | | | | | |
| Ivarsson  2015 | 269  (NA) | | | NA  (7-17) | OCD  NOR | K-SADS-PL, CBCL | **Severity in OCD.** F-M Cohen’s d: Sleep problems = 0.07 p = 0.63. | Sleep problems: M ~ F |
| Nabinger de Diaz 2019 | 96  (53) | | | 12 ± 3  (7-17) | OCD AUS | ADIS-IV-P, CYBOCS, CBCL | **Presence in OCD.** Parental report: Overtired = 1.07 [95%CI, 0.45-2.50], Sleeps less = [95%CI, 0.45-2.35], Sleeps more = [95%CI, 0.41-4.48], Sleep Talks or walks = [95%CI, 0.54-4.23], Has trouble sleeping = [95%CI, 0.62-3.28]. Self-report: ‘*I have trouble sleeping’* = 1.59 [95%CI, 0.70-3.58], ‘*I sleep next to someone*’ = 1.51 [95%CI, 0.66-3.42]. | Sleep problems: M ~ F |
| Williams  2019 | 206  (99) | | | 11 ± 3  (4-18) | OCD,  USA | EHR | **Presence in OCD of IgA deficiency, OR**. vs. TS: M=1.89 [95%CI, 0.97-4.99], F = 0.64 [95%CI, 0.24-1.69]; vs. ADHD: M = 1.94 [95%CI, 1.07-3.52], F = 0.81 [95%CI, 0.38-1.74]; vs. ASD: M = 2.75 [95%CI, 1.23-4.28], F = 1.55 [95%CI, 0.62-3.85]; vs. anx: M = 2.13 [95%CI, 1.19-3.81], F = 1.79 [95%CI, 0.87-3.30] | IgA deficiency↑↑ OCD: M > F |
| Zahed  2020 | 399 (194) | | | 8 ± 4  (5-12) | cancer pts  IRN | CSI-4 | **Presence of OCD in cancer patients:** M = 0.208 F = 0.211 (2^nd^ highest after SPe), F:M OR = 0.89 [95%CI, 0.60-1.33]. | Cancer: M ~ F |
| Wang  2021 | 78  (38) | | | 15-17 | GEN/OCD  CHN | OCI-CV, MINI, YBOCS | **Presence in OCD of facial changes.** Lower lip height: M OCD=7.86 ± 1.83, HC=9.12 ± 1.44, p=0.020, F OCD=7.51 ± 1.56, HC=7.94 ± 1.39, p=0.379, Nasolabial Angle: M OCD=94.82 ± 10.06, M HC=93.50 ± 10.59, p=0.716, F OCD=92.56 ± 8.35, F HC=101.03 ± 11.48, p=0.013 | In M Only OCD=Lip height ↓  In F Only OCD= Nasolabial Angle ↓ |
| Westwell-Roper  2022 | 112  (56) | | | 14 ± 3  (7-20) | OCD | VSS, CY-BOCS, COIS-R | **Presence in OCD of Bowel/Bladder Sx**. F = 9.8 ± 5., M= 8.2 ± 5.2, *p* = 0.0117  VSS < 11 %F=43.6%, VSS>11 %F=64.7%, OR=1.5, p=0.040 | Bowel/Bladder Sx in OCD:  M < F |
| Herdi  2024 | 348  (195) | | | 14.34±1.56  (12-18) | MIS  TUR | MOCI, MCL, CPRS | **Severity of OCs in Misophonia, Pearlson r.** M = 0.19 p < 0.05, F= 0.42, p < 0.01; Fisher r-to-z = 2.32, p = 0.02. | OCD ↑↑ Misophonia: M < F |
| DeWitt  2025 | 121  (69) | | | 12 ± NA  (<18) | CCS | Database Review | **Presence of OCD in cancer survivors.** Female sex RR= 1.39, [95%CI 1.17–1.61] | OCD in CCS: M < F |
| Jari  2025 | 1284 (683) | | 11±2  (8–15) | | GEN  IRA | SCAS  Beighton | **Comorbid OCD.** In generalized joint hypermobility (GJH, Beighton ≥6) vs non-GJH controls (CT, Beighton <6), OCD: F+GJH = 5.6%, F-GJH = 4.9% OR=1.15 p > 0.05; M+GJH = 7.1% M-GJH =7.5% OCD OR = 0.94, p > 0.05. | Comorbid OCD in GJH, M ~ F |
| *In adults* | | | | | | | | |
| Hamed  2013 | 295 (133) | | | 32 ± 6  (20-49) | EPI EGY | MINI, YBOCS | **Presence of OCs/OCD in idiopathic epilepsy, F:M OR.** OCs = 1.24 [95%CI, 0.74-2.07]. OCD = 1.16 [95%CI, 0.61-2.21]. | OCs/ OCD in Epilepsy: M ~ F |
| Goulart  2014 | 15105  (5864) | | | NA  (35-74) | MIG  BRA | HS-2004, CIS-R | **Presence of OCD in migraine, PPV by severity.** 1x/Month: M = 3.29 [95%CI, 0.77-14.10], F = 2.25 [95%CI, 1.14-4.43]; 1x/Month-1x/Week: M = 5.57 [95%CI, 1.93-16.08], F = 3.24 [95%CI, 2.05-5.13]; 2-6x Week: M = 2.33 [95%CI, 0.31-17.63], F = 4.7 [95%CI, 2.83-7.81]; Daily: M = 29.86 [95%CI, 4.66-191.43], F = 3.2 [95%CI, 1.39-7.38]. | OCD **↑↑** MIG 1x/Month: M ~ F  1-6x Week: M ≥ F  Daily: M > F |
| Latvala  2016 | 1039443M | | | 18→ up to 45 f/u | GEN  (SWE) | Nationwide registries | **Longitudinal.** 10 unit increase in rHR at 18 ~ OCD at > 18: HR = 1.16 [95%CI, 1.12-1.19]; 10 unit increase in BP at 18~ OCD at > 18: systolic HR = 1.01 [95%CI, 1.00-1.02], diastolic HR = 1.06 [95%CI, 1.05-1.07]. | heart rate/blood pressure at 18 ↑↑ OCD at >18 |
| Aguglia  2017 | 162  (66) | | | 36 ± 14 (18+) | OCD ITA | SCID,  YBOCS | **Presence of general medical conditions in OCD, F:M OR.** GMC= 3.65 [95%CI, 1.88-7.06]. | **GMCs**: M < F |
| Husky  2018 | 17,237  (10,262) | | | (18+) | GEN FRA | CIDI-SF, BMI | **Presence of assess weight in OCD, PPV.** M = 0.76 [95%CI, 0.48-1.21], F = 1.51 [95%CI, 1.12-2.02], Obesity x gender p = 0.15. | Weight gain: M ≤ F |
| Ghorayeb 2019 | 105  (62) | | | 61 ± 15  (18+) | RLS  FRA | ASRS, MOCI | **Presence of OCD in restless leg syndrome.** F:M OR **=** 1.19 [95%CI, 0.40-3.55]. | OCD in RLS: M ~ F |
| Wang  2019 | 378990  (126406) | | | NA  (25 - 50+) | SAD  TWN | NHIP Review | **Presence of OCD in systemic autoimmune disorder, HR.** M = 1.43 [95%CI, 0.82-2.47], F = 2.15 [95%CI, 1.57-2.95], F x HR p = 0.13. | OCD in SAD: M ≤ F |
| Westwell-Roper 2019 | 1401  (790) | | | 28 ± 14  (6-82) | OCD  USA | SCID-I,  SADS-LA-R | **Presence in OCD of Rheumatic fever, %.** M=0.0 [95%CI, 0.0—0.5], F = 1.0 [95%CI, 0.5-2.1], p=0.011; NS when corrected for multiple comparisons. | RF: M ≤ F |
| Kilicaslan 2020 | 130  (49) | | | 34 ± 10  (18+) | EPI-OCD  TUR | SCID-CV, OCI-R | **Presence of OCD in temporal lobe epilepsy.** F:M OR = 0.85 [95%CI, 0.38-1.89]. | In TLE: M ~ F |
| Saadat  2020 | 125  (59) | | | 68 ± 10  (NA) | PrkD IRN | SLC-25 | **Severity of OCs in Parkinson’s Disorder.** M v F Cohen’s d = 0.18 p = 0.31. | OCs in PrkD: M ~ F |
| Chang  2021 | 5034  (2515) | | | 37 ± 17  (18+) | OCD/GEN  TWN | Database Review (NHIRD) | **Presence in OCD of Sjögren’s syndrome, HR.** F = 2.84 [95% CI: 1.35–5.94], M = HR = 5.24 [95% CI: 1.39–19.73]. p = 0.43 | SS ↑↑OCD: M ~ F |
| Chen  2021 | 14817  (2250) | | | 56 ± 9  (45+) | OCD  TWN | Database Review (NHIRD) | **Presence in OCD of Dementia, HR.** Alzheimer’s Disease**:** M = 6.43; [95%CI 1.42–29.13]; Vascular Dementia: F = 4.76; [95%CI 1.54–14.74], Unspecified Dementia**:** M = 3.85 [95%CI 1.96-7.58], F = 5.46 [95%CI = 2.85 – 10.50] | OCD ↑↑ dementia  M: OCD ↑↑ AD  F: OCD ↑↑vasc.dem |
| Viticchi  2022 | 60  (40) | | | 50 ± 11  (18+) | MIG  ITA | OCI-R | **Presence of OCD in migraine.** At T0, with migraine: Pathologic OCD: M=60%, F=22.5%, p=0.016, Borderline OCD: M=5%, F=27.5%, Normal: M=25%, F=37.5%; p=0.016  At T1 (post OBT-A sessions): Pathologic OCD: M=40%, F=15%, Borderline: M=5%, F=12.5%, Normal M=30%, F=60%, p=0.041 | Pathological OCD in MIG: M > F  OCI-R ↓ in both M,F with treatment of MIG |
| Li  2022 | 80  (58) | | | medn =15  (12-18) | JIA  (CAN) | RCADS | **Presence of OCD in juvenile idiopathic arthritis.** F = 6.9% M = 0% [95%CI, 0.18–76.9], p = 0.57 | OCs ↑↑ rheumatic arthritis: M ~ F |
| Nováková  2023 | 167  (119) | | | 44 12  (18+) | FMD  (CZE) | OCI-R, S-FMDRS | **Severity of OCs in functional movement disorders (FMD).** OCI-R ≥ 21: F=49/70, OCI-R<21: F=70/127, p=0.90 | In FMD, OCD Severity: M~ F |
| Yin  2023 | 1497949  (617747) | | | NA  (0-25) | GEN  (SWE) | Database review | **Risk of parental OCD on TIDM in kids:** Maternal OCD RR 1.62 [1.02–2.58], paternal OCD – n.s. | If Mother has OCD, T1DM ↑ in offspring |
| Chen  2024 | 148929  (75450) | | | NA | GEN  (EUR) | Mendelian Randomization of GWAS | **Presence of Infertility in OCD. M:** MR Egger =1.43 [95%CI, 0.56–3.66] p = 0.46; Weighted median=0.90 [95%CI, 0.75–1.08) p=0.26; Inverse variance weighted = 0.82 [95%CI, 0.72–0.94] p = 0.005  **F:** MR Egger = 1.02(0.91–1.13), p=0.76; Weighted median = 1.00 [95%CI, 0.94–1.06], p = 0.87; Inverse variance weighted = 1.0 [95%CI, 0.95–1.05] p=0.98 | M: OCD ↓↓ Infertility  F: No effect |
| Holmberg  2024 | 496  (391) | | | 36±12.5  (18-79) | OCD  (INT) | Survey | **Presence in OCD.** Any: F=74.4%, M=57.6%, χ2 =10.51, p=0.005; gastroint.: F=30.2%, M=15.2%, χ2 = 9.37, p=0.009; cardiometab.: F=18.2%, M=28.3%, χ2 = 8.08, p=0.018; hypertension: F=9.2%, M=23.9%, χ2 =17.37, p<0.001, migraine: F=18.9%, M=7.6%, χ2 =6.86, p=0.032; autoimmune: F=10.5%, M=1.1%, χ2 =8.84, p=0.012, Other: F=20.2%, M=9.8%, χ2 =6.45, p=0.040.  Allergies, cholesterol, diabetes, cardiovascular, Previous acute, vascular, musculoskeletal, Respiratory, Neurological, Cancer, Obesity: n.s. | Any Somatic M<F  Gastrointestinal, Migraine, Autoimmune, Other M < F  cardiometabolic, Hypertension: M > F |
| İnci  2024 | 524  (321) | | | 60 ± 12  (18+) | GEN  (TUR) | MOCI | **Severity of OCs in cancer patients.** M = 21.77±10.26, F=26.81±7.44, p<0.001. | OCD in Cancer Patients M < F |
| Jüres  2024 | 328  (141) | | | 32 ± 11  (18-65) | GEN  (DEU) | SCID, OCI-R, | **Presence in OCD of elevated heart rate (HR).** HR ~ OCD: 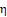^2^_p_ = 0.03 p = 0.02; *M*_Diff_ = 3.55, [95% CI, 0.50-6.60]*.* HR ~ OCD x F: 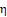^2^_p_ = 0.002 p = 0.44*.* | HR↑↑OCD: M ~ F |
| Mazurie  2024 | 386  (240) | | | NA  (18+) | RLS  (FRA) | *MOCI* | **Presence of OCs in restless leg syndrome, %.** M = 0.15; F = 0.18; p = 0.13.  **Severity of OCs in restless leg syndrome.** M = 2 ± 4; F = 2 ± 4; p = 0.26. | OCs ↑↑ RLS: M~F |
| Bhatnagar 2025 | 216794  (98232) | | | 4 ± 5  (0-21) | CP  USA | ICD-10-CM, | **Presence of OCD in Cerebral Palsy.** F:M OR = 0.83 [95% CI: 0.65–1.08] | OCD in CP: M ~ F |
| Grotteschi  2025 | 70  (28) | | | 40 ± 12  (21 – 74) | Down  Synd.  ITA | neuro. exam DSM-5 | **Presence of OCD.** In Down Syndrome: OCD = 11/70; M = 21.4%, F = 7.1%, p > 0.05 | OCD in Down: M ~ F |
| Kang  2025 | 88341 (47970) | | | 20+ | IID  GEN  KOR | Database  Review | **Presence of OCD.** In adults with recurrent intestinal infectious disease (IID):  OCD HR: M = 1.98 [95%CI, 1.33−2.93], F = 1.65 [95%CI, 1.20−2.29]. | OCD in IID:  M ~ F |
| Karahan  2025 | 200 (169) | | | 47 ± 11  (18+) | FIBRO  TUR | SCL-90-R | **Presence of OCS.** In Fibromyalgia (FIBRO): M = 0.84 ± 0.7, F= 0.80 ± 0.7, p = 0.68 | OCS in Fibromyalgia: M ~ F |
| Sun  2025 | 154  (82) | | | 37 ± 8  (18+) | OCD  GEN  TUR | YBOCS, ASEX | **Presence of OCS.** In Sexual Dysfunction (SD): M SD+ = 19 ± 40.4, SD- =14 ± 56; F SD+ = 28 ± 59.6, SD- = 11 ± 44, p = 0.31. | OCS in SDs: M ~ F |
| Weinstock  2025 | 1111 (915) | | | 50 ± 16  (18 – 85) | MCAS CT  USA | MCMRS, health survey | **Presence of OCD.** In Mast Cell Activation Syndrome (MCAS): F MCAS+ = 25.5%, MCAS- = 4.8%, OR = 6.1 [95%CI, 3.8–10.3], p < 0.001; M MCAS+ = 31.6%, MCAS- = 4.9%, OR = 8.0 [95%CI, 3.2–22.2] p < 0.001. | OCD↑↑MCAS; M ~ F |
| Ma  2026 | 4484 (919) | | | NA  18-23 | GEN  CHN | SCL-90, BMI | **Presence in OCS.** Overweight/obesity: M β = 0.26 p = 0.20 OR=1.29 [95%CI, 0.871-1.909]; F β =-0.72 p = 0.35 OR=0.49 [95%CI, 0.11-2.21] | Weight status–OCS: n.s. in M&F |
| *Single gender studies* | | | | | | | | |
| Rezaeimehr  2022 | 128F | | | 45 ± NA  (21 - 74) | LUTS  (IRN) | AUASS | **Presence in OCD:** Lower Urinary Tract Symptoms: Duration of LUTS in OCD vs GEN 10.7 vs 5.2, p=0.006 [95%CI 1.049-1.214], Stress Urinary Incontinence OCD vs GEN 57.8% vs 23.4%, p<0.0001 [95%CI 0.077-0.539], Bladder Outlet Obstruction (BOO) p=0.006 [95%CI 1.538-12.781], Functional Bladder Score p=0.038 [95%CI 0.994-1.000] | LUTS in OCD:  Onset Time ↓↓  Duration ↑↑  SUI ↑↑  BOO ↑↑  Bladder Score ↓↓ |
| Perrotta 2024 | 208F | | | 39 ± 15  (18+) | OCD (ITA) | Medical  Records | **Presence of OCD.** In secondary anorgasmia, OCD-primary vs OCD-secondary. PSM-Q profiles did not differ between groups, all p > 0.05. | F in secondary anorgasmia, OCD-primary = OCD-secondary |
| Zhou  2024 | 78061F | | | NA  (18+) | GEN  (FIN) | Medical records | **Presence of OCD.** Genetically Predicted Female Genital Prolapse OR = 0.98 [95%CI, 0.80-1.20] p = 0.84 | no relation between OCD and FGP |
| He  2026 | 325M | | | NA  (0–15) | DMD USA | Medical records, genetic testing | **Presence of OCD.** In Duchenne muscular dystrophy (DMD): Distal vs proximal DMD variants: OR=1.60 [95%CI, 0.46–5.58]; very distal exons 63–79 vs 45–62: OR=4.97 [95%CI, 0.87–28.36]. | OCD: exons 63–79 ≥ exons 45–62 |

**Abbreviations used in Table A4.**

**Instruments: ADIS-P/ ADIS-IV-P** **/ ADIS-C / ADIS-CP** = Anxiety Disorders Interview Schedule for Parent version /DSM-IV Parent versions/ Child version/ Child-and-Parent version;^1-3^ AICA-S=Assessment of Internet and Computer Game Addiction;^4^ **AQ** = Autism-Spectrum Quotient;^5^ **ASSIST** = Alcohol, Smoking and Substance Involvement Screening Test;^6^ **ASSQ** = Autism Spectrum Screening Questionnaire;^7^ **A-TAC** = Autism–Tics, ADHD and Other Comorbidities Inventory;^8^ ATQ=Adult Temperament Questionnaire;^9^ **AUASS** = American Urological Association Symptom Score/Index;^10^ AUDIT=Alcohol Use Disorders Identification Test;^11^ BABS=Brown Assessment of Beliefs Scale;^12^ BAI=Beck Anxiety Index;^13^ BCQ=Body Checking Questionnaire;^14^ **BDDQ-A** = Body Dysmorphic Disorder Questionnaire–Adolescent Version;^15^ **BDD-YBOCS** = Yale-Brown Obsessive Compulsive Scale Modified for Body Dysmorphic Disorder;^16^ BDI=Beck Depression Index;^13, 17^ **BDI-II** = Beck Depression Inventory-II;^18^ **BDI-Y** = Beck Depression Inventory for Youth;^19^ BI-AAQ=Body Image Acceptance and Action Questionnaire;^20^ **BIQ-C** = Body Image Questionnaire–Child and Adolescent Version;^21^ BIS=Barrat Impulsiveness Scale;^22^ BMAS=Bergen Social Media Addiction Scale;^23^ BFRB=Body-Focused Repetitive Behaviors Questionnaire;^24^ BRIEF-A=Behavior Rating Inventory of Executive Function;^25^ BSAS= Bergen Shopping Addiction Scale;^26^ BSI=Brief Symptoms Index;^27^ BSIS=Beck Scale for Suicide Ideation;^28^ BCL=Child Behavior Checklist;^29^ **CBCL/ CBCL-OC** = Child Behavior Checklist/ Obsessive-Compulsive scale;^29, 30^ CBMS=Compulsive Buying Measurement Scale;^31^ CDI=Children’s Depression Inventory;^32^ CGI-S=Clinical Global Illness – Severity;^33^ **ChOCI-R** = Children’s Obsessional Compulsive Inventory–Revised;^34^ CIA=Clinical Impairment Assessment;^35^ **CIDI-SF** = Composite International Diagnostic Interview–Short Form;^36^ **CIS-R** = Clinical Interview Schedule–Revised;^37^ COIS-C/P/-R=Child OCD Impact Scale-Child Report and Parent Report/Revised;^38, 39^ **COPER / COGDIS** = Cognitive-Perceptive Basic Symptoms / Cognitive Disturbances criteria;^40^ **CPRS / CPRS-R-S** = Conners’ Parent Rating Scale / Conners’ Parent Rating Scale–Revised, Short Form;^41^ CPQ-A-S=Cannabis Problems Questionnaire-Short Form;^42^ CSSRS=Columbia Suicide Severity Rating Scale;^43^ **mini-CTIM** = Mini-Child Tourette Syndrome Impairment Scale;^44^ CTQ=Childhood Trauma Questionnaire;^45^ **CY-BOCS** = Children’s Yale-Brown Obsessive Compulsive Scale;^46^ DAWBA=Development and Wellbeing Assessment;^47^ **DCQ** = Dysmorphic Concern Questionnaire;^48^ DEBQ=Dutch Eating Behavior Questionnaire;^49^ **DEQ-A** = Depressive Experiences Questionnaire for Adolescents;^50^ **DES-II** = Dissociative Experiences Scale-II;^51^ **DIAMOND**/**OCD module** = Diagnostic Interview for Anxiety, Mood, and OCD and Related Neuropsychiatric Disorders;^52^ DIGS/DIGS-K=Diagnostic Interview for Genetic Studies;^53^ **DIP** = Diagnostic Interview for Psychosis;^54^ **DOCS** = Dimensional Obsessive-Compulsive Scale;^55^ **DPSS-R** = Disgust Propensity and Sensitivity Scale–Revised;^56^ DSED=Distrust of the Senses in Eating Disorders Scale;^57^ **EAT-26** = Eating Attitudes Test-26;^58^ EDDS=Eating Disorder Diagnosis Scale;^59^ EDE-Q: Eating Disorder Examination-Questionnaire;^60, 61^ EDI=Eating Disorder Inventory;^62^ EMCDDA=European School Survey Project on Alcohol and Other Drugs Student Questionnaire;^63^ **EPQ-R** = Eysenck Personality Questionnaire–Revised;^64^ FAS- Family Accommodation Scale;^65^ FMPS=Frost Multidimensional Perfectionism Scale;^66^ FOCI= Florida Obsessive Compulsive Inventory;^67^ FRS=Stunkard Figure Rating Scale;^68^ FSFI=Female Sexual Function Index;^69^ **FSQ** = Fear of Self Questionnaire;^70^ **GAD-7** = Generalized Anxiety Disorder-7;^71^ GAS=Game Addiction Scale;^72^ **GHQ-28** = General Health Questionnaire-28;^73^ HADS=Hospital Anxiety and Depression Scale;^74^ HAM-A=Hamilton Anxiety Rating Scale;^75^ HAM-D=Hamilton Depression Rating Scale;^76^ HRS-I=Hoarding Rating Scale Interview;^77^ HRS-SR=Hoarding Rating Scale-Self Report;^78^ IAT=Youngs 20-item Internet Addiction Test;^79^ **IDS** = Inventory of Depressive Symptomatology;^80^ IIEF=International Index of Erectile Function;^81^ K-SADS-PL=Kiddle Schedule for Affective Disorders and Schizophrenia for School Age-Children-Present and Lifetime Version;^82^ LOI-CV=Leyton Obsessional Inventory-Child Version;^83^ MASC=Multidimensional Anxiety Scale for Children;^84^ **MBSRQ/BASS** = Multidimensional Body-Self Relations Questionnaire/ Body Areas Satisfaction Subscale;^85^ **MCQ-30** = Metacognitions Questionnaire-30;^86^ **MGH-HPS** = Massachusetts General Hospital Hairpulling Scale;^87^ MINI=Mini International Neuropsychiatric Interview;^88^ **MINI-KID** = Mini International Neuropsychiatric Interview for Children and Adolescents;^89^ Mini-IPIP=Mini-International Personality Item Pool;^90^ MOCI=Maudsley obsessional-compulsive inventory;^91, 92^ **MOODS-SR** = Mood Spectrum Self-Report;^93^ NEO-PI/NEO-FFI=Five Factor Model of personality (Neuroticism, Extraversion, Openness, Agreeableness, and Conscientiousness);^94, 95^ NI-MH-GOCS=National Institute of Mental Health Global Obsessive-Compulsive Scale;^96^ **OBS-SR** = Obsessive-Compulsive Spectrum Self-Report;^97^ OCD-NHQ=OCD Natural History Questionnaire;^98^ **OCI-R** = Obsessive-Compulsive Inventory–Revised;^99^ **OCI-CV** = Obsessive-Compulsive Inventory–Child Version;^100^ **OVIS** = Overvalued Ideas Scale;^101^ **PANAS-X** = Positive and Negative Affect Schedule–Expanded Form;^102^ **PANSS** = Positive and Negative Syndrome Scale;^103^ **PARDI** = Pica, ARFID, and Rumination Disorder Interview;^104^ PBI=Parental Bonding Instrument;^105^ PI-R=Padua Inventory-Revised/–Abbreviated;^106^ **PHQ-9** = Patient Health Questionnaire-9;^107^ PS=Psychache Scale;^108^ PTQ=Parent Tic Questionnaire;^109^ PUTS=Premonitory Urge for Tics Scale;^110^ RAPI=Rutgers Alcohol Problem Index;^111^ **RAS-25** = Recovery Assessment Scale-25;^112^ **RCADS** = Revised Child Anxiety and Depression Scale;^113^ **RMQ** = Readiness and Motivation Questionnaire;^114^ **SADS-LA/** **SADS-LA-R** = Schedule for Affective Disorders and Schizophrenia–Lifetime Anxiety version/Revised;^115^ **SAM** = Self-Ambivalence Measure;^116^ **SCARED** = Screen for Child Anxiety Related Emotional Disorders;^117^ **SCAS** = Spence Children’s Anxiety Scale;^118^ SCID-I=Structured Clinical interview for DSM-IV-TR Axis Disorders;^119^ **SCL-90 / SCL-90-R** = Symptom Checklist-90 / Symptom Checklist-90-Revised/ **GSI**=General Symptomatic Index, a component of the Brief Symptom Inventory (BSI) and the Symptom Checklist-90 (SCL-90);^120^ **SCQ** = Social Communication Questionnaire;^121^ **SDQ** = Strengths and Difficulties Questionnaire;^122^ SDS=Schedule for Deficit Syndrome;^123^ **SIAB / SIAB-EX** = Structured Interview for Anorexic and Bulimic Disorders / Expert Rating;^124^ SIAS=Social Interaction Anxiety Scale;^125^ **SIDP-IV** = Structured Interview for DSM-IV Personality;^126^ **SIPS** = Structured Interview for Psychosis-Risk Syndromes;^127^ SIQ-TR=Self-Injury Questionnaire-Treatment Related;^128^ SOCS= Short OCD Screener;^129^ **SPI-CY** = Schizophrenia Proneness Instrument–Child and Youth Version;^40^ SPQ=Shorter PROMIS Questions;^130^ SPS=Social Phobia Scale;^131^ SPSQ=Skin Picking Stanford Questionnaire;^132^ SSAI=Spielberger State Anxiety Inventory;^133^ **SSI** = Scale for Suicide Ideation;^28^ STAGE=Swedish Twin Study of Adults: Genes and Environments;^134^ **TEC** = Traumatic Experiences Checklist;^135^ USP-SPS=Sensory Phenomena Scale;^136^ WMH-CIDI=World Mental Health Composite International Diagnostic Interview;^36^ **YASR-OCS** = Young Adult Self-Report Obsessive-Compulsive Symptoms subscale;^137^ **YBOCS** = Yale-Brown Obsessive Compulsive Scale;^138, 139^. YDQ=Young Diagnostic Questionnaire;^140^ Y-GTSS – Yale Global Tic Severity Scale;^141^ YRBSSQ=Youth Risk Behavior Surveillance System Questionnaire.^142^

**Comorbidities:** **ADHD** = Attention-Deficit/Hyperactivity Disorder; **Ago** = Agoraphobia; **AN** = Anorexia Nervosa; **AN-BP** = Anorexia Nervosa Binge/Purging Type; **AN-R** = Anorexia Nervosa Restrictive Type; **ANX** = anxiety; **ARFID** = Avoidant/Restrictive Food Intake Disorder; **ASD** = Autism Spectrum Disorder; **AUD** = Alcohol Use Disorder; **BDD** = Body Dysmorphic Disorder; **BED** = Binge Eating Disorder; **BFRB** = Body-Focused Repetitive Behavior; **BN** = Bulimia Nervosa; **BPD** = Bipolar Disorder; **BPD-I** = Bipolar I Disorder; **BPD-II** = Bipolar II Disorder; **CB** = Compulsive Buying; **CAN** = Cannabis Dependence; **CUD** = Cannabis Use Disorder; **ED** = Eating Disorder; **Exter** = externalizing; **GAD** = Generalized Anxiety Disorder; **IA** = Internet Addiction; **IC** = internalizing conditions; **ICD** = Impulse Control Disorder; **MDD** = Major Depressive Disorder; **MDE** = Major Depressive Episode; **NDC** = neurodevelopmental conditions; **NPSD** = Non-Psychotic Spectrum Disorder; **ODD** = Oppositional Defiant Disorder; **OP** = Opioid Dependence; **PD** = Panic Disorder; **PG** = Pathological Gambling; **PH** = Hypersomnia; **PI** = Insomnia; **PTSD** = Post-Traumatic Stress Disorder; **SA** = Suicide Attempt; **SAD** = Social Anxiety Disorder; **SCZ** = Schizophrenia; **SD** = Substance Dependence; **SED** = Sedative Dependence; **SepA** = Separation Anxiety; **SI** = Suicide Ideation; **SMD** = Somatization Disorder; **SP** = Social Phobia; **SPD** = Skin-Picking Disorder; **SPe** = Specific Phobia; **SSD** = Somatic Symptom Disorder; **ST** = Stimulant Dependence; **SUD** = Substance Use Disorder; **TD** = Tic Disorder; **TS** = Tourette Syndrome; **TTM** = Trichotillomania.

**Other abbreviations:** Other abbreviations: **ARR** = adjusted relative risk; **β** = regression coefficient; **CC** = comorbid or co-occurring condition; **CI** = confidence interval; **clin interv** = clinical interview; **CT** = controls; **FDR** - false discovery rate; **FDRs** = first-degree relatives; **GEN** = general population; **HC** = healthy control; **HR** = hazard ratio; **IR** = incidence rate; **LHID** = Longitudinal Health Insurance Database (Taiwan)^143^; **MDiff** = mean difference; **med records** = medical records; **NA** = not available/not reported; **n.s.** = nonsignificant; **OCS** = obsessions and compulsions; **OR** = odds ratio; **PAF** = population attributable fraction; **PP** = postpartum; **q** = FDR-adjusted p-value; **RR** = relative risk; **SD** = standard deviation; **SE** = standard error; **study-spec** = study specific; **Sx** = symptoms; **Sx CL** = symptom checklist; **W/O** = without.; **M** – males ; **F** – females.

**Appendix 5.** Papers that addressed gender differences in genetics of obsessive-compulsive disorder.

Table A5. Gender differences in genetics and familiality of OCD.

| **Author** | **N (F)** | **Age** | **Pop.** | **Study design / target** | **gender/sex effects (F vs M)** | |
| --- | --- | --- | --- | --- | --- | --- |
|  |  |  |  |  | **stats** | **direction** |
| **Family studies** |  |  |  |  |  |  |
| Murphy  2010 | 107  (42) | 4 – 17  9±2 | mothers of kids with OCD/TD  (USA) | Structured interview study | Maternal history. Autoimmune conditions: OCD mothers = 17.8%, gen population = 5% | Autoimmune  OCD mothers > GEN |
| Mataix-Cols  2013 | 24,768 (13,943) ^1st – 3d^R  twn: 16,383 | twn: NA  18 - 44 | SNPR, MGR, STAGE  (SWE) | population based family clustering study | Familiarity ~ OCD. M/F: Similar ORs across M:M, M:F, F:M, and F:F dyads comparing proband to relatives (large # of stats, see eTable3) | **M ~ F:** genetics/ environment ↑↑OCD |
| Steinhausen  2013 | 8,112 (4,154),  1^st^R: 27,396 (13,562) | NA  13-61 | DPCRR  (DNK) | family aggregation case-control study | Familiarity ~ OCD (OCD OR). Paternal OCD OR =34.04 p < 0.01, maternal OCD OR = 11.21 p < 0.002, maternal age >35 OR = 1.25 p < 0.01 | paternal OCD ↑↑ OCD > maternal OCD ↑↑ OCD  maternal age >35 ↑↑ OCD |
| Sackl-Pammer 2015 | 60 (22)  60 mothers  38 fathers | kid: 11±2 | parents of kids with SAD (AUS) | Case-control study | Familiarity of SAD ~ OCD (% OCD). Fathers: SAD = 35% noSAD = 0% p = 0.04; mothers: SAD = 20% noSAD = 7%, p = 0.76. | **M:** Familiarity of SAD ↑↑ OCD |
| Brander  2021 | 22232  (12752) | median = 25  IQR = 16-36 | TPR, SNPR, PDR, MGR  (SWE) | population based family clustering study | Familiarity ~ OCD. OCD-noTD: M-M HR=6.32 [95% CI, 5.37-7.45], M-F HR=4.09 [95% CI, 3.46-4.83], F-M HR= 4.26 [95% CI, 3.63-5.00], F-F HR=4.15 [95% CI, 3.63-4.75].  OCD-TD: M-M HR= 13.02 [95% CI, 8.98-18.87], M-F HR= 7.56 [95% CI, 5.02-11.39], F-M HR= 10.75 [95% CI, 5.78-20.00], F-F HR= 13.95 [95% CI, 8.66-22.45]. | **M-M > F-F/mixed:** familiarity for OCD-noTD |
| Balachander  2021 | 330  (164)  from 153 families | 36 ± 14 | ABDS (IND) | family aggregation study in multiplex (2+) families | Familiarity ~ OCS (ICC). Same-sex pairs: symmetry/ arranging ~0.48, contamination/washing ~0.34, doubts/ checking ~0.38, forbidden thoughts (ns).  Mixed-sex: symmetry/arranging ~0.30, contamination/ washing ~0.34, doubts/checking ~0.18, forbidden thoughts (ns).  Same-sex pairs > mixed-sex (ΔICC ≈ 0.10–0.15). | **same sex > mixed sex:** OCS familiarity |
| He  2022 | 2254234 (1098252) | 17±NA  (0–38) | GEN (DNK) | National registry cohort` | Maternal autoimmune disease (prenatal exposure) ~ OCD. HR = 1.42 [95% CI 1.24–1.63]. | Maternal autoimmune disease ↑ OCD |
| Lin  2022 | 202310  (100670) | 34 ± 18 | ^1st^R of SLE (TWN) | population registry study | Familiarity ~ OCD. ^1st^R of individuals with Lupus (SLE) RR for OCD: F =1.46 [95% CI 1.17-1.82]; M =1.00 [95% 0.48-2.07] | **F**: ^1st^R of SLE↑↑OCD |
| Kendler  2023 | 2413128 (1154458) | 40 ± 11  23 -58 | OCD parents → offspring (SWE) | registry based family aggregation study | Mother → offspring OCD (tetrachoric r). Genes + rearing = 0.19 [95% CI 0.17–0.21], Genes only = 0.26 [95% CI 0.08–0.44], Rearing only: insufficient data.  Father → offspring OCD (tetrachoric r). Genes + rearing = 0.19 [95% CI 0.17–0.22], Genes only = 0.17 [95% CI, 0.10–0.24], Rearing only = 0.04 [95% CI −0.10–0.19]. | Genes + rearing → OCD: paternal ~ maternal  Genes only →OCD: paternal < maternal |
| Zhang  2023 | 2949080  (1433667) | age 3 → up to 37 years f/u | GEN born 1973–2003  (SWE) | Registry study,  Exposure to prenatal and early infections | Risk factors for OCD after age 6. Prenatal infection: maternal HR=1.33 [95% CI, 1.12–1.57], paternal – n.s. Early childhood infections HR=1.19 [95% CI, 1.14–1.25].  Adjusted for parental psychiatric history and autoimmune diseases No longer significant in the sibling analyses. | Prenatal infection: maternal ↑↑ risk for OCD. |
| Zhou  2023 | 2947703  (≈ 1437000) | 29 ± 9  19–49 | OCD parents → offspring (SWE) | registry based family study | Parent psychopathology→ OCD. M ≈ F (no stats). | **M ~ F:** parental psychopathology ↑↑ OCD |
| Hsu  2024 | 767,943 (331,547) | 35 ± 10  12–64 | parents → OCD/SCZ offspring (TWN) | family aggregation study nested in registry cohort | Parent psychopathology→ OCD (F & M OR). *SCZ*: F=1.37 [95% CI, 1.02–1.85], M=1.67 [95% CI, 1.31–2.13]; *BPAD*: F=2.49 [95% CI, 2.01–3.08], M=2.36 [95% CI, 1.95–2.86]; *MDD*: F=2.61 [95% CI, 2.34–2.92], M=2.53 [95% CI, 2.29–2.80]; *AUD*: F= 1.73 [95% CI, 1.39–2.14], M= 1.71 [95% CI, 1.40–2.08]; *SUD*: F= 2.36 [95% CI, 1.91–2.93], M= 1.56 [95% CI, 1.24–1.97]; *OCD*: F= 8.09 [95% CI, 6.31–10.36], M= 7.55 [95% CI, 5.96–9.55]. | **M ~ F:** parental psychopathology ↑↑ OCD |
| Pol-Fuster  2025 | 4,916,898 (2391146) | 24 (IQR 17–32)- age of Dx | relatives of individuals with severe infections (SWE) | Population-based registry family study | Severe infection → OCD in relatives after age 6. Mothers HR=1.49 [95% CI, 1.43–1.55], fathers HR=1.25 [95% CI, 1.18–1.33], maternal half-siblings HR=1.18 [95% CI, 1.13–1.22], paternal half-siblings HR=1.08 [95% CI, 1.04–1.12], aunts HR=1.20 [95% CI, 1.15–1.24], uncles HR=1.11 [95% CI, 1.06–1.16]. | **F > M** relatives: increased OCD risk when proband has serious infection |
| Jo  2026 | 2571290 (1272206) | 32 ± 4  13–58 | mother–child pairs (KOR; replication JPN) | Population-based registry study | Maternal diabetes → OCD in offspring. Pregestational diabetes: aHR=1.16 [95% CI, 0.90–1.50]; gestational diabetes: aHR=0.97 [0.92–1.02]; both n.s. | Maternal diabetes ↛ OCD in offspring |
| Hsu  2026 | 765455 (382385) | NA | FDRs of PD, matched controls (TWN) | Registry family study | PD proband → OCD in first-degree relatives. ARR: M=1.56 [95% CI, 1.37–1.77], F=1.95 [1.70–2.23]. | **F > M:** OCD risk ↑ in first-degree relatives of probands. |
| Li  2026 | 223982 (110363) | 20 ± 3 disease onset | offspring with BPAD (TWN) | Registry based family study | Parental OCD → BPAD offspring. Mania-onset: mother OR=3.31 [95% CI, 2.38–4.60], father OR=4.74 [3.43–6.56]; depression-onset: mother OR=5.12 [3.74–7.01], father OR=3.87 [2.57–5.83]; unspecified: mother OR=3.72 [2.33–5.93], father OR=3.13 [1.75–5.62]. | **OCD:** paternal > maternal for mania-onset; maternal > paternal for depression-onset/unspecified BPAD |
| **Twin studies** |  |  |  |  |  |  |
| Moore  2010 | 508  (262) | 11–16  13±1 | CHSD  (GB-NIR) | twin modeling study | OCD heritability. M: obsess/incompl = 60%; numbers/luck = 65%. | **M:** heritable obsess/incompl/ numbers/luck |
| Anckarsater  2011 | 17,220  (NA) | 9, 12, 15 | CATSS, same-sex twins (SWE) | longitudinal twin study | environ/genes (ICC) ~Sx. Comp. ~ identical (MZ): F = 0.22 M = 0.36; comp. ~ fraternal (Dz): F = 0.16 M = 0.09.  <stats not reported> | **F < M**: comp ~ MZ  **F > M**: comp ~ DZ |
| Bolhuis  2014 | **1:** 2651→ 1597  **2:** 1319F | **1:** 12–21, 15±NA →14–23, 17±NA  **2:** 17- 87, 55±NA | 1: Genesis 12–19  2: UK-ATR  (GBR) | longitudinal twin study | Familiarity ~ Sx. M/F: ‘similar trends in both sexes’ <stats not reported> | **M ~ F:** genetic/environ factors ~ OCS |
| Lopez-Sola  2014 | 2495  (1468) | 18–45  34±8 | ATR (AUS) | twin registry study | Environ/genes ~Sx. MZ M = 0.41 [95%CI, 0.27-0.51], MZ F= 0.42 [95%CI, 0.32-0.51], DZ M = 0.16 [95%CI, -0.03-0.33], DZ F=0.23 = [95%CI, 0.09-0.36] | **M ~ F:** heritability in twins |
| Zilhao  2015 | 20,376  (13,264) | NA  34±15 | NTR (NLD) | population based twin modeling study | Environ/genes ~Sx. M/F: MZ male 0.400 (0.324-0.468), MZ female (0.308-0.400), DZ male/brothers 0.130 (0.090-0.169), DZ female/sisters 0.084 (0.036-0.131) | **M ~ F**: heritability in twins |
| Krebs  2015 | 14,743  (NA) | 4, 7, 9, 16 | TEDS (GRB) | longitudinal population based study | Familiarity ~ Sx. M/F: Multivariate modeling of genetic and environmental influences using the ACE & ADE models examined with and without sex differences with similar trends (ACE sex diff. p<0.05 AIC 25884.28, ACE no sex diff. p<0.05 AIC 26509.63, ADE sex diff p,.05 AIC 25982.94, ADE no sex diff p<.05 AIC=29810.05) | **M ~ F:** Familiarity ~ Sx |
| Pinto  2016 | 21,911  (12,006) | 19 – 47  34±8 | STAGE (SWE) | twin modeling study | Environ/genes ~Sx. M/F: ‘similar trends in both sexes’ <stats not reported> | **M ~ F:** Environ/genes ~Sx |
| **Candidate gene** |  |  |  |  |  |  |
| Alonso  2011 | 508  (227) | 18 – 65  37±11 | OCD, HC  (ESP) | 20 ESR2 SNPs; 9 ESR1 SNPs | Variant ~ OCD. ESR1/ ESR2: F~M (data not reported)  Variant ~ OCD contam/no contam. ESR1/ ESR2: F~M (data not reported) | **F ~ M:** ESR1/ ESR2 ~OCD, ESR1/ ESR2 ~ OCD contam/non-contam |
| Liu  2011a | 482  (231) | ≥ 18  36±16 | OCD, HC  (CHN) | 5-HTTLPR | Variant ~ OCD. 5-HTTLPR: F *Ꭓ^2^*= 6.04 p = 0.01, M *Ꭓ^2^*= 0.21 p = 0.65 | **F:** 5-HTTLPR ↑↑ OCD |
| Liu  2011b | 603  (226) | ≥ 18  29±19 | (CHN) | COMT | Variant ~ OCD. 5-HT2A locus ~ OCD: M p = 0.0255, F n.s.; HTR1B locus ~ OCD: M p = 0.0208 F n.s. | **M:** 5-HT2A/HTR1B ↑↑ OCD |
| Liu  2011c | 103 trios  (41) | NA | OCD+prts  (CHN) | HT2A, HTR1B, DRD2, COMT | Variant ~ OCD (*TDT*). AAIXUtion bwtqwwn 5-HT2A and HTR1B: M p < 0.05; F n.s. | **M:** 5-HT2A, HTR1B ↑↑ OCD |
| Samuels  2011 | 999  (622) | ≥ 18  NA | (USA) | near SLC1A1 | Variant ~ OCD. SLC1A! (& haplotype): M families p = 0.002, F families p n.s. | **M:** SLC1A! (& haplotype) ↑↑ OCD |
| Voyiaziakis  2011 | 1241  (NA) | 7+  NA | OCD + probands  (USA) | SLC6A4 | Variant ~ OCD. Genotype 9/10 of the Int2 VNTR: F p < 0.01. | **F:** SLC6A4 ↑↑ OCD |
| Alonso  2012 | 225 (109)  HC: 279 (NA) | 18–65  37±11 | OCD, HC  (ESP) | 10 GRIN2B SNPs | Variant ~ OCD contam. rs1805476 (GT/TT): M OR = 4.13 p < 0.001, F OR 1.55 p = 0.27.  Haplotype ~ OCD contam/no contam: 4-SNP rs1805476, s1805501, rs1805502, rs180547 (a composite A-A-T-C): OR 1.92 p = 0.023, M ~ F (data not shown). | **M:** rs1805476↑↑ contam  **M ~ F:** 4-SNP ↑↑ OCD |
| Lei  2012 | 572  (312) | 18 – 26  20±1 | STU  (CHN) | 9 variants in SLC6A4 | Variant ~ OCS. 5-HTTLPR: M p = 0.148, F p = 0.004; haplotype CGAAGG/CGAAGG: M p = 0.024 F p > 0.10.  Variant ~ obsess. 5-HTTLPR: M p > 0.10 F p = 0.029.  Variant ~ contam. 5-HTTLPR: M p > 0.10 F p = 0.004; STin2: M p = 0.014 F p > 0.10; haplotype CGAAGG/CGAAGG: M p = 0.002 F p > 0.10. | **F:** 5-HTTLPR ↑↑ OCS/ obsess/ contam.  **M:** haplotype CGAAGG/CGAAGG ↑↑ OCS/contam; STin2↑↑ contam |
| Tukel  2012 | 210  (130) | 18+  28±8 | OCD, HC  (TUR) | BDNF | Variant ~ OCD. BDNF: M n.s., F p = 0.06. | **F:** BDNF↑↑ OCD (trend) |
| Wu  2013 | 1598  (501) | NA | OCD, HC  (CHN) | SLC1A1 | Variant ~ OCD. GACG: M OR = 0.29 [95%CI, 0.15-0.57], F OR = 0.37 [95%Ci, 0.19-0.73]; GGTC: M OR = 0.66 [95%Ci, 0.47-0.91], F OR = 0.63 [95%CI, 0.44-0.90]. | **F ~ M:** SLC1A1 ↑↑ OCD |
| Lennertz  2013 | 540  (229) | ≥ 18  37±13 | OCD, HC  (DEU) | NPSR1 variant | Variant ~ sex in OCD. NPSR1/Asn107Il: n.s. (data not reported) | **F ~ M:** NPSR1 ↑↑ OCD |
| Liu  2013 | 654  (230) | ≥ 18  30±11 | OCD, HC  (CHN) | MAO-A | Variant ~ OCD. Allele frequencies: no association M ~ F | **M ~ F**: MAO-A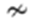 OCD |
| Marquez  2013 | 515 (255)  111 1^st^R, | 26±9  HC: 46±14 | OCD, 1^st^R, HC  (MEX) | BDNF | Variant ~ OCD. BDNF: M ~ F <data not reported> | **M ~ F**: BDNF ↑↑ OCD |
| Mas  2013 | 281  (209) | 10 – 18  16±2 | OCD, AN, HC  (ESP) | 28 candidate genes | Variant ~ OCD. GRIA3: F p < 0.05, M p > 0.05 <similar trend, data not shown>; other 27 variants: M ~ F. | **F:** GRIA3 ↑↑ OCD |
| Taj  2013 | 374  (129) | 29±10  HC: NA | OCD, HC  (IND) | DRD4 | Variant ~ OCD. DRD4: M p = 0.34, F p = 0.02. | **F:** DRD4 ↑↑ OCD |
| Mas  2014 | 64  (37)  trios | NA  15±3 | OCD + prts  (ESP) | 35 genes (frontal-striatal-thalamic system) | Variant ~ OCD. rs2000292 (HTR1B): M p = 0.0006, F p < 0.05; s8190748 and rs992990 (GAD2): M p=0.01, F p=0.0006; other 33 variants: no association M ~ F. | **M > F:** HTR1B ↑↑ OCD  **F > M:** GAD2 ↑↑ OCD |
| Li  2015 | 751  (334) | ≥ 18  29±9 | OCD, HC  (CHN) | DLGAPI, EFNA5 | Variant ~ OCD. EFNA5 rs26728: M p > 0.10 F p > 0.10. | **F ~ M:** DLGAPI, EFNA5 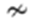 OCD |
| Liu  2015 | 747  (333) | ≥ 18  29±14 | OCD, HC  (CHN) | BDNF | Variant ~ OCD. Val66Met: F p < 0.01; M p > 0.40.  Variant ~ TIC. Val66Met: M p < 0.03 F p > 0.85. | **F:** BDNF ↑↑ OCD  **M**: BDNF ↑↑ TIC |
| Zhang  2015 | 859  (340) | 6-77  23±13 | OCD, HC  (CHN) | OLIG2 | Variant ~ OCD. rs762178: M OR = 1.19 [95%CI, 0.85-1.66], F OR = 1.62 [95%CI, 1.08-2.42] | M < F: OLIG2 ↑↑ OCD |
| Lochner  2016 | 451  (114) | 18+  35±13 | OCD  (ZAF) | dopaminergic genes | Variant ~ OCS. A MAOA ~ symm: M p=0.58, F = 0.16 | **M ~ F:** A MAOA 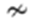symm |
| Kim  2016 | 1195  (412) | 19 – 63  26±11 | OCD, HC  (KOR) | 10 variants in HTR3B | Variant ~ OCD subtype. rs3758987 ~ EO: M OR = 0.49, [CI95% = 0.31–0.79] p = 0.0031.  rs6766410 /rs6443930 ~ contam: F OR = 0.36 [CI95%, 0.18–0.69] p = 0.0016; F OR = 0.47 [CI95%, 0.29–0.79] p = 0.003. | **M:** rs3758987 ↑↑ EO  **F:** rs6766410 /rs6443930 ↑↑ contam |
| Melo-Felippe  2016 | 199 (103)  HC: 200 (NA) | 18 – 75  35±13 | OCD, HC  (BRA) | 2 variants in COMT | Variant ~ OCD. Val158Met (rs4680) alleles: M p = 0.04, F = 0.97.  Variant ~ OCS. rs2075507 ~ order: M OR = 4.25 [CI95%, 1.05-6.57] p = 0.03, F p > 0.10; rs2075507 ~ contam: M p > 0.10, F OR 4.17 [CI95%, 1.55-11.22] p = 0.01. | **M:** COMT ↑↑ OCD, COMT ↑↑ order  **F:** COMT ↑↑ contam |
| Grünblatt  2017 | 245 (94) | NA  13±3 | OCD, HC  (CHE) | Large copy number variations (CNVs) | Variant ~ OCD. Different rare CNVs in males and females with OCD. | **M, F:** rare CNVs ↑↑ OCD |
| Kim  2018 | 1123  (439) | 19 – 63  26±11 | OCD, HC  (KOR) | 6 variants in SLC1A1 | Variant ~ OCD. No association: F ~ M | **F ~ M:** SLC1A1 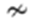 OCD |
| Sina  2018 | 293 (204)  HC: 245 (NA) | 18 – 65  35 ± 10 | OCD, HC  (IRN) | 2HTR2A SNPs | Variant ~ OCD subtype. rs6313 ~ familial OCD (recessive): F p = 0.005, M n.s. | **F:** 2HTR2A varnt. ↑↑ familial OCD |
| Taj  2018 | 377 (147)  HC: 449 (NA) | ≥ 18  29±10  HC (NA) | OCD, HC  (IND) | BDNF | Variant ~ OCD. F/M “No association” <stats not reported>. | **F ~ M:** BDNF 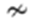 OCD |
| Abdolhosseinzadeh  2019a | 243 (171)  HC: 221 (NA) | 18 – 65  35±10 | OCD, HC  (TUR) | 3 SLC1A1 SNPs | Variant ~ OCD subtype. rs2228622 ~ familial form of OCD: M p = 0.03, F n.s. | **M:** SLC1A1 varnt. ↑↑ familial OCD |
| Abdolhosseinzadeh  2019b | 235 (163)  HC: 225 (NA) | 18 – 65  34±11 | OCD, HC  (IRN) | 2 BDNF SNPs | Variant ~ OCD. rs6265 ~ OCD: M p = 0.01, F n.s.; rs2883187 ~ OCD: M p < 0.001, F p < 0.001. | **M:** rs6265 ↑↑ OCD  **M/F:** rs2883187 ↑↑ OCD |
| Alizadeh  2019 | 205 (143)  HC: 207 (NA) | ≥ 18  34 ± 10 | OCD, HC  (IRN) | 3 HTR1A SNPs | Variant ~ OCD. Genotypic frequency of rs10042486 ~ OCD: M p = 0.020, F p = 0.036; allelic frequency of rs10042486 ~ OCD: F p = 0.019, M n.s.  SNPs ~ AO. C-1019G ~ late age of onset /positive family history: F p < 0.05, M n.s. | **F:** C-1019G ↑↑ LO, C-1019G ↑↑ familial OCD |
| Boloc  2019 | 101 (51)  trio | NA  15±2 | OCD+prts  (ESP) | HTR1B, SLC18A1, GAD1, GAD2 SNPs | Variant ~ OCD (TDT). HTR1B: M p = 0.019, F p = 0.07; GAD2: M p = 0.047, F p = 0.006; GAD1: M p = 0.021 F p > 0.10; SLC18A1: M p = 0.0009 F p > 0.10. | **M > F:** HTR1B↑↑OCD  **F > M:** GAD2 ↑↑ OCD  **M:** GAD1, SLC18A1 |
| Caliskan  2019 | 128  (90) | ≥ 18  34±12 | OCD, HC  (TUR) | 2 MTHFR SNPS | Variant ~ OCD. MTHFR: F ~ M p > 0.05 | **F ~ M:** MTHFR 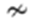 OCD |
| Cotrin  2019 | 400  (228) | 18 - 75  38±14 | OCD, HC  (BRA) | 40 bp VNTR and SNP in DAT | Variant ~ OCD. 10R-carriers: M OR = 0.42 [95% CI, 0.19–0.94] p = 0.02, F p = 0.53.  Variant ~ OCS. 10R-carriers ~ checking: F OR = 4, [95% CI 1.85–8.65] p < 0.001, M p > 0.10; 9R-carriers ~ no checking: F OR = 0.25 [CI95%, 0.12–0.54] p < 0.001, M p > 0.10; T allele of −839C/T~ obsess: M OR = 3.53 [CI 95%, 1.23–10.15] p = 0.02, F p > 0.10; 9R/9R ~ neutral: M OR = 3.38 [95%CI, 1.03–11.11] p = 0.04, F p > 0.10. | **M:** 10R-carriers ↑↓ OCD, 839C/T↑↑ obsess, 9R/9R ↑↑ neutral  **F:** 10R ↑↑ checking, 9R↑↓no checking |
| Huang  2019 | 859  (340) | 9-77  30 ± 15 | OCD, HC  (CHN) | PDE4D polym. rs1838733 | Variant ~ OCD. Female late onset OCD vs. control: p = 0.021; males early/males late/female early– p > 0.16. | **F:** PDE4D ↑↑LO |
| Melo-Felippe  2019 | 402  (228) | 18 - 75  35±13 | OCD, HC  (BRA) | SAPAP3, SLITRK1, PBX1, LMX1A, RYR3 | Variant ~ OCD. PBX1 (GG, G): M p = 0.0003 F p > 0.56.  Variant ~ OCD subtype. LMX1A ~ wash M p > 0.10 F p = 0.01; SLTRK1~check M p = 0.04, F p > 0.10. | **M > F:** PBX1 ↑↑ OCD  **F:** LMX1A ↑↑ wash  **M:** SLTRK1↑↑check |
| Salles-Andrade  2019 | 399  (227) | 18 – 73  38±13  HC (NA) | OCD, HC  (BRA) | SLC1a1 | Variant ~ OCD. rs2072657 (GG): M p > 0.10, F OR = 0.23 [95%CI, 0.04–0.88] p = 0.02; s301443 (G-carrier): M OR = 3.06, [95%CI 1.10–8.4] p = 0.04, F p > 0.10; s301443 (G-carrier): M OR = 1.73, [95%CI, 1.08–2.76] p = 0.03,  Variant ~ OCS. s3780412 ~ hoard (G): M p > 0.10, F OR = 2.46 [95%CI, 1.05–5.80] p = 0.04, rs3780412 ~ hoard (AA): M p > 0.10, F OR = 2.46 [95%CI, 1.03- 5.87] p = 0.04; 301434 ~ order: M p > 0.10, F OR = 2.46 [95%CI, 1.03- 5.87] p = 0.04; s2072657 (T) ~ neutral: M p > 0.10, F OR = 1.98 [95%CI, 1.04–3.75] p = 0.04. | **M:** s301443 ↑↑ OCD  **F:** rs2072657 (GG) ↑↓ OCD, s3780412 ↑↑ hoard, 301434 ↑↑ order, s2072657↑↑neutral |
| Sinopoli  2019a | 2,018  (968) | 6 – 18  11±3 | OCD traits (CAN) | Serotonin genes | Variant ~ OC traits. 5-HTTLRP~ hoard: M p=0.003 and 0.004, F n.s; HTR2A~rumin: M p=1.04E-6 to 5.20E-6, F n.s. | **M:** 5-HTTLRP↑↑ hoard, HTR2A↑↑rumin |
| Sinopoli  2019b | 200  (103) | 6 – 19  13±4 | OCD, HC  (USA) | 5-HTTLPR, SLC6A4, HTR2A, HTR1B, HTR2C | Variant ~ GMV x OCD. Two variants in HTR2C: F rs12860460, p=9.7E-8, n.s. M rs12854485, p=2.07E-6). | **F:** HTR2C ↑↑GMV x OCD |
| Vieira-Fonseca  2019 | 205  (105) | 18+  38±13 | OCD, HC  (BRA) | OLIG2 | Variant ~ OCD. rs762178-rs965371 M OR = 3.17 [95%CI, 2.01-4.96], F OR = 1.57 [95%CI, 1.08-2.28]. | **M~F:** OLIG2 ↑↑ OCD |
| Park  2020 | 259  (91) | 18+  22±2 | OCD, HC  (KOR) | DNA methylation of oxytocin receptor gene | Methylation profiles of genes ~ OCD. M~F: levels of methylation in cpg1 and cpg2 sites of UTR OXTR1↓↓ | Methylation profiles of OXTR1  M ~ F |
| Huang  2021 | 1102  (367) | 18+  29±13 | OCD, HC  (CHN) | 4 SLCIAI SNPs | Variant ~ OCD. rs301434: M p = 0.013, F p = 0.31.  Haplotypes ~ OCD. G-A-G-G: M OR = 1.737 [95% CI: 1.134–2.66] p = 0.01, F p > 0.10; G-G-A-G: M p > 0.10, F OR = 1.67, [95% CI: 1.021–2.730] p = 0.03. | **M:** rs301434/G-A-G-G ↑↑ OCD  **F:** G-G-A-G↑↑ OCD |
| Siu  2021 | 254  (NA) | 2– 18  NA | OCD, HC  (CAN) | DNA methylation of OXTR gene | OXTR methylation ~ OCD.: two CpG sites hypomethylated in OCD vs HC, Δβ. Pooled: site 1 = 1.37% site = 1.62%; M: site 1 = 1.45%, site 2= 1.83%; p < 0.05. | **M:** OXTR CpG hypomethylation ↑↑ OCD |
| Seo  2023 | 458  (180) | 18– 40  NA | OCD, HC  (TWN) | DNA methylation of HPA axis-related gene FKBP5 | Variant ~ OCD. FKBP5 intron 7 CpG hypomethylation, OCD vs HC (Δβ): M = 1.33%, [95% CI 2.11-0.55], p < 0.001, F =0.44%, [95% CI –1.51-0.63], p=0.418).  “*Finding in males remained significant in drug naive individuals and those without MDD, and no mediation effect was observed between early-life trauma and OCD*.” | **M:** FKBP5 hypomethylation ↑ OCD |
| Akkus  2024 | 195  (108) | 18– 55  32±10 | OCD, HC  (TUR) | SNPs in DLGAP1 and SLC1A1 | Variant ~ OCD. DLGAP1 rs11081062: F OR = 3.01 [95% CI, 1.04–8.71] p = 0.042 | **F:** DLGAP1 rs11081062↑ OCD |
| Bellia  2025 | 52  (24) |  | OCD. HC (ITA) | DNA methylation of SLC6A4, MAOB, COMT | Methylation profiles ~ OCD. OCD vs HC: M ~ F: SLC6A4 and MAOB methylation ↓; MB-COMT methylation ↑ in OCD. | **M ~ F**: SLC6A4, MAOB, COMT methylation profiles. |
| Wang  2025 | 82  (65) | 30 ± 14 | OCD, HC (CHN) | DNA methylation of TPH2 and parenting | TPH2 methylation ~ parental factors. CpG2 differed by emotional warmth (t=−3.82, p<.001), punishment (t=3.04, p=.006), and rejection (t=2.28, p=.032); CpG5 (Z=−2.268, p=.023) and CpG9 (Z=−2.15, p=.031) were associated with F overprotection. For M, CpG5 was associated with excessive interference (t=−2.43, p=.023), CpG10 with rejection (Z = −2.24, p=.025), and CpG4 with preference (Z=−2.19, p=.029). | In OCD: F overprotection ~ CpG5/9; M parenting factors ~ CpG4/5/10 |
| **Genome-wide molecular studies** | | | | | | |
| Lionel  2014 | 64,114  (23,676) | NA | NDD  10 countr | rare CNVs at 9q33.1 | CNV disruption ~ OCD. ASTN2/TRIM32 locus with cooccurring OCD: M p < 0.01, F p > 0.10. | **M:** ASTN2/TRIM32 ↑↑ OCD |
| Khramtsova  2019 | 9,870  (5,832) | 18 + | IOCDF-GC  multiple | GWAS study meta-analysis | Genome-wide SNPs. GRID2: F p =1.07E-07, M p = 0.72; GRP135: F p = 1.55E-06, M p = 0.70.  OCD heritability estimates: M~F: Strong genetic correlation between M and F (r_g_=1.04 ± 0.51 p = 0.04) | **F:** GRID2/ GRP135 ↑↑ OCD  OCD genetic correlation **F↑↑M** |
| Ohi  2022 | ~ 337000  (~172000) | 40–69 | UK Biobank  (GBR) | GWAS study and genetic correlations | GWAS genetic correlation (rg) ~ OCD.  Later age at first sexual intercourse ↑↑ OCD: M & F p < 0.005; Age at first birth ↑↑OCD: F only p<0.005  Number of children ever born ↑↓OCD: M p<0.005, F p<0.01. | **M & F:** reproductive timing ↑ OCD; fertility ↓ OCD |
| Kutzner  2023 | 547  (301) | 16-17 → 18-19 | longitudinal trial (USA) | PRS analysis in a longitudinal cohort | PRS ~ OCS/cannabis. Pooled and EA males only: OCS PRS ↓ cannabis use at 17 and ↑ OCS at 19. | **EA M:** OCS PRS ↓ cannabis use |
| Aicoboaie  2025 | 4729  (2343) | 8-21 | PNC (USA) | polygenic risk for OCD, AN, SCZ ~ phenotypes, neurocognitive measures, imaging | OCD PRS ~ phenotypes. OCD PRS was not associated with mental-health phenotypes, neurocognitive measures, or cortical phenotypes in M or F; all FDR-adjusted P>.05. | **M ~ F:** no detectable OCD PRS associations. |
| Halvorsen  2025 | 5856  (NA) | 18–65 | OCD, HC (NOR & SWE) | CNV analysis | CNV burden ~ OCD. OCD cases had increased rare CNV burden (OR=1.12, P=1.77×10⁻³), particularly CNVs overlapping protein-coding regions (OR=1.19, P=3.07×10⁻⁴). Deleterious CNV burden: M ~F OCD cases. | **M ~ F:** deleterious CNV burden in OCD. |
| Sakurai  2025 | 28  (11) | 36 ± 11 | OCD, OCD+NDD (JAP) | RNA sequencing study | Gene expression ~ OCD/NDD. Differentially expressed genes (P<.01): 431 in M and 268 in F, with 9 genes commonly differentially expressed in M and F. | **M > F:** number of differentially expressed genes; limited overlap between sexes. |
| Hoffler  2026 | 798  (572) | 18-14 | OCD, HC (NOR) | Methylome-wide association study | DNA methylation ~ OCD. Identified 1 significant DMP and 14 DMRs in F and 7 DMRs in M; most DMRs showed sex-by-diagnosis interactions. Associated genes included AKAP12, PIWIL1. | **F > M DMRs:** Sex-specific OCD-associated methylation |
| **Parental age studies** | | | | | | |
| Wu  2012 | 711  (305) | 18+  24±8 | SCZ, OCD, HC  (CHN) | cohort study | Relative risk for OCD (RR, reference= age 25-29). Paternal age 30-34 RR = 2.225 p=0.005, paternal age >=35 RR = 5.413 p<0.001; Maternal age 30-34 RR = 0.698 p=0.294, maternal age >=35 RR =0.398 p=0.214. | **M > F:**  Advanced paternal age↑↑OCD |
| Chudal  2017 | 1,358 individuals with OCD matched with 4 controls | 3-13  13±3  TS: 0-19  9±3 | FHDR (FIN) | nested case-control study | Relative risk for OCD (reference =age 25-29). Maternal age: 35-39 OR = 1.31 [95% CI 1.03-1.66], paternal age n.s. in final model | Advanced maternal age ↑↑ OCD |
| Janecka  2019 | 1,490,745  (NA) | 6 - 33 | DMBR, DCRS  (DNK) | birth cohort study | Relative risk for OCD. Higher maternal age vs. higher paternal age HR = 1.23 [95%CI, 1.07-1.42].  Paternal age: M HR = 1.14 [95%CI, 1.00-1.31]; F HR = 1.04 [95%CI, 0.92-1.16]. | Parental age ↑↑ OCD: paternal ~ maternal |
| Biria  2019 | 85  (18) | 18+  21±5 | SCZ + clozap (GBR) | cohort study | Relative risk for OCD. Father’s age at birth: OCD = 29.6 ± 6.1 non-OCD = 33.52 ± 8, d = 0.58 p = 0.028; Mother’s age at birth: OCD = 27.02 ± 5.06 non-OCD = 30.17 ± 6.33, d = 0.58 p = 0.056. | SCZ + clozap father/mother age at birth ↑↑OCD |
| Wang  2022 | 7264788  (NA) | 0-38 | GEN (TWN) | Single-payer compulsory insurance registry | Relative risk for OCD. Paternal age (25+) OR = 1.13 per 10-year increase [95% CI: 1.09–1.17]. Maternal age (25+) OR per 10-year increase = 1.28, [95% CI, 1.21–1.36]. | Paternal & maternal age ↑↑ OCD |
| Yilmaz  2022 | 1154067 | 6-29 | GEN  (DNK) | birth cohort study | Relative risk for OCD. Older maternal age: M p < 0.001, F p = 0.15; M vs F p=0.01; Older parental age: M p=0.01, F p=0.91, M vs F p=0.09. | **M:** Paternal & maternal age ↑↑ OCD |
| **Other** | | | | | | |
| Schneider  2016 | 131  (41) | 32±9 (18+) | FMR1 (USA) | Case-control study | Comorbid OCS, severity. M: HC = 48.27 FMR1 = 45.04, p=0.618; F: HC = 44.67 FMR1 = 66.91, p = 0.012. | FMR1 ↑↑ OCS: M < F |
| Loesch  2021 | 22  (13) | M:62 → 67±8  F:53 → 63±13 | FMR1  USA  AUS | SCL-90-R, genetics | FMR1 ~ Sx progression, Δ T-scores. M = - 6 p > 0.05, F = 7 p < 0.001;  F x Δ T-scores p > 0.05 | FMR1 ↑↑ Δ Sx: M ≤ F |
| Kang  2021 | 469  (188) | 28±8  (18+) | OCD, HC (KOR) | mtDNA copy number and telomere length study | mtDNAcn / telomere length ~ OCD. mtDNA copy number ↓ OCD in M & F (p < 0.001); telomere length ↓ OCD in F only (p = 0.023). \| M & F: mtDNAcn ↓ OCD; F: telomere ↓ OCD. | **M ~ F**: mtDNAcn ↓OCD;  **F**: telomere length ↓ OCD |
| Zhang  2022 | 3345  (1746) | 12-20 | GEN (CHN) | Cohort study of middle school students | Parental rearing factors ~ OCS, r. Paternal warmth = −0.13, maternal warmth = −0.07; paternal punitive = 0.23, paternal over-involved = 0.20, paternal rejection = 0.26, paternal overprotection = 0.24; maternal over-involved/overprotection = 0.32, maternal rejection = 0.34, maternal punitive = 0.25. | Paternal/maternal: Warmth ↓ OCS; punitive/rejection/overprotection ↑ OCS |
| Salvatore  2024 | 2828777  (1404283) | 52 ± 11  21–68 | GEN  (DNK) | Registry study, family genetic risk | Familial genetic risk score (OCD) ~ divorce: F > M (no stats reported, only figures) | divorce↑↑genetic risk for OCD: **M < F** |
| Kohlrausch  2025 | 210  (118) | 37 ± 11  18-73 | OCD, HC (BRA) | Telomere length (TL) study | TL ~ OCD/sex. Age was negatively correlated with TL in F OCD (ρ=−0.349, P=.002) and M OCD (ρ=−0.422, P=.001), but not HC. TL was shorter in both M and F OCD vs HC. TL correlated negatively with OCD severity in F (ρ=−0.341, P=.009), but not M (ρ=−0.228, P=.152); association in F was n.s. after controlling for age. | TL ~ OCD severity before age adjustment; M ~ F: shorter TL in OCD vs HC. |
| **Single Gender studies** | | | | | | |
| Iervolino  2011 | 4355F | 17-86  56±13 | TwinsUK  (GBR) | Multivariate twin modeling | Environ/genes (ICC) ~OCS, genetic/environment. Common: Checking = 0.27/0.27; Hoarding = 0.20/0.24; Obsessing = 0.29/0.14; Ordering = 0.29/0.21; Washing = 0.35/0.25. Specific: Checking = 0.11/0.35; Hoarding = 0.24/0.27; Obsessing = 0.18/0.34; Ordering = 0.15/0.35; Washing = 0.04/0.25. | **F:** common genetic factors → risk for broadly defined OCD,  specific environmental factors → specific Sx. |
| Monzani  2012 | 2148F twins | 16 – 90  60±13 | TwinsUK  (GBR) | Twin registry study | Common OCD/BDI phenotypes ~ genetics. F = 64% [95%CI, 50% – 80%] | **F:** genetic → OCD/BDI phenotypic overlap |
| Taberner  2012 | 184F | 18 – 35  22±3 | STU + parents  (ESP) | Familial predictors of OCS | Familiarity ~ OCS. Contam: Hygiene in fathers β=0.22, t(155)=3.5, p<.001; symm/order in mother β=0.20, t(155)=3.1, p<.05. | fathers: Hygiene→ contam  mothers: symm/order → contam |
| Veenstra  2012 | 184M | 7 – 53  20±13 | OCD  (USA  CAN) | Rare variation in SLC1A1 | Variant ~ OCD. No significant associations. | **M:** SLC1A1≁ OCD |
| Bergin  2014 | 1382F twins | 29 ± 8 → 36 ± 8 | Twins  (USA) | Longitudinal twin study, Padua, Eysenck Person., | Cross Twin Correlations. neurot. r = 0.15 p < 0.05; contam r = 0.43 p < 0.05; aggress obsess r = 0.38 p < 0.05; check r = 0.39 p < 0.05. | **F:** Cross twin corr: neurot, contam, aggress obsess, check. |
| Monzani  2014 | 5409F twins | 16 – 90  NA | TwinsUK  (GBR) | Twin registry study | OCD specific genetic factors (EFA on phenotypic factors). The best fitting model = 2-factor latent model; factor 1 = OCD, BDD, HD; factor 2 = TTM and ED. | **F:** 2 liability factors - OCD/BDD/HD and TTM/ED |
| Cederlof  2015 | 28,276  (19,118)  twn: 8,550F | ≥ 18  NA  twn: 33±8 | OCD, AN SNPR/ STR  (SWE) | Registry longitudinal and twin study | Familiarity of OCD ~ AN in F (RR). M RR = 36.9 [95%CI, 20.0-68.1], F RR=16.1 [95%CI, 14.0-18.5].  OCD/AN genetic overlap. F: ‘moderate degree’ | **M > F:**  familiarity OCD ↑↑ AN |
| Scherrer  2015 | 1675M twins | 45 – 60  NA | VETR  (USA) | Twin registry study | OCD specific genetic factors (LCA on phenotypic factors). The best fitting model = 4-factor latent model; factor 1 = unaffected, factor 2 = ritual/symm, factor 3= germ/contam, factor 4 = severe OC. | **M:** 4 genetic classes – unaffect, ritual/symm, germ/contam, severe OC |
| Campbell  2016 | 47F | NA  44±13 | FMR1 prem carriers  (SCO) | Special pop. study | Variant x phenotypic factors ~ OCD (%). FMR1 prem in F: endocrine PMC OCD = 44.4%, non-endocrine PMC OCD = 23.1%, no PMC OCD = 0%, p = 0.009. | **F:** FMR1 premutation + endocrine PMC↑↑ OCS |
| Nissen  2016 | 33F | NA  13±3 | OCD, HC  (DEN) | DNA methylation study, 14 genes | Methylation profiles of genes ~ OCD. *“No statistically significant differences”* | Methylation profiles  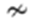 OCD |
| Lee  2018 | 107M | 5 – 23  12±6 | DMD (USA) | Retrospective chart review | DMD ~ OCS. M: OCS in 14% of DMD | **M:** DMD ↑↑ OCS |
| Jones  2021 | 400F | 4–18  11 ± 3 | Tic/OCD, HC, ND  (AUS) | Cytokine and transcriptome analysis | Parental autouimmune disease ~ tic/OCD. Maternal autoimmune neurological disorder ↑ tic/OCD (p = 0.05 vs controls; p = 0.0004 vs HC). | Maternal autoimmunity ↑ tic/OCD |
| Korkmaz  2025 | 42F | 18-65  39 ± 13 | OCD, HC (TUR) | Biomarker study, 3 miRNAs | miRNAs ~ OCD. In OCD vs HC, miR-132 was ↑ (P=.006), miR-125b-5p was ↓ (P<.001), and serotonin was ↑ (P<.05). | F only: miR-132 and serum serotonin higher in OCD; miR-125b-5p: OCD < HC. |

**Abbreviations used in Table A5: 1stR** – first degree relative; **ACE** – Additive genetic (A), Common environmental (C), and Unique environmental € influences model; **ADE** – Additive genetic (A), Dominant genetic (D), and Environmental € influences model; **aggress** – aggressive symptoms; **AIC –** Akaike Information Criterion; **AN –** anorexia nervosa; **ATR** – Australian Twin Registry^226^; **BDD –** body dysmorphic disorder; **CATSS** – the Child and Adolescent Twin Study in Sweden^227^; **check –** checking symptoms; **CHSD –** the Child Health System Database^228^; **CI –** credible interval; **contam –** contamination symptoms; **corr** – correlation; **DCRS –** the Danish Civil Registration System^229^; **diff –** difference; **DMBR –** the Danish Medical Birth Registry^230^; **DMD –** Duchenne Muscular Dystrophy; **DPCRR –** the Danish Psychiatric Central Research Register^231^; **DZ** – dizygotic twins; **ED –** eating disorder; **environ –** environment; **ESR1** – the estrogen receptor 1 gene; **ESR2: -** the estrogen receptor 2 gene; **FHDR** – Finnish Hospital Discharge Register^232^; **Genesis 12–19 –** a twin and sibling study of gene–environment interplay and adolescent development in the UK^233^; **HC** – healthy control; **HD** – hoarding disorder; **hoard** – hoarding symptoms; **HR** – hazard rate; **incompl** - incompleteness symptoms; **IOCDF-GC –** International OCD Foundation – Genetics Collaborative; **IQR -** Interquartile Range; **MAOA** – monoamine oxidase A; **MGR –** the Multi-Generation Register^234^; **MZ –** monozygotic twins; **n.s.** – not significant; **NDD –** Neurodevelopmental Disorders; **neurot –** neuroticism; **obsess** – obsessive symptoms; **OCD –** obsessive compulsive disorder; **order -** ordering symptoms; **PDR –** Prescribed Drug Register^235^; **prem -** premutation; **prts –** parents; **RR –** risk rate; **rumin –** ruminative thoughts; **SAD –** separation anxiety disorder; **SCZ –** schizophrenia; **SNP –** Single Nucleotide Polymorphism; **SNPR –** The Swedish National Patient Register^236^; **STAGE –** Swedish Twin Study of Adults: Genes and Environments^219^; **STR –** Swedish Twin Register^237^; **STU –** students; **symm –** symmetry symptoms; **TD –** tic disorder; **TDT –** Transmission Disequilibrium Test; **TEDS –** Twins Early Development Study^238^; **TPR –** Sweden Total Population Register^239^; **TTM –** Trichotillomania; **TwinsUK –** the United Kingdom adult twin registry^240^; **UK-ATR – U**K Autism Twin Registry^241^; **unaffect –** unaffected; **VETR –** The Vietnam era twin registry^242^; **wash –** washing symptoms**. M –** male**; F –** female.

**Appendix 6.** Papers that addressed gender differences in altered functioning in obsessive-compulsive disorder.

Table A6. Gender differences in neurocognitive, neurobiological, and hormonal systems functioning.

| **Author** | **N**  **(F)** | **Age** | **Pop.** | **Process** | | **Measures** | | **gender/sex effects (F vs M)** | | | | |
| --- | --- | --- | --- | --- | --- | --- | --- | --- | --- | --- | --- | --- |
|  |  |  |  |  |  |  |  | **stats** | **stats** | | | |
| ***Cognition*** | | | | | | | | | | | | |
| *pooled* | | | | | | | | | | | | |
| Xiong  2021 | 227 (107) | 13–45 | OCD  HC  (CHN) |  | cognitive fusion, experiential avoidance | CFQ, AAQ-II | | CFQ. OCD-M = 42.07 ± 8.31 OCD-F = 44.23 ± 8.99 d = –0.25, p = 0.18  AAQ-II. OCD-M = 34.97 ± 5.44, OCD-F = 34.88 ± 6.34, d = 0.02, p = 0.93 | Cognitive fusion/ experiential avoidance: M ~ F | | | |
| *children/adolescents* | | | | | | | | | | | | |
| Esbjørn 2013 | 974 (538) | 9-17  NA | GEN  (DNK) |  | Metacognition about OCs | MCQ-C30, SCARED-R | | Total effect: M → MCQ-C: β=-0.07 p<0.05; Direct effect: n.s.  Indirect effect: M → SCARED OCD β=-0.2 p<0.001 & SCARED OCD → MCQ-C: β=03 p<0.001 | Subclinical OCs: F > M *which leads to*  MCQ-C: F > M | | | |
| Shaw 2021 | 277 (145) | 8-17 13±3 | OCD DEP ANX  (USA) |  | Affect intolerance (AI) | DTS, CASI, RCADS | | M: AI ~ OCs (M: β =0.86 p <0.001;  F: β =0.79 p <0.001) | **F:** OCs↑↑AI  **M:** OCs↑↑AI | | | |
| Barcaccia  2026 | 950  (485) | 8-14  11 ± 1 | school  (ITA) |  | obsessive beliefs | OBQ-CV, OCI-CV, SCAS, CDI | | Obsessive beliefs ~ OCS. *Cross-sectional partial* *r*: t1: M = 0.37 [0.29–0.45], F = 0.29 [0.21–0.37]; t2: M = 0.33 [0.24–0.41], F = 0.30 [0.22–0.38]; t3: M = 0.36 [0.27–0.44], F = 0.31 [0.22–0.39]. sex-difference posterior prob. = 0.07–0.34.  Longitudinal panel, β. *Obsessive beliefs → later OCS*: M = 0.20, p < 0.05; F n.s. (β/p not reported). *OCS → later obsessive beliefs*: n.s. in both sexes. | Obsessive beliefs ~ OCS: M ~ F  Obsessive beliefs → later OCS: M > F | | | |
| *adults* |  |  |  |  |  |  | |  |  | | | |
| Gülüm 2014 | 992 (661) | 17-29 21±2 | GEN  (TUR) |  | Locus of control (LOC), Repetitive Thinking (REP) | RTQ, LCS, ECR-R, MOCSL | | LOC ~OCs F: β=0.16, p<0.001; M: β=-0.03 p>0.05; *Mediation by attach anx.* F: Sobel test=3.6 p<0.001 M: Sobel test = 0.9 p > 0.1  REP~OCs F: β=0.28 p<0.001 M: β=0.22 p<0.001; *Mediation by attach anx.* F: Sobel test = 5.3 p<0.001 M: Sobel test =3.2 p<0.001 | M&F: REP → OCs ↑↑ mediated attach anx.  F: LOC → OCs ↑↑ mediated attach anx. | | | |
| Whitehead 2016 | 199 (119) | 18-24 20±1 | STU  (USA) |  | Impulse control (ImpC), Goal directed (GoalD) | SCL-90-R; OC- modf. DERS-modf. | | DEP=70 (30F), pOCD=58 (43F), pOCD/DEP=71 (46F) p<0.01  DEP > pOCD: ImpC=1.15 p<0.05; F=0.20 p<0.01;  pOCD/DEP > OCD: ImpC=1.21, GoalD=1.10 p<0.05, F=0.34 p<0.05; pOCD/DEP > DEP: GoalD x M=1.17 p<0.05 | M&F:  ImpC: DEP, pOCD/DEP > pOCD  M:  GoalD: pOCD/DEP > DEP;  F: pOCD > pOCD/DEP, DEP | | | |
| Vivas 2019 | 100 (71) | 18-29 21±3 | STU  (ESP) |  | Emotional processing (Arousal, Valence), personality | IAPS, DOCS, NEO-FFI, SCID–I | | M: Neuro high OCs > Low OCs d = 0.84, p<0.05, DOCs-unaccept ~ Agree β_std_ = -0.39 p = 0.037,  DOCs-unaccept ~ Arousal β_std_ = 0.52 p = 0.007,  DOCS-cont ~ Valence β = -0.42 p = 0.03  F: Valence hOCs > lOCs d =0.52 p < 0.001; Agree hOCs < lOCs, d = 0.59 p <0.05;  DOCS-resp ~ Valence β=0.25 p =0.04; DOCS ~ Neuro β=0.44 p <0.001, DOCS ~ Agree β=-0.32 p=0.007 | M&F: OCs↑↑Neuro;  F: OCs↑↑Valence,  OCs ↑↓Agree | | | |
| Bezahler 2024 | 470  (118 SM) | 29±11 | OCD  (USA) |  | Emotion regulation | Y-BOCS-SR, DERS-16, DTS-SF | | DERS-16 total × SM: d = −0.25 p = 0.028  DERS-Strategies × SM: d = −0.26 p = 0.023  DERS-Nonacceptance × SM: d = −0.30 p = 0.011  DTS-SF × SM: d = 0.28 p = 0.015 | Emotion reg in OCD: M/F > SM | | | |
| Ding  2025 | 1529 (919) | 18–25 20 ± 1 | STU (CHN) |  | Intolerance of uncertainty (IU) –OCs network | C-IUS-12, OCI-R | | Network structure invariance = 0.22, p = 0.053. Global strength = 12.90, F = 12.25; S = 0.65, p = 0.09. | IU–OC symptom network: M ~ F | | | |
| Wagner 2025 | 1345 (939) | 18–83 31 ± 11 | current/  past OCS (HISP; USA/  LATAM) |  | Reward responsiveness | PVSS-21,  Y-BOCS-II, PHQ-9 | | OCS ~ reward responsiveness. **M**: OCS ↑↓ social responsiveness, edge=−0.11 [95% CrI, −0.22, −0.01], OCS ↑↓ hobbies = −0.12 [95% CrI, −0.23, −0.02].  **F**: OCD severity ↑↓ social responsiveness, −0.09 [95% CrI, −0.15, −0.02], OCS↑↑ goal responsiveness, 0.08 [95% CrI, 0.01, 0.14]. | M&F: OCS ↑↓ soc. responsiveness  M: OCS ↑↓ hobbies  F: OCS↑↑ goal responsiveness | | | |
| Yong  2026 | 2364 (1324) | 18–30 22±NA | STU (SGP) |  | OCD-related beliefs; conformity, religiosity, fear of losing out | SCOPI, OBQ, FoLO, CS, DUREL, PHQ-8, GAD-7 | | R/TE: M>F, W1 t(2340)=4.41, d=.18; W2 t(1857)=4.63, d=.22, ps<.004. Conformity: F>M, t(2341)=5.15, d=.06, p<.004. Religiosity: F>M, t(2341)=3.00, d=.13, p<.004. | responsibility/threat estimation: M > F conformity, religiosity: F > M | | | |
| ***Behavior*** | | | | | | | | | | | | |
| *children/adolescents* | | | | | | | | | | | | |
| Eyüpoğlu 2026 | 100 (46) | 12–18 16 ± 2 | OCD (TUR) |  | Sensory processing, executive functioning,  cognitive flexibility | AASP, WCST, SCWT, CCTT, VFT, CY-BOCS | | WCST performance, β_F_-_std_. Failure to maintain set = −0.00, p = 0.99. Categories completed = 0.06, p = 0.55. CCTT-2 completion time = −0.07, p = 0.51 | WCST performance: M ~ F | | | |
| *adults* | | | | | | | | | | | | |
| Cougle 2013 | 167 (107) | 18-30 19±1 | GEN  (USA) |  | Persistence, distress intolerance | MTPT HGPT; OCI-R | | HGPT: M x obss. β=0.21 p<0.02; in M persistence ~ obss. β=-0.42 p <0.01, in F – n.s.  Distress intolerance & MTPT: obss. x M - n.s. | M: persistence↓↑ obss.; F: n.s. | | | |
| Martoni 2015 | 84  (36) | 18+ 34±12 | OCD  HC  (DEU) |  | Visuospatial working memory (VSWM) | CANTAB battery: SWM, SRM, DMS; DYBOCS | | SWM errors: F x Dx p <0.0005; F x Dx x load F(3,79)=4.8 p<0.005;  SWM strategy: F x Dx p <0.005;  VS storing and recognition: F x Dx p > 0.1; VSM: delay x F x Dx: p >0.1 | SWM errors: OCD-F > OCD-M = HC-F = HC-M, strategy: HC-M = HC-F = OCD-M > OCD-F;  VS storing capacity & recognition: OCD-M = OCD-F = HC-M = HC-F | | | |
| Steinman 2016  (& Ahmari 2016) | 170 (102) | 18–50 27±6 | OCD  SAD AN HC  (USA) |  | Prepulse inhibition (PPI) | PPI at 74, 78, or 86 dB | | Dx x F: p = 0.04  OCD-F: 19.7±6.0%; HC-F: 35.6±4.8%; p = .039, ES = 0.57 | PPI: OCD-F < HC-F = HC-M = OCD-M | | | |
| Cillo 2019 | 205 (101) | 18+ 33±11 | OCD HC  (USA) |  | Habitual risk taking | Ordered CGT | | OVERALL: OCD-M=0.54±0.15, OCD-F= 0.47±0.15, HC-F =0.44±0.14, p<0.05; Ascend: OCD-M =0.43±0.20, OCD-F=0.35±0.19, HC-M=0.39±0.15, HC-F =0.35±0.15, p<0.05 | OCD-M > OCD-F = HC-M = HC-F | | | |
| Ma  2021 | 58  (31) | 18 ≤  31±11 | OCD HC  (USA) |  | Information processing, decision formation | PVDM | | HDDM model: Decision threshold (a): OCD-M > HC-M across perceptual easy & difficult conditions (p <0.05), OCD-F ≈ HC-F (n.s)  Drift rate (v): OCD-M < HC-M across perceptual and value-based tasks (p <0.01), OCD-F ≈ HC-F (no effect) | Decision threshold: OCD-M > HC-M, OCD-F ≈ HC-F  Drift rate: OCD-M < HC-M, OCD-F ≈ HC-F | | | |
| Bednarek 2024 | 97  (59) | 18–63 32±12 | OCD HC  (USA) |  | Alerting and executive attention | ANT-R | | Alerting benefits. HC ~OCD p=0.53. OCD x F p > 0.10  Simon/Flanker costs: HC < OCD p<0.001, OCD x F p >0.10 | M ~ F: executive attention ↓↓ OCD | | | |
| Mısır  2025 | 135 (99) | 18 + 29±9 | OCD  (TUR) |  | Neurocognitive functioning | CVLT, WCST, TMT-A/B, COWAT, DST, ACT, Y-BOCS, HDRS-17 | | 2 clusters: (1) Globally Impaired (GI); (2) Cognitively Intact (CI)  GI: F > M p = 0.015 | Globally Impaired: F > M | | | |
| Gambolò 2025 | 31  (11) | 18 +  45 ± 20 | OCD  (ITA) |  | Motor structure of compulsive rituals | Video/BORIS; Y-BOCS, CTQ, FCQ, SOFAS | | Ritual subtypes in OCD: iterative rituals = 13 (F= 53.8%); idiosyncratic rituals = 14 (F=28.6%); F:M OR = 2.92 [95% CI: 0.59–14.33], p = 0.182. | Iterative versus idiosyncratic ritual subtype: M ~ F | | | |
| Puialto 2026 | 90  (39) | 18–62 39 ± NA | OCD HC (ESP) |  | Executive function, non-verbal memory; 10.6-y f/u | Digit Span, RCFT, Y-BOCS, DY-BOCS, HDRS | | Working memory/organizational strategy composite: sex = 2.10 [95% CI, 0.20–4.00], p<.05; age×sex = −0.056 [−0.104 to −0.008], p<.05. With increasing age, women showed poorer performance relative to men.  Non-verbal memory composite: sex = 0.41 [−2.66 to 3.48], n.s.; age×sex = −0.03 [−0.10 to 0.05], n.s. | Working memory/organizational strategy: age-related decline F > M; non-verbal memory: M ~ F | | | |
| Shankar  2026 | 119 (46) | 18–45  28 ± 8 | OCD  (IND) |  | Neuropsychological functioning | DST, CTT-2, CFT-IR, TOH-3, WCST | | Performance-dependent sex differences in performance, M vs F. *DST:* M > F at the 25th–75th percentiles (β=2.04–4.16, p=.01–<.001); 5th/95th: M ~ F. *TOH:* M > F at the 50th–95th percentiles (fewer moves, p=.01–.05); 5th/25th: M ~ F.  *CTT-2, CFT-IR, WCST-PR, WCST-TTCC1*: M ~ F across performance levels, all p > 0.05 | | | | |
| ***Neurophysiology*** | | | | | | | | | | | | |
| *pooled* | | | | | | | | | | | | |
| Weeland  2022 | 5423  (2713) | 6–69 | OCD HC (INT) |  | GM volume, thalamic subnuclei | | Thalamus (anterior, medial, pulvinar, ventral, whole) ~ age × M × Dx: <12: d = −0.32 to −0.42, p ≈ 0.006–0.045  (12–17): ps > 0.10  18+: Anterior only: d = −0.07 p = 0.03. | | | | | <12: Thalamus ↑↑ age: M > F  12–17: Thalamus ~ age: M ~ F  18+: Anterior thalamus F-OCD > M-OCD |
| *children* | | | | | | | | | | | | |
| Suñol  2018 | 255 (125) | 8-12 10±1 | GEN  (ESP) |  | GM /WM volume, OCI-CV, | | OCI-CV dimension ~ brain volume x F n.s. | | | | | M = F |
| Suñol  2020 | 227 (118) | 8-12 10±1 | GEN  (ESP) |  | Resting state fc,  OCI-CV | | left medial dorsal thalamus↔ STN ~ hoarding differed by F (girls: r=-0.34 boys r = -0.82 p =0.04); medial dorsal thalamus ↔PCC ~ hoarding x F x age (girls >9.67 y: r = -0.507, other groups r = -0.112 p =0.004) | | | | | |
| Hanna 2026 | 210 (130) | 8–18 14 ± 3 | OCD HC (USA) |  | ERP brain potentials during flanker task | | ERN, CRN, ΔERN, Pe, Pc, ΔPe, and standardized residual scores: all sex effects p > 0.05. Exact statistics not reported. | | | | | |
| Li  2026 | 100 (43) | 10–18  16 ± 1 | OCD HC  (CHN) |  | Resting state fc: fALFF, scan average, dynamic | | fALFF, Dx×sex. Left insula: ηp²=0.078 p=0.006; M OCD < HC d=0.75 t=2.62, pFDR=0.02, F p=0.24.  Right MCC: ηp²=0.114, p<0.001; F OCD > HC d=0.88, t=2.64, pFDR=0.02; M p=0.06.  Static FC, Dx×sex. Insula–SMG: ηp²=0.042, p=0.042; M OCD < HC d=1.08, t=3.75, pFDR<0.001; F ns.  Insula–DLPFC: ηp²=0.098, p=0.002; M OCD < HC d=1.12, t=3.90, pFDR<0.001; F ns.  Dynamic FC, Dx×sex. Insula–OFC: ηp²=0.060, p=0.016; M OCD < HC d=0.73, t=2.54, pFDR=0.03.  Insula–precuneus: ηp²=0.077, p=0.006; M OCD < HC d=0.83, t=2.89, pFDR=0.01.  MCC–precentral gyrus: ηp²=0.041, p=0.046; F OCD > HC d=0.85, t=2.57, pFDR=0.02.  MCC–precuneus: ηp²=0.088, p=0.003; F OCD > HC d=1.04, t=3.14, pFDR=0.004.  F OCD: MCC fALFF–CY-BOCS total ρ=0.80, p<0.001; MCC fALFF–compulsions ρ=0.59, p=0.02.  M OCD: insula–SMG sFC–CY-BOCS total ρ=−0.40, p=0.04; insula–SMG sFC–compulsions ρ=−0.44, p=0.02. | | | | | |
| *adults* | | | | | | | | | | | | |
| Park  2011 | 138 (48) | 18 +  26 ± 7 | OCD HC  (KOR) |  | corpus callosum (CC) thickness | | Unmedicated Dx x CC: p = 0.043;  M: Un OCD > HC, p = 0.032 | | | | | unmed OCD-M > OCD-F = HC-M = HC-F |
| Braber 2013 | 203 (131) | 18-60 | GEN /Sib  (NLD) |  | GM volume, YBOCS | | Left middle temporal gyrus (BA21): hOCs-M > lOCs-F = hOCs-F = lOCs-M, p <0.001;  Right middle temporal gyrus (BA39): hOCs-M = lOCs-F> hOCs-F > lOCs-M, p <0.001;  Right precunes (BA7): hOCs-M < lOCs-F < hOCs-F = lOCs-M, p<0.001 | | | | | |
| Real  2016 | 236 (112) | 18+ 35±10 | OCD HC  (ESP) |  | SLEs, GM volume, YBOCS | | nonSLE vs SLE F:M OR = 2.9 p<0.007;  SLE OCD > nonSLE OCD ~HC: right cerebellum p < 0.0005  nonSLE OCD > SLE OCD >HC: central tegmental tract, p < 0.05 | | | | | SLE: F > M  SLE →GM volume |
| Hawco 2016 | 131 (60) | 18+ 35±11 | OCD SCZ HC  (CAN) |  | WM Fractional anisotropy (FA), YBOCS | | The corpus callosum FA: OCD-F < OCD-M < HC: more in OCD and SCZ than in HC (DX x F p=0.036) but YBOCS F = M | | | | | OCD-F < OCD-M<HC-F = HC-M |
| Reess  2018 | 132 (92) | 32±10 | OCD HC  (DEU) |  | Hippocampus volume, YBOCS, OCI-R | | 3 clusters: (1) washing/obsessing; (2) ordering/ checking /obsessing, (3) obsessing/neutralizing;  M: (1) < (2) = (3) p = 0.035 | | | | | Contamination: F > M  Type of OCs→ hippocampal volume |
| Ma  2022 | 62  (31) | 18+ 29±7 | OCD  HC  (CHN) |  | Resting state fc, ALFF | | ALFF. OCD-F > OCD-M in right parahippocampal gyrus (F=28.48, p<0.001).  FC (seed parahippocampal Gyrus): OCD-F < HC to postcentral/ precentral/ STG/ paracentral/cuneus (p<0.01, AlphaSim); OCD-M > HC to right insula; OCDs × sex: significant (Wilks’ Λ=.196, F=4.60, p<0.001). | | | | | ALFF: OCD-F > OCD-M  Rs-fMRI: OCD-F ≠ OCD-M |
| Han  2023 | 228  (117) | 18+ 22±9 | non-med OCD  HC  (CHN) |  | GM volume  (VBM normative + latent factors) | | 4 latent factors, individual-specific weights x F.  (↑ bilateral thalamus, putamen, MTG, postcentral gyrus): F < M, d = −0.47, FDR p = 0.047  (↓ vmPFC, insula, mid-cingulum, MTG): F > M, d = 0.47, FDR p = 0.047  (↑ vmPFC, striatum, insula, MTG, occipital): F ~ M (ns)  (↓ right putamen, hippocampus, right inferior temporal gyrus): F ~ M (ns) | | | | | |
| Kapıcı 2023 | 116 (66) | 33–36 34±10 | OCD  HC  (TUR) |  | Incidental MRI findings | | Incidental finding. Fazekas WMH grades, CVI, CSP/CV, mucosal thickening, cysts), Y-BOCS OCD > HC: Fazekas-1 (17.2% vs 1.7%, p=0.002), CVI (20.7% vs 1.7%, p=0.001), generalized atrophy (p=0.008), ethmoidal thickening (p=0.004). Sex effects (OCD-M vs OCD-F): all comparisons ps > .05 (no differences; CVI raw p=0.021 but ns after Bonferroni) | | | | | |
| Mitiureva 2024 | 103  (83) | 18+  30±11 | OCD, MDD, HC  (RUS) |  | Resting state fc EEG; BDI | | Fc ~ Group*Sex*Frequency: DMN: F(2.71, 129.97) = 0.44, p =0.71. | | | | | |
| Xu  2024 | 110 (51) | 18–45 27±NA | OCD HC  (CHN) |  | Multimodal LICA (VBM, FA/MD, ALFF/fALFF/ReHo)  med records | | 4 latent components:  IC1 (94% VBM; widespread cortical GM): F < M, r = –0.42, FDR p = 0.011  IC15 (mixed structural–functional; frontal/temporal/cerebellar): F ~ M (ns).  IC23 (multimodal age-linked cortical decreases): F ~ M (ns).  IC8 (FA-dominated; striatum–thalamus–temporal WM tracts): F ~ M (ns). | | | | | |
| Wen  2025 | 206 (100) | 18–40 22±8 | OCD  HC  (CHN) |  | GM volume, indiv. z-maps | | Two subtypes: (1) ↑GMV in frontal/precuneus/insula/hippocampus/amygdala/temporal; (2) ↓GMV in frontal/precuneus/insula/superior parietal/fusiform/temporal.  GMV1: M > F χ²=16.161, p<0.001. | | | | | |
| Yi  2026 | 245 (115) | 18+  29 ± 6 | OCD  HC  (CHN) |  | VBM/SBM; cortical complexity, thickness, gyrification, sulcal depth, GMV | | Right paracentral lobule cortical complexity Dx×sex: F=22.70, pFWE=0.012; OCD: F<M, t(145)=−4.28, d=−0.71, p<0.001; HC: F>M, t(96)=2.50, d=0.51, p=0.014.  Other morphometric measures: no significant Dx×sex effects. | | | | | |
| ***Hormones and growth factors*** | | | | | | | | | | | | |
| *Children/adolescents* | | | | | | | | | | | | |
| Bilgiç 2021 | 117 (71) | 8-18 13±3 | OCD  HC  (TUR) | serum HPA axis: BDNF, GDNF, NGF, NTF3, ACTH, cortisol | | BDNF. OCD-M > HC-M (d = 0.88 p = 0.005), OCD-F > HC-F (d=0.37 p =0.002)  GDNF. OCD-M = OCD-F = HC-M = HC-F (d < 0.10 p > 0.50)  NGF. OCD-M = OCD-F = HC-M = HC-F (d < 0.15 p > 0.19)  NTF3. OCD-M = OCD-F = HC-M = HC-F (d < 0.14 p > 0.53)  ACTH. OCD-M > HC-M (d = 0.37 p = 0.04), OCD-F >= HC-F (d = 0.39 p = 0.106)  Cortisol. OCD-M > HC-M (d = 0.63 p = 0.046), OCD-F = HC-F (d = 0.001 p = 0.995) | | | | | | |
| *adults* |  |  |  |  | |  | | | | | | |
| Fontenelle 2012 | 80  (38) | 18-65 | OCD HC  (BRA) | OCI-R, serum BDNF, NGF, GNDF | | BDNF, NGF, GDNF: OCD < HC;  OCD: BDNF, NGF, GDNF x F: z=-1.69 p =0.91 | | | | M = F | | |
| Kawano 2013 | 120 (47) | 18 ≤ 33±13 | OCD HC  (JPN) | YBOCS, salivary a-amylase (sAA) cortisol | | sAA: F: OCD > HC p <0.01; M: OCD > HC, p < 0.01;  Cortisol: F: OCD=HC p > 0.1; M: OCD=HC, p > 0.1; | | | | F: OCD > HC  M: OCD > HC | | |
| Erbay 2015 | 60  (40) | 18-49  32±10 | OCD  HC  (TUR) | serum neurosteroid levels | | DHEA: OCD-F > HC-F (Δ=6.76 p= 0.009), OCD-M > HC-F (Δ=6.98 p= 0.037);  DHEA-S: OCD-M > HC-F (Δ=102.17 p= 0.027), HC-F < HC-M (Δ=-100.67 p= 0.030);  Cortisol: OCD-F > HC-F (Δ=4.76 p= 0.008), OCD-F~ HC-M (Δ=3.8 p = 0.14), OCD-M > HC-F (Δ=7.98 p< 0.001), OCD-M > HC-M (Δ=6.98 p=0.005).  Progesterone: OCD-M < HC-F (Δ=-4.54 p=0.041).  Testosterone: OCD-M < HC-M (Δ=-89.20 p<0.047), OCD-F < OCD-M (Δ=-344.76 p<0.001), OCD-F < HC-M (Δ=-433.96 p<0.001), HC-F < OCD-M (Δ=-358.14 p<0.001), HC-F < HC-M (Δ=-447.34 p<0.001) | | | | | | |
| Marazziti 2015 | 88  (42) | 18 ≤ 29±7 | unmed OCD  HC  (ITA) | Plasma Oxytocin Level (OT), ECR, YBOCs | | OT: OCD-M = OCD-F (d=0.22 p =0.46); OCD-M < HC-M (d=1.72 p <0.001); OCD-F > HC-F (d=0.73 p=0.025); HC-M =< HC-F (d=0.49 p = 0.11);  M: OT↑↑fearful -avoidant (r=0.35 p =0.05), dismissing (r=0.38 p =0.04) attachment | | | | OT: HC-M=< HC-F < OCD-F = OCD-M;  OT↑↑Attach: M only | | |
| O’Neill 2016 | 56  (25) | 18 ≤ 33±11 | OCD HC  (USA) | Metabolites: Glx, Cr, Cho, GAS, YBOCS | | Metabolite levels: OCD-M = OCD-F p > 0.05 | | | | | | |
| Melia  2019 | 180 (103) | 18 ≤ 41±18 | GEN  (ESP) | OCI-R; cortisol awakening response (CAR), diurnal cortisol slope (DCS), Holmes-Rahe Scale (HRS) | | CAR. OCI-R: β = 0.23 p = 0.06; OCI-R x F: β = -0.32 p = 0.02; OC-obsessing x F: β = -0.25 p = 0.03; OC-ordering: β = 0.365, p = 0.006; OC-ordering x F: β = -0.37 p = 0.02.  **M:** # SLE: r = 0.30 p < 0.01; HRS: r = 0.29 p = 0.02; **F:** OC-ordering: r = -0.22 p = 0.03;  DCS. **M:** washing: r = 0.24 p = 0.03, hoarding: r = 0.26 p = 0.02. | | | | | | |
| Labad 2021 | 155 (73) | 18 ≤ 43±13 | OCD  HC  (ESP) | YBOCS, DYBOCS; Awakening /fixed point diurnal cortisol slope (AWE/FPT DCS), the cortisol suppression ratio (DSTR) | | AWE DCS. OCD-M =HC-F = HC-M = OCD-F p = 0.34;  ordering/ symmetry: β = 0.57 p = 0.012; ordering/ symmetry x F: β= -0.68 p = 0.015.  FTP DCS. OCD-M < HC-F < HC-M < OCD-F p = 0.014;  FTP DCS. Hoarding β = -0.21 p = 0.23; hoarding x F: β = 0.42 p = 0.029.  DSTR. OCD-F < HC-M < OCD-M = HC-F p = 0.031. | | | | | | |
| Kılıç  2022 | 36  (21) | 31±9 | OCD  HC  (TUR) | Serum claudin-5 & zonulin (ELISA). | | Zonulin, pg/mL. OCD-F= 191.0 OCD-M= 110.7 d = 1.69; p < 0.001.  Claudin-5, pg/mL. OCD-F = 2556.1 OCD-M = 2222.0 d = 0.30; p < 0.10. | | | | | | |
| Marazziti 2023a | 50  (11) | 18–65  32±11 | OCD  (ITA) | serum vitamin D; Y-BOCS; HRSD | | Vitamin D. M=15.8, F=16.2; p=ns.  Vitamin D ~ Sx. M: “interference from obsessions r=−0.352, p=0.028, “distress from obsessions r=−0.381, p=0.017, “time spent on compulsions r=−0.357, p=0.026.  F: compulsion subscale r=−0.607, p=0.048, resistance to compulsions r=−0.637, p=0.035, control of compulsions r=−0.620, p=0.042, insight r=−0.774, p=0.005. | | | | | | |
| Marazziti 2023b | 40  (20) | 23–37  29±7 | OCD HC  PTSD  MDD  (ITA) | plasma oxytocin | | Plasma oxytocin. OCD-M = 5.81 OCD-F = 6.93 p > 0.10  HC-M = 5.02 HC-F = 6.46 p < .05 | | | | | | |
| Mina  2023 | 90  (39) | 18+  28±11 | OCD, FDR, HC  (IND) | serum BDNF (ELISA); Y-BOCS; DY-BOCS | | BDNF, pg/mL. OCD-M = 1441.71 ± 294.71, OCD-F = 1301.92 ± 372.85, p = 0.26 | | | | | | |
| Sarmin  2024 | 58  (24) | 18–60 | OCD  HC  (BGD) | serum IL-1β/IL-6 (ELISA), YBOCS | | IL-1β, pg/mL OCD-M = 25.41 HC-M = 15.02 p = 0.006, OCD-F = 21.28 HC-F = 16.57 p = 0.14; OCD-M vs OCD-F p > 0.10.  IL-6, pg/mL. OCD-M = 45.51 HC-M = 37.43 p < 0.001, OCD-F = 44.35 HC-F = 36.69 p < 0.001; OCD-M vs OCD-F p > 0.10. | | | | | | |
| Salimi 2026 | 140 (56) | 18–65 40±10 | SCZ BMD (IRN) | Serum inflammatory markers, OCI-R | | GLMtree, IL-17A→OCS differed by sex, p=.050. M: β=−0.471, p=.148; F: β=−0.135, p=.540. CCL20 sex effect not reported. | | | | | | |
| Algin  2025 | 300 (141) | 18–60  27 ± 9 | OCD  (BGD) | serum Vitamin B12, Folate, Homocysteine; Y-BOCS; DOCS | | Vitamin B12. F: β = 96.1 p < 0.001.  Folate. F: β = 1.09 p = 0.023.  Homocysteine. F: β = –4.97 p < 0.001. | | | | | M > F: Homocysteine,  M < F: Vitamin B12, Folate | |
| Marazziti  2025 | 42  (13) | 18+ | ASO+ OCD (Italy) | Folate | | Folate: F > M, Z=2.37, p=0.018. | | | | | Folate in ASO+ OCD: M < F | |
| Okumuş 2026 | 62F | 10–18  15 ± 5 | OCD, TTM, HC (TUR) | Plasma untargeted metabolomics; LC-Q-TOF/MS | | OCD vs HC: linoleic acid FC=0.02, p=3.34×10⁻⁵; trihexosylceramide FC=15.48, p=2.37×10⁻⁷; 7α,12α-dihydroxy-cholestene-3-one FC=4.33, p=1.34×10⁻⁴.  TTM vs OCD: linoleic acid FC=51.39, p=.0016; arachidonic acid FC=3.55, p=.0197; biocytin FC=0.10, p=.0146. | | | | | F: OCD vs HC: ↓ linoleic acid; ↑ trihexosylceramide, 7α,12α-dihydroxy-cholestene-3-one. OCD and TTM show distinct metabolic profiles. | |
| ***Single gender studies*** | | | | | | | | | | | | |
| *Cognition* | | | | | | | | | | | | |
| Egan 2013 | 226M | 18-62 34 ± 9 | GEN  (GBR) | Online Porn Use, Personality, Obsessionality, Compulsivity | | Neuroticism. Checking: β_std_ = 0.44, p < 0.001; Cleaning: β_std_ = 0.20 p < 0.01;  Extraversion. Checking: β_std_ = -0.18 p < 0.01;  Internet addiction. Checking: β_std_ = 0.33 p <0.001; Cleaning: β_std_ = 0.21 p <0.01; Pornography use. Checking: β_std_ = 0.16 p <0.05; sexual addiction: β_std_ = 0.33 p <0.001;  Checking. Sex addiction β_std_ = 0.20, Internet addiction β_std_ = 0.34, Addiction β_std_ = 0.50, controlling for Big-5 | | | M:  Sex/ Internet addiction ~ checking;  Checking ~ Neuroticism | | | |
| Claus  2026 | 447 F | 18–30  23 ± 3 | GEN  (DEU) | Perfectionism ~ OCs | | Network Analysis, baseline (BL) + 6m f/u. perfectionistic concerns ↑↑ obsessions: nonzero edge at BL/ 6m; perfectionistic strivings ↑↑ ordering: nonzero edge at BL/ 6m. Each edge was stronger than ≥10% of network edges; BL–6m edge changes p > 0.05. Exact stats not reported. | | | F: perfectionism ↑↑ obs/order – temporary stable | | | |
| Claus  2025 | 499F | 18–30  23 ± 3 | GEN  (DEU) | Perfectionism → OCs | | 3-wave SEM (baseline, 6m, 12m). Perfectionistic concerns → later OCs: β = 0.10 [95% CI, 0.04–0.17], p < 0.01. Perfectionistic strivings → later OCs: β = −0.10 [95% CI, −0.20 to −0.01], p < 0.05. OCs → later perfectionistic concerns: β = 0.00 [95% CI, −0.03–0.03], p = 1.0. OCs → later perfectionistic strivings: β = 0.01 [95% CI, −0.01–0.03], p = 0.29. | | | F: Perfectionistic concerns ↑↑ later OCs; perfectionistic strivings ↑↓ later OCs; OCs ↛ later perfectionism. | | | |
| *Behavior* | | | | | | | | | | | | |
| *children* |  |  |  |  | |  | | |  | | | |
| Bohon  2019 | 49F | 12-19 | OCD  HC, AN  (USA) | WCST during fMRI scan, C-YBOCS | | Perseverative errors: OCD > HC, AN: p < 0.047, d > 0.81 | | | F:  executive function OCD < HC | | | |
| *adults* |  |  |  |  | |  | | |  | | | |
| Abramovitch  2011 | 60M | 18 – 60  32±8 | OCD, HC  (ISR) | Broad neurocognitive functioning, Mind-streams battery, MINI | | M-OCD < M-HC: Composite Performance Index d = 2, Attention d = 1.4, Executive Functions d=1.39, Information Processing Speed d = 1.33, Motor skills d = 1.28, Verbal Function d = 1.17, Memory d = 1.05, Visual-Spatial d = 1; ps < 0.0001. | | | M:  Attention, Executive Functions, Information Processing, Memory, Visual-Spatial, Verbal and Motor skills OCD < HC | | | |
| Boisseau 2013 | 57F | 18 ≤ (23±5) | OCD  ED  (USA) | decision making, IGT | | OCD-F ~ HC-F, Stats not reported | | | F:  decision making OCD ~ HC | | | |
| Altman  2013 | 114F | 18 – 43  20±3 | GEN  (USA) | Startle response Picture-viewing task, EDI-3, SCOPI | | EMG startle response. *Contam. img:* SCOPI-contam. β = 0.22, t (113) = 2.36, p < 0.05, SCOPI-contam. x EDI β = 0.21, t (113) = 2.30, p < 0.05; *Gen. threat img:* SCOPI-contam. β = 0.23, t (113) = 2.47, p < 0.05, SCOPI-contam. x EDI β = 0.19, t (113) = 2.00, p < 0.05; *Food images:* SCOPI-contam. & SCOPI-contam. x EDI p > 0.10. | | | F:  Startle response + Sx provoc. & Startle response + gen. threat OCD+BN > OCD | | | |
| Sternheim  2016 | 228F | 18-35  22±2 | (NLD) | OCI-R, BCST, DFlex | | SubOCs > HC, effect size η**.** BCST: total errors = 0.08 p = 0.01; category completed = 0.05 p = 0.06, perserv. Error = 0.08 p = 0.01. | | | F: Cogn. Inflex ↑↑ subOCs | | | |
| *Neurophysiology* | | | | | | | | | | | | |
| *children/adolescents* | | | | | | | | | | | | |
| Buse 2016 | 50M | 11- 17 15±2 | OCD  HC  (DEU) | The harmonic expectancy violation paradigm during fMRI session, OCTCDQ | | Harmonic vs. disharmonic chord sequences reversal effect (p < 0.001):  the hippocampus and the parahippocampal gyrus (BA 27): OCD < HC  the precuneus (BA 7): HC < OCD  the superior temporal/inferior frontal cluster (BA 22/ BA 47): opposite in HC and OCD | | | | | | |
| Carlisi 2017 | 64M | 11-17 (15±2) | OCD  ASD HC  (GBR) | IGT, fMRI | | Choice sensitivity HC > OCD (mean HDI = 2.83, [95% HDI, 1.44- 4.22], p < 0.001); reinforcement learning weights (ω) HC> OCD (mean HDI = 0.71[95% HDI, 0.45 -0.97], p < 0.001).  Choice, safe vs. risky: Left DLPFC, right IFG/insula, left IFG, left IFG/insula: HC > OCD p < 0.01. | | | | | | |
| Becker 2023 | 93F | 8–21 14±4 | OCD Anx. HC (USA) | rs-fMRI, OCI-R, CYBOCS, MASC | | Rs-fc. Within-Cingulo-Opercular: OCD > ANX (p=0.003), OCD > HC (p=0.027); Cingulo-Opercular –CSTC: OCD > ANX (p=0.004), OCD > HC (p=0.043); | | | | | | |
| *adults* |  |  |  |  | |  | | | | | | |
| Lee 2017 | 12M | 18 + 25 ± 5 | EO/LO OCD  (KOR) | plasma escitalopram during [11C] DASB PET scan; self-reports | | EO OCD >LO OCD: Putamen U = 4 p = 0.026; dorsal raphe nucleus (DRN): U = 7, p = 0.093; SERT BP ~ age of onset: Putamen r = −0.580, p = 0.048; but not with the duration of illness: r = 0.422, p = 0.172 | | | | | | |
| Serrano 2019 | 30F | 18 + 22±4 | GEN  (ESP) | BAT with stimulus evoking disgust; heart rate (HR); subjective reports (SR) of emotions;  OCI–R | | HR. time x Dx η^2^_p_ = 0.16 p = 0.04; HR decreased pre task→task subOC-cont > HC;  SR during BAT. subOC > HC: anx (η^2^_p_ = 0.38 p <0.001), urge to vomit (η^2^_p_ = 0.19 p 0.03), urge to wash (η^2^_p_ = 0.56 p <0.001), contam (η^2^_p_ = 0.39 p = 0.001), disgust (η^2^_p_ = 0.31 p = 0.003);  subOC group. contam η^2^_p_ = 0.17 p = 0.02 & disg η^2^_p_ = 0.22 p =0.006, valence η^2^_p_ = 0.35 p = 0.002, dominance η^2^_p_ = 0.16 p = 0.04 increased during task; HR variability ~ need to wash r > 0.54 p < 0.05; HFnu ~ control r > 0.53 p < 0.05 and activation r = 0.56 p < 0.05 | | | | | | |
| Lee  2021 | 31M | 18 +  23±2 | GEN  (KOR) | TAF-task, fMRI, TAFS, OCI-R, DOCS, BDI | | Task-based fMRI. Negative-TAF > Positive-TAF: ↑ precuneus (BA7), mPFC/SFG/SMA, insula, caudate, globus pallidus, thalamus, midbrain, IPL, STG/MTG, lateral OFC (FDR p<.05); Common PS+NS: fusiform/lingual gyri, vlPFC/dlPFC/dmPFC, mid-cingulate, TPJ, IPL/MTG; activity in precuneus ↑ with TAFS; inferior OFC/insula/SFG/MFG/mPFC/IPL ↑ with DOCS (inferior OFC survives Bonferroni). | | | | | | |
| Çınaroğlu  2025 | 71M | 27±5  (18+) | BDD/MD, TUR | OCI-R, skin conductance | | Comorbid OCS, OCI-R. In BDD/muscle dysmorphia: OCI-R~ correlated with skin conductance level ρ = 0.73 and response frequency ρ = 0.76, ps < 0.05. | | | | | | |
| *Hormones and growth factors* | | | | | | | | | | | | |
| Lord  2012 | 39F | 18+ | PP 6w-12w OCD,  HC  (CAN) | MIST, saliva cortisol | | Cortisol stress response**:** PP- OCD > PP HC~ OCD: t(26) = 2.0 p,p = 0.05 | | | | | | |
| Korkmaz  2025 | 42F | 18–65  39 ± 13 | OCD, HC  (TUR) | Plasma miR-132, miR-21-5p, miR-125b-5p; serotonin, dopamine; Y-BOCS | | Plasma microRNAs and serotonin, OCD > HC, serotonin p=0.046; dopamine p > 0.05; miR-132 p=0.006, AUC=0.75, [95% CI, 0.59, 0.86]; miR-21-5p p=0.077, AUC=0.66, [95% CI, 0.496, 0.804]; miR-125b-5p p<0.0001, AUC=0.89, [95% CI, 0.75, 0.96]. Within OCD, dopamine–Y-BOCS: r=0.44, p=0.04. | | | | | | |

**Abbreviations used in Table A.6:**

Instruments: **ACT –** Auditory Consonant Trigram test ^243, 244^; **Agree** – Agreeableness, subscale of the NEO Five-Factor Inventory^245^; **ANT-R** – Attention Network Test–Revised (Bednarek 2024); **BAT** - behavioral avoidance task^246^; **Big-5** - Revision of the NEO Five-Factor Inventory^245^; **CANTAB battery** - the Cambridge Automated Neuropsychological Test Battery^247^; **CASI** - Child Anxiety Sensitivity Index^248^; **COWAT** – Controlled Oral Word Association Test (Mısır 2025); **CY-BOCS** - the Children’s Yale-Brown Obsessive-Compulsive Scale^17^; **DERS-modf** – modified Difficulties in Emotion Regulation Scale^249^; **DMS** - delayed matching to sample task from CANTAB battery^247^; **DOCS** - Dimensional Obsessive-Compulsive Scale^25^; **DOCS**-**resp** - responsibility for harm subscale of DOCS^25^; **DOCS-unaccept** – unacceptable thoughts subscale of DOCS^25^; **DTS -** Distress Tolerance Scale^250, 251^; **DYBOCS** - The Dimensional Yale–Brown Obsessive–Compulsive Scale^29^; **ECR** - Experiences in Close Relationships’ questionnaire^252^; **ECR-R** - Experiences in Close Relationships Scale – Revised^253^; **EDI-3 (EDI)** - Eating Disorder Inventory-3^254^; **GAS** - Global Assessment Scale^176^; **HDDM** – hierarchical drift diffusion model; **HDRS-17** – Hamilton Depression Rating Scale, 17-item version (Mısır 2025); **HGPT** - Handgrip dynamometer endurance task^255^; **HRS** - Holmes-Rahe Scale^256^; **IAPS** - International Affective Picture System^257^; **IGT** – Iowa Gambling Task^258^; **LCS** - Locus of Control Scale^259^; **MASC** – Multidimensional Anxiety Scale for Children (Becker 2023); **MCQ-C30** (**MCQ-C**)- Metacognitions Questionnaire, a 30-item version^260^; **MINI** - the Mini-International Neuropsychiatric Interview^261^; **MIST** - Montreal Imaging Stress Task.^130^ **MOCSL** - Maudsley Obsessive-Compulsive Symptom Checklist^262^; **MTPT** - mirror tracing persistence task^263^; **NEO-FFI** - NEO Five-Factor Inventory^264^; **Neuro** – neuroticism subscale of NEO Five-Factor Inventory^264^; **OC- modf** – modified OC subscale of Symptom checklist‐90‐revised ^265^; **OCI-CV** - Obsessive Compulsive Inventory-Child Version^52^; **OCI-R** - The obsessive–compulsive inventory-revised^53^; **OCTCDQ** - The Obsessive–Compulsive Trait Core Dimensions Questionnaire —Revision^266^ **Ordered CGT** – Cambridge Gambling Task,^267^ with trials ordered by increasing level of risk ; **RCADS** - Revised Child Anxiety and Depression Scale^64^; **RTQ** – Repetitive Thinking Questionnaire^268^; **SCARED-R** - revised screen for child anxiety related emotional disorders^144^; **SCID-I** - the Structured Clinical Interview for DSM-IV^68^; **SCL-90-R** - Symptom checklist‐90‐revised^265^; **SCOPI** - Schedule of Compulsions, Obsessions, and Pathological Impulses^269^; **SCOPI-contam** – contamination subscale of the Schedule of Compulsions, Obsessions, and Pathological Impulses^269^; **SLEs** – The Paykel Scale of Stressful Life Events^270^; **SRM** - spatial recognition memory task from CANTAB battery^247^; **SWM** - spatial working memory task from CANTAB battery^247^; **TAFS** – Thought-Action Fusion Scale (Lee 2021); **PVDM** – perceptual-value decision making task; **WCST** - Wisconsin card sorting test^271^; **YBOCS** – the Yale-Brown Obsessive Compulsive Scale^82, 83^.

Other abbreviations: **βstd –** standardized parameter estimate; **95% HDI –** contains 95% of the distribution of the value of the parameters; **ACTH -** adrenocorticotropic hormone

**AI -** affect intolerance; **ALFF** – amplitude of low-frequency fluctuations; **AlphaSim (α)** – cluster-level correction method; **AN –** anorexia nervosa; **ANX -** anxiety ; **ASD -** asparagus spectrum disorder; **Attach –** attachment; **Attach anx –** attachment anxiety; **AWE DCS -** awakening diurnal cortisol slope; **BA –** Broadman Area; **BDNF -** brain-derived neurotrophic factor; **CAR -** cortisol awakening response; **CC -** corpus callosum; **Cho -** choline-compounds; **Contam –** contamination obsessions; **Contam. Img –** images provoking contamination symptoms; **Cr -** creatine + phosphocreatine; **CSP** – cavum septi pellucidi; **CSTC** – cortico-striatal-thalamo-cortical circuit; **CVI** – cavum veli interpositi; **d –** effect size Cohen’s d; **dB –** decibels; **DCS -** diurnal cortisol slope; **DEP –** depression; **DHEA –** dehydroepiandrosterone; **DHEA-S -** dehydroepiandrosterone sulfate; **Disg –** disgust; **DLPFC –** dorsolateral prefrontal cortex; **DSTR -** cortisol suppression ratio after dexamethasone; **DSTR -** the cortisol suppression ratio; **Dx –** diagnosis; **ED -** eating disorder; **ELISA** – enzyme-linked immunosorbent assay; **EO OCD –** early onset OCD; **ES –** effect size, mean percent PPI differences/ the standard deviation of percent PPI in the overall sample; **FA -** Fractional anisotropy; **FA –** WM Fractional anisotropy; **fALFF** – fractional amplitude of low-frequency fluctuations; **Fc –** functional connectivity; **fMRI –** functional magnetic resonance imaging; **FPT DCS -** fixed point diurnal cortisol slope; **FTP DCS -** fixed point diurnal cortisol slope; **GDNF -** glial cell line-derived neurotrophic factor; **GEN –** general population; **Gen. threat img –** general threat image condition in the picture viewing task; **Glx -** glutamine/glutamate; **GM/ WM volume –** gray matter/white matter volume; **GNDF -** glial cell line-derived neurotrophic factor; **GoalD -** goal directed activity; **HC –** healthy control; **HDI -** highest density interval; **HFnu -** normalized high frequency; **hOCS –** high severity of obsessions and compulsions; **HPA axis -** hypothalamic-pituitary-adrenal axis; **HR -** heart rate; **IFG –** inferior frontal gyrus; **IL-1β** – interleukin-1 beta; **IL-6** – interleukin-6; **ImpC -** impulse control; **LICA** – linked independent component analysis; **LO OCD –** late onset OCD; **LOC –** locus of control; **lOCS –** low severity of obsessions and compulsions; **MD** – mean diffusivity; **MFG** – middle frontal gyrus; **MTG** – middle temporal gyrus; **n.s. –** non-significant; **NGF -** nerve growth factor; **NJRE -** not-just-right experiences; **nonSLE OCD –** no stressful event preceding OCD; **NTF3 -** Neurotrophin-3; **Obss –** obsessions; **OCD –** obsessive compulsive disorder; **OCD+BN –** comorbid OCD and Bulimia nervosa; **OCS –** obsessions and compulsions; **OR -** odds ratio; **OT -** plasma oxytocin level; **PCC** – posterior cingulate cortex; **PET -** positron emission tomography; **pg/mL** – picograms per milliliter; **Plasma [11C] DASB -** [(11) C]-3-amino-4-(2-dimethylaminomethyl-phenylsulfanyl)-benzonitrile; **Plasma OT -** plasma oxytocin level; **pOCD –** provisional OCD (based on elevated self-reported obsessions and compulsions); **pOCD/DEP –** comorbid provisional OCD and depression; **PPI -** pre-pulse inhibition; **ReHo** – regional homogeneity; **REP -** repetitive thinking; **sAA -** salivary a-amylase cortisol; **SAD –** social anxiety disorder; **SCZ –** schizophrenia; **SERT BP -** serotonin transporter binding potential; **Sex/ internet addiction –** sexual and porno internet addiction; **Sib –** siblings; **SLE OCD –** stressful event preceding OCD; **SM** – sexual minority; **SMA** – supplementary motor area; **SR -** subjective reports; **STG** – superior temporal gyrus; **STN** – subthalamic nucleus; **STU –** college students; **subOC-cont –** subclinical contamination‒OCD; **Sx** – symptoms; **Sx provoc. –** symptom provocation; **TAF** – thought–action fusion; **TPJ** – temporoparietal junction; **Unmed (un) –** unmedicated; **VS storing and recognition –** visuospatial storing and recognition; **VSM -** visuospatial memory; **VSWM -** visuospatial working memory;

**M** – male; **F** – female.

**Appendix 7.** Papers that addressed gender differences in treatment use and response individuals with obsessive-compulsive disorder.

Table A7. Gender differences in treatment of OCD.

| **First auth**  **year** | **N**  **(F)** | **Age**  **mean** ± **SD**  **(range)** | **Pop.** | **Design** | **Therapies’ details** | **Treatment variables of interest** | **Gender effects stats (F/M)** | |
| --- | --- | --- | --- | --- | --- | --- | --- | --- |
|  |  |  |  |  |  |  | **Stats** | **Direction** |
| Naturalistic treatment | | | | | | | |  |
| *all ages* | | | | | | | |  |
| Isomura  2016 | 10523  (5998) | all ages | SWE | medical records | person therapy and meds | Prescribed, F:M OR:  SRI=1.27 [95%CI, 1.12-1.44], non-benzo anxiolytics = 1.26 [95%CI, 1.16–1.38], benzo sedatives = 1.21 [95%CI, 1.10–1.33], analgesics = 1.64 [95%CI, 1.49–1.80]  AAP. = 0.68 [95%CI, 0.61-9.76], 1^st^ gen antypsych. = 0.86 [95%CI, 0.75-0.98],  Other: SNRI, non-SRI/SNRI antidepr., benzo anxiolytics, non-benzo sedatives, antiepileptics ~ 1 | | **SRI:**  **M < F**  **AAP:**  **M > F**  Other:  **M ~ F** |
| *children/adolescents* | | | | | | | |  |
| Rudy  2014 | 78  (38) | 13 ± 3  (7 – 18) | USA | observ | Daily CBT up to 90-min; 14 sessions in 3w, include family; % on meds NA | Responder: GCI-I =1-2  Improve: post-T C-YBOCS | Responder: **F** OR=0.33 [95%CI=0.13-0.86]  Improve: **F:** β= 0.32 t=2.82 p<0.05. **F- M:** d = 0.69 p<0.01 | **M > F** |
| Højgaard  2020 | 314  (157) | NA  (13 – 17) | USA | observ | ERP + person meds; 75.43±34.24 d (5-213 d) | Responder: C-YBOCS-SR ≤15 + ΔC-YBOCS-SR≥7.15  Improve: post-T C-YBOCS-SR | Responder: **M:** β>−0.19 OR>0.83, p>0.45  Improve: **M:** β>−0.046 p>0.47 | **M ~ F** |
| Conelea  2023 | 36  (18) | 12 ± 3  (≤17) | USA | observ | intens ERP and meds;  (5d/w, 4h/d; ~40d) | Improve: ΔCYBOCS **F** vs. **M**: d = 0.22 p =0.5  Baseline SSRT → ΔCYBOCS: **F** R²=0.53 p<0.001; **M** R² =0.01 p=0.72  ΔSSRT → ΔCYBOCS: **F** R²=0.40 p=0.01, **M** R² =0.01 p=0.76 | | base SSRT/ ΔSSRT → improve:  **F** only |
| Franklin  2024 | 1286 (784) | 14 ± 2  (7-17) | USA | open label | IOP: ~3 hours/day, 5 d/week; PHP: ~6 h/day, 5 d/week, person meds | Responder: ΔCYBOCS-SR ≥35%  Remission: CYBOCS-SR ≤12 | Responder ~ F: β=0.09 p =0.54; Remission ~ F: β=0.09 p =0.55 | **M ~ F** |
| Kardaş  2024 | 50  (22) | 14 ± 2  (12–18) | TUR | observ | daily ERP + GCBT + family work + meds (SSRIs, antipsych); mean stay 8.3 w | Responder: CGI-S<5, YBOCS≤15, MOCI<18, CGAS>60;  Improve: Δ YBOCS | Responder: F=64% M=61% χ² p = 0.83  Improve: F=14.6 ± 9.7 M= 13.3 ± 8.7 p = 0.61 | **M ~ F** |
| *adults* | | | | | | | |  |
| Eisen  2013 | 213  (120) | 40 ± 13  (18 +) | USA | observ | 3+ annual assess, in person/ by phone | Remission: LIFE OCD = 1-2 for 8w; Relapse: LIFE OCD = 4-6 for 4w. | Remission: **F** HR=0.71 [95%CI=0.45,1.11]  Relapse: **F** HR=1.20 [95%CI=0.68,2.11] | **M ~ F** |
| Semiz  2013 | 120  (93) | 35 ± 10  (19 – 62) | DEU | observ | 10 w person treatment, 1yr follow up | Responder: Δ YBOCS≥ 35 % for ≥12 mo | Responder:  β_M_ = -3.9 Wald = 2.34, p = 0.13, controlling for dur, anx, depress, relig, insight, dissoc, trauma | **M ~ F** |
| Batmaz  2016 | 180  (109) | 33 ± 12  (18 +) | TUR | retro observ | person meds | A-OCD vs. R-OCD: F: M OR = 0.31 [95%CI, 0.16-0.60]  Prescription: AAP β_A-OCD_ = 0.83 p = 0.025 OR = 2.3 [95%CI, 1.11-4.77], β_F_ = -0.17 p = 0.64 OR = 0.85 [95%CI, 0.42-1.70]; A-OCD x F not tested | | **A-OCD:**  **↑↑AAP**  **↑↑M** |
| Lisoway  2018 | 112  (67) | 37 ± 11  (18 +) | CAN | observ | person meds, ≥ 8w | Responder: CGI-I = 1-2 | *“The mean gender did not differ significantly between responders and nonresponders (p > .05).”* | **M ~ F** |
| Falkenstein  2019 | 305  (139) | 30 ± 11  (18 +) | USA | observ | 4h ERP daily + symptom speciﬁc groups + behavior family therapy, psychiatrists; 95% on med | Subtypes: YBOCS trajectories over time. | **F** proportion across subtypes: 0.46:0.46:0.46  χ2 = 0.002 p= 0.999 | **M ~ F** |
| Albert  2019 | 251  (116) | NA  (18 +) | ITA | observ | person meds | Responder: Δ YBOCS≥25%  Improve: post-T YBOCS | Responder: **F:** β=0.31 p=0.27  Improve: **F:** β=-1.22 p=0.16 | **M ~ F** |
| Samuels  2019 | 705  (405) | 37 ± 13  (18 – 75) | USA | observ | CBT or SSRI; no details | Responder: SR = “poor” / “good”. Control for personality traits | **M:** OR= 1.10 [95CI=0.77–1.57] | **M ~ F** |
| Nanjundaswamy 2020 | 58  (24) | 30 ± 12  (17+) | IND | observ | person ERP and meds | Responder: ≥35% ΔYBOCS | **M:** OR = 1.15; χ2=0.07 p=0.79 | **M ~ F** |
| Bezahler  2022 | 191  (95) | 29 ± 10  (18+) | USA | observ | IRT: 4h ERP/day +family+meds+group  mean stay = 62 ± 23d | Improve: pre-post Y-BOCS | Sexual orient minority vs heterosex: $\eta_{p}^{2}=0.017$ p=0.07 | sex minority ≥ heterosexual |
| CBT studies | | | | | | | |  |
| *children/adolescents* | | | | | | | |  |
| Olino  2011 | 41  (22) | 12 ± 3  (6 – 17) | USA | open label | Up to 4 per w, up to 2.25h; Up and Down the Worry Hill protocol | Improve: Post-T DYBOCS subscales | M: βϵ[-0.84 - 1.11] across subscales p > 0.05 | **M ~ F** |
| Torp  2015 | 269  (131) | 13 ± 3  (7 – 17) | DMK, SWE,  NOR | open label | Exposure based CBT: 14, weekly, 75min; in 8s – 30min with parents, in 6s 75min with parents | Post-T C-YBOCS | **M:** β<0.35 p>0.75 | **M ~ F** |
| Højgaard  2018 |  |  |  |  |  | Responder: CY-BOCS ≤15  Improve: ΔC-YBOCS, Control for comorbidities | Responders: **M:** β=-0.47 p = 0.108  Improve: **M:** β= -.005 p = 0.996 | **M ≤ F** |
| Storch  2018 | 142  (74) | 12 ± 3  (7-17) | USA | open label | CBT + ERP: 10 weekly sessions | QoL – kid: PedsQL, COIS-C  QoL parent: CBCL, COIS-P | social QoL-kid & parent total QoL: n.s. (no stats reported) | **M ~ F** |
| Peris  2020 | 62  (35) | 13 ± NA  (8 – 17) | USA | open label | Child: 12 weekly 1h ERP. Family: CBT - 30 min E weekly; EFT – bi-weekly 1h, distress tolerance, problem- solve. | Responder: CGI-I =1-2  Improve: ΔC-YBOCS | Responder:  F p = 0.76; F x T p = 0.22  Improve:  F p=0.97, F x T p = 0.86 | **M ~ F** |
| Duholm  2024 | 147  (68) | NA  (7-17) | SWE, NOR, DEN | open label; 6, 12, 24, 36 mo f/up | Child: 14 weekly exposure based CBT, 16 CBT +/- SSRI for non-responders | # contamination over time ~ F: n.s. (stats not reported) | | **M ~ F** |
| Belschner 2025 | 98  (53) | 14 ± 3  (8–19) | CAN | Naturalistic longitudinal open-label | 12-wk group-based family CBT + ERP; weekly 1.5-h sessions; endpoint (12 wk) | Response = ≥35% ↓ CY-BOCS baseline→EOT; remission = ≥55% ↓ CY-BOCS baseline→f/up or CY-BOCS ≤11 | Completers vs noncompleters: F 54.2% vs 54.1%, p=.206. No gender effect on CBT response/remission reported. | **M ~ F** |
| *adults* | | | | | | | |  |
| Maher  2010 | 108  (46) | 39 ± 14  (18 – 70) | USA | open label, controlled | ERP: 2 planning + 15 exposure. SMT: 2 intro +15 training; 2 weekly, for 90–120 min + HW | Improve: Post-T YBOCS | **F x T:** β=5.60 (2.14) p=0.01;  **M-F** (ERP-SMT) = 5.50; d ≈ 0.8 p=0.01 | **M > F** |
| Safak  2014 | 29  (NA) | 32 ± 10  (14 - 65) | TUR | open label | GCBT: 4 weekly 90-120-min | Improve: ΔYBOCS | *“the difference between the genders was not significant”* | **M ~ F** |
| Sunde  2019 | 40  (NA) | 44 ±NA  (18 +) | NOR | open label | ERP: 12 weeks 2.5 h; ~8 yr follow up | Responder: YBOCS ≤14 & ΔYBOCS ≥8 | *“no significant differences with respect to gender (p = 1.00) between the recovered and the non-recovered group.”* | **M ~ F** |
| Blakey  2019 | 90  (44) | 29 ± 11  (18 +) | USA | open label | ERP: inpatient, 60±30 days, + additional CBT as needed | *A series of independent samples t-tests did not detect a significant effect of gender on any study variables (all ps N .15).”* | | **M ~ F** |
| Wootton  2024a | 157  (123) | 35 ± 11  (18–64) | worldwide | open label | iCBT: 5 mod over 8 w: psychoedu, behavioral experiments, arousal reduction, ERP, relapse prevention; no clinician contact | Intent-to-Treat (ITI): all enrolled Responder: ≥35% ΔYBOCS | ITI ~ F: β ≈ −0.05 p = 0.39  Responder: n.s. (stats not reported) | **M ~ F** |
| Wootton  2024b | 216  (157) | 34 ± 13  (18–78) | worldwide | open label |  | Improve: YBOCS, post, 3m | post ~ F: expβ = -7.5% p = 0.015; 3mo ~ F: expβ = 7.0% p = 0.075 | post: **M < F**  3mo: **M ≥ F** |
| Tjelle  2024 | 163  (117) | 35 ± 11  (18+) | NOR | open label | Bergen 4-Day ERP, 8 hour per day; for refractory OCD | Improve: Y-BOCS (post, 3-month, 1-year), WSAS (1year) | 12mo Y-BOCS ~ F:  β = −0.01, p = 0.96  12mo WSAS ~ F:  β = −0.10 p = 0.33 | **M ~ F** |
| Wootton 2025 | 323  (235) | 33 ± 12 (18–78) | Various | Retro chart review of open label trials | Self-guided internet-delivered CBT (ICBT); 5 modules over 8 weeks | DOCS, YBOCS, DIAMOND improvement | Gender was not significant for any dropout definition: early dropout p=.525; treatment commencement p=.851; treatment completion p=.115; post-treatment questionnaire completion p=.413 | **M ~ F** |
| SSRIs - studies | | | | | | | |  |
| *children/adolescents* | | | | | | | |  |
| Blazquez  2014 | 74  (50) | NA  (10-17) | ESP | open label | FLX in OCD and GAD: 10 mg/d 1w→ 20 mg/d 2-8w; low improve → 40 mg/d; no improve→60 mg/d 8-12 w. | Improve: ΔC-YBOCS  Meds serum concentration to respond: FLX, NORFLX | FLX. w8: **M** = 61.7±52.3, **F** = 111.4 ± 92.9 d= 0.66 p =0.02; w12: **M** = 82.2±85.8, **F** = 154.3 ± 150.2 d= 0.53 p = 0.03.  NORFLX. w8: **M** = 82.6±54.1, **F** = 128.5 ± 96.6 d= 0.53 p =0.03; w12: **M** = 98.2±64.5, **F** = 150.7 ± 108.9 d= 0.53 p = 0.03.  FLX+ NORFLX. w8: **M** = 144.3±100.8, **F** = 237.9 ± 172.8 d= 0.60 p =0.02; w12: **M** = 180.4±132.0, **F** = 304.9 ± 240.8 d= 0.58 p = 0.02. | FLX  **M < F** |
| Cifter  2022 | 102  (43) | 12 ± 3  (6-17) | TUR | open label | SRI (personalized): ≥3 mo | Responder: CGI-I = 1–2 | Responder: F = 69.5%, M = 62.8%; χ²(1) = 0.48, d = 0.14 p = 0.50 | **M ~ F** |
| Bharthi  2024 | 90  (56) | 14 ± 3  (7–18) | CAN | naturalistic, retrospective report | FLX: mean = 152 d; mean = 41 ± 22 mg/d | Improve: yes/no  Side effect: yes/no | Improve: M : F OR = 2.4 [95% CI, 0.83–6.98], p = 0.108;  Side effects: M : F OR = 0.3 [95% CI, 0.08–1.16] p=0.08 | FLX:  **M ≥ F;**  Side/eff:  **M ≤ F** |
| *adults* |  |  |  |  |  |  |  |  |
| Selvi  2011 | 57  (37) | 32 ±10  (18 – 65) | TUR | open label | Meds increase 4w; at max 8w. Sertraline: 50mg/day→200mg/day, Paroxetine: 20mg/day→ 60mg/day; FLX: 20mg/day→80mg/day | Responder: ≥35% ΔYBOCS | *Ꭓ*^2^(1) =0.41 p = 0.52;  M OR=0.63 | **M ~ F** |
| Marazziti  2012 | 20  (10) | 30 ± 8  (18+) | ITA | open label | FLX: person to achieve improvement; 4w, 7w | Improve: ΔYBOCS, ΔYBOCS-comp | FLX. w3: M = 247.3±74.7 F = 430.0±230.2 d= 1.07 p = 0.03; w7: M = 250.5±74.7 F = 429.2±223.3 d= 1.07 p = 0.03;  ΔYBOCS-comp: M = 9.9 p = 0.011 F = 4.5 p > 0.10 | FLX  **M < F**  Δcomp  **M > F** |
| Belotta-Silva  2012 | 158  (87) | 34±11  (18 – 65) | BRA | open label, controlled | FLX: 12 w, 20mg/day→ 80mg/day at w4 (or w8). GCBT: groups 6-8, 2h, weekly, 12w + 3 extra | Improve: % ΔYBOCS | **M:** β=-0.093 (0.045), p=0.042;  **M x T**: was not tested | **M < F** |
| Hasanpour  2019 | 151  (64) | 34 ± 10  (18 – 65) | IRN | open label | 12-weeks, 150–300 mg | Apriory algorithm for association rules: Responder = Female + Married + Late onset + YBOCS comp <10: 0.85 chance to respond;  Refractory = Female + Early onset: ≥0.71 chance to not respond | | **F+LO > F+EO** |
| Astaneh  2022 | 69  (36) | 32 ±NA  (20-50) | IRN | naturalistic  open-label | FLX: 6 mo, 50-100 | Responder: post YBOCS < 17 (≥50% ΔYBOCS) | Responders: F:M OR = 1.98;  χ²(1) = 1.21, p = 0.27 | FLX  **M ≤ F** |
| Tyler  2024 | 101  (55) | NA  (18-75) | USA | RCT | 12 weeks SRI → 25 ERP; continue vs. discontinue SRI → 24 w f/up | Responder: YBOCS ≤14  Worsening: CGI-I “much worse” or “very much worse” for 2 cons weeks | Worsening: F = 51%, M = 17%; F:M OR = 2.42 [95% CI, 1.13–6.17], p = 0.02 | Worse after SRI disc:  **M < F** |
| *Studies of other meds* | | | | | | | |  |
| Tural  2019 | 36  (22) | 37 ± 11  (NA) | TUR | double blind, placebo controlled | FLX: 40 mg/day;  + Folic acid: 5mg/day or placebo. | Improve: ΔYBOCS over 6 assessments (12w) | **T:** F(15,480) = 0.88, p=0.58, ƞ2=0.03 F**:** F(15,480) = 0.30, p=0.996, ƞ2=0.01; **T x F:** F(15,480) = 0.61 p =0.86 ƞ2=0.02 | **M ~ F** |
| *Neuromodulation* | | | | | | | |  |
| Bation  2019 | 21  (12) | 45 ± 20  (16-70) | FRA | random, sham controlled | Transcranial direct current stimulation: 2 20 min sessions/day, 5 consecutive days | Improve: ΔYBOCS | *"sex was not associated with improvement in YBOCS in the active group", stats NA* | **M ~ F** |
| Metin  2019 | 50  (24) | 33 ± 12  (18 +) | TUR | open label | rTMS: 1 Hz to the rDLPFC; 1000 pulses with 110% of the motor thresh; mean 22 s (12-40) | Responder: clinical evaluation interview conducted by 2 psychiatrists | **M** OR=0.92 χ2 = 0.015 p = 0.90 | **M ~ F** |
| Storch  2021 | 100  (45) | 39 ± NA  (22 – 68) | USA | random, sham controlled | 3–5 min Sympt.-Prov. →TMS /sham; w1-5: 5 a week; w6: 4 a week. TMS: 20 Hz dTMS at 100% of RMT, 2s pulse trains/20s inter-train intervals (50 trains and 2000 pulses/session) | Improve: ΔYBOCS over 6 assessments | **F:** d 6w = -0.15, 10w =-0.18, p>0.05 | **M ~ F** |
| Mar-Barrutia  2024 | 25  (12) | 48 ± 11  (NA) | ESP | open label | DBS with target vALIC; severe refractory OCD | Responder: ≥35% ΔY-BOCS | **M:** 76.9%, **F:** 33.3%;  OR = 10.05, p = 0.038 | **M > F** |
| Asik  2024 | 32  (14) | 35 ± 13  (18 +) | TUR | retrospective naturalistic case-control | Deep TMS (H7): 30 sessions over 6 weeks (5 days/week); 20 Hz, 2-s trains, 20-s intertrain intervals, 50trains/2,000 pulses/session | Responder = ≥50% reduction in Y-BOCS from baseline to endpoint | Responders: F = 8/14 (57.1%); M = 17/18 (94.4%). Non-responders: F = 6/14 (42.9%); M = 1/18 (5.6%) | **M > F** |
| Mudunuru 2025 | 378  (178) | 35 ± 12 (12–89) | INDIA | Retro observational | observational study; consecutive clinical sample. Accelerated deep repetitive TMS (aDTMS), H7 coil: 15 sessions over 8 days (~2/day); each session iTBS600 followed by 20-Hz HFS; 39,000 total pulses | OCD symptom response to aDTMS, measured by Y-BOCS Day 1 → Day 8 | Y-BOCS % reduction: M 46.11 ± 22.44% vs F 42.40 ± 25.31%. Response: M 67.14% vs F 59.32%. Remission: M 52.14% vs F 38.90%. Severe: M 45.13% Y-BOCS reduction vs F 39.64%. Extreme: M 50.85% vs F 35.72%. | **M > F** |
| Elvan  2026 | 60  (35) | Med 30, IQR 19, (13–68) | TUR | Retro, tx | 29 sessions of high-frequency dTMS using H7 coil; individualized symptom provocation before sessions; patients continued pharmacoth | YBOCS | dTMS response ≥35% reduction in Y-BOCS. TRS: 9 F, 11 M; TRP: 26 F, 14 M. Female response = 26/35 (74.3%); male response = 14/25 (56.0%) | **M < F** |
| Swisher  2026 | 712  (425) | 27 ± 9  (18–72) | USA | retrospective, naturalistic IOP | Intensive outpatient ERP, 3 h/day, 5 d/week; mean 11.11 ± 4.57 weeks; 82.7% in person | Responder: ≥35% ↓ YBOCS-SR | Responder status × gender: χ²(1)=0.17, p=.683. | **M ~ F** |
| *In children* |  |  |  |  |  |  |  |  |
| Ingram 2025 | 98  (47) | 13 ± 3  (7–17) | USA | observ | Differential fear-conditioning/extinction task with skin conductance response | CY-BOCS, OCI-CV CGI-S | For dropout, sex was tested but was not significant; only age significantly predicted dropout (β = −2.13, p < .001). | **M ~ F** |
| *Other treatments* | | | | | | | |  |
| Shams  2019 | 40  (24) | NA  (18 – 55) | IRN | double-blind, placebo | Caffeine: “low dose” → 300 mg/day | Improve: ΔYBOCS | β_M_ = 2.89, t = 2.24, p = 0.03 | **M > F** |
| Rück  2012 | 24  (10) | 41 ± 11  (25-68) | GBR | open label | Capsulotomy: 12- 20 mm lesion in the middle portion of the internal capsules at the level of the posterior part of the medial putamen, via radiofrequency heating or gamma radiation. | Improve: ΔYBOCS | *"female sex predicting worse outcome on the YBOCS at both follow-up points at a trend level"* stats NA | **M ≥ F** |
| Del Casale 2025 | 30  (11) | 36 ± 15  (18+) | ITA | Retro, observ | <25% Y-BOCS reduction after ≥12w of an SSRI or clomipramine at the highest tolerated dose & ≥30 hours of CBT | TR-OCD vs other TRMDs. | TR-OCD: 19 M/11 F; other TRMDs: 80 M/100 F; χ²(1)=3.682, p=.055 | M ~ F |
| *Treatment preferences* | | | | | | | |  |
| *children/adolescents* | | | | | | | |  |
| Negreiros  2019 | 94  (57) | 14 ± 3  (8-18) | CAN | open label | GF-CBT: 12 weekly sessions with family → monthly boosters drop-ins | Attendees of booster sessions | Attendees: F:M OR = 0.92 [95%CI, 0.35-2.43] | **M ~ F** |
| Mennies  2020 | 11875  (5687) | 10 ± 1  (9 – 10) | USA | retro history of treatment | NA | OCD dx: F OR =0.72 [95%CI=0.62-0.84]; Seeking treatment once diagnosed: F OR=0.5 [95%CI=0.44-0.56] | | **M > F** |
| Alt  2025 | 20  (11) | 16 ± 2  (12–18) | GER | Quant/qual; focus groups and surveys | 14 sessions (vCBT) with exposure, augmented by ECG chest strap measuring heart rate/HRV, eye-tracking glasses, and movement sensors for compulsive behavior. | Technology affinity: t(16.5) = −0.09, p = .93, NS. Technology acceptance: t(9.5) = 3.09, p = .01, d = 1.51, significant. | | M > F tech acceptance  M ~ F tech affinity  M ~ F tx response, OCD severity |
| Baweja  2025 | 37355 (18423) | 11 ± 4  (6–18) | USA | Observ of tx; retro | Psychotherapy, antidepressants, antipsychotics | Gender differences in psychotherapy, antidepressant, and antipsychotic utilization after OCD Dx | | M < F psychotherapy, antidepressant use  M > F antipsychotic use |
| *adults* |  |  |  |  |  |  | |  |
| Stengler  2012 | 84  (50) | 39±13  (18 – 77) | DEU | retro reports | NA | T onset to help seeking: M =10±10.5 yr F=6.4±8 yr (M-F=3.6±6.4 yr; d = 0.4; p = 0.079); time onset to diagnosis M= 13.2 yr and F= 8.6yr. | | **M > F** |
| Jakubovski  2013 | 196  (109) | 34 ± 11  (18 +) | BRA | random open label trial | FLX: 12 weeks, 20mg/day → 80mg/day, weekly by 20 mg/day; psychiatric consultations every 4w. GCBT: groups of 6–8, 2-h, weekly, 12 weeks + 3 extra | Completers vs. dropouts | **M:** OR=1.12, χ2=0.15 p = 0.70 | **M ~ F** |
| Patel  2017 | 216  (157) | 34 ± 13  (18+) | USA | online survey | First line: SSRI vs ERP; augmentation: antipsychotic vs ERP. Details NA | Preferences for (1) first line, (2) augmented treatments | First line: F prefer ERP OR = 1.5 p = 0.21; Augmented: F prefer ERP OR = 2.87 p = 0.02 | **F: ERP>med** |
| Li  2022 | 1343  (824) | 34 ± 12  (18+) | AUS | observ | iCBT; 2019–2020 | Use of iCBT (pre→COVID) | Covid F:M OR = 1.57 [95% CI; 1.19–2.06] | COVID↑↑iCBT:  **M < F** |
| Sun  2023 | 1767→  2648 | 23 ± 7→  41 ± 15 | CHN | medical records | Psychiatric emergency room visits 2018-2020 | Pre → pandemic. | F:M OR: 2018 = 0.50, 2020 = 0.68, p > 0.6 | **M ~ F** |
| Tabeleão  2024 | 90  (64) | NA  (18 - 60) | BRA | open label | CBT: 7-10 sessions | Completers vs. dropouts | **F:** OR = 1.2, **M:** OR = 3.3, **F:M** OR = 2.8, χ² p = 0.05; Cramer’s V = 0.207 | **M > F** |
| Bey  2025 | 276  (199) | 33 ± 9  (18-65) | GER | online survey | NA | Delay to seeking treatment: M = 5 ± 7 yr F = 4 ±6 yr; d = 0.21 p = 0.04.  Delay to recognition: M = 5.6 ± 7.2 yr; F = 3.9 ± 5.6 yr, d = 0.27 p = 0.03. | | **M > F** |
| George  2025 | 978 (654) | 18-44 | USA,  Afr Amer | medical records | NA | Spoke to professionals: about obsess: F:M OR = 0.93 [0.47–1.89]; about compuls: F:M OR = 0.71 [0.38–1.30] | | **M ~ F** |
| Girone  2025 | 316 (155) | 38 ± NA  (18-65) | ITA | retrospective interviews | consecutive OCD outpatients at two tertiary OCD clinics | Duration of untreated illness (DUI): M 49.1%, F 50.9%; lDUI: M 52.7%, F 47.3%; gender n.s. in multivariable model (exact statistic not reported) | | **M ~ F** |
| Kaewpila  2025 | 81  (63) | 26 ± 6 | THAI | Retro reports | mixed | Treatment seekers **F: 17/63 (27.0%); M: 10/18 (55.6%)** sought treatment | | **M > F** |
| Stein  2025 [A3] | 26136 | NA  (18+) | GEN  (woldwide) | multistage clustered-area probability household sampling | clinical interviews | Treatment use within 12m: FM OR = 1.30 [95%CI, 0.70–2.30], χ2 = 0.7 p > 0.05 | | **M ~ F** |
| Woodson  2026 | 191  (108) | 29 ± 12 (18–74) | USA | natural | residential vs. virtual | Men more in residential; women more in virtual tx χ²(1) = 3.75, p = .053. | | M > F residential  M < F virtual |
| *All ages* |  |  |  |  |  |  |  |  |
| Mathai  2026 | 2639 (1826) | 31 ± 12  (18+) | 11 countr | Cross-sectional, secondary; self-report | Naturalistic psychedelic/related substance use for OCD: psilocybin, LSD, DMT, mescaline, MDMA, ketamine, THC | Psych use vs. Non-use | Males use psychedelics more (OR = 1.41, 95% CI [1.02, 1.90], p = .035). | **M > F** |
| Pelligrini  2025 | 19  (6) | 38 ± NA  (20-60) | GBR | fixed order, within-subjects | 2 doses of oral psilocybin, 1 mg followed by 10 mg, separated by 4 weeks | % change in Y-BOCS after 10-mg psilocybin | Gender × Time NS at 1 week. At 4 weeks post-10 mg, significant M/F difference: β = −6.22, p=.009, males showing greater sustained response. | **M > F long-term improvement** |
| *Other topics:* | | | | | | | |  |
| Chu  2016 | 527  (287) | NA  (20 +) | TWN | observ | SSRI: paroxetine, fluoxetine, sertraline, citalopram, escitalopram, fluvoxamine | SRI decrease risk of stroke (HR = 0.30 [95% CI = 0.10–0.86] p = 0.02). **M** HR = 1.42 [95%CI = 0.55,3.68] | | **M ~ F** |
| Dominick  2021 | 975  (220) | NA  (0-25+) | USA,  fragile X | registry records | person meds | Med use: med prescribed for OCD | OCD meds: F=10.6% vs M=7.7%, p > 0.05 | **M ~ F** |
| Cohen  2025 | 673  (410) | 37 ± 13  (18+) | NLD | retro chart review | SSRIs, clomipramine, antipsychotic augm | Medication adequacy ~ F: β = 0.21, p = 1.0; ≥2 SSRIs ~ F: β = 0.27, p = 1.0; Max dose ~ F: β = 0.18, p = 1.0 | | **M ~ F** |
| Alqahtani 2026 | 64  (23) | <30 = 20%;  30–40 = 48%;  40–50 = 16%;  >50 = 16% | SAU | Cross-sect; online survey | Senior psychiatry residents and board-certified psychiatrists on referrals for neurosurgery | Referral: B = 1.864, OR = 6.450, 95% CI [1.078, 38.573], p = .041. Positive attitude: B = −0.372, OR = 0.690, 95% CI [0.191, 2.489], p = .570 | | M > F referring for surgery |
| Berg  2025 | 163  (117) | 35 ± 11  (18+) | NOR | Sec analysis of RCT | concentrated exposure-based CBT/ERP delivered over 4 consecutive days | Sex did not significantly predict OCD severity/outcome: posttreatment β = .00, t = −0.02, p = .984; 3-month β = .03, t = 0.47, p = .642; 12-month β = .03, t = 0.41, p = .679. For 12-month WSAS: β = .02, t = 0.22, p = .827. | | M ~ F |
| Bakhshineejad 2026 | 51  (34) | 37 ± 8  (18–70) | IRAN | Random, triple-blind, placebo-control | Both groups received fluvoxamine 100 mg twice daily. Experimental group received DPHP 250 mg twice daily 8 wks | Gender significantly affected QoL (p=.040). \| OCD: F = M (NS). No significant sex difference in OCD severity. QoL: M > F, significant | | M ~ F OCD severity  M > F QOL |
| *Females only* | | | | | | | |  |
| Timpano  2011 | 71F | 27±4  (NA) | USA | random, controlled, open label | CBE with OCD prevention & CBE: 6 weekly group sessions prior to childbirth. | Improve: post-T YBOCS across 3 assessments | time x T: F(1, 34) = 4.83, p < 0.01 | PP + E > E |
| Sabetnejad  2016 | 58F | 33 ± 6  (21 – 46) | IRN | retro observ | FLX: mean dose of 60 mg/day for at least 3 months. GCBT group: 10s 45-min weekly. | Improve: ΔYBOCS, ΔFSFI | Time x T  YBOCS: p = 0.15  FSFI: p = 0.003 | side effects  FLX > CBT |
| Sayyah  2016 | 30F | 30 ± NA  (20 – 45) | IRN | open label | GCBT: 11sessions; SSRI: homogenized | Improve: post-T YBOCS | F(1,27) = 47.63 p = 0.001 | GCBT >0 |
| Challacombe  2017 | 65F | 33 ± NA  NA | GBR | random, HC controlled | PP < 6 mo → PP 9 mo  iCBT: 4 x 3 h over 2 w  TAU: naturalistic CBT | Improve: ΔYBOCS, ΔOCI-R | Time x T x Dx:  YBOCS: d = 1.32  OCI-R: d = 1.90 | iCBT > TAU |
| Conrad  2020 | 5F | NA  (11-13) | USA | open label | GCBT: 12 weeks | *“During COVID-19 pandemic this group process repeatedly recovered and strengthened”* | | GCBT > 0 |
| Trinh  2023 | 1317F | 28 ± 5  (18+) | DEN | registry, medical records | SSRI/SNRI/other antidepress | Unexposed =34.1%  Discontinued before pregn= 26.3%  Continued during pregn= 39.6% | Postpartum OCD Visit: Continuers vs discontinuers  HR=1.3 [95%CI,0.7–2.6]  Continuers vs unexposed  HR = 1.7 [95%CI,0.8–3.6]  Continuers ≥2 fills  HR = 1.3 [95%CI,0.6–2.5]  1 fill pregn vs unexposed: HR = 3.4 [95%CI, 1.2–9.5]  vs discontinuers: HR = 2.49 [95%CI, 0.91–6.83] | unstable treatment in pregn ↑↑ PP OCD |
| Zhang  2023 | 78F | 19±1  (18–20 | CHI | RCT | Baduanjin Qigong exercise: 12 w; 1hx 3×/w; vs usual lifestyle | Improve: SCL-90 -OS | Post: Baduanjin = 0.78 ± 0.33 Control = 1.05 ± 0.57  d = 0.57 p < 0.05 | exercise↑↓OCs |
| Levy  2024 | 35F | 29 ± 7  (19-48) | USA | open label | Single-session ERP | Improve: DIAMOND  High estradiol: > 1.60 pg/mL | Time x high estradiol:  $\eta_{p}^{2}$ = 0.34, p = 0.01  Pre–post x high estradiol:  $\eta_{p}^{2}$ = 0.40, p = 0.02  Post–recall x high estradiol: $\eta_{p}^{2}$ =0.40, p = 0.02 | **ERP x estradiol:**  immediate: high < low  recall:  high > low  durability:  high > low |
| Monaco  2025 | 45F | 19 ± 7 (16+) | ITA | Naturalistic observ, longitudinal | Lurasidone + fluvoxamine + multidisciplinary rehabilitation (n=14) vs heterogeneous SSRI/antipsychotic regimens + same rehabilitation (n=31); ~6 mo | Improve YBOCS-II | RAS: time BF10 = 7.15×10⁹ (MissForest), 1.68×10¹² (MissRanger).  BUT-GSI: time BF10 = 5.20×10³, 3.68×10³.  Group × time: BF < 1 across primary outcomes.  Y-BOCS-II: inconclusive/exploratory. | Tx ~ control |
| Steinman  2025 | 15F | 28 ± 5  (18+) | USA | open label | Ante: 3rd tri, high anx: online CBT, with partner | Improve: OBQ, STAI | Pre-post: OBQ d=–0.92 p=0.003; trait anxiety d= –1.29 p<.001, interpretation bias d=–1.02 p=0.001 | online Ante CBT↓ risk OCD |

**Abbreviations used in Table A.7**

Instruments: **CBCL** - Child Behavior Checklist^14^;**CGI-I** - Clinical Global Impression – Severity and Clinical Global Impression –Improvement^272^; **COIS-C** - The Child Obsessive-Compulsive Impact Scale-Child reports^273^; **COIS-P** - The Child Obsessive-Compulsive Impact Scale-Parent reports^273^; **CY-BOCS** - the Children’s Yale-Brown Obsessive-Compulsive Scale^17^; **CY-BOCS-SR** - the Children’s Yale-Brown Obsessive-Compulsive Scale,^17^ self-report version; **DYBOCS** - The Dimensional Yale–Brown Obsessive–Compulsive Scale^29^; **LIFE OCD** - Longitudinal Interval Follow-Up Evaluation^274^ of OCD symptoms; **PedsQL** - The Pediatric quality of life Scale^275^; **QoL-kid** - The Pediatric quality of life Scale Child Report^275^; **QoL-parent** - The Pediatric quality of life Scale Parent Report^275^; **YBOCS** – the Yale-Brown Obsessive Compulsive Scale^82, 83^; **YBOCS-comp** – compulsion subscale of the Yale-Brown Obsessive Compulsive Scale^82, 83^.

Other abbreviations: **95%CI** – 95% confidence interval; **AAP** -Atypical antipsychotics; **Antidepr** – antidepressants; **Antypsych** – antipsychotics; **Anx** – anxiety; **A-OCD** – autogenous obsessions; **Assess** – assessments; **CBE** – childbirth education classes; **CBT** – cognitive behavioral therapy; **d** – effect size Cohen’s d; **Depress** – depression; **Dissoc** – dissociation; **dTMS** - deep transcranial magnetic stimulation; **Dur** – duration; **ERP** – exposure and response prevention therapy; **FLX** – Fluoxetine; **GCBT** – group based cognitive behavioral therapy; **h** – hours; **HR** – hazard rate; **HW** – homework; **Hz** – hertz; iCBT – time intensive cognitive behavioral therapy; **Meds** – medications; **Min** – minute; **Mo** – month; **NA** – not available, not reported; **non-SRI** – alternatives to serotonin reuptake inhibitors; **Observ** – observational; **OR** – odds ratio; **Person ERP** – personalized exposure and response prevention; **Person meds** – personalized medications; **Person therapy and meds** - personalized therapy and medications; **Person treatment** – personalized treatment; **post-T** – post treatment; **Random** – randomized; **rDLPFC** - right dorsolateral prefrontal cortex; **Relig** – religious obsessions; **Retro** – retrospective; **Retro observ** – retrospective observational; **RMT** - resting motor threshold; **R-OCD** – reactive obsessions; **rTMS** - repetitive transcranial magnetic stimulation; **SMT** – stress management; **SNRI** - Serotonin and norepinephrine reuptake inhibitors; **SR** – self-reports; **SSRI** - selective serotonin reuptake inhibitor; **Sym.-Prov.** – symptoms provocation; **T** – treatment; TAU – treatment as usual; **TMS** - transcranial magnetic stimulation; **w** – week; **yr** – year; **M** – male; **F** – female.

**Appendix 8.** Papers that employ animal models approaches.

Table A8. Sex differences revealed by animal model studies.

| **Author**  **Year** | **N** | **Age** | **Spec** | **Methods** | **Gender effects stats (F/M)** | |
| --- | --- | --- | --- | --- | --- | --- |
|  |  |  |  |  | **Stats** | **Effect** |
| Samocha  2010 | F2 M=237, F=235, F34 M=56, F=79 | F2=89±0.25d, F34= 92±0.29d | MS | PPI, QTL Analysis | **Startle:** M SM/J>LG/J, F SM/J=LG/J, p<0.05  **Genome-wide LOD Scores for Startle:** Genome-wide significance threshold for marker rs31102161 (Chromosome 7) and rs29530504 (Chromosome 17) when sex is interactive covariate, p<0.05 | M and F mice show unique QTL for startle. |
| Haas  2012 | N not reported | 3-4mo | MS | Quinpirole  OF | **Rate of Repetition:** *A/J Females* Saline<QNP p<0.05, *C57BL/6J Females* p<0.05, **Behavioral Repertoire (# Distinct Behavioral Patterns):** *A/J Females* Saline<QNP p<0.05, *C57BL/6J Females* Saline>QNP p<0.0125, | **Quinpirole=Repetition ↑** |
| Hoffman  2012 | 60 PREG F | 6-18mo | RAB | Straw Carrying, SCH23390 (D1 Ant), Raclopride (D2 Ant), OF | **Straw Carrying (total straw carried): (Mann-Whitney)** SCH23390 Dose vs Baseline (0µg/kg) 5 µg/kg p=0.06, 12.5 µg/kg p<0.005, 50 µg/kg p<0.005, 100 µg/kg p<0.05; Raclopride Dose vs Baseline 50 µg/kg, 200 µg/kg, 500 µg/kg p>0.05, 1000 µg/kg p<0.005  **# of Cycles:** vs Saline, both SCH23390 and Raclopride p<0.005  **Latency to begin carrying, Max Cycle Frequency, Max Bout Length, Total # Bouts:** p>0.05 | **D1/D2 Inhibition ↑ = Repetitive Straw Carrying ↓** |
| Olvera- Hernández  2013 |  | 11-14mo | RAT | 8-OH-DPAT, fluoxetine  T-Maze | **T-Maze Repetitive Choices:** 2-Way ANOVA Treatment p<0.001, Sex/Endocrine condition p>0.05, no interaction,  8-OH-DPAT F in Persistent Diestrus > Veh, p<0.001, vs 8-OH-DPAT Irregular Cycle F p<0.01, vs 8-OH-DPAT M p<0.01  Fluoxetine treat in 8-OH-DPAT Per. Diest. F < 8-OH-DPAT, p<0.01  1-Way ANOVA 8-OH-DPAT Males > Veh, p<0.05, 8-OH-DPAT Irregular Cycle Females=Veh, p>0.05, no effect of fluoxetine  **Spontaneous Activity Counts:** 8-OH-DPAT>Veh p<0.001 for Per.Diest. F and M, p<0.05 for irreg. F, no effect of fluoxetine | **T-Maze Repetition:**  8-OH-DPAT = ↑ repetitive choices in Persistent Diestrus F only  **Spontaneous Activity:**  M=F |
| Arvidsson  2014 | Young M=7, F=8, Adult M=6, F=8 | 3-4 wk, 16-20 wk | MS | Chronoamperometry | **DA Amplitude:** (3-way repeated measure ANOVA) AF vs AM p<0.01 (all stim), p<0.001 (Stim1), p>0.05 (Stim 6), p<0.05 (Stim7,15min rest), YF vs YM p<0.01 (all stim), p<0.001 (Stim1), p<0.05 (Stim6), p<0.001 (Stim7); AM vs YM p<0.01, AF vs YF p<0.01, AM vs YF p=0.3244, AF vs YM p<0.0001 | **DA Amplitude:** AF>AM, YF>YM  AF>YM, AM=YF |
| Kalinichev  2014 | 11-17 sex/geno/treat | Adult | MS, RAT | ADX88178 (mGlu_4_ receptor agonist)  EPM | **EPM:** *Open Arm Entries* Mouse: M ADX >Veh=ADX+KO=Veh+KO, p<0.05, F ADX >Veh=ADX+KO=Veh+KO, p<0.01, Effect of treatment p<0.001  *Time Spent Open Arms* Effect Sex p<0.001, Effect Treatment/Genotype NS  **MBur** | **MGlu4 Activity ↑**  **= EPM Open Arm Entries ↑, M=F**  **Time Open Arms: M=F** |
| Brydges  2015 | F=19, M=33 | 60 PND | RAT | Prepubertal Stress (PPS), DDT | **# sessions to learn nosepoke training** (Box-Cox Transformed) Group x Sex p=0.04, PPSF vs CTF p=0.035, PPSM vs CTM p>0.05, PPSF vs M p=0.002  **AVG Trials Initiated:** Delay p<0.001, Group x Sex p=0.85  **AVG Trials Responded To:** Group x Sex x Delay Block p=0.006  **Task Response Latency:** Group x Sex p<0.0001, Group x Delay Block x Sex p=0.045  **Nosepoking during choice phase:** Group x Delay Block x Sex p=0.04 | **Nosepoke Training:**  CTM=CTF=PPSM<PPSF  **Trial Initiate: M=F**  **Trial Response:** PPSF<CTM,CTF,PPSM  **Laten** PPSM,CTM,CTF  **Time Nosepoke:**  PPSF>PPSM,CTM,CTF |
| Freund  2015 | M (by age) CMI=7,6,12, CT=7,10,9  F (by age) CMI=7,8,13, T=7,7,10 | 28PND, 60PND, 90PND | RAT | Neonatal CMI  SpA, OF, MBur | **Treatment with Clomipramine at PND 9-15**  **SpA** (3-way ANOVA): Drug p<0.006, Drug x Sex NS  **MBur:** Sex x Age p<0.0001, CMI vs CT M p<0.05, F p>0.05  **EPM: *Time Open Arm*** Sex x Age x Drug p<0.05, Male x Drug p<0.05 | **MBur:** CMI M at 100d = MBur ↑, F CMI=CT  **EPM:** CMIM < CTM (all age)  CMIF = CTF (25d, 60d), CMIF<CTF (100d) |
| Dries  2016 | N=4/sex/geno | 18-24wk | MS | *Nicastrin* cKO in *olig-cre*, OF, EPM, PPI, Grooming | **OF, EPM, PPI:** (Student’s t-test) M vs F p>0.05 | M vs F NS |
| Mitra  2016 | Female Only | 80-90d | MS | Non-Induced Model of Compulsive Nest Building (BIG), OVX  Nest Building, MBur, OF, EPM, NOR | **Nesting Score:** OVX in BIG1/BIG2 Compulsive Mice > SHAM, p<0.0001, BIG1>BIG2, p<0.001, NS for CT1, CT2, SML1, SML2  **MBur:** OVX BIG1**/*BIG2>SHAM BIG1/BIG2, **p<0.01, *p<0.05, SHAM=OVX CT1, CT2, SML1, SML2  **OF:** *Time Center* OVX<SHAM for BIG1***, BIG2***, CT1***, CT2***, SML1*, ***p<0.0001, *p<0.05  **EPM:** *Time Open Arms* OVX BIG1/BIG2***<SHAM, ***p<0.0001, OVX=SHAM for CT1, CT2, SML1, SML2  **NOR:** OVX = SHAM, P>0.05  **E2 Levels in BIG Mice:** OVX<SHAM, BIG1 p<0.001, BIG2 p<0.0001, **Progesterone:** OVX<SHAM, BIG2 p<0.0001  **E2 Administration in BIG OVX Mice:** *Nesting Scores* E2<Veh in BIG1 p<0.001, BIG2 p<0.0001, *MBur* E2<Ven in BIG1/BIG2, p<0.0001, *OF Time Center* E2>Veh in BIG1, p<0.0001 | **OVX in Compulsive BIG Mice:**  =Nesting ↑  =MBur ↑  =OF Center ↓  =EPM Open Arm ↓  **Estradiol (E2) Administration:**  Normalizes behavioral values in BIG mice |
| Cano-Ramírez  2017 | PREG=12, Estrous (EST)=12 | 6-24mo | RAB | Straw Carrying, C-Fos | **Latency to Straw:** Preg vs EST p<0.01, **# Times Drink/Eat:** Preg vs EST p<0.05, **Entrance to Nest Box:** p=0.06,  **cFos Expression** *Orbitofrontal* Preg+Straw vs EST+Straw&Preg p<0.05, *Premotor* Preg+Straw vs Preg p<0.05, *Ant Cingulate* Preg+Straw vs Est+Straw&Preg p<0.05, *Prelimbic* Preg+Straw vs Preg p<0.05, *Piriform Cortex* Preg+straw vs Est+Straw&Preg p<0.05, EST+Straw vs EST p<0.05, Preg vs EST p<0.05, *Infralimbic* Preg+Straw vs Preg p<0.05 | **Preg + Repetitive Straw Carrying =**  **cFos ↑**  (for Orbitofrontal, Ant. Cingulate, Premotor, Prelimbic, Piriform, Infralimbic) |
| Mitra  2017 | Female Only | >60d | MS | Non-Induced Model of Compulsive Nest Building (BIG), Fluoxetine Treatment  MBur, Nest Building, OF, TST | **MBur:** Lactating (LAC) Veh < Non LAC = CT for all three weeks p<0.05 Wk1, Wk2, p<0.01 Wk3; Fluoxetine < Veh for LAC, p<0.001 Wk1-3; Fluoxetine < Veh for nonLAC, p<0.05 Wk2, p<0.001 Wk3, Fluoxetine < Veh for Nulliparous Wk3 p<0.001  **Nesting Score:** LAC Veh<NonLAC=Null Wk1-3 p<0.001, Fluoxetine LAC<Veh LAC Wk1-3 p<0.001, Fluoxetine NonLAC<Veh NonLAC Wk3 p<0.01, Fluoxetine Null<VehNull Wk3, p<0.001  **OXTR Antagonist:** *MBur* In Lac MBur > Veh p<0.001, *Nesting Scores* > Veh for 0-1 and 1-2 Hr post administration p<0.001  **Serum 5HT Levels:** LAC > NonLAC=Null, p<0.05  **OF:** LAC Veh<Non Lac=Null, p<0.05, Fluoxetine LAC > Veh LAC, p<0.01, Fluoxetine Null > Veh Null, p<0.01, *Dist Travelled* NS All | **Lactation in BIG Compulsive Mice:**  =MBur ↓  =Nesting ↓  **Fluoxetine Administration**  =MBur ↓↓  =Nesting ↓↓  **5HT Levels ↑ in LAC** |
| Mitra  2017 | Proestrus Females Onlhy | >60d | MS | Non-Induced Model of Compulsive Nest Building (BIG); Nest Building, MBur, OF, EPM, FST, ELISA | **Nest Building:** Effect size Strain p<0.001, Replicate p<0.02, Sex p<0.0001 (BIG M > BIG F), Strain x Sex Interaction p<0.0001  **MBur:** Strain p=0.054, trend BIG>SM  **OF:** NS Strain p>0.40, Sex p>0.15, Strain x Sex p=0.08  **EPM:** Strain p<0.0001, Sex NS p>0.30, Strain x Sex p>0.40  **NOR:** Strain NS p>0.09, Strain x Sex p>0.05  **FST:** Strain p>0.25, Sex x Strain p>0.15, Sex p<0.05 CT + SML F > M | **Nest Building:**  BIG > CT=SML  BIG M > BIG F  **MBur:**  BIG > CT = SML, M=F |
| Taylor  2017 | 35/sex | 64d at pretest | MS | Diazepam (GABA R target), Escitalopram (serotonin transporter inhibitor), mBur | **MBur:** Diazepam < Escitalopram < Vehicle, p<0.05, M=F NS | M=F |
| Reimer  2018 | ***N’s differ by test*** | 8wks | RAT | mCPP  FCon, Cued FExt, Groom | **Grooming:** mCPP 3.0mg/kg > mCPP0.5mg/kg=Saline, P<0.05, M=F (n’s SalM=19, mCPP0.5M=12, mCPP3.0M=13, SalF=14, mCPP0.5F=12, mCPP3.0F=14)  **Cued-Fear:** *Extinction Retention*: Fear Extinction Index (FEI) mCPP=Sal, M=F p>0.05, Estrous Cycle NS  **Contextual-Fear:** Extinction Retention: 2-way ANOVA Sex p<0.05, Treatment p>0.05, sex x treatment p>0.05  FEI M mCPP=SAL, p>0.05, F mCPP 3.0mg/kg<mCPP 0.5mg/kg=Saline, 1-Way ANOVA Treatment p<0.05  *Estrous*: 2-Way ANOVA Phase p<0.05, treatment p<0.05, phase x treatment p<0.05, mCPP 3.0mg/kg in Met/Di FEI < Pro/Est = all phase SAL=all phase mCPP 0.5mg/kg (SalM=11, mCPP0.5M=10, mCPP3.0M=11, SalF=13, mCPP0.5F=12, mCPP3.0F=15) | **Grooming**: mCPP>Sal, M=F  **Cued Fear:** M=F  **Contextual Fear:**  FEI M>F, mCPP 3.0 F<Sal F=mCPP 0.5 F  **Contextual Fear Est.:**  Met/Di w mCPP 3.0 FEI < Pro/Est=all Sal=all mCPP 0.5 |
| Baig  2018 | ***N’s differ by test, see stats*** | 2-7 mo | MS | *Akap13* Deletion  MBur, Grooming Assay, EPM, OF | **MBur:** (ANOVA) Genotype x Sex p=0.01, *Akap13*^+/-^ F (9) vs WT F (8) p=0.0025, *Akap13*^+/-^ M (4) vs WT M (6) p=1.00  **Grooming: (Complete)**Genotype x Sex p=0.01, *Akap13*^+/-^ F (4) vs WT F (4) p=0.06, *Akap13*^+/-^ M (5) vs WT M (7) p=1.00, **(Incomplete)** Genotype x Sex p=0.01, *Akap13*^+/-^ F (4) vs WT F (4) p=0.06, *Akap13*^+/-^ M (5) vs WT M (7) p=1.00  **OF:** Genotype x Sex p=0.54, **EPM:** Genotype x Sex p=0.47 | **M-Bur:** *Akap13*^+/-^ F>WT F  *Akap13*^+/-^ M=WT M  **Grooming:**  *Akap13*^+/-^ F>WT F  *Akap13*^+/-^ M=WT M  **OF/EPM:** M=F, *Akap13*^+/-^ = WT |
| Coba  2018 | 16/sex/  genotype | 8-11wk | MS | *DlGap1* KO, OF, Dig test, plash, social approach, nest building, PPI, FS | **Dig Test:** *Latency to Dig:* Genotype x Sex p<0.05, WTM vs KO-M p<0.05, KO-F vs KO-M p<0.05; *# Dig Bouts:* WT vs KO p<0.05, *Time Digging, AVG Bout Duration* NS  **Nest Building:** NS, **PPI:** NS  **Splash Test:** *Grooming Bout Duration* Geno x Sex p<0.01, HT-M vs HT-F p<0.05, HT-M vs KO-M p<0.05, *Latency to groom* KO>HT&WT, p<0.05, *Time Groom* HT-M vs HT-F p<0.05  **Social Approach:** *# Entries* Sex x Geno x Condition (Mouse or Object) p<0.01, in M KO or HT vs WT p<0.05, F p>0.05  *Time near Mouse:* Sex x Geno x Cond. P<0.05, M HT&KO<WT | **Dig Test Latency ↑** in KO-M only  **Splash Test (bout duration):** HT-M>HT-F, KO-F, KO-M  **Social Approach:** # Entries M KO&HT<WT,  F KO=HT=WT  Time near MS  M WT>HT & KO  F WT=HT=KO |
| Babbs  2019 | >20 mice/Genotype/Treat | 50-100d | MS | *Cyfip1,2* gene deletion, MBur, OF, hole board, induced grooming, CPP | **MBur:** (Mann-Whitney) *Cyfip1^N/-^* mice > Ctrl, p=0.031, (2-Way ANOVA) Effect of Genotype p=0.009, *Cyfip1^J/-^* mice > Ctrl p=0.019, (2-Way ANOVA) Effect of genotype p=0.042, NS for sex  **BE in CPP (PF Intake):** *Cyfip1^J/-^* mice Paternal deletion F > Ctrl, Maternal depletion F, p<0.001; *Cyfip1^J/-^* mice Maternal deletion M < Ctrl, p<0.05 | **BE in CPP:**  Paternal depletion F *Cyfip1^J/-^* mice > CT  Maternal deletion M *Cyfip1^J/-^* mice < CT |
| Gannon  2019 | ***N’s differ by test, see stats*** | 90-100d Immuno | MS | Hindbrain GR Ablation, Barbering Analyses, Plasma steroid analyses | **Grooming (Barbering):** HB-GRKO>Het>WT, M=F  **Adrenal Weight:** (ANOVA n=8/group) MWT<Het p<0.001, WT<HB-GRKO p<0.0001; F NS  **Corticosterone Levels:** (ANOVA n=8/group) M WT<Het=HB-GRKO p<0.05, F WT=Het=HB-GRKO p>0.05  **Cleaved Caspase (Adrenal Cortex) (Apoptosis):** (ANOVA n=5/group) M WT=Het=HB-GRKO p>0.05), F WT<Het<HB-GRKO p<0.0001 | **Grooming Severity:** M=F  **Corticosterone:** MWT<Het=KO  FWT=Het=KO  **Apoptosis:** M WT=KO  F WT<Het<KO |
| Kim  2019 | M KCNQ^+/+^=12, KCNQ^+/-^=11, F KCNQ^+/+^=5, KCNQ^+/-^=6 | 4-5mo | MS | Kv7.2 Channel KD  OF, Home Cage Activity, Rotarod, EPM, Grooming, MBur, 3c Soc, Tube Dom, Urine Marking | **Home Cage Activity:** *Distance* Q2^+/-^ > WT, P<0.05, M=F  **OF:** *# Entries Center* Q2^+/-^ > WT, p<0.05, M=F, *Time Center* Q2^+/-^ > WT, p<0.05, M=F, *Velocity Center* WT F > WTM; Q2M^+/-^ > WTM, p<0.05, Q2F^+/-^ < WTF, p<0.05, **EPM** *#Entries/Time/Distance in Open Arms*, Q2^+/-^ > WT, p<0.05, M=F  **Repetetive Grooming:** *Duration* Q2^+/-^ > WT, p<0.01, Sex Effect M>F, p=0.001  **MBur:** Q2M^+/-^ > WTM, p<0.005, Q2F^+/-^ = WTF, p>0.05  **Sociability:** *Duration in Mouse Chamber* Q2M^+/-^ < WTM, p<0.05, Q2F^+/-^ = WTF, *Duration Sniff Mouse* Q2M^+/-^ < WTM, p<0.05, Q2F^+/-^ = WTF, *Latency to Sniff Mouse* Q2M^+/-^ > WTM, p<0.05, Q2F^+/-^ = WTF  **Tube Dom:** *Avg Score* Q2M^+/-^ > WTM, p<0.005, *% Wins* Q2M^+/-^ < WTM, p<0.05, **Urine Mark:** *# Urine Marks:* Q2^+/-^ > WT, sex x genotype p=0.032, *Area of Urination* Q2^+/-^ > WT, sex effect p=0.003 | **Kv7.2 channel KD**  =Activity ↑, **M=F**  =Grooming ↑, **M>F**  =MBur ↑, **M Only**  **=**Sociability ↓, **M>F**  =Tube Dom ↑, **M>F**  =Urine Mark ↑, **M>F** |
| Lei  2019 | KO M=20, WTM=21, KO F=4, WTF=2 | 3-10 mo | MS | Sapap3 KO  Orbitofrontal Cortex (OFC) Recording | **OFC LFP:** Power KO<WT, no effect of sex, data pooled | **M=F** |
| Rohbani  2019 | N=16/group/test (male) | 60-65d | RAT | Parental Morphine Exposure – Paternal (P.ME) and Maternal (M.ME)  MBur, mRNA analysis | **MBur:** P.ME**, M.ME*, P.ME+M.ME*** > CT (1-Way ANOVA)  **Grooming:** *# Bouts:* P.ME*, M.ME*, P.ME+M.ME* > CT, *Time*: P.ME*, M.ME*, P.ME+M.ME* > CT (1-Way ANOVA)  **Fold Change BDNF mRNA Expression:** P.ME*, M.ME*, P.ME+M.ME**<CT  **p-CREB** P.ME**, M.ME***, P.ME+M.ME*** > CT (1-Way ANOVA), **Total CREB** P.ME=M.ME=P.ME+M.ME=CT (1-Way ANOVA)  **D2 Dopamine Receptor:** P.ME***, M.ME***, P.ME+M.ME*** > CT (1-Way ANOVA); *p<0.05, **p<0.01, ***p<0.001 | **Both P.ME and M.ME**  MBur ↑  BDNF mRNA ↓  p-CREB ↑  Total CREB =  D2R levels in NAc ↑ |
| Scoenberg  2019 | F=26, M=21 | 75-90d | RAT | Operant training followed by reinforcer devaluation (RD) | **Acquisition:** (RM ANOVA) F Effect Session p<0.001, M Effect Session p<0.001  Devaluation group NS for differences prior to RD for both M + F  **Consumption Test (Dev vs NonDev):** (RM ANOVA) F within subject time p=0.002, between subject dev.group p<0.001, time x dev.group p<0.001, M within subject time<0.001, between subject dev.group p<0.001, time x dev.group p<0.001  **Habit Test (Responses/min):** (2x2 ANOVA – training group x devaluation group) F training p=0.334, dev group p=0.930, train x dev=0.954; M training p=0.654, dev group p=0.025, train x dev p=0.948  **Extinction test nosepokes/min:** Devalued F=8.19±2,28, NonDevalued F=6.64±4.38, p=0.286, Dev.M=3.71±1.23, NonDev.M=5.26±1.43, p=0.018 | **Behavioral Types:**  F=Habitual (nosepokes Dev=NonDev)  M=Goal-Directed (nosepokes Dev<NonDev) |
| Tränkner  2019 | ***N’s differ by test***  See Stats | >3mo | MS | *Hoxb8* microglia or gene ablation. OVX, grooming, EPM | ***Hoxb8* KO Grooming:** *Coat Loss* 2 Way ANOVA Effect Genotype p<0.0001 and Sex p=0.003, KO F (n=29) > WT F (n=11) p<0.0001, KO M (n=13) < WT M (n=16) p<0.0001, *Hours per Day* 2-Way ANOVA Effect Genotype p<0.0001, Sex p=0.001, KO F(n=8) >WT F(n=11) p<0.0001, KO M(n=10) >WT M(n=9) p=0.008  ***Hoxb8* KO EPM:** *Time Closed Arms* KO F(n=11) > WT F(n=11) =WT M(n=6) =KO M(n=7), 2-Way ANOVA Effects Genotype p=0.0005 and Sex p<0.0001, KO F vs WT F p<0.0001, KO M vs WT M p>0.05 **Cortisol Levels: :** KO F(n=9) > WT F(n=13) =WT M(n=7) =KO M(n=7), 2-Way ANOVA Effects Genotype p=0.025 and Sex p=0.001, KO F vs WT F p=0.007, KO M vs WT M p=0.21  **Trilostane or OVX in *Hoxb8* KO F:** *Coat loss* Tri(n=7) =OVX(n=11)=M<DMSO(n=10) or Sham(n=6) p<0.014, *Groom Time* Tri=OVX<Sham (p<0.005) trend for DMSO (p=0.09), *EPM Time Closed Arms* Tri=OVX<DMSO or Sham p<0.0025, *Cortisol Levels*: Tri=OVX<DMSO or Sham p<0.0025  **Hormones in *Hoxb8* KO M:** *Coat Loss* Progesterone + Est. (n=6) > DMSO(n=12) p=0.0002, *EPM Closed Arms:* Prog. + Est.>DMSO, p<0.05 | ***Hoxb8* KO Coat Loss:**  KO > WT, F > M  ***Hoxb8* KO Anxiety (EPM):** KO F > KO M=WT  ***Hoxb8* KO Cortisol:** KO F > KO M=WT  **OVX in *Hoxb8* KO F**  = Coat Loss ↓, Anxiety (EPM) ↓, Cortisol ↓  **Est. + Prog. in *Hoxb8* KO M**  = Coat Loss ↑, Anxiety (EPM) ↑, Cortisol ↑ |
| Nagarajan  2024 |  |  | MS | Optogenetic activation of *Hoxb8* microgila, grooming, freezing, EPM, OF, ephys | No sex bias seen in elevated grooming induced by *Hoxb8* microglia activation in the DMS or mPFC | Elevated grooming with *Hoxb8*  microglia stimulation: M = F |
| Lustberg 2020 |  | 3-8 mo | MS | *Dbh -/-* mice, EZM, Nestlet shredding, MB, cFos | **Pilot work indicated no differences between sexes** |  |
| Muzyko  2020 | Behavior 41  GABA: CT=59, EP=61, Succi 39, Salifen 32, Pheni 61, Panto 30 | 40d, 70d, 6mo, 12mo | RAT | Preeclampsia (EP)  GABA derivative administration  OF, EPM, MBur | **OF:** M=F, Fecal Bolus EP>CT p=0.05, Grooming M Only p<0.05, Rearings M=F EP<CT p<0.05  **EPM:** *Time Open Arm* 40d F EP< CT p<0.05 (2.3x less), 40d M<CT p<0.05, *# Entries* F EP<CT p<0.05, *Rearings* F EP<CT, p<0.05, *Edge Exploration* F EP<CT, p<0.05  **MBur:** M=F, 40d M&F EP>CT, p<0.05  **Treatment with GABA derivatives PND 40-70**  Treatment w Succi/Sali/Pheni reverses behavioral deficits in both groups | **OF: (Fecal Bolus)** M=F, EP>CT  **OF: (Rearing)** M=F, EP>CT  **Grooming:** M>F following EP  **EPM:** M<F for #Entries, Rearings, Edge Explore  **MBur:** M=F, EP>CT |
| Odland  2021 | Females Only  ***N=9-10/group*** | 17-26 wk | MS | 5-HT_2A_R antagonists (M100907, SB242084)  DOI, citalopram, psilocybin  MBur | **Psilocybin (agonist):** *MBur:* 2.0 and 1.0 mg/kg < 0.5mg/kg and Veh, p<0.01, *Distance Travelled*: NS across all doses  **DOI:** *MBur*: DOI<VEH, p<0.001, DOI<M100907+DOI=VEH, p<0.001, SB+DOI=DOI  **Citalopram:** *MBur:* CIT<VEH p<0.01, M+CIT=CIT, CIT<SB+CIT=VEH, p<0.05  **Psilocybin:** PSIL=M+PSIL=SB+PSIL<VEH, p<0.01 | **In Females:**  5-HT_2A_R agonist = MBur ↓  M100907 blocks DOI  SB242084 blocks CIT  None block PSIL |
| Milton  2022 | Females Only  N=44 | 6 wk | RAT | ABA paradigm, Running wheel, plasma analysis, EPM, MBT | **Compulsive Marble Burying (MBT):** For ABA resistant rats, trend increased marbles buried p=0.0551, not correlated with Running Wheel Activity (RWA) p>0.187, positive association w food intake (p=0.034, vs ABA susceptible rats p=0.139) | Compulsive MBT not associated with ABA susceptiblity |
| O’Reilly  2021 | Males Only | 8-14wks | MS | SERT Ala56^+/+^  5-HT_1B_ Agonist (RU24969)  OF, PPI, Startle, cFOS | **Receptor Binding:** SERT Ala56 > WT in lateral (p<0.01) and ventral (p<0.05) OFC, NS in medial OFC  **Hyperlocomotion:** Effect of time p<0.0001 and RU24969 admin p<0.0001, time x drug interaction p<0.0001, genotype p>0.93, time x genotype p=0.56, drug x genotype p=0.56  **PPI:** Effect Prepulse Intesnity p<0.0001, RU24969 p<0.001, Intensity x Drug p<0.0001, genotype p=0.26, genotype x intensity p=0.26, genotype x drug p=0.15, genotype x intensity x drug p=0.30 | No genotype effect of SERT Ala56 in males with 5-HT_1B_ activation |
| de Brouwer  2021 | N=100 | 12 wks | Deer MS | Deer Mouse Model of Stereotypical Behavior | **No Statistics Reported.**  However, in graphs, female and male points are indicated by different colors, and female and male behavior appear to overlap completely for spontaneous alternation behavior (both N and H mice, all drug treatments), T-CAT, Stereotypical Activity, Vertical and Horizontal Activity in the Dark Phase, | No differences in behavior in Deer Mouse Model between M and F |
| Subbanna  2022 | N=8ms/sex/group | 90d | MS | Fetal Alcohol Spectrum Disorder Model | **Y-Maze:** Control vs GDE: Arm entry: male: F_3,21_ = 27; female: F_3,21_ = 20; *p* < 0.05, Dwell time: male: F_3,21_ = 24, *p* < 0.05; female: F_3,21_ = 22; *p* < 0.05, Preference for Novel Arm: Male: F_3,21_ = 18, *p* < 0.05, Female: F_3,21_ = 16, *p* < 0.05, Male vs Female: p>0.05  **Social Recognition Memory:** Control vs GDE: Male: F_3,21_ = 16, *p* < 0.05, Female: F_3,21_ = 18, *p* < 0.05, Male vs Female: p>0.05  **Nestlet Shredding:** Control vs GDE: male, F_3,21_ = 13; female, F_3,21_ = 12; *p* < 0.05, Male vs Female: p > 0.05  **Marble** **Burying**: Control vs GDE: male, F_3,21_ = 8; female, F_3,21_ = 9; *p* < 0.05, Male vs Female: p > 0.05  **Elevated Plus**: Control vs GDE: Open Arm Entries: male, F_3,21_ = 12; female, F_3,21_ = 11; *p* < 0.05, Time Open Arms: male, F_3,21_ = 19; female, F_3,21_ = 16; *p* < 0.05, Male vs Female: p > 0.05 | Behavioral Deficits Caused by GDE (Y-Maze, Social Recognition, Nestlet Shredding, Marble Burying, Elevated Plus):  M = F |
| Grant  2022 | N=35 (18F) | 3.5 - 5.6 yr | MK | Attentional set-shifting task analysis with fMRI | **ASST Acquisition:** High vs Low Performance Groups: *F*_(1,30)_ = 178.2, *p* < 0.0001; sex, *F*_(1,30)_ = 0.45, *p* = 0.5; group × sex, *F*_(1,30)_ = 1.7, *p* = 0.2 | For ASST Training Acquisition: M = F |
| Casado-Sainz  2021 | N=14  7 mDS-DREADD, 4F/3M  7 mDS-mCherry, 4F/3M | Adult (≥250g) | RAT | DREADD activation of DA neurons in SN, PPI, OF | **PPI of Acoustic Startle Response:**  Startle Response (SR): Males: 58.1 ± 3.8%, Females: 48.6 ± 4.3%, *P* < 0.001, *F*(1,84) = 30.64; Short Term Habituation (STH): In combined female groups: 19.7 ± 6.2%, *P* = 0.008, *F*(1,86) = 7.3; SR following CNO treatment in mDS-DREADD females: Ave. Diff. = 3.3 ± 1.5%, *P* = 0.0501 (*F*(1,9) = 5.11)  PPI: In mDS-DREADD female rats: 24% increase, *P* = 0.022, *F*(1,9) = 7.6; 58% increase at 69dB PPI condition with CNO, *P* = 0.018, *n* = 10, Male *P* > 0.05  Baseline PPI following 69dB prepulse: F > M, *P* = 0.006, *F*(1,84) = 7.86 | SR: M > F  Mean STH Significant in all female groups combined  PPI Increases with bilateral activation of nigro-striatal dopamine projections innervating the rat mDS:  M < F |
| Lustberg  2023 | 38 *Dbh*+/- (14F), 21 *Dbh*-/- (12F) | 8–16 mon | MS | *Dbh*-/- mice (Dop ↑, NE ↓) | **Odorant-induced defensive digging/grooming:** (M vs F) p > 0.05  **Novel odorant evoked CORT response:** *Dbh*-/- and *Dbh*+/- F greater than males, F(1,12) = 36.98, *p* < 0.0001, | Behavioral changes in *Dbh*-/- mice: M = F  Cortisol response to novel odorant: M < F |
| Ayub  2022 | 370 (340F) Swiss mice | 8wk | MS | S-Ketamine  MBT, NBT, OFT | **OFT:** In females, total distance travelled S-Ket 30mg/kg = 10mg/kg = 3mg/kg = Saline, p>0.05  **MBT:** In females # of Marbles Buried S-Ket 30mg/kg < 10mg/kg, 3mg/kg, saline, p<0.05  **Behavior in pre-selected compulsive females (>5 marbles buried in MBT at baseline): OFT** Total distance travelled S-Ket 30mg/kg < Saline, p<0.05, **MBT** # of marbles buried, S-Ket 30mg/kg < Saline, p<0.05, **NST** Cotton shredded: 1 hr S-Ket 30mg/kg < Saline, p<0.05; 24 hrs S-Ket = Saline, p>0.05  **With chronic subthreshold dose of fluoxetine:** *In F*: Only Fluoxetine+S-ketamine group reduces MBT compared to veh+veh control (p<0.05) on all days, and veh+ket on D1, 7 and 21 (p<0.05)(2 way repeated measures anova p<0.05), In OFT no difference between treatment groups, all decrease by D21; *In M*: Veh+S-Ket, Flu+Veh, and Flu+S-Ket all decrease MBT on D7 and D21, Veh+S-Ket and Flu+S-Ket decrease MBT on D14 (p<0.05), In OFT, no effects seen  No effects of estrous cycle or OVX on the effects of S-Ket on MBT or OFT | **In Females:**  S-Ket F OFT =  S-Ket F 30mg/kg MBT ↓  In Compulsive Females:  S-Ket OFT Distance ↓  MBT Marbles Buried ↓    **Subthreshold Chronic Doses of S-Ket or Flu**  In F: MBT ↓ in S-Ket+Flu group only  In M: MBT ↓ in all treatment groups compared to control  **No effect of Estrous or OVX** |
| Meinke  2022 | All n sex/group  Mendelian:3-6, mRNA: 2-5  WB: 4  OFT 6-20  EZM: 8-23  TST: 4-9  FST: 4-13  SocDom: 10-15  MBT: 5-27 | 4-10 wks | MS | SERT Thr276Ala Knock-in Mice | **Analysis of Mendelian Genetics of SERT Ala276 mice:** χ^2^ test, male vs. female n.s, **mRNA and protein expression of SERT:** mRNA one-way ANOVA; midbrain males *P* = 0.21; midbrain females *P* = 0.39; hippocampus males *P* = 0.66; hippocampus females *P* = 0.62, protein levels same effects  **OFT/EZM**: NS in both M and F for SERT Ala276 homo and het mice  **TST/FST**: NS in both M and F for SERT Ala276 homo and het mice  **Social Dominance**: In M, Ala276 < WT, χ^2^ test **P* < 0.05; 80 bouts, *n* = 10–15, NS in F  **MBT**: NS in M, Ala276 Het and Homo Mice < WT, one-way ANOVA followed by Tukey’s multiple comparisons test, **P* < 0.05, *n* = 5–27 | **No effect of SERT Ala276:**  M = F, SERT mRNA/protein  M = F, mendelian genes  M = F, OFT, EPM, TST, FST    In M only, Social Dominance ↓ in SERT Ala276 mice  In F only, MBT ↓ in SERT Ala276 mice |
| Hatayama  2022 | Several time points P3, P7, P21, 6M | Newborn - 6mo | MS | Slitrk1-KO  USV, OFT, FST, Mouse Pup Behavior | **Body Weight:** KO Males < WT Males on P3 (p=0.022), NS on P7, P14, KO M < WT M on P21 and later (p<0.01), KO F < WT F only on P14  **Ultrasonic Vocalizations of isolated pups:** Call Rate KO F < WT F at P7 (p=0.0029), P10 (p=0.0012); Max Vocalization Amplitude KO < WT (male, *P* = 0.035; mixed-sex, *P* = 0.023), Vocalization syllables for U-shaped KO < WT P7 (M p=0.027, F p=0.013), P10 (male, *P* = 0.026)  **increased density of the NET-positive fibers in KO**: At P7, male, *P* = 0.0040; female, *P* = 0.045  varicosity size of NET fibers: P7 9.2% larger in KO males (p=0.044) and 7.7% smaller at 6M (p=0.0086)  **NET-Positive locus coeruleus in KO mice at P7:** male, +36%, *P* = 0.036; female, +17%, *P* = 0.10  **Varicosity size of SERT+ fibers**: +21% in KO M at P7 (p=0.0087), NS in KO F  **Varicosity size of DAT+ dopaminergic fibers in NAC**: NS all groups  **Monoamine transmitters at P7**: in PFC, NA+89% in KO M (p=0.0063) NS in F (+12%, p=0.62), *F*(1,22) = 1.96, *P*_genotype×sex_ = 0.18 in genotype × sex two-way analysis of variance [ANOVA], for MHPG, both KO M and F decrease MHPG, −81%, *F*(1,21) = 6.12, *P*_genotype_ = 0.022; MHPG/NA, −78%, *F(*1,21) = 5.64, *P*_genotype_ = 0.027 in genotype × sex two-way ANOVA, NS for dopamine, homovanillic acid +75% in KO M mice (p-0.017)  **Administration of Clomipramine or Clonidine**: At P7, serotonergic varicosity increase, male, +2.6%, *P* = 0.32; female, +4.1%, *P* = 0.0098; mixed-sex, +3.3%, *P* = 0.028 | Weight loss with KO: M > F  USV Call Rate M = F ↓  USV Voc Amplitude ↓ All  Voc Syllables ↓ All P7, M Only P10  NET+ Fibers ↑ M=F  Varicosity Size of Net Fibers: In M only, P7 ↑, 6M ↓  NET+ increase in LC: M > F  Varicosity size increase of SERT Fibers at P7: M > F  Varicosity size of dopaminergic fibers: KO=WT, M=F  Transmitters:  PFC NA ↑ M > F  MHPG ↓ M = F  Dopamine M=F, KO=WT  Homovanillic Acid ↑ M > F  Serotonic varicosity ↑ with clomipramine/conlidine M = F |
| Izyurov  2023 | A^Y^/a M 8  A^Y^/a F 8  CT M 8  CT F 8 | 6 mo | MS | *Lethal yellow* mutation mice  OFT, EPM, MBT, qPCR | **OPM:** A^Y^/a M = A^Y^/a F = CT M = CT F, Distance travelled F(3,28)=2.55, p=0.08, time center F(3,28)=1.06, p=0.38, vertical postures F(3,28)<1, grooming bouts F(3,28)<1; **EPM**: Distance travelled F(3,28)=6.45, p=0.002, time spent in open F(3,28)=11.08, p<0.001, closed arm F(3,28)=7.67, p<0.001, effect of genotype F(1,28)=9.61, p=0.004, effect of sex F(1,28)=9.08, p=0.005, no interaction, A^Y^/a M distance < CT M p<0.001; for open arm time sex x genotype F(1,28)=9.49, p=0.005, no sex effect alone, A^Y^/a M < a/a M p<0.001  **MBT:** Buried Marbles F(3,28)=6.15, p<0.03, effect of genotype F(3,28)=6.15 p<0.003, but not sex F(1,28)<1 or interaction F(1,28)=2.67, p=0.11, A^Y^/a F > CT F, **Gene Expression mRNA:** no sex effect, only genotype for *Agouti* and *Raly* for frontal cortex, hypothalamus, hippocampus; *Agrp* Four groups comparison Frontal Cortex F3,28 = 23.1,p<0.001, Hippocampus F3,28 = 18.26, p<0.001, Hypothalamus F3,28 < 1. Effect of Sex in frontal cortex F1,28=  44.24, p<0.001, Effect of sex F1,28= 13.81, p<0.001 and interaction F1,28= 14.68, p<0.001 in hippocampus; A^Y^/a M > CT M, no effect in F, *Pomc* Difference in Frontal Cortex F3,28 =4.6, p=0.01 but not hypothalamus and hippocampus, Interaction of genotype x Sex F1,28=11.86, p=0.0018, A^Y^/a F > CT F (p=0.013); *Mc3r* difference in hypothalamus F3,28 = 4.74, p=0.008 but not other regions, effect of genotype F1,28=7.45, p=0.011, A^Y^/a M < CT M, p=0.0024, *Mc4r* Difference in frontal cortex F3,28 =4.51, p=0.01, but not hippocampus or hypothalamus, A^Y^/a M < CT M, p=0.018 | No effect OPM  EPM: A^Y^/a M < CTM, no effect in F  MBT no effect of sex  Gene expression:  *Agrp* A^Y^/a M > CT M for frontal cortex and hippocampus  *Pomc* A^Y^/a F > CT F for frontal cortex  *Mc3r* A^Y^/a M < CT M in hypothalamus  *Mc4r* A^Y^/a M < CT M in frontal cortex |
| Partridge  2023 | 92 ICR (CD-1)  (45F) | 8 wk+ | MS | Buspirone, Ket, Flu  MBT | **Comparing bedding:** Effect of digging medium F(1, 36) = 6.91, *P* = 0.01), no main effect of sex F(1, 36) = 0.06, *P* = 0.81 or interaction F(1, 36) = 0.20, *P* = 0.66, Locomotor activity no interaction F(1, 36) = 0.00, *P* = 0.97, no effect of medium F(1, 36) = 0.07, *P* = 0.80 or sex F(1,36) = 1.76, *P* = 0.19  **Repeated Test Area Exposure:** no significant main effect of test exposures (F(3, 42) = 1.88, *P* = 0.15), no main effect of sex (F(1, 14) = 2.35, *P* = 0.15) or interaction (F(3, 42) = 0.40, *P* = 0.75)  **Effects of Drug**: *Buspirone*: # Marbles Buried: significant effect of drug dose (F(3, 27) = 10.74, *P* < 0.001), no main effect of sex (F(1, 9) = 0.23, *P* = 0.64) or interaction (F(3, 27) = 0.74, *P* = 0.54); Distance Travelled: significant effect of drug dose (F(3, 27) = 6.30, *P* < 0.01), no main effect of sex (F(1, 9) = 0.01, *P* = 0.92) or interaction (F(3, 27) = 1.05, *P* = 0.39); *Ketamine*: # Marbles Buried: significant effect of drug dose (F(4, 40) = 6.23, *P* < 0.001); no main effect of sex (F(1, 10) = 1.25, *P* = 0.28) or interaction (F(4, 40) = 0.06, *P* = 0.99); Distance Travelled: no significant effect of drug dose (F(4, 40) = 0.59, *P* < 0.01), no main effect of sex (F(1, 9) = 1.25, *P* = 0.28) or interaction (F(4, 40) = 1.01, *P* = 0.41), *Fluoxetine:* # Marbles Buried: significant effect of drug dose (F(3, 27) = 5.42, *P* < 0.01), no main effect of sex (F(1, 9) = 0.18, *P* = 0.68) or interaction (F(3, 27) = 0.445, *P* = 0.73); Distance Travelled: no significant effect of drug dose (F(3, 27) = 2.78, *P* = 0.06), no main effect of sex (F(1, 9) = 1.37, *P* = 0.27) or interaction (F(3, 27) = 0.28, *P* = 0.84) | For MBT with different bedding: M = F  Effects of Drug: M = F |
| Ho  2023 | M: 16 WT, 16 HT, 11 KO  F: 16 WT, 16 HT, 9 KO | 8-11 wk | MS | Protein Tyrosine Phosphatase Receptor Type D (PTPRD) – KO  OFT, Dig Test, grooming, PPI, nest building | **OFT:** Total Distance Travelled, Center Time, Center Distance Travelled, % Time in Center, Rearing, Spatial *d*: no effect of genotype, sex, or interaction  **Dig Test:** Time spent digging, latency to dig, number of dig bouts, and average duration of dig bout: no effect of genotype or sex, no interaction, F(1,2) = 2.00, p = 0.14  Splash Test: Time grooming, latency to groom, # groom bouts, average groom bout duration: no effect of genotype or sex  PPI: genotype by sex interaction [F(2, 78) = 3.31] (p<0.05) for percent prepulse inhibition, no other effects found. Effect of *Ptprd* genotype within females [F(2, 38) = 3.58] (p = 0.038) but not in males. Startle Magnitude main effect of sex [F(1, 78) = 8.51] (p<0.01), effect across blocks F(1, 78) = 8.78] (p < 0.01) | No Differences with *PTPRD*-KO in OFT, Dig Test, Splash Grooming Test    For PPI, *PTPRD*-KO in F ↓ PPI    For Startle Magnitude M > F |
| Caddedu  2023 | N = 8/group | 2-4 mo | MS | Cholinergic interneuron (CIN) depletion  Grooming, PPI | **Cholinergic Interneuron depletion:** CIN-d in M and F ChAT+ cells < WT, p<0.0001, overall effect of sex for CINs, p<0.0001  Baseline Behavior: CIN-d = WT for OFT, spontaneous grooming  PPI: Startle no main effect of ablation, sex, or interaction  **Acoustic PPI**: CIN-d × sex interaction: *F*(1,28) = 6.21, *P* = 0.02; partial *η*^2^ = 0.33  **Tactile Startle**: No effect CIN-d, Main effect of sex: *F*(1,28) = 4.64, *P* = 0.04; partial *η*^2^ = 0.14  **Cross-Modal PPI**: Sensorimotor gating CIN-d × sex interaction: *F*(1,28) = 6.43, *P* = 0.02; partial *η*^2^ = 0.38  **Spatial confinement effects on Grooming**: Increase in CIN-d M CIN-d × sex interaction: *F*(1,27) = 16.38, *P* = 0.0004; partial *η*^2^ = 0.34, digging CIN-d × sex interaction: F(1,28) = 10.99, *P* = 0.003; partial *η*^2^ = 0.28, head jerks CIN-d × sex interaction: *F*(1,28) = 15.00, *P* = 0.0006; partial *η*^2^ = 0.35, Acoustic Startle Amplitude Main effect of CIN-d: *F*(1,28) = 12.98, *P* = 0.001; partial *η*^2^ = 0.32, PPI Deficit CIN-d × sex interaction: *F*(1,28) = 8.02, *P* = 0.008; partial *η*^2^ = 0.25, Cross Modal PPI CIN-d × sex interaction: *F*(1,28) = 4.23, *P* = 0.04; partial *η*^2^ = 0.34, CIN-d M vs CT M p<0.05, CIN-d M vs CIN-d F p<0.01  **Marble Burying**: CIN-d × sex interaction: *F*(1,28) = 7.61, *P* = 0.01; partial *η*^2^ = 0.21  **Open Field**: No effect of CIN-d, M vs F p<0.01  **Treatment of male CIN-d mice**: *Haloperidol*: CIN-d × HAL grooming interaction: *F*(2,42) = 6.80, *P* = 0.003; partial *η*^2^ = 0.24, PPI deficits *F*(2,42) = 4.71, *P* = 0.01; partial *η*^2^ = 0.20, head jerks *F*(1,28) = 9.62, *P* = 0.004; partial *η*^2^ = 0.26, eye blinks *F*(1,28) = 4.23, *P* = 0.04; partial *η*^2^ = 0.13, *Clonidine* and *ecopipam*: similar effects to HAL  **Systemic AP in CIN-d Mice**: Grooming: Time × AP interaction: *F*(4,42) = 2.77, *P* = 0.04; partial *η*^2^ = 0.21, PPI *F*(1,26) = 4.50, *P* = 0.04; partial *η*^2^ = 0.15  **Direct injection of AP**: *mPFC:* Grooming Time × AP interaction: *F*(2,28) = 3.64, *P* = 0.04; partial *η*^2^ = 0.21, PPI Deficits (1,20) = 4.70, *P* = 0.04; partial *η*^2^ = 0.19, *DLS*: Grooming p<0.05, no effect PPI  **Blocking AP synthesis with FIN:** Grooming *F*(2,38) = 12.69, *P* < 0.0001; partial *η*^2^ = 0.40, PPI *F*(2,38) = 3.94, *P* = 0.03; partial *η*^2^ = 0.17 | Striatal CIN density: M > F  Acoustic PPI ↓ in CIN-d M, no effect in CIN-d F  Tactile Startle M < F  Sensorimotor Gating ↓ M only    **Spatial confinement effects in CIN-d males only:**  Grooming ↑, Digging ↑, Head Jerks ↑, Deficit in Cross-Modal PPI ↑, Marble Burying ↑    Acoustic startle amplitude ↑ M = F, EPM no effect of CIN-d, Baseline open arm M < F    Treatment of M CIN-d with Haloperidol, clonidine, or ecopipam: Symptoms ↓    Systemic AP administration in CIN-d Males: Worsens abnormal grooming, PPI deficits,  mPFC AP: Deficits worsen in both abnormal grooming and PPI  DLS: mild increase grooming, no effect PPI  Blocking AP synthesis in M: restores grooming and PPI |
| Huffstetler  2023 | 98 | 3mo | MS | Cannabidol (CBD) admin, Kv1.3 (Kv1.3−/−) mice, NOR, EPM, Object based attention test, MBT  LDB | **MBT:** WT M CT buries more than WT M CBD 10 mg/kg (p=0.0364), WT M CT = WT M CBD 20mg/kg, Kv1.3 M CT trends towards increased marbles vs Kv1.3 M CBD 10mg/kg (p=0.1078), Kv 1.3 M CT buries more than Kv1.3 M CBD 20mg/kg (p=0.0156); no effect of genotype or treatment in females WT F CT vs WT F CBD 10mg/kg (p=0.603), WT F CT vs WT F CBD 20mg/kg (p=0.8716), Kv1.3 F CT vs Kv1.3 F CBD 10mg/kg (p=0.6704), Kv1.3 F CT vs Kv1.3 F CBD 20mg/kg (p=0.5336)  **EPM**: no significant drug treatment x location interaction was observed (F 10 mg/kg - F(2,30) = 1.353, F 20 mg/kg - F(2, 24) = 1.390, M 10 mg/kg - F(2, 24) = 1.864, M 20 mg/kg - F(2,25) = 0.2323  **EPM in Kv1.3-/-:** F 10 mg/kg - F(2,21) = 6.380, *p* = 0.0251 closed arms, *p* = 0.0357 open arms; F 20 mg/kg - F(2,24) = 1.390; M 10 mg/kg - F(2,24) = 1.864, M 20 mg/kg - F(2,32) = 0.4156  **LDB**: In F drug x location interaction (F(1,20) = 5.507, *p* = 0.0294 (more time in light box) (CBD vs Veh 10 mg/kg - *p* < 0.001 vehicle, *p* > 0.05 CBD), no effect with 20mg/kg F(1,10) = 0.1573, *p* = 0.7000; In M no effect with 10mg/kg F(1,22) = 3.000, *p* = 0.0973, increased time in light with 20mg/kg F(1,18) = 8.237, *p* = 0.0102, (M 20 mg/kg - *p* < 0.0001 vehicle, *p* > 0.05 CBD)  **LDB in Kv1.3**: At 10mg/kg not a significant drug x location interaction for either male or female Kv1.3−/− mice (female, F(1,16) = 0.0009, *p* = 0.9763; male, F(1,16) = 0.0089, *p* = 0.9257, with 20 mg/kg more time in dark F 20 mg/kg - *p* > 0.05 vehicle, *p* < 0.01 CBD; M 20 mg/kg - *p* > 0.05 vehicle, *p* < 0.01 CBD  **NOR**: in M, 10mg/kg CBD in WT mice reduces recognition index (p=0.0484)  **Object Based Attention**: No effect in WT by sex or CBD treatment  In Kv1.3 F, CBD reduces recognition index p=0.0079, no effect of 20mg/kg  In Kv1.3 M, CBD enhances recognition index in 20mg/kg, p=0.037, NS in 10mg/kg | MBT WT:  CBD 10mg/kg M ↓, F no eff  MBT Kv1.3 -/-:  CBD 20mg/kg M ↓, F no eff  EPM WT:  M + F time in Closed Arm ↑  No effect of CBD  EPM Kv1.3 -/-:  CBD 10mg/kg F Closed Arm ↑ vs Veh, no effect M  LDB in WT:  10mg/kg in F ↓ time in dark  20mg/kg in M ↓ time in dark  LDB in Kv1/3 -/-:  20mg/kg in M + F ↑ time in dark  NOR: 1 hr, M = F, n no effect of genotype or CBD  NOR: 24 hour WT M CT > WT M CBD, no effect in F, or Kv1.3 -/- M  Attention: no effects in WT  In Kv1.3 F, CBD ↓ attention at 10mg/kg only  In Kv1.3 M CBD ↑ attention at 20mg/kg only |
| Abbasi  2024 | 72F | 7-8 wk | RAT | REM Sleep Deprivation (SD), OFT, MBT, FST, Hot Plate, Shuttle Box | **OFT:** Locomotor Activity: Sham vs CTRL NS, Sham vs REM SD, the effect of sleep (F _1,40_ = 164.24, P<0.001); the effect of lithium (F _3,40_ = 29.22, P<0.001);  the effect of sleep*lithium (F _3,40_ = 18.83, P<0.001)  **Anxiety:** (Time in Middle) Control Groups F 3,23 = 10.85, P<0.001, +lithium p<0.001, Sham vs REM SD: the effect of sleep (F _1,40_ = 58.05, P<0.001); the effect of lithium (F _3,40_ = 12.93, P<0.001); the effect of sleep*lithium (F _3,40_ = 2.56, P>0.05)  **MBT:** Sham vs CTRL NS; REM SD vs CTRL: the effect of sleep (F _1,40_ = 70.94, P<0.001);the effect of lithium (F _3,40_ = 13.18, P<0.001); the effect of sleep*lithium (F _3,40_ = 12.48, P<0.001)  **Immobility**: Control groups NS; Control vs REM SD NS  **Climbing**: +lithium in CTRL F _3,23_ =6.90, P<0.01, Sham vs REM SD the  effect of sleep (F _1,40_ = 8.80, P<0.01); the effect of lithium (F _3,40_ = 5.13, P<0.01); the effect of sleep*lithium (F _3,40_= 9.27, P<0.001)  **Pain Subthreshold**: in CTRL lithium 100mg/kg p<0.05; for REM SD vs Sham the effect of sleep (F _1,40_ = 90.59, P<0.001); the effect of lithium (F_3,40_ = 17.80, P<0.001); the effect of sleep*lithium (F _3,40_ = 1.11, P>0.05)  **Passive Avoidance:** Control vs Sham NS, Sham vs REM SD F _1,40_ = 12.51, P<0.001); the effect of lithium (F _3,40_ = 3.19, P<0.05); the effect of sleep*lithium (F_3,40_ = 1.43, P>0.05)  **Hippocampal BDNF**: CTRL +lithium F _3,23_ = 62.35, P<0.001; the effect of lithium (F 3_,40_ = 442.89, P<0.001) and the effect of lithium*sleep (F _3,40_ =  11.29, P<0.001), while the effect of sleep (F _1,40_ = 2.99, P>0.05)] | OFT: Locomotion: REM SD ↑, Lithium 50mg/100mg reduces  OFT: Time in Middle (Anxiety) REM SD ↑, Lithium 50/100mg reduces  MBT: REM SD Marbles Buried ↑, Lithium 50mg reduces, 100mg normalizes  Immobility: REM SD ↓, Lithium 50/100mg increases  Climbing REM SD = CTRL, Lithium 10mg ↑  Pain Subthreshold: REM SD ↓, 50/100mg increases  Passive Avoidance Memory: REM SD latency to entrance ↓, Lithium 50/100mg normalizes  BDNF: REM SD ↓, for all subgroups (CTRL, Sham, REM SD) +lithium 100mg BDNF ↑ |
| Houshyar  2024 | 72F | 7-8 wk | RAT | REM SD  OFT, MBT, Hot plate, FST, | **OFT:** *Locomotion:* Sham vs REM SD the effect of sleep (F_1,48_ = 93.69, *P* < 0.001); the effect of crocin (F_3,48_ = 36.10, *P* < 0.001); the effect of sleep*crocin (F_3,48_ = 33.93, *P* < 0.001), REM SD no treatment vs REM SD + crocin 25 mg/kg (*P* < 0.01), REM SD + crocin 50 mg/kg (*P* < 0.001), and REM SD + crocin 75 mg/kg (*P* < 0.001); *Time in Center*: Sham vs REM SD the effect of sleep (F_1,48_ = 27.16, *P* < 0.001); the effect of crocin (F_3,48_ = 5.54, *P* < 0.005); the effect of sleep*crocin (F_3,48_ = 4.29, *P* < 0.025), REM SD vs REM SD + crocin 25 mg/kg (*P* < 0.01), REM SD + crocin 50 mg/kg (p < 0.001), REM SD + crocin 75mg/kg (p<0.001); **MBT:** Sham vs REM SD the effect of sleep (F_1,48_ = 41.88, *P* < 0.001); the effect of crocin (F_3,48_ = 15.31, *P* < 0.001); the effect of sleep*crocin (F_3,48_ = 12.11, *P* < 0.001), REM SD vs all REM SD + crocin groups (25, 50, and 75 mg/kg) (p<0.001), **Hot Plate Pain Threshold:** Sham vs REM SD the effect of sleep (F_1,48_ = 16.17, *P* < 0.001); the effect of crocin (F_3,48_ = 10.54, *P* < 0.001); the effect of sleep*crocin (F_3,48_ = 6.10, *P* < 0.01), REM SD vs all REM SD groups+crocin (25mg/kg, *P* < 0.05; 50mg/kg, *P* < 0.001; 75mg/kg, *P* < 0.001)  **FST**: Sham vs REM SD effect of sleep (F_1,48_ = 3.95, *P* < 0.05); the effect of crocin (F_3,48_ = 10.00, *P* < 0.001); the effect of sleep*crocin (F_3,48_ = 12.25, *P* < 0.001), REM SD vs REM SD Crocin 25 mg/kg (*P* < 0.05), 50 mg/kg (*P* < 0.01), and 75 mg/kg (*P* < 0,001), **Climbing**: effect of sleep (F_1,48_ = 0.43, NS); the effect of sleep*crocin (F_3,48_ = 0.56, NS)] but the effect of crocin (F_3,48_ – 5.81, P < 0.05, only effect in Sham + Crocin 75mg/kg compared to Sham CT (p<0.05) | OFT: in REM SD Locomotion ↑, Crocin treatment in REM SD Locomotion normalized  Center Time ↑ in REM SD, Center Time normalize with crocin  MBT: Marbles buried ↑ in REM SD, ↓ with Crocin treatment  Pain Threshold in REM SD ↓ Normalized with Crocin  FST: Immobility ↓ in REM SD, normalized with Crocin |
| Salloum  2024 | 30F | 2mo | MS | 5-HT1B agonist RU24969  MBT, TST, Sucrose Preference, FST, NST | **MBT:** # Buried Marbles: 15mg/kg and 20mg/kg dose of RU vs CT, p<0.05  NST: Remaining Nestlet Weight 15mg/kg and 20mg/kg > 5mg/kg and 10mg/kg (p<0.05)  **FST and TST**: Immobility time, no effect of RU  **Sucrose Intake**: On D2, 15mg/kg and 20mg/kg > control (p<0.05)  **Gabra1 gene expression**: 15 (-1.01) and 20 mg/kg (-2.35) fold decrease (p<0.05), No significant effects noted for serotonin transporter  **GABA protein expression**: lowest value at 15mg/kg (2.7-fold decrease), *p* > 0.05 | MBT: # Marbles buried ↑ in F w/ 15mg/kg and 20mg/kg  NST: Remaining nestlet weight ↑ in 15 and 20mg/kg  FST and TST: No Effect  Some increase in sucrose intake in 15 and 20 mg/kg on D2 vs control  Gabra 1 gene ↓ with dose ↑  GABA protein ↓ with RU |
| Wilson  2024 | Baseline  Groom N=21-32/group  Environ Groom  N=6-13/group  Loco  N=6-12/group  Anxiety  N=6-12/group  NSFT  N=6-12/group | 8 wk | MS | SAPAP3 KO  Grooming, Locomotion, LDB, EPM, Gut analysis, NSFT | **Groomed Body Region:** significant effect of region (*F*_(3, 420)_ = 41.72, *p* < .0001, *η^2^* = 0.22), along with region x sex (*F*_(3, 420)_ = 3.38, *p* = .018, *η^2^* = 0.02) and region x genotype (*F*_(3, 420)_ = 3.83, *p* = .010, *η^2^* = 0.02; males: *F*_(3,232)_ = 15.51, *p* < .0001, *η^2^* = 0.16; females: *F*_(3, 141)_ = 19.62, *p* < .0001, *η^2^* = 0.29, Males: paws vs nose/face head p<0.0001, vs body p<0.0001, genitals vs nose/face/head p<0.0001, vs body p<0.005; Females: paws vs nose/face/head or body p<0.0001 or legs/tail/genitals p=0.004, legs/tails/genitals vs nose/face/head p=0.004  Females Only: Interaction region x genotype *F*_(3, 141)_ = 2.79, *p* = .043, *η^2^* = 0.04, KO vs WT p=0.042; **Grooming Time:** WT vs SAPAP3 KO *F*_(1, 105)_ = 27.51, *p* < .0001, *η^2^* = 0.18, genotype x sex interaction *F*_(1, 105)_ = 7.03, *p* = .009, *η^2^* = 0.05, M KO vs M CT p<0.0001, F KO vs F WT p=0.162  Bout Duration (trend): effect of sex *F*_(1, 105)_ = 3.26, *p* = .07, *η^2^* = 0.03, no interaction; **Locomotor Activity**: Overall SAPAP3 KO < WT *estimate* = −325.99, *SE* = 40.52, *t*_(548.62)_ = −8.05, *p* < .0001, and F > M *estimate* = 195.80, *SE* = 54.62, *t*_(227.99)_ = 3.59, *p* < .001; **Spatial *d*** : Increase in female mice *F*_(1, 99)_ = 14.72, *p* < .001, *η^2^* = 0.06, reduction in KO vs WT, *F*_(1, 99)_ = 104.20, *p* < .0001, *η^2^* = 0.43 sex x genotype x housing interaction (*F*_(2, 99)_ = 3.55, *p* = .033, *η^2^* = 0.03, effect of housing present in females *F*_(2, 45)_ = 4.68, *p* = .014, *η^2^* = 0.09, not males *F*_(2, 54)_ = 0.06, *p* = .941, *η^2^* < 0.01; **LDB:** significant effects of genotype (*F*_(1, 99)_ = 44.49, *p* < .0001, *η^2^* = 0.28) and housing (*F*_(2, 99)_ = 3.91, *p* = .023, *η^2^* = 0.05), but no effect of sex (*F*_(1, 99)_ = 0.04, *p* = .839, *η^2^* < 0.01, **EPM: s**ignificant effects of genotype (*F*_(1, 99)_ = 19.59, *p* < .0001, *η^2^* = 0.12) and housing (*F*_(2, 99)_ = 17.64, *p* < .0001, *η^2^* = 0.22), but no effect of sex (*F*_(1, 99)_ = 1.64, *p* = .203, *η^2^* = 0.01), **NSFT**: Rate of Pellet Consumption sex x housing(EX) effect (*HR* = 0.110, 95 % *CI* [0.026, 0.462, *p* = .003, M KO vs WT *HR* = 0.237, 95 % *CI* [0.119, 0.472], *p* < .001, no effect of housing, F KO vs WT *HR* = 0.229, 95 % *CI* [0.120, 0.435], *p* < .001, F Exercise vs Standard Housing *HR* = 6.214, 95 % *CI* [2.707, 14.264], *p* < .001, no interaction | Grooming Region:  M: Paws = Genitals > nose/face/head = body  F: Paws > legs/tails/genitals > nose/face/head  F KO vs WT: Paw grooming ↑  Grooming Time:  KO M > WT M, KO F = WT F    Exercise in F KO Mice reduces Spatial *d* more than standard housing or EE, effect not in M    No sex effect LDB  No sex effect EPM    NSFT Food Consumed  In SH M WT vs KO ↓  No effect of housing  In SH F WT vs KO ↓  EX WT ↑ in F than all others |
| Macedo  2024 | Exp1  N=10-12/group  Exp 2  CT M 17  CT F 15  Mem5 M 16  Mem5 F 16  Mem10 M 18  Mem10 F 18 | 8-12 wk | MS | Memantine administration  MBT, OFT, NBT | **Memantine in the MBT:** Effect of treatment F_(4.107)_ = 10.746; p < 0.001; ηp2 = 0.287, no effect of sex F(_1.107_)= 0.560; p = 0.456; ηp2 = 0.005; interaction between treatment and sex: F(_4.107_)= 1.437; p = 0.227; ηp2 = 0.051  Flu 10mg/kg reduces total distance in M (p<0.05) but not F, interaction between treatment and sex: F_(4.107)_ = 1.790; p = 0.136; ηp2 = 0.063  **Memantine in NBT:** % of cotton with Memantime, treatment effect: F_(2.94)_ = 7.054; p = 0.001; ηp2 = 0.130; interaction between treatment and section: F_(2.94)_ = 5.885; p = 0.004; ηp2 = 0.115; interaction between treatment, sex, and repeated measure: F_(2.94)_ = 3.161; p = 0.047; ηp2 = 0.060; At 1 hour: M/Mem 5mg/kg vs M/Veh p = 0.073; Cohen’s d = 1.20, 10mg/kg p = 0.015; Cohen’s d = 1.47), F/Mem 5mg/kg p = 0.463; Cohen’s d = 0.30, 10mg/kg p = 0.001; Cohen’s d = 0.98; 24 hours F/Mem 10mg/kg vs F/Veh p = 0.060; Cohen’s d = 0.74, M/Mem 10mg/kg vs M/Veh p = 1.000; Cohen’s d = 0.017  **L-ARG (NO precursor) on Mem in MBT**: interaction between sex and L-arginine: F_(1.91)_ = 4.627; p = 0.034; ηp2 = 0.048), in M/Mem + L-ARG vs M/Veh p = 0.904; Cohen’s d = 0.24, in F/Mem + L-ARG vs F/Veh p = 0.001; Cohen’s d = 2.40  **NBQX (AMPA receptor antagonist):** No effect on MEM5mg/kg or by sex | MBT: effect of mem M=F  Distance Traveled: Flu in M < all other sex/treatment  NBT: Removed cotton  1 hr 10mg/kg M = F, 5mg/kg M > F  24 hr 10mg/kg M < F  L-ARG mitigates effect of Mem 5mg/kg in M but not F |
| Minagawa  2025 | SHIRPA  12M, 12F  OF  10/group | 2mo – 12 mo | MS | Dlgap1 KO Mice  Grooming and Skin Lesions, OFT, SHIRPA | **Open Field Test:** Main effect of genotype (locomotor activity, *F*_2, 36_ = 67.27, *p* < 0.001; time spent in the center, *F*_2, 36_ = 48.72, *p* < 0.001; time spent in the periphery, *F*_2, 36_ = 18.01, *p* < 0.001; number of rearings, *F*_2, 36_ = 13.15, *p* < 0.001; number of grooming bouts, *F*_2, 36_ = 55.19, *p* < 0.001; grooming time, *F*_2, 36_ = 81.28, *p* < 0.001), no effect of sex or two-way interaction  **Grooming behavior following treatment with Fluvoxamine:** Non-Scratching bouts main effect of treatment (*F*_1, 11_ = 29.50, *p* < 0.001) and a significant interaction of sex and treatment (*F*_1, 11_ = 10.64, *p* < 0.001), Flu F vs Veh F p=0.021, Veh M vs Veh F p=0.022,  **Behavior of vehicle treated mice across sex and genotype:** main effects of genotype (non-scratching time, *F*_2, 16_ = 13.27, *p* < 0.001; number of non-scratching bouts, *F*_2, 16_ = 24.10, *p* < 0.001; number of scratching bouts, *F*_2, 16_ = 41.76, *p* < 0.001) and sex (non-scratching time, *F*_1, 8_ = 8.70, *p* = 0.018; number of non-scratching bouts, *F*_1, 8_ = 14.34, *p* = 0.005; number of scratching bouts, *F*_1, 8_ = 37.95, *p* < 0.001)**,** interaction genotype x sex for number of scratching bouts (*F*_2, 16_ = 16.79, *p* < 0.001), but not for the others, for bouts smaller contract in male vs female (vs. WT, *p* = 0.024; vs. HT, *p* = 0.048) | # of non-scratching bouts M < F in Veh treatment, Veh F > Fluvoxamine F, Veh M = Fluvoxamine M |
| Crummy  2025 | 24 (12F) | 8wk | MS | Operative Negative Reinforcement Paradigm, OFT, EPM, LDT, EZM | **Acquisition of avoidance:** main effect of sex *F*_(1,29)_ = 1.03, *p* = 0.32; main effect of session *F*_(4.51,130.6)_ = 15.08, *ε* = 0.32, *p* < 0.0001; interaction of sex × session *F*_(14,406)_ = 0.58, *p* = 0.88, **Acquisition Rates**: chi-squared Fisher's exact test *p* = 0.69 | M = F on Operant Negative Reinforcement Learning for avoidance |
| Ratsuki  2025 | 7007  (917 female cages, 588 breeders) | N/A | MS | Barbering analysis in home cage | **Barbering risk factors:** In female cages LR χ^2^ = 3.89, DF = 1, P = 0.0485 | Abnormal grooming/barbering behaviors: M < F |
| Brownstien  2024 | 50 (22F) | 6-7m | MS | Sapap3-KO  Grooming Analysis, MBT, OFT, EPM, | **Grooming Analysis:** NS between M and F for Baseline Grooming Duration (F = 569.2 +184.1, M = 721.4 + 279.6), Baseline Grooming Bouts (F = 104.2 +23.15, M = 138.6 + 44.64), Baseline Short Grooming Bouts (F = 45.8 + 19.84, M = 38.40 + 12.84). Baseline Long Grooming Bouts (F = 23.40 + 8.649, M = 31.60 + 9.290), Baseline Head-Body Twitch (F = 6.60 + 4.519, M = 5.40 + 4.669)  **Treatment on Grooming Phenotypes**: no difference between M and F | Excessive self-grooming and head-body twitches in Sapap3-KO mice: M = F  Treatment effects of PSIL and PME: M + F |
| Coppola   2025 | M = 21  F = 17 | 5-8 wks | MS | MBT, EPM, Grooming, Hole Board Test, Y-Maze | **MBT Baseline: # Marbles Buried**: M > F t = -2.935, p = 0.0058, All time intervales sex × interval interaction: F (2,70) = 2.86, p = 0.0642, post hoc M > F p<0.01; Timing sex × intervals interaction: F (2,72) = 6.978, p = 0.0017, Females consistent across 30 m, Males start after 10 m  **Digging Behaviors**: *M > F* Snout Digging (t = 2.729, p = 0.0098), Paw Digging (t = 3.179, p = 0.003), Marble Directed Digging (t = 2.275, p = 0.0290), Displacement Digging (t = 2.565, p = 0.0148), Marble Sniffing (t = 3.743, p = 0.0006); *F > M* Unsupported (t = 2.961, p = 0.0055) and Supported Rearing (t = 3.068, p = 0.0041); *M = F* Self Grooming (t = 0.2815, p = 0.7799)  No sex differences in other behavioral tests except Total # Arm Entries in Y Maze: M > F (t = -2.210, p = 0.0336)  **PCA Analysis on MBT and EPM Measures**: Based on PC1 and PC2 scores, Male Scores Lower along PC1 (more compulsive/anxiety-like behaviors) and Females Higher (exploratory behaviors), t = 3.117, p = 0.0036, PC2 scores equivalent | For MBT behavioral analysis, M > F for marble related behaviors and anxiety/compulsive behaviors, M < F for exploratory behaviors |
| Gattuso  2025 | 6-8 M/group  7-11 F/group | 4-7 mo | MS | SAPAP3 KO, Psilocybin  Locomotor, Head-twitch, Soc. Interact Test, LDB, PPI, Gut Microbiota Analysis, FST | **Locomotion**: Psilocybin > Veh in WT but not SAPAP3 KO F_2,87_ = 4.55; *p* = 0.013, η^2^ = 0.050, p<0.001; WT F > WT M (F_1,87_ = 7.387, *p* = 0.008, η^2^ = 0.049, p=0.017; SAPAP3 KO < WT (F_1,87_ = 45.18, *p* < 0.0001) regardless of sex  **Head Twitch**: M Psilocybin (0.1mg/kg; F_1,40_ = 21.40; *p* < 0.0001; and 1.0mg/kg; F_1,42_ = 31.43; *p* < 0.0001) vs Vehicle; F Psilocybin (0.1 mg/kg; F_1,49_ = 10.4; *p* = 0.002; 1.0mg/kg; F_1,53_ = 34.6; *p* < 0.0001)  **Sociability**: SAPAP3 < WT in both M and F for sociability (F_1,72.67_ = 27.90, *p* < 0.0001) and social novelty (F_1,72.89_ = 6.172, *p* = 0.015); Effect of genotype x treatment x sex (F_2,71.18_ = 3.846, *p* = 0.026) where Male WT with psilocybin > M WT vehicle  **Compulsive Behavior**: , enhanced compulsive behavior in SAPAP3 K); No sex effect (F_1,83_ = 1.251, *p* = 0.266, η^2^ = 0.011)  **LDB**: Increased anxiety-like behavior in SAPAP3 KO; No sex effect (F_1,83_ = 1.525, *p* = 0.220, η^2^ = 0.013)  **PPI**: No effects on PPI in SAPAP3 KO mice, no sex effects (F _1,83_ = 0.323; *p* = 0.571, η^2^ = 0.004); for startle M with psilocybin 1.0mg/kg < all vehicle controls (F_2,83_ = 5.134; *p* = 0.008, η^2^ = 0.069, p<0.017)  **Gut transit time**: NS for M vs F; Water content M > F (F_1,80_ = 48.50; *p* < 0.0001, η^2^ = 0.345); no interaction sex x treatment; Fecal output Sex x Genotype (F_1,83_ = 7.180; *p* = 0.009, η^2^ = 0.068) SAPAP3 KO M > WT (p<0.017) | For locomotion: Distance Travelled WT F > WT M; SAPAP3 KO < WT with M = F  For Head Twitch: Psilocybin = Head Twitch ↑; M = F  For Sociability: SAPAP3 < WT; M = F; Psilocybin treatment in WT M Soc ↑  For Compulsive Behavior, LDB, PPI, no effect of sex shown  For Fecal output: SAPAP3 KO > WT in M only |
| Gorburg  2025 | n’s not reported | 6 wks | MS | 2,5-dimethoxy-4-iodoamphetamine (DOI) model of TS/OCD-like behavior  HTR, ESR, Grooming | **Effects of raloxifene on DOI-induced Behavior:**  **Head Twitch Response (HTR)**: In M: At 5mg/kg HTR ↓ 16%, At 10mg/kg HTR ↓ 29%; In F: At 5mg/kg HTR ↓ 12%, At 10mg/kg HTR ↓ 23%; F (1, 40) = 6.140, p = 0.0175 for 5 mg/kg; F (1, 40) = 6.758, p = 0.0130 for 10 mg/kg  HSR at 20mg/kg High Dose: In M HTR ↑ 25%; In F HTR ↓ 16%; F (1, 40) = 5.243, p = 0.0274  **Ear Scratch Response (ESR)**: In M: NS on ESR for 5, 10, 20mg/kg; In F: ESR ↓ 61% for 10mg/kg, ↓ 52% for 20mg/kg (F (1, 40) = 6.088, p = 0.0180, for 10 mg/kg; F (1, 40) = 7.556, p = 0.0089, for 20 mg/kg)  **Grooming:** In M, 10mg/kg DOI-induced grooming ↓ 45%; In F, 10mg/kg DOI-induced grooming ↓ 36% (F (1, 40) = 11.03, p = 0.0019)  **In healthy (no DOI) mice**, NS effect of raloxifene on HTR, ESR for both M and F. In 10mg/kg grooming ↓ in F only (F (1, 45) = 5.590, p = 0.0224) | In both M and F: Raloxifene reduced HTR at 5mg/kg and 10 mg/kg  At 20mg/kg: M HTR ↑, F HTR ↓  For ESR: Raloxifene NS in M, ESR ↓ in F  For grooming, DOI-induced groom ↓, M=F  In non DOI mice, groom ↓ at 10mg/kg in F only |
| Kajita  2025 | 10-14 F rat/group | MS at PND 1-9 or 10-20  Behav. PND 49-74 | RAT | Maternal Separation (MS)  SPT, MBT, OFT, LDT, Social Interact, EPM, FST | **Comparative Analysis of Female (Kajita, 2025) with male rats from previous studies:**  **FST, Immobility Score:** For F: CT 34.0 ± 1.6 , Early MS 41.1 ± 1.8, Late MS 42.4 ± 1.1; For M: CT 38.3 ± 1.9, Early MS 41.3 ± 1.8, Late MS 44.8 ± 1.6  **FST, Climbing Score:** For F: CT 11.0 (11.5), Early MS 6.5 (7.0), Late MS 8.0 (10.0); For M: CT 8.92 ± 0.87, Early MS 8.61 ± 1.07, Late MS 8.33 ± 1.38  **SPT Rate**: For F: CT 0.93 (0.04), Early MS 0.94 (0.05), Late MS 0.92 (0.05), For M: CT 0.98 (0.03), Early MS 0.96 (0.05), Late MS 0.96 (0.03)  **MBT (# Marbles):** For F: CT 7.0 (6.0), Early MS 8.0 (5.0), Late MS 11.0 (2.25); For M: CT 7.31 ± 0.81, Early MS 7.75 ± 0.72, Late MS 6.18 ± 0.80  **Spine Density (Basal Dendrites):** For F: CT 0.76 ± 0.04, Early MS 1.00 ± 0.07, Late MS 0.70 ± 0.05; For M: CT 1.66 ± 0.04, Early MS 1.82 ± 0.05, Late MS 1.50 ± 0.06  **Spine Density (Apical Dendrites):** For F: CT 0.94 (0.35), Early MS 1.00 (0.50), Late MS 0.52 (0.25); For M: CT 1.66 ± 0.04, Early MS 1.74 ± 0.05, Late MS 1.36 ± 0.05  Reported anxiety behaviors in OFT, LDB, EPM, and Soc unaffected in both M and F | FST: Immobility:  For Early MS ↑, F only  For Late MS ↑, F = M  Climbing: For Early MS, ↓ for F only  SPT: For Late MS, ↓ for M only  MBT: For Late MS, ↑ F Only  Spine Density:  Basal Dendrites: In Early MS ↑, F Only  Apical Dendrites: For Late MS, ↓ M = F  Anxiety Behaviors: M=F, NS |
| Lazar  2025 | Study 1: SAPAP3 KO 20M, 24F; HET 21M, 21F; WT 26M, 29F  Study 2: SAPAP3-KO HOM + WT, 16/group | Study1: 11.03 ± 0. 071 wks; Study 2: 11.85 ± 0.87 wks | MS | MBT, OFT, EPM, Y-Maze, NOR, FST, Tube Dom, Oreo Test, SOC, Synaptic Protein Analysids | **Study 1:** Evaluation of anxiety and depression-like phenotypes in juvenile SAPAP3 KO: Sex NS for OFT, EPM, MBT, Tube Dominance  **For Oreo Test**: *Found Oreo*: M WT: 88.96%, M HET: 81.82%, M SAPAP3 HOM: 35%, χ^2^ (df)=16.74 (2), p=0.0002; F WT: 62.07%, F HET: 61.9%, F SAPAP3 HOM: 58.33%, χ^2^ (df)=0.10(2), p=0.93; *Ate Oreo*: M WT: 40%, M HET: 33.3%, M SAPAP3 HOM: 57.14%, χ^2^ (df)=1.25(2), p=0.54; F WT: 50%, F Het: 76.92%, F SAPAP3 HOM: 64.29%, χ^2^ (df)=2.49(2), p=0.28  S  **Study 2:** No effect of sex on any measures.  **Study 2: Oreo Test:** *Found Oreo*: M WT=100%, M HOM=50%, p<0.05; F WT=87.5%, F HOM=25%, p<0.01; Psilocybin treated M WT=87.5%, M HOM=25%, p<0.01, F WT = 100%, F HOM=37.5%, p<0.01 | No effect of sex on SAPAP3 juveniles for OFT, EPM, MBT, Tube Dom  M SAPAP3 HOM KO less likely to find buried sweet  Psilocybin no effect by sex on found oreos |
| Pandey  2025 | 6-12/group | 8-10 wk | MS | NL2 StopKI mice (genetic deletion of NL2 C-tail)  OFT, EPM, MBT, Nestlet shred, Grooming, SOC Pref, SOC Novelty | For both NOR and OFT Distance travelled, no effect of StopKI or sex.  For OFT Center Time: main effect of genotype, *F*_(1,43)_ = 15.47; *p* = 0.0003, no sex or interaction; For # of center zone entries: main effect of genotype, *F*_(1,43)_ = 26.33; *p* < 0.0001, no sex or interaction  For EPM: Time in Open Arm: main effect of genotype, *F*_(1,46)_ = 54.98; *p* < 0.0001; main effect of gender, *F*_(1,46)_ = 0.7074; *p* = 0.4047; interaction, *F*_(1,46)_ = 2.647; *p* = 0.1106, # Entries Open Arm: main effect of genotype, *F*_(1,46)_ = 35.42; *p* < 0.0001; main effect of gender, *F*_(1,46)_ = 7.605; *p* = 0.0083; interaction, *F*_(1,46)_ = 2.148; *p* = 0.1495  For MBT: # Marbles Buried: main effect of genotype, main effect of genotype, F_(1,22)_ = 42.73; p < 0.0001; main effect of gender, F_(1,22)_ = 9.322; p = 0.0058; interaction, F_(1,22)_ = 0.2589; p = 0.6159 Latency to Bury: main effect of genotype, *F*_(1,22)_ = 53.85; *p* < 0.0001; main effect of gender, *F*_(1,22)_ = 6.592; *p* = 0.0176; interaction, *F*_(1,22)_ = 0.01141; *p* = 0.9159  For Nestlet Shredding: main effect of genotype, *F*_(1,22)_ = 18.89; *p* = 0.0003; main effect of gender, *F*_(1,22)_ = 0.04089; *p* = 0.8416; interaction, *F*_(1,22)_ = 0.004615; *p* = 0.9465  For Grooming: Time: main effect of genotype, *F*_(1,41)_ = 21.56; *p* < 0.0001; main effect of gender, *F*_(1,41)_ = 2.547; *p* = 0.1181; interaction, *F*_(1,41)_ = 0.4044; *p* = 0.5284, # Bouts: main effect of genotype, *F*_(1,41)_ = 36.04; *p* < 0.0001; main effect of gender, *F*_(1,41)_ = 2.032; *p* = 0.1616; interaction, *F*_(1,41)_ = 1.763; *p* = 0.1916  For Social Preference: Interaction time male, main effect of novelty, *F*_(1,48)_ = 32.91; *p* < 0.0001; main effect of genotype, *F*_(1,48)_ = 3.058; *p* = 0.0867; interaction, *F*_(1,48)_ = 10.55; *p* = 0.0021; for Interaction time female, main effect of novelty, *F*_(1,46)_ = 36.47; *p* < 0.0001; main effect of genotype, *F*_(1,46)_ = 8.530; *p* = 0.0054; interaction, *F*_(1,46)_ = 25.83; *p* < 0.0001, For Social Novelty: Interaction time male, main effect of preference, *F*_(1,48)_ = 12.45; *p* = 0.0009; main effect of genotype, *F*_(1,48)_ = 4.005; *p* = 0.0510; interaction, *F*_(1,48)_ = 14.58; *p* = 0.0004; for Interaction time female, main effect of preference, *F*_(1,46)_ = 8.652; *p* = 0.0051; main effect of genotype, *F*_(1,46)_ = 5.062; *p* = 0.0293; interaction, *F*_(1,46)_ = 4.476; *p* = 0.0398 | For OFT, Anx ↑ for StopKI, M = F  For EPM: Anx ↑ for StopKI, for # entries, Diff in M < F, but both exhibit phenotype  For MBT, Nestlet shred and Grooming: ↑ for StopKI, M > F for effect of gender, but no interaction of genotype x sex  For Social Preference: StopKI show no preference for MS vs object; M = F  For Social Novelty: StopKI show no preference for new MS vs familiar MS; M = F |
| Stoppiglia  2025 | 73M, 72F | 10-12 wks | RAT | mCPP Model, T-Maze | **Acquisition and Reversal Learning in T-Maze:** *Acquisition Learning: Accuracy*: NS for effect of sex and sex x session interaction. *Reaction Time*: NS for effect of sex and sex x session interaction. *Omissions*: NS for effect of sex and sex x session interaction. *Reversal Learning*: *Accuracy*: NS for effect of sex and sex x session interaction. *Reaction Time*: NS for effect of sex and sex x session interaction. *Omissions*: NS for effect of sex and sex x session interaction. *Flexibility Score: Accuracy:* Effect of Sex F1,22 = 2.46, p = 0.13, Effect of Session **F2,44 = 115.14, p < 0.001**, Sex x Session **F2,44 = 3.42, p = 0.04**; *Reaction Time*: Effect of Sex: NS for effect of sex and sex x session interaction.  **mCPP in Males**: *Acquisition Learning: Accuracy; Reaction Time, Omissions*, effect of Session only, no effect of treatment; *Reversal Learning*: *Accuracy*: NS for treatment and treatment x session; *Reaction Time*: Effect of Treatment F4,56 = 3.56, p = 0.01, Effect of Session F2,112 = 9.60, p < 0.001, Treatment x Session F8,112 = 2.49, p = 0.02; *Omissions* Effect of Treatment F4,56 = 3.13, p = 0.02, Effect of Session F2,112 = 11.60, p < 0.001, Treatment x Session F8,112 = 4.33, p < 0.001; *Flexibility Score: Accuracy,* no effect of treatment or treatment x session; *Reaction Time*: Effect of treatment F4,54 = 4.78, p = 0.002, Effect of Session F2,108 = 3.34, p = 0.04, Session x Treatment F8,108 = 4.49, p < 0.001  **mCPP in Females:** *Acquisition Learning:* *Accuracy*: Effect of Session F4,55 = 8.10, p < 0.001, Effect of Treatment F4,220 = 59.36, p < 0.001 , NS for Treatment x Session; *Reaction Time*: Effect of Treatment F4,51 = 8.91, p < 0.001, Effect of Session F4,204 = 44.30, p < 0.001, NS Treatment x Session; *Omissions*: Effect of Treatment F4,55 = 10.10, p < 0.001, Effect of Session F4,220 = 15.87, p < 0.001, NS for Treatment x Session*; Reversal Learning: Accuracy*: NS for Treatment, Effect of Treatment NS, Effect of Session F2,110 = 111.69, p < 0.001, Treatment x Session F8,110 = 2.64, p = 0.01; *Reaction Time*; Effect of Treatment F4,52 = 6.31, p < 0.001, Effect of Session F2,104 = 14.97, p < 0.001, Treatment x Session F8,104 = 2.28, p = 0.03; *Omissions*: Effect of Treatment F4,55 = 7.43, p < 0.001, Effect of Session F2,110 = 8.98, p < 0.001, Treatment x Session F8,110 = 6.55, p < 0.001, *Flexibility Score: Accuracy:* Effect of Treatment: F4,55 = 3.73, p = 0.01, NS for Effect of Session, Session x Treatment; *Reaction Time*: Effect of Treatment F4,51 = 6.53, p < 0.001, Effect of Session F2,102 = 5.19, p = 0.01, NS for Treatment x Session Effect of Sex, Effect of Session, Sex x Session | In T-Maze: No sex effects for Acquisition Learning.  No sex effects for Reversal learning.  For Accuracy in flexibility scores, sex x session detected, but no post-hoc analysis revealed.  In M with mCPP:  Acquisition Learning: No effect of treatment, no interaction  Reversal Learning:  sal-mCPP2 longer reaction time + more omissions vs sal-sal and mCPP0.5-sal  Flexibility Scores: Longer reaction times in sal-mCPP2 vs mCPP0.5-sal and mCPP2-sal; sal-mCPP2 longer reaction time than mCPP0.5-sal and MCPP2-sal.   In F with mCPP:  Acquisition learning: Decreased accuracy, increase reaction time and omissions in mCPP2-sal vs all other groups.  Reversal Learning: More omissions in sal-mCPP2 group vs all other groups, increased reaction time in sal-mCPP2 vs sal-sal, mCPP0.5-sal and mCPP2-sal. Increased time in sal-mCPP2 vs other groups in multiple sessions.  Flexibility Scores: Higher accuracy in mCPP2-sal and longer reaction in sal-mCPP0.5 and sal-MCPP2 |
| Wilson  2025 | Grooming/  Anxiety/EPM  n=6-12/group  NSFT n=5-11  Protein n=6-7 | SIS at 5wks, Behav at 5wk, 8wk, 11wks | MS | SAPAP3 Model  Grooming assessment, LDB, EPM, NSFT, Proteomics | **Behavior experiments timeline:** 5wks, 8wks, 11wks of age, Social Isolation Stress (SIS) start at 5 wks baseline  **Time Spent Grooming**: Males: Standard Housing: SAPAP3 KO > WT CTRL at 5wks p<0.01 and 8wks p<0.05, SIS SAPAP3 KO > WT at 11 wk p<0.001; Females: Standard Housing SAPAP3 KO = WT CTRL, p>0.05; SIS SAPAP3 KO = WT CTRL, p>0.05; **Average Grooming Bout Length**: For M SAPAP3 KO > WT CT (p<0.01), For F SAPAP3 KO = WT CT (p>0.05); **Interrupted Grooming Bouts**: SAPAP3 KO > WT CT, M p<0.05, F p<0.001  **For NSFT**: Both M and F SAPAP3 KO Mice eat slower than WT CTRL regardless of SI-Stress males: *HR* = 0.181, 95 % *CI* [0.041, 0.806], *p* = .025; females: *HR* = 0.056, 95 % *CI* [0.011, 0.275], *p* < .001  **EPM**: In both M (p<0.05) and F (p<0.01), SIS SAPAP3 KO < Standard SAPAP3 KO for Time in Open Arms, For both M (p<0.01) and F (p<0.001), SIS SAPAP3 KO < SIS WT CT for Time. For both SAPAP3 KO Standard = WT CT Standard.  **Anxiety in Light Dark Box, Time in Light Side**: Males: At 5wk/8wk, both Standard/SIS SAPAP3 KO=WT CT, At 11 wks WT CT > SAPAP3 KO for Standard p<0.001 and SIS p<0.05; Females: At 5wk, WT CT = SAPAP3 KO for Standard and SIS (trend SIS p=0.07); At 8 wks, WT CT > SAPAP3 KO Standard (p<0.05) and SIS (p<0.01), At 11 wks WT CT > SAPAP3 KO Standard (p<0.01) and SIS (p<0.001)  **Protein expression**: In M PFC of SIS SAPAP3 KO: Downregulation of sialin (−log10(p) = 1.88, *FC* = 3.91), dual specificity calcium/calmodulin-dependent 3′,5′-cyclic nucleotide phosphodiesterase 1C (−log10(p) = 2.38, *FC* = 2.89), mediator of RNA polymerase II transcription subunit 12 (−log10(p) = 2.28, *FC* = 2.34), endothelin receptor type B (−log10(p) = 2.87, *FC* = 2.16), EF-hand domain-containing protein D1 (−log10(p) = 3.20, *FC* = 1.96), and protein noxp20 (−log10(p) = 3.67, *FC* = 1.71; In F PFC of SIS SAPAP3 KO: Downregulation SAPAP3 (−log10(p) = 8.03, *FC* = 4.04) and pulmonary surfactant-associated protein B (−log10(p) = 4.98, *FC* = 2.42); Upregulation neutrophilic granule protein (−log10(p) = 3.36, *FC* = 0.48), sulfotransferase 1A1 (−log10(p) = 4.10, *FC* = 0.48), and ethanolamine-phosphate phospho-lyase (−log10(p) = 2.69, *FC* = 0.36) | **For Grooming**: % Time SAPAP3 KO M > CT, SIS worsen SAPAP3 KO M over time, F NS; Average Bout Length SAPAP3 KO M > M CT; F NS; Interrupted bouts both M and F SAPAP3 KO > WT  **For Anxiety**: In *EPM*, SIS worsens anxiety in SAPAP3 KO, M=F; for *NSFT*, SAPAP3 KO reduces food consumption regardless of SIS, M=F; For *LDB*: anxiety worsens over time in SAPAP 3 KO regardless of stress, F effect occurs earlier (in M, 11 wk, in F, 8 wk)  **Changes in protein expression** in the PFC of SIS SAPAP3 KO mice different by sex, M proteins mostly involved in nervous system development and function |
| Payamandi  2025 | 9/group (6/group for behavior, 3/group for gene expr.) | Age | Rat | REM Sleep Deprivation (SD), Fear Conditioning (FC), OFT, MBT, PCR | Groups: CT, 1wk REM SD (6h/d), 2wk REM SD, 3wk REM SD; CTwFC, 1wk w FC, 2wk w FC, 3wk w FC  **Freezing (PTSD-like):** effect of REM SD (F_3,80_ = 98.60, *P* < 0.001), FC (F_1,80_ = 3202.50, P < 0.001), sex (F_1,80_ = 10.43, *P* < 0.01), REM SD*FC (F_3,80_ = 95.99, P < 0.001), REM SD *sex (F_3,80_ = 3.06, P < 0.05), FC*sex (F_1,80_ = 8.06, *P* < 0.01), REM SD*FC*sex (F_3,80_ = 3.61, P < 0.05); Male vs Female REM SD 1w and 2w: P < 0.05, 3w: *P* < 0.01  **Locomotor:** effect of REM SD (F_3,80_ = 280.10, *P* < 0.001), FC (F_1,80_ = 338.84, P < 0.001), sex (F_1,80_ = 1.40, *P* > 0.05), REM SD*FC (F_3,80_ = 54.49, P < 0.001), REM SD *sex (F_3,80_ = 22.00, *P* < 0.001), FC*sex (F_1,80_ = 0.16, P > 0.05), REM SD*FC*sex (F_3,80_ = 21.01, *P* < 0.001); 2wk and 3wk both sexes *P* < 0.001; 3wk F p<0.001, 3wk M p<0.01; FC in both sexes locomotion down P < 0.001; effect of 1w REM in FC stronger in M than F *P* < 0.01  **Anxiety in OFT:** effect of REM SD (F_3,80_ = 9.30, *P* < 0.001), FC (F_1,80_ = 65.50, P < 0.001), sex (F_1,80_ = 6.61, *P* < 0.05), REM SD*FC (F_3,80_ = 22.01, *P* < 0.001), REM SD *sex (F_3,80_ = 6.15, *P* < 0.001), FC*sex (F_1,80_ = 2.94, *P* > 0.05), REM SD*FC*sex (F_3,80_ = 1.47, P > 0.05). In REM SD F only, anxiety like behavior up (p<0.01)  **Grooming (OCD-Like):** effect of REM SD (F_3,80_ = 152.20, *P* < 0.001), FC (F_1,80_ = 215.52, P < 0.001), sex (F_1,80_ = 8.62, *P* < 0.01), REM SD*FC (F_3,80_ = 19.90, P < 0.001), REM SD *sex (F_3,80_ = 1.39, *P* > 0.05), FC*sex (F_1,80_ = 0.34, P > 0.05), REM SD*FC*sex (F_3,80_ = 3.03, P < 0.05)  **MBT:** effect of REM SD (F_3,80_ = 384.11, P < 0.001), FC (F_1,80_ = 45.00, P < 0.001), sex (F_1,80_ = 245.00, P < 0.001), REM SD*FC (F_3,80_ = 3.07, P < 0.05), REM SD *sex (F_3,80_ = 28.91, P < 0.001), FC*sex (F_1,80_ = 0.31, P > 0.05), REM SD*FC*sex (F_3,80_ = 4.53, P < 0.01)  **BDNF in hippocampus:** effect of REM SD (F_3,80_ = 68.45, P < 0.001), FC (F_1,80_ = 13.23, P < 0.001), sex (F_1,80_ = 25.94, P < 0.001), REM SD*FC (F_3,80_ = 106.10, P < 0.001), REM SD *sex (F_3,80_ = 1.16, P > 0.05), FC*sex (F_1,80_ = 1.47, P > 0.05), REM SD*FC*sex (F_3,80_ = 2.41, P > 0.05). | Freezing: ↑ in both M and F FC rats, decrease with increased REM SD in both M and F; M > F for freezing  Locomotion: For both M and F REM SD mice at 2 and 3 wks, ↑  Anxiety-like behavior: in REM SD, only 1wk F ↑  For FS+REM SD Anx in OFT: REM SD counteracts FC induced anxiety in both M and F  For Grooming: Both M and F grooming ↑ with REM SD for both REM SD alone and FC + REM SD  For MBT:  REM SD = # Marbles ↑  Females = # Marbles ↑  Changes in BDNF show same patterns in M and F |
| Wang  2025 | 5M/5F for both WT + SAPAP3 KO for behavior  10/sex/grp for tube dom and oxytocin | 4MO | MS | SAPAP3 KO, OFT, Aversive stimulus, Tube Dom, Oxytocin treatment intervention, Grooming | **Grooming Behavior**: SAPAP3 grooming up: 467.43 ± 94.98 s vs. WT 391.62 ± 86.44 s, *p* < 0.05; No interaction of sex x genotype F*₁*,*₈*= 0.1948, *p* > 0.05, KO F vs KO M: 526.59 ± 25.69 s vs. 408.26 ± 104.33 s, *p* < 0.05  **Tube Dominance Test:** Effect of Sex x Gene *F (1*, *36)* = 8.471, p=0.0062; Effect of Sex *F (1*, *36)* = 0.000, p>0.9999; Effect of Gene *F (1*, *36)* = 8.471, p=0.0062  Tube Dominance with Oxytocin: NS for Sex x Gene, NS for Sex, Effect of Gene p=0.0658 | Grooming: M < F for SAPAP3 KO   For Tube Dominance: SAPAP3 more aggressive.  No effect of oxytocin by sex. |
| Sachse  2026 | 9M, 8F | 3MO+ | RAT | DBS, Set-Shift Operant Test | **Effects of Mid-Striatal Deep Brain Stimulation (DBS) on Set-Shift Operant Tast:**  **Baseline:** Response Time (RT): light rule: *p* = 0.0466, side rule: *p* = 0.0486; Accuracy: Side: *p* < 0.00001, Light: *p* = 1.431, OFF Trial Omissions: p>0.05  **Stimulation Effects:** RT: In M, no stimulation condition reduced RT (p>0.05), In F, bilateral: *p* < 0.00001, right: *p* < 0.00001, left: *p* = 0.0099; bilateral vs left *p* = 0.0162; M vs F Bilateral/Right stim greater effect in F bilateral: *p* = 0.0084, right: *p* = 0.0478; Left same *p* = 0.07437; Accuracy, no sex differences; Omissions, only males with left stim *p* = 0.0291, all other *p*s > 0.05 | RT in Set-Shift: F slower than M; Accuracy in F lower for Side rule, but M=F for Light Rule  OFF Trial Omission M=F  In M, no effect of stim on RT.  In F, stim all types = RT ↓  For Bilateral Stim and Right Stim RT ↓ M < F |
| Laurent  2025 | PMA: GCaMP6f WT 4M/4F, KO 4M/4F; mCherry  WT 6M/6F;  KO 5M/5F; eNpHR3.0 WT 4M/4F, KO 5M/4F; ChR2  WT 6M/5F,  KO 4M/6F | 106 – 141 days on Ext. Day 2 | MS | SAPAP3 Null Mice, Platform mediated avoidance, fiber photometry | All data pooled for male and female mice after assessing for sex differences. Assumption of no significant differences between M and F SAPAP3-Null mice in intercalated nuclei of amygdala hyperexcitability interfering with platform-mediated avoidance behavior. No statistics reported. |  |

Abbreviations for Table A7: T-M=T-Maze, MBur=Marble Burying, EPM=Elevated Plus Maze, OF=Open Field, DDT=Delay Discounting Task, PPI=Prepulse Inhibition, FS= Forced Swim, SpA=Spontaneous Alternation, ConAvo=Conditioned Avoidance, FCon=Fear Conditioning, FExt=Fear Extinction, TST=Tale suspension Test, NOR=Novel Object Recognition

**References:**

1.	Gooding, D.C. and M.J. Pflum, *Further validation of the ACIPS as a measure of social hedonic response.* Psychiatry research, 2014. **215**(3): p. 771-777.

2.	Brown, T.A. and D.H. Barlow, *Anxiety and related disorders interview schedule for DSM-5 (ADIS-5)-adult and lifetime version: Clinician manual*. 2014: Oxford University Press.

3.	A. McGahuey, A.J.G., Cindi A. Laukes, Francisco A. Moreno, Pedro L. Delgado, Kathy M. McKnight, Rachel Manber, Cynthia, *The Arizona sexual experience scale (ASEX): reliability and validity.* Journal of Sex &Marital Therapy, 2000. **26**(1): p. 25-40.

4.	Abdel-Khalek, A.M., *The construction and validation of the revised Arabic Scale of Obsession-Compulsion (ASOC).* Online Journal of Neurology and Brain Disorders, 2018. **1**(2): p. 1-7.

5.	Sperling, M.B. and W.H. Berman, *Attachment in adults: Clinical and developmental perspectives*. 1994: Guilford Press.

6.	Stanford, M.S., et al., *Fifty years of the Barratt Impulsiveness Scale: An update and review.* Personality and individual differences, 2009. **47**(5): p. 385-395.

7.	Kubany, E.S., et al., *Development and preliminary validation of a brief broad-spectrum measure of trauma exposure: the Traumatic Life Events Questionnaire.* Psychological assessment, 2000. **12**(2): p. 210.

8.	Beck, A.T., et al., *An inventory for measuring clinical anxiety: psychometric properties.* Journal of consulting and clinical psychology, 1988. **56**(6): p. 893.

9.	Sanavio, E., *Obsessions and compulsions: the Padua Inventory.* Behaviour research and therapy, 1988. **26**(2): p. 169-177.

10.	Barratt, E.S., *Impulsiveness and aggression.* 1994.

11.	Buss, A.H. and M. Perry, *The aggression questionnaire.* Journal of personality and social psychology, 1992. **63**(3): p. 452.

12.	Lebowitz, E.R., et al., *Family accommodation in obsessive–compulsive disorder.* Expert review of neurotherapeutics, 2012. **12**(2): p. 229-238.

13.	Achenbach, T.M. and L. Rescorla, *Child behavior checklist for ages 6-18*. 2001: University of Vermont Burlington, VT.

14.	Achenbach, T.M. and C. Edelbrock, *Child behavior checklist.* Burlington (vt), 1991. **7**: p. 371-392.

15.	Guy, W., *Clinical global impression. ECDEU assessment manual for psychopharmacology.* Rockville: US Dept. of Health, Education, and Welfare, 1976.

16.	Piacentini, J., et al., *BRIEF REPORT: Functional impairment in childhood OCD: Development and psychometrics properties of the child obsessive-compulsive impact scale-revised (COIS-R).* Journal of Clinical Child and Adolescent Psychology, 2007. **36**(4): p. 645-653.

17.	Scahill, L., et al., *Children's Yale-Brown obsessive compulsive scale: reliability and validity.* Journal of the American Academy of Child & Adolescent Psychiatry, 1997. **36**(6): p. 844-852.

18.	Storch, E.A., et al., *Development and psychometric evaluation of the children’s Yale-Brown obsessive-compulsive scale second edition.* Journal of the American Academy of Child & Adolescent Psychiatry, 2019. **58**(1): p. 92-98.

19.	Helweg-Larsen, K., *The Danish register of causes of death.* Scandinavian journal of public health, 2011. **39**(7_suppl): p. 26-29.

20.	Mors, O., G.P. Perto, and P.B. Mortensen, *The Danish psychiatric central research register.* Scandinavian journal of public health, 2011. **39**(7_suppl): p. 54-57.

21.	Andersen, T.F., et al., *The Danish National Hospital Register. A valuable source of data for modern health sciences.* Danish medical bulletin, 1999. **46**(3): p. 263-268.

22.	Henry, J.D. and J.R. Crawford, *The short‐form version of the Depression Anxiety Stress Scales (DASS‐21): Construct validity and normative data in a large non‐clinical sample.* British journal of clinical psychology, 2005. **44**(2): p. 227-239.

23.	Nurnberger, J.I., et al., *Diagnostic interview for genetic studies: rationale, unique features, and training.* Archives of general psychiatry, 1994. **51**(11): p. 849-859.

24.	De Jong-Gierveld, J. and F. Kamphuls, *The development of a Rasch-type loneliness scale.* Applied psychological measurement, 1985. **9**(3): p. 289-299.

25.	Abramowitz, J.S., et al., *Assessment of obsessive-compulsive symptom dimensions: development and evaluation of the Dimensional Obsessive-Compulsive Scale.* Psychological assessment, 2010. **22**(1): p. 180.

26.	Fergus, T.A. and D.P. Valentiner, *The Disgust Propensity and Sensitivity Scale–Revised: An examination of a reduced-item version.* Journal of anxiety disorders, 2009. **23**(5): p. 703-710.

27.	Bell, C.C., *DSM-IV: diagnostic and statistical manual of mental disorders.* Jama, 1994. **272**(10): p. 828-829.

28.	Piqueras, J.A., M. Garcia-Olcina, and P. Cuijpers, *DetectaWeb-Distress Scale: A global and multidimensional web-based screener for emotional disorder symptoms in children and adolescents.* Frontiers in Psychology, 2021. **12**: p. 627604.

29.	Rosario-Campos, M., et al., *The Dimensional Yale–Brown Obsessive–Compulsive Scale (DYBOCS): an instrument for assessing obsessive–compulsive symptom dimensions.* Molecular psychiatry, 2006. **11**(5): p. 495-504.

30.	Cox, J.L., J.M. Holden, and R. Sagovsky, *Detection of postnatal depression: development of the 10-item Edinburgh Postnatal Depression Scale.* The British journal of psychiatry, 1987. **150**(6): p. 782-786.

31.	Boterhoven de Haan, K.L., et al., *Reliability and validity of a short version of the general functioning subscale of the McMaster Family Assessment Device.* Family process, 2015. **54**(1): p. 116-123.

32.	Calvocoressi, L., et al., *Family accommodation in obsessive-compulsive disorder.* The American Journal of Psychiatry, 1995. **152**(3): p. 441-443.

33.	Wu, M., *FAMILY ACCOMMODATION SCALE FOR OBSESSIVE-COMPULSIVE DISORDER.*

34.	Calvocoressi, L., et al., *Family accommodation of obsessive-compulsive symptoms: instrument development and assessment of family behavior.* The Journal of Nervous and Mental Disease, 1999. **187**(10): p. 636-642.

35.	Evans, D.E. and M.K. Rothbart, *Developing a model for adult temperament.* Journal of research in personality, 2007. **41**(4): p. 868-888.

36.	Hamilton, M., *The assessment of anxiety states by rating.* British journal of medical psychology, 1959.

37.	Hamilton, M., *A rating scale for depression.* Journal of neurology, neurosurgery, and psychiatry, 1960. **23**(1): p. 56.

38.	Habarth, J.M., *Development of the heteronormative attitudes and beliefs scale.* Psychology & Sexuality, 2015. **6**(2): p. 166-188.

39.	Rosen, R.C., et al., *The international index of erectile function (IIEF): a multidimensional scale for assessment of erectile dysfunction.* Urology, 1997. **49**(6): p. 822-830.

40.	Keller, M.B., et al., *The longitudinal interval follow-up evaluation: A comprehensive method for assessing outcome in prospective longitudinal studies.* Archives of general psychiatry, 1987. **44**(6): p. 540-548.

41.	Montgomery, S.A. and M. Åsberg, *A new depression scale designed to be sensitive to change.* The British journal of psychiatry, 1979. **134**(4): p. 382-389.

42.	Sheehan, D.V., et al., *The Mini-International Neuropsychiatric Interview (MINI): the development and validation of a structured diagnostic psychiatric interview for DSM-IV and ICD-10.* Journal of clinical psychiatry, 1998. **59**(20): p. 22-33.

43.	Butcher, J.N., et al., *Development and use of the MMPI-2 Content Scales*. 1990: University of Minnesota Press.

44.	Hodgson, R.J. and S. Rachman, *Obsessional-compulsive complaints.* Behaviour research and therapy, 1977. **15**(5): p. 389-395.

45.	Ware, J.E., *SF-36 health survey. Manual and interpretation guide.* The health institute, 1993: p. 6: 1-6: 22.

46.	Coles, M.E., et al., *“Not just right experiences”: perfectionism, obsessive–compulsive features and general psychopathology.* Behaviour research and therapy, 2003. **41**(6): p. 681-700.

47.	Dorz, S., et al., *Presentazione della versione italiana dell’Obsessive Beliefs Questionnaire (OBQ): struttura fattoriale e analisi di attendibilità (parte I).* Psicoterapia cognitiva e comportamentale, 2009. **15**(2): p. 139-70.

48.	Coles, M.E., et al., *Development and initial validation of the obsessive belief questionnaire‐child version (OBQ‐CV).* Depression and Anxiety, 2010. **27**(10): p. 982-991.

49.	Group, O.C.C.W., *Development and initial validation of the obsessive beliefs questionnaire and the interpretation of intrusions inventory.* Behaviour Research and Therapy, 2001. **39**(8): p. 987-1006.

50.	Melli, G., et al., *Assessing Core Affective Dimensions in OCD: Development and Validation of the Obsessive–Compulsive Core Dimensions Scale (OC‐CDS).* Clinical Psychology & Psychotherapy, 2025. **32**(4): p. e70113.

51.	Abramovitch, A., J.S. Abramowitz, and D. McKay, *The OCI-4: An ultra-brief screening scale for obsessive-compulsive disorder.* Journal of anxiety disorders, 2021. **78**: p. 102354.

52.	Foa, E.B., et al., *Development and validation of a child version of the obsessive compulsive inventory.* Behavior therapy, 2010. **41**(1): p. 121-132.

53.	Foa, E.B., et al., *The Obsessive-Compulsive Inventory: development and validation of a short version.* Psychological assessment, 2002. **14**(4): p. 485.

54.	Hills, P. and M. Argyle, *The Oxford Happiness Questionnaire: a compact scale for the measurement of psychological well-being.* Personality and individual differences, 2002. **33**(7): p. 1073-1082.

55.	Burns, G.L., et al., *Revision of the Padua Inventory of obsessive compulsive disorder symptoms: Distinctions between worry, obsessions, and compulsions.* Behaviour research and therapy, 1996. **34**(2): p. 163-173.

56.	Spence, S.H., *Structure of anxiety symptoms among children: a confirmatory factor-analytic study.* Journal of abnormal psychology, 1997. **106**(2): p. 280.

57.	Allison, K.C., et al., *Development of a brief measure of postpartum distress.* Journal of Women's Health, 2011. **20**(4): p. 617-623.

58.	Abramowitz, J.S., et al., *Religious obsessions and compulsions in a non-clinical sample: The Penn Inventory of Scrupulosity (PIOS).* Behaviour research and therapy, 2002. **40**(7): p. 825-838.

59.	Lord, C., et al., *Piloting the perinatal obsessive-compulsive scale (POCS): development and validation.* Journal of anxiety disorders, 2011. **25**(8): p. 1079-1084.

60.	Doron, G., et al., *Flaws and all: Exploring partner-focused obsessive-compulsive symptoms.* Journal of Obsessive-Compulsive and Related Disorders, 2012. **1**(4): p. 234-243.

61.	Corrigan, P. and J.D. Calabrese, *Toolkit.* APA PsycNet website. XX <https://psycnet>. apa. org/doiLanding, 2005.

62.	Usami, M., et al., *The reliability and validity of the Questionnaire-Children with Difficulties (QCD).* Child and adolescent psychiatry and mental health, 2013. **7**(1): p. 11.

63.	Pascual-Vera, B., et al., *The Mediating Role of Misinterpretations and Neutralizing Responses to Unwanted Intrusive Thoughts in Obsessive-Compulsive Spectrum Disorders.* European Journal of Investigation in Health, Psychology and Education, 2025. **15**(7): p. 135.

64.	Chorpita, B.F., et al., *Assessment of symptoms of DSM-IV anxiety and depression in children: A revised child anxiety and depression scale.* Behaviour research and therapy, 2000. **38**(8): p. 835-855.

65.	Doron, G., D.S. Derby, and O. Szepsenwol, *Relationship obsessive compulsive disorder (ROCD): A conceptual framework.* Journal of Obsessive-Compulsive and Related Disorders, 2014. **3**(2): p. 169-180.

66.	Purdon, C. and D.A. Clark, *Obsessive intrusive thoughts in nonclinical subjects. Part I. Content and relation with depressive, anxious and obsessional symptoms.* Behaviour research and therapy, 1993. **31**(8): p. 713-720.

67.	Weissman, M.M., et al., *Social adjustment by self-report in a community sample and in psychiatric outpatients.* The Journal of Nervous and Mental Disease, 1978. **166**(5): p. 317-326.

68.	Sanchez-Villegas, A., et al., *Validity of a self-reported diagnosis of depression among participants in a cohort study using the Structured Clinical Interview for DSM-IV (SCID-I).* BMC psychiatry, 2008. **8**: p. 1-8.

69.	Derogatis, L.R., M. Fitzpatrick, and M. Maruish, *The SCL-90-R, the brief symptom inventory (BSI), and the BSI-18.* The use of psychological testing for treatment planning and outcomes assessment: Instruments for adults, 2004. **3**: p. 1-41.

70.	Sheehan, D.V., M. Giddens, and P. Lisensi, *Sheehan disability scale (SDS).* International Clinical Psychopharmacology, 1983. **11**: p. 89-95.

71.	Hatch, R.L., et al., *The spiritual involvement and beliefs scale.* Journal of family Practice, 1998. **46**(6): p. 476-486.

72.	Janssen, E., et al., *The sexual inhibition/sexual excitation scales (SIS/SES): Validity, reliability, and scoring.* 2010.

73.	Uher, R., et al., *Screening young people for obsessive-compulsive disorder.* The British Journal of Psychiatry, 2007. **191**(4): p. 353-354.

74.	Williams, M.T., et al., *Assessing sexual orientation symptoms in obsessive-compulsive disorder: Development and validation of the Sexual Orientation Obsessions and Reactions Test (SORT).* Behavior Therapy, 2018. **49**(5): p. 715-729.

75.	Angst, J., A. Dobler-Mikola, and J. Binder, *The Zurich study—A prospective epidemiological study of depressive, neurotic and psychosomatic syndromes: I. Problem, methodology.* European Archives of Psychiatry and Neurological Sciences, 1984. **234**(1): p. 13-20.

76.	Park, L.S., et al., *The Toronto obsessive-compulsive scale: psychometrics of a dimensional measure of obsessive-compulsive traits.* Journal of the American Academy of Child & Adolescent Psychiatry, 2016. **55**(4): p. 310-318. e4.

77.	Sampaio, A.S., et al., *Validation of the University of São Paulo’s sensory phenomena scale—English version.* Comprehensive psychiatry, 2014. **55**(5): p. 1330-1336.

78.	Thordarson, D.S., et al., *The Vancouver obsessional compulsive inventory (VOCI).* Behaviour research and therapy, 2004. **42**(11): p. 1289-1314.

79.	Group, W., *Development of the World Health Organization WHOQOL-BREF quality of life assessment.* Psychological medicine, 1998. **28**(3): p. 551-558.

80.	Makkar, J.K., et al., *Cross Cultural Adaptation and Validation of Hindi Version of WHOQOL-BREF in Patients With Chronic Low Back Pain.* 2022.

81.	Mundt, J.C., et al., *The Work and Social Adjustment Scale: a simple measure of impairment in functioning.* The British Journal of Psychiatry, 2002. **180**(5): p. 461-464.

82.	Goodman, W.K., et al., *The yale-brown obsessive compulsive scale: II. Validity.* Archives of general psychiatry, 1989. **46**(11): p. 1012-1016.

83.	Goodman, W.K., et al., *The Yale-Brown obsessive compulsive scale: I. Development, use, and reliability.* Archives of general psychiatry, 1989. **46**(11): p. 1006-1011.

84.	Storch, E.A., et al., *Development and psychometric evaluation of the Yale–Brown obsessive-compulsive scale—second edition.* Psychological assessment, 2010. **22**(2): p. 223.

85.	Thomas, A., et al., *Manual for the Youth Self-Report and 1991 profile.* Burlington, VT: University of Vermont, Department of Psychiatry, 1991.

86.	Koss, M. and J. Marks, *Relationship of childhood abuse and household dysfunction to many of the leading causes of death in adults: The adverse childhood experiences (ACE) study.* American journal of preventative medicine, 1998. **14**(4): p. 245-258.

87.	Pace, C.S., et al., *The Adverse Childhood Experiences–International Questionnaire (ACE-IQ) in community samples around the world: A systematic review (part I).* Child Abuse & Neglect, 2022. **129**: p. 105640.

88.	Spielberger, C.D., *State‐Trait anger expression inventory.* The Corsini encyclopedia of psychology, 2010: p. 1-1.

89.	Beck, A., et al., *An Inventory for Measuring Depression. Archives of General Psychiatry, Vol. 4.* 1961.

90.	Beck, A.T., R.A. Steer, and G.K. Brown, *Beck depression inventory (BDI-II)*. Vol. 10. 1996: Pearson London, UK.

91.	Association, A.P., *Diagnostic and statistical manual of mental disorders.* Text revision, 2000.

92.	Pallesen, S., et al., *A new scale for measuring insomnia: the Bergen Insomnia Scale.* Perceptual and motor skills, 2008. **107**(3): p. 691-706.

93.	Snell Jr, W.E., et al., *The clinical anger scale: Preliminary reliability and validity.* Journal of clinical psychology, 1995. **51**(2): p. 215-226.

94.	Bordin, I.A., et al., *Child Behavior Checklist (CBCL), Youth Self-Report (YSR) and Teacher's Report Form (TRF): an overview of the development of the original and Brazilian versions.* Cadernos de saúde pública, 2013. **29**: p. 13-28.

95.	Kovacs, M., *Manual of the Children‘s Depression Inventory. 1992.* New York: Multi-Health Systems.

96.	Shaffer, D., et al., *A children's global assessment scale (CGAS).* Archives of General psychiatry, 1983. **40**(11): p. 1228-1231.

97.	Andrews, G. and L. Peters, *The psychometric properties of the composite international diagnostic interview.* Social psychiatry and psychiatric epidemiology, 1998. **33**: p. 80-88.

98.	Wittchen, H.U., et al., *Measuring symptoms and diagnosing mental disorders in the elderly community: the test–retest reliability of the CIDI65+.* International Journal of Methods in Psychiatric Research, 2015. **24**(2): p. 116-129.

99.	Wittchen, H.U., et al., *Screening for mental disorders: performance of the Composite International Diagnostic–Screener (CID–S).* International Journal of Methods in Psychiatric Research, 1999. **8**(2): p. 59-70.

100.	Stoddard, J. and J. Kaufman, *Coronavirus impact scale.* Johns Hopkins School of Medicine, 2020. **2**.

101.	Lewis, G. and A. Pelosi, *Manual of the revised clinical interview schedule (CIS-R).* London: Institute of Psychiatry, 1990.

102.	Carver, C.S., M.F. Scheier, and J.K. Weintraub, *Assessing coping strategies: a theoretically based approach.* Journal of personality and social psychology, 1989. **56**(2): p. 267.

103.	EA, H. *A standard stimulus for measuring vasomotor reactions: its application in the study of hypertension*. in *Mayo Clin Proc*. 1932.

104.	Lovallo, W., *The cold pressor test and autonomic function: a review and integration.* Psychophysiology, 1975. **12**(3): p. 268-282.

105.	Gadow, K. and J. Sprafkin, *Child Symptom Inventory-4: Teacher Checklist.* Stony Brook, NY: Checkmate Plus, 1998.

106.	Schwab-Stone, M., et al., *Reliability of diagnostic reporting for children aged 6-11 years: a test-retest study of the Diagnostic Interview Schedule for Children-Revised.* The American journal of psychiatry, 1994. **151**(7): p. 1048-1054.

107.	Wittchen, H.-U., et al., *Test-retest reliability of the computerized DSM-IV version of the Munich-Composite International Diagnostic Interview (M-CIDI).* Social psychiatry and psychiatric epidemiology, 1998. **33**: p. 568-578.

108.	Harvey, P.D., *Domains of cognition and their assessment.* Dialogues in clinical neuroscience, 2022.

109.	Group, T.E., *EuroQol-a new facility for the measurement of health-related quality of life.* Health policy, 1990. **16**(3): p. 199-208.

110.	Fei, L., et al., *Preliminary evaluation of Chinese version of FACES II and FES: comparison of normal families and families of schizophrenic patients.* Chinese Mental Health Journal, 1991. **5**(5): p. 198-202.

111.	Andreasen, N.C., et al., *The family history method using diagnostic criteria: reliability and validity.* Archives of general psychiatry, 1977. **34**(10): p. 1229-1235.

112.	Weissman, M.M., et al., *Brief screening for family psychiatric history: the family history screen.* Archives of general psychiatry, 2000. **57**(7): p. 675-682.

113.	Shirzadi, M., H. Khazaie, and S. Farhang, *Mental health survey of adult population in Kermanshah County, 2015: Preliminary report.* Asian journal of psychiatry, 2017. **29**: p. 103-105.

114.	Calkins, M.E., et al., *The Philadelphia Neurodevelopmental Cohort: constructing a deep phenotyping collaborative.* Journal of Child Psychology and Psychiatry, 2015. **56**(12): p. 1356-1369.

115.	Rust, J., et al., *The golombok rust inventory of marital state (GRIMS).* Sexual and Marital Therapy, 1986. **1**(1): p. 55-60.

116.	Organization, W.H., *Mental and behavioral disorders: diagnostic criteria for research.* International Statistical Classification of Diseases, 10th Revision (ICD-10), 1992.

117.	Watson, D., et al., *Development and validation of new anxiety and bipolar symptom scales for an expanded version of the IDAS (the IDAS-II).* Assessment, 2012. **19**(4): p. 399-420.

118.	Bates, J.E., C.A.B. Freeland, and M.L. Lounsbury, *Measurement of infant difficultness.* Child development, 1979: p. 794-803.

119.	Werner, P.D., *Implications of attitude-behavior studies for population research and action.* Studies in Family Planning, 1977. **8**(11): p. 294-299.

120.	Lee, T., et al., *Development of North Korean version of the composite international diagnostic interview.* Journal of Korean Neuropsychiatric Association, 2015. **54**(2): p. 228-235.

121.	Kaufman, J., et al., *Schedule for affective disorders and schizophrenia for school-age children-present and lifetime version (K-SADS-PL): initial reliability and validity data.* Journal of the American Academy of Child & Adolescent Psychiatry, 1997. **36**(7): p. 980-988.

122.	Brugha, T.S. and D. Cragg, *The list of threatening experiences: the reliability and validity of a brief life events questionnaire.* Acta Psychiatrica Scandinavica, 1990. **82**(1): p. 77-81.

123.	McHugo, G.J., et al., *The assessment of trauma history in women with co-occurring substance abuse and mental disorders and a history of interpersonal violence.* The journal of behavioral health services & research, 2005. **32**: p. 113-127.

124.	Duncan, L., et al., *Psychometric evaluation of the Mini International Neuropsychiatric Interview for Children and Adolescents (MINI-KID).* Psychological Assessment, 2018. **30**(7): p. 916.

125.	Belza, B., et al., *A systematic review of studies using the multidimensional assessment of fatigue scale.* Journal of Nursing Measurement, 2018. **26**(1): p. 36-74.

126.	Organization, W.H., *The ICD-10 classification of mental and behavioural disorders: clinical descriptions and diagnostic guidelines*. Vol. 1. 1992: World Health Organization.

127.	Hoyer, J., et al., *Test-retest reliability of the computer-assisted DIA-X-5 interview for mental disorders.* BMC psychiatry, 2020. **20**: p. 1-16.

128.	Elliott, C.M. and A.S. Radomsky, *Analyses of mental contamination: Part I, experimental manipulations of morality.* Behaviour Research and Therapy, 2009. **47**(12): p. 995-1003.

129.	Miller, C.J., et al., *Sensitivity and specificity of the Mood Disorder Questionnaire for detecting bipolar disorder.* Journal of affective disorders, 2004. **81**(2): p. 167-171.

130.	Dedovic, K., et al., *The Montreal Imaging Stress Task: using functional imaging to investigate the effects of perceiving and processing psychosocial stress in the human brain.* Journal of Psychiatry and Neuroscience, 2005. **30**(5): p. 319-325.

131.	Pederson, F., et al., *Construction of self and offspring in the pregnancy and early infancy period.* Society for Research in Child Development, 1989.

132.	Rachman, S., et al., *Mental contamination: The perpetrator effect.* Journal of Behavior Therapy and Experimental Psychiatry, 2012. **43**(1): p. 587-593.

133.	Armour, C., et al., *Factor structure of the PTSD checklist for DSM-5: Relationships among symptom clusters, anger, and impulsivity.* The Journal of nervous and mental disease, 2016. **204**(2): p. 108-115.

134.	Kroenke, K., R.L. Spitzer, and J.B. Williams, *The PHQ‐9: validity of a brief depression severity measure.* Journal of general internal medicine, 2001. **16**(9): p. 606-613.

135.	Rubio-Aparicio, M., et al., *The Padua Inventory–Washington State University Revision of obsessions and compulsions: A reliability generalization meta-analysis.* Journal of personality assessment, 2020. **102**(1): p. 113-123.

136.	Fairbrother, N. and S.R. Woody, *Fear of childbirth and obstetrical events as predictors of postnatal symptoms of depression and post-traumatic stress disorder.* Journal of Psychosomatic Obstetrics & Gynecology, 2007. **28**(4): p. 239-242.

137.	Bayrampour, H., et al., *Pregnancy-related anxiety: A concept analysis.* International journal of nursing studies, 2016. **55**: p. 115-130.

138.	Buysse, D.J., et al., *The Pittsburgh Sleep Quality Index: a new instrument for psychiatric practice and research.* Psychiatry research, 1989. **28**(2): p. 193-213.

139.	Marshall, N.L. and R.C. Barnett, *Work‐family strains and gains among two‐earner couples.* Journal of Community Psychology, 1993. **21**(1): p. 64-78.

140.	Abramowitz, J.S., S.A. Schwartz, and K.M. Moore, *Obsessional thoughts in postpartum females and their partners: content, severity, and relationship with depression.* Journal of Clinical Psychology in Medical Settings, 2003. **10**: p. 157-164.

141.	Salkovskis, P.M., et al., *Responsibility attitudes and interpretations are characteristic of obsessive compulsive disorder.* Behaviour research and therapy, 2000. **38**(4): p. 347-372.

142.	von Soest, T., et al., *A validation study of the resilience scale for adolescents (READ).* Journal of Psychopathology and Behavioral assessment, 2010. **32**: p. 215-225.

143.	Miller, F. and L. Lazowski, *The adult SASSI-3 manual.* Springville, IN: SASSI Institute, 1999.

144.	Birmaher, B., et al., *The screen for child anxiety related emotional disorders (SCARED): Scale construction and psychometric characteristics.* Journal of the American Academy of Child & Adolescent Psychiatry, 1997. **36**(4): p. 545-553.

145.	Osório, F.L., et al., *Clinical validity and intrarater and test–retest reliability of the Structured Clinical Interview for DSM‐5–Clinician Version (SCID‐5‐CV).* Psychiatry and clinical neurosciences, 2019. **73**(12): p. 754-760.

146.	Ventura, J., et al., *Training and quality assurance with the Structured Clinical Interview for DSM-IV (SCID-I/P).* Psychiatry research, 1998. **79**(2): p. 163-173.

147.	Gutiérrez, F., et al., *Measuring the core features of personality disorders in substance abusers using the Temperament and Character Inventory (TCI).* Journal of personality disorders, 2002. **16**(4): p. 344-359.

148.	Derogatis, L.R., R.S. Lipman, and L. Covi, *SCL-90: an outpatient psychiatric rating scale–preliminary report.* Psychopharmacol bull, 1973. **9**(1): p. 13-28.

149.	Derogatis, L.R., *SCL-90-R: Symptom Checklist-90-R: Administration, scoring, and procedures manual*. 1975: NCS Pearson.

150.	Derogatis, L.R., et al., *The Hopkins Symptom Checklist (HSCL): A self‐report symptom inventory.* Behavioral science, 1974. **19**(1): p. 1-15.

151.	Jacobsson, L., et al., *Perceived parental behaviour and psychogenic needs.* Archiv für Psychiatrie und Nervenkrankheiten, 1980. **228**: p. 21-30.

152.	Ware, J.E., M. Kosinski, and S.D. Keller, *A 12-Item Short-Form Health Survey: construction of scales and preliminary tests of reliability and validity.* Medical care, 1996. **34**(3): p. 220-233.

153.	Spielberger, C.D., et al., *The state-trait anxiety inventory.* Revista Interamericana de Psicologia/Interamerican journal of psychology, 1971. **5**(3 & 4).

154.	Dembińska-Krajewska, D. and J. Rybakowski, *The Temperament Evaluation of Memphis, Pisa and San Diego Autoquestionnaire (TEMPS-A)--an important tool to study affective temperaments.* Psychiatr Pol, 2014. **48**(2): p. 261-76.

155.	Whooley, M.A., et al., *Case‐finding instruments for depression: Two questions are as good as many.* Journal of general internal medicine, 1997. **12**(7): p. 439-445.

156.	Gadow, K. and J. Sprafkin, *Youth’s inventory-4 manual.* Stony Brook, NY: Checkmate Plus, 1999.

157.	Leckman, J., *Yale OCD natural history questionnaire.* Unpublished manuscript, 2002.

158.	Dempster, E.L., et al., *Genome-wide methylomic analysis of monozygotic twins discordant for adolescent depression.* Biological psychiatry, 2014. **76**(12): p. 977-983.

159.	Klein, R., *The first wave of NHS foundation trusts*. 2004, British Medical Journal Publishing Group. p. 1332.

160.	Silverman, W., *Anxiety disorders interview schedule for children (parent version).* Albany, NY Greywind, 1991.

161.	Wolfling, K., M. Beutel, and K. Muller, *OSV-S-Skala zum Onlinesuchtverhalten [AICA-S: scale for the assessment of Internet and computer game addiction].* Diagnostische Verfahren in der Psychotherapie (Diagnostik fur Klinik und Praxis)[Diagnostic measures for psychotherapy], 2016: p. 362-366.

162.	Dawson, D.A., et al., *Effectiveness of the derived Alcohol Use Disorders Identification Test (AUDIT‐C) in screening for alcohol use disorders and risk drinking in the US general population.* Alcoholism: Clinical and Experimental Research, 2005. **29**(5): p. 844-854.

163.	Eisen, J.L., et al., *The brown assessment of beliefs scale: reliability and validity.* American Journal of Psychiatry, 1998. **155**(1): p. 102-108.

164.	Beck, A.T., et al., *Beck anxiety inventory.* Journal of consulting and clinical psychology, 1993.

165.	Calugi, S., et al., *Validation of the Body Checking Questionnaire (BCQ) in an eating disorders population.* Behavioural and Cognitive Psychotherapy, 2006. **34**(2): p. 233-242.

166.	Beck, A.T., R.A. Steer, and G.K. Brown, *Beck depression inventory-II.* San Antonio, TX: Psychological Corporation, 1996: p. b9.

167.	Sandoz, E.K., et al., *Assessment of body image flexibility: The body image-acceptance and action questionnaire.* Journal of Contextual Behavioral Science, 2013. **2**(1-2): p. 39-48.

168.	Gomez, R., et al., *The Bergen–Social Media Addiction Scale (BSMAS): longitudinal measurement invariance across a two-year interval.* Clinical Psychologist, 2024. **28**(2): p. 185-194.

169.	Bienvenu, O.J., et al., *The relationship of obsessive–compulsive disorder to possible spectrum disorders: results from a family study.* Biological psychiatry, 2000. **48**(4): p. 287-293.

170.	Isquith, P.K. and G.A. Gioia, *BRIEF-A: Behavior Rating Inventory of Executive Function-Adult Version*. 2005: PAR Psychological Assessment Ressources.

171.	Andreassen, C.S., et al., *The Bergen Shopping Addiction Scale: Reliability and validity of a brief screening test.* Frontiers in psychology, 2015. **6**: p. 1374.

172.	Derogatis, L.R., *Brief symptom inventory: BSI*. 1993: Pearson.

173.	Beck, A.T., M. Kovacs, and A. Weissman, *Assessment of suicidal intention: the Scale for Suicide Ideation.* Journal of consulting and clinical psychology, 1979. **47**(2): p. 343.

174.	Valence, G., A. d'Astous, and L. Fortier, *Compulsive buying: Concept and measurement.* Journal of consumer policy, 1988. **11**(4): p. 419-433.

175.	Kovacs, M., *Children’s Depression Inventory.* CDI, Manual/Multi-Health Systems Inc, 1992.

176.	Endicott, J., et al., *The Global Assessment Scale: A procedure for measuring overall severity of psychiatric disturbance.* Archives of general psychiatry, 1976. **33**(6): p. 766-771.

177.	Bohn, K. and C.G. Fairburn, *The clinical impairment assessment questionnaire (CIA).* Cognitive behavioral therapy for eating disorders, 2008: p. 315-317.

178.	Storch, E.A., et al., *The children's Yale-Brown obsessive–compulsive scale: Psychometric properties of child-and parent-report formats.* Journal of anxiety disorders, 2006. **20**(8): p. 1055-1070.

179.	Proudfoot, H., et al., *Development of a short cannabis problems questionnaire for adolescents in the community.* Addictive Behaviors, 2010. **35**(7): p. 734-737.

180.	Posner, K., et al., *Columbia-suicide severity rating scale (C-SSRS).* New York, NY: Columbia University Medical Center, 2008. **10**: p. 2008.

181.	Bernstein, D.P., et al., *Initial reliability and validity of a new retrospective measure of child abuse and neglect.* The American journal of psychiatry, 1994. **151**(8): p. 1132-1136.

182.	Goodman, R., et al., *The development and well‐being assessment: Description and initial validation of an integrated assessment of child and adolescent psychopathology.* Journal of child psychology and psychiatry, 2000. **41**(5): p. 645-655.

183.	Van Strien, T., et al., *The Dutch Eating Behavior Questionnaire (DEBQ) for assessment of restrained, emotional, and external eating behavior.* International journal of eating disorders, 1986. **5**(2): p. 295-315.

184.	Wilson, S., *The role of doubt in bulimia nervosa.* 2018.

185.	Stice, E., M. Fisher, and E. Martinez, *Eating disorder diagnostic scale: additional evidence of reliability and validity.* Psychological assessment, 2004. **16**(1): p. 60.

186.	Fairburn, C.G. and S.J. Beglin, *Eating disorder examination questionnaire.* Cognitive behavior therapy and eating disorders, 2008. **309**: p. 313.

187.	Garner, D.M., M.P. Olmstead, and J. Polivy, *Development and validation of a multidimensional eating disorder inventory for anorexia nervosa and bulimia.* International journal of eating disorders, 1983. **2**(2): p. 15-34.

188.	Gmel, G., et al., *Das European School Survey Project on Alcohol and Other Drugs (ESPAD) in der Schweiz: Wichtigste Ergebnisse im Vergleich 2003 und 2007*. 2009, Institut suisse de prévention de l'alcoolisme et autres toxicomanies.

189.	Frost, R.O. and P.A. Marten, *Perfectionism and evaluative threat.* Cognitive therapy and research, 1990. **14**: p. 559-572.

190.	Storch, E.A., et al., *Florida obsessive‐compulsive inventory: Development, reliability, and validity.* Journal of clinical psychology, 2007. **63**(9): p. 851-859.

191.	Stunkard, A., et al., *Use of theDanish Adoption Register for the study of obesity and thinness*. 1983, Raven Press New York. p. 115-120.

192.	Rosen, C.B., J. Heiman, S. Leiblum, C. Meston, R. Shabsigh, D. Ferguson, R. D'Agostino, R, *The Female Sexual Function Index (FSFI): a multidimensional self-report instrument for the assessment of female sexual function.* Journal of sex & marital therapy, 2000. **26**(2): p. 191-208.

193.	Lemmens, J.S., P.M. Valkenburg, and J. Peter, *Development and validation of a game addiction scale for adolescents.* Media psychology, 2009. **12**(1): p. 77-95.

194.	Snaith, R.P., *The hospital anxiety and depression scale.* Health and quality of life outcomes, 2003. **1**: p. 1-4.

195.	Tolin, D.F., R.O. Frost, and G. Steketee, *A brief interview for assessing compulsive hoarding: the Hoarding Rating Scale-Interview.* Psychiatry research, 2010. **178**(1): p. 147-152.

196.	Tolin, D.F., et al., *Hoarding Rating Scale--Self-Report.* Cognitive Therapy and Research, 2010.

197.	Young, K.S., *Internet addiction test.* Center for on-line addictions, 2009.

198.	Berg, C.J., J.L. Rapoport, and M. Flament, *The Leyton obsessional inventory-child version.* Journal of the American Academy of Child Psychiatry, 1986. **25**(1): p. 84-91.

199.	March, J.S., et al., *The Multidimensional Anxiety Scale for Children (MASC): factor structure, reliability, and validity.* Journal of the American academy of child & adolescent psychiatry, 1997. **36**(4): p. 554-565.

200.	Sheehan, D.V., et al., *The Mini-International Neuropsychiatric Interview (MINI): the development and validation of a structured diagnostic psychiatric interview for DSM-IV and ICD-10.* Journal of clinical psychiatry, 1998. **59**: p. 22-33.

201.	Donnellan, M.B., et al., *Mini-International Personality Item Pool.* Psychological Assessment, 2006.

202.	Sternberger, L.G. and G.L. Burns, *Maudsley Obsessional-Compulsive Inventory: Obsessions and compulsions in a nonclinical sample.* Behaviour research and therapy, 1990. **28**(4): p. 337-340.

203.	Costa, P. and R. McCrae, *NEO five-factor inventory (NEO-FFI).* Odessa, FL: Psychological Assessment Resources, 1989. **3**.

204.	McCrae, R.R. and P.T. Costa Jr, *The NEO Personality Inventory: Using the five‐factor modeI in counseling.* Journal of Counseling & Development, 1991. **69**(4): p. 367-372.

205.	Guy, W., *Clinical global impressions scale.* Psychiatry, 1976.

206.	Parker, G., H. Tupling, and L.B. Brown, *A parental bonding instrument.* British journal of medical psychology, 1979.

207.	Holden, R.R., et al., *Development and preliminary validation of a scale of psychache.* Canadian Journal of Behavioural Science/Revue canadienne des sciences du comportement, 2001. **33**(4): p. 224.

208.	Chang, S., et al., *Initial psychometric properties of a brief parent-report instrument for assessing tic severity in children with chronic tic disorders.* Child & Family Behavior Therapy, 2009. **31**(3): p. 181-191.

209.	Woods, D.W., et al., *Premonitory Urge for Tics Scale (PUTS): initial psychometric results and examination of the premonitory urge phenomenon in youths with Tic disorders.* Journal of Developmental & Behavioral Pediatrics, 2005. **26**(6): p. 397-403.

210.	White, H.R. and E.W. Labouvie, *Towards the assessment of adolescent problem drinking.* Journal of studies on alcohol, 1989. **50**(1): p. 30-37.

211.	First, M., et al., *SCID-I. Structured Clinical Interview for DSM-IV-TR Axis I Disorders, Research Version*. 2014, Pracownia Testów Psychologicznych PTP Warszawa, Poland.

212.	Kirkpatrick, B., et al., *The schedule for the deficit syndrome: an instrument for research in schizophrenia.* Psychiatry research, 1989. **30**(2): p. 119-123.

213.	Mattick, R.P. and J.C. Clarke, *Development and validation of measures of social phobia scrutiny fear and social interaction anxiety.* Behaviour research and therapy, 1998. **36**(4): p. 455-470.

214.	Claes, L. and W. Vandereycken, *The Self-Injury Questionnaire—Treatment Related (SIQ-TR): Construction, reliability, and validity in a sample of female eating disorder patients.* 2007.

215.	Piqueras, J.A., et al., *Validation of the Short Obsessive–Compulsive Disorder Screener (SOCS) in children and adolescents.* BJPsych open, 2015. **1**(1): p. 21-26.

216.	Christo, G., et al., *The Shorter PROMIS Questionnaire: Further validation of a tool for simultaneous assessment of multiple addictive behaviours.* Addictive behaviors, 2003. **28**(2): p. 225-248.

217.	Peters, L., *Discriminant validity of the social phobia and anxiety inventory (SPAI), the social phobia scale (SPS) and the social interaction anxiety scale (SIAS).* Behaviour research and therapy, 2000. **38**(9): p. 943-950.

218.	Keuthen, N.J., et al., *The prevalence of pathologic skin picking in US adults.* Comprehensive psychiatry, 2010. **51**(2): p. 183-186.

219.	Larsson, H., et al., *Genetic and environmental influences on adult attention deficit hyperactivity disorder symptoms: a large Swedish population-based study of twins.* Psychological medicine, 2013. **43**(1): p. 197-207.

220.	Rosario-Campos, M., et al., *Protocolo de pesquisa do ambulatório de adultos do PROTOC.* São Paulo: Depto. de Psiquiatria da Faculdade de Medicina da Universidade Federal de São Paulo, 2000.

221.	Kessler, R.C. and T.B. Üstün, *The world mental health (WMH) survey initiative version of the world health organization (WHO) composite international diagnostic interview (CIDI).* International journal of methods in psychiatric research, 2004. **13**(2): p. 93-121.

222.	Young, K.S., *Internet addiction: The emergence of a new clinical disorder.* Cyberpsychology & behavior, 1998. **1**(3): p. 237-244.

223.	Kircanski, K., et al., *Cluster analysis of the Yale Global Tic Severity Scale (YGTSS): Symptom dimensions and clinical correlates in an outpatient youth sample.* Journal of abnormal child psychology, 2010. **38**: p. 777-788.

224.	Brener, N.D., et al., *Reliability of the Youth Risk Behavior Survey Questionnaire.* American Journal of Epidemiology, 1995. **141**(6): p. 575-580.

225.	Hsieh, C.-Y., et al., *Taiwan’s national health insurance research database: past and future.* Clinical epidemiology, 2019: p. 349-358.

226.	Hopper, J.L., et al., *Australian Twin Registry: 30 years of progress.* Twin Research and Human Genetics, 2013. **16**(1): p. 34-42.

227.	Anckarsäter, H., et al., *The child and adolescent twin study in Sweden (CATSS).* Twin Research and Human Genetics, 2011. **14**(6): p. 495-508.

228.	McDaid, D., et al., *Ireland: Health system review.* 2009.

229.	Pedersen, C.B., *The Danish civil registration system.* Scandinavian journal of public health, 2011. **39**(7_suppl): p. 22-25.

230.	Knudsen, L.B. and J. Olsen, *The Danish medical birth registry.* Danish medical bulletin, 1998. **45**(3): p. 320-323.

231.	Munk-Jørgensen, P. and P.B. Mortensen, *The Danish psychiatric central register.* Danish medical bulletin, 1997. **44**(1): p. 82-84.

232.	Sund, R., *Quality of the Finnish Hospital Discharge Register: a systematic review.* Scandinavian journal of public health, 2012. **40**(6): p. 505-515.

233.	McAdams, T.A., et al., *The genesis 12–19 (G1219) study: a twin and sibling study of gene–environment interplay and adolescent development in the UK.* Twin Research and Human Genetics, 2013. **16**(1): p. 134-143.

234.	Ekbom, A., *The Swedish multi-generation register.* Methods in biobanking, 2011: p. 215-220.

235.	Wettermark, B., et al., *The new Swedish Prescribed Drug Register—opportunities for pharmacoepidemiological research and experience from the first six months.* Pharmacoepidemiology and drug safety, 2007. **16**(7): p. 726-735.

236.	Waldenlind, K., et al., *Validation of the rheumatoid arthritis diagnosis in the Swedish National Patient Register: a cohort study from Stockholm County.* BMC musculoskeletal disorders, 2014. **15**: p. 1-6.

237.	Lichtenstein, P., et al., *The Swedish Twin Registry: a unique resource for clinical, epidemiological and genetic studies.* Journal of internal medicine, 2002. **252**(3): p. 184-205.

238.	Haworth, C.M., O.S. Davis, and R. Plomin, *Twins Early Development Study (TEDS): a genetically sensitive investigation of cognitive and behavioral development from childhood to young adulthood.* Twin Research and Human Genetics, 2013. **16**(1): p. 117-125.

239.	Ludvigsson, J.F., et al., *Registers of the Swedish total population and their use in medical research.* European journal of epidemiology, 2016. **31**: p. 125-136.

240.	Verdi, S., et al., *TwinsUK: the UK adult twin registry update.* Twin Research and Human Genetics, 2019. **22**(6): p. 523-529.

241.	Reed, Z.E., et al., *Mapping the genetic and environmental aetiology of autistic traits in Sweden and the United Kingdom.* JCPP advances, 2021. **1**(3): p. e12039.

242.	Goldberg, J., et al., *The Vietnam era twin registry.* Twin Research and Human Genetics, 2002. **5**(5): p. 476-481.

243.	Stuss, D., et al., *Traumatic brain injury: A comparison of three clinical tests, and analysis of recovery.* The Clinical Neuropsychologist, 1989. **3**(2): p. 145-156.

244.	Stuss, D., L. Stethem, and G. Pelchat, *Three tests of attention and rapid information processing: An extension.* The Clinical Neuropsychologist, 1988. **2**(3): p. 246-250.

245.	McCrae, R.R. and P.T. Costa Jr, *A contemplated revision of the NEO Five-Factor Inventory.* Personality and individual differences, 2004. **36**(3): p. 587-596.

246.	García-Soriano, G., V. Rosell-Clari, and M.Á. Serrano, *Emotional and cognitive variables associated with contamination-related obsessive-compulsive symptoms.* The Spanish journal of psychology, 2016. **19**: p. E25.

247.	Robbins, T.W., et al., *Cambridge Neuropsychological Test Automated Battery (CANTAB): a factor analytic study of a large sample of normal elderly volunteers.* Dementia and geriatric cognitive disorders, 1994. **5**(5): p. 266-281.

248.	Silverman, W.K., et al., *Childhood anxiety sensitivity index.* Journal of Clinical Child and Adolescent Psychology, 1991. **20**(2): p. 162-168.

249.	Gratz, K.L. and L. Roemer, *Multidimensional assessment of emotion regulation and dysregulation: Development, factor structure, and initial validation of the difficulties in emotion regulation scale.* Journal of psychopathology and behavioral assessment, 2004. **26**: p. 41-54.

250.	Simons, J.S. and R.M. Gaher, *The Distress Tolerance Scale: Development and validation of a self-report measure.* Motivation and emotion, 2005. **29**(2): p. 83-102.

251.	Tonarely, N.A. and J. Ehrenreich-May, *Confirming the factor structure and validity of the distress tolerance scale (DTS) in youth.* Child Psychiatry & Human Development, 2020. **51**(4): p. 514-526.

252.	Brennan, K.A., C.L. Clark, and P.R. Shaver, *Self-report measurement of adult attachment: An integrative overview.* Attachment theory and close relationships, 1998. **46**: p. 76.

253.	Fraley, R.C. and P.R. Shaver, *Adult romantic attachment: Theoretical developments, emerging controversies, and unanswered questions.* Review of general psychology, 2000. **4**(2): p. 132-154.

254.	Garner, D.M. and T. Van Strien, *EDI-3.* Eating disorder inventory-3: professional manual, 2004. **35**: p. 479.

255.	Vohs, K.D., R.F. Baumeister, and N.J. Ciarocco, *Self-regulation and self-presentation: regulatory resource depletion impairs impression management and effortful self-presentation depletes regulatory resources.* Journal of personality and social psychology, 2005. **88**(4): p. 632.

256.	Noone, P.A., *The holmes–rahe stress inventory.* Occupational medicine, 2017. **67**(7): p. 581-582.

257.	Lang, P.J., M.M. Bradley, and B.N. Cuthbert, *International affective picture system (IAPS): Technical manual and affective ratings.* NIMH Center for the Study of Emotion and Attention, 1997. **1**(39-58): p. 3.

258.	Bechara, A., *Iowa gambling task*. 2007: Psychological Assessment Resources.

259.	Dağ, İ., *Kontrol Odaği Ölçeği (KOÖ): Ölçek geliştirme, güvenirlik ve geçerlik çalişmasi.* Türk Psikoloji Dergisi, 2002.

260.	Wells, A. and S. Cartwright-Hatton, *A short form of the metacognitions questionnaire: properties of the MCQ-30.* Behaviour research and therapy, 2004. **42**(4): p. 385-396.

261.	Lecrubier, Y., et al., *The Mini International Neuropsychiatric Interview (MINI). A short diagnostic structured interview: reliability and validity according to the CIDI.* European psychiatry, 1997. **12**(5): p. 224-231.

262.	Sanavio, E. and G. Vidotto, *The components of the Maudsley obsessional-compulsive questionnaire.* Behaviour Research and Therapy, 1985. **23**(6): p. 659-662.

263.	Quinn, E.P., T.H. Brandon, and A.L. Copeland, *Is task persistence related to smoking and substance abuse? The application of learned industriousness theory to addictive behaviors.* Experimental and Clinical Psychopharmacology, 1996. **4**(2): p. 186.

264.	Costa Jr, P.T. and R.R. McCrae, *Four ways five factors are basic.* Personality and individual differences, 1992. **13**(6): p. 653-665.

265.	Derogatis, L.R. and R. Unger, *Symptom checklist‐90‐revised.* The Corsini encyclopedia of psychology, 2010: p. 1-2.

266.	Summerfeldt, L., et al. *Assessing and validating the obsessive-compulsive-related construct of incompleteness*. in *Poster presented at the 62nd annual convention of the Canadian Psychological Association, Ste-Foy, Quebec*. 2001.

267.	Rogers, R.D., et al., *Dissociable deficits in the decision-making cognition of chronic amphetamine abusers, opiate abusers, patients with focal damage to prefrontal cortex, and tryptophan-depleted normal volunteers: evidence for monoaminergic mechanisms.* Neuropsychopharmacology, 1999. **20**(4): p. 322-339.

268.	McEvoy, P.M., A.E. Mahoney, and M.L. Moulds, *Are worry, rumination, and post-event processing one and the same?: Development of the Repetitive Thinking Questionnaire.* Journal of anxiety disorders, 2010. **24**(5): p. 509-519.

269.	Watson, D. and K.D. Wu, *Development and validation of the Schedule of Compulsions, Obsessions, and Pathological Impulses (SCOPI).* Assessment, 2005. **12**(1): p. 50-65.

270.	Paykel, E.S., B.A. Prusoff, and E. Uhlenhuth, *Scaling of life events.* Archives of general psychiatry, 1971. **25**(4): p. 340-347.

271.	Heaton, R.K. and P. Staff, *Wisconsin card sorting test: computer version 2.* Odessa: Psychological Assessment Resources, 1993. **4**: p. 1-4.

272.	Guy, W., *ECDEU assessment manual for psychopharmacology*. 1976: US Department of Health, Education, and Welfare, Public Health Service ….

273.	Piacentini, J., et al. *Measuring impairment in childhood OCD: Psychometric properties of the COIS*. in *Proceedings of the American academy of child and adolescent psychiatry meeting*. 2001.

274.	Marcks, B.A., et al., *Longitudinal course of obsessive-compulsive disorder in patients with anxiety disorders: a 15-year prospective follow-up study.* Comprehensive psychiatry, 2011. **52**(6): p. 670-677.

275.	Storch, E.A., et al., *Quality of life in children and youth with obsessive-compulsive disorder.* Journal of child and adolescent psychopharmacology, 2018. **28**(2): p. 104-110.
